# Efficacy of Melflufen in Patients with Relapsed/Refractory Multiple Myeloma and Mutated or Deleted *TP53*

**DOI:** 10.1101/2024.12.02.24318289

**Authors:** Klara Acs, Juho J. Miettinen, Philipp Sergeev, Tobias Heckel, Yumei Diao, Kristina Witt-Mulder, Marcus Thureson, Thorsten Bischler, Maiju-Emilia Huppunen, Jakob Obermüller, Umair Munawar, Ana Slipicevic, Ralf C. Bargou, Fredrik Lehmann, Stefan Svensson Gelius, Stefan Norin, Fredrik Schjesvold, Pieter Sonneveld, Thorsten Stühmer, Caroline A. Heckman

**Author notes:** K.A., J.J.M and P.S. contributed equally to this study.

## Abstract

**Background:** Patients with relapsed/refractory multiple myeloma (RRMM) and high-risk genetic abnormalities such as del(17p) and *TP53* mutation have poor response to standard therapies and shorter survival compared to patients without these aberrations. Here, we investigated the activity and mechanism of action of peptide-drug conjugate melphalan flufenamide (melflufen) in *TP53* wild type (*TP53*wt) and mutant (*TP53*mut) myeloma models and assessed the efficacy of melflufen in patients with del(17p) and/or *TP53* mutation.

**Methods:** We evaluated melflufen activity *ex vivo* in 24 myeloma bone marrow (BM) samples and explored indicators of response from single cell RNA sequencing (scRNAseq) profiles. The efficacy of melflufen *vs.* control treatments was further investigated in *TP53*^−/−^ and parental *TP53*wt myeloma cell lines. DNA damage, apoptosis kinetics, mitochondrial function, plus transcriptomic and metabolic data were analyzed to understand the mechanisms responsible for melflufen activity in the absence of p53. Patient outcome data from the OCEAN phase III clinical trial (NCT03151811), which investigated the clinical activity of melflufen in RRMM, were statistically analyzed to assess the impact of del(17p) and *TP53* mutation on clinical response.

**Results:** BM plasma cell (PC) response to melflufen was independent of *TP53* mutation status, with melflufen active in del(17p), *TP53*mut, and *TP53*wt samples. Differential analysis of scRNAseq data demonstrated that melflufen sensitive PCs had lower expression of p53 target genes and higher expression of genes associated with DNA damage repair and cell cycle checkpoints. Analysis of *TP53*^−/−^ and *TP53*wt cell lines showed superior efficacy of melflufen in comparison to melphalan or cyclophosphamide. In the presence and absence of functional p53, melflufen robustly induced apoptosis, DNA damage, and mitochondrial dysfunction. In *TP53*^−/−^ cells, melflufen treatment led to distinct changes in expression of genes associated with cell cycle checkpoint and apoptosis, which were not observed with melphalan treatment. Notably, post-hoc analysis of the OCEAN trial del(17p) patient population demonstrated favorable progression free survival in the del(17p) subgroup treated with melflufen plus dexamethasone compared to the pomalidomide plus dexamethasone arm.

**Conclusions:** Our insights into the molecular mechanisms of melflufen activity in *TP53*mut myeloma support its clinical efficacy and application in the del(17p) and *TP53*mut patient population.

**Trial registration:** NCT03151811, registration 2017-05-09.

## BACKGROUND

The number of targeted therapies developed in the last decades has enormously increased the treatment possibilities for patients with multiple myeloma (MM) and has prolonged life expectancies.^1–4^ However, after initial response to early line therapies, patients suffer from multiple relapses and eventually develop resistance as the disease progresses. In current clinical practice, therapy regimens that include proteasome inhibitors (PIs), immunomodulatory drugs (IMIDs), monoclonal antibodies (mAbs), and dexamethasone (Dex) are commonly used as early line treatments.^5^ The development of resistance to these therapies has resulted in a patient population with heavily pre-treated relapsed/refractory MM (RRMM). As the disease progresses, patients develop enhanced clonal heterogeneity, with their tumor genome often accumulating more mutations and structural changes. Patients belonging to this population have benefited only modestly from the approval of novel targeted drugs and have very limited treatment options.^6–9^

The loss of parts of the short arm of chromosome 17 (del(17p)) in myeloma is one of the most important prognostic factors for patient response. The *TP53* gene is located at chromosome 17p13.1 and encodes the p53 tumor suppressor protein, which is important for maintaining genome integrity and initiation of apoptosis.^10^ Patients with del(17p) are assigned high-risk status according to the International Myeloma Working Group (IMWG) and based on several studies.^11–13^ In case a deletion of the chromosome 17 small arm is associated with *TP53* mutation on the remaining allele, the patients are grouped into the so-called “double-hit” population with very poor survival.^14^ The reason for the worse outcome is that no functional p53 protein remains in the tumor cells of these patients. However, del(17p) alone also confers poor outcome compared with patients without this deletion, and del(17p) cytogenetics is considered as a prognostic indicator for poor outcome.^14–21^

Melphalan flufenamide (melflufen) is a novel peptide-drug conjugate recently approved by the European Medical Agency (EMA) for the treatment of RRMM.^22^ The lipophilicity of melflufen facilitates rapid cellular uptake and the antineoplastic activity of the drug depends on high expression of aminopeptidases in the malignant myeloma cells.^23^ After hydrolysis by aminopeptidases, a robust increase of the alkylating metabolites occurs inside the tumor cells leading to induction of apoptosis with remarkably fast kinetics.^22–27^

In the OCEAN trial, a randomized, head-to-head, open-label, phase III clinical study, melflufen combined with dexamethasone (mfldex) or pomalidomide plus dexamethasone (pomdex) was studied in patients who had received 2-4 lines of previous treatments, were refractory to lenalidomide and their last treatment, and had not previously received pomalidomide treatment. The primary endpoint was progression free survival (PFS), and key secondary endpoints included overall survival (OS) and overall response rate (ORR). Results from the trial showed that patients on the melflufen arm had superior PFS compared to the pomalidomide arm patients.^28,29^ In a post-hoc analysis, patients on the melflufen arm that either had no prior ASCT or progressed >36 months after a prior ASCT had a better PFS and OS than patients progressing <36 months after an ASCT.^30^

The robust cellular efficacy of melflufen led us to compare the mechanism of action in different myeloma models with or without a mutated *TP53* genetic background, including bone marrow (BM) CD138+ plasma cells (PCs) from RRMM patients positive for del(17p)/*TP53* mutation, isogenic myeloma cell lines with double-mutated *TP53*, and patients with the same background enrolled in the OCEAN trial.

## MATERIALS AND METHODS

### Compounds and stock solutions

Melflufen (Oncopeptides AB) was freshly prepared in DMSO before application. Melphalan (cat. no. M2011, Sigma-Aldrich) stock solution was prepared in acidified ethanol. Cyclophosphamide (N,N-bis(2-chloroethyl)-4-hydroperoxy-2-oxo-1,3,25-oxazaphosphinan-2-amine,TRC Canada) was freshly prepared in DMSO.

### *Ex vivo* drug sensitivity testing of myeloma bone marrow samples

Viably cryopreserved BM mononuclear cell (BM-MNC) samples (n=24) from 23 patients with MM were obtained from the Finnish Hematology Registry and Biobank (Table 1). The samples were collected after informed consent using approved protocols and in accordance with the Declaration of Helsinki. The study was approved by an ethical committee of the Helsinki University Hospital (Ethical Committee Statement 303/13/03/01/201, latest amendment 7 dated 15 June 2016; latest HUS study permit HUS/395/2018 dated 13 February 2018; Permit numbers 239/13/03/00/2010, 303/13/03/01/2011). For *ex vivo* drug sensitivity testing, BM-MNCs were thawed, resuspended in conditioned medium from the HS-5 human BM stromal cell line, and seeded on 96-well plates. Resuspended drugs were added to the wells and cells incubated at 37°C for 72h. Labeled antibodies specific for CD138 and CD38 (BD Biosciences) were used to detect plasma cells, while Annexin V and 7AAD (BD Biosciences) were used to detect apoptotic and dead cells, respectively. The cells were analyzed on the iQue PLUS flow cytometer (Sartorius). Additional details are included in the Supplementary Materials and Methods.

### Single-cell gene expression analysis

For single cell RNA sequence (scRNAseq) analysis, the above-mentioned 24 BM-MNCs were sorted based on CD138+ expression and libraries prepared with 10x Genomics reagents and sequenced on the Illumina NovaSeq 6000 instrument. The samples were grouped based on *ex vivo* drug sensitivity to melflufen and melphalan. Cell clusters were identified based on gene expression patterns and differential gene expression (DGE) and gene set enrichment analysis (GSEA) performed on the plasma cell clusters of the highly sensitive and low sensitive samples for both drugs (see Supplementary Materials and Methods for details). Data link NCBI GEO (http://www.ncbi.nlm.nih.gov/geo) accession number GSE263201.

### Cell lines, cell culture, and cytotoxicity

The AMO-1 cell line was obtained from DSMZ (Braunschweig, Germany). Generation and characterization of *TP53* deficient AMO-1 clones is described in Munawar et al., 2019.^31^ Cell viability was determined after 72h incubation using the CellTiter-Glo® 2.0 assay (Promega) or AlamarBlue^TM^ reagent (Invitrogen) according to the manufacturers’ protocols.

### DNA damage signaling and cell apoptosis analyses

AMO-1 *TP53*wt and AMO-1 *TP53*^−/−^ cells were treated with melflufen, melphalan or cyclophosphamide at the indicated concentrations and time points. H2AX phosphorylation at Ser139(γ-H2AX) for DNA damage signal was analyzed using flow cytometry. For apoptosis analysis cells were stained for Annexin V APC and PI (Invitrogen). All samples were analyzed with the FACSCanto II flow cytometer (BD Biosciences). Data were analyzed using FlowJo software (BD Biosciences).

### JC-1 mitochondria membrane potential assay

AMO-1 *TP53*wt and AMO-1 *TP53*^−/−^ cells were treated with melflufen, melphalan or cyclophosphamide at the indicated concentrations and time points. The JC-1 mitochondrial membrane potential assay was performed according to the manufacturer’s protocol (MitoProbe™ JC-1 Assay Kit for Flow Cytometry (M34152), Molecular Probes).

### RNA preparation and RNA sequence analysis

AMO1 *TP53*wt cells or the *TP53^−/−^* knock out clonal subline AMO-1 *TP53*^−/−^ were treated with melphalan or melflufen at different concentrations and timepoints. RNA was isolated according to the RNeasy Plus protocol, quantified with Nanodrop (Thermo Scientific), and quality assessed with the Agilent 2100 Bioanalyzer. DNA libraries were prepared using the TruSeq Stranded mRNA Library Preparation Kit (Illumina) and sequencing performed on the NextSeq-500 platform (Illumina). Sequences were mapped to the human genome (GRCh38.p14) with STAR aligner and gene level based read counts generated with RefSeq. Differential gene expression (DGE) analysis was performed using DESeq2 with log-fold change shrinkage using beta Prior. Cluster profiler was used for gene set enrichment analysis (GSEA) and for KEGG pathway analysis. R was applied to generate heatmaps. Data link NCBI GEO (http://www.ncbi.nlm.nih.gov/geo) accession number GSE254959.

### Statistical analyses of the OCEAN trial data

Primary analysis of tumor response and progression-dependent endpoints of patients enrolled in the OCEAN trial (NCT03151811) were based on response assessments by an Independent Review Committee. All tumor response and progression-dependent endpoints were assessed using the IMWG Uniform Response Criteria (IMWG-URC). PFS was defined as time (months) from date of randomization to either confirmed disease progression or death due to any cause. ORR was defined as the proportion of patients for whom the best overall confirmed response was stringent complete response (sCR), complete response (CR), very good partial response (VGPR), or partial response (PR). Posthoc analyses were performed to examine associations between PFS and chromosome 17 deletion status and/or *TP53* gene mutation status. Furthermore, another post-hoc analysis was carried out to study the patient responder status in the chromosome 17 deletion population.

### Assessment of del(17p) status

The del(17p) status had been determined from bone marrow at baseline level (entry) of the clinical study by interphase Fluorescence In-Situ Hybridization (iFISH) in accordance with Good Clinical Practice (GCP). No cutoff applied, all patients with del(17p) cytogenetics had been included into the analyses. The presence of del(17p) in plasma cells varied between 8-90%. The del(17p) status of the patient samples used for the *ex vivo* drug sensitivity testing was determined using iFISH as previously described.^32^

### Assessment of *TP53* mutation status

*TP53* mutation status of OCEAN trial patients was determined by next generation sequencing (NGS) using Almac Genomic Services. Briefly, DNA was extracted from BM derived CD138+ cells and processed using Illumina TruSeq Custom Amplicon Kit Dx. A custom *TP53* oligo panel detecting pan-cancer single nucleotide polymorphisms was applied and the samples sequenced on the Illumina MiSeq instrument. All detected single nucleotide variants classified. The *TP53* mutation status of the patient samples used for the *ex vivo* drug sensitivity testing was determined using exome sequencing of DNA from CD138+ cells as previously described.^32^

Detailed materials and methods are provided in the supplementary information.

## RESULTS

### Melflufen targets MM plasma cells *ex vivo* regardless of *TP53* mutation status

To assess the efficacy of melflufen in high-risk myeloma *ex vivo*, we conducted flow cytometry-based drug sensitivity testing of 24 myeloma BM-MNC samples obtained from 23 patients with various clinical and molecular backgrounds (Figure 1A). The cohort included samples from newly diagnosed and relapsed patients. After treatment of the BM-MNCs with melflufen or control drugs for 72h, the viability of the CD138+CD38+ cells was assessed by Annexin V and 7AAD staining. All samples showed reduced viability after exposure to melflufen, but sensitivity varied between samples from extremely sensitive to less sensitive, which was reflective of the heterogeneity of the patients (Figure 1B). For subsequent analyses, the samples were divided into different melflufen sensitivity groups based on the melflufen DSS values of the samples: high (HS, DSS>40; n=8), intermediate (IS, 40<DSS<31; n=8), and low (LS, DSS>31; n=8). Confirming our earlier observations, a significant indicator of melflufen sensitivity was disease stage as all but one of the HS samples were from RRMM patients.^26^ A gain of chromosome 1q (+1q) and loss of chromosome 13 (−13/−13q) were the most frequent abnormalities observed in 22/24 samples individually or in combination. Statistical analysis showed tendencies of samples with +1q to have higher sensitivity to melflufen, and the opposite trend for samples with −13/−13q although significance was not obtained due to the limited sample number. In addition, 4 of the 8 LS samples and only 1 of the 8 HS samples had t(4;14), while 3 of the HS, 1 of the IS and 2 of the LS samples were positive for del(17p). *TP53* mutation status was available for 14 of the 24 tested samples. Of the 5 HS samples with *TP53* sequence data, 3 were mutation positive, while *TP53* mutations were not detected in any of the 5 LS samples with sequence information. These results suggested that melflufen was highly active despite the presence of *TP53* mutation or del(17p). At the same time, comparison of the *ex vivo* drug sensitivity of samples to melflufen, melphalan, and cyclophosphamide clearly showed the superior potency of melflufen in *TP53*wt, del17p, and del17p + *TP53*mut patient samples (Figure 1C). For instance, the sensitivity to melflufen was significantly higher in the *TP53*wt (melflufen: n = 8, melphalan: n = 8, cyclophosphamide: n = 2) and del(17p) + *TP53*mut groups (melflufen: n = 4, melphalan: n = 4, cyclophosphamide: n = 2). Although the threshold for statistical significance was not achieved in the del(17p) cohort (n = 2 for all drugs), the trend was similar.

**Figure 1.**
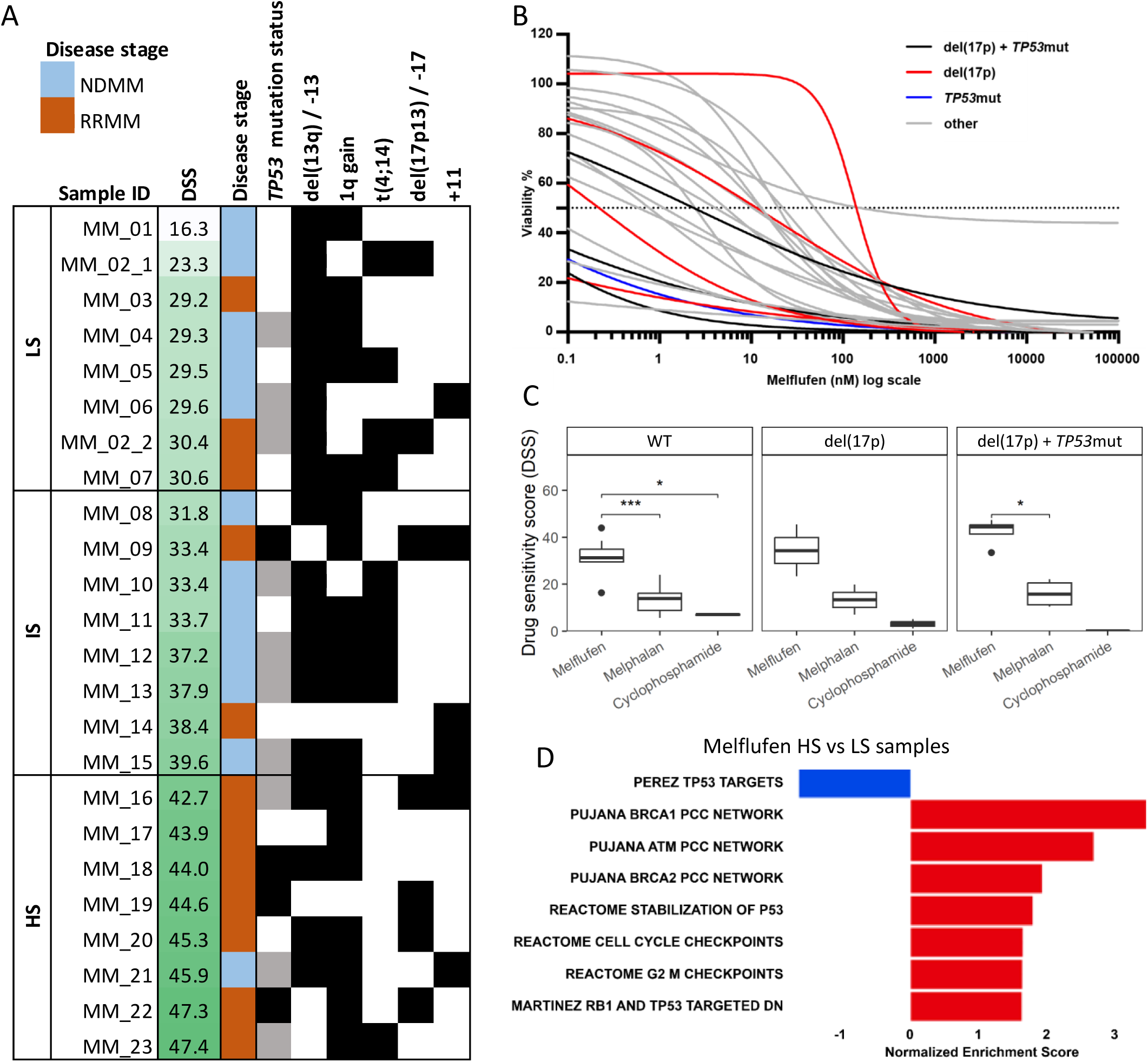
Melflufen *ex vivo* drug sensitivity testing results from the 24 myeloma bone marrow samples. **A)** Patient sample characteristics. **B)** Melflufen dose response curves for the 24 MM patient samples. Samples having both del(17p) and *TP53* mutation (black), having only del(17p) (red), having only *TP53* mutation (blue), other (grey). **C)** Drug sensitivity scores of samples with different molecular background for three alkylators – melflufen, melphalan, and cyclophosphamide. P-values are indicated above the brackets, with the corresponding comparison. * – p<0.05, *** – p<0.001. **C**) Bar plot representing normalized enrichment scores for selected gene sets derived from the most differentially expressed genes in the plasma cell populations between the melflufen high sensitive (HS) and low sensitive (LS) groups.

To investigate molecular differences between groups with different sensitivity to melflufen, we analyzed the transcriptomic profiles of the plasma cell populations, obtained by scRNA sequencing, and conducted differential gene expression (DGE) and gene set enrichment (GSEA) analyses. GSEA of genes detected in HS and LS samples showed a decrease in p53 downstream pathways and targets in the HS group, and, at the same time, higher enrichment of pathways associated with DDR genes *BRCA1*, *ATM*, and *CHEK2* (Figure 1D).

### Loss of *TP53* compromises multiple pathways

To better understand the mechanisms responsible for the superior activity of melflufen compared to other standard of care alkylators observed from the *ex vivo* drug sensitivity testing analysis, we used the myeloma cell line AMO-1 with intact WT *TP53* or with deletion of both alleles (*TP53*^−/−^). To validate the functional significance of these cell line models and their relevance with the patient samples, we investigated transcriptomic alterations caused by mutation of *TP53* by comparing baseline gene expression of *TP53*^−/−^ *vs*. *TP53*wt patient samples and cell lines. This analysis yielded 1485 differentially expressed genes in patient samples (Supplementary Table S1) and 5357 of those in cell lines (Supplementary Table S2). In both comparisons, there were 252 similarly deregulated genes, such as *IL6R*, *JUN*, *DUSP5*, *AREG*, and *FOS* (Supplementary Table S3). Subsequently, gene set enrichment analysis (GSEA), when applied to these cohorts of genes, gave us multiple gene sets, associated with *TP53* and cell cycle progression, as well as pathways, associated with signaling and DNA damage (Supplementary Figure S1). These results confirmed a similar impact of *TP53* dysfunction in the patient samples and cell line models and illustrated the exploitability of *TP53* loss for treatment purposes by a drug such as melflufen.

### Melflufen is cytotoxic and induces early apoptosis in cells without functional p53

Earlier studies had demonstrated that deletion of one or both alleles of *TP53* in AMO-1 cells by CRISPR/Cas9 resulted in significant loss of sensitivity to melphalan or doxorubicin.^31,33^ In support of the earlier report,^33^ double mutant *TP53*^−/−^ AMO-1 cells displayed profound loss of sensitivity to melphalan with EC50 values close to 20µM (Figure 2A, B, Supplementary Table S4). However, melflufen was highly active towards both *TP53*wt and *TP53*^−/−^ AMO-1 cells and compared to melphalan exhibited 30-fold more inhibition of cell viability with EC50 0.6μM. Importantly, melflufen-induced apoptosis as measured by Annexin V was similar in *TP53*wt and *TP53*^−/−^ cells (Figure 2B).

**Figure 2.**
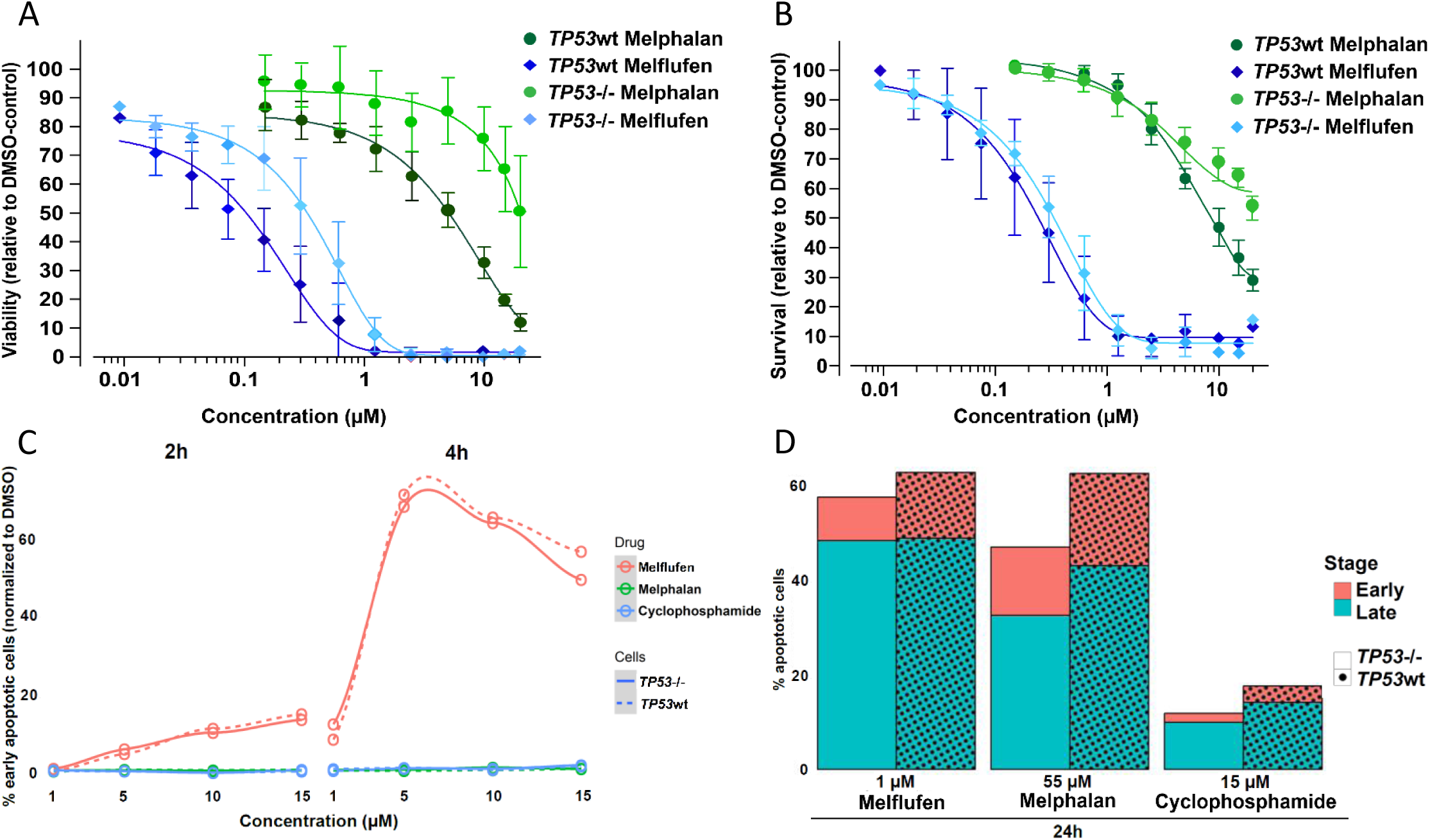
Cytotoxicity and apoptosis detection in the AMO-1 *TP53* isogenic cell lines. **A, B)** Melflufen and melphalan cytotoxicity dose effect curves in parental AMO-1 cells (*TP53*wt) and in its bi-allelic-double mutant *TP53* gene harbouring clonal cell line (*TP53*^−/−^) using either the alamarBlue metabolic activity assay (A) or the apoptosis measurement Annexin V/propidium iodide staining (B). The curves represent three independent experiments, error bars reflect standard deviations. **C)** Melflufen, melphalan or cyclophosphamide induced early apoptosis in AMO-1 *TP53*wt and AMO-1 *TP53*^−/−^ isogenic cell lines. Cells were treated with the indicated drug concentrations and time points, melflufen (red), melphalan (green), or cyclophosphamide (blue). Early apoptosis was detected with Annexin V and PI staining using flow cytometry (Annexin V positive, PI negative). **D)** Both early and late apoptosis were measured in AMO-1 *TP53*wt and AMO-1 *TP53*^−/−^ isogenic cell lines after 24h treatment with the indicated drugs and drug concentrations (late apoptotic cells: Annexin V positive, PI positive).

Earlier studies have shown that melflufen has more efficient kinetics compared to melphalan,^34,35^ we therefore investigated the induction of apoptosis in *TP53*wt and *TP53*^−/−^ cells after treatment with melflufen or melphalan. Even at 2h, induction of apoptosis was apparent in both *TP53*wt and *TP53*^−/−^ cells after melflufen but not melphalan treatment (Figure 2C). At 4h, early induction of apoptosis was as prominent in the mutant *TP53*^−/−^ AMO-1 cells as in *TP53*wt cells. In contrast to melflufen, melphalan and cyclophosphamide failed to induce early apoptosis (Figure 2D), although the apoptotic signal increased later and at higher IC50 values (55μM and 15μM, respectively) compared to melflufen (IC50 1μM) (Figure 2D). Overall, these results demonstrate that melflufen treatment resulted in complete inhibition of viability and induction of apoptosis at sub-to low micromolar concentrations, and indicate that, in contrast to melphalan, melflufen remains active despite compromised p53 function.

### Melflufen induces DNA damage and mitochondrial membrane disruption in *TP53* deficient cells

As the main cellular target for alkylating agents is genomic DNA, we aimed to gain insight into the kinetics of DNA damage induction for cells harboring *TP53*wt or *TP53*^−/−^ cells. Cells were treated with either melflufen, melphalan or cyclophosphamide and induction of early DNA damage was measured by phosphorylation of H2AX (γH2AX (ser139)). Melflufen treatment induced yH2AX signal after 30 min in the *TP53*wt cells in a concentration dependent manner, while in the *TP53*^−/−^ mutant cells maximal DNA damage induction occurred already at the lowest concentration. *TP53*^−/−^ cells exhibited a higher baseline level of yH2AX which is in line with loss of p53 protein function in the feed-back loops for DNA replication and repair protein functions.^10^ Exposure to melphalan resulted in little induction of early DNA damage in *TP53*^−/−^ cells while in *TP53*wt cells DNA damage induction was apparent only at a 10-fold higher concentration compared to melflufen. Similarly, there was little change in γH2AX levels in *TP53*^−/−^ cells treated with cyclophosphamide, and the yH2AX signal induction by cyclophosphamide in the *TP53*wt cells appeared later in comparison to the other drugs (Figure 3A).

**Figure 3.**
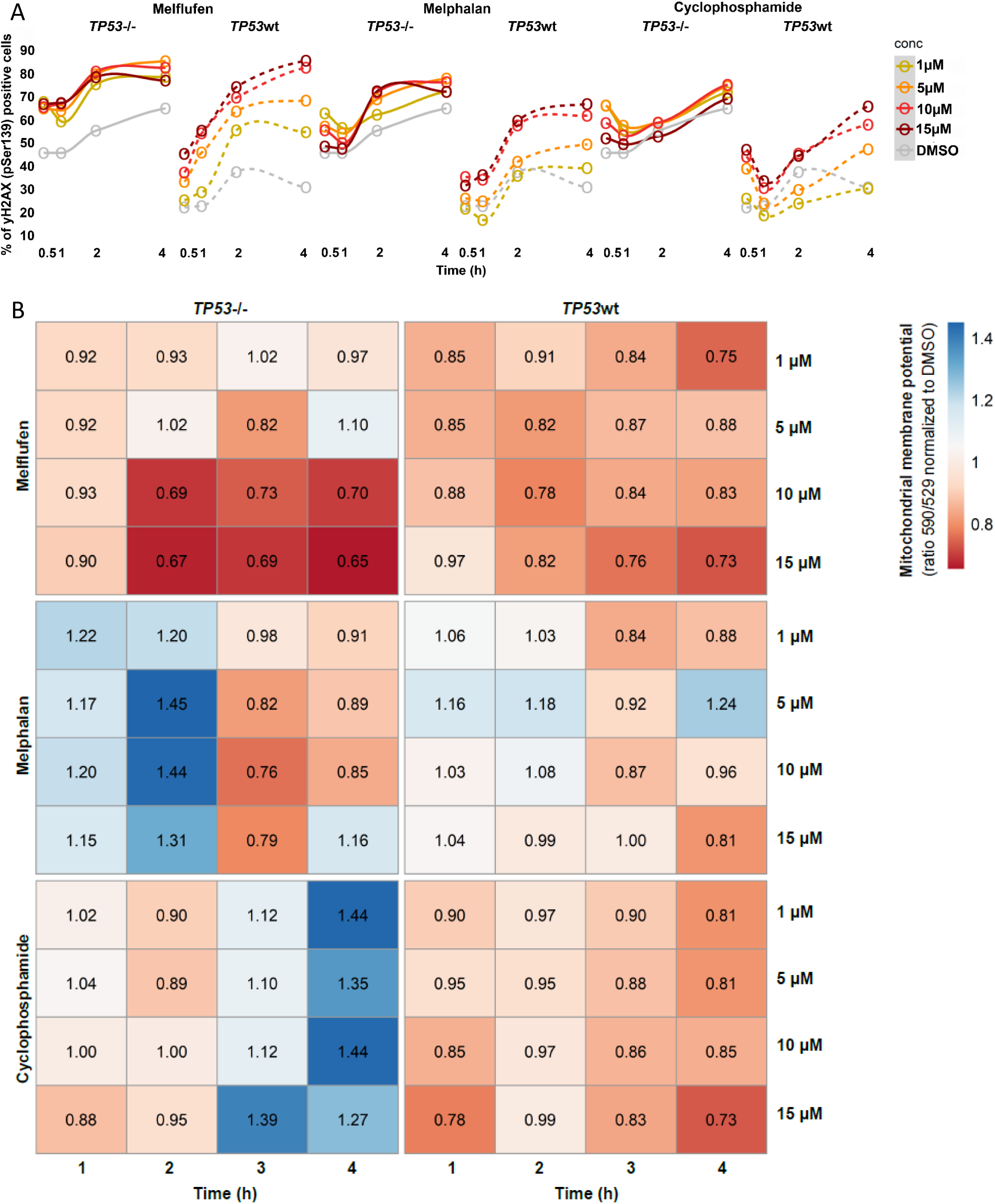
DNA damage and mitochondrial membrane potential detection in AMO-1 *TP53*wt and *TP53* knock out cells. **A)** Melflufen, melphalan or cyclophosphamide induced DNA damage in AMO-1 *TP53*wt and AMO-1 *TP53*^−/−^ isogenic cell lines. Early DNA damage was detected as yH2AX signal by flow cytometry after the indicated treatment concentrations and time points. yH2AX: phosphorylated histone protein 2AX. **B)** Melflufen, melphalan or cyclophosphamide treatment induced mitochondrial disfunction in AMO-1 *TP53* isogenic cell line. Melflufen, melphalan or cyclophosphamide treatment were applied in the indicated concentrations, and after 1, 2, 3 and 4h time points mitochondrial function of the treated cells was measured using JC-1 mitochondrial membrane potential assay. PI: propidium iodide.

We further investigated whether other cellular targets such as mitochondria^27^ are affected by melflufen with similar efficacy in the *TP53*wt and *TP53*^−/−^ cells. Mitochondrial functionality by membrane integrity measurements of the AMO-1 isogenic cell lines showed that melflufen induced a drop of membrane potential in the *TP53*wt cells within 2 hours after treatment, and the effect on mitochondria function was even more pronounced in the mutant *TP53*^−/−^ cells (Figure 3B). In contrast, melphalan and cyclophosphamide had no measurable effect on mitochondrial function in *TP53*^−/−^ cells at the 4h treatment timepoint. Altogether, these data demonstrate that the activity of melflufen compared to other alkylators is associated with more efficient induction of cellular DNA damage and mitochondrial membrane disruption, and that melflufen can induce these processes with or without functional p53.

### Gene expression analysis reveals differences in *TP53*wt *vs*. *TP53*^−/−^ cells after treatment with melflufen and melphalan

To understand if the rapid kinetics of melflufen intracellular accumulation and DNA damage induction uniquely influenced gene expression patterns compared to melphalan, RNA was collected from AMO-1 *TP53*wt and *TP53*^−/−^ cells after 2h exposure to the IC90 concentrations of melflufen or melphalan (Supplementary Table S4) and sequenced. Gene expression analysis showed distinct differences in the p53 pathway between melflufen and melphalan treatment of the *TP53*^−/−^ AMO-1 cells that were not apparent in *TP53*wt cells (Figure 4A). For example, melflufen treatment of the *TP53*^−/−^ cells resulted in significant upregulation of genes including the cyclin dependent kinase inhibitor 1A (*CDKN1A*), cyclin G2 (*CCNG2*), and DNA damage inducible alpha, beta, and gamma genes (*GADD45A, B* and *G*), as well as upregulation of the apoptosis inducing *PMAIP1* (NOXA) gene. At the same time melflufen treatment caused significant downregulation in apoptosis inhibition and stress response genes including the well-known *BCL2* and *BCL2L1* genes. The primary target of alkylators is genomic DNA and treatment results in the upregulation of DNA damage repair (DDR) signaling as a cellular response to genomic stress. Importantly, the p53 protein is a key factor in the maintenance of DNA repair protein homeostasis^10^, and base excision repair (BER) and nucleotide excision repair (NER) are the main DNA repair pathways activated upon alkylator induced DNA damage.^36^ We therefore investigated genes belonging to these pathways and analyzed their expression levels (Figure 4B and Supplementary Figure S2). In the DNA replication pathway analysis, these genes tended to be upregulated in *TP53*^−/−^ cells after melflufen treatment. Besides well-known replication guarding genes such as *RPA1*, *POLD1,3,4* and *POLE2,3*, the upregulation of a cluster of genes that includes several micro-chromosome maintenance genes (*MCM7*, *MCM3*, *MCM5*) was also observed. Importantly, BER associated genes such as *PARP1*, *PARP2*, *RCF4* and *XRCC1*, and NER pathway genes including the hallmark ERCC genes (*ERCC1,2,5*) and *RAD23A* were upregulated in *TP53*^−/−^ cells after melflufen but not melphalan treatment. The differential gene expression patterns between melflufen and melphalan treated *TP53*^−/−^ cells were also apparent after 12h incubation with drug (Supplementary Figure S2). In contrast, there was little difference in the gene expression patterns induced by melflufen and melphalan in *TP53*wt cells at 2 or 12h. The distinct gene expression patterns induced by melflufen in the *TP53*^−/−^ cells compared to melphalan suggest a clear mechanistic difference between the two alkylators.

**Figure 4.**
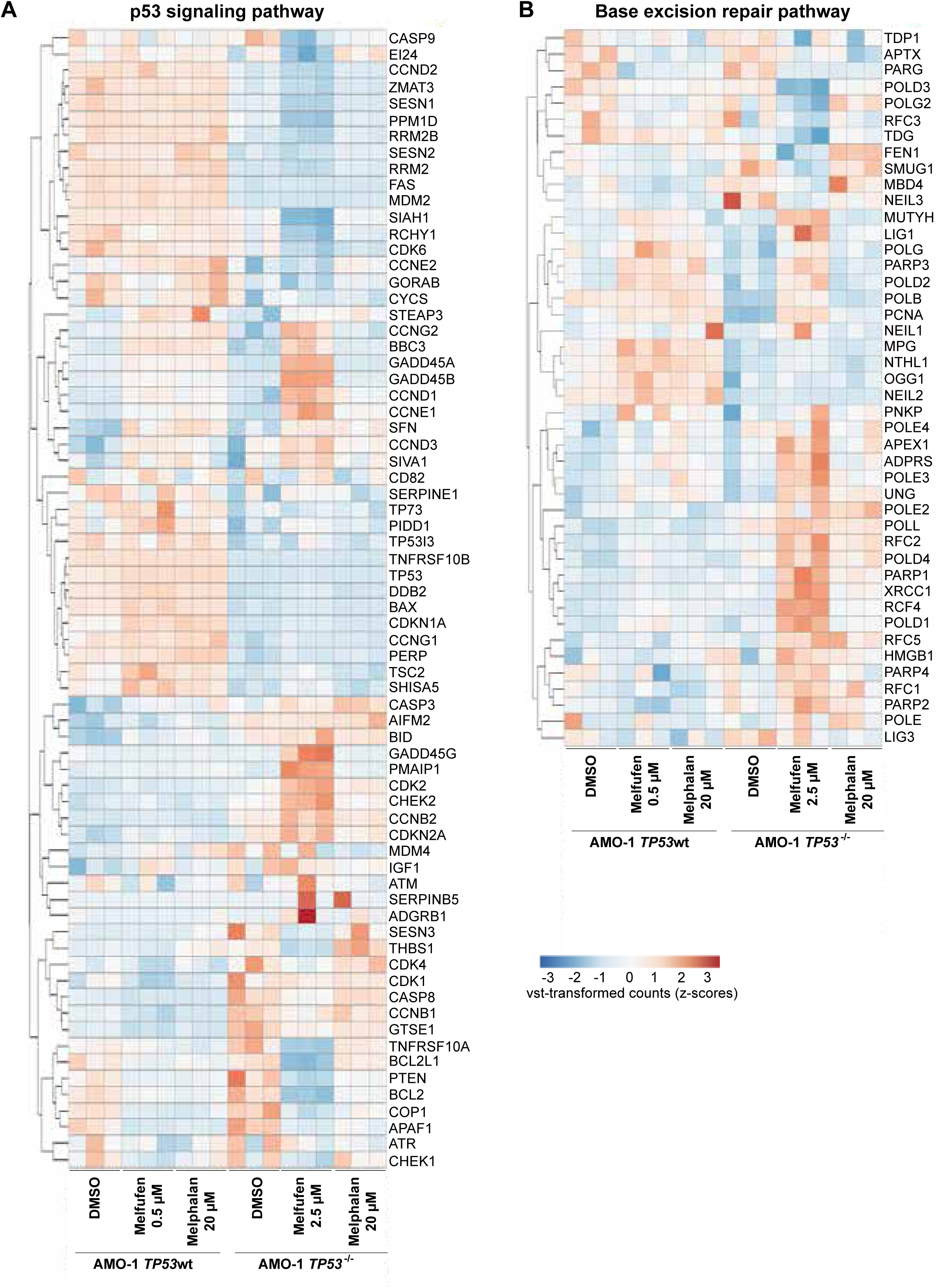
Gene expression analysis in *TP53* wild type versus *TP53* mutant genetic background AMO-1 cells. **A, B)** Heatmaps of gene expression of p53 signalling (A) and the Base Excision Repair (BER) DNA repair pathway (B) genes after 2h treatment of *TP53*wt or *TP53^−/−^ cells* with 2.5 μM melflufen versus 20 μM melphalan, respectively.

### Patient PFS, OS and response in the del(17p) population

To confirm our preclinical observations and determine if melflufen is active in patients with dysfunctional *TP53*, we conducted a post-hoc analysis of the OCEAN head-to-head open label clinical trial where the primary endpoint was PFS in the mfldex *vs*. pomdex treatment arms. Earlier, the PFS of 6.8 months in the mfldex group was shown to be significantly better than the PFS of 4.9 months in the pomdex group (stratified log-rank p value: p=0.03; HR 0.792 [95% CI: 0.640, 0.981]).^28^ In the post-hoc analysis for patient responder status in the chromosome 17 deletion population study, 70 patients characterized with del(17p) were included in this analysis from the total enrolled 495 patients in the OCEAN trial. Notably, the del(17p) patient population size from the OCEAN trial is in line with the published and commonly detected del(17p) cytogenetics rate where del(17p) is detected in ∼10-15% from total MM trial populations.^18,37,38^

For PFS analysis of the del(17p) subpopulation, the patient distribution was as follows; in the mfldex arm 33 patients had del(17p) chromosomal aberration and 213 had no del(17p) while in the pomdex arm 37 patients were del(17p) mutant and 212 patients had no deletion (Supplementary Table S5). In the del(17p) subpopulation there was a trend towards improved PFS in the mfldex group with 7.1 months *vs*. 2.9 months in the pomdex group (HR 0.45 [95% CI: 0.26, 0.79], p=0.006) (Figure 5A). These data suggest that melflufen treatment is efficient in this high-risk subpopulation and can substantially prolong patient PFS. OS in the del(17p) subpopulation indicated a favourable OS in the pomdex arm versus mfldex arm, with 11.5 months in the mfldex arm and 15.9 months in the pomdex arm (HR 1.31 [95% CI: 0.76, 2.26], p=0.33) (Figure 5B). The different PFS and OS outcomes between the treatment groups (mfldex arm vs. pomdex arm) might be explained based on post-hoc subgroup analyses which indicated that OS outcomes may have been driven primarily by patients who had received previously high-dose melphalan followed by an autologous haematopoietic stem-cell transplantation (ASCT).^28^ These earlier post-hoc analyses determined that melflufen plus dexamethasone should be indicated for RRMM, but exclude patients with a prior ASCT, whose time to progression is less than 3 years from transplantation.^39^

**Figure 5.**
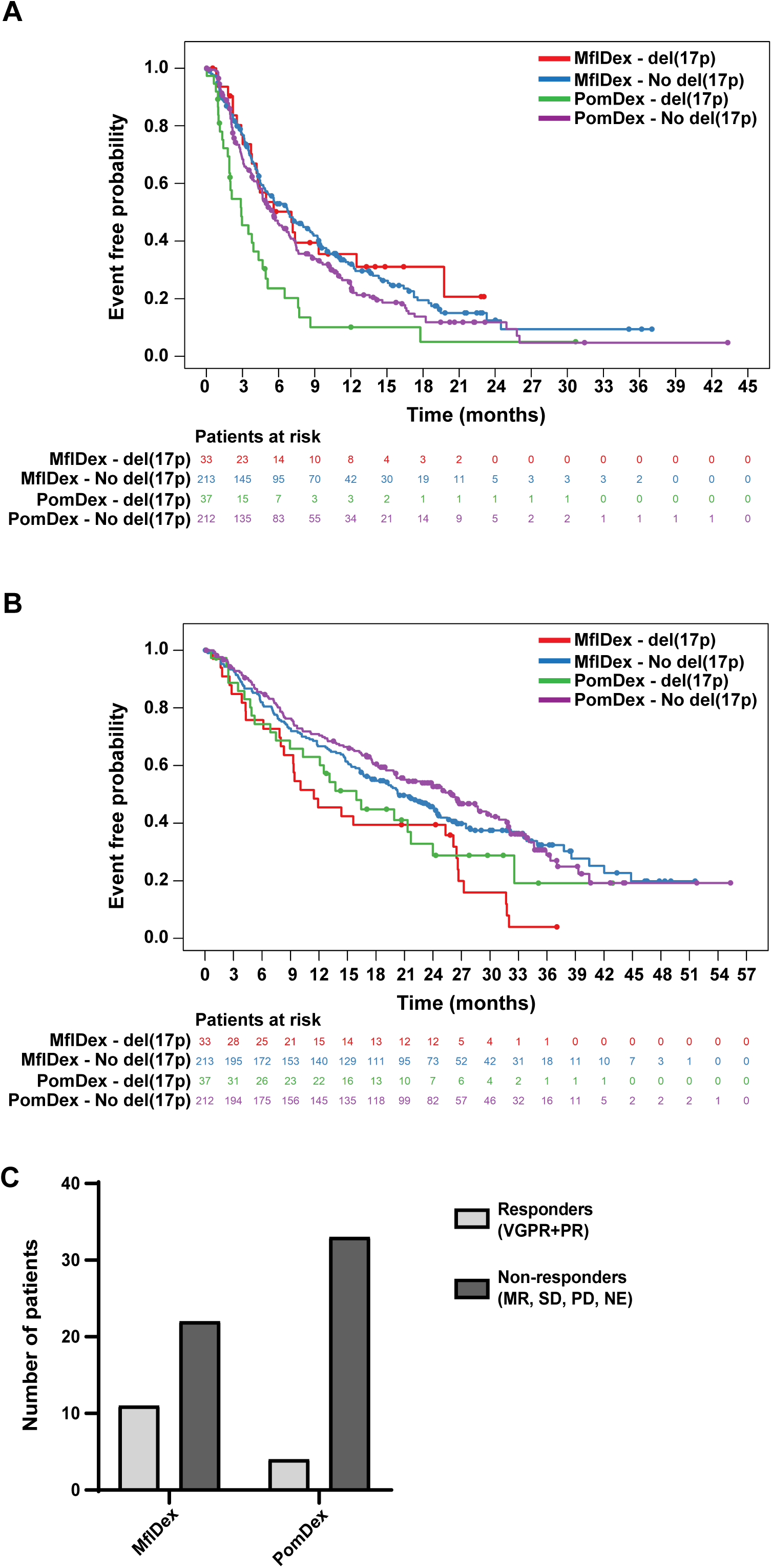
Progression-free survival, overall survival, and overall response rate in del(17p) patient population from the OCEAN trial. **A**) Kaplan-Meier progression-free survival curve, **B**) overall survival curve, and **C**) overall response rate for the investigator assessed all-treated del(17p) population. Analysis set of 70 del(17p) patients and 425 patients not harbouring del(17p). MflDex: melflufen + dexamethasone; PomDex: pomalidomide + dexamethasone; VGPR: very good partial response; PR: partial response; MR: minimal response; SD: stable disease; PD: progressive disease; NE: not evaluable

Based on analysis of response to treatment in the del(17p) subgroup, the ORR results are as follows; the mfldex arm had 33.3% ORR (11 responders from 33 patients) *vs*. pomdex which had an ORR of 10.8% (4 responders from 37 patients) (33.3% vs 10.8%, p=0.028) (Figure 5C). The observed ORR data in the del(17p) population showed that the mfldex treatment arm had a significantly higher number of responders compared to the pomdex treatment arm and suggest that melflufen treatment can induce responses in this high-risk population. In addition, the ORR to melflufen treatment in the del(17p) population was comparable to the ORR in the general patient population of the OCEAN trial, which included heavily pre-treated R/R MM patients. In the OCEAN study, the ORR for mfldex was 33% (95% CI, 27-39) *vs*. an ORR of 27% (95% CI, 22-33) for pomdex.

The del(17p) aberration often co-occurs with other high-risk cytogenetic abnormalities, which may also impact patient response to treatment and outcome. The distribution of these other high-risk changes within the del(17p) population and the outcome of these del(17p) subgroups are shown in Supplementary Table S6. The most common co-occurring high-risk cytogenetic abnormality with del(17p) was gain of 1q (+1q), which appeared to negatively impact response to both mlfdex and pomdex in the del(17p) population; however, statistical analysis of these further subdivided cytogenetic groups would have been underpowered.

### Melfufen shows favorable clinical efficacy in del(17p) and/or *TP53* mutation patients from the OCEAN trial

To better understand mfldex clinical efficacy in the population with dysfunctional *TP53*, which included a mixed population of patients comprised of del(17p) and non-del(17p) patients, we performed a NGS exploratory study to determine *TP53* gene mutation status for both mfldex and pomdex treatment arms. For both arms, most sequenced patient samples contained silent single nucleotide polymorphism alterations to *TP53* and thus retained p53 function. From the mfldex arm, 15 samples that were sequenced had del(17p) and/or contained a pathogenic mutation to *TP53*, while in the pomdex arms 18 patients were identified with del(17p) and/or pathogenic/loss-of-function mutation to *TP53*. Analyzing PFS in the sequenced population, the patient distribution was as follows; in the mfldex arm, 62 patients had no functional alteration to *TP53,* and 15 patients had either del(17p) and/or *TP53* pathogenic mutation. In the pomdex arm, 50 patients had no functional *TP53* impairment, and 18 patients displayed del(17p) and/or *TP53* pathogenic mutation (Supplementary Table S7). The PFS analysis showed superiority of mfldex treatment *vs*. pomdex (Figure 6A). In the impaired *TP53* group (del(17p) and/or *TP53*mut) the PFS in the mfldex group was 6.7 months *vs*. 4.7 months in the pomdex group (HR 0.57 [95% CI: 0.25, 1.29], p=0.2). These data indicate that in the OCEAN trial, mfldex treatment resulted in a better PFS in the *TP53* gene mutated patient population.

**Figure 6.**
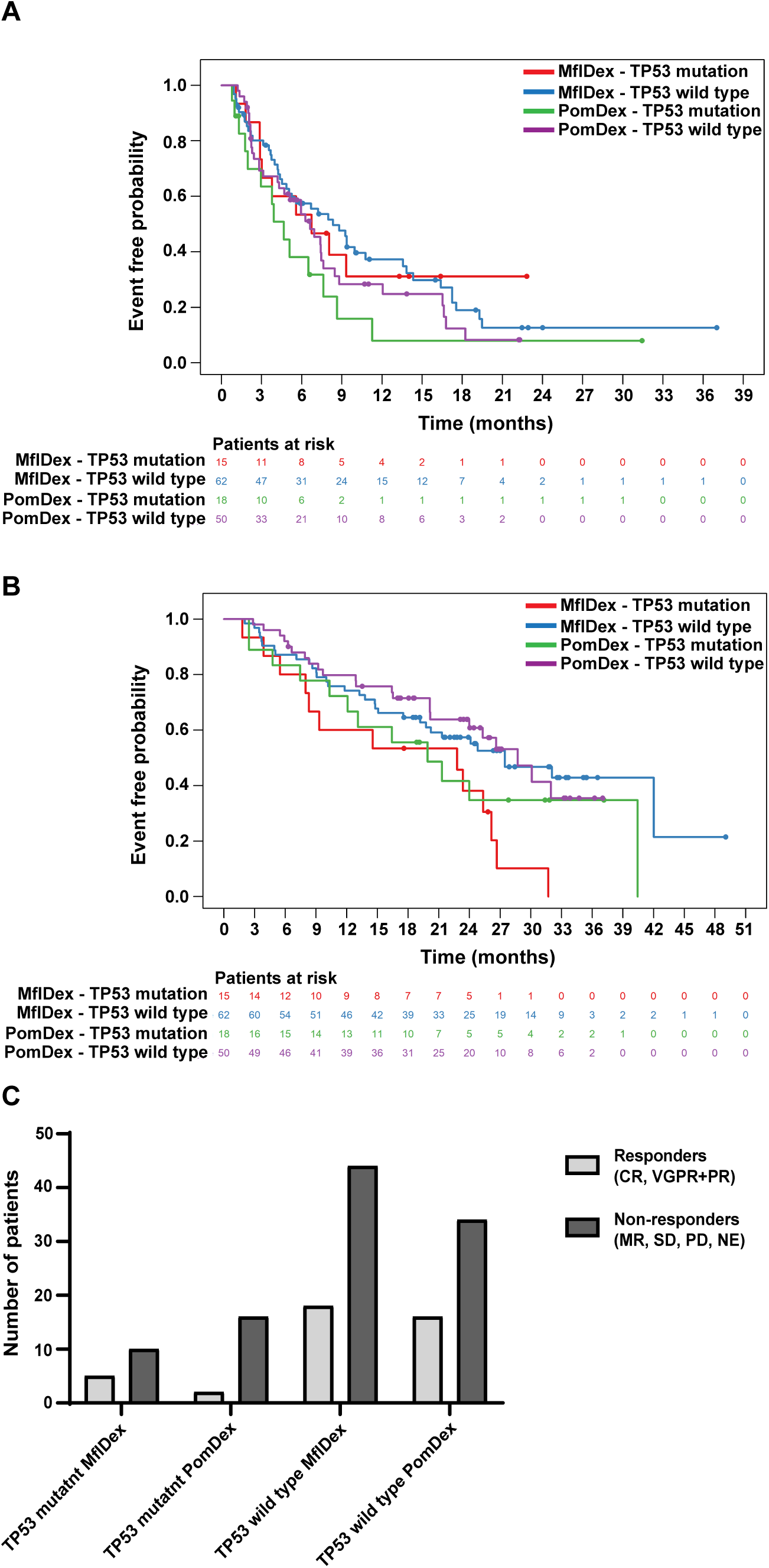
Progression-free survival, overall survival and overall response rate in del(17p) and/or *TP53* gene mutated patient population from the OCEAN trial. **A)** Kaplan-Meier progression-free survival curve, **B**) overall survival curve, and **C**) overall response rate for all del(17p) and/or *TP53* gene sequenced population. Analysis set of 33 del(17p) and/or *TP53* gene mutated patients and 112 patients not harbouring del(17p) or *TP53* gene mutation. MflDex: melflufen + dexamethasone; PomDex: pomalidomide + dexamethasone; CR: complete response; VGPR: very good partial response; PR: partial response; MR: minimal response; SD: stable disease; PD: progressive disease; NE: not evaluable

Additionally, OS analysis for the *TP53* sequenced subpopulation indicated not significant but prolonged OS in the mfldex arm *vs*. pomdex arm (Figure 6B) with 22.8 months *vs*. 19.9 months, respectively (HR 1.54 [95% CI: 0.69, 3.47], p=0.30).

Based on analysis of response to treatment in the *TP53* gene sequenced patient population, the ORR results are as follows; the mfldex arm had 33.3% ORR *vs*. pomdex which had an ORR of 11.1% (Figure 6C). The observed ORR data in the *TP53* gene sequenced population showed that the mfldex treatment arm had a higher number of responders compared to the pomdex treatment arm. Only 3 patients were double hit patient with del(17p) and *TP53* mutation. Their distribution was the following, 1 patient with partial response (PR) in the mfldex arm and 2 patients with no respond (stable disease (SD)) in the pomdex arm.

Notably, both post-hoc analyses (del(17p) population or the del(17p) and/or *TP53* mutation sequenced population) included the melflufen treatment target (>36 months post ASCT) and non-target populations (<36 months post ASCT).^38^ Inclusion of both populations resulted in a less favorable outcome for the mfldex arm. However further dividing the del(17p) or the del(17p) and/or *TP53* sequenced populations would result in very small patient groups with very limited statistical power.

## DISCUSSION

In myeloma, the sequential appearance of small mutations and larger chromosomal structural changes are acquired with treatment course and ultimately result in progressive disease driven by drug resistant clones. The deletion of chromosome 17p13 and mutation to *TP53* are the most deleterious aberrations described in myeloma and predict poor outcome and limited responses to most available therapies. Analysis of late line RRMM patients treated with lenalidomide and dexamethasone showed that del(17p) patients had low response rate and significantly shorter survival compared to patients without the deletion, indicating an unmet clinical need for this subpopulation of patients.^40^

Our earlier preclinical investigations of melflufen activity in myeloma indicated that plasma cells from relapsed/refractory patients were more sensitive than those from newly diagnosed patients.^26^ We confirmed those observations in this study but also demonstrated that melflufen is active in patient-derived plasma cells with del(17p) and/or mutated *TP53*. Although this cohort was too small to demonstrate statistical significance across all subpopulations, we found that melflufen was clearly more effective than standard of care alkylators melphalan and cyclophosphamide when tested against patient plasma cells with WT *TP53*, del(17p), or del(17p) plus mutated *TP53*. Although we lacked *TP53* mutation status for all the tested samples, comparative gene expression analysis showed decreased enrichment of gene sets associated with p53 downstream pathways and targets in samples highly sensitive to melflufen. These results indicated that melflufen retains its activity despite compromised p53 function.

Using isogenic myeloma cell lines with intact *TP53* or loss of both alleles, we were able to confirm our results from the patient samples and demonstrate that melflufen is highly effective in the presence or absence of functional p53. Although melflufen should have a similar mechanism of action as melphalan, our results showed that melflufen treatment resulted in rapid and robust induction of apoptosis in both *TP53*wt and *TP53^−/−^* cells compared to melphalan, which especially had reduced effect on *TP53*^−/−^ cells. In addition, melflufen rapidly induced DNA damage and disrupted mitochondrial function. These results further strengthened the applicability of melflufen as an alkylating drug in the RRMM population as several studies indicate increased mitochondria number and function with myeloma disease progression.^41–43^

Although the fast kinetics of melflufen may be in part due to its lipophilic profile,^22,24^ melflufen treatment was associated with unique differences in the gene expression profiles of treated cells. Notably, melflufen treatment of *TP53*^−/−^ cells resulted in increased expression of genes associated with DNA replication and DNA repair and downregulation of cell cycle checkpoint and anti-apoptosis genes (e.g., *CHEK1, PTEN, BCL2*, *BCL2L1*). These changes were not observed in melphalan treated *TP53^−/−^* cells indicating distinct mechanistic differences between these drugs.

Our preclinical data were supportive of post-hoc analyses of the OCEAN trial. The original analyses of the OCEAN study cohort showed that RRMM patients treated with the combination mfldex had a longer PFS compared to patients that received pomdex (6.8 months *vs*. 4.9 months).^28^ In this follow-up study, a similar difference in PFS for the two treatment arms was reflected in high-risk patients with del(17p) cytogenetics, where PFS of the mfldex group was superior to the pomdex group. The del(17p) patients treated with mfldex also had a greater ORR (33%) compared to patients that received pomdex (ORR 10.8%). Furthermore, the PFS for the del(17p) and/or *TP53* gene mutated subgroup in the mfldex arm was 6.74 months compared to 4.67 months for the pomdex arm. Although OS analyses showed prolonged survival of patients with *TP53* mutation who received mfldex compared to pomdex, the results were not statistically significant. In addition, OS was better for patients with del(17p) who had received pomdex compared to those receiving mfldex, but this also lacked significance. These analyses included all patients who had received high dose melphalan prior to ASCT, although the indication of the melflufen plus dexamethasone combination is for patients who progress >36 months after ASCT. Thus, inclusion of non-indicated patients who progressed <36 months after receiving high dose melphalan and ASCT could have affected the results. However, dividing the patients into even smaller subgroups would not have been statistically meaningful. Nevertheless, these results demonstrated that melflufen had superior efficacy in the del(17p) and *TP53* mutant patient population.

Although our preclinical studies did not include a comparison between melflufen and pomalidomide, we could demonstrate that melflufen had superior activity compared to that of the standard of care alkylators melphalan and cyclophosphamide, even without functional p53. In addition, the analyses of the OCEAN trial patient cohort have demonstrated better efficacy of the mfldex combination in heavily pretreated patients including those with del(17p) and *TP53* mutation. Future studies could investigate the impact of other genetic abnormalities or lines of treatment on response to melflufen.

## CONCLUSION

Our findings underline melflufeńs unique mechanism of action in which intracellular accumulation of the alkylating payload efficiently causes cytotoxic effects by rapid induction of DNA damage and mitochondrial dysfunction independent of *TP53* gene status and protein function. These results together with the post-hoc analysis of the OCEAN trial data demonstrate that melflufen is particularly effective in high-risk myeloma with del(17p) and/or *TP53* mutation.

## DECLARATIONS

### Ethics approval and consent to participate

Bone marrow (BM) aspirates were collected from multiple myeloma (MM) patients after informed consent using protocols approved by the ethical committee of Helsinki University Hospital (Ethical Committee Statement 303/13/03/01/201, latest amendment 7 dated 15 June 2016; latest HUS study permit HUS/395/2018 dated 13 February 2018; Permit numbers 239/13/03/00/2010, 303/13/03/01/2011) and in accordance with the Declaration of Helsinki.

### Consent for publication

Not applicable.

### Availability of data and materials

Bulk RNAseq data from AMO1 *TP53*wt cells and the *TP53^−/−^* knock out clonal subline AMO-1 TP53^−/−^ is available at the NCBI GEO (http://www.ncbi.nlm.nih.gov/geo) accession number GSE254959. Single cell RNA sequencing data from multiple myeloma patient samples is available at the NCBI GEO (http://www.ncbi.nlm.nih.gov/geo) accession number GSE263201. All other data are available upon request from the corresponding author.

### Authors’ contributions

Conceptualization – K.A., J.J.M., T.S., S.N., C.A.H.

Data Curation – K.A., P.Se., J.J.M., T.S., T.B., T.H., J.O., M.T.

Formal Analysis – K.A., P.Se., J.J.M., T.H., T.B., T.S., J.O., M.T.

Funding Acquisition – S.N., T.S., C.A.H.

Investigation – K.A., P.Se., J.J.M., M.-E.H., U.M., T.B., T.H.

Methodology – K.A., P.Se., J.J.M., Y.D., K.W.-M., M.T., J.O., U.M.

Project administration – K.A., C.A.H.

Resources – K.A., C.A.H., T.S.

Software – P.Se., T.B., J.O., M.T.

Supervision – K.A., C.A.H.

Validation – K.A., C.A.H., T.S., S.N.

Visualization – K.A., P.Se., J.J.M., T.B.

Writing (Original Draft Preparation) – K.A., P.Se., J.J.M., C.A.H.

All authors participated in paper reviewing and editing.

