## Supplementary files for "Efficacy of Melflufen in Patients with Relapsed/Refractory Multiple Myeloma and Mutated or Deleted *TP53*"

### SUPPLEMENTARY MATERIALS AND METHODS

#### Patient samples

Bone marrow (BM) aspirates (n=24) were collected from 23 multiple myeloma (MM) patients after informed consent using protocols approved by an ethical committee and in accordance with the Declaration of Helsinki. BM mononuclear cells (BM-MNCs) were purified by Ficoll gradient centrifugation and cryopreserved prior to analyses.

#### Ex vivo drug sensitivity testing with patient cells by flow cytometry

For flow cytometry (FC)-based drug sensitivity testing, viably frozen MM patient BM-MNC were thawed, freezing solution washed away, cell pellet suspended in conditioned medium (RPMI 1640 medium supplemented with 10% FBS, 2 mM L-glutamine, penicillin (100 U/ml), streptomycin (100 μg/ml) and 25% conditioned medium from the HS-5 human bone marrow stromal cell line),[1] DNase I (Promega) treated for 60 min, and cultured in conditioned medium overnight. Cells were filtered through a 70 μm cell strainer (Fisher Scientific), cell viability measured and cells plated in parallel on pre-drugged 96-well plates (100,000 viable cells/well in 100 μl). If cell viability was poor (<50%), the Dead Cell Removal kit (Miltenyi Biotec) was used prior to plating the cells. Stock solutions (10 000 μM) of melflufen (Oncopeptides AB) and melphalan (Sigma-Aldrich), that had been prepared in advance and stored at -80 °C, were thawed and diluted in conditioned medium to desired concentrations shortly before the drug testing to minimize auto-hydrolysis of melflufen and melphalan (exchange of Cl with OH at the *N*-mustard part of the molecules). Melflufen and melphalan were added to the 96-well plates by manually pipetting only after the cells had been plated, just prior to the start of the drug sensitivity test. The cells were incubated with the drugs for 72 h at 37 °C and 5% $CO_2$.

Following 72 h incubation with the drugs, cells were centrifuged (500xg, 6 min) in the 96-well plates and media discarded by inverting the plate. Cells were suspended in 25 µl of antibody mix containing staining buffer (5% FBS in Dulbecco's phosphate-buffered saline) and the following two antibodies (BD Biosciences): CD138 (BV605, clone MI15, dilution 1:100) and CD38 (BV786, clone HIT2, 1:100). Cells were stained for 30 min at room temperature in the dark and subsequently washed with 100 µl staining buffer followed by centrifugation (500xg, 6 min) and supernatant removal by inverting the plate. Apoptotic and dead cells were discriminated by 7-aminoactinomycin D (7-AAD) and PE-Annexin V (BD Biosciences) staining with both dyes diluted 1:50 in 25 µl annexin V binding buffer. The plates were incubated for 10 min at room temperature before FC analysis. FC analysis was performed using the IntelliCyt iQue Screener PLUS instrument (Sartorius). MM patient sample BM-MNCs were acquired for analysis from each 96-well plate well by the iQue Screener PLUS instrument using a 17 s sip time per well and a pump speed of 32 revolutions per minute. ForeCyt software (Sartorius) was used to gate cells and acquire population counts. Analysis was done from viable singlet cells. The cell count of a well was normalized to its 6 adjacent DMSO controls, from which the highest and lowest value were excluded and normalized values were used for calculating cell viability percentages for each tested drug concentration, and dose response curves drawn. Drug sensitivity score (DSS) values were calculated based on the dose-response curves.[2]

**Single-cell gene expression analysis**

For single cell RNA sequence (scRNAseq) analysis, the MM patient BM-MNCs were sorted based on cell viability (7-AAD, BD Biosciences) and CD138+ surface expression (APC, clone MI15, BD Biosciences) using fluorescence-activated cell sorting (BD Influx Cell Sorter, BD Biosciences). CD138+ and CD138− sorted cell fractions were mixed at a 1:1 ratio with a maximum of 800,000 cells per sample. If there were fewer than 400,000 CD138+ cells, then all CD138+ cells were mixed with 400,000 CD138− cells. scRNAseq library preparation was performed on the mixed CD138+ and CD138+ cell samples using the Chromium Single Cell

3′ Gene Expression v3 reagent kit (10x Genomics). The gel beads in emulsion (GEM) generation, cDNA amplification and library preparation were performed according to the manufacturer's instructions. The prepared libraries were then sequenced on the Illumina NovaSeq 6000 instrument (Illumina) with paired-end reads 28 bp and 89 bp.

Analysis of scRNA-seq data was performed in R Studio environment, using Seurat package (version 4.0.1).[3] Upon data uploading, quality control of cells was done. We used percentage of mitochondrial genes (< triple median or 30%, whichever is less), number of unique genes (>500), number of transcripts (< triple median), and log10 transformed ratio of number of transcripts to number of unique genes (>0.8) to remove dead and late apoptotic cells, as well as multiplets from the subsequent analysis. Samples were integrated, using anchoring procedure, normalized and variance stabilized, and clustered, based on construction of a Shared Nearest Neighbour Graph, which utilized dataset with dimensions reduced by the prior principal component analysis. Clusters were identified, based on the gene expression patterns, and divided into plasma cells (CD138+ pre-sorted fraction) and cells of other types (CD138- pre-sorted fraction) for further analysis. After the division of the initial dataset, new ones were re-integrated, normalized, and clustered. Additionally, for plasma cells dataset, we filtered out immunoglobulin genes. Final clustering results were checked for the distribution of cells, based on cell cycle phase and mitochondrial content, and annotated, using the genes expressed differently between the given and all the other clusters.

For the analysis of differentially expressed genes (DGE), we sequentially compared HS vs IS, IS vs LS, and HS vs LS samples, filtering out non-monotonous changes in gene expression between those groups, and using the threshold of 0.2 for log2-transfored fold change values. The identification of DGE was done by the means of MAST framework. [4]

For the gene set enrichment analysis, clusterProfiler (v.3.18.1)[5] and ReactomePA (v. 1.34.0)[6] packages were used. We utilized Reactome[7], KEGG[8] and MSigDB[9,10] datasets. Differentially expressed genes from melflufen comparison, with preliminary subtracted genes from DGE analysis for melphalan, and retained genes altered due to the disease stage, were used as an input to cluster genes, based on their belonging to the gene sets, respectively to their functions, regulation, or expression patterns.

**Cell lines, cell culture and cytotoxicity**

The AMO-1 cell line was obtained from DSMZ (Braunschweig, Germany). Generation and characterization of *TP53* deficient AMO-1 clones is described in Munawar et al., 2019.[11] Standard cell culture was performed in RPMI-1640 medium supplemented with 10% fetal bovine serum (FBS), 1 mM sodium pyruvate, 2 mM glutamine, 100 U/ml penicillin, and 100 µg/ml streptomycin. Cells were kept at 5% $CO_2$ and 37 °C. Cells were cultured on 96well plates in 200 µl culture medium for 72 h exposure period before measuring the number of viable cells. Cell viability was determined using the AlamarBlue™ reagent (Invitrogen) or cell death assays (annexin V/propidiumiodide) according to the manufacturers' protocols.

**Compounds and stock solutions**

Melflufen (Oncopeptides AB) was freshly prepared in DMSO before application. Melphalan (Sigma-Aldrich, M2011) stock solution was prepared in acidified ethanol. Cyclophosphamide (N,N-bis(2-chloroethyl)-4-hydroperoxy-2-oxo-1,3,25-oxazaphosphinan-2-amine,TRC (product number TRC-H714675) was freshly prepared in DMSO.

**RNA preparation and RNA-seq analysis**

AMO1 *TP53* wild type cells or the *TP53*$^{-/-}$ knock out clonal subline AMO-1 TP53$^{-/-}$ were treated with melphalan or melflufen at different concentrations and timepoints. RNA was isolated according to the RNeasy plus protocol, quantified with Nanodrop (Thermo Scientific), and quality assessed with the Agilent 2100 Bioanalyzer. DNA libraries were prepared using the TruSeq Stranded mRNA Library Preparation Kit (Illumina) and sequencing performed on the NextSeq 500 platform (Illumina). Base calling was performed by Real-Time Analysis (RTA) version 2.4.11 or 2.11.3 and demultiplexed FASTQ files were generated with bcl2fastq2 v2.20.0.422 (Illumina). To assure high sequence quality, Illumina reads were quality- and adapter-trimmed via Cutadapt[12,13] version 2.5 using a cutoff Phred score of 20 in NextSeq mode and reads without any remaining bases were discarded (command line parameters: --nextseq-trim=20 -m 1 -a AGATCGGAAGAGCACACGTCTGAACTCCAGTCAC). Processed reads were subsequently mapped to the human genome (GRCh38.p14) using STAR v2.7.2b with default parameters based on RefSeq annotation version 110 for GRCh38.p14.[14] Read counts on exon level summarized for each gene were generated using featureCounts v1.6.4 from the Subread package.[15] Multi-mapping and multi-overlapping reads were counted strand-specific and reversely stranded with a fractional count for each alignment and overlapping feature (command line parameters: -s 2 -t exon -M -O --fraction). The count output was utilized to identify differentially expressed genes using DESeq2[15] version 1.24.0. Read counts were normalized by DESeq2 and fold-change shrinkage was applied by setting the parameter “betaPrior=TRUE”. Differential expression of genes was assumed at an adjusted p-value (padj) after Benjamini-Hochberg correction < 0.05 and |log2FoldChange| ≥ 0.5. clusterProfiler version 3.12.0 was used to perform functional enrichment analyses based on Kyoto Encyclopedia of Genes and Genomes (KEGG) pathways and Gene Ontology (GO) terms.[5] The *enricher* function was used to perform hypergeometric tests based on lists of significant genes and the *GSEA* function was applied for gene set enrichment analysis considering the DESeq2 log2FoldChange of all analyzed genes. R was applied to generate heatmaps.

**DNA damage signaling and cell apoptosis analyses**

AMO-1 *TP53* wild type and AMO-1 *TP53*$^{-/-}$ cells were treated with melflufen, melphalan or cyclophosphamide at the indicated concentrations and time points. H2AX phosphorylation at Ser139 (γ-H2AX) for DNA damage signal was analyzed using flow cytometry. The following protocol was applied for *γ-H2AX staining;* Cells were fixed in Flow Cytometry Fixation Buffer (R&D Systems) and stored in 70% ethanol at -20°C overnight. Prior to incubation with Alexa Fluor 488-conjugated phosphorylated H2AX antibody (Sigma-Aldrich) or Mouse IgG1 (R&D Systems), cells were blocked with Human Seroblock (Bio-Rad Laboratories).

For apoptosis analysis cells were stained for annexin V-APC and PI (Invitrogen) as follows; cells were suspended in staining mix containing Annexin V APC (Invitrogen) and PI (Invitrogen), fixed in Flow Cytometry Fixation Buffer and resuspended in annexin V binding buffer (Invitrogen).

All samples were analyzed with the FACSCanto II flow cytometer (BD Biosciences). Data were analyzed using FlowJo software (BD Biosciences).

**JC-1 mitochondria membrane potential assay**

AMO-1 *TP53* wild type and AMO-1 *TP53*$^{-/-}$ cells at 1x10$^{5}$/mL were treated with melflufen, melphalan or cyclophosphamide at the indicated concentrations and cells were harvested after 2h or 4h. The JC-1 mitochondrial membrane potential assay was performed according to the manufacturers protocol (MitoProbe™ JC-1 Assay Kit for Flow Cytometry (M34152), Molecular Probes, Eugene, OR). Following drug treatment, the cells were stained with JC-1 (5,5',6,6'-tetrachloro-1,1',3,3'-tetra-ethyl-benzimidazolyl-carbocyanine iodide) reagent for 15 min at 37 °C, pelleted by centrifugation (4 min/250xg), washed with PBS and resuspended in PBS. The ratio of green to red fluorescence was determined with a Spectra Max i3X (Molecular Devices) plate reader. Data was generated with 488 nm excitation and using 529 nm and 590 nm emission filters. Obtained measurement data was analyzed to calculate the mitochondria membrane potential changes upon drug treatment.

**Statistical analyses of the OCEAN trial data**

Primary analysis of tumor response and progression-dependent endpoints of patients enrolled in the OCEAN trial (NCT03151811) were based on response assessments by an Independent Review Committee. All tumor response and progression-dependent endpoints were assessed using the IMWG Uniform Response Criteria (IMWG-URC). The overall response rate (ORR) was defined as the proportion of patients for whom the best overall confirmed response was stringent complete response (sCR), complete response (CR), very good partial response

(VGPR), or partial response (PR). Post-hoc analyses were performed to examine associations between progression-free survival (PFS) and chromosome 17 deletion status and/or *TP53* gene mutation status. Furthermore, another post-hoc analysis was carried out to study the patient responder status in the chromosome 17 deletion population.

**Assessment of *TP53* mutation status of OCEAN trial patients**

*TP53* mutation status of OCEAN trial patients was determined by next generation sequencing (NGS) using Almac Genomic Services. In brief, Almac´s NGS v2 TP53 Assay has been performed what detects germline and somatic single nucleotide variants (SNV), small insertion and small deletion events within the promoter region, the coding and non-coding regions of exons one to ten, a portion of exon eleven, and the intron-exon boundaries of TP53 isoform a (NM_000546) using the Illumina MiSeq platform. Sample preparation for the sequencing has been performed as followed; from patients, whom consented, bone marrow aspirates CD138+ purified plasma cells had been frozen shipped to Almac Genomic Services. From CD138+ cells, DNA was extracted using QIAamp® DNA Blood Midi Kit (QIAGEN©), DNA quality was checked according to Almac`s Qubit quality control. NGS v2 TP53 assay was performed using the TruSeq Custom Amplicon Kit Dx and custom TP53 oligo panel. Analysis was performed as follows: sequencing, demultiplexing, FASTQ file generation, sequence alignment and variant calling was automatically performed onboard the MiSeq system. Sequencing run, sample and control QC was performed using the Almac Genomic Services developed software, named Almac TP53 CTA v2 Analysis. All detected SNVs were classified and aligned with the National Library of Medicine, NIH database, for ClinVar genomic variation database and clinical relevance to classify mutation type into benign or pathogenic.

### Supplementary Figure Legends

**Supplementary Figure 1. GSEA results of *TP53*mut vs *TP53*wt cells in cell lines and patient samples.** Bar plots, representing normalized enrichment score for top 20 deregulated gene sets, consisting of the most deregulated genes between *TP53* mutated and *TP53*wt cell lines (A), patient samples (B), and commonly deregulated genes (C) in both tested models.

**Supplementary Figure 2. Gene expression analysis in *TP53*wt versus *TP53*$^{-/-}$ genetic background AMO-1 cells.** Functional and signalling pathways gene expression heat maps after 2h and 12h treatment with melflufen or melphalan at their IC90 values in *TP53*wt or *TP53*$^{-/-}$ AMO-1 cells.

**Supplementary Table 1. Differentially expressed genes between *TP53*wt and *TP53*mut cohorts of patient samples.**

**Supplementary Table 2. Differentially expressed genes between *TP53*wt and *TP53*$^{-/-}$ cell line models.**

**Supplementary Table 3. Commonly deregulated differentially expressed genes between *TP53*wt and *TP53*mut cohorts of patient samples and cell line models.**

**Supplementary Table 4. Melflufen and melphalan EC50 and EC90 values in the AMO-1 isogenic wild type (*TP53*wt) or mutant (*TP53*$^{-/-}$) cell lines.** Cell lines were treated with either melflufen or melphalan at 8-point drug concentration range to determine EC50 or EC90 values using viability assay.

**Supplementary Table 5. Patients baseline demographic and diseases characteristics in del(17p) patient subpopulation from the OCEAN trial.**

**Supplementary Table 6. Cytogenetics data of del(17p) subpopulation from the OCEAN trial.**

**Supplementary Table 7. Patients baseline demographic and diseases characteristics in del(17p) and/or *TP53* gene mutated patient subpopulation from the OCEAN trial.**

**Supplementary Figure S1**

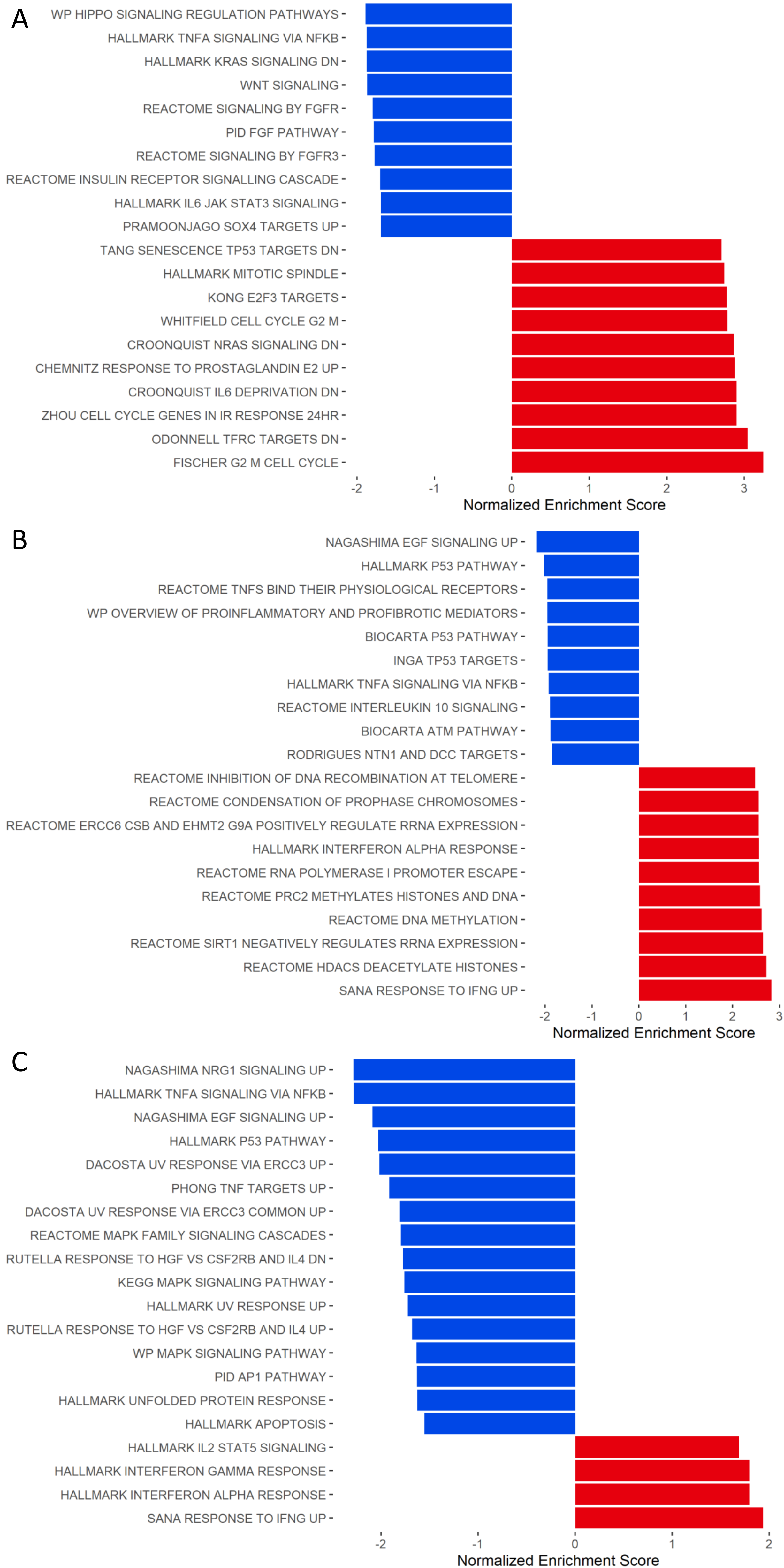

**Supplementary Figure S2**

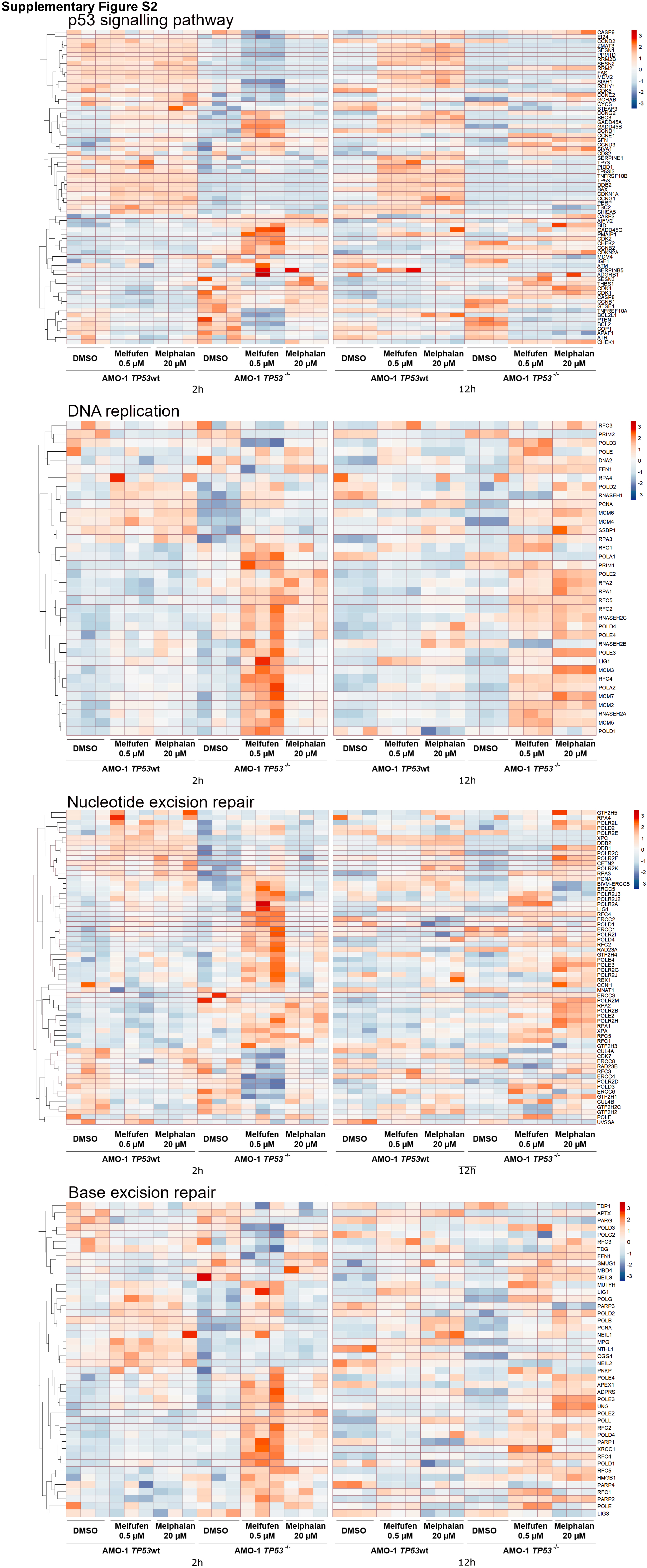

**Supplementary Table S1**

| | p_val | avg_log2FC | pct.1 | pct.2 | p_val_adj |
|---|---|---|---|---|---|
| PDLIM1 | 0 | 3.664687853 | 0.681 | 0.117 | 0 |
| C2orf76 | 0 | 3.420471839 | 0.583 | 0.091 | 0 |
| TEX9 | 0 | 2.286099642 | 0.715 | 0.235 | 0 |
| TMSB4X | 0 | 0.90976914 | 0.97 | 0.511 | 0 |
| SNRPA1 | 0 | 2.156806557 | 0.678 | 0.23 | 0 |
| CCT5 | 0 | 2.684253142 | 0.792 | 0.345 | 0 |
| TIMP1 | 0 | -1.743381244 | 0.116 | 0.55 | 0 |
| CTSC | 0 | 1.843922557 | 0.617 | 0.194 | 0 |
| ADAM28 | 0 | 2.202429331 | 0.436 | 0.056 | 0 |
| LSM6 | 0 | 1.682444727 | 0.657 | 0.277 | 0 |
| AREG | 0 | -3.813613138 | 0.06 | 0.438 | 0 |
| ACADM | 0 | 1.785230408 | 0.578 | 0.2 | 0 |
| PTTG1 | 0 | 2.500333653 | 0.448 | 0.074 | 0 |
| CKLF | 0 | 1.349794982 | 0.692 | 0.323 | 0 |
| IFT57 | 0 | 1.490791577 | 0.707 | 0.344 | 0 |
| MRPL16 | 0 | 1.654690202 | 0.563 | 0.2 | 0 |
| MRPL3 | 0 | 1.513319351 | 0.67 | 0.309 | 0 |
| PSMG1 | 0 | 2.247866535 | 0.492 | 0.131 | 0 |
| COPS5 | 0 | 1.46056881 | 0.653 | 0.294 | 0 |
| SDHAF3 | 0 | 1.905224517 | 0.469 | 0.11 | 0 |
| CCT2 | 0 | 1.379970259 | 0.69 | 0.336 | 0 |
| PSMA1 | 0 | 1.563550789 | 0.784 | 0.431 | 0 |
| MRPL36 | 0 | 1.842694291 | 0.586 | 0.233 | 0 |
| PSMA2 | 0 | 1.403057147 | 0.752 | 0.4 | 0 |
| GAS2 | 0 | 1.797866305 | 0.493 | 0.142 | 0 |
| MRPL13 | 0 | 1.593190446 | 0.573 | 0.223 | 0 |
| STMP1 | 0 | 1.557474358 | 0.566 | 0.217 | 0 |
| ANXA5 | 0 | 1.465860868 | 0.783 | 0.435 | 0 |
| APOL3 | 0 | 3.309008404 | 0.363 | 0.017 | 0 |
| SPOCK2 | 0 | 3.31339059 | 0.367 | 0.022 | 0 |
| FAM30A | 0 | 0.788409479 | 0.59 | 0.246 | 0 |
| CCDC34 | 0 | 2.532285529 | 0.421 | 0.078 | 0 |
| HDDC2 | 0 | 1.394602557 | 0.602 | 0.259 | 0 |
| GLRX | 0 | 0.63081179 | 0.743 | 0.401 | 0 |
| TCF4 | 0 | 0.381815775 | 0.778 | 0.439 | 0 |
| OLA1 | 0 | 1.364450273 | 0.6 | 0.262 | 0 |
| LPIN1 | 0 | 1.009935629 | 0.523 | 0.188 | 0 |
| NIT2 | 0 | 1.622802067 | 0.525 | 0.19 | 0 |
| SSB | 0 | 1.295647837 | 0.73 | 0.397 | 0 |
| PSMD14 | 0 | 1.215588037 | 0.598 | 0.267 | 0 |
| MPC2 | 0 | 1.654053609 | 0.749 | 0.42 | 0 |
| THOC7 | 0 | 1.210747362 | 0.696 | 0.367 | 0 |
| RHOBTB3 | 0 | 2.31633227 | 0.414 | 0.086 | 0 |
| THYN1 | 0 | 1.372417273 | 0.534 | 0.211 | 0 |
| GTF3C6 | 0 | 1.250404124 | 0.623 | 0.3 | 0 |
| MRPL21 | 0 | 1.303532592 | 0.574 | 0.251 | 0 |
| OTUD1 | 0 | -2.44605744 | 0.274 | 0.595 | 0 |
| DECR1 | 0 | 1.054851728 | 0.686 | 0.367 | 0 |
| BUB3 | 0 | 1.18576582 | 0.623 | 0.305 | 0 |

| | | | | | |
|---|---|---|---|---|---|
| SNRPF | 0 | 1.202622728 | 0.708 | 0.39 | 0 |
| PSMD13 | 0 | 1.1514223 | 0.664 | 0.35 | 0 |
| ELOC | 0 | 0.970902058 | 0.692 | 0.381 | 0 |
| ATP5F1C | 0 | 1.226032061 | 0.756 | 0.446 | 0 |
| ANXA2 | 0 | 0.676031905 | 0.714 | 0.405 | 0 |
| NDUFB6 | 0 | 0.932838533 | 0.668 | 0.361 | 0 |
| METTL5 | 0 | 0.98402097 | 0.641 | 0.334 | 0 |
| PMPCB | 0 | 1.114328174 | 0.573 | 0.267 | 0 |
| GTF2A2 | 0 | 1.0028673 | 0.683 | 0.379 | 0 |
| UTP3 | 0 | 1.503341195 | 0.508 | 0.206 | 0 |
| PSMB2 | 0 | 1.042296855 | 0.686 | 0.386 | 0 |
| HNRNPR | 0 | 0.984679112 | 0.732 | 0.432 | 0 |
| SRP72 | 0 | 1.41385347 | 0.832 | 0.533 | 0 |
| GLRX3 | 0 | 1.38538194 | 0.48 | 0.184 | 0 |
| PSMA5 | 0 | 1.05021204 | 0.683 | 0.39 | 0 |
| UBE2N | 0 | 1.006253542 | 0.52 | 0.227 | 0 |
| PSMA4 | 0 | 0.884032593 | 0.751 | 0.458 | 0 |
| RAN | 0 | 1.38593788 | 0.831 | 0.538 | 0 |
| ATP6V1A | 0 | 1.51589263 | 0.421 | 0.129 | 0 |
| ZNF791 | 0 | 0.85764303 | 0.774 | 0.483 | 0 |
| VDAC3 | 0 | 1.224637695 | 0.568 | 0.277 | 0 |
| CBX3 | 0 | 1.081237502 | 0.726 | 0.436 | 0 |
| HSD17B10 | 0 | 0.987361002 | 0.605 | 0.316 | 0 |
| MRPL42 | 0 | 0.870777225 | 0.565 | 0.276 | 0 |
| NDUFA12 | 0 | 0.956450717 | 0.694 | 0.407 | 0 |
| RBX1 | 0 | 0.924528882 | 0.697 | 0.41 | 0 |
| CCT8 | 0 | 0.970724725 | 0.762 | 0.476 | 0 |
| MINOS1 | 0 | 0.958039869 | 0.722 | 0.437 | 0 |
| MACF1 | 0 | 0.750819688 | 0.59 | 0.306 | 0 |
| SEC11A | 0 | 1.215686599 | 0.822 | 0.538 | 0 |
| IL6R | 0 | -2.223770292 | 0.087 | 0.37 | 0 |
| SF3B5 | 0 | 0.961625156 | 0.807 | 0.524 | 0 |
| COA4 | 0 | 0.89016747 | 0.614 | 0.332 | 0 |
| G3BP1 | 0 | 0.931977622 | 0.597 | 0.315 | 0 |
| LSM3 | 0 | 1.169657544 | 0.717 | 0.435 | 0 |
| EIF4G2 | 0 | 1.084394557 | 0.837 | 0.555 | 0 |
| TOMM22 | 0 | 0.861471114 | 0.645 | 0.364 | 0 |
| VPS29 | 0 | 0.872379523 | 0.619 | 0.338 | 0 |
| PFDN2 | 0 | 0.834537105 | 0.652 | 0.372 | 0 |
| DCAF13 | 0 | 1.076419382 | 0.5 | 0.221 | 0 |
| HCLS1 | 0 | 0.829974636 | 0.677 | 0.398 | 0 |
| PRPF40A | 0 | 0.845869429 | 0.681 | 0.403 | 0 |
| MRPL22 | 0 | 0.97644171 | 0.559 | 0.282 | 0 |
| PSMB7 | 0 | 0.823471266 | 0.703 | 0.427 | 0 |
| ENY2 | 0 | 0.649801371 | 0.761 | 0.49 | 0 |
| PPHLN1 | 0 | 0.773548088 | 0.641 | 0.37 | 0 |
| PRDX3 | 0 | 0.93832879 | 0.567 | 0.297 | 0 |
| ARPC3 | 0 | 1.064958717 | 0.848 | 0.579 | 0 |
| ATP6V1F | 0 | 0.867465927 | 0.659 | 0.39 | 0 |
| PDHB | 0 | 1.01645015 | 0.507 | 0.238 | 0 |

| | | | | | |
|---|---|---|---|---|---|
| CRNDE | 0 | 1.420347084 | 0.416 | 0.148 | 0 |
| HSPE1 | 0 | 0.834593454 | 0.812 | 0.545 | 0 |
| SRP9 | 0 | 0.716154404 | 0.708 | 0.441 | 0 |
| EID1 | 0 | 1.339332374 | 0.835 | 0.569 | 0 |
| JPT1 | 0 | 1.527763733 | 0.383 | 0.117 | 0 |
| MRPL32 | 0 | 0.856546289 | 0.588 | 0.322 | 0 |
| POMP | 0 | 0.76886582 | 0.698 | 0.433 | 0 |
| FRG1 | 0 | 0.823408354 | 0.572 | 0.308 | 0 |
| CISD1 | 0 | 0.892524441 | 0.508 | 0.246 | 0 |
| EIF2S2 | 0 | 0.555351976 | 0.762 | 0.501 | 0 |
| ITGB3BP | 0 | 1.162445862 | 0.386 | 0.125 | 0 |
| NUDT5 | 0 | 0.925290381 | 0.524 | 0.264 | 0 |
| NKAPD1 | 0 | 1.378014218 | 0.387 | 0.128 | 0 |
| NUDT21 | 0 | 1.140671716 | 0.47 | 0.212 | 0 |
| SOD1 | 0 | 0.612248875 | 0.758 | 0.5 | 0 |
| PSMD7 | 0 | 0.701598921 | 0.687 | 0.43 | 0 |
| CMC2 | 0 | 1.174191778 | 0.433 | 0.176 | 0 |
| KLF2 | 0 | -2.631480419 | 0.626 | 0.883 | 0 |
| PTPN1 | 0 | -2.318727925 | 0.053 | 0.31 | 0 |
| SAMD9L | 0 | 0.718269431 | 0.475 | 0.22 | 0 |
| SDHB | 0 | 0.987982169 | 0.523 | 0.268 | 0 |
| NDUFB3 | 0 | 0.571046993 | 0.736 | 0.483 | 0 |
| CEBPB | 0 | -1.346547891 | 0.157 | 0.409 | 0 |
| PSMB5 | 0 | 0.740236937 | 0.609 | 0.357 | 0 |
| AHNAK | 0 | -1.079306698 | 0.237 | 0.488 | 0 |
| CDC123 | 0 | 1.203378247 | 0.43 | 0.179 | 0 |
| PSMA7 | 0 | 0.776689293 | 0.82 | 0.569 | 0 |
| TXN | 0 | 1.799974964 | 0.885 | 0.635 | 0 |
| SDHD | 0 | 0.84881851 | 0.552 | 0.303 | 0 |
| ANP32A | 0 | 0.80148512 | 0.607 | 0.358 | 0 |
| ANXA6 | 0 | 0.764570228 | 0.525 | 0.276 | 0 |
| LEO1 | 0 | 1.460147888 | 0.359 | 0.11 | 0 |
| MLKL | 0 | 1.561079169 | 0.338 | 0.09 | 0 |
| BCCIP | 0 | 1.126951643 | 0.425 | 0.177 | 0 |
| ELL2 | 0 | 0.524548059 | 0.822 | 0.574 | 0 |
| APLP2 | 0 | 1.462768014 | 0.431 | 0.184 | 0 |
| PRDX6 | 0 | 0.674852293 | 0.712 | 0.465 | 0 |
| CFAP298 | 0 | 0.988893075 | 0.436 | 0.19 | 0 |
| TRIAP1 | 0 | 1.126280277 | 0.418 | 0.172 | 0 |
| XIST | 0 | -13.95297672 | 0 | 0.246 | 0 |
| S100A10 | 0 | 0.48841121 | 0.649 | 0.404 | 0 |
| TENT5C | 0 | -1.662438698 | 0.451 | 0.695 | 0 |
| MRPL18 | 0 | 0.927109251 | 0.487 | 0.244 | 0 |
| SUMO1 | 0 | 0.661717428 | 0.617 | 0.374 | 0 |
| OSTC | 0 | 0.67029785 | 0.805 | 0.564 | 0 |
| SNRPD2 | 0 | 1.160330683 | 0.865 | 0.624 | 0 |
| CD47 | 0 | 0.529481375 | 0.664 | 0.425 | 0 |
| GNAI2 | 0 | 0.664290612 | 0.659 | 0.42 | 0 |
| B3GALNT2 | 0 | 1.643477689 | 0.32 | 0.081 | 0 |
| DYNLL1 | 0 | 0.926556981 | 0.841 | 0.602 | 0 |

| | | | | | |
|---|---|---|---|---|---|
| NAE1 | 0 | 1.033296003 | 0.417 | 0.178 | 0 |
| PTTG1IP | 0 | 1.202318621 | 0.411 | 0.172 | 0 |
| POLR2G | 0 | 0.995183242 | 0.466 | 0.228 | 0 |
| MTCH2 | 0 | 1.14247988 | 0.42 | 0.182 | 0 |
| SNRPC | 0 | 0.66770827 | 0.616 | 0.378 | 0 |
| RWDD1 | 0 | 0.47185114 | 0.704 | 0.466 | 0 |
| SMC2 | 0 | 1.435595237 | 0.365 | 0.128 | 0 |
| ZNHIT1 | 0 | 0.511049194 | 0.688 | 0.451 | 0 |
| PRKCB | 0 | -1.484646642 | 0.057 | 0.292 | 0 |
| SF3A3 | 0 | 1.100754027 | 0.425 | 0.19 | 0 |
| KRR1 | 0 | 0.628046074 | 0.554 | 0.32 | 0 |
| GGH | 0 | 1.49406422 | 0.371 | 0.137 | 0 |
| PLA2G16 | 0 | 0.379213971 | 0.64 | 0.406 | 0 |
| SLC38A1 | 0 | 0.534210175 | 0.578 | 0.344 | 0 |
| PSMB3 | 0 | 0.541269169 | 0.733 | 0.501 | 0 |
| ECHDC1 | 0 | 0.94022131 | 0.454 | 0.223 | 0 |
| RARS | 0 | 0.992282583 | 0.413 | 0.182 | 0 |
| UBE2V2 | 0 | 0.78534755 | 0.491 | 0.26 | 0 |
| TIAL1 | 0 | 0.650532935 | 0.521 | 0.292 | 0 |
| LYRM7 | 0 | 1.399069983 | 0.323 | 0.094 | 0 |
| APOL6 | 0 | -1.029192407 | 0.177 | 0.405 | 0 |
| HSBP1 | 0 | 1.142084327 | 0.463 | 0.235 | 0 |
| NBN | 0 | 0.932663059 | 0.436 | 0.208 | 0 |
| TMEM126B | 0 | 0.949670764 | 0.428 | 0.2 | 0 |
| ATP5MF | 0 | 0.534577875 | 0.787 | 0.561 | 0 |
| UNG | 0 | 2.06903716 | 0.285 | 0.059 | 0 |
| CCDC91 | 0 | 0.771219411 | 0.471 | 0.246 | 0 |
| DHX30 | 0 | 1.002081767 | 0.374 | 0.15 | 0 |
| GOLGA7 | 0 | 1.143295454 | 0.393 | 0.169 | 0 |
| COX7B | 0 | 0.615745033 | 0.834 | 0.61 | 0 |
| MED12L | 0 | 2.737022052 | 0.241 | 0.018 | 0 |
| SAMSN1 | 0 | -2.36556343 | 0.033 | 0.256 | 0 |
| BRIX1 | 0 | 0.822793763 | 0.427 | 0.205 | 0 |
| BAG2 | 0 | 1.971472105 | 0.284 | 0.062 | 0 |
| SETD7 | 0 | 1.03036481 | 0.369 | 0.148 | 0 |
| AC006058.1 | 0 | 3.689492925 | 0.228 | 0.009 | 0 |
| PARK7 | 0 | 0.626186949 | 0.802 | 0.584 | 0 |
| FAM210A | 0 | 1.382340305 | 0.316 | 0.098 | 0 |
| GTF2H5 | 0 | 0.848038326 | 0.437 | 0.219 | 0 |
| RAD51C | 0 | 1.707064756 | 0.303 | 0.086 | 0 |
| ATP5PF | 0 | 0.781948155 | 0.849 | 0.633 | 0 |
| C4orf46 | 0 | 2.224799477 | 0.259 | 0.044 | 0 |
| MAPRE2 | 0 | 1.868629771 | 0.257 | 0.044 | 0 |
| MRPL39 | 0 | 1.665907052 | 0.307 | 0.094 | 0 |
| NSMCE3 | 0 | -2.339083822 | 0.094 | 0.306 | 0 |
| CKS1B | 0 | 1.265088089 | 0.391 | 0.182 | 0 |
| CTR9 | 0 | 0.894626148 | 0.381 | 0.172 | 0 |
| PRPS2 | 0 | 1.230407496 | 0.332 | 0.125 | 0 |
| EXOSC7 | 0 | 1.108314041 | 0.357 | 0.151 | 0 |
| PKN1 | 0 | 0.291677929 | 0.48 | 0.274 | 0 |

| | | | | | |
|---|---|---|---|---|---|
| SEC61G | 0 | 0.533418637 | 0.874 | 0.669 | 0 |
| MGST2 | 0 | 1.394664583 | 0.345 | 0.141 | 0 |
| CREB5 | 0 | 1.104029616 | 0.283 | 0.082 | 0 |
| MRPS9 | 0 | 0.831429722 | 0.359 | 0.159 | 0 |
| FAM111A | 0 | 1.164817089 | 0.311 | 0.113 | 0 |
| PRDX2 | 0 | 0.813581728 | 0.816 | 0.62 | 0 |
| AC098850.3 | 0 | -4.415245629 | 0.011 | 0.206 | 0 |
| CUTC | 0 | 1.310053625 | 0.284 | 0.091 | 0 |
| DLD | 0 | 1.008961399 | 0.307 | 0.115 | 0 |
| KLF3 | 0 | -1.932565569 | 0.117 | 0.309 | 0 |
| PHF19 | 0 | 1.882016652 | 0.264 | 0.073 | 0 |
| RASD1 | 0 | -5.067100576 | 0.009 | 0.198 | 0 |
| HOPX | 0 | 1.685368902 | 0.256 | 0.069 | 0 |
| MCM6 | 0 | 1.462979157 | 0.279 | 0.093 | 0 |
| GPN3 | 0 | 1.264026689 | 0.282 | 0.099 | 0 |
| CXCR4 | 0 | -1.492094528 | 0.288 | 0.467 | 0 |
| MKI67 | 0 | 2.464828436 | 0.204 | 0.026 | 0 |
| NLRP2 | 0 | 3.920228433 | 0.182 | 0.012 | 0 |
| ENAH | 0 | 2.350844766 | 0.193 | 0.024 | 0 |
| GLUL | 0 | -0.463432602 | 0.057 | 0.225 | 0 |
| TUT4 | 0 | -1.571893356 | 0.277 | 0.445 | 0 |
| HSPA1B | 0 | -3.589401052 | 0.195 | 0.362 | 0 |
| C12orf73 | 0 | 1.616174412 | 0.218 | 0.052 | 0 |
| LY96 | 0 | -1.565670048 | 0.166 | 0.332 | 0 |
| EPHA6 | 0 | 2.090576232 | 0.217 | 0.052 | 0 |
| PARPBP | 0 | 2.61523168 | 0.183 | 0.018 | 0 |
| PIAS2 | 0 | 0.327765857 | 0.317 | 0.152 | 0 |
| CITED2 | 0 | -2.144712212 | 0.243 | 0.408 | 0 |
| MFSD8 | 0 | 1.041863032 | 0.25 | 0.087 | 0 |
| SLC8A1 | 0 | -1.931455671 | 0.012 | 0.175 | 0 |
| MAGEC2 | 0 | 2.282034077 | 0.186 | 0.024 | 0 |
| DEPTOR | 0 | -2.365696156 | 0.105 | 0.266 | 0 |
| CCNB2 | 0 | 3.152638657 | 0.176 | 0.015 | 0 |
| TENM1 | 0 | 4.496324121 | 0.163 | 0.004 | 0 |
| DUSP5 | 0 | -1.597128069 | 0.477 | 0.634 | 0 |
| EIF3E | 0 | 0.903193582 | 0.876 | 0.72 | 0 |
| CDC6 | 0 | 1.950497599 | 0.195 | 0.041 | 0 |
| PTCH1 | 0 | 1.288135132 | 0.192 | 0.041 | 0 |
| ESR2 | 0 | 2.609705587 | 0.159 | 0.013 | 0 |
| IGSF10 | 0 | 1.837159353 | 0.159 | 0.016 | 0 |
| PIK3AP1 | 0 | 2.26692331 | 0.169 | 0.026 | 0 |
| ATP5MG | 0 | 1.151892968 | 0.916 | 0.774 | 0 |
| FAM216A | 0 | 1.998150699 | 0.172 | 0.03 | 0 |
| FCRL2 | 0 | -4.307206178 | 0.023 | 0.163 | 0 |
| CD86 | 0 | -1.630197142 | 0.024 | 0.163 | 0 |
| SGO1 | 0 | 2.880703864 | 0.151 | 0.015 | 0 |
| BIRC5 | 0 | 2.55268575 | 0.153 | 0.018 | 0 |
| TPX2 | 0 | 2.306901266 | 0.15 | 0.02 | 0 |
| TP53INP1 | 0 | -0.452118395 | 0.556 | 0.428 | 0 |
| CCNB1 | 0 | 2.61060301 | 0.149 | 0.021 | 0 |

| | | | | | |
|---|---|---|---|---|---|
| DSCR8 | 0 | 1.660452231 | 0.148 | 0.027 | 0 |
| ZWINT | 0 | 2.132863684 | 0.139 | 0.02 | 0 |
| TCTEX1D2 | 0 | 1.934298372 | 0.146 | 0.028 | 0 |
| MND1 | 0 | 2.9110809 | 0.128 | 0.011 | 0 |
| PALLD | 0 | 4.268485092 | 0.118 | 0.003 | 0 |
| DEPDC1 | 0 | 3.002123388 | 0.121 | 0.009 | 0 |
| NCAPG | 0 | 2.607586159 | 0.126 | 0.015 | 0 |
| PRC1 | 0 | 2.104210214 | 0.13 | 0.019 | 0 |
| ATP5MPL | 0 | -0.303410403 | 0.675 | 0.565 | 0 |
| JTB | 0 | -0.436865905 | 0.743 | 0.633 | 0 |
| TPM4 | 0 | -0.501102314 | 0.768 | 0.659 | 0 |
| MGLL | 0 | 1.943367778 | 0.126 | 0.019 | 0 |
| EVI2B | 0 | -0.515715983 | 0.678 | 0.574 | 0 |
| JUN | 0 | -2.86322098 | 0.864 | 0.962 | 0 |
| H3F3A | 0 | 0.586282753 | 0.928 | 0.831 | 0 |
| FGFR3 | 0 | -13.3903156 | 0 | 0.096 | 0 |
| PBK | 0 | 3.080433975 | 0.103 | 0.008 | 0 |
| APOBEC3B | 0 | 3.611632696 | 0.1 | 0.006 | 0 |
| PLK4 | 0 | 3.277419858 | 0.101 | 0.007 | 0 |
| AC006058.3 | 0 | 3.802352802 | 0.088 | 0.004 | 0 |
| CD63 | 0 | -0.990615603 | 0.711 | 0.63 | 0 |
| HSPB1 | 0 | -2.768301324 | 0.445 | 0.523 | 0 |
| SSR4 | 0 | -2.131363247 | 0.897 | 0.973 | 0 |
| RASL10B | 0 | 6.652886174 | 0.076 | 0.001 | 0 |
| FOS | 0 | -0.797007171 | 0.846 | 0.92 | 0 |
| AC136475.3 | 0 | 5.776966913 | 0.074 | 0.001 | 0 |
| ZFP36L2 | 0 | -1.280421039 | 0.534 | 0.607 | 0 |
| TXNIP | 0 | -1.197487688 | 0.676 | 0.742 | 0 |
| SRRM2 | 0 | -0.425819357 | 0.765 | 0.701 | 0 |
| CD53 | 0 | -0.754251856 | 0.475 | 0.537 | 0 |
| PNRC1 | 0 | -0.358745578 | 0.844 | 0.786 | 0 |
| DNAJB9 | 0 | -0.782020261 | 0.564 | 0.516 | 0 |
| CPEB4 | 0 | -1.019197606 | 0.513 | 0.557 | 0 |
| MDM4 | 0 | -1.054296841 | 0.591 | 0.626 | 0 |
| FBXW7 | 0 | -0.542880132 | 0.74 | 0.707 | 0 |
| CIRBP | 0 | -1.121610131 | 0.76 | 0.792 | 0 |
| IER2 | 0 | -1.506783145 | 0.851 | 0.82 | 0 |
| IER5 | 0 | -0.679708655 | 0.574 | 0.545 | 0 |
| DDX24 | 0 | -0.889866647 | 0.549 | 0.521 | 0 |
| DDIT4 | 0 | -1.705858436 | 0.296 | 0.317 | 0 |
| STAT3 | 0 | -1.009047391 | 0.515 | 0.535 | 0 |
| KLF13 | 0 | -0.646721913 | 0.62 | 0.605 | 0 |
| JUND | 0 | -0.936465279 | 0.937 | 0.951 | 0 |
| SQSTM1 | 0 | -0.867244032 | 0.507 | 0.52 | 0 |
| H1FX | 0 | -1.051421957 | 0.762 | 0.751 | 0 |
| HLA-E | 0 | -1.405035856 | 0.834 | 0.845 | 0 |
| TSTD1 | 0 | -1.012818196 | 0.561 | 0.572 | 0 |
| GNAS | 0 | -0.623316243 | 0.893 | 0.884 | 0 |
| SMAP2 | 0 | -0.98527339 | 0.365 | 0.374 | 0 |
| EEF1A1 | 0 | 0.370415577 | 0.991 | 0.996 | 0 |

| | | | | | |
|---|---|---|---|---|---|
| EEF1D | 0 | -0.52823342 | 0.92 | 0.923 | 0 |
| TPT1 | 0 | -0.614602609 | 0.983 | 0.985 | 0 |
| DPM1 | 9.88131291682493e-324 | 0.877727298 | 0.422 | 0.208 | 2.1103519996463e-319 |
| SEL1L3 | 4.94065645841247e-323 | 0.508172254 | 0.41 | 0.211 | 1.05517599982315e-318 |
| TPD52 | 1.03753785626662e-322 | 0.511372681 | 0.77 | 0.548 | 2.21586959962862e-318 |
| COPB1 | 2.07507571253324e-322 | 0.506250612 | 0.611 | 0.381 | 4.43173919925723e-318 |
| PRR11 | 3.50786608547285e-322 | 2.775994029 | 0.106 | 0.012 | 7.49174959874437e-318 |
| BUB1 | 4.15015142506647e-322 | 1.917431182 | 0.131 | 0.023 | 8.86347839851446e-318 |
| NSD2 | 6.37344683135208e-322 | -1.987601073 | 0.2 | 0.176 | 1.36117703977186e-317 |
| GCA | 3.98710976193886e-321 | 0.936177892 | 0.281 | 0.114 | 8.51527031857282e-317 |
| ATP5PD | 4.72820823070073e-321 | 0.332120158 | 0.737 | 0.517 | 1.00980343183075e-316 |
| SGO2 | 7.30723090199204e-321 | 1.94729937 | 0.135 | 0.024 | 1.56060530373844e-316 |
| PCCB | 1.47528001848196e-320 | 0.34962619 | 0.375 | 0.195 | 3.15075553547193e-316 |
| ASAH1 | 7.31217155845045e-320 | 0.70117916 | 0.558 | 0.329 | 1.56166047973826e-315 |
| NDUFA8 | 3.22185148309535e-318 | 0.719965471 | 0.443 | 0.229 | 6.88090821244675e-314 |
| STIP1 | 4.50656050066343e-318 | 0.794700927 | 0.441 | 0.226 | 9.62466126126689e-314 |
| CCDC85B | 1.05325655478906e-317 | 0.451836371 | 0.843 | 0.684 | 2.24944002406299e-313 |
| SEC24D | 2.12213398310268e-317 | 0.522711745 | 0.573 | 0.346 | 4.53224154771239e-313 |
| HIGD1A | 1.76769745471543e-315 | 0.949194347 | 0.359 | 0.162 | 3.77527145403574e-311 |
| APP | 2.24545392441826e-314 | 0.594529126 | 0.386 | 0.189 | 4.79561594638008e-310 |
| MRPS23 | 4.4179662651399e-314 | 0.895635527 | 0.42 | 0.209 | 9.43545055245929e-310 |
| DTNA | 1.12E-306 | 0.434759761 | 0.151 | 0.053 | 2.39E-302 |
| AKAP13 | 4.26E-306 | 0.461951287 | 0.691 | 0.465 | 9.11E-302 |
| MAN1A1 | 5.42E-306 | -0.315836679 | 0.516 | 0.384 | 1.16E-301 |
| KIF15 | 2.58E-305 | 2.380129835 | 0.101 | 0.012 | 5.51E-301 |
| ILF2 | 3.71E-303 | 0.455223739 | 0.643 | 0.42 | 7.92E-299 |
| MYC | 6.07E-303 | -1.519126404 | 0.167 | 0.345 | 1.30E-298 |
| IRF1 | 3.32E-302 | -0.518406639 | 0.853 | 0.841 | 7.09E-298 |
| PSMC5 | 5.77E-302 | 0.610395344 | 0.622 | 0.395 | 1.23E-297 |
| CALU | 1.54E-301 | 0.446080075 | 0.621 | 0.401 | 3.28E-297 |
| MAT2A | 5.89E-300 | 0.759320104 | 0.435 | 0.228 | 1.26E-295 |
| SLC23A1 | 2.86E-299 | 2.291526726 | 0.108 | 0.015 | 6.10E-295 |
| HSP90AA1 | 1.11E-297 | -0.357998727 | 0.896 | 0.784 | 2.36E-293 |
| DDX1 | 5.84E-297 | 0.709561758 | 0.433 | 0.227 | 1.25E-292 |
| COA3 | 1.17E-295 | 0.723999881 | 0.471 | 0.258 | 2.50E-291 |
| FAM227B | 6.55E-295 | 1.011737726 | 0.176 | 0.052 | 1.40E-290 |
| RBM8A | 7.50E-294 | 0.328849066 | 0.712 | 0.501 | 1.60E-289 |
| MED23 | 2.48E-293 | 0.663527052 | 0.271 | 0.115 | 5.30E-289 |
| ZNF22 | 5.47E-291 | 0.691076831 | 0.427 | 0.225 | 1.17E-286 |
| ST6GAL1 | 6.15E-290 | -0.855007798 | 0.447 | 0.475 | 1.31E-285 |
| ECHS1 | 8.59E-289 | 0.756542091 | 0.465 | 0.255 | 1.83E-284 |
| MPHOSPH10 | 1.15E-288 | 0.66713347 | 0.446 | 0.241 | 2.45E-284 |
| SNTB1 | 1.26E-288 | 1.705938989 | 0.102 | 0.016 | 2.69E-284 |
| TRAPPC4 | 2.90E-288 | 0.84268709 | 0.422 | 0.219 | 6.19E-284 |
| ATF3 | 5.35E-288 | -0.451269471 | 0.457 | 0.32 | 1.14E-283 |
| IKBIP | 1.92E-287 | 0.916482329 | 0.372 | 0.179 | 4.11E-283 |
| NDUFB5 | 2.94E-287 | 0.556154812 | 0.623 | 0.401 | 6.29E-283 |
| MT1X | 6.40E-287 | 1.511774189 | 0.222 | 0.073 | 1.37E-282 |
| CEP55 | 2.40E-286 | 2.99652056 | 0.092 | 0.009 | 5.12E-282 |
| UBE2K | 2.95E-286 | 0.530583901 | 0.54 | 0.328 | 6.30E-282 |

| | | | | | |
|---|---|---|---|---|---|
| FUT8 | 7.03E-286 | 0.381302474 | 0.574 | 0.362 | 1.50E-281 |
| COBLL1 | 1.76E-285 | -0.741038412 | 0.48 | 0.435 | 3.76E-281 |
| IL6ST | 8.23E-285 | -1.075453332 | 0.353 | 0.494 | 1.76E-280 |
| VPS11 | 2.18E-284 | 1.118103057 | 0.204 | 0.066 | 4.66E-280 |
| POLB | 5.97E-284 | 1.183501504 | 0.218 | 0.074 | 1.28E-279 |
| TMEM65 | 1.17E-283 | 0.387761212 | 0.261 | 0.121 | 2.50E-279 |
| NAV1 | 1.22E-282 | 0.970902208 | 0.186 | 0.062 | 2.62E-278 |
| ERN1 | 1.76E-282 | -0.332802981 | 0.557 | 0.466 | 3.76E-278 |
| TTK | 1.07E-281 | 2.732618224 | 0.088 | 0.009 | 2.28E-277 |
| TIPRL | 1.87E-280 | 0.645512898 | 0.455 | 0.252 | 4.00E-276 |
| TIMM13 | 7.14E-280 | 0.417330988 | 0.718 | 0.503 | 1.52E-275 |
| UBE2B | 2.02E-279 | -0.272531114 | 0.575 | 0.486 | 4.32E-275 |
| RGS9 | 3.33E-279 | 3.376322132 | 0.072 | 0.004 | 7.11E-275 |
| IFRD2 | 3.64E-278 | 0.360075003 | 0.47 | 0.277 | 7.78E-274 |
| PGK1 | 5.75E-278 | 0.425722968 | 0.671 | 0.462 | 1.23E-273 |
| TTLL7 | 3.75E-277 | -1.490371681 | 0.239 | 0.351 | 8.00E-273 |
| YWHAE | 3.29E-276 | 0.465152343 | 0.807 | 0.612 | 7.02E-272 |
| ALCAM | 1.33E-274 | 0.887967955 | 0.384 | 0.192 | 2.84E-270 |
| IFIH1 | 1.88E-274 | 0.698713062 | 0.358 | 0.178 | 4.02E-270 |
| RAB7A | 2.12E-274 | 0.692152488 | 0.504 | 0.295 | 4.52E-270 |
| SEPTIN2 | 5.42E-274 | 0.431442916 | 0.693 | 0.48 | 1.16E-269 |
| ADGRG5 | 6.43E-274 | 1.640460094 | 0.179 | 0.05 | 1.37E-269 |
| NUP107 | 6.60E-274 | 0.735268548 | 0.256 | 0.109 | 1.41E-269 |
| AHSA1 | 1.82E-273 | 0.934052226 | 0.349 | 0.165 | 3.89E-269 |
| MRPS14 | 2.22E-273 | 0.625716804 | 0.458 | 0.257 | 4.75E-269 |
| IER3IP1 | 5.95E-273 | 0.625332917 | 0.515 | 0.305 | 1.27E-268 |
| COX6B1 | 6.40E-273 | 0.436265576 | 0.864 | 0.683 | 1.37E-268 |
| NUDT6 | 6.53E-273 | 1.207330369 | 0.196 | 0.063 | 1.40E-268 |
| COQ3 | 4.64E-271 | 1.653691357 | 0.129 | 0.027 | 9.91E-267 |
| NEDD8 | 6.57E-271 | 0.309701157 | 0.731 | 0.534 | 1.40E-266 |
| PDCD4-AS1 | 1.32E-270 | 2.008322852 | 0.136 | 0.029 | 2.83E-266 |
| PSMA3 | 1.50E-270 | 0.30964857 | 0.565 | 0.372 | 3.21E-266 |
| TMEM184C | 2.01E-270 | 1.250320913 | 0.213 | 0.073 | 4.29E-266 |
| CYLD | 3.07E-270 | 0.702451094 | 0.462 | 0.259 | 6.56E-266 |
| CYP20A1 | 7.22E-270 | -1.263301275 | 0.289 | 0.33 | 1.54E-265 |
| UMPS | 6.10E-269 | 1.068562934 | 0.226 | 0.083 | 1.30E-264 |
| UQCRC2 | 7.93E-269 | 0.470572543 | 0.595 | 0.385 | 1.69E-264 |
| ABRACL | 2.23E-268 | 0.805719438 | 0.396 | 0.207 | 4.77E-264 |
| HOOK2 | 6.78E-268 | 0.313757459 | 0.292 | 0.149 | 1.45E-263 |
| DLGAP5 | 7.27E-268 | 2.096493 | 0.098 | 0.014 | 1.55E-263 |
| COMMD1 | 7.07E-267 | 0.626463103 | 0.452 | 0.253 | 1.51E-262 |
| USP10 | 1.17E-266 | 0.658794373 | 0.311 | 0.147 | 2.50E-262 |
| AC245014.3 | 3.23E-266 | -1.036894586 | 0.581 | 0.579 | 6.89E-262 |
| NDUFAF4 | 3.89E-266 | 0.483027803 | 0.47 | 0.275 | 8.30E-262 |
| MTIF2 | 9.22E-265 | 0.81172755 | 0.365 | 0.183 | 1.97E-260 |
| NAPG | 1.46E-264 | 0.513750631 | 0.382 | 0.205 | 3.12E-260 |
| LMO4 | 1.94E-264 | -0.562456796 | 0.573 | 0.51 | 4.13E-260 |
| DNAJC12 | 6.61E-264 | 1.006659612 | 0.258 | 0.103 | 1.41E-259 |
| GK5 | 6.75E-264 | 0.351133299 | 0.211 | 0.093 | 1.44E-259 |
| NDNF | 7.89E-263 | -2.272029138 | 0.079 | 0.182 | 1.68E-258 |

| | | | | | |
|---|---|---|---|---|---|
| DCAF7 | 9.88E-263 | 0.883927117 | 0.35 | 0.17 | 2.11E-258 |
| BUB1B | 1.17E-262 | 2.357876507 | 0.094 | 0.013 | 2.50E-258 |
| ZNF714 | 1.37E-262 | 1.809726182 | 0.11 | 0.02 | 2.92E-258 |
| TSPYL1 | 2.65E-262 | 0.347372349 | 0.478 | 0.29 | 5.65E-258 |
| NDUFV3 | 6.07E-262 | 0.427790243 | 0.565 | 0.361 | 1.30E-257 |
| SDC1 | 7.17E-262 | -0.836448893 | 0.576 | 0.536 | 1.53E-257 |
| CCDC66 | 1.25E-261 | 0.412647228 | 0.38 | 0.21 | 2.66E-257 |
| ELMO1 | 2.30E-261 | 0.78250876 | 0.289 | 0.129 | 4.91E-257 |
| SUV39H2 | 3.05E-261 | 1.661342621 | 0.115 | 0.023 | 6.51E-257 |
| CDCA8 | 1.35E-260 | 2.308397527 | 0.09 | 0.011 | 2.88E-256 |
| BAZ2B | 1.69E-259 | -1.207162114 | 0.28 | 0.396 | 3.60E-255 |
| GMDS | 1.69E-259 | 0.58601898 | 0.341 | 0.171 | 3.61E-255 |
| SUMF2 | 2.24E-259 | -1.03496673 | 0.378 | 0.437 | 4.79E-255 |
| TFG | 2.12E-258 | 0.347837432 | 0.52 | 0.332 | 4.54E-254 |
| UBE2L6 | 3.42E-258 | 0.899486838 | 0.421 | 0.227 | 7.31E-254 |
| SPC25 | 2.10E-257 | 2.747418046 | 0.077 | 0.007 | 4.49E-253 |
| KIF23 | 2.51E-257 | 2.633037163 | 0.084 | 0.009 | 5.36E-253 |
| DDX3Y | 3.37E-257 | -1.141576514 | 0.097 | 0.26 | 7.21E-253 |
| GNG5 | 9.93E-257 | 0.374482758 | 0.726 | 0.524 | 2.12E-252 |
| REXO1 | 1.85E-256 | 0.341979844 | 0.295 | 0.151 | 3.96E-252 |
| MCTS1 | 2.30E-254 | 0.347718328 | 0.534 | 0.339 | 4.91E-250 |
| RGS1 | 2.31E-254 | -0.662425942 | 0.422 | 0.317 | 4.94E-250 |
| RPF2 | 1.72E-252 | 0.573084806 | 0.368 | 0.195 | 3.68E-248 |
| TATDN1 | 1.25E-251 | 0.380149217 | 0.493 | 0.302 | 2.67E-247 |
| TRIM28 | 2.47E-251 | 0.26456391 | 0.49 | 0.309 | 5.27E-247 |
| CNKSR1 | 1.39E-250 | 0.620254283 | 0.113 | 0.034 | 2.97E-246 |
| AC109830.1 | 1.92E-249 | 3.604984065 | 0.058 | 0.003 | 4.10E-245 |
| ZFAND1 | 1.57E-248 | 0.447981895 | 0.469 | 0.279 | 3.36E-244 |
| TWSG1 | 2.47E-247 | 1.299730588 | 0.203 | 0.07 | 5.28E-243 |
| LINC02551 | 4.92E-247 | 1.577747691 | 0.107 | 0.022 | 1.05E-242 |
| CYB5B | 4.98E-246 | 0.930446393 | 0.242 | 0.1 | 1.06E-241 |
| ILF3-DT | 1.04E-245 | -0.456067486 | 0.521 | 0.432 | 2.23E-241 |
| MRPS25 | 7.64E-245 | 0.391612407 | 0.382 | 0.218 | 1.63E-240 |
| C11orf58 | 1.58E-242 | 0.43786268 | 0.742 | 0.544 | 3.37E-238 |
| CDKN1C | 1.41E-241 | -1.439594373 | 0.009 | 0.096 | 3.02E-237 |
| MRPL50 | 8.32E-241 | 0.769736823 | 0.372 | 0.194 | 1.78E-236 |
| CCDC107 | 1.88E-240 | -1.450210497 | 0.171 | 0.312 | 4.02E-236 |
| DAP3 | 8.51E-240 | 0.29156205 | 0.565 | 0.379 | 1.82E-235 |
| SLC12A2 | 1.51E-239 | 0.434601661 | 0.222 | 0.102 | 3.23E-235 |
| POLR1E | 3.41E-239 | 0.576017067 | 0.251 | 0.115 | 7.29E-235 |
| POLR3K | 5.36E-239 | 1.035858809 | 0.251 | 0.103 | 1.14E-234 |
| VEGFA | 7.90E-239 | -1.375142619 | 0.059 | 0.197 | 1.69E-234 |
| CEP57L1 | 1.25E-237 | 0.693558555 | 0.197 | 0.078 | 2.67E-233 |
| SNAPC5 | 6.07E-237 | 0.761781951 | 0.337 | 0.169 | 1.30E-232 |
| GPR132 | 3.30E-236 | 3.863545301 | 0.056 | 0.002 | 7.05E-232 |
| ASF1B | 1.06E-235 | 1.688010041 | 0.1 | 0.019 | 2.26E-231 |
| TEDC1 | 1.64E-235 | 1.700586771 | 0.135 | 0.034 | 3.50E-231 |
| TIMP2 | 3.71E-235 | 0.305146475 | 0.386 | 0.238 | 7.93E-231 |
| RAB11A | 4.88E-235 | 0.710842692 | 0.409 | 0.227 | 1.04E-230 |
| COMMD6 | 8.17E-235 | 0.350772003 | 0.663 | 0.475 | 1.75E-230 |

| | | | | | |
|---|---|---|---|---|---|
| CKAP2L | 1.54E-234 | 2.287351814 | 0.081 | 0.01 | 3.29E-230 |
| TERF1 | 4.41E-234 | 0.662282055 | 0.421 | 0.239 | 9.41E-230 |
| TM7SF2 | 2.55E-233 | 0.314258475 | 0.24 | 0.122 | 5.44E-229 |
| KIFC1 | 8.68E-233 | 2.061111091 | 0.089 | 0.014 | 1.85E-228 |
| IRF4 | 1.12E-231 | -0.412114016 | 0.574 | 0.51 | 2.38E-227 |
| INSR | 1.55E-231 | -0.853305693 | 0.389 | 0.417 | 3.31E-227 |
| MNAT1 | 1.66E-231 | 0.526047365 | 0.317 | 0.162 | 3.54E-227 |
| CARMIL1 | 1.73E-231 | -0.86477133 | 0.383 | 0.383 | 3.70E-227 |
| C18orf54 | 1.40E-230 | 2.563386748 | 0.064 | 0.005 | 3.00E-226 |
| IFI27L2 | 4.35E-229 | -0.640908026 | 0.353 | 0.356 | 9.30E-225 |
| RACGAP1 | 3.85E-228 | 2.366925087 | 0.092 | 0.015 | 8.21E-224 |
| SAP30 | 4.16E-228 | 1.076957314 | 0.281 | 0.127 | 8.89E-224 |
| DRG1 | 1.06E-227 | 0.85445376 | 0.299 | 0.142 | 2.26E-223 |
| NCAPH | 2.00E-227 | 2.813927719 | 0.074 | 0.008 | 4.26E-223 |
| KCNQ5 | 3.57E-227 | 0.331819906 | 0.14 | 0.057 | 7.63E-223 |
| IRAK1 | 4.86E-227 | -0.410216977 | 0.493 | 0.414 | 1.04E-222 |
| TK1 | 6.58E-227 | 1.192220862 | 0.102 | 0.023 | 1.40E-222 |
| VAMP2 | 1.49E-226 | -0.310315628 | 0.547 | 0.474 | 3.18E-222 |
| EYA3 | 1.56E-226 | 1.036483305 | 0.19 | 0.069 | 3.33E-222 |
| KIAA1217 | 4.36E-226 | 0.274752639 | 0.534 | 0.356 | 9.31E-222 |
| PHF1 | 1.61E-223 | -1.620503287 | 0.135 | 0.275 | 3.43E-219 |
| ZNF250 | 5.00E-223 | -3.00093543 | 0.036 | 0.139 | 1.07E-218 |
| GATB | 4.32E-222 | 1.255355292 | 0.154 | 0.047 | 9.22E-218 |
| ADAR | 5.09E-221 | -0.533070596 | 0.588 | 0.555 | 1.09E-216 |
| HAUS1 | 3.41E-220 | 0.645570633 | 0.299 | 0.151 | 7.28E-216 |
| RIOX2 | 5.11E-220 | 0.596333944 | 0.356 | 0.192 | 1.09E-215 |
| ECI2 | 7.85E-220 | 0.652257731 | 0.402 | 0.227 | 1.68E-215 |
| OSGEP | 1.59E-219 | 0.643893869 | 0.264 | 0.125 | 3.40E-215 |
| ANKS6 | 2.83E-219 | 0.614829524 | 0.138 | 0.049 | 6.03E-215 |
| TXNDC11 | 2.04E-218 | -0.331871414 | 0.616 | 0.551 | 4.36E-214 |
| NUF2 | 3.35E-218 | 1.948897741 | 0.087 | 0.015 | 7.16E-214 |
| NDUFS1 | 5.64E-217 | 0.427073274 | 0.391 | 0.23 | 1.20E-212 |
| UBB | 7.43E-217 | 0.257188019 | 0.88 | 0.724 | 1.59E-212 |
| SNRNP40 | 2.69E-216 | 0.812347635 | 0.294 | 0.141 | 5.74E-212 |
| JMJD1C | 3.14E-216 | -0.42578567 | 0.617 | 0.548 | 6.71E-212 |
| RTF2 | 5.07E-216 | 0.549099739 | 0.463 | 0.283 | 1.08E-211 |
| CMTM3 | 8.66E-216 | 2.666021342 | 0.081 | 0.011 | 1.85E-211 |
| KMT2B | 1.49E-215 | 0.301558541 | 0.351 | 0.205 | 3.17E-211 |
| TSG101 | 3.51E-215 | 0.706643959 | 0.388 | 0.217 | 7.50E-211 |
| OGFOD1 | 5.89E-214 | 0.896870225 | 0.212 | 0.086 | 1.26E-209 |
| PHAX | 2.26E-213 | 0.476606492 | 0.395 | 0.23 | 4.82E-209 |
| FRAT2 | 2.19E-212 | 0.489963847 | 0.262 | 0.132 | 4.68E-208 |
| DDIAS | 2.47E-212 | 2.186745893 | 0.076 | 0.011 | 5.26E-208 |
| IFI16 | 1.50E-211 | -0.3768509 | 0.621 | 0.563 | 3.20E-207 |
| CDC20 | 3.30E-211 | 1.801284022 | 0.084 | 0.014 | 7.04E-207 |
| EIF5 | 5.30E-211 | -0.326837252 | 0.624 | 0.549 | 1.13E-206 |
| MAP1LC3B | 1.27E-210 | -0.696358405 | 0.397 | 0.431 | 2.72E-206 |
| LYRM1 | 1.41E-210 | 0.519279898 | 0.325 | 0.174 | 3.01E-206 |
| LINC00582 | 6.79E-210 | 0.491760126 | 0.15 | 0.06 | 1.45E-205 |
| PRIM2 | 7.44E-210 | 0.748482732 | 0.157 | 0.057 | 1.59E-205 |

| | | | | | |
|---|---|---|---|---|---|
| COA6 | 9.10E-210 | 0.646385043 | 0.342 | 0.185 | 1.94E-205 |
| TIMMDC1 | 2.41E-209 | 0.520804002 | 0.417 | 0.247 | 5.15E-205 |
| PRPS1 | 8.83E-209 | 0.430000143 | 0.301 | 0.159 | 1.89E-204 |
| AZGP1 | 2.15E-208 | -4.130104481 | 0.013 | 0.095 | 4.59E-204 |
| CENPA | 4.29E-208 | 2.574646706 | 0.066 | 0.007 | 9.16E-204 |
| EXOSC3 | 4.73E-208 | 1.044028548 | 0.225 | 0.094 | 1.01E-203 |
| SLFN13 | 1.34E-206 | 0.355759275 | 0.186 | 0.086 | 2.85E-202 |
| PLGRKT | 4.01E-206 | 0.699697319 | 0.286 | 0.141 | 8.56E-202 |
| ABCG1 | 4.47E-206 | 2.456787824 | 0.065 | 0.007 | 9.55E-202 |
| ISY1 | 2.48E-205 | 0.956362837 | 0.258 | 0.118 | 5.30E-201 |
| EIF4E2 | 9.63E-205 | 0.84816742 | 0.322 | 0.166 | 2.06E-200 |
| NKG7 | 3.97E-203 | 0.478109068 | 0.06 | 0.136 | 8.48E-199 |
| IL5RA | 5.53E-203 | -2.619848221 | 0.027 | 0.127 | 1.18E-198 |
| VPS35 | 8.72E-203 | 0.78763874 | 0.376 | 0.21 | 1.86E-198 |
| EIF3B | 1.39E-202 | 0.344289814 | 0.536 | 0.36 | 2.96E-198 |
| UBALD2 | 2.55E-202 | -0.382577497 | 0.779 | 0.71 | 5.44E-198 |
| CPQ | 1.41E-201 | -1.451709939 | 0.014 | 0.1 | 3.00E-197 |
| MRPS17 | 3.71E-201 | 1.174993539 | 0.184 | 0.069 | 7.92E-197 |
| RTL8A | 1.65E-200 | 0.941020299 | 0.228 | 0.099 | 3.53E-196 |
| AIM2 | 1.87E-200 | -0.33819723 | 0.267 | 0.184 | 3.98E-196 |
| XRN1 | 1.59E-199 | -0.412818783 | 0.59 | 0.523 | 3.40E-195 |
| FMNL2 | 3.74E-199 | 1.560876971 | 0.137 | 0.04 | 7.98E-195 |
| APIP | 9.52E-199 | 0.71644971 | 0.309 | 0.159 | 2.03E-194 |
| SETBP1 | 1.10E-198 | -0.621324859 | 0.375 | 0.333 | 2.36E-194 |
| ZMYND8 | 9.72E-198 | 0.294818578 | 0.404 | 0.255 | 2.08E-193 |
| PDK1 | 9.86E-198 | -0.699909145 | 0.419 | 0.36 | 2.11E-193 |
| CDK1 | 3.75E-197 | 2.072129767 | 0.083 | 0.015 | 8.00E-193 |
| DIAPH3 | 4.21E-197 | 2.476585319 | 0.073 | 0.01 | 8.99E-193 |
| COQ8A | 8.59E-197 | 0.268303708 | 0.322 | 0.187 | 1.84E-192 |
| FNDC3B | 5.81E-196 | -0.426935177 | 0.577 | 0.544 | 1.24E-191 |
| CNN3 | 9.05E-196 | 1.882059866 | 0.118 | 0.03 | 1.93E-191 |
| FAM173B | 9.91E-196 | 0.945147859 | 0.176 | 0.067 | 2.12E-191 |
| MEIS1 | 5.81E-195 | 2.254435529 | 0.064 | 0.008 | 1.24E-190 |
| WASHC3 | 8.40E-195 | 0.550864685 | 0.43 | 0.263 | 1.79E-190 |
| BRD2 | 8.25E-194 | -0.390686753 | 0.619 | 0.563 | 1.76E-189 |
| MFAP1 | 3.60E-192 | 0.906842404 | 0.229 | 0.101 | 7.69E-188 |
| EXOSC8 | 8.22E-192 | 0.639852845 | 0.272 | 0.137 | 1.76E-187 |
| CPSF3 | 1.64E-191 | 0.776894559 | 0.228 | 0.105 | 3.51E-187 |
| BTBD10 | 1.89E-191 | 1.063607044 | 0.141 | 0.047 | 4.04E-187 |
| GLA | 2.04E-191 | 0.750300886 | 0.228 | 0.103 | 4.35E-187 |
| DEPDC1B | 6.79E-191 | 1.966139028 | 0.074 | 0.012 | 1.45E-186 |
| DST | 1.19E-190 | -0.437580971 | 0.701 | 0.62 | 2.53E-186 |
| TMEM70 | 1.35E-190 | 0.521242348 | 0.388 | 0.233 | 2.89E-186 |
| RWDD2B | 2.43E-190 | 0.905728985 | 0.199 | 0.081 | 5.18E-186 |
| GLO1 | 2.68E-190 | -1.008825066 | 0.266 | 0.347 | 5.71E-186 |
| NIP7 | 4.87E-190 | 0.849583734 | 0.253 | 0.119 | 1.04E-185 |
| TSEN34 | 5.59E-190 | 0.424141331 | 0.322 | 0.181 | 1.19E-185 |
| NDUFB1 | 1.09E-189 | -0.420934918 | 0.458 | 0.421 | 2.34E-185 |
| ANKRD36C | 5.61E-188 | -0.377406325 | 0.251 | 0.182 | 1.20E-183 |
| NUDC | 8.76E-188 | 0.393537926 | 0.549 | 0.378 | 1.87E-183 |

| | | | | | |
|---|---|---|---|---|---|
| SLC26A2 | 2.16E-187 | 0.674342163 | 0.193 | 0.084 | 4.62E-183 |
| ARPC1A | 2.22E-187 | 0.521717641 | 0.404 | 0.243 | 4.74E-183 |
| SLC4A1AP | 5.60E-187 | 0.680030339 | 0.262 | 0.13 | 1.20E-182 |
| WDR61 | 2.08E-186 | 0.441275308 | 0.374 | 0.223 | 4.44E-182 |
| MRPL48 | 2.34E-186 | 0.668826929 | 0.286 | 0.147 | 4.99E-182 |
| CHRM3 | 1.04E-185 | 0.262730052 | 0.125 | 0.056 | 2.22E-181 |
| ACP5 | 1.37E-184 | 0.328120367 | 0.103 | 0.04 | 2.93E-180 |
| PPP3CA | 3.67E-184 | 0.283010282 | 0.501 | 0.343 | 7.84E-180 |
| PTPRA | 3.81E-184 | -1.365789203 | 0.14 | 0.26 | 8.14E-180 |
| EBP | 1.20E-182 | 0.54750568 | 0.358 | 0.206 | 2.57E-178 |
| NUP37 | 2.25E-182 | 0.778140669 | 0.234 | 0.109 | 4.80E-178 |
| F2R | 4.43E-182 | 0.386953588 | 0.083 | 0.022 | 9.45E-178 |
| CHCHD1 | 4.99E-182 | 0.449010966 | 0.458 | 0.297 | 1.07E-177 |
| NGRN | 2.67E-180 | 1.090138273 | 0.151 | 0.054 | 5.69E-176 |
| KIAA1143 | 5.34E-180 | 0.587957791 | 0.335 | 0.189 | 1.14E-175 |
| C18orf25 | 3.60E-179 | 0.274794713 | 0.234 | 0.128 | 7.69E-175 |
| DGKZ | 1.35E-178 | 0.452238992 | 0.246 | 0.129 | 2.89E-174 |
| NR4A1 | 1.55E-178 | -1.004626805 | 0.171 | 0.319 | 3.32E-174 |
| ELP6 | 4.59E-178 | 0.795312106 | 0.22 | 0.102 | 9.80E-174 |
| NCAM1 | 5.61E-178 | -0.872196021 | 0.093 | 0.216 | 1.20E-173 |
| JAM2 | 8.69E-178 | 5.579010095 | 0.036 | 0 | 1.86E-173 |
| KIAA1549 | 4.03E-177 | 0.46209321 | 0.081 | 0.027 | 8.61E-173 |
| ACTL6A | 5.80E-177 | 0.533480612 | 0.281 | 0.149 | 1.24E-172 |
| KIF4A | 1.72E-176 | 2.247209078 | 0.064 | 0.009 | 3.68E-172 |
| AF279873.3 | 4.96E-176 | 1.968985804 | 0.068 | 0.011 | 1.06E-171 |
| CPSF1 | 5.91E-176 | 0.313283699 | 0.226 | 0.115 | 1.26E-171 |
| VTA1 | 6.78E-175 | 0.970925125 | 0.216 | 0.096 | 1.45E-170 |
| CDC23 | 6.87E-175 | 0.961530276 | 0.15 | 0.055 | 1.47E-170 |
| ME1 | 1.52E-174 | 1.529053059 | 0.081 | 0.017 | 3.25E-170 |
| MRPS27 | 3.52E-174 | 0.315108844 | 0.245 | 0.132 | 7.51E-170 |
| RPE | 1.39E-173 | 1.099866652 | 0.171 | 0.068 | 2.97E-169 |
| AK6 | 2.42E-173 | 0.459308372 | 0.361 | 0.214 | 5.17E-169 |
| PLEKHB2 | 7.05E-173 | 0.543987966 | 0.264 | 0.142 | 1.51E-168 |
| ZNF331 | 1.05E-172 | -1.601770776 | 0.109 | 0.218 | 2.25E-168 |
| QRSL1 | 3.69E-170 | 0.796641216 | 0.168 | 0.07 | 7.87E-166 |
| JMY | 5.01E-170 | -0.310570747 | 0.431 | 0.344 | 1.07E-165 |
| LINC00920 | 2.04E-169 | 2.083495499 | 0.103 | 0.027 | 4.35E-165 |
| TEX13C | 2.25E-169 | 2.607613552 | 0.051 | 0.005 | 4.81E-165 |
| SLCO3A1 | 3.10E-168 | -0.316300882 | 0.223 | 0.161 | 6.62E-164 |
| HSD17B11 | 1.46E-167 | -0.866617083 | 0.096 | 0.223 | 3.12E-163 |
| BX004987.1 | 9.67E-167 | 0.305537652 | 0.111 | 0.047 | 2.07E-162 |
| LIN7C | 8.58E-166 | 0.455568651 | 0.295 | 0.168 | 1.83E-161 |
| FAM200B | 8.99E-166 | 1.163932978 | 0.149 | 0.056 | 1.92E-161 |
| RAMMET | 2.22E-165 | 0.474984495 | 0.326 | 0.19 | 4.73E-161 |
| CIAPIN1 | 2.88E-165 | 0.925415176 | 0.212 | 0.096 | 6.14E-161 |
| ELP3 | 5.24E-165 | 0.866752415 | 0.162 | 0.065 | 1.12E-160 |
| DIRAS2 | 6.72E-164 | -4.629068099 | 0.001 | 0.055 | 1.43E-159 |
| HPF1 | 7.20E-164 | 0.722795521 | 0.268 | 0.139 | 1.54E-159 |
| FAM96A | 8.37E-164 | 0.336766074 | 0.426 | 0.28 | 1.79E-159 |
| NOB1 | 9.59E-164 | 0.260428175 | 0.329 | 0.204 | 2.05E-159 |

| | | | | | |
|---|---|---|---|---|---|
| MYNN | 1.73E-163 | 0.35538199 | 0.252 | 0.142 | 3.69E-159 |
| AGK | 1.44E-162 | 0.910500019 | 0.154 | 0.061 | 3.08E-158 |
| DCXR | 2.76E-162 | -0.585621364 | 0.363 | 0.361 | 5.90E-158 |
| MPV17 | 1.25E-161 | 0.67911144 | 0.235 | 0.119 | 2.67E-157 |
| AC109588.1 | 1.54E-161 | 1.723364868 | 0.055 | 0.008 | 3.29E-157 |
| ARF3 | 4.32E-161 | 0.608858456 | 0.259 | 0.139 | 9.23E-157 |
| NNT-AS1 | 3.38E-160 | 0.417362482 | 0.206 | 0.107 | 7.23E-156 |
| OSGIN2 | 8.34E-160 | 0.986521812 | 0.149 | 0.057 | 1.78E-155 |
| BDH2 | 3.85E-159 | 0.656994033 | 0.144 | 0.06 | 8.23E-155 |
| PTCH2 | 7.83E-159 | -1.703423067 | 0.085 | 0.175 | 1.67E-154 |
| KANK2 | 8.49E-159 | 1.270270356 | 0.069 | 0.015 | 1.81E-154 |
| MRPL44 | 8.75E-159 | 0.728092717 | 0.248 | 0.127 | 1.87E-154 |
| CENPN | 1.34E-158 | 1.100162464 | 0.123 | 0.042 | 2.86E-154 |
| FNIP1 | 1.76E-158 | -0.453642353 | 0.329 | 0.277 | 3.75E-154 |
| GBP4 | 1.35E-157 | 0.725850105 | 0.13 | 0.049 | 2.88E-153 |
| SYPL1 | 3.43E-157 | 0.316516739 | 0.38 | 0.242 | 7.32E-153 |
| WASHC4 | 4.78E-157 | 0.752789602 | 0.306 | 0.171 | 1.02E-152 |
| AL139099.1 | 6.11E-157 | -2.857695343 | 0.061 | 0.116 | 1.31E-152 |
| USP39 | 7.03E-157 | 1.139177362 | 0.174 | 0.073 | 1.50E-152 |
| ISLR | 8.39E-157 | -7.109377038 | 0.001 | 0.051 | 1.79E-152 |
| FASN | 1.06E-156 | 0.299733337 | 0.12 | 0.051 | 2.27E-152 |
| CD33 | 2.98E-155 | 3.251457503 | 0.054 | 0.006 | 6.37E-151 |
| SMARCE1 | 5.50E-155 | 0.430813327 | 0.309 | 0.183 | 1.18E-150 |
| TROAP | 8.26E-155 | 1.843016427 | 0.063 | 0.012 | 1.77E-150 |
| SLC16A14 | 1.04E-154 | 1.524765338 | 0.081 | 0.019 | 2.23E-150 |
| HMGCS1 | 1.15E-154 | 0.797313467 | 0.277 | 0.148 | 2.46E-150 |
| CENPS | 1.33E-154 | 1.453943495 | 0.099 | 0.028 | 2.83E-150 |
| MRPL27 | 2.60E-154 | 0.397515714 | 0.352 | 0.219 | 5.54E-150 |
| ARHGAP4 | 6.22E-153 | -0.50625973 | 0.324 | 0.282 | 1.33E-148 |
| KMT2E-AS1 | 2.20E-152 | -1.480924046 | 0.127 | 0.222 | 4.70E-148 |
| SHC1 | 2.83E-152 | -0.66060915 | 0.317 | 0.31 | 6.05E-148 |
| ARL4C | 5.17E-152 | 0.341665125 | 0.414 | 0.274 | 1.10E-147 |
| ITPR1 | 6.99E-152 | 0.832081696 | 0.099 | 0.033 | 1.49E-147 |
| AL391069.3 | 1.44E-151 | 1.968697793 | 0.058 | 0.009 | 3.08E-147 |
| POLD1 | 1.55E-151 | 0.409186556 | 0.098 | 0.037 | 3.30E-147 |
| PTPN18 | 3.61E-151 | 0.732420305 | 0.267 | 0.144 | 7.70E-147 |
| UBXN8 | 5.20E-151 | 0.610609095 | 0.137 | 0.056 | 1.11E-146 |
| SV2C | 6.41E-151 | -2.002518057 | 0.008 | 0.064 | 1.37E-146 |
| CASP1 | 8.62E-151 | 0.292435966 | 0.104 | 0.187 | 1.84E-146 |
| SRP14-AS1 | 2.66E-150 | 0.935424535 | 0.147 | 0.059 | 5.67E-146 |
| PPIL3 | 2.86E-150 | 0.669036171 | 0.205 | 0.1 | 6.10E-146 |
| ARIH1 | 1.19E-149 | -0.397499869 | 0.41 | 0.349 | 2.53E-145 |
| VASH2 | 1.32E-149 | 2.375007384 | 0.052 | 0.006 | 2.82E-145 |
| SYNJ2BP | 1.93E-149 | 0.690966561 | 0.181 | 0.083 | 4.12E-145 |
| ARL15 | 2.06E-149 | 1.618433519 | 0.065 | 0.012 | 4.40E-145 |
| RIN3 | 5.16E-149 | 0.327095043 | 0.011 | 0.056 | 1.10E-144 |
| MELK | 2.28E-148 | 2.693173552 | 0.053 | 0.007 | 4.88E-144 |
| C16orf54 | 2.54E-148 | -1.29687247 | 0.151 | 0.255 | 5.42E-144 |
| C1orf56 | 2.78E-148 | -1.312334964 | 0.157 | 0.266 | 5.93E-144 |
| RALB | 8.82E-148 | 1.591530575 | 0.134 | 0.047 | 1.88E-143 |

| | | | | | |
|---|---|---|---|---|---|
| GGCX | 1.60E-147 | 0.440524621 | 0.238 | 0.128 | 3.41E-143 |
| HACD3 | 3.04E-147 | 0.281258743 | 0.363 | 0.236 | 6.50E-143 |
| WDR92 | 7.38E-147 | 0.872115814 | 0.126 | 0.047 | 1.58E-142 |
| POP4 | 9.25E-147 | 0.5335257 | 0.322 | 0.19 | 1.98E-142 |
| TSFM | 1.29E-145 | 0.773896837 | 0.232 | 0.118 | 2.76E-141 |
| CDK5RAP1 | 7.59E-145 | 0.456882518 | 0.177 | 0.086 | 1.62E-140 |
| PAAF1 | 8.52E-145 | 0.5985223 | 0.235 | 0.123 | 1.82E-140 |
| ARL2BP | 3.14E-144 | 0.472118444 | 0.32 | 0.192 | 6.71E-140 |
| LYRM2 | 3.40E-144 | 0.273698381 | 0.349 | 0.226 | 7.26E-140 |
| ARID5A | 1.10E-143 | -1.048312441 | 0.23 | 0.294 | 2.35E-139 |
| FGFR1OP | 1.22E-143 | 1.050118939 | 0.132 | 0.05 | 2.60E-139 |
| METTL7A | 1.48E-142 | -0.266729327 | 0.578 | 0.519 | 3.17E-138 |
| ID1 | 2.08E-142 | -0.926981631 | 0.253 | 0.275 | 4.45E-138 |
| TBC1D23 | 4.36E-142 | 0.618531913 | 0.246 | 0.132 | 9.32E-138 |
| RFC5 | 6.72E-142 | 1.104194609 | 0.107 | 0.035 | 1.43E-137 |
| AC084782.3 | 7.54E-142 | 2.9142379 | 0.041 | 0.003 | 1.61E-137 |
| NARS2 | 1.42E-141 | 0.827942702 | 0.136 | 0.056 | 3.02E-137 |
| AP3M1 | 3.28E-141 | 0.603264424 | 0.203 | 0.102 | 7.01E-137 |
| PRIM1 | 4.88E-141 | 0.875228541 | 0.174 | 0.078 | 1.04E-136 |
| EXOSC10 | 8.16E-141 | 0.414945011 | 0.185 | 0.094 | 1.74E-136 |
| SAXO2 | 1.78E-140 | 0.507409202 | 0.076 | 0.024 | 3.81E-136 |
| SSBP3 | 7.48E-140 | -1.452107257 | 0.091 | 0.186 | 1.60E-135 |
| LINC02506 | 8.37E-140 | 1.257649647 | 0.052 | 0.011 | 1.79E-135 |
| DROSHA | 1.23E-139 | 0.651411471 | 0.132 | 0.054 | 2.62E-135 |
| INTS14 | 1.74E-139 | 0.730933238 | 0.167 | 0.075 | 3.73E-135 |
| BICD1 | 3.14E-139 | -0.304987142 | 0.302 | 0.233 | 6.71E-135 |
| PEX3 | 4.90E-138 | 0.870026789 | 0.17 | 0.076 | 1.05E-133 |
| ARMC1 | 1.75E-137 | 0.560011785 | 0.305 | 0.18 | 3.74E-133 |
| ABCF2.1 | 3.89E-137 | 0.526510067 | 0.194 | 0.098 | 8.30E-133 |
| BMP8B | 2.43E-136 | -1.044465093 | 0.218 | 0.249 | 5.19E-132 |
| TGFBR2 | 2.47E-136 | 0.858625057 | 0.143 | 0.058 | 5.28E-132 |
| CDHR1 | 3.68E-136 | -3.783167263 | 0 | 0.043 | 7.86E-132 |
| ARNTL2 | 5.54E-136 | 4.050272217 | 0.041 | 0.003 | 1.18E-131 |
| GNL1 | 1.23E-135 | -0.301318693 | 0.37 | 0.299 | 2.62E-131 |
| METTL6 | 3.69E-135 | 0.878470357 | 0.167 | 0.075 | 7.88E-131 |
| CA13 | 1.94E-134 | 0.282977667 | 0.093 | 0.041 | 4.15E-130 |
| SLIT2 | 3.53E-134 | 1.927218572 | 0.048 | 0.007 | 7.53E-130 |
| SLC12A9 | 4.58E-134 | 0.306741195 | 0.146 | 0.074 | 9.79E-130 |
| TSPAN3 | 5.30E-134 | -0.743451786 | 0.328 | 0.35 | 1.13E-129 |
| SLC10A7 | 9.66E-134 | 0.457203378 | 0.198 | 0.103 | 2.06E-129 |
| SCN9A | 2.54E-133 | 0.252576785 | 0.165 | 0.087 | 5.42E-129 |
| SCAMP1 | 5.18E-133 | 0.289081439 | 0.246 | 0.144 | 1.11E-128 |
| ZDHHC24 | 5.28E-133 | 0.266870779 | 0.292 | 0.181 | 1.13E-128 |
| E2F7 | 3.57E-132 | 1.51607586 | 0.053 | 0.01 | 7.63E-128 |
| FBXW9 | 7.52E-132 | 0.512613677 | 0.11 | 0.045 | 1.61E-127 |
| TIMM22 | 4.26E-131 | 0.723126263 | 0.188 | 0.092 | 9.10E-127 |
| AL441992.1 | 9.06E-131 | 0.92404528 | 0.116 | 0.044 | 1.93E-126 |
| TIPARP | 1.02E-130 | -0.301460275 | 0.398 | 0.337 | 2.17E-126 |
| NCKAP1 | 1.07E-130 | -2.983578268 | 0.013 | 0.074 | 2.29E-126 |
| SMAD3 | 1.56E-130 | -2.017930744 | 0.034 | 0.109 | 3.34E-126 |

| | | | | | |
|---|---|---|---|---|---|
| FANCL | 1.59E-130 | 0.989124337 | 0.123 | 0.047 | 3.39E-126 |
| P2RY11 | 4.63E-130 | 0.362041651 | 0.107 | 0.048 | 9.88E-126 |
| HADHB | 5.15E-130 | 0.330882119 | 0.37 | 0.25 | 1.10E-125 |
| ANKRD36 | 1.44E-129 | -0.351609472 | 0.266 | 0.208 | 3.08E-125 |
| RGS16 | 2.10E-129 | 0.745766736 | 0.177 | 0.085 | 4.48E-125 |
| AP1S1 | 2.11E-129 | 0.564983219 | 0.262 | 0.15 | 4.50E-125 |
| P2RX5-TAX1BP3 | 2.71E-129 | 1.894722864 | 0.064 | 0.013 | 5.79E-125 |
| CD1D | 4.85E-129 | -1.210531923 | 0.109 | 0.128 | 1.04E-124 |
| LGALS1 | 1.54E-128 | -0.307736502 | 0.455 | 0.465 | 3.28E-124 |
| ZNF90 | 1.56E-128 | 0.321647775 | 0.08 | 0.033 | 3.33E-124 |
| SPAG5 | 1.83E-128 | 1.289127444 | 0.063 | 0.014 | 3.91E-124 |
| RRM2B | 6.70E-128 | 0.954303528 | 0.147 | 0.062 | 1.43E-123 |
| TTN-AS1 | 1.59E-127 | -0.26570996 | 0.087 | 0.049 | 3.39E-123 |
| ARF6 | 4.28E-127 | -0.386755667 | 0.362 | 0.374 | 9.13E-123 |
| AP002748.3 | 4.67E-126 | 1.154497134 | 0.098 | 0.032 | 9.97E-122 |
| DLST | 7.50E-126 | 0.784237785 | 0.176 | 0.083 | 1.60E-121 |
| PBXIP1 | 8.83E-126 | -1.127597744 | 0.187 | 0.233 | 1.89E-121 |
| CADM2 | 2.26E-125 | -6.896708522 | 0 | 0.038 | 4.84E-121 |
| FGD2 | 2.46E-125 | -1.399779621 | 0.04 | 0.121 | 5.26E-121 |
| ERVMER61-1 | 2.60E-125 | 1.768972121 | 0.043 | 0.007 | 5.55E-121 |
| CFAP97 | 9.49E-125 | -0.283381836 | 0.376 | 0.323 | 2.03E-120 |
| CSNK1G3 | 1.07E-124 | -0.542677467 | 0.308 | 0.28 | 2.30E-120 |
| GTF2F1 | 1.11E-124 | -0.342732197 | 0.327 | 0.269 | 2.37E-120 |
| ASF1A | 1.24E-124 | 0.806312452 | 0.187 | 0.091 | 2.65E-120 |
| MPHOSPH6 | 2.51E-124 | 0.825950815 | 0.149 | 0.067 | 5.37E-120 |
| TMEM64 | 2.88E-123 | 0.637529014 | 0.231 | 0.124 | 6.16E-119 |
| SMIM12 | 4.27E-123 | 0.499122402 | 0.246 | 0.14 | 9.13E-119 |
| SNRNP35 | 5.43E-123 | 0.432794419 | 0.181 | 0.098 | 1.16E-118 |
| PKNOX1 | 5.84E-123 | 0.63806744 | 0.181 | 0.091 | 1.25E-118 |
| TRMU | 1.68E-122 | 0.29455292 | 0.143 | 0.073 | 3.58E-118 |
| SELPLG | 1.73E-122 | -0.684908612 | 0.142 | 0.253 | 3.70E-118 |
| B4GALT7 | 2.15E-122 | -1.357837395 | 0.092 | 0.185 | 4.58E-118 |
| SLC30A9 | 2.32E-122 | 0.459795071 | 0.222 | 0.125 | 4.96E-118 |
| RPP25 | 1.15E-121 | -1.688982944 | 0.021 | 0.087 | 2.46E-117 |
| CLIC2 | 2.06E-121 | -1.20567701 | 0.027 | 0.094 | 4.41E-117 |
| SLC30A7 | 3.96E-121 | 0.294954946 | 0.282 | 0.177 | 8.45E-117 |
| PIGF | 5.84E-121 | 0.376922275 | 0.28 | 0.173 | 1.25E-116 |
| ALAS1 | 1.47E-120 | 0.693922585 | 0.162 | 0.078 | 3.14E-116 |
| INTS4 | 1.81E-120 | 0.791989314 | 0.124 | 0.052 | 3.87E-116 |
| PLEKHA7 | 2.13E-120 | 0.631775306 | 0.133 | 0.058 | 4.56E-116 |
| CHMP5 | 2.16E-120 | 0.45573438 | 0.446 | 0.313 | 4.61E-116 |
| CBX5 | 1.14E-119 | 0.258152313 | 0.32 | 0.211 | 2.44E-115 |
| ABCD3 | 6.40E-119 | 0.559968118 | 0.136 | 0.065 | 1.37E-114 |
| AL133467.1 | 3.59E-118 | -2.036691884 | 0.028 | 0.098 | 7.66E-114 |
| C22orf39 | 7.68E-118 | 0.369240903 | 0.263 | 0.16 | 1.64E-113 |
| AC104024.1 | 7.75E-118 | -2.409397448 | 0.033 | 0.1 | 1.65E-113 |
| RARS2 | 1.36E-117 | 0.42910486 | 0.244 | 0.144 | 2.90E-113 |
| SAC3D1 | 2.16E-117 | 0.26149293 | 0.187 | 0.108 | 4.61E-113 |
| RANGAP1 | 2.29E-117 | 0.475695977 | 0.181 | 0.095 | 4.90E-113 |
| GNB5 | 1.24E-116 | -0.649105434 | 0.196 | 0.17 | 2.65E-112 |

| | | | | | | |
|---|---|---|---|---|---|---|
| STYXL1 | 2.29E-116 | 0.652896852 | 0.2 | 0.106 | | 4.89E-112 |
| GRIP1 | 2.59E-116 | 0.463926894 | 0.079 | 0.031 | | 5.54E-112 |
| SAP30BP | 3.84E-116 | 0.329418777 | 0.307 | 0.199 | | 8.20E-112 |
| RAB28 | 6.21E-116 | 0.658333805 | 0.164 | 0.08 | | 1.33E-111 |
| SLC9A9 | 9.51E-116 | -2.192980825 | 0.019 | 0.08 | | 2.03E-111 |
| GIT2 | 9.80E-116 | 0.319963046 | 0.189 | 0.107 | | 2.09E-111 |
| WDR4 | 2.07E-115 | 0.289362629 | 0.131 | 0.066 | | 4.43E-111 |
| PDHX | 2.79E-115 | 0.569793826 | 0.137 | 0.064 | | 5.96E-111 |
| MYO1B | 2.85E-115 | -3.289723187 | 0.004 | 0.049 | | 6.09E-111 |
| ALOX12-AS1 | 3.12E-115 | 0.908879893 | 0.102 | 0.037 | | 6.66E-111 |
| VWDE | 8.13E-115 | 0.288920797 | 0.052 | 0.018 | | 1.74E-110 |
| PCNA | 6.21E-114 | 0.918548488 | 0.248 | 0.14 | | 1.33E-109 |
| KPNA5 | 1.61E-113 | 0.398489791 | 0.13 | 0.067 | | 3.43E-109 |
| SEC23IP | 1.76E-113 | 0.575773106 | 0.188 | 0.1 | | 3.75E-109 |
| SNX18 | 3.54E-113 | -0.794969998 | 0.101 | 0.202 | | 7.56E-109 |
| CNN2 | 4.09E-113 | -0.301559006 | 0.412 | 0.389 | | 8.74E-109 |
| ANKRD18B | 6.06E-113 | 4.260079394 | 0.031 | 0.002 | | 1.29E-108 |
| TNFRSF10D | 9.84E-113 | 1.043733533 | 0.09 | 0.031 | | 2.10E-108 |
| PDK4 | 1.35E-112 | -1.585635039 | 0.002 | 0.035 | | 2.88E-108 |
| ANLN | 3.59E-112 | 1.704160244 | 0.045 | 0.008 | | 7.67E-108 |
| AC132192.1 | 5.92E-112 | 1.146764508 | 0.042 | 0.008 | | 1.26E-107 |
| AURKA | 8.41E-112 | 1.830316785 | 0.057 | 0.013 | | 1.80E-107 |
| CCDC15 | 1.43E-111 | 1.981154596 | 0.052 | 0.01 | | 3.06E-107 |
| ADAMTS15 | 2.23E-111 | 0.647027198 | 0.042 | 0.01 | | 4.76E-107 |
| MUT | 1.26E-110 | 0.506566994 | 0.184 | 0.098 | | 2.69E-106 |
| GBP3 | 1.34E-110 | -1.873075516 | 0.032 | 0.099 | | 2.86E-106 |
| RSBN1L | 2.15E-110 | -0.437951247 | 0.377 | 0.353 | | 4.60E-106 |
| MTCH1 | 1.11E-109 | -0.686012376 | 0.275 | 0.316 | | 2.38E-105 |
| IFT22 | 1.75E-109 | 1.163623099 | 0.078 | 0.025 | | 3.73E-105 |
| RGS13 | 2.69E-109 | -4.288124745 | 0.008 | 0.046 | | 5.74E-105 |
| KLHL24 | 7.21E-109 | -0.549806838 | 0.348 | 0.341 | | 1.54E-104 |
| MTFR2 | 7.91E-109 | 1.857583693 | 0.049 | 0.009 | | 1.69E-104 |
| BCAS2 | 8.07E-109 | -0.767614298 | 0.277 | 0.303 | | 1.72E-104 |
| EFCAB7 | 1.92E-108 | 0.515469317 | 0.096 | 0.04 | | 4.09E-104 |
| SQOR | 2.03E-108 | 0.482535942 | 0.314 | 0.203 | | 4.33E-104 |
| XRRA1 | 2.49E-108 | -1.022660317 | 0.187 | 0.237 | | 5.32E-104 |
| RAB5A | 2.96E-108 | 0.394264009 | 0.325 | 0.213 | | 6.32E-104 |
| SLC25A37 | 5.00E-108 | -0.327788771 | 0.232 | 0.221 | | 1.07E-103 |
| FBXO41 | 8.67E-108 | 0.566760663 | 0.087 | 0.034 | | 1.85E-103 |
| MRPL45 | 1.63E-107 | 0.253627853 | 0.274 | 0.175 | | 3.48E-103 |
| EIF4E | 9.07E-107 | -1.042966335 | 0.222 | 0.297 | | 1.94E-102 |
| SAMHD1 | 1.10E-106 | 1.26427067 | 0.211 | 0.115 | | 2.36E-102 |
| STN1 | 1.27E-106 | 0.545315138 | 0.176 | 0.093 | | 2.70E-102 |
| ACSF2 | 1.82E-106 | 1.157629051 | 0.058 | 0.015 | | 3.89E-102 |
| CCDC141 | 6.42E-106 | 2.31371171 | 0.032 | 0.003 | | 1.37E-101 |
| KIF20A | 1.28E-105 | 2.093690288 | 0.034 | 0.004 | | 2.74E-101 |
| C11orf54 | 2.11E-105 | 0.456352901 | 0.181 | 0.101 | | 4.50E-101 |
| NEGR1 | 2.23E-105 | 4.088276994 | 0.025 | 0.001 | | 4.76E-101 |
| DUSP26 | 3.67E-105 | -3.159885614 | 0.011 | 0.061 | | 7.83E-101 |
| APTX | 4.78E-105 | 0.37684666 | 0.219 | 0.132 | | 1.02E-100 |

| | | | | | |
|---|---|---|---|---|---|
| APOOL | 6.31E-105 | 0.361332453 | 0.211 | 0.125 | 1.35E-100 |
| CHAF1B | 9.79E-105 | 1.739200647 | 0.052 | 0.011 | 2.09E-100 |
| MT2A | 1.14E-104 | 1.815782631 | 0.167 | 0.085 | 2.44E-100 |
| INTS7 | 1.47E-104 | 0.435609684 | 0.111 | 0.053 | 3.15E-100 |
| MRPS28 | 7.59E-104 | 0.795240631 | 0.173 | 0.088 | 1.62E-99 |
| LINC01287 | 8.23E-104 | 2.479423479 | 0.05 | 0.01 | 1.76E-99 |
| AASDH | 2.24E-103 | 0.495883527 | 0.151 | 0.078 | 4.78E-99 |
| PRRG2 | 3.38E-103 | 0.311371874 | 0.068 | 0.027 | 7.22E-99 |
| NXT2 | 3.86E-103 | 0.724099527 | 0.122 | 0.054 | 8.24E-99 |
| RGPD2 | 4.72E-103 | -4.523270929 | 0.002 | 0.04 | 1.01E-98 |
| TSPYL2 | 1.26E-102 | -1.05738125 | 0.192 | 0.243 | 2.68E-98 |
| TST | 1.48E-102 | 0.267279389 | 0.203 | 0.122 | 3.16E-98 |
| KIAA0586 | 3.15E-102 | -1.050496259 | 0.15 | 0.195 | 6.73E-98 |
| FUCA1 | 3.57E-102 | 0.457376332 | 0.163 | 0.089 | 7.61E-98 |
| MAP1A | 4.06E-102 | 1.394061771 | 0.044 | 0.01 | 8.66E-98 |
| DHRS11 | 5.14E-102 | 0.334252556 | 0.135 | 0.069 | 1.10E-97 |
| SCAMP5 | 1.27E-101 | -0.924081449 | 0.15 | 0.149 | 2.70E-97 |
| CNTNAP1 | 1.49E-100 | 1.666192219 | 0.044 | 0.008 | 3.18E-96 |
| ARHGEF2 | 1.78E-100 | -0.80486538 | 0.231 | 0.271 | 3.80E-96 |
| RNF14 | 2.24E-100 | 0.276632426 | 0.176 | 0.102 | 4.78E-96 |
| AHI1 | 5.39E-100 | -1.203849105 | 0.157 | 0.184 | 1.15E-95 |
| SLC19A1 | 1.22E-99 | 0.568147104 | 0.08 | 0.033 | 2.60E-95 |
| AC008074.3 | 1.66E-99 | -1.865839527 | 0.048 | 0.111 | 3.54E-95 |
| TFB1M | 2.50E-99 | 0.725513017 | 0.137 | 0.064 | 5.35E-95 |
| AC079630.1 | 1.33E-98 | -3.462498609 | 0.003 | 0.04 | 2.85E-94 |
| MXI1 | 1.54E-98 | -0.418953266 | 0.296 | 0.267 | 3.30E-94 |
| PBX2 | 3.17E-98 | -1.136396052 | 0.115 | 0.188 | 6.77E-94 |
| AC010343.3 | 7.56E-98 | 1.303420855 | 0.034 | 0.006 | 1.61E-93 |
| AL135791.1 | 1.16E-97 | -2.885813567 | 0.009 | 0.055 | 2.47E-93 |
| DYNC2H1 | 1.29E-97 | 1.425932051 | 0.039 | 0.007 | 2.76E-93 |
| AC005261.1 | 8.71E-97 | -0.272933272 | 0.443 | 0.398 | 1.86E-92 |
| FOXM1 | 1.25E-96 | 1.423635826 | 0.046 | 0.01 | 2.67E-92 |
| DBN1 | 1.29E-96 | 0.729192328 | 0.165 | 0.084 | 2.75E-92 |
| TRMT9B | 1.30E-96 | -3.925776419 | 0.006 | 0.046 | 2.78E-92 |
| SPARC | 3.31E-96 | 0.879209763 | 0.115 | 0.05 | 7.07E-92 |
| BCR | 4.91E-96 | 0.264729728 | 0.151 | 0.084 | 1.05E-91 |
| PTMS | 9.31E-96 | 0.336486584 | 0.22 | 0.136 | 1.99E-91 |
| B3GALNT1 | 9.33E-96 | 1.296936223 | 0.046 | 0.011 | 1.99E-91 |
| TTC5 | 2.84E-95 | 0.402173406 | 0.132 | 0.07 | 6.06E-91 |
| MOB3B | 6.11E-95 | 1.822037765 | 0.036 | 0.006 | 1.31E-90 |
| AL451085.1 | 2.94E-94 | -2.262350573 | 0.03 | 0.082 | 6.29E-90 |
| TRIP4 | 3.23E-94 | 0.744308769 | 0.133 | 0.064 | 6.91E-90 |
| YPEL3 | 3.62E-94 | -0.466162306 | 0.33 | 0.388 | 7.74E-90 |
| FGFR1OP2 | 6.62E-94 | -0.740225687 | 0.252 | 0.313 | 1.41E-89 |
| MELTF-AS1 | 6.68E-94 | 0.540053293 | 0.049 | 0.016 | 1.43E-89 |
| PHOSPHO2 | 1.86E-93 | 0.367157758 | 0.075 | 0.032 | 3.97E-89 |
| PIAS4 | 2.69E-93 | 0.306502877 | 0.127 | 0.069 | 5.75E-89 |
| ETFRF1 | 2.75E-93 | -0.810495154 | 0.213 | 0.271 | 5.87E-89 |
| GGT1 | 3.77E-93 | 0.534563417 | 0.064 | 0.025 | 8.04E-89 |
| SUN1 | 4.47E-93 | -0.764122276 | 0.219 | 0.223 | 9.54E-89 |

| | | | | | |
|---|---|---|---|---|---|
| RSU1 | 5.08E-93 | 0.491940683 | 0.251 | 0.157 | 1.08E-88 |
| HIC2 | 6.07E-93 | 0.285312827 | 0.09 | 0.043 | 1.30E-88 |
| A1BG | 6.59E-93 | -1.225866279 | 0.058 | 0.133 | 1.41E-88 |
| TSIX | 1.37E-92 | -5.056375028 | 0.001 | 0.033 | 2.92E-88 |
| CBX4 | 2.50E-92 | -0.679626279 | 0.234 | 0.247 | 5.34E-88 |
| SERINC1 | 3.42E-92 | -0.503599752 | 0.289 | 0.336 | 7.31E-88 |
| NUP85 | 4.00E-92 | 0.803423243 | 0.091 | 0.038 | 8.53E-88 |
| PSMD10 | 1.39E-91 | 0.471090034 | 0.21 | 0.122 | 2.98E-87 |
| TXNDC9 | 2.19E-91 | 0.475581356 | 0.24 | 0.148 | 4.67E-87 |
| CIT | 3.64E-91 | 1.730693483 | 0.037 | 0.007 | 7.77E-87 |
| KCTD10 | 1.07E-90 | 0.746613685 | 0.09 | 0.036 | 2.28E-86 |
| GTF2H3 | 2.67E-90 | 0.666595546 | 0.201 | 0.115 | 5.70E-86 |
| WDSUB1 | 4.70E-90 | 0.933224082 | 0.083 | 0.032 | 1.00E-85 |
| ZC2HC1A | 4.85E-90 | -0.900651647 | 0.033 | 0.092 | 1.04E-85 |
| UBE2Z | 1.48E-89 | 0.31396253 | 0.265 | 0.175 | 3.16E-85 |
| PRKAB2 | 3.57E-89 | 0.341187134 | 0.094 | 0.046 | 7.63E-85 |
| TNIP1 | 4.85E-89 | -0.578271279 | 0.272 | 0.302 | 1.04E-84 |
| C11orf49 | 1.76E-88 | 0.415435133 | 0.107 | 0.053 | 3.75E-84 |
| XKR9 | 4.96E-88 | -1.85938902 | 0.019 | 0.07 | 1.06E-83 |
| ANXA1 | 4.48E-87 | 0.608538841 | 0.067 | 0.103 | 9.58E-83 |
| KCNMB3 | 1.42E-86 | -2.523739102 | 0.009 | 0.05 | 3.04E-82 |
| CYB5A | 1.80E-86 | -1.158325656 | 0.073 | 0.144 | 3.85E-82 |
| BRMS1L | 2.42E-86 | 0.637418469 | 0.108 | 0.05 | 5.17E-82 |
| ARHGAP5 | 2.58E-86 | -0.81009234 | 0.241 | 0.293 | 5.51E-82 |
| ZNF275 | 5.08E-86 | -0.923790795 | 0.178 | 0.226 | 1.09E-81 |
| LINC00865 | 7.14E-86 | 2.053654329 | 0.035 | 0.006 | 1.53E-81 |
| XPR1 | 1.17E-85 | -1.292113309 | 0.088 | 0.149 | 2.50E-81 |
| TNFRSF14 | 1.64E-85 | -0.657973829 | 0.267 | 0.323 | 3.49E-81 |
| RFWD3 | 2.18E-85 | 0.854882973 | 0.111 | 0.051 | 4.65E-81 |
| SNTB2 | 3.79E-85 | 1.060106778 | 0.079 | 0.029 | 8.10E-81 |
| TRIM35 | 5.49E-85 | 0.936706461 | 0.066 | 0.023 | 1.17E-80 |
| TMEM8B | 1.26E-84 | 0.419688615 | 0.071 | 0.031 | 2.68E-80 |
| STEAP1B | 1.91E-84 | 1.359894259 | 0.035 | 0.007 | 4.07E-80 |
| AC005726.5 | 2.69E-84 | 0.687387403 | 0.054 | 0.018 | 5.75E-80 |
| NR1D1 | 3.96E-84 | -1.729154414 | 0.051 | 0.102 | 8.45E-80 |
| ARL6 | 1.63E-83 | 1.161102838 | 0.045 | 0.011 | 3.47E-79 |
| GMPR | 3.37E-83 | 0.852533024 | 0.066 | 0.027 | 7.20E-79 |
| AC005670.2 | 6.60E-83 | 1.283492521 | 0.039 | 0.009 | 1.41E-78 |
| PDLIM2 | 1.51E-82 | 1.479749247 | 0.056 | 0.017 | 3.23E-78 |
| CREM | 3.93E-82 | 0.291428166 | 0.23 | 0.148 | 8.39E-78 |
| APOO | 4.11E-82 | 0.571610116 | 0.124 | 0.062 | 8.79E-78 |
| JRK | 6.31E-82 | 0.327524911 | 0.066 | 0.028 | 1.35E-77 |
| LRR1 | 1.44E-81 | 0.91501511 | 0.094 | 0.04 | 3.07E-77 |
| MRPL58 | 3.24E-81 | 0.611706603 | 0.235 | 0.146 | 6.91E-77 |
| MRM2 | 4.84E-81 | 0.696817996 | 0.146 | 0.078 | 1.03E-76 |
| GPAT3 | 6.86E-81 | -0.793274484 | 0.002 | 0.027 | 1.47E-76 |
| RAB3B | 1.73E-80 | -3.555537591 | 0.006 | 0.04 | 3.70E-76 |
| HMGB1 | 3.04E-80 | 0.380163132 | 0.861 | 0.768 | 6.50E-76 |
| PEPD | 4.29E-80 | 0.450117367 | 0.259 | 0.171 | 9.15E-76 |
| NUDT16 | 4.36E-80 | 0.594978901 | 0.216 | 0.135 | 9.31E-76 |

| | | | | | |
|---|---|---|---|---|---|
| SEC22A | 7.90E-80 | 0.566206674 | 0.107 | 0.051 | 1.69E-75 |
| KLHDC2 | 7.91E-80 | -0.636159667 | 0.263 | 0.274 | 1.69E-75 |
| SUCLA2 | 8.32E-80 | 0.551803433 | 0.129 | 0.067 | 1.78E-75 |
| ATF7 | 9.38E-80 | 0.374364532 | 0.159 | 0.094 | 2.00E-75 |
| TCN2 | 1.12E-79 | -0.647000955 | 0.014 | 0.055 | 2.39E-75 |
| SSPN | 2.81E-79 | -1.720923662 | 0.038 | 0.095 | 6.00E-75 |
| CLP1 | 3.39E-79 | 1.027395693 | 0.078 | 0.03 | 7.25E-75 |
| DCLK1 | 3.44E-79 | 0.662640882 | 0.037 | 0.011 | 7.34E-75 |
| TMEM214 | 5.66E-79 | -0.508880187 | 0.28 | 0.272 | 1.21E-74 |
| ZBED5-AS1 | 6.84E-79 | 0.626575209 | 0.152 | 0.082 | 1.46E-74 |
| LAMP2 | 8.19E-79 | -0.375079636 | 0.301 | 0.313 | 1.75E-74 |
| ATP9B | 8.56E-79 | -0.334277669 | 0.153 | 0.115 | 1.83E-74 |
| NOL4L | 1.16E-78 | 0.648691111 | 0.075 | 0.03 | 2.48E-74 |
| TYW1 | 2.58E-78 | 0.380429192 | 0.169 | 0.101 | 5.50E-74 |
| CYP2J2 | 2.62E-78 | 0.272542338 | 0.054 | 0.023 | 5.61E-74 |
| PNPLA8 | 2.13E-77 | -0.285535866 | 0.35 | 0.322 | 4.54E-73 |
| BAG4 | 2.67E-77 | 0.58684068 | 0.154 | 0.086 | 5.70E-73 |
| LRIF1 | 3.15E-77 | -0.839554951 | 0.11 | 0.189 | 6.72E-73 |
| NFE2L2 | 1.63E-76 | -0.603293418 | 0.272 | 0.312 | 3.49E-72 |
| NR4A2 | 7.30E-76 | -0.494141535 | 0.351 | 0.384 | 1.56E-71 |
| ZMYM2 | 7.94E-76 | -0.411732024 | 0.188 | 0.168 | 1.70E-71 |
| CTNNA2 | 1.77E-75 | 1.601255931 | 0.028 | 0.005 | 3.77E-71 |
| AP000894.4 | 2.18E-75 | 0.560617213 | 0.041 | 0.014 | 4.67E-71 |
| UVSSA | 8.57E-75 | -1.20206835 | 0.099 | 0.149 | 1.83E-70 |
| TTI2 | 8.80E-75 | 0.719159091 | 0.155 | 0.084 | 1.88E-70 |
| TRIM33 | 1.35E-74 | -0.844536593 | 0.185 | 0.222 | 2.89E-70 |
| BCL9 | 3.85E-74 | -0.778886798 | 0.086 | 0.079 | 8.22E-70 |
| CXCL8 | 4.19E-74 | 1.74532697 | 0.043 | 0.04 | 8.94E-70 |
| P4HA2 | 4.35E-74 | 0.778969351 | 0.142 | 0.074 | 9.29E-70 |
| ETHE1 | 6.43E-74 | 0.419572451 | 0.244 | 0.16 | 1.37E-69 |
| POLL | 1.08E-73 | -1.506319251 | 0.055 | 0.11 | 2.30E-69 |
| ATG5 | 1.19E-73 | -0.28670941 | 0.29 | 0.253 | 2.55E-69 |
| AAR2 | 1.64E-73 | 0.76176525 | 0.096 | 0.044 | 3.50E-69 |
| RANBP6 | 1.69E-73 | -0.656105866 | 0.16 | 0.147 | 3.60E-69 |
| LEPROT | 2.65E-73 | -0.325300252 | 0.325 | 0.333 | 5.66E-69 |
| RNF5 | 4.79E-73 | 0.27609703 | 0.232 | 0.161 | 1.02E-68 |
| KBTBD4 | 1.25E-72 | 0.449059821 | 0.085 | 0.041 | 2.67E-68 |
| FBXW4 | 1.29E-72 | -1.363781378 | 0.088 | 0.129 | 2.77E-68 |
| ATP6V1E1 | 2.62E-72 | 0.310224989 | 0.298 | 0.216 | 5.60E-68 |
| ZNF667-AS1 | 4.37E-72 | -6.122374494 | 0.001 | 0.025 | 9.33E-68 |
| MAATS1 | 5.03E-72 | 0.799611795 | 0.025 | 0.005 | 1.08E-67 |
| CIDEB | 9.80E-72 | 0.730952784 | 0.068 | 0.028 | 2.09E-67 |
| TMC4 | 9.85E-72 | 0.281471126 | 0.047 | 0.02 | 2.10E-67 |
| THAP1 | 1.40E-71 | 0.403735517 | 0.21 | 0.133 | 2.98E-67 |
| CLUAP1 | 1.73E-71 | 0.296266303 | 0.099 | 0.051 | 3.70E-67 |
| MAN1C1 | 2.72E-71 | -2.253947043 | 0.013 | 0.053 | 5.81E-67 |
| SYNE3 | 4.66E-71 | -1.235126334 | 0.089 | 0.154 | 9.95E-67 |
| ZNF93 | 1.84E-70 | 1.070284684 | 0.042 | 0.012 | 3.92E-66 |
| EMSY | 2.73E-70 | 0.48464859 | 0.141 | 0.08 | 5.84E-66 |
| PFN2 | 4.83E-70 | -1.518772448 | 0.021 | 0.067 | 1.03E-65 |

| | | | | | |
|---|---|---|---|---|---|
| DIP2C | 6.23E-70 | -5.500363904 | 0.001 | 0.024 | 1.33E-65 |
| CDC25C | 1.07E-69 | 2.273983231 | 0.028 | 0.004 | 2.29E-65 |
| AC091563.1 | 1.66E-69 | -2.73319098 | 0.008 | 0.043 | 3.54E-65 |
| CD180 | 3.00E-69 | -0.810733678 | 0.076 | 0.083 | 6.41E-65 |
| NID2 | 3.06E-69 | 1.922705942 | 0.055 | 0.019 | 6.54E-65 |
| RELL2 | 4.83E-69 | 0.519816618 | 0.066 | 0.03 | 1.03E-64 |
| TCEAL4 | 1.27E-68 | -0.646129235 | 0.208 | 0.234 | 2.71E-64 |
| SLC25A45 | 1.49E-68 | -0.316265944 | 0.212 | 0.176 | 3.19E-64 |
| HIST1H2AG | 4.03E-68 | -0.43770934 | 0.107 | 0.078 | 8.61E-64 |
| TDP2 | 6.07E-68 | 0.332267687 | 0.156 | 0.094 | 1.30E-63 |
| GOLM1 | 9.14E-68 | 0.278935764 | 0.124 | 0.07 | 1.95E-63 |
| SDHAF2 | 1.77E-67 | 0.500581406 | 0.207 | 0.132 | 3.78E-63 |
| ZSCAN16 | 1.82E-67 | 0.498153563 | 0.089 | 0.043 | 3.89E-63 |
| CCP110 | 2.16E-67 | 0.51507479 | 0.124 | 0.068 | 4.61E-63 |
| C8orf76 | 1.10E-66 | 0.53373941 | 0.154 | 0.088 | 2.35E-62 |
| TNRC6C | 5.29E-66 | -0.680421522 | 0.086 | 0.075 | 1.13E-61 |
| CASP6 | 8.30E-66 | 0.495766672 | 0.134 | 0.075 | 1.77E-61 |
| DNAAF2 | 1.96E-65 | -0.749656753 | 0.169 | 0.18 | 4.18E-61 |
| DUSP8 | 2.82E-65 | -0.581913879 | 0.066 | 0.046 | 6.02E-61 |
| TTC26 | 4.23E-65 | 1.158949622 | 0.039 | 0.011 | 9.03E-61 |
| CLCN4 | 7.50E-65 | 1.419365805 | 0.062 | 0.022 | 1.60E-60 |
| NCOA7 | 1.51E-64 | -0.489051389 | 0.232 | 0.246 | 3.22E-60 |
| NSL1 | 2.06E-64 | 0.282201765 | 0.231 | 0.16 | 4.40E-60 |
| AL080276.2 | 2.14E-64 | 0.894204859 | 0.058 | 0.022 | 4.57E-60 |
| KIN | 4.17E-64 | 0.298049719 | 0.172 | 0.112 | 8.90E-60 |
| AL450998.2 | 9.26E-64 | -0.504647403 | 0.109 | 0.091 | 1.98E-59 |
| AL591846.2 | 1.47E-63 | -1.021683858 | 0.066 | 0.062 | 3.14E-59 |
| GATAD2B | 2.48E-63 | -0.42611449 | 0.216 | 0.204 | 5.30E-59 |
| AC025423.4 | 2.72E-63 | 2.671816912 | 0.024 | 0.003 | 5.80E-59 |
| P2RY8 | 2.94E-63 | -0.439113239 | 0.164 | 0.142 | 6.27E-59 |
| ZNF599 | 3.10E-63 | 0.612720286 | 0.055 | 0.022 | 6.62E-59 |
| GBP2 | 3.12E-63 | -1.045008058 | 0.014 | 0.051 | 6.67E-59 |
| AL121603.2 | 3.65E-63 | -1.178308718 | 0.088 | 0.125 | 7.80E-59 |
| TLR7 | 4.03E-63 | 2.744640318 | 0.037 | 0.014 | 8.62E-59 |
| CAMK2D | 4.98E-63 | -0.863772569 | 0.126 | 0.145 | 1.06E-58 |
| AC099778.1 | 1.61E-62 | -0.538736654 | 0.154 | 0.137 | 3.44E-58 |
| MEPCE | 2.94E-62 | -0.625079018 | 0.171 | 0.17 | 6.29E-58 |
| RNF34 | 3.52E-62 | 0.454316547 | 0.181 | 0.114 | 7.51E-58 |
| CISH | 3.89E-62 | 0.478543679 | 0.098 | 0.054 | 8.32E-58 |
| CORO7 | 1.58E-61 | -0.57024629 | 0.207 | 0.208 | 3.38E-57 |
| ADGRA3 | 1.75E-61 | 0.898072326 | 0.093 | 0.044 | 3.73E-57 |
| SYDE2 | 1.76E-61 | 0.307714174 | 0.031 | 0.011 | 3.75E-57 |
| TLE1 | 4.14E-61 | -1.114972686 | 0.085 | 0.148 | 8.85E-57 |
| GPHN | 5.80E-61 | -0.493821927 | 0.09 | 0.067 | 1.24E-56 |
| RAB26 | 5.91E-61 | 0.832607653 | 0.033 | 0.01 | 1.26E-56 |
| FAM118B | 6.49E-61 | 0.874332698 | 0.083 | 0.038 | 1.39E-56 |
| PRDM8 | 1.22E-60 | -2.057412549 | 0.003 | 0.029 | 2.61E-56 |
| CMIP | 1.41E-60 | 0.737700435 | 0.128 | 0.073 | 3.01E-56 |
| NUAK2 | 2.98E-60 | -1.216940441 | 0.059 | 0.094 | 6.37E-56 |
| TRIP11 | 7.02E-60 | -0.634451986 | 0.217 | 0.276 | 1.50E-55 |

| | | | | | |
|---|---|---|---|---|---|
| ULK1 | 9.86E-60 | -0.314847034 | 0.122 | 0.092 | 2.11E-55 |
| USP13 | 1.12E-59 | 0.63478085 | 0.073 | 0.033 | 2.40E-55 |
| PPP1R3D | 1.36E-59 | -1.431722044 | 0.022 | 0.063 | 2.91E-55 |
| RMI1 | 2.36E-59 | 0.895460982 | 0.08 | 0.037 | 5.03E-55 |
| CTBP1 | 2.65E-59 | -0.557642239 | 0.218 | 0.251 | 5.66E-55 |
| BABAM2 | 2.98E-59 | 0.278578239 | 0.15 | 0.097 | 6.37E-55 |
| FAM43A | 3.65E-59 | -1.85960241 | 0.006 | 0.035 | 7.79E-55 |
| PAXX | 6.38E-59 | -0.411903946 | 0.267 | 0.289 | 1.36E-54 |
| SLC16A6 | 8.82E-59 | 0.933292364 | 0.057 | 0.021 | 1.88E-54 |
| AAMDC | 1.82E-58 | 0.324140758 | 0.192 | 0.129 | 3.88E-54 |
| NMNAT1 | 2.93E-58 | 0.74933297 | 0.066 | 0.029 | 6.25E-54 |
| MGAT4A | 3.23E-58 | -1.036274114 | 0.024 | 0.067 | 6.89E-54 |
| CYSLTR1 | 4.90E-58 | -0.731639782 | 0.121 | 0.184 | 1.05E-53 |
| TLDC2 | 6.00E-58 | -3.700310208 | 0.001 | 0.022 | 1.28E-53 |
| ZSCAN18 | 7.16E-58 | -0.594119654 | 0.056 | 0.039 | 1.53E-53 |
| ZNF720 | 7.77E-58 | 0.335545546 | 0.127 | 0.078 | 1.66E-53 |
| AC004865.2 | 1.42E-57 | 0.527978225 | 0.058 | 0.025 | 3.04E-53 |
| NAGK | 1.93E-57 | 0.598672697 | 0.228 | 0.153 | 4.13E-53 |
| HEXIM1 | 2.40E-57 | -0.866525515 | 0.161 | 0.187 | 5.13E-53 |
| CDK11A | 2.59E-57 | -0.941549279 | 0.123 | 0.163 | 5.54E-53 |
| MIDN | 7.16E-57 | -0.498446679 | 0.209 | 0.262 | 1.53E-52 |
| HSPH1 | 7.93E-57 | -0.251252973 | 0.291 | 0.264 | 1.69E-52 |
| HAR1B | 9.87E-57 | -5.495773601 | 0 | 0.019 | 2.11E-52 |
| FAM193A | 1.06E-56 | -0.317780907 | 0.134 | 0.108 | 2.27E-52 |
| ATG101 | 2.06E-56 | -0.542409148 | 0.229 | 0.245 | 4.40E-52 |
| RBM5-AS1 | 5.66E-56 | 1.224699084 | 0.032 | 0.008 | 1.21E-51 |
| ENTPD6 | 5.88E-56 | -1.213946453 | 0.043 | 0.091 | 1.26E-51 |
| PLEKHG7 | 8.88E-56 | 1.675356946 | 0.024 | 0.005 | 1.90E-51 |
| SMARCAL1 | 1.00E-55 | 0.331472456 | 0.077 | 0.041 | 2.14E-51 |
| XYLT1 | 1.64E-55 | -0.829050799 | 0.022 | 0.059 | 3.51E-51 |
| WDR35 | 1.75E-55 | 0.408600498 | 0.045 | 0.019 | 3.73E-51 |
| MYLK-AS1 | 2.22E-55 | 0.859969242 | 0.037 | 0.013 | 4.74E-51 |
| CRB3 | 6.47E-55 | 0.464320207 | 0.041 | 0.017 | 1.38E-50 |
| EMC1 | 7.21E-55 | 0.301410374 | 0.153 | 0.1 | 1.54E-50 |
| NSMF | 7.69E-55 | 0.479244766 | 0.05 | 0.023 | 1.64E-50 |
| STAP1 | 1.19E-54 | -1.086607419 | 0.075 | 0.078 | 2.54E-50 |
| CIC | 1.28E-54 | -0.372648031 | 0.155 | 0.132 | 2.73E-50 |
| SLC25A15 | 1.47E-54 | 1.7872003 | 0.024 | 0.005 | 3.14E-50 |
| SOS1 | 1.58E-54 | -0.646753222 | 0.184 | 0.197 | 3.38E-50 |
| DCTD | 1.87E-54 | -0.736580394 | 0.169 | 0.213 | 4.00E-50 |
| TCEAL9 | 4.64E-54 | -1.187320406 | 0.017 | 0.053 | 9.92E-50 |
| AL035563.1 | 4.79E-54 | -1.679906532 | 0.024 | 0.062 | 1.02E-49 |
| STAM-AS1 | 1.66E-53 | -0.752358647 | 0.043 | 0.03 | 3.55E-49 |
| LINC01374 | 1.86E-53 | 1.201142076 | 0.026 | 0.006 | 3.97E-49 |
| BEX4 | 1.86E-53 | -0.903978387 | 0.062 | 0.113 | 3.97E-49 |
| ZDHHC4 | 1.99E-53 | -0.262983995 | 0.196 | 0.164 | 4.25E-49 |
| LEMD3 | 2.21E-53 | -0.319875526 | 0.186 | 0.157 | 4.72E-49 |
| SLC37A1 | 6.19E-53 | -0.420252885 | 0.076 | 0.057 | 1.32E-48 |
| PKP2 | 6.87E-53 | 0.370424157 | 0.163 | 0.104 | 1.47E-48 |
| CRIP1 | 9.15E-53 | 0.912976198 | 0.05 | 0.019 | 1.95E-48 |

| | | | | | |
|---|---|---|---|---|---|
| MED27 | 3.28E-52 | 0.26699022 | 0.157 | 0.101 | 7.02E-48 |
| POU5F2 | 5.20E-52 | -0.702980361 | 0.048 | 0.035 | 1.11E-47 |
| LINC00685 | 5.54E-52 | -1.968020396 | 0.024 | 0.056 | 1.18E-47 |
| PARD6G-AS1 | 7.91E-52 | -0.750290237 | 0.035 | 0.021 | 1.69E-47 |
| NAV2 | 2.26E-51 | -2.26215705 | 0.01 | 0.04 | 4.82E-47 |
| GRIA3 | 2.80E-51 | -7.2379963 | 0 | 0.015 | 5.97E-47 |
| ZFAND2B | 6.07E-51 | -0.703619351 | 0.158 | 0.184 | 1.30E-46 |
| RGS2 | 8.16E-51 | -0.601916592 | 0.251 | 0.288 | 1.74E-46 |
| IFT88 | 1.29E-50 | 0.581878263 | 0.064 | 0.031 | 2.76E-46 |
| AC083798.2 | 1.55E-50 | 0.403451789 | 0.07 | 0.037 | 3.30E-46 |
| KIF19 | 1.63E-50 | -0.682256997 | 0.051 | 0.042 | 3.48E-46 |
| TRIT1 | 2.25E-50 | 0.594056924 | 0.101 | 0.056 | 4.81E-46 |
| ARG2 | 3.05E-50 | 0.756989677 | 0.039 | 0.014 | 6.52E-46 |
| AC104653.1 | 6.86E-50 | 1.033210306 | 0.024 | 0.006 | 1.47E-45 |
| MYO5B | 9.42E-50 | 0.533097751 | 0.119 | 0.068 | 2.01E-45 |
| RNF144B | 1.16E-49 | 0.681272897 | 0.008 | 0.023 | 2.49E-45 |
| KHDRBS2 | 1.46E-49 | -1.772539397 | 0.028 | 0.04 | 3.12E-45 |
| CCDC80 | 1.49E-49 | 1.926455174 | 0.022 | 0.004 | 3.18E-45 |
| PRR3 | 2.17E-49 | -0.700021454 | 0.097 | 0.086 | 4.64E-45 |
| ZNF83 | 2.74E-49 | -1.342798551 | 0.027 | 0.066 | 5.85E-45 |
| FAM102A | 3.63E-49 | 0.363838358 | 0.117 | 0.071 | 7.76E-45 |
| RIMS1 | 3.66E-49 | -7.010767045 | 0 | 0.014 | 7.83E-45 |
| C3orf70 | 4.95E-49 | -1.500112698 | 0.028 | 0.068 | 1.06E-44 |
| HOMER1 | 7.72E-49 | -0.766265678 | 0.059 | 0.05 | 1.65E-44 |
| IQSEC1 | 8.88E-49 | -0.676724714 | 0.121 | 0.132 | 1.90E-44 |
| AC026979.2 | 1.12E-48 | -0.960292549 | 0.064 | 0.071 | 2.40E-44 |
| VPS13D | 1.35E-48 | -0.595593681 | 0.093 | 0.086 | 2.88E-44 |
| ZNF667 | 4.30E-48 | -3.931634979 | 0.001 | 0.018 | 9.18E-44 |
| ZFYVE28 | 5.06E-48 | 0.786543151 | 0.054 | 0.024 | 1.08E-43 |
| THUMPD2 | 9.67E-48 | -0.582632172 | 0.101 | 0.094 | 2.07E-43 |
| BHLHB9 | 1.46E-47 | 0.870476548 | 0.025 | 0.008 | 3.12E-43 |
| ARHGAP6 | 1.58E-47 | -2.793765872 | 0.004 | 0.026 | 3.37E-43 |
| ELAC1 | 1.62E-47 | 0.272302714 | 0.135 | 0.086 | 3.46E-43 |
| ECHDC2 | 1.84E-47 | -1.086102043 | 0.075 | 0.085 | 3.92E-43 |
| TNC | 2.95E-47 | -2.093405729 | 0 | 0.014 | 6.31E-43 |
| AP001059.2 | 3.91E-47 | 2.53392521 | 0.014 | 0.001 | 8.35E-43 |
| RBM4B | 4.36E-47 | 0.323477866 | 0.095 | 0.055 | 9.32E-43 |
| EDIL3 | 4.48E-47 | 0.325709867 | 0.022 | 0.008 | 9.56E-43 |
| RAB11FIP2 | 4.87E-47 | -0.729306074 | 0.111 | 0.129 | 1.04E-42 |
| TM2D2 | 7.36E-47 | 0.327903134 | 0.187 | 0.129 | 1.57E-42 |
| ELOA-AS1 | 9.67E-47 | 0.622077531 | 0.094 | 0.051 | 2.07E-42 |
| AC013394.1 | 1.18E-46 | -0.304773993 | 0.118 | 0.091 | 2.52E-42 |
| OASL | 1.62E-46 | -0.684575703 | 0.023 | 0.058 | 3.46E-42 |
| AC087623.3 | 1.77E-46 | -1.115300576 | 0.075 | 0.092 | 3.78E-42 |
| CCDC115 | 2.45E-46 | 0.260044458 | 0.147 | 0.098 | 5.24E-42 |
| GCHFR | 4.34E-46 | -0.674180248 | 0.11 | 0.162 | 9.27E-42 |
| CCM2 | 6.38E-46 | -0.362531333 | 0.224 | 0.212 | 1.36E-41 |
| GSN | 1.41E-45 | 2.115799411 | 0.037 | 0.012 | 3.01E-41 |
| RHOC | 3.88E-45 | -0.283392759 | 0.2 | 0.183 | 8.28E-41 |
| CDK7 | 5.27E-45 | 0.440429609 | 0.099 | 0.056 | 1.13E-40 |

| | | | | | |
|---|---|---|---|---|---|
| TBXAS1 | 5.79E-45 | -0.443369748 | 0.087 | 0.143 | 1.24E-40 |
| CFAP54 | 1.24E-44 | -0.616451847 | 0.038 | 0.074 | 2.64E-40 |
| NAT14 | 3.36E-44 | 0.302322917 | 0.043 | 0.02 | 7.17E-40 |
| AC006449.6 | 3.68E-44 | -0.733162621 | 0.107 | 0.115 | 7.86E-40 |
| WDPCP | 7.72E-44 | -0.38518247 | 0.076 | 0.058 | 1.65E-39 |
| ARSD | 8.15E-44 | -0.558397258 | 0.007 | 0.026 | 1.74E-39 |
| MID1 | 8.91E-44 | 1.994171913 | 0.016 | 0.002 | 1.90E-39 |
| GATA3 | 1.66E-43 | 1.387247794 | 0.02 | 0.004 | 3.54E-39 |
| AL645728.1 | 2.42E-43 | -0.702100594 | 0.061 | 0.057 | 5.17E-39 |
| STX11 | 2.45E-43 | -0.448804856 | 0.087 | 0.14 | 5.23E-39 |
| PIK3CA | 4.73E-43 | -0.601640048 | 0.13 | 0.131 | 1.01E-38 |
| PTPDC1 | 6.11E-43 | 0.318941296 | 0.029 | 0.012 | 1.30E-38 |
| FILIP1L | 7.80E-43 | 1.153087575 | 0.015 | 0.002 | 1.67E-38 |
| NR4A3 | 9.03E-43 | 0.778526319 | 0.043 | 0.018 | 1.93E-38 |
| EGLN1 | 1.20E-42 | -0.275692336 | 0.193 | 0.167 | 2.55E-38 |
| PANX2 | 1.84E-42 | -6.52939948 | 0 | 0.012 | 3.92E-38 |
| FAAH2 | 2.06E-42 | -0.557593779 | 0.045 | 0.034 | 4.41E-38 |
| CALCOCO2 | 2.93E-42 | -0.323735713 | 0.25 | 0.259 | 6.25E-38 |
| MAP2K1 | 3.62E-42 | 0.304437323 | 0.218 | 0.16 | 7.74E-38 |
| ARF5 | 3.66E-42 | 0.523709081 | 0.167 | 0.113 | 7.82E-38 |
| ATRNL1 | 5.20E-42 | -1.227363141 | 0.026 | 0.023 | 1.11E-37 |
| AC093772.1 | 9.59E-42 | 1.552752299 | 0.031 | 0.009 | 2.05E-37 |
| AC099518.5 | 1.87E-41 | -1.460376048 | 0.001 | 0.013 | 4.00E-37 |
| ISPD | 2.25E-41 | -4.927291889 | 0 | 0.015 | 4.80E-37 |
| EME1 | 2.43E-41 | 1.424905039 | 0.022 | 0.005 | 5.19E-37 |
| CPAMD8 | 2.53E-41 | -1.83655675 | 0.004 | 0.025 | 5.39E-37 |
| AC116366.1 | 4.58E-41 | -0.758022644 | 0.141 | 0.19 | 9.79E-37 |
| SPECC1 | 5.54E-41 | 0.874966473 | 0.147 | 0.094 | 1.18E-36 |
| CSTB | 7.71E-41 | 0.521091381 | 0.436 | 0.364 | 1.65E-36 |
| OR2A1-AS1 | 1.18E-40 | 1.355622171 | 0.022 | 0.005 | 2.53E-36 |
| ACTR8 | 1.55E-40 | 0.313031036 | 0.102 | 0.062 | 3.30E-36 |
| TCEAL2 | 1.82E-40 | -7.519542217 | 0 | 0.012 | 3.89E-36 |
| PIGK | 2.49E-40 | -0.636806793 | 0.163 | 0.173 | 5.32E-36 |
| CCDC51 | 2.85E-40 | 0.675284982 | 0.091 | 0.051 | 6.08E-36 |
| MED15 | 5.28E-40 | 0.318360278 | 0.103 | 0.068 | 1.13E-35 |
| MAGI2 | 7.00E-40 | -2.625585389 | 0.011 | 0.033 | 1.50E-35 |
| HYLS1 | 7.40E-40 | 0.62062617 | 0.055 | 0.026 | 1.58E-35 |
| ZNF236 | 1.02E-39 | -0.314595609 | 0.073 | 0.055 | 2.18E-35 |
| PRKACB | 2.16E-39 | -0.6568928 | 0.136 | 0.169 | 4.61E-35 |
| AC073389.1 | 2.37E-39 | -0.907098358 | 0.049 | 0.048 | 5.06E-35 |
| PAN3 | 2.65E-39 | -0.320920488 | 0.139 | 0.131 | 5.67E-35 |
| INO80E | 2.98E-39 | -0.342401562 | 0.138 | 0.122 | 6.36E-35 |
| RGS5.1 | 3.86E-39 | 0.522256743 | 0.036 | 0.016 | 8.24E-35 |
| LINC00909 | 4.19E-39 | -0.5141059 | 0.128 | 0.132 | 8.96E-35 |
| PEX13 | 5.39E-39 | 0.402709387 | 0.125 | 0.079 | 1.15E-34 |
| MB21D2 | 6.33E-39 | 0.802207082 | 0.078 | 0.041 | 1.35E-34 |
| GSTA4 | 6.72E-39 | -1.984165861 | 0.008 | 0.031 | 1.44E-34 |
| MICALL1 | 1.11E-38 | 0.311750475 | 0.044 | 0.022 | 2.37E-34 |
| TUBB | 1.18E-38 | 0.590637256 | 0.501 | 0.435 | 2.51E-34 |
| F8A3 | 1.27E-38 | 0.851188809 | 0.07 | 0.035 | 2.72E-34 |

| | | | | | |
|---|---|---|---|---|---|
| ARHGEF37 | 2.99E-38 | -1.730521971 | 0.025 | 0.05 | 6.38E-34 |
| PHLDB2 | 3.94E-38 | -2.287730838 | 0.007 | 0.029 | 8.41E-34 |
| BCL2L10 | 5.26E-38 | 1.006337074 | 0.017 | 0.004 | 1.12E-33 |
| ZNF625 | 8.82E-38 | 0.966933587 | 0.028 | 0.009 | 1.88E-33 |
| CCR2 | 9.05E-38 | -0.574336839 | 0.046 | 0.087 | 1.93E-33 |
| TMBIM1 | 1.28E-37 | -0.603069472 | 0.116 | 0.161 | 2.73E-33 |
| TUBA1B | 1.44E-37 | 0.659132995 | 0.594 | 0.519 | 3.08E-33 |
| TMEM74B | 2.96E-37 | -4.557758864 | 0.002 | 0.016 | 6.32E-33 |
| ATP10D | 2.97E-37 | -0.524239065 | 0.039 | 0.077 | 6.35E-33 |
| ZNF880 | 3.04E-37 | -1.389084739 | 0.023 | 0.05 | 6.49E-33 |
| CYB5R3 | 3.16E-37 | -0.599974791 | 0.111 | 0.154 | 6.74E-33 |
| KLHL28 | 6.77E-37 | -0.635463933 | 0.064 | 0.109 | 1.45E-32 |
| C14orf119 | 9.15E-37 | 0.360209923 | 0.198 | 0.142 | 1.95E-32 |
| USP54 | 9.78E-37 | 0.312857009 | 0.027 | 0.011 | 2.09E-32 |
| NLGN4X | 1.42E-36 | -1.719207371 | 0.017 | 0.045 | 3.02E-32 |
| DSTYK | 2.28E-36 | -0.549674682 | 0.096 | 0.104 | 4.88E-32 |
| AMN | 2.89E-36 | -4.193408112 | 0.001 | 0.015 | 6.16E-32 |
| DRAM2 | 3.89E-36 | -0.557821707 | 0.16 | 0.179 | 8.30E-32 |
| GON7 | 4.36E-36 | 0.389144744 | 0.164 | 0.113 | 9.31E-32 |
| AC004816.1 | 4.67E-36 | 1.620605243 | 0.021 | 0.005 | 9.98E-32 |
| ZBTB16 | 5.63E-36 | -1.375071287 | 0.003 | 0.019 | 1.20E-31 |
| NOVA1 | 1.55E-35 | -7.494176142 | 0 | 0.01 | 3.31E-31 |
| ARFGEF2 | 1.58E-35 | -0.624141537 | 0.144 | 0.181 | 3.37E-31 |
| CCDC28B | 2.84E-35 | 0.80554635 | 0.038 | 0.016 | 6.06E-31 |
| CCDC171 | 3.97E-35 | 0.35863065 | 0.023 | 0.01 | 8.48E-31 |
| MYL5 | 4.88E-35 | -1.038843673 | 0.038 | 0.073 | 1.04E-30 |
| PDGFC | 6.10E-35 | -1.015587941 | 0.003 | 0.019 | 1.30E-30 |
| CCNE2 | 7.19E-35 | -0.321877566 | 0.052 | 0.036 | 1.54E-30 |
| MPPE1 | 9.01E-35 | -0.308069291 | 0.148 | 0.14 | 1.92E-30 |
| ABCD2 | 9.41E-35 | -0.339182077 | 0.086 | 0.07 | 2.01E-30 |
| ZNF274 | 9.98E-35 | -0.386195491 | 0.128 | 0.112 | 2.13E-30 |
| GREB1 | 1.17E-34 | -3.846689194 | 0.003 | 0.018 | 2.50E-30 |
| PRICKLE2 | 1.27E-34 | -1.013606099 | 0.051 | 0.055 | 2.71E-30 |
| FAT4 | 1.36E-34 | -3.157107191 | 0.003 | 0.019 | 2.90E-30 |
| TMUB2 | 1.37E-34 | -0.484597649 | 0.135 | 0.144 | 2.92E-30 |
| ZNF284 | 1.59E-34 | 0.795839038 | 0.014 | 0.003 | 3.39E-30 |
| AC245140.2 | 1.65E-34 | -0.702826835 | 0.044 | 0.034 | 3.53E-30 |
| ARHGAP33 | 1.79E-34 | 0.30846063 | 0.025 | 0.011 | 3.82E-30 |
| FAM213A | 2.25E-34 | -0.445802096 | 0.107 | 0.101 | 4.80E-30 |
| FAM174B | 2.48E-34 | -0.655304613 | 0.04 | 0.032 | 5.30E-30 |
| CDK17 | 4.03E-34 | -0.787810638 | 0.112 | 0.132 | 8.61E-30 |
| ATF1 | 7.96E-34 | 0.254153508 | 0.162 | 0.12 | 1.70E-29 |
| BAG3 | 8.11E-34 | -0.377626995 | 0.098 | 0.082 | 1.73E-29 |
| AL359232.1 | 9.85E-34 | 0.384158598 | 0.032 | 0.014 | 2.10E-29 |
| CTNS | 1.11E-33 | 0.701644248 | 0.067 | 0.036 | 2.37E-29 |
| CDKL5 | 1.23E-33 | 0.287491575 | 0.081 | 0.048 | 2.62E-29 |
| AC092053.2 | 1.51E-33 | -1.576353561 | 0.016 | 0.039 | 3.22E-29 |
| AC024257.3 | 1.84E-33 | -1.54770569 | 0.016 | 0.038 | 3.93E-29 |
| ZNF540 | 3.21E-33 | -3.204557804 | 0.002 | 0.015 | 6.85E-29 |
| SPON2 | 4.54E-33 | -0.584898576 | 0.019 | 0.045 | 9.69E-29 |

| | | | | | |
|---|---|---|---|---|---|
| MYADM | 4.80E-33 | -0.625434386 | 0.122 | 0.167 | 1.03E-28 |
| SIK1B | 8.79E-33 | -0.722492923 | 0.035 | 0.024 | 1.88E-28 |
| PHF12 | 9.05E-33 | -0.396470732 | 0.128 | 0.116 | 1.93E-28 |
| AC243964.2 | 1.02E-32 | 0.985113221 | 0.02 | 0.006 | 2.17E-28 |
| AL365259.1 | 1.06E-32 | 1.098070068 | 0.015 | 0.004 | 2.26E-28 |
| APEX2 | 1.84E-32 | 0.381735332 | 0.063 | 0.038 | 3.92E-28 |
| SPAG16 | 2.33E-32 | 0.253871435 | 0.101 | 0.067 | 4.97E-28 |
| COCH | 3.15E-32 | -0.437553294 | 0.057 | 0.046 | 6.73E-28 |
| BANP | 3.39E-32 | -0.471443994 | 0.08 | 0.072 | 7.25E-28 |
| SPINK5 | 4.07E-32 | 2.513360262 | 0.025 | 0.01 | 8.70E-28 |
| AC009318.1 | 5.13E-32 | 0.860214542 | 0.019 | 0.006 | 1.09E-27 |
| APOF | 5.21E-32 | -3.779736689 | 0.002 | 0.016 | 1.11E-27 |
| THRIL | 7.58E-32 | 1.571839624 | 0.016 | 0.004 | 1.62E-27 |
| WNT10A | 8.24E-32 | -0.930773276 | 0.058 | 0.082 | 1.76E-27 |
| RTN4IP1 | 1.26E-31 | 0.490049362 | 0.042 | 0.021 | 2.70E-27 |
| LINC01686 | 1.41E-31 | 0.912819478 | 0.018 | 0.005 | 3.02E-27 |
| AP001107.1 | 1.81E-31 | 0.80631627 | 0.016 | 0.004 | 3.86E-27 |
| ZNF460 | 1.95E-31 | -0.556054441 | 0.153 | 0.155 | 4.16E-27 |
| CENPI | 2.10E-31 | 1.477204705 | 0.015 | 0.003 | 4.48E-27 |
| HIST1H4A | 2.38E-31 | -2.069273502 | 0.011 | 0.029 | 5.08E-27 |
| CAPN10-DT | 2.50E-31 | -1.667611555 | 0.026 | 0.048 | 5.35E-27 |
| MFSD11 | 5.38E-31 | -0.610941095 | 0.141 | 0.153 | 1.15E-26 |
| SEC11C | 6.27E-31 | -0.282711963 | 0.875 | 0.845 | 1.34E-26 |
| AC084018.2 | 9.29E-31 | -0.409739281 | 0.046 | 0.035 | 1.98E-26 |
| AL158152.1 | 1.30E-30 | -0.517459623 | 0.054 | 0.045 | 2.78E-26 |
| AGO4 | 1.71E-30 | -0.875310215 | 0.035 | 0.063 | 3.65E-26 |
| RGPD5 | 1.81E-30 | -0.740435409 | 0.123 | 0.145 | 3.87E-26 |
| AC092803.2 | 2.10E-30 | -1.277564346 | 0.024 | 0.034 | 4.49E-26 |
| ZNF17 | 3.58E-30 | 0.564620071 | 0.04 | 0.019 | 7.64E-26 |
| AC022098.4 | 4.63E-30 | -1.075657178 | 0.027 | 0.027 | 9.88E-26 |
| LINC01967 | 6.99E-30 | 0.442445641 | 0.023 | 0.01 | 1.49E-25 |
| TUBGCP5 | 8.23E-30 | -0.839307551 | 0.079 | 0.101 | 1.76E-25 |
| PIBF1 | 8.52E-30 | -0.386187195 | 0.096 | 0.086 | 1.82E-25 |
| NME7 | 1.31E-29 | -0.342373301 | 0.083 | 0.07 | 2.79E-25 |
| AC009133.1 | 2.35E-29 | -0.491328922 | 0.059 | 0.05 | 5.02E-25 |
| HELZ2 | 3.02E-29 | -0.606855095 | 0.074 | 0.075 | 6.45E-25 |
| LINC02413 | 3.07E-29 | 1.123089825 | 0.015 | 0.004 | 6.57E-25 |
| NRG1 | 3.29E-29 | -4.948711808 | 0 | 0.011 | 7.03E-25 |
| PAPOLG | 5.89E-29 | 0.366352507 | 0.099 | 0.065 | 1.26E-24 |
| APOBEC3F | 7.33E-29 | 0.658999765 | 0.047 | 0.024 | 1.56E-24 |
| RDH5 | 7.39E-29 | 1.362576604 | 0.021 | 0.007 | 1.58E-24 |
| WNT5B | 1.25E-28 | -0.964123217 | 0.07 | 0.099 | 2.67E-24 |
| CGRRF1 | 1.31E-28 | -0.739024795 | 0.085 | 0.126 | 2.80E-24 |
| CD69 | 2.02E-28 | 0.934448434 | 0.205 | 0.167 | 4.32E-24 |
| AL031316.1 | 2.61E-28 | 1.873479008 | 0.019 | 0.005 | 5.57E-24 |
| AC243829.4 | 3.63E-28 | -0.897110655 | 0.012 | 0.032 | 7.76E-24 |
| NKILA | 3.82E-28 | -2.112758349 | 0.005 | 0.021 | 8.16E-24 |
| RIDA | 3.90E-28 | 0.421220042 | 0.136 | 0.095 | 8.34E-24 |
| AC040970.1 | 3.94E-28 | 0.391646368 | 0.114 | 0.076 | 8.42E-24 |
| TCEAL8 | 5.49E-28 | -0.319228032 | 0.189 | 0.191 | 1.17E-23 |

| | | | | | |
|---|---|---|---|---|---|
| SNX22 | 1.24E-27 | 0.288509044 | 0.039 | 0.02 | 2.64E-23 |
| CSTF2 | 1.68E-27 | 0.627544306 | 0.044 | 0.023 | 3.59E-23 |
| SPAST | 1.98E-27 | -0.61092024 | 0.094 | 0.108 | 4.22E-23 |
| ZNF180 | 3.43E-27 | 0.327097717 | 0.033 | 0.017 | 7.33E-23 |
| ST7L | 3.68E-27 | -0.805712236 | 0.07 | 0.095 | 7.85E-23 |
| MSANTD4 | 4.05E-27 | 0.355925674 | 0.087 | 0.056 | 8.65E-23 |
| HOXB7 | 9.32E-27 | 0.353872048 | 0.026 | 0.013 | 1.99E-22 |
| AC006480.2 | 1.06E-26 | -0.702544821 | 0.033 | 0.026 | 2.26E-22 |
| NBPF26 | 1.70E-26 | 0.290363598 | 0.016 | 0.005 | 3.63E-22 |
| NDST1 | 1.89E-26 | 0.5663282 | 0.029 | 0.014 | 4.03E-22 |
| C20orf96 | 2.40E-26 | -0.339429623 | 0.041 | 0.031 | 5.12E-22 |
| FCRLA | 2.58E-26 | -0.592745583 | 0.176 | 0.191 | 5.50E-22 |
| AGPAT2 | 3.53E-26 | -0.65579135 | 0.091 | 0.129 | 7.54E-22 |
| RBBP9 | 5.38E-26 | 0.711270947 | 0.04 | 0.02 | 1.15E-21 |
| GTF2E1 | 5.76E-26 | 0.448656683 | 0.079 | 0.05 | 1.23E-21 |
| PHLPP2 | 8.27E-26 | -0.750843395 | 0.046 | 0.048 | 1.77E-21 |
| TRIM13 | 8.30E-26 | -0.253188636 | 0.082 | 0.069 | 1.77E-21 |
| RBBP5 | 9.76E-26 | -0.349640218 | 0.134 | 0.127 | 2.08E-21 |
| AC009309.1 | 1.31E-25 | 0.494996642 | 0.019 | 0.007 | 2.79E-21 |
| LRIG3 | 1.53E-25 | -1.54947189 | 0.021 | 0.039 | 3.27E-21 |
| ASB1 | 3.49E-25 | -0.357085561 | 0.076 | 0.06 | 7.45E-21 |
| METTL27 | 3.91E-25 | -3.491127678 | 0.002 | 0.013 | 8.36E-21 |
| MAVS | 4.32E-25 | -0.510517538 | 0.122 | 0.135 | 9.22E-21 |
| MOK | 4.89E-25 | 0.368361998 | 0.022 | 0.01 | 1.04E-20 |
| PTPRN2 | 1.16E-24 | -2.43381538 | 0.005 | 0.018 | 2.48E-20 |
| AC037459.3 | 1.20E-24 | -0.46875344 | 0.023 | 0.015 | 2.57E-20 |
| ZNF426 | 1.24E-24 | -0.428394238 | 0.073 | 0.066 | 2.65E-20 |
| GLS2 | 1.29E-24 | 0.263975046 | 0.021 | 0.01 | 2.75E-20 |
| CXCR3 | 1.58E-24 | 2.032899187 | 0.011 | 0.002 | 3.37E-20 |
| BLOC1S3 | 2.23E-24 | 0.528663432 | 0.044 | 0.023 | 4.77E-20 |
| HIST1H2BE | 5.75E-24 | -1.386474239 | 0.022 | 0.037 | 1.23E-19 |
| ZNF672 | 6.86E-24 | -0.663230491 | 0.074 | 0.087 | 1.47E-19 |
| SPHK1 | 7.65E-24 | 0.652747348 | 0.016 | 0.006 | 1.63E-19 |
| TCF7L1 | 8.55E-24 | -1.850428093 | 0.002 | 0.012 | 1.83E-19 |
| PANX1 | 1.03E-23 | 0.5216883 | 0.048 | 0.026 | 2.20E-19 |
| RPH3AL | 1.39E-23 | 0.90069312 | 0.016 | 0.006 | 2.96E-19 |
| PODXL2 | 1.92E-23 | -1.723160636 | 0.013 | 0.019 | 4.11E-19 |
| LEMD2 | 1.99E-23 | -0.79819488 | 0.062 | 0.081 | 4.26E-19 |
| PPIL4 | 2.19E-23 | -0.296703982 | 0.168 | 0.158 | 4.68E-19 |
| AC008079.1 | 2.29E-23 | 1.159924197 | 0.012 | 0.003 | 4.88E-19 |
| CACNA1A | 2.34E-23 | -1.383624261 | 0.025 | 0.031 | 4.99E-19 |
| AC010761.1 | 2.60E-23 | 0.990105326 | 0.017 | 0.006 | 5.54E-19 |
| RHOG | 3.29E-23 | 0.646200003 | 0.177 | 0.135 | 7.03E-19 |
| ZFP14 | 4.13E-23 | -0.940333716 | 0.029 | 0.037 | 8.83E-19 |
| CALD1 | 7.66E-23 | -2.969461724 | 0.004 | 0.015 | 1.64E-18 |
| AC104794.2 | 1.17E-22 | -1.133592535 | 0.005 | 0.017 | 2.51E-18 |
| CEBPB-AS1 | 1.42E-22 | -2.316179001 | 0.003 | 0.015 | 3.03E-18 |
| AL137003.2 | 1.63E-22 | -1.286834851 | 0.028 | 0.036 | 3.48E-18 |
| SLC25A44 | 3.96E-22 | -0.30962257 | 0.069 | 0.058 | 8.46E-18 |
| GDPGP1 | 4.45E-22 | 0.538089426 | 0.045 | 0.026 | 9.49E-18 |

| | | | | | |
|---|---|---|---|---|---|
| ARSB | 4.55E-22 | 0.448355119 | 0.079 | 0.053 | 9.71E-18 |
| HIST2H2AC | 4.78E-22 | -0.387210993 | 0.185 | 0.197 | 1.02E-17 |
| ESAM | 5.05E-22 | 0.487915129 | 0.022 | 0.011 | 1.08E-17 |
| PCYOX1 | 5.26E-22 | -0.507741017 | 0.159 | 0.167 | 1.12E-17 |
| CEP126 | 6.69E-22 | -1.555110345 | 0.012 | 0.028 | 1.43E-17 |
| SHISAL2A | 1.27E-21 | -0.549445883 | 0.081 | 0.08 | 2.71E-17 |
| FOXB1 | 1.37E-21 | -2.461336894 | 0.003 | 0.015 | 2.93E-17 |
| AL603756.1 | 1.42E-21 | -2.900123227 | 0.004 | 0.016 | 3.03E-17 |
| NFATC4 | 1.76E-21 | -1.721396051 | 0.008 | 0.02 | 3.77E-17 |
| ZNF134 | 1.93E-21 | 0.402773464 | 0.058 | 0.035 | 4.11E-17 |
| KIF3C | 2.07E-21 | 0.279181038 | 0.02 | 0.01 | 4.42E-17 |
| AC012645.3 | 2.16E-21 | -0.507123288 | 0.139 | 0.175 | 4.62E-17 |
| SCUBE1 | 2.73E-21 | -3.280175013 | 0.002 | 0.012 | 5.83E-17 |
| TBC1D32 | 2.93E-21 | -1.544213089 | 0.018 | 0.024 | 6.25E-17 |
| ZNF227 | 2.95E-21 | 0.295773451 | 0.068 | 0.045 | 6.30E-17 |
| MID2 | 3.02E-21 | -2.717163737 | 0.002 | 0.012 | 6.45E-17 |
| HIST1H4E | 3.21E-21 | -0.828063111 | 0.043 | 0.052 | 6.85E-17 |
| EXD3 | 3.55E-21 | -1.259235788 | 0.023 | 0.042 | 7.58E-17 |
| AC068633.1 | 4.38E-21 | -0.618720147 | 0.023 | 0.018 | 9.34E-17 |
| FAM49A | 5.20E-21 | 0.40386806 | 0.177 | 0.137 | 1.11E-16 |
| ACBD4 | 5.37E-21 | -0.994156234 | 0.049 | 0.071 | 1.15E-16 |
| EPB41L4A | 7.09E-21 | -0.59099279 | 0.051 | 0.08 | 1.51E-16 |
| TRAF2 | 1.31E-20 | -0.593165036 | 0.054 | 0.054 | 2.80E-16 |
| DYNLT3 | 1.60E-20 | -0.48987141 | 0.125 | 0.152 | 3.41E-16 |
| C5orf30 | 2.49E-20 | -0.353322312 | 0.081 | 0.071 | 5.33E-16 |
| SP4 | 2.79E-20 | -0.466223812 | 0.075 | 0.077 | 5.95E-16 |
| USP20 | 3.01E-20 | -0.43894061 | 0.042 | 0.033 | 6.44E-16 |
| MAN1B1-DT | 3.13E-20 | -0.551313667 | 0.046 | 0.042 | 6.69E-16 |
| CLMP | 3.74E-20 | -2.843204815 | 0.004 | 0.014 | 7.99E-16 |
| PTPRJ | 4.13E-20 | 0.671773686 | 0.008 | 0.017 | 8.83E-16 |
| ATG10 | 7.72E-20 | -0.35551682 | 0.094 | 0.085 | 1.65E-15 |
| VPS8 | 1.18E-19 | -0.473954361 | 0.074 | 0.075 | 2.52E-15 |
| AL390208.1 | 1.71E-19 | -1.637154433 | 0.013 | 0.019 | 3.65E-15 |
| OSGEPL1 | 2.20E-19 | 0.402617489 | 0.052 | 0.034 | 4.70E-15 |
| EFEMP1 | 2.64E-19 | 1.112113586 | 0.012 | 0.003 | 5.64E-15 |
| PLEKHB1 | 3.16E-19 | -0.730260961 | 0.018 | 0.014 | 6.76E-15 |
| MBIP | 3.51E-19 | -0.67505647 | 0.084 | 0.103 | 7.50E-15 |
| SMN2 | 3.67E-19 | 1.196189544 | 0.023 | 0.01 | 7.83E-15 |
| HCG27 | 4.66E-19 | -1.026069115 | 0.024 | 0.028 | 9.96E-15 |
| MYL9 | 8.03E-19 | 0.396813017 | 0.019 | 0.01 | 1.72E-14 |
| OSBPL6 | 8.37E-19 | -2.26042849 | 0.003 | 0.012 | 1.79E-14 |
| VWA7 | 9.61E-19 | -2.513021251 | 0.005 | 0.015 | 2.05E-14 |
| ICK | 9.75E-19 | -0.43369418 | 0.094 | 0.094 | 2.08E-14 |
| C15orf53 | 1.12E-18 | -0.690039763 | 0.018 | 0.014 | 2.39E-14 |
| AC093525.6 | 1.13E-18 | -0.347339245 | 0.022 | 0.014 | 2.42E-14 |
| AL606760.3 | 1.50E-18 | -2.838754858 | 0.004 | 0.014 | 3.19E-14 |
| SPA17 | 3.21E-18 | -0.549540178 | 0.028 | 0.023 | 6.86E-14 |
| AL135925.1 | 4.31E-18 | -0.491776799 | 0.054 | 0.047 | 9.20E-14 |
| NECAB1 | 4.56E-18 | 1.369305329 | 0.011 | 0.003 | 9.75E-14 |
| AC005618.1 | 6.10E-18 | -0.380949428 | 0.018 | 0.01 | 1.30E-13 |

| | | | | | |
|---|---|---|---|---|---|
| GTPBP2 | 1.42E-17 | -0.621682497 | 0.059 | 0.081 | 3.03E-13 |
| LILRB3 | 1.80E-17 | 2.126609829 | 0.025 | 0.013 | 3.85E-13 |
| AC008083.2 | 2.18E-17 | 0.408973051 | 0.013 | 0.005 | 4.66E-13 |
| ZNF117 | 2.86E-17 | -0.343022158 | 0.071 | 0.067 | 6.10E-13 |
| AC074044.1 | 3.82E-17 | -0.566145786 | 0.023 | 0.018 | 8.16E-13 |
| FAR2 | 4.35E-17 | -0.851595646 | 0.042 | 0.054 | 9.29E-13 |
| JMJD1C-AS1 | 5.21E-17 | 0.797499372 | 0.011 | 0.004 | 1.11E-12 |
| AC105254.1 | 5.97E-17 | -2.689080008 | 0.004 | 0.011 | 1.27E-12 |
| CA5B | 6.49E-17 | -0.332480872 | 0.074 | 0.065 | 1.39E-12 |
| AC048341.2 | 6.80E-17 | -0.955112078 | 0.026 | 0.031 | 1.45E-12 |
| AC005838.2 | 7.96E-17 | -0.283725663 | 0.111 | 0.101 | 1.70E-12 |
| GIMAP8 | 8.51E-17 | 2.863699212 | 0.015 | 0.006 | 1.82E-12 |
| RASGEF1A | 8.58E-17 | -1.091532925 | 0.002 | 0.01 | 1.83E-12 |
| GLYCTK | 8.69E-17 | -0.788475515 | 0.039 | 0.041 | 1.86E-12 |
| AC005480.1 | 1.17E-16 | 0.902187945 | 0.014 | 0.005 | 2.50E-12 |
| ALG1L | 1.30E-16 | -1.026141539 | 0.017 | 0.019 | 2.78E-12 |
| AC112907.3 | 1.63E-16 | -1.109671288 | 0.019 | 0.023 | 3.49E-12 |
| AC092910.3 | 2.06E-16 | -1.323156095 | 0.015 | 0.021 | 4.41E-12 |
| TRIM58 | 2.62E-16 | -0.74258233 | 0.007 | 0.02 | 5.60E-12 |
| AP003352.1 | 2.85E-16 | 0.293211751 | 0.019 | 0.009 | 6.10E-12 |
| ARRDC4 | 3.06E-16 | 1.504957892 | 0.026 | 0.012 | 6.55E-12 |
| AC011446.2 | 3.15E-16 | -1.312209376 | 0.016 | 0.029 | 6.72E-12 |
| AC010883.1 | 3.66E-16 | 0.433765005 | 0.026 | 0.014 | 7.82E-12 |
| HIST1H2AB | 4.17E-16 | -1.028073155 | 0.015 | 0.013 | 8.90E-12 |
| GOLPH3L | 5.46E-16 | -0.523952 | 0.13 | 0.147 | 1.17E-11 |
| GALNT13 | 6.88E-16 | -2.131649457 | 0.006 | 0.015 | 1.47E-11 |
| AC133550.2 | 7.48E-16 | -0.646266129 | 0.019 | 0.017 | 1.60E-11 |
| CHODL | 9.19E-16 | 0.404838572 | 0.011 | 0.004 | 1.96E-11 |
| STRADB | 1.50E-15 | -0.509866809 | 0.047 | 0.07 | 3.21E-11 |
| CDC25B | 2.38E-15 | -0.508830357 | 0.078 | 0.088 | 5.08E-11 |
| SPDYA | 2.93E-15 | 0.977016772 | 0.016 | 0.007 | 6.27E-11 |
| ITPKB | 3.30E-15 | -0.275520918 | 0.059 | 0.049 | 7.05E-11 |
| SLC19A2 | 3.45E-15 | -0.781931884 | 0.035 | 0.039 | 7.36E-11 |
| AC092040.1 | 4.06E-15 | -1.060759297 | 0.024 | 0.029 | 8.67E-11 |
| AC009318.3 | 4.11E-15 | -0.49828727 | 0.019 | 0.014 | 8.79E-11 |
| RETREG1 | 8.82E-15 | -1.296738853 | 0.002 | 0.01 | 1.88E-10 |
| AP000542.2 | 1.07E-14 | 0.447085351 | 0.014 | 0.006 | 2.30E-10 |
| NR1H3 | 1.15E-14 | 1.234280607 | 0.021 | 0.009 | 2.45E-10 |
| NPC1 | 1.46E-14 | -0.822126162 | 0.033 | 0.047 | 3.11E-10 |
| COPS4 | 1.53E-14 | -0.250987605 | 0.133 | 0.129 | 3.27E-10 |
| RAD21-AS1 | 3.20E-14 | -2.080608647 | 0.007 | 0.013 | 6.84E-10 |
| MORF4L2-AS1 | 3.24E-14 | -1.293047341 | 0.018 | 0.025 | 6.92E-10 |
| SSTR3 | 4.00E-14 | -2.224211965 | 0.005 | 0.012 | 8.55E-10 |
| FOXO4 | 5.22E-14 | -0.674050126 | 0.022 | 0.023 | 1.11E-09 |
| RNASEL | 5.67E-14 | -0.593232389 | 0.013 | 0.027 | 1.21E-09 |
| PAG1 | 9.38E-14 | 1.111736401 | 0.027 | 0.023 | 2.00E-09 |
| METTL18 | 9.50E-14 | -0.642456422 | 0.053 | 0.072 | 2.03E-09 |
| AL354920.1 | 1.31E-13 | -1.20157483 | 0.017 | 0.031 | 2.81E-09 |
| KIF1BP | 2.27E-13 | -0.380310064 | 0.052 | 0.075 | 4.85E-09 |
| BLZF1 | 2.58E-13 | -0.388529314 | 0.114 | 0.135 | 5.52E-09 |

| | | | | | |
|---|---|---|---|---|---|
| AC009133.3 | 3.87E-13 | -1.589147104 | 0.013 | 0.02 | 8.27E-09 |
| KBTBD8 | 8.79E-13 | -0.711424894 | 0.051 | 0.054 | 1.88E-08 |
| EPSTI1 | 1.01E-12 | 0.994251705 | 0.114 | 0.099 | 2.16E-08 |
| ARNT2 | 1.66E-12 | -1.493535052 | 0.003 | 0.011 | 3.55E-08 |
| ANKDD1B | 2.84E-12 | 1.234093496 | 0.014 | 0.006 | 6.06E-08 |
| BX284668.6 | 3.45E-12 | -1.038754549 | 0.022 | 0.029 | 7.37E-08 |
| EXO5 | 3.82E-12 | 0.731354046 | 0.017 | 0.008 | 8.15E-08 |
| LRRC7 | 4.19E-12 | -1.177893851 | 0.013 | 0.02 | 8.95E-08 |
| C17orf67 | 4.28E-12 | -0.774069894 | 0.04 | 0.056 | 9.15E-08 |
| AC072061.1 | 4.52E-12 | -1.178069727 | 0.016 | 0.024 | 9.64E-08 |
| AL137077.2 | 5.77E-12 | -1.957932156 | 0.004 | 0.011 | 1.23E-07 |
| AC008378.1 | 5.83E-12 | -1.977782727 | 0.005 | 0.01 | 1.25E-07 |
| ANKRD33B | 7.47E-12 | -0.642996765 | 0.07 | 0.089 | 1.60E-07 |
| ILDR1 | 8.04E-12 | -1.159275033 | 0.017 | 0.03 | 1.72E-07 |
| AC136475.5 | 9.16E-12 | -0.580665094 | 0.009 | 0.019 | 1.96E-07 |
| CNTN1 | 1.00E-11 | -1.234882488 | 0.007 | 0.015 | 2.14E-07 |
| C2CD5 | 1.04E-11 | -0.309873211 | 0.079 | 0.086 | 2.23E-07 |
| KLRF1 | 1.23E-11 | 0.625650228 | 0.007 | 0.012 | 2.63E-07 |
| AL135905.1 | 1.39E-11 | -0.952075693 | 0.015 | 0.013 | 2.97E-07 |
| MAP2 | 1.61E-11 | 0.259649733 | 0.022 | 0.013 | 3.44E-07 |
| TNFSF9 | 1.95E-11 | -1.192449855 | 0.011 | 0.012 | 4.17E-07 |
| ANKRD20A4 | 2.10E-11 | -1.761380659 | 0.005 | 0.013 | 4.48E-07 |
| GPRASP1 | 2.11E-11 | -1.385440537 | 0.009 | 0.015 | 4.51E-07 |
| AC120193.1 | 2.43E-11 | 0.2542149 | 0.012 | 0.007 | 5.20E-07 |
| BCAS3 | 3.58E-11 | -0.461570523 | 0.053 | 0.053 | 7.64E-07 |
| PKD2L2 | 4.04E-11 | -1.344281004 | 0.011 | 0.014 | 8.62E-07 |
| FCRL1 | 4.33E-11 | -0.908285875 | 0.012 | 0.018 | 9.24E-07 |
| AC009093.2 | 4.52E-11 | -0.895532961 | 0.026 | 0.027 | 9.65E-07 |
| ZC3H12D | 5.26E-11 | -0.399828711 | 0.01 | 0.006 | 1.12E-06 |
| SYTL3 | 7.35E-11 | -1.308557033 | 0.016 | 0.028 | 1.57E-06 |
| INPP4B | 8.75E-11 | -1.290338869 | 0.004 | 0.011 | 1.87E-06 |
| WDR11-AS1 | 9.53E-11 | -1.815872512 | 0.004 | 0.011 | 2.04E-06 |
| ZC3H11A | 1.31E-10 | -0.653306684 | 0.023 | 0.021 | 2.80E-06 |
| ETV3 | 1.50E-10 | -0.305821616 | 0.115 | 0.117 | 3.21E-06 |
| PRICKLE1 | 1.96E-10 | -0.93943407 | 0.011 | 0.021 | 4.19E-06 |
| TCTN2 | 2.10E-10 | -0.452131779 | 0.014 | 0.01 | 4.48E-06 |
| LINC01465 | 3.34E-10 | -0.689464877 | 0.032 | 0.036 | 7.13E-06 |
| PTPRD | 3.64E-10 | -1.558083902 | 0.007 | 0.015 | 7.77E-06 |
| AC012368.2 | 3.98E-10 | -2.111237619 | 0.004 | 0.01 | 8.49E-06 |
| HIST2H2AB | 4.59E-10 | -1.478416159 | 0.01 | 0.016 | 9.81E-06 |
| AC079305.1 | 6.24E-10 | -1.096504341 | 0.014 | 0.021 | 1.33E-05 |
| AC108134.2 | 7.14E-10 | -1.877310395 | 0.004 | 0.011 | 1.53E-05 |
| AL021068.1 | 7.17E-10 | -1.480952938 | 0.006 | 0.013 | 1.53E-05 |
| AC073195.1 | 7.63E-10 | -0.450062691 | 0.014 | 0.013 | 1.63E-05 |
| DUSP19 | 8.44E-10 | -0.684265592 | 0.014 | 0.013 | 1.80E-05 |
| HEXA-AS1 | 9.18E-10 | -1.805989725 | 0.007 | 0.012 | 1.96E-05 |
| GAS7 | 2.22E-09 | 1.978198128 | 0.011 | 0.007 | 4.74E-05 |
| TXLNB | 2.32E-09 | -0.36952895 | 0.025 | 0.023 | 4.94E-05 |
| TTC9 | 2.77E-09 | 0.721645272 | 0.034 | 0.023 | 5.91E-05 |
| ARHGEF3 | 3.07E-09 | -0.336961001 | 0.072 | 0.076 | 6.56E-05 |

| | | | | | |
|---|---|---|---|---|---|
| AC092069.1 | 3.23E-09 | -0.331626912 | 0.103 | 0.099 | 6.90E-05 |
| ZFYVE1 | 4.67E-09 | 0.878865557 | 0.03 | 0.019 | 9.98E-05 |
| PLLP | 5.84E-09 | 0.373837569 | 0.011 | 0.005 | 0.000124643 |
| AC078883.3 | 7.06E-09 | -0.555686825 | 0.05 | 0.056 | 0.000150863 |
| TMEM53 | 7.34E-09 | 0.324398435 | 0.024 | 0.016 | 0.00015678 |
| AHDC1 | 9.29E-09 | -0.344701685 | 0.017 | 0.015 | 0.000198327 |
| TPP1 | 1.24E-08 | 0.29754227 | 0.158 | 0.145 | 0.000263952 |
| ADPRM | 1.59E-08 | 0.255393364 | 0.032 | 0.023 | 0.000339229 |
| MYOF | 2.19E-08 | 1.207492075 | 0.009 | 0.01 | 0.000467756 |
| POLD4 | 2.45E-08 | -0.365416206 | 0.071 | 0.072 | 0.000524118 |
| ECHDC3 | 2.98E-08 | 0.592388488 | 0.029 | 0.018 | 0.000635767 |
| TMEM250 | 5.10E-08 | -0.319603117 | 0.071 | 0.08 | 0.001089133 |
| PLPPR2 | 5.77E-08 | 0.826785702 | 0.022 | 0.014 | 0.00123216 |
| RNF43 | 5.80E-08 | -1.848085545 | 0.006 | 0.01 | 0.001238983 |
| STAT4 | 6.76E-08 | -0.463342895 | 0.113 | 0.125 | 0.001443189 |
| LINC01801 | 1.14E-07 | 1.337291083 | 0.014 | 0.007 | 0.002427537 |
| ARHGAP31 | 1.23E-07 | -0.338148638 | 0.035 | 0.044 | 0.00263632 |
| CFH | 1.35E-07 | -1.628983835 | 0.004 | 0.01 | 0.002884652 |
| SPRY1 | 1.70E-07 | -0.860754433 | 0.024 | 0.031 | 0.00363448 |
| PRCD | 1.74E-07 | -1.350078161 | 0.005 | 0.011 | 0.003722707 |
| AC016708.1 | 1.94E-07 | 0.292529157 | 0.01 | 0.006 | 0.00413625 |
| VAMP3 | 2.23E-07 | 0.323248493 | 0.155 | 0.135 | 0.004753593 |
| ZNF571 | 2.44E-07 | -0.948475408 | 0.009 | 0.016 | 0.005203746 |
| DDIT4-AS1 | 3.77E-07 | -1.456821792 | 0.007 | 0.013 | 0.008049164 |
| AL590764.1 | 3.97E-07 | 0.263083034 | 0.024 | 0.016 | 0.008475883 |
| LINC00511 | 4.84E-07 | 0.31711566 | 0.01 | 0.006 | 0.01033449 |
| FPGT | 5.34E-07 | -0.730186648 | 0.015 | 0.021 | 0.011412105 |
| FGF13 | 8.43E-07 | -0.703258677 | 0.015 | 0.022 | 0.017993475 |
| MOSPD2 | 8.57E-07 | 0.282183783 | 0.061 | 0.049 | 0.018300689 |
| NOTCH1 | 9.94E-07 | -0.268401016 | 0.032 | 0.043 | 0.021219851 |
| AL033527.5 | 1.35E-06 | -0.340762743 | 0.039 | 0.037 | 0.028865485 |
| B3GNT5 | 1.36E-06 | 0.976337068 | 0.012 | 0.006 | 0.029010055 |
| TRMT10A | 1.54E-06 | -0.438978327 | 0.044 | 0.054 | 0.032911923 |
| LINC01358 | 1.95E-06 | -1.012967031 | 0.009 | 0.016 | 0.041619793 |
| AC092718.1 | 2.04E-06 | -1.262300739 | 0.006 | 0.011 | 0.04364017 |
| ADAMTSL4 | 2.18E-06 | 0.654453909 | 0.023 | 0.016 | 0.046526383 |

**Supplementary Table S2**

| | baseMean | log2FoldChange | lfcSE | pvalue | padj |
|---|---|---|---|---|---|
| SSX2 | 607.6591199 | -13.99023503 | 3.221299021 | 1.25E-26 | 1.16E-24 |
| SSX2B | 607.5012824 | -13.98608237 | 3.216838876 | 1.26E-26 | 1.16E-24 |
| LOC124900602 | 78.17232375 | -10.65095038 | 2.896069032 | 8.48E-16 | 3.77E-14 |
| MAGEC1 | 72.3464631 | -10.52775074 | 2.890605973 | 1.53E-15 | 6.59E-14 |
| C9orf64 | 50.57205003 | -9.925657648 | 2.832116071 | 7.34E-14 | 2.65E-12 |
| BOK-AS1 | 43.63127328 | -9.674814242 | 2.806864959 | 3.19E-13 | 1.08E-11 |
| ZNF649 | 32.74059288 | -9.179566034 | 2.75988136 | 6.22E-12 | 1.79E-10 |
| NFILZ | 29.1743931 | 9.168690742 | 2.834312436 | 8.97E-12 | 2.54E-10 |
| MEIOC | 24.54395457 | -8.682369362 | 2.711551187 | 9.15E-11 | 2.20E-09 |
| GABRA4 | 17.87267797 | 8.300754376 | 2.746517139 | 1.02E-09 | 2.13E-08 |
| SSX7 | 16.85776637 | -8.012102686 | 2.652060924 | 3.03E-09 | 5.91E-08 |
| LOC105373974 | 103.9609559 | -7.986298434 | 0.971724042 | 2.05E-15 | 8.68E-14 |
| LIN28B | 134.3910221 | -7.836140936 | 0.821069078 | 7.19E-21 | 4.73E-19 |
| ARHGAP25 | 566.1053463 | -7.834781969 | 0.395246504 | 9.17E-87 | 6.35E-84 |
| PYDC2-AS1 | 118.1003418 | -7.658666318 | 0.813029854 | 3.68E-20 | 2.35E-18 |
| LINC01224 | 12.76072694 | -7.488133763 | 2.617147458 | 4.57E-08 | 7.36E-07 |
| KLRK1 | 10.54278571 | 7.295887433 | 2.670110575 | 1.61E-07 | 2.36E-06 |
| ANGPT1 | 60.54952367 | 7.281939681 | 0.991624802 | 8.37E-13 | 2.70E-11 |
| LOC613266 | 33.26022514 | 7.19044007 | 1.306986694 | 5.69E-10 | 1.24E-08 |
| LOC105371197 | 34.41934565 | -7.10736594 | 1.300565902 | 8.35E-10 | 1.77E-08 |
| TRPM6 | 31.72444154 | -6.999708718 | 1.296444211 | 1.43E-09 | 2.94E-08 |
| PARVG | 9.172881757 | -6.858489051 | 2.575612646 | 6.85E-07 | 8.93E-06 |
| STYK1 | 8.407820331 | 6.854868862 | 2.640966466 | 1.03E-06 | 1.29E-05 |
| TTC22 | 9.149853854 | -6.832877562 | 2.586991861 | 9.58E-07 | 1.21E-05 |
| CTSZ | 59.86535699 | -6.64505066 | 0.833665227 | 4.76E-15 | 1.95E-13 |
| TIGIT | 8.08871283 | -6.604009015 | 2.565060398 | 1.95E-06 | 2.30E-05 |
| LOC124905234 | 7.983722107 | -6.582488622 | 2.564010854 | 2.11E-06 | 2.48E-05 |
| SSX4 | 24.34229343 | -6.557724925 | 1.302713369 | 1.31E-08 | 2.28E-07 |
| SSX4B | 24.34229343 | -6.557724925 | 1.302713369 | 1.31E-08 | 2.28E-07 |
| ZNF577 | 7.114475873 | -6.342888465 | 2.554325167 | 5.09E-06 | 5.48E-05 |
| NUDT13 | 6.434213872 | 6.295449031 | 2.616139774 | 8.13E-06 | 8.36E-05 |
| GBP4 | 270.1721206 | 6.224469149 | 0.335263296 | 5.90E-77 | 3.00E-74 |
| RN7SL128P | 6.645976486 | 6.194107059 | 2.726696592 | 3.47E-05 | 0.000310725 |
| LOC105376418 | 6.399156423 | -6.102190256 | 2.557410284 | 1.31E-05 | 0.000129485 |
| PTCHD4 | 6.201125813 | -6.041265107 | 2.545269483 | 1.46E-05 | 0.000142586 |
| SOX5 | 71.43361708 | -5.954465024 | 0.617125918 | 4.27E-22 | 3.08E-20 |
| ATP10A | 13.41926784 | 5.729004419 | 1.343466044 | 8.32E-07 | 1.07E-05 |
| TLR7 | 1163.001096 | -5.677144388 | 0.156903116 | 1.09E-287 | 9.40E-284 |
| GBP2 | 157.3680793 | 5.563622191 | 0.359109345 | 2.09E-54 | 6.34E-52 |
| TMEM144 | 21.70722213 | -5.558768133 | 1.023689182 | 5.01E-08 | 8.01E-07 |
| GBP1 | 139.9652218 | 5.537263811 | 0.379426296 | 1.64E-48 | 4.23E-46 |
| SSX8P | 4.895625284 | -5.511213196 | 2.554818584 | 7.71E-05 | 0.000632724 |
| SSX6P | 4.877484123 | -5.505734251 | 2.553774264 | 7.77E-05 | 0.00063635 |
| ATOH8 | 4.649734149 | 5.48511437 | 2.655972575 | 0.000126776 | 0.000978573 |
| LOC105375451 | 4.669079349 | -5.368615222 | 2.583100164 | 0.000132065 | 0.001013725 |
| RGS9 | 78.18348611 | 5.251009442 | 0.464125806 | 5.64E-30 | 6.51E-28 |
| SPOCK2 | 191.3768802 | -5.103777048 | 0.296035159 | 8.70E-67 | 3.68E-64 |
| FCRL4 | 31.79219861 | 5.094621986 | 0.710904755 | 3.52E-13 | 1.18E-11 |
| KLRC4-KLRK1 | 15.02508889 | 5.087950713 | 1.075097004 | 1.12E-06 | 1.39E-05 |

| | | | | | |
|---|---|---|---|---|---|
| LOC105373480 | 4.192959574 | -5.040581759 | 2.641225627 | 0.00034124 | 0.002332904 |
| GNLY | 70.47253012 | 4.981481296 | 0.462752643 | 1.79E-27 | 1.80E-25 |
| ZNF350 | 14.21579001 | -4.877670478 | 1.062514028 | 2.19E-06 | 2.55E-05 |
| COL22A1 | 17.23030521 | 4.79996079 | 0.910051888 | 4.97E-08 | 7.97E-07 |
| CLIC2 | 223.9301862 | -4.76969591 | 0.266602428 | 1.53E-72 | 7.38E-70 |
| LOC124900856 | 3.621247249 | -4.719451399 | 2.624860294 | 0.0005431 | 0.003498835 |
| NLRP5 | 35.93408695 | -4.691680198 | 0.614838186 | 7.36E-15 | 2.94E-13 |
| BMP7 | 7.471211332 | 4.655784858 | 1.430715346 | 5.85E-05 | 0.000494581 |
| DPEP1 | 3.721078143 | -4.634848446 | 2.728905876 | 0.000825357 | 0.005043301 |
| MIR548XHG | 3.482854459 | -4.593887565 | 2.648557047 | 0.000703442 | 0.004389733 |
| CRIM1 | 33.08609925 | -4.580509757 | 0.621427003 | 5.91E-14 | 2.17E-12 |
| EPHX2 | 53.86339735 | -4.573539951 | 0.484005282 | 1.34E-21 | 9.40E-20 |
| SMIM3 | 3.550259275 | -4.573083332 | 2.693281339 | 0.000815219 | 0.004993683 |
| LOC105372349 | 36.75532589 | -4.519757687 | 0.618871545 | 5.29E-14 | 1.96E-12 |
| SSX3 | 3.33278392 | -4.453518182 | 2.677144167 | 0.000914537 | 0.005508535 |
| PHLDA3 | 576.280596 | -4.452670047 | 0.171064795 | 1.48E-150 | 2.56E-147 |
| ENPP2 | 45.95731689 | 4.401749381 | 0.516428622 | 2.19E-18 | 1.16E-16 |
| MACROD2 | 62.44963196 | 4.381931044 | 0.443249778 | 7.63E-24 | 6.21E-22 |
| CD3G | 20.90511018 | -4.292274519 | 0.751132915 | 2.44E-09 | 4.84E-08 |
| LGALS3BP | 16.0897291 | 4.288764423 | 0.839346148 | 6.54E-08 | 1.02E-06 |
| H2BC14 | 14.12490052 | 4.247511901 | 1.193269524 | 3.19E-05 | 0.000288728 |
| AICDA | 6.385108971 | -4.203192326 | 1.481688335 | 0.00025743 | 0.001821788 |
| P3H3 | 119.6859233 | -4.16041281 | 0.309399661 | 5.61E-42 | 1.08E-39 |
| SIT1 | 25.45949437 | -4.15789638 | 0.649066259 | 3.40E-11 | 8.76E-10 |
| KRT8 | 380.849156 | -4.148623852 | 0.185450696 | 1.81E-111 | 1.74E-108 |
| GDF15 | 190.2237265 | -4.146386601 | 0.246423902 | 1.78E-64 | 6.99E-62 |
| TNFRSF1A | 18.95780705 | -4.136798725 | 0.75904995 | 1.04E-08 | 1.86E-07 |
| LRRC43 | 6.235038432 | -4.134984488 | 1.506910796 | 0.000332431 | 0.002281694 |
| H2AC4 | 13.61104812 | 4.128758528 | 1.238666079 | 6.22E-05 | 0.000523061 |
| MIR23AHG | 21.97938011 | -4.121448079 | 0.700982236 | 8.09E-10 | 1.72E-08 |
| ZNF883 | 12.06292674 | -4.08139674 | 0.965755206 | 5.27E-06 | 5.64E-05 |
| LRRK2 | 8.582551949 | 4.058641468 | 1.172800909 | 9.67E-05 | 0.000772784 |
| CRMP1 | 27.87503874 | 4.042502769 | 0.589839343 | 1.09E-12 | 3.50E-11 |
| FST | 5.681459867 | 4.039192528 | 1.523991995 | 0.000372644 | 0.002520733 |
| SIGLEC17P | 5.510550827 | 4.031381526 | 1.516526861 | 0.0003899 | 0.002625153 |
| CD4 | 5970.284299 | -4.031028476 | 0.089575497 | 0 | 0 |
| LMO2 | 5.840280796 | -4.023639706 | 1.503867981 | 0.000412191 | 0.002758083 |
| CCL4L2 | 229.8945406 | -4.010173296 | 0.215994358 | 6.59E-78 | 3.56E-75 |
| MYLK-AS1 | 11.32921682 | -3.942427124 | 1.003607087 | 1.44E-05 | 0.000140994 |
| MAP2 | 13.45375074 | 3.933701071 | 0.894120317 | 1.43E-06 | 1.74E-05 |
| CHI3L2 | 27.74653317 | -3.93288793 | 0.614695231 | 2.76E-11 | 7.20E-10 |
| SCARNA9L | 9.305884295 | 3.913445799 | 1.477228795 | 0.000587593 | 0.00376029 |
| LOC124903989 | 17.65184257 | 3.886047268 | 0.743521502 | 2.48E-08 | 4.15E-07 |
| SCN3A | 316.6222878 | 3.864490757 | 0.186972192 | 5.16E-96 | 4.06E-93 |
| TERC | 8.155136105 | 3.842709422 | 1.294746577 | 0.000329916 | 0.002268024 |
| SPRR1A | 5.603403455 | -3.835483742 | 1.511530946 | 0.000513901 | 0.003346556 |
| CCL3L3 | 2425.785006 | -3.833737967 | 0.117432352 | 3.09E-234 | 1.07E-230 |
| BABAM2-AS1 | 71.53777806 | -3.818300363 | 0.36469362 | 1.00E-26 | 9.46E-25 |
| MS4A1 | 19.75236089 | 3.814789124 | 0.723900639 | 1.55E-08 | 2.67E-07 |
| COL4A5 | 79.8732555 | -3.765177796 | 0.344201302 | 1.23E-28 | 1.29E-26 |

| | | | | | |
|---|---|---|---|---|---|
| DTX3L | 125.2926269 | 3.752239384 | 0.277861061 | 1.15E-42 | 2.28E-40 |
| LGALS9 | 38.71071591 | 3.731700094 | 0.477827737 | 7.11E-16 | 3.22E-14 |
| GBP5 | 4.857755924 | 3.726306195 | 1.595871829 | 0.000880804 | 0.005324436 |
| LINC00668 | 4.907815117 | 3.722078818 | 1.593746563 | 0.000837506 | 0.00510313 |
| TNFSF9 | 1991.064812 | -3.712068328 | 0.119712967 | 1.12E-211 | 3.23E-208 |
| EDA2R | 102.1796753 | -3.711800637 | 0.293482817 | 1.40E-37 | 2.38E-35 |
| LOC124904613 | 7.601851883 | 3.647213949 | 1.366665338 | 0.000657117 | 0.004142447 |
| PARP9 | 91.50215039 | 3.635896044 | 0.299055061 | 6.62E-35 | 9.55E-33 |
| COL6A2 | 15.51761783 | 3.610408687 | 0.802254001 | 6.45E-07 | 8.45E-06 |
| LOC728989 | 15.07504937 | 3.60860605 | 0.745294941 | 1.58E-07 | 2.31E-06 |
| KRT8P3 | 25.60712409 | -3.608328585 | 0.57891755 | 5.34E-11 | 1.32E-09 |
| KCNK1 | 241.5284903 | -3.570696542 | 0.191628355 | 1.86E-78 | 1.07E-75 |
| DMD | 12.791182 | 3.566770599 | 0.834888128 | 2.18E-06 | 2.55E-05 |
| CDKN1A | 7930.94239 | -3.563453643 | 0.102330815 | 2.01E-266 | 1.16E-262 |
| ISLR | 4.493598438 | 3.555558041 | 1.620601087 | 0.001226983 | 0.007059027 |
| PKHD1L1 | 20.1123848 | -3.542911804 | 0.66018529 | 9.42E-09 | 1.69E-07 |
| KLRC4 | 4.497122434 | 3.531460378 | 1.641784016 | 0.001329347 | 0.007555071 |
| CD96 | 759.0548862 | 3.528819831 | 0.126703981 | 6.96E-172 | 1.51E-168 |
| TCEAL9 | 16.0544389 | -3.527944821 | 0.762629516 | 3.67E-07 | 5.05E-06 |
| MIR9898 | 13.32291774 | -3.518656063 | 0.83791721 | 3.18E-06 | 3.57E-05 |
| EPSTI1 | 25.94066101 | 3.510372488 | 0.570481798 | 8.34E-11 | 2.01E-09 |
| LINC01781 | 20.24261091 | 3.502530903 | 0.630085907 | 2.84E-09 | 5.56E-08 |
| TP53 | 902.0498131 | -3.4719142 | 0.113053972 | 3.22E-208 | 7.96E-205 |
| PRKAG3 | 57.68421695 | -3.468386216 | 0.374201097 | 2.95E-21 | 2.00E-19 |
| TLR9 | 70.44119782 | -3.459011458 | 0.332764645 | 3.18E-26 | 2.87E-24 |
| CNGB3 | 4.784396866 | -3.430867681 | 1.600292844 | 0.001275907 | 0.007293012 |
| MB | 43.02147064 | 3.403973101 | 0.438175675 | 7.25E-16 | 3.27E-14 |
| NPM2 | 12.12691951 | -3.384180503 | 0.832230755 | 5.98E-06 | 6.31E-05 |
| KLHL4 | 11.64081369 | 3.376928318 | 0.868759259 | 9.43E-06 | 9.54E-05 |
| KIF13A | 18.55257482 | 3.342599294 | 0.649627949 | 2.45E-08 | 4.10E-07 |
| LURAP1L | 47.11277006 | 3.335838299 | 0.412718056 | 4.87E-17 | 2.35E-15 |
| JUP | 34.74992117 | -3.287332481 | 0.491702756 | 3.10E-12 | 9.32E-11 |
| LINC01446 | 11.84272569 | -3.26494818 | 0.854276481 | 1.24E-05 | 0.000122861 |
| LOC124906331 | 6.006122794 | -3.240284975 | 1.289893432 | 0.001286714 | 0.007344107 |
| MS4A7 | 5.813954698 | 3.213473807 | 1.325978658 | 0.001318469 | 0.007498173 |
| LOC105373296 | 4.191893455 | -3.189252212 | 1.715929279 | 0.002572525 | 0.013262404 |
| MIR3648-1 | 17.81522304 | 3.145150455 | 0.983199951 | 5.63E-05 | 0.000478677 |
| PLK2 | 14.85921041 | 3.138296566 | 0.697484885 | 5.94E-07 | 7.84E-06 |
| PARP14 | 196.3774676 | 3.134508206 | 0.20321981 | 8.22E-55 | 2.54E-52 |
| LOC105376360 | 5.760132113 | -3.130058003 | 1.329051584 | 0.001744968 | 0.009574656 |
| HBE1 | 228.7909782 | -3.118141866 | 0.185059492 | 1.34E-64 | 5.38E-62 |
| SYT11 | 671.1834726 | -3.069681931 | 0.121465688 | 2.10E-142 | 2.59E-139 |
| OR51B5 | 97.43331453 | -3.069207626 | 0.267056247 | 1.40E-31 | 1.80E-29 |
| ANO4 | 19.08834871 | 3.066783665 | 0.648377247 | 1.64E-07 | 2.39E-06 |
| SVIL | 8.364838579 | 3.045724689 | 1.002827249 | 0.000177757 | 0.001316603 |
| LINC02709 | 22.89081421 | -3.028262203 | 0.552490927 | 3.85E-09 | 7.38E-08 |
| HAPLN3 | 322.5771833 | 3.027829892 | 0.17523822 | 5.05E-68 | 2.19E-65 |
| TINAGL1 | 14.52302593 | 3.025075864 | 0.760812364 | 4.53E-06 | 4.93E-05 |
| LOC124903859 | 5.708655425 | -3.0073718 | 1.312079651 | 0.001741567 | 0.009562052 |
| LY96 | 117.1518154 | -2.913856489 | 0.266836338 | 7.21E-29 | 7.61E-27 |

| | | | | | |
|---|---|---|---|---|---|
| CCL4 | 466.0539558 | -2.893238627 | 0.151917745 | 6.87E-82 | 4.25E-79 |
| BLK | 120.1793898 | -2.883380152 | 0.259229417 | 6.31E-30 | 7.23E-28 |
| NR4A3 | 1134.645525 | -2.873566775 | 0.296159184 | 1.60E-23 | 1.27E-21 |
| NTNG1 | 8.265582734 | 2.872139994 | 1.09428483 | 0.000465031 | 0.003067784 |
| CYS1 | 41.62443353 | 2.869944128 | 0.394855023 | 2.96E-14 | 1.12E-12 |
| GABRB3 | 20.67659039 | 2.860217881 | 0.557726914 | 2.24E-08 | 3.78E-07 |
| LOC107985968 | 12.4055384 | -2.856310261 | 0.755343719 | 1.22E-05 | 0.000121095 |
| LTA | 20.90696771 | -2.837080308 | 0.579039244 | 7.09E-08 | 1.10E-06 |
| AREG | 12.30180851 | -2.835708899 | 0.810172717 | 3.36E-05 | 0.000301899 |
| SLC27A6 | 5.22498211 | 2.802857274 | 1.509484804 | 0.003387667 | 0.016708697 |
| GRAMD2B | 13.26655235 | -2.792862028 | 0.735304793 | 1.12E-05 | 0.00011262 |
| LRRC38 | 24.58079843 | 2.772591803 | 0.506413044 | 3.33E-09 | 6.46E-08 |
| SLITRK2 | 8.523851534 | 2.768335987 | 0.938837408 | 0.000213405 | 0.001545604 |
| RTKN2 | 43.91815173 | 2.733675041 | 0.387739128 | 1.22E-13 | 4.34E-12 |
| H4C9 | 16.93522744 | 2.727580781 | 0.951787161 | 0.000160699 | 0.001204161 |
| ELAVL2 | 32.58039223 | 2.714958819 | 0.448987801 | 1.01E-10 | 2.42E-09 |
| RGS2 | 1218.630973 | -2.689502122 | 0.124430322 | 1.00E-104 | 8.69E-102 |
| APOL3 | 375.2064875 | 2.689404919 | 0.138083543 | 1.11E-85 | 7.09E-83 |
| CLEC7A | 64.28622785 | 2.664280406 | 0.311701138 | 8.52E-19 | 4.69E-17 |
| LINC00665 | 4.95800586 | -2.660045711 | 1.408306137 | 0.003661353 | 0.017764342 |
| EPHA3 | 31.69791287 | -2.643863518 | 0.460786612 | 6.25E-10 | 1.35E-08 |
| MT1G | 196.8912637 | -2.635623832 | 0.206247972 | 2.20E-38 | 3.85E-36 |
| GPR162 | 9.666850925 | -2.629988053 | 0.884214731 | 0.000202705 | 0.001479242 |
| MBOAT2 | 329.1156999 | 2.622568051 | 0.158974247 | 2.44E-62 | 8.99E-60 |
| A4GALT | 18.64414587 | 2.618981292 | 0.618336751 | 1.36E-06 | 1.66E-05 |
| KLRF2 | 4.735300747 | 2.618505229 | 1.551755022 | 0.004576444 | 0.021401643 |
| LINC02446 | 17.44660666 | 2.61356587 | 0.63128475 | 2.07E-06 | 2.44E-05 |
| LPCAT2 | 82.78386115 | -2.60764829 | 0.281070625 | 1.35E-21 | 9.40E-20 |
| ESRP1 | 13.73467434 | 2.603325258 | 0.703937917 | 1.35E-05 | 0.000132433 |
| LOC102724340 | 40.55606316 | -2.554208655 | 0.405388564 | 2.02E-11 | 5.40E-10 |
| H2BU1 | 21.08631037 | 2.551508452 | 0.918565096 | 0.000194161 | 0.001422284 |
| APOL4 | 4.506821846 | 2.546459854 | 1.549821994 | 0.004964942 | 0.022939673 |
| NR4A1 | 5775.123951 | -2.546387393 | 0.129822738 | 7.83E-87 | 5.65E-84 |
| LOC105373582 | 6.611797347 | 2.543623146 | 1.070769737 | 0.001004419 | 0.005966905 |
| BCYRN1 | 15.0715307 | -2.540725303 | 0.648811567 | 6.59E-06 | 6.90E-05 |
| EEF1A2 | 6.620267096 | 2.536286174 | 1.106531766 | 0.001206656 | 0.00696784 |
| LOC107985211 | 29.79619695 | -2.498872118 | 0.476428625 | 1.12E-08 | 1.98E-07 |
| TUSC3 | 14.11071627 | 2.495691304 | 0.697651988 | 2.04E-05 | 0.0001931 |
| PLA2G2D | 38.52629357 | 2.481467488 | 0.427908756 | 3.99E-10 | 8.86E-09 |
| RNF43 | 25.3605665 | -2.476777415 | 0.512135711 | 8.53E-08 | 1.31E-06 |
| NR4A2 | 219.4195219 | -2.456137346 | 0.174743932 | 4.64E-46 | 1.11E-43 |
| CA9 | 102.582592 | 2.456006288 | 0.24432595 | 5.65E-25 | 4.77E-23 |
| CES3 | 110.9387223 | -2.447181219 | 0.241026044 | 2.38E-25 | 2.04E-23 |
| ARHGEF6 | 236.0562548 | -2.446327451 | 0.177432824 | 1.70E-44 | 3.72E-42 |
| PHACTR1 | 66.79356164 | -2.438509553 | 0.30501343 | 1.08E-16 | 5.09E-15 |
| BNC2 | 28.10738223 | -2.433444204 | 0.464279921 | 1.03E-08 | 1.84E-07 |
| SNORD14E | 17.22867341 | -2.419605966 | 0.633147661 | 7.48E-06 | 7.74E-05 |
| CYP26A1 | 22.33421733 | 2.416928736 | 0.524186848 | 2.64E-07 | 3.73E-06 |
| SMOC1 | 58.41864379 | -2.404939842 | 0.313057469 | 1.20E-15 | 5.25E-14 |
| PLK3 | 264.516415 | -2.39861159 | 0.164643462 | 3.95E-49 | 1.05E-46 |

| | | | | | |
|---|---|---|---|---|---|
| ID2B | 10.02939062 | 2.393172275 | 0.945220013 | 0.000495024 | 0.003234843 |
| STAT1 | 11326.45231 | 2.388936891 | 0.073002103 | 4.63E-236 | 2.01E-232 |
| ASS1P3 | 5.974964526 | -2.388282822 | 1.423535834 | 0.003855898 | 0.018542791 |
| LOC105377663 | 5.291434213 | 2.386628735 | 1.313458252 | 0.003231764 | 0.016081742 |
| CRYAB | 8.073983908 | -2.386335078 | 1.126094741 | 0.001577039 | 0.00878065 |
| LOC105376505 | 56.96983694 | -2.381539664 | 0.324810347 | 1.92E-14 | 7.39E-13 |
| ANXA9 | 25.96872745 | -2.371631297 | 0.480459505 | 5.24E-08 | 8.34E-07 |
| ACKR3 | 44.23799267 | 2.370540923 | 0.358617438 | 2.66E-12 | 8.00E-11 |
| ASS1 | 1087.854414 | -2.365162153 | 0.092559081 | 3.02E-145 | 4.36E-142 |
| BASP1 | 441.6574293 | 2.357939904 | 0.131552806 | 4.64E-73 | 2.29E-70 |
| CYSLTR1 | 123.2469847 | -2.339376787 | 0.220946785 | 2.86E-27 | 2.81E-25 |
| TTC39C | 30.03957644 | 2.335115536 | 0.447196406 | 1.17E-08 | 2.05E-07 |
| BCYRN1P1 | 14.64135267 | -2.333365744 | 0.645610725 | 2.00E-05 | 0.000190141 |
| LINC01053 | 7.598863908 | -2.321754591 | 1.067790094 | 0.001484571 | 0.008325234 |
| CBR3 | 44.93768414 | 2.317641947 | 0.356879283 | 5.65E-12 | 1.64E-10 |
| RAPGEF4 | 20.14428696 | -2.300938091 | 0.556080592 | 2.09E-06 | 2.46E-05 |
| SELENOP | 9.42760388 | -2.297820402 | 0.85176415 | 0.000403252 | 0.00270349 |
| BCAM | 9.95804338 | 2.29351186 | 0.816687049 | 0.000260424 | 0.001840719 |
| H2AC16 | 25.23265059 | 2.291303089 | 0.750603591 | 8.78E-05 | 0.000707799 |
| SLC12A7 | 133.1477459 | -2.28169315 | 0.216826067 | 4.35E-27 | 4.26E-25 |
| EVPL | 8.980472097 | -2.272546053 | 0.952064729 | 0.000769335 | 0.004753171 |
| SNORD14D | 39.84445708 | -2.236442096 | 0.387468445 | 5.27E-10 | 1.15E-08 |
| COL6A1 | 25.00882029 | 2.236416426 | 0.483149891 | 2.28E-07 | 3.26E-06 |
| ARID5B | 45.41588402 | 2.235605956 | 0.356248348 | 2.10E-11 | 5.57E-10 |
| KRT8P33 | 14.43656207 | -2.234835283 | 0.66105866 | 4.12E-05 | 0.000361306 |
| AMZ1 | 107.6429486 | 2.233400014 | 0.232414493 | 4.58E-23 | 3.56E-21 |
| SNORD78 | 20.26249648 | -2.230886154 | 0.534604651 | 1.85E-06 | 2.20E-05 |
| DUSP4 | 1690.301843 | -2.227679988 | 0.093484445 | 1.47E-126 | 1.60E-123 |
| LOC124905129 | 51.92114478 | 2.222080934 | 0.324269542 | 4.67E-13 | 1.56E-11 |
| SLC8A1-AS1 | 11.60094057 | -2.221869225 | 0.763331281 | 0.000185041 | 0.001365301 |
| RNF144B | 11.22799544 | 2.209670383 | 0.756955306 | 0.000189098 | 0.001390492 |
| NALT1 | 4.388857945 | -2.19705561 | 1.832559648 | 0.008402268 | 0.035281782 |
| SNORD79 | 35.96943852 | -2.19428254 | 0.433987526 | 2.17E-08 | 3.67E-07 |
| CLEC9A | 5.836749353 | 2.186500802 | 1.187937445 | 0.002834491 | 0.014411298 |
| ANKRD20A12P | 4.493915209 | -2.183632081 | 1.697047686 | 0.00723475 | 0.03133671 |
| ZNF222 | 8.17701125 | -2.173931506 | 0.933030915 | 0.000985157 | 0.005868581 |
| ACTA2 | 31.37384657 | -2.171995152 | 0.443167874 | 6.62E-08 | 1.03E-06 |
| RTP4 | 11.0206025 | 2.15498843 | 0.756445536 | 0.000229424 | 0.001651946 |
| H2BC9 | 66.17928415 | 2.150924859 | 1.053400567 | 0.001049967 | 0.006194985 |
| ASTN1 | 267.0272031 | -2.150250472 | 0.153842033 | 1.81E-45 | 4.12E-43 |
| LOXL4 | 951.2555315 | -2.14750902 | 0.096296599 | 2.85E-111 | 2.60E-108 |
| LOC105370956 | 16.93748166 | -2.138665756 | 0.623391151 | 3.21E-05 | 0.000290012 |
| TNFRSF10B | 1943.431794 | -2.128675111 | 0.089128569 | 4.12E-127 | 4.75E-124 |
| LAMP5 | 11.53188999 | 2.125255672 | 0.887454264 | 0.000693697 | 0.004339864 |
| H3C1 | 8.797684456 | 2.120757892 | 1.267901457 | 0.002697235 | 0.013814852 |
| NRP2 | 346.0563669 | -2.120629633 | 0.145992601 | 4.73E-49 | 1.24E-46 |
| DHRS2 | 994.4340626 | -2.118141234 | 0.106340986 | 1.38E-89 | 1.04E-86 |
| RFLNB | 262.2396557 | -2.115324 | 0.150774353 | 8.30E-46 | 1.94E-43 |
| SELL | 139.3570833 | 2.113276056 | 0.201905263 | 8.73E-27 | 8.32E-25 |
| FOSL2 | 6777.374927 | -2.106877563 | 0.082486719 | 4.67E-145 | 6.23E-142 |

| | | | | | |
|---|---|---|---|---|---|
| LOC124903001 | 38.9976981 | -2.099718707 | 0.375883066 | 1.51E-09 | 3.08E-08 |
| H4C12 | 48.98013219 | 2.08654831 | 1.10952454 | 0.001435549 | 0.00808962 |
| SLFN5 | 122.0880813 | 2.081382574 | 0.25089228 | 6.83E-18 | 3.50E-16 |
| MYO3B | 4.985763685 | -2.077876717 | 1.436868225 | 0.005912597 | 0.026450126 |
| HOXB6 | 20.59797775 | 2.074034595 | 0.533264226 | 5.83E-06 | 6.19E-05 |
| ATF3 | 2453.113568 | -2.073807503 | 0.0791779 | 2.36E-152 | 4.53E-149 |
| ELAPOR1 | 25.71161842 | 2.071394217 | 0.467865026 | 5.58E-07 | 7.40E-06 |
| H2AC14 | 50.2363233 | 2.057794383 | 1.217826462 | 0.001932328 | 0.01042434 |
| RNA45SN2 | 311131.212 | 2.055034424 | 0.95601145 | 0.000834601 | 0.005089012 |
| CMBL | 668.0447829 | -2.046072299 | 0.116852811 | 6.18E-70 | 2.74E-67 |
| PELI2 | 50.78690879 | 2.044897085 | 0.367384545 | 1.55E-09 | 3.16E-08 |
| RNA45SN3 | 312291.9349 | 2.043675997 | 0.960959598 | 0.000877109 | 0.005307092 |
| RNA45SN5 | 313407.9539 | 2.040772744 | 0.960430518 | 0.000881341 | 0.005324436 |
| RNA45SN1 | 314658.569 | 2.039653265 | 0.959833523 | 0.000881513 | 0.005324436 |
| LOC105372717 | 120.5361927 | -2.037610287 | 0.270263344 | 3.17E-15 | 1.32E-13 |
| H4C11 | 48.7841051 | 2.028891603 | 1.016300714 | 0.001204304 | 0.006958602 |
| PLEK | 756.0058823 | 2.027263529 | 0.136673859 | 5.28E-51 | 1.45E-48 |
| LINC02253 | 26.4483915 | 2.022068149 | 0.45816119 | 6.15E-07 | 8.09E-06 |
| APOBEC3H | 247.2003554 | -2.012971488 | 0.152187854 | 4.00E-41 | 7.53E-39 |
| ZNF165 | 143.2473952 | -2.00702228 | 0.203618783 | 4.40E-24 | 3.61E-22 |
| AOC1 | 113.0989267 | 1.993723798 | 0.225537225 | 6.38E-20 | 3.94E-18 |
| MX1 | 44.00718407 | 1.990815247 | 0.340589678 | 3.32E-10 | 7.42E-09 |
| PMP22 | 30.33235972 | 1.988609448 | 0.47575738 | 1.53E-06 | 1.84E-05 |
| ELF3 | 587.4337175 | -1.9844583 | 0.120659925 | 8.45E-62 | 2.99E-59 |
| RNA45SN4 | 326266.0926 | 1.975293047 | 0.968818712 | 0.001066975 | 0.006284946 |
| SNORD44 | 30.20033283 | -1.974896765 | 0.425376738 | 2.02E-07 | 2.90E-06 |
| RPS27L | 1821.407173 | -1.974855414 | 0.087873283 | 4.62E-113 | 4.71E-110 |
| H2AC13 | 27.67822344 | 1.971626452 | 0.777785764 | 0.000398004 | 0.002672442 |
| FOXP2 | 49.82385192 | 1.966302276 | 0.344713873 | 7.43E-10 | 1.59E-08 |
| H3C3 | 46.64159012 | 1.963245035 | 1.258447027 | 0.002376262 | 0.012424433 |
| SLC45A3 | 13.98730096 | 1.96130651 | 0.643873001 | 0.000123239 | 0.000954015 |
| EVI2B | 1685.180504 | 1.96099725 | 0.111269711 | 9.51E-71 | 4.45E-68 |
| GATA3 | 31.71946557 | 1.956167325 | 0.493815511 | 3.82E-06 | 4.21E-05 |
| TNFRSF10D | 798.6283855 | -1.955629073 | 0.103820662 | 2.91E-80 | 1.74E-77 |
| CKMT1B | 173.2142723 | -1.953798369 | 0.185715351 | 5.74E-27 | 5.55E-25 |
| WNT11 | 165.5184063 | -1.947694751 | 0.224560227 | 2.87E-19 | 1.66E-17 |
| FLT1 | 8.296836003 | 1.942814033 | 1.038900234 | 0.002179978 | 0.011562697 |
| TMEM132D | 7.831871127 | 1.938314354 | 0.955731237 | 0.00176199 | 0.009646658 |
| PPIAP14 | 13.94733422 | 1.934147666 | 0.66477602 | 0.000183783 | 0.001357172 |
| CELF6 | 7.10208132 | -1.933808007 | 1.177286554 | 0.003407268 | 0.016786248 |
| HHEX | 133.6152759 | 1.924948805 | 0.208717682 | 1.84E-21 | 1.27E-19 |
| FAM167A | 273.0778366 | -1.91931946 | 0.146861669 | 3.94E-40 | 7.25E-38 |
| GPR174 | 11.92111845 | 1.915154516 | 0.816250683 | 0.000802516 | 0.004924581 |
| PTP4A3 | 890.8913282 | -1.913514227 | 0.119908763 | 2.00E-58 | 6.30E-56 |
| TGIF1 | 16.02258781 | 1.907823018 | 0.672438627 | 0.000207062 | 0.001506597 |
| H3P4 | 23.71743578 | 1.907485675 | 0.859012829 | 0.000857043 | 0.005203861 |
| IGHG3 | 27.72021585 | -1.906968442 | 0.452154812 | 1.55E-06 | 1.87E-05 |
| GIMAP7 | 11.53108593 | 1.896912325 | 0.751116464 | 0.000538772 | 0.003477416 |
| IL32 | 62.52712574 | 1.895830277 | 0.291824949 | 5.23E-12 | 1.52E-10 |
| RPL22L1 | 4460.314002 | -1.893926068 | 0.088155613 | 1.39E-103 | 1.15E-100 |

| | | | | | |
|---|---|---|---|---|---|
| ASB2 | 105.0019139 | 1.87723614 | 0.226614195 | 8.35E-18 | 4.26E-16 |
| NFKBID | 86.43216596 | -1.874423769 | 0.263313237 | 7.65E-14 | 2.74E-12 |
| LOC105371457 | 33.4723402 | 1.874139692 | 0.400991019 | 1.82E-07 | 2.64E-06 |
| FSTL4 | 56.47756737 | -1.871523739 | 0.318291267 | 2.47E-10 | 5.58E-09 |
| MT1E | 144.316286 | -1.868444879 | 0.203258171 | 2.77E-21 | 1.89E-19 |
| BTBD3 | 99.09552731 | -1.86824223 | 0.244852048 | 1.54E-15 | 6.63E-14 |
| TYRP1 | 6.832309778 | 1.865287615 | 1.066643305 | 0.003030559 | 0.01521165 |
| CKMT1A | 160.6636241 | -1.860593583 | 0.187477606 | 2.52E-24 | 2.09E-22 |
| MARCHF3 | 106.5597357 | 1.857601546 | 0.223199552 | 5.76E-18 | 2.96E-16 |
| THEMIS2 | 303.2437641 | -1.847184295 | 0.16282913 | 6.11E-31 | 7.51E-29 |
| GCSAM | 266.4549295 | 1.842079189 | 0.156296205 | 2.95E-33 | 4.09E-31 |
| ST14 | 28.13653835 | 1.841262365 | 0.450800182 | 2.55E-06 | 2.93E-05 |
| PTGER4 | 67.78906492 | 1.836613943 | 0.290569316 | 1.56E-11 | 4.28E-10 |
| RASSF2 | 102.1384833 | 1.833265965 | 0.244367791 | 4.18E-15 | 1.72E-13 |
| BTK | 968.7968956 | -1.826688858 | 0.098462793 | 4.52E-78 | 2.53E-75 |
| GAS1 | 8.689285099 | 1.820332435 | 1.017295415 | 0.002531232 | 0.01309099 |
| ADORA2A-AS1 | 5.582712182 | -1.814935983 | 1.315761703 | 0.005665287 | 0.025549629 |
| MCAM | 220.2000879 | -1.803557261 | 0.157869374 | 2.39E-31 | 3.02E-29 |
| LOC124904529 | 118.7467419 | -1.803374859 | 0.233685101 | 6.96E-16 | 3.16E-14 |
| TTC39A | 22.08661327 | -1.80113274 | 0.517629204 | 2.82E-05 | 0.000258107 |
| LTF | 455.8884022 | -1.789998159 | 0.116820257 | 3.63E-54 | 1.08E-51 |
| LOC124903462 | 6.519916694 | -1.787318107 | 1.226687971 | 0.004749637 | 0.022121962 |
| SNORD47 | 57.46347329 | -1.784010767 | 0.351787903 | 2.12E-08 | 3.59E-07 |
| LINC00997 | 49.04461707 | -1.781683811 | 0.354515921 | 2.70E-08 | 4.50E-07 |
| TSPAN13 | 652.6864252 | -1.780081556 | 0.163965313 | 1.05E-28 | 1.10E-26 |
| DPYSL4 | 287.6052991 | -1.773190175 | 0.150830163 | 4.89E-33 | 6.66E-31 |
| DOCK2 | 73.05872819 | -1.768576796 | 0.264976138 | 1.70E-12 | 5.28E-11 |
| GRAMD1B | 8.922156905 | 1.760405572 | 0.910151299 | 0.002045068 | 0.010927011 |
| LOC124905968 | 29.14116432 | -1.755230024 | 0.48727784 | 1.92E-05 | 0.000183401 |
| H2BP1 | 17.09107295 | 1.752319268 | 1.072499021 | 0.002624462 | 0.013470008 |
| CLDN14 | 8.730214677 | -1.749310014 | 1.047001584 | 0.0029622 | 0.014907414 |
| SAMD11 | 20.69726842 | 1.748678095 | 0.55974898 | 9.37E-05 | 0.000751481 |
| MT1X | 360.2456956 | -1.747159243 | 0.158759036 | 2.66E-29 | 2.89E-27 |
| DUSP26 | 19.24542606 | -1.745926356 | 0.541916725 | 6.98E-05 | 0.000579125 |
| MME | 188.1859982 | 1.738340413 | 0.168764328 | 4.69E-26 | 4.19E-24 |
| NMI | 142.0308359 | 1.735399251 | 0.196532183 | 6.88E-20 | 4.22E-18 |
| NLRP11 | 164.3778261 | -1.729260978 | 0.18145238 | 1.05E-22 | 7.94E-21 |
| CXCR1 | 5.217613354 | 1.726267588 | 1.518902292 | 0.006897184 | 0.030104554 |
| TAGAP | 97.22407738 | 1.721181918 | 0.227834421 | 2.80E-15 | 1.17E-13 |
| PRLR | 9.242239071 | 1.711067452 | 1.027635631 | 0.003069568 | 0.015380704 |
| ARL11 | 45.01143195 | -1.708126331 | 0.339195418 | 3.17E-08 | 5.23E-07 |
| UBD | 36.19663993 | 1.701512446 | 0.389071401 | 7.58E-07 | 9.80E-06 |
| IFIT5 | 22.8708764 | 1.700720888 | 0.498121612 | 3.53E-05 | 0.000315778 |
| CMAHP | 62.37403338 | 1.700171051 | 0.276995321 | 5.55E-11 | 1.37E-09 |
| CRABP2 | 24.38624934 | -1.699377388 | 0.48601722 | 2.79E-05 | 0.000255913 |
| SNORD99 | 5.877681683 | -1.697853909 | 1.488144942 | 0.006475116 | 0.028553497 |
| BCL9L | 183.0705869 | 1.696883541 | 0.174458883 | 1.54E-23 | 1.23E-21 |
| MDM2 | 14157.60344 | -1.69675931 | 0.106293258 | 1.76E-58 | 5.66E-56 |
| B3GALT4 | 12.10681343 | 1.696662993 | 0.740804625 | 0.000966244 | 0.00577379 |
| IRF1-AS1 | 33.91401702 | 1.693977956 | 0.385261296 | 6.87E-07 | 8.95E-06 |

| | | | | | |
|---|---|---|---|---|---|
| LOC105370449 | 8.098638687 | 1.693714163 | 1.279770606 | 0.004693926 | 0.021903722 |
| AHR | 30.32659886 | 1.690322582 | 0.415456768 | 2.74E-06 | 3.12E-05 |
| SHTN1 | 56.44007008 | -1.675281691 | 0.301212895 | 1.73E-09 | 3.50E-08 |
| MAGEA9 | 54.27611259 | -1.670746742 | 0.305049317 | 2.94E-09 | 5.75E-08 |
| ID2 | 1078.517629 | 1.669634665 | 0.103920895 | 2.75E-59 | 9.54E-57 |
| NSG2 | 8.440283715 | -1.666906386 | 0.973966036 | 0.003239865 | 0.016108164 |
| MIR34AHG | 14.75436958 | -1.66360688 | 0.624482635 | 0.000398568 | 0.002675195 |
| IGHG1 | 51.39846722 | -1.660960172 | 0.318644505 | 1.30E-08 | 2.27E-07 |
| CLGN | 127.7786441 | -1.658373445 | 0.203560762 | 2.66E-17 | 1.32E-15 |
| GADD45A | 281.744787 | -1.652707224 | 0.142981128 | 4.67E-32 | 6.22E-30 |
| LYPD6B | 49.09759412 | 1.64394022 | 0.320555309 | 1.87E-08 | 3.19E-07 |
| OGFRL1 | 1651.04419 | 1.643271307 | 0.159710185 | 5.07E-26 | 4.50E-24 |
| MAGEA9B | 68.28275371 | -1.632405997 | 0.270182397 | 1.05E-10 | 2.48E-09 |
| DTX4 | 201.1945481 | 1.631071677 | 0.166471969 | 8.32E-24 | 6.73E-22 |
| LINC02909 | 278.0738195 | 1.627117859 | 0.144394285 | 1.32E-30 | 1.60E-28 |
| HIVEP1 | 974.2170232 | -1.622841243 | 0.122204752 | 1.90E-41 | 3.62E-39 |
| LMO3 | 112.0142918 | 1.620572779 | 0.247153107 | 3.51E-12 | 1.04E-10 |
| IGHG2 | 716.3910183 | -1.617497016 | 0.107156271 | 1.47E-52 | 4.30E-50 |
| CCDC187 | 95.48742829 | -1.616714123 | 0.255350882 | 1.77E-11 | 4.79E-10 |
| APOBEC3G | 1390.474284 | 1.6144241 | 0.091740018 | 1.88E-70 | 8.56E-68 |
| CLCF1 | 26.76743524 | -1.613802827 | 0.476768082 | 4.02E-05 | 0.000353152 |
| SPRR2F | 10.88273138 | -1.607945653 | 0.756738434 | 0.001526816 | 0.008528993 |
| SLC6A5 | 27.11123183 | 1.606968134 | 0.452895624 | 2.23E-05 | 0.000208913 |
| FMNL3 | 284.4817312 | 1.601551352 | 0.139651265 | 1.37E-31 | 1.78E-29 |
| RNASE6 | 7.410390118 | 1.599884625 | 1.198964972 | 0.005077549 | 0.023378866 |
| SLC7A8 | 6.25996869 | -1.598223881 | 1.368154 | 0.006579442 | 0.028939854 |
| MUC3A | 11.5817597 | 1.598033471 | 0.770369941 | 0.001603768 | 0.008912854 |
| RPS6KA2 | 14.27892953 | 1.589612265 | 0.718062232 | 0.001166503 | 0.006778637 |
| CA2 | 263.6214067 | 1.588240864 | 0.14473558 | 3.72E-29 | 4.00E-27 |
| SYK | 35.07033498 | 1.586195323 | 0.394025826 | 3.46E-06 | 3.85E-05 |
| CD79A | 22.10207194 | -1.583473835 | 0.51375648 | 0.000106087 | 0.000838481 |
| WARS1 | 20714.34058 | 1.580150026 | 0.06093493 | 7.76E-150 | 1.22E-146 |
| LOC107985079 | 31.92451882 | 1.579093574 | 0.401650435 | 4.98E-06 | 5.36E-05 |
| TRAV22 | 14.7485584 | 1.570582478 | 0.639163626 | 0.000654539 | 0.004130704 |
| NCALD | 264.9870797 | 1.56945026 | 0.152072556 | 4.10E-26 | 3.68E-24 |
| SPRY2 | 101.4922937 | -1.55576882 | 0.230346226 | 9.94E-13 | 3.18E-11 |
| SFXN3 | 45.27220805 | 1.555279868 | 0.388358823 | 3.57E-06 | 3.97E-05 |
| SPINK2 | 171.591708 | 1.55229198 | 0.203830998 | 1.88E-15 | 7.99E-14 |
| CCDC122 | 9.985651362 | 1.551182275 | 0.982375025 | 0.003659136 | 0.017759322 |
| ST8SIA4 | 1510.25339 | 1.549179466 | 0.113855816 | 2.48E-43 | 5.17E-41 |
| ZNF829 | 26.71492826 | -1.548971193 | 0.443097555 | 2.75E-05 | 0.000252202 |
| KIAA1522 | 12.43751721 | 1.547856211 | 0.785281809 | 0.001886863 | 0.010236467 |
| GBP3 | 86.28111639 | 1.536069822 | 0.250667429 | 5.99E-11 | 1.47E-09 |
| RSPH1 | 11.36615287 | -1.531125174 | 0.788137136 | 0.002206322 | 0.011676917 |
| IRF7 | 69.91285935 | 1.524053472 | 0.26845499 | 9.56E-10 | 2.01E-08 |
| IGHG4 | 44.12770379 | -1.521471446 | 0.335669587 | 3.83E-07 | 5.23E-06 |
| CAMKV | 266.153161 | -1.51834464 | 0.144850342 | 7.80E-27 | 7.50E-25 |
| MARCKS | 2947.925889 | -1.51723896 | 0.117453679 | 2.47E-39 | 4.46E-37 |
| C4orf50 | 18.62752023 | 1.515078617 | 0.557441613 | 0.000336641 | 0.00230602 |
| LBH | 377.7955767 | 1.512135055 | 0.138018789 | 4.78E-29 | 5.11E-27 |

| | | | | | |
|---|---|---|---|---|---|
| IRF1 | 677.7656015 | 1.510081665 | 0.109272139 | 1.45E-44 | 3.21E-42 |
| IER5 | 872.2276816 | -1.509266925 | 0.12386976 | 3.02E-35 | 4.43E-33 |
| MGC70870 | 25.49281994 | -1.50753626 | 0.464786215 | 6.78E-05 | 0.000564758 |
| DIO2 | 34.26698411 | -1.502508785 | 0.449665642 | 4.83E-05 | 0.000416163 |
| ARHGAP44 | 51.58601946 | 1.501888114 | 0.348846996 | 1.02E-06 | 1.27E-05 |
| PCBP3 | 8.051910892 | 1.499098445 | 0.977824441 | 0.004274707 | 0.020203357 |
| ADPGK-AS1 | 8.893484886 | -1.497044987 | 1.124586601 | 0.004963934 | 0.022939673 |
| CYP19A1 | 25.0482103 | 1.49543897 | 0.468049144 | 7.96E-05 | 0.000650229 |
| BAMBI | 137.3132889 | 1.492610645 | 0.189976239 | 2.85E-16 | 1.31E-14 |
| F2RL3 | 21.49816547 | 1.483238097 | 0.509391714 | 0.000193684 | 0.001419392 |
| DUSP6 | 679.968549 | 1.47999342 | 0.121303934 | 2.08E-35 | 3.22E-33 |
| FOS | 668.7186523 | -1.475895341 | 0.121111581 | 2.75E-35 | 4.11E-33 |
| IGHGP | 49.26109975 | -1.475346136 | 0.325324129 | 3.81E-07 | 5.21E-06 |
| PDGFRB | 18.87715308 | 1.470564742 | 0.533323326 | 0.000306009 | 0.002122209 |
| RNA28SN2 | 220696.1314 | 1.468945779 | 0.991819202 | 0.003289985 | 0.016301191 |
| NCF1 | 11.54590463 | 1.467028632 | 0.728138614 | 0.001896198 | 0.010264604 |
| RTN4RL1 | 11.83537296 | 1.466411971 | 0.779735848 | 0.002378662 | 0.012429481 |
| TERT | 824.8776787 | 1.465598786 | 0.138123781 | 1.89E-27 | 1.88E-25 |
| NEAT1 | 447.9517935 | -1.463333893 | 0.192790724 | 2.02E-15 | 8.59E-14 |
| RNA28SN1 | 224842.5779 | 1.463259365 | 0.997429297 | 0.003355659 | 0.01657916 |
| LOC124906134 | 14.42530239 | -1.462089495 | 0.680189719 | 0.001459955 | 0.008208454 |
| RNA28SN5 | 223634.9035 | 1.461423108 | 0.998969147 | 0.003375474 | 0.016653298 |
| HID1 | 327.172936 | -1.460771779 | 0.163248247 | 2.90E-20 | 1.87E-18 |
| SERPINB9 | 146.5965016 | 1.458221366 | 0.204350076 | 6.93E-14 | 2.52E-12 |
| RHOD | 70.15363657 | -1.457411257 | 0.279023776 | 1.30E-08 | 2.26E-07 |
| RASL11A | 45.80454524 | -1.456890469 | 0.337702365 | 1.03E-06 | 1.29E-05 |
| PLAC8 | 275.1342475 | -1.456854178 | 0.151359353 | 4.92E-23 | 3.80E-21 |
| SMIM10 | 22.91101503 | 1.456062829 | 0.485386577 | 0.000151087 | 0.001142523 |
| GGT7 | 127.7526452 | 1.456007117 | 0.205132041 | 9.34E-14 | 3.34E-12 |
| NAMPTP1 | 2029.567927 | -1.45569668 | 0.111271711 | 2.93E-40 | 5.45E-38 |
| RNA28SN3 | 221948.604 | 1.453905689 | 1.004843864 | 0.003454299 | 0.016969672 |
| DNAJC12 | 148.5225697 | -1.452410041 | 0.182479974 | 1.31E-16 | 6.15E-15 |
| PLAG1 | 98.46136385 | -1.448795227 | 0.239275885 | 9.28E-11 | 2.23E-09 |
| HPSE | 48.94096784 | 1.448504677 | 0.34935841 | 2.16E-06 | 2.53E-05 |
| LINC01160 | 8.823995201 | -1.448348551 | 1.003284932 | 0.004876722 | 0.022616547 |
| LOC124905332 | 373262.0111 | 1.445396497 | 1.003597188 | 0.003497656 | 0.017143761 |
| LOC124905315 | 373029.8804 | 1.444755016 | 1.003774599 | 0.00350274 | 0.017160348 |
| KMT2E-AS1 | 40.51392386 | -1.442585046 | 0.354633889 | 3.27E-06 | 3.67E-05 |
| HCN3 | 25.53960022 | -1.441585812 | 0.476643958 | 0.000140151 | 0.001066339 |
| SNORA33 | 17.46160447 | -1.439833726 | 0.634843433 | 0.001076165 | 0.006327994 |
| PDE4DIPP1 | 14.35720492 | 1.438988008 | 0.641954452 | 0.00118516 | 0.006868612 |
| ABCF2P1 | 14.36274879 | -1.438224984 | 0.67182816 | 0.001488879 | 0.008343988 |
| HERPUD1 | 39937.12035 | -1.438102388 | 0.073617189 | 3.46E-86 | 2.30E-83 |
| LOC107985563 | 7.658674627 | 1.43775512 | 1.204081624 | 0.006177682 | 0.02748268 |
| S1PR1 | 779.446799 | 1.437046817 | 0.096027069 | 9.16E-52 | 2.60E-49 |
| COLEC11 | 15.83726398 | -1.434469692 | 0.637313414 | 0.001144159 | 0.006671182 |
| NAMPT | 6486.546094 | -1.433266152 | 0.11339274 | 9.31E-38 | 1.60E-35 |
| RDH10 | 504.3775721 | -1.431517555 | 0.11741819 | 2.31E-35 | 3.53E-33 |
| RELT | 409.5621891 | -1.431078977 | 0.141438513 | 3.73E-25 | 3.16E-23 |
| KDM7A-DT | 99.82768482 | 1.42517014 | 0.228937985 | 3.27E-11 | 8.44E-10 |

| | | | | | |
|---|---|---|---|---|---|
| FLJ32255 | 37.79362274 | 1.424979842 | 0.391855704 | 1.65E-05 | 0.000159889 |
| IL21R | 707.6112031 | -1.422320277 | 0.11833494 | 2.29E-34 | 3.28E-32 |
| RIPOR2 | 1997.695036 | 1.419801178 | 0.076838332 | 2.32E-77 | 1.22E-74 |
| HOXC10 | 8.077726264 | 1.419057718 | 1.044605452 | 0.005297123 | 0.024201205 |
| DDB2 | 804.417876 | -1.418332875 | 0.105694255 | 4.03E-42 | 7.94E-40 |
| PBX3 | 140.0164727 | 1.417123001 | 0.204758474 | 3.22E-13 | 1.08E-11 |
| DSE | 213.3682213 | -1.414867434 | 0.154966696 | 5.28E-21 | 3.51E-19 |
| DMC1 | 29.64389334 | -1.410387428 | 0.547047599 | 0.000476856 | 0.003129108 |
| PRRG3 | 6.376352327 | -1.410149461 | 1.649395535 | 0.007881244 | 0.033500123 |
| NEDD9 | 28.13787245 | 1.405600935 | 0.445052946 | 8.90E-05 | 0.000716297 |
| MARCKSP1 | 229.4457327 | -1.405178346 | 0.270290843 | 1.28E-08 | 2.24E-07 |
| LOC400499 | 87.57038461 | -1.403420016 | 0.242093289 | 5.26E-10 | 1.15E-08 |
| ENO3 | 266.3428345 | -1.400243105 | 0.157220418 | 4.58E-20 | 2.88E-18 |
| RNA28SN4 | 235798.2855 | 1.399951423 | 1.025947671 | 0.003929399 | 0.01883296 |
| EPHB2 | 116.9380939 | -1.392890457 | 0.245567967 | 1.11E-09 | 2.31E-08 |
| RASSF6 | 70.03707171 | 1.390994828 | 0.292581021 | 1.36E-07 | 2.03E-06 |
| LOC100420587 | 72.19511979 | 1.387921339 | 0.285371615 | 8.01E-08 | 1.23E-06 |
| INSIG1 | 46831.31311 | -1.387743907 | 0.087686662 | 9.58E-59 | 3.13E-56 |
| GJA3 | 376.7604669 | -1.383472525 | 0.132320809 | 1.13E-26 | 1.06E-24 |
| ACOXL | 7.719464807 | -1.381027984 | 1.171056 | 0.006832983 | 0.029895594 |
| LINC01511 | 31.85997956 | 1.379641019 | 0.395163919 | 2.98E-05 | 0.00027137 |
| ZBTB46 | 94.14200075 | -1.378061553 | 0.25574726 | 5.55E-09 | 1.03E-07 |
| MIR3667HG | 53.51833282 | -1.375402761 | 0.317723894 | 1.01E-06 | 1.27E-05 |
| FDXR | 478.4357801 | -1.3724279 | 0.133424247 | 7.23E-26 | 6.33E-24 |
| LINC00958 | 85.61912093 | -1.370449355 | 0.246030601 | 1.87E-09 | 3.78E-08 |
| PREX1 | 1597.157634 | 1.368758528 | 0.083165736 | 5.09E-62 | 1.84E-59 |
| TMEM45A | 76.48424419 | -1.367162756 | 0.275234871 | 4.87E-08 | 7.80E-07 |
| CIBAR1 | 39.00428283 | -1.359430233 | 0.418673498 | 7.13E-05 | 0.000590036 |
| ADGRA2 | 99.22336593 | -1.355040601 | 0.265099499 | 2.44E-08 | 4.10E-07 |
| TPPP | 160.8403164 | -1.352527068 | 0.193618221 | 2.16E-13 | 7.47E-12 |
| LOC105370384 | 206.4814493 | -1.350575368 | 0.171704175 | 2.82E-16 | 1.30E-14 |
| ZNF230 | 27.45755017 | -1.343685157 | 0.445425887 | 0.000151343 | 0.001143831 |
| LIMA1 | 39.33209708 | -1.338721346 | 0.358599474 | 1.31E-05 | 0.000129469 |
| MCF2L | 128.2241197 | 1.337786304 | 0.196342034 | 7.02E-13 | 2.29E-11 |
| MIR1204 | 16.23666141 | -1.337519262 | 0.622567026 | 0.001541164 | 0.008598047 |
| B3GNT4 | 30.38754103 | -1.336418467 | 0.405085975 | 6.29E-05 | 0.000528942 |
| PER1 | 908.6014911 | -1.335790831 | 0.104994071 | 3.51E-38 | 6.08E-36 |
| CSMD1 | 29.16822752 | -1.332956981 | 0.453255197 | 0.000192598 | 0.001413232 |
| MSX1 | 172.4024787 | -1.331066943 | 0.221100613 | 1.40E-10 | 3.27E-09 |
| PTPRS | 50.33621243 | 1.328933762 | 0.331982367 | 4.06E-06 | 4.46E-05 |
| SLC4A8 | 164.0024505 | 1.326343614 | 0.172977971 | 1.35E-15 | 5.84E-14 |
| NDRG4 | 126.57861 | -1.325703783 | 0.220699798 | 1.45E-10 | 3.39E-09 |
| BCL11B | 8.303522078 | 1.322907348 | 1.197877351 | 0.006673895 | 0.029288353 |
| APOL6 | 589.7969667 | 1.322273771 | 0.115661601 | 2.12E-31 | 2.69E-29 |
| FAIM | 78.17793376 | -1.322006212 | 0.266328787 | 4.85E-08 | 7.79E-07 |
| MYOM2 | 354.9434904 | -1.321263924 | 0.133175162 | 2.58E-24 | 2.13E-22 |
| TMEM52 | 111.5735296 | -1.317255937 | 0.219161124 | 1.50E-10 | 3.49E-09 |
| NMUR1 | 34.20194581 | 1.315947498 | 0.390479946 | 4.80E-05 | 0.000414226 |
| LINC01634 | 12.39517898 | -1.311966515 | 0.796050081 | 0.003993732 | 0.019078472 |
| LOC124904428 | 181.0957608 | -1.311950648 | 0.190872025 | 5.15E-13 | 1.70E-11 |

| | | | | | |
|---|---|---|---|---|---|
| DMRT2 | 16.32067674 | 1.311073 | 0.659665131 | 0.002100315 | 0.011180802 |
| SLC46A3 | 21.53943841 | 1.309824785 | 0.50459008 | 0.000521861 | 0.003387204 |
| C4A | 120.5247661 | 1.305384327 | 0.210873521 | 4.65E-11 | 1.16E-09 |
| RBM33 | 2227.190221 | -1.303843384 | 0.088337453 | 1.97E-50 | 5.34E-48 |
| RCN3 | 1162.859033 | -1.302263441 | 0.106730444 | 2.70E-35 | 4.06E-33 |
| MXRA7 | 27.10758059 | 1.299630306 | 0.444438066 | 0.000203863 | 0.001485424 |
| LOC105374382 | 133.16 | -1.298948657 | 0.21762833 | 1.88E-10 | 4.31E-09 |
| DUSP9 | 22.53265337 | 1.297028447 | 0.475757006 | 0.000365285 | 0.002476759 |
| OTULINL | 288.8817302 | 1.296189355 | 0.136371158 | 1.59E-22 | 1.18E-20 |
| UNC13C | 550.7796173 | 1.294583422 | 0.195031971 | 2.22E-12 | 6.73E-11 |
| ERICH6-AS1 | 37.16794264 | 1.293106923 | 0.365096403 | 2.61E-05 | 0.00024119 |
| APLN | 63.77144833 | 1.292779165 | 0.289619077 | 5.50E-07 | 7.30E-06 |
| C4B | 121.5408777 | 1.289470436 | 0.204362136 | 2.20E-11 | 5.82E-10 |
| SNORD80 | 29.00026532 | -1.287098581 | 0.424667862 | 0.000151635 | 0.001144676 |
| PTGS1 | 48.98000554 | 1.28666596 | 0.346047083 | 1.33E-05 | 0.000130668 |
| TAP1 | 7643.945079 | 1.284113853 | 0.090800551 | 3.05E-46 | 7.43E-44 |
| DUSP5 | 18739.78783 | -1.281344761 | 0.0851316 | 2.85E-52 | 8.22E-50 |
| FGR | 557.937847 | -1.278517533 | 0.121914115 | 8.74E-27 | 8.32E-25 |
| P2RY10 | 1007.452644 | 1.278348828 | 0.112083746 | 3.16E-31 | 3.96E-29 |
| CRIP2 | 47.77148705 | 1.276315325 | 0.324920674 | 5.94E-06 | 6.28E-05 |
| LINC03016 | 7.803664375 | 1.275634949 | 1.147932372 | 0.007181423 | 0.03113688 |
| CFHR1 | 15.27132728 | 1.273142051 | 0.612470907 | 0.001804155 | 0.00983971 |
| ANKRD50 | 151.1945144 | -1.272946305 | 0.201616009 | 2.00E-11 | 5.34E-10 |
| LOC101928156 | 264.0386369 | -1.272270796 | 0.144262131 | 9.63E-20 | 5.83E-18 |
| MAPK15 | 8.888963925 | -1.271765498 | 1.113110864 | 0.007275524 | 0.031458239 |
| MBP | 522.9199825 | 1.270625102 | 0.114491249 | 9.97E-30 | 1.12E-27 |
| FAM78A | 420.0590166 | 1.270461502 | 0.127356766 | 1.62E-24 | 1.35E-22 |
| NOVA1-DT | 21.68463094 | -1.268306764 | 0.51038754 | 0.000728604 | 0.00453694 |
| RGS3 | 121.5744923 | 1.265793261 | 0.204682833 | 4.87E-11 | 1.22E-09 |
| SPAG4 | 422.9891547 | -1.263185451 | 0.129076208 | 1.17E-23 | 9.40E-22 |
| DUSP10 | 68.70117929 | -1.258370488 | 0.28244407 | 5.88E-07 | 7.77E-06 |
| BEST4 | 25.61293592 | -1.257882287 | 0.486359597 | 0.000587501 | 0.00376029 |
| TMEM132D-AS1 | 261.9557469 | 1.257060264 | 0.144406968 | 2.58E-19 | 1.50E-17 |
| TUBB3 | 1216.21307 | -1.245270958 | 0.101679388 | 1.57E-35 | 2.47E-33 |
| FBXO32 | 2030.265841 | -1.24262128 | 0.104621765 | 1.32E-33 | 1.86E-31 |
| KISS1R | 77.96161019 | -1.241176582 | 0.32324015 | 8.97E-06 | 9.11E-05 |
| LOC100419583 | 578.0968529 | 1.23763078 | 0.104552507 | 2.11E-33 | 2.95E-31 |
| ANKRD37 | 674.4096297 | -1.236945451 | 0.133398706 | 1.65E-21 | 1.14E-19 |
| JUN | 3289.24555 | -1.235457871 | 0.087080724 | 9.61E-47 | 2.41E-44 |
| FXYD7 | 24.07430871 | 1.23440814 | 0.59750379 | 0.001800946 | 0.009831071 |
| DNAAF1 | 10.92207667 | -1.233494085 | 0.839143481 | 0.005373909 | 0.024489471 |
| FAM114A1 | 45.41520411 | -1.23333135 | 0.339048056 | 2.01E-05 | 0.000190751 |
| NRIP1 | 168.4242054 | 1.232430762 | 0.206765291 | 1.90E-10 | 4.35E-09 |
| LOC107986898 | 42.68109034 | -1.231196397 | 0.358567882 | 3.88E-05 | 0.000342564 |
| IRF9 | 547.8147056 | 1.230247582 | 0.110423968 | 6.83E-30 | 7.79E-28 |
| FCRLA | 244.7829949 | 1.226456121 | 0.159055305 | 1.04E-15 | 4.56E-14 |
| LOC105372141 | 47.11729514 | 1.221646905 | 0.330874864 | 1.52E-05 | 0.000147907 |
| KANK1 | 1043.910657 | 1.221587465 | 0.097901391 | 8.07E-37 | 1.34E-34 |
| LOC124906307 | 15.80146109 | -1.221019677 | 0.63751328 | 0.002601334 | 0.013375087 |
| MT1F | 44.61990886 | -1.219806027 | 0.3517883 | 3.78E-05 | 0.000334934 |

| | | | | | |
|---|---|---|---|---|---|
| SNORD14C | 49.40928429 | -1.218122535 | 0.341671167 | 2.40E-05 | 0.000223389 |
| SLA | 69.8474631 | 1.215318028 | 0.271196591 | 5.40E-07 | 7.20E-06 |
| DUSP1 | 681.2786487 | -1.213918152 | 0.107435242 | 1.11E-30 | 1.35E-28 |
| FZD8 | 77.48883991 | 1.211358667 | 0.254037594 | 1.44E-07 | 2.13E-06 |
| ABRAXAS1 | 387.8135571 | -1.209432828 | 0.150768927 | 8.73E-17 | 4.16E-15 |
| BAX | 1680.046368 | -1.20755961 | 0.105853468 | 3.38E-31 | 4.19E-29 |
| PLD4 | 237.5413852 | 1.206176656 | 0.176796952 | 7.50E-13 | 2.43E-11 |
| TRAC | 125.2889658 | 1.20575858 | 0.22059637 | 3.69E-09 | 7.10E-08 |
| NIBAN2 | 43.56522563 | 1.2049954 | 0.379490401 | 9.98E-05 | 0.000793613 |
| UBXN10 | 70.15242014 | 1.204572137 | 0.274261974 | 8.44E-07 | 1.08E-05 |
| LOC124904791 | 15.17682818 | -1.200469638 | 0.650941717 | 0.003138549 | 0.015680975 |
| CEMIP2 | 227.9138173 | -1.20040838 | 0.178511433 | 1.42E-12 | 4.46E-11 |
| TLR1 | 38.6449516 | 1.200123586 | 0.363883884 | 6.46E-05 | 0.000541027 |
| GPR155 | 586.5595768 | 1.199680608 | 0.108687007 | 2.13E-29 | 2.34E-27 |
| LRIG1 | 1360.541014 | 1.1980347 | 0.086031389 | 3.90E-45 | 8.78E-43 |
| TXNIP | 30129.1074 | -1.196427569 | 0.070633041 | 2.18E-65 | 8.99E-63 |
| LINC02055 | 109.9543594 | -1.195997165 | 0.21957846 | 4.28E-09 | 8.14E-08 |
| SPIRE2 | 22.89771148 | -1.195418498 | 0.501680046 | 0.000948289 | 0.005682187 |
| VAV3 | 154.9416577 | 1.192797007 | 0.185550082 | 1.08E-11 | 3.02E-10 |
| TASL | 428.3227605 | 1.192405066 | 0.125991087 | 2.56E-22 | 1.88E-20 |
| LYSMD2 | 72.61013544 | 1.191924359 | 0.253300509 | 1.95E-07 | 2.81E-06 |
| SOAT2 | 16.51089422 | -1.187183833 | 0.645884106 | 0.003010166 | 0.015118053 |
| ATL1 | 28.68761097 | -1.184908397 | 0.447909224 | 0.000481407 | 0.003152999 |
| KLHL3 | 90.4570701 | -1.184045199 | 0.250195563 | 1.64E-07 | 2.40E-06 |
| SNORA63B | 13.72480219 | -1.183987322 | 0.725434128 | 0.004380627 | 0.020619547 |
| LOC124904704 | 11.67116956 | 1.182711281 | 1.22861239 | 0.00746845 | 0.032123981 |
| AEN | 2518.837195 | -1.181390923 | 0.086013668 | 5.84E-44 | 1.25E-41 |
| SYNGR1 | 153.456763 | -1.177224654 | 0.2163625 | 4.67E-09 | 8.86E-08 |
| HVCN1 | 2554.481817 | -1.175995122 | 0.091714697 | 1.18E-38 | 2.10E-36 |
| LOC105374933 | 32.50078852 | -1.173322694 | 0.406064125 | 0.000256938 | 0.001819792 |
| CCDC65 | 39.32236366 | 1.172317814 | 0.36319634 | 8.43E-05 | 0.000682821 |
| ZNF225 | 56.4209952 | 1.171225485 | 0.327735264 | 2.41E-05 | 0.000224537 |
| TLE1 | 527.9842599 | 1.169612413 | 0.132675431 | 1.00E-19 | 6.05E-18 |
| RASA4B | 203.919905 | -1.169363887 | 0.158010772 | 1.19E-14 | 4.68E-13 |
| MOB3B | 7.065698503 | 1.167674385 | 1.379428826 | 0.008979153 | 0.037252513 |
| TSPAN6 | 32.40323192 | 1.167381927 | 0.403481003 | 0.000244556 | 0.001747106 |
| TUSC1 | 37.53320846 | 1.165865766 | 0.366741013 | 9.95E-05 | 0.000791915 |
| PLAAT4 | 1535.771629 | 1.165443624 | 0.082843686 | 5.54E-46 | 1.31E-43 |
| LRRC2 | 27.8227498 | -1.164439167 | 0.471420459 | 0.000788322 | 0.004854686 |
| KCNS3 | 2619.501857 | -1.164269101 | 0.077698734 | 9.41E-52 | 2.63E-49 |
| KDM7A | 644.8251159 | 1.163410064 | 0.13121887 | 6.45E-20 | 3.97E-18 |
| TSC22D1 | 1911.391291 | -1.163130511 | 0.10393255 | 4.09E-30 | 4.82E-28 |
| ITGA6 | 1941.823734 | -1.163112542 | 0.089319058 | 7.51E-40 | 1.37E-37 |
| YWHAEP7 | 52.54108477 | 1.162872816 | 0.300762305 | 8.27E-06 | 8.49E-05 |
| BCL11A | 25.6289495 | 1.160204325 | 0.472023389 | 0.000820196 | 0.005020621 |
| JOSD1 | 4050.025325 | -1.156247233 | 0.099285536 | 2.21E-32 | 2.97E-30 |
| NR3C2 | 21.87129484 | 1.156180404 | 0.55086768 | 0.00184786 | 0.01004375 |
| TM7SF3 | 6551.078733 | -1.154989617 | 0.071841702 | 3.58E-59 | 1.22E-56 |
| GAS2L1 | 49.49587517 | 1.152731742 | 0.326415679 | 3.00E-05 | 0.000273245 |
| LINC01480 | 1404.191455 | -1.152694344 | 0.085963118 | 4.86E-42 | 9.46E-40 |

| | | | | | |
|---|---|---|---|---|---|
| LY6G5C | 11.66097312 | -1.152074204 | 0.830029455 | 0.006030817 | 0.026930288 |
| CXCR3 | 133.6168394 | 1.150463326 | 0.239009126 | 1.16E-07 | 1.76E-06 |
| DYRK2 | 631.1078366 | 1.150159965 | 0.127943577 | 2.10E-20 | 1.36E-18 |
| RNVU1-15 | 13.87683522 | 1.14993668 | 0.789699336 | 0.005277824 | 0.024128201 |
| LOC124901002 | 100.4769694 | -1.146785205 | 0.230710283 | 5.31E-08 | 8.45E-07 |
| CRYBG3 | 656.84303 | 1.145283921 | 0.124913875 | 3.90E-21 | 2.61E-19 |
| LOC390876 | 9.5004034 | 1.139729851 | 0.927217989 | 0.00723834 | 0.031344422 |
| BOLA3-DT | 48.61154743 | -1.137450067 | 0.333591781 | 4.74E-05 | 0.00040954 |
| MT2A | 309.217417 | -1.13627451 | 0.21081949 | 6.07E-09 | 1.12E-07 |
| CCL3 | 15273.16124 | -1.136092137 | 0.101024988 | 1.88E-30 | 2.25E-28 |
| SLC9A3R2 | 296.3425791 | -1.135752756 | 0.160694381 | 1.49E-13 | 5.22E-12 |
| S1PR1-DT | 18.1754605 | 1.134058013 | 0.637966989 | 0.003470114 | 0.017032872 |
| OAS2 | 866.1161874 | 1.133356404 | 0.107016132 | 2.84E-27 | 2.81E-25 |
| ZNF224 | 106.9877674 | 1.132098021 | 0.214364355 | 1.05E-08 | 1.87E-07 |
| EFR3B | 310.9775328 | -1.130135126 | 0.13169004 | 8.47E-19 | 4.67E-17 |
| TLN2 | 41.14798874 | -1.12865392 | 0.371789475 | 0.000174771 | 0.001297814 |
| FAM234B | 209.2711702 | -1.125991364 | 0.15602744 | 4.88E-14 | 1.82E-12 |
| PDZK1 | 15.56755761 | 1.125229328 | 0.621526813 | 0.003369146 | 0.016626815 |
| LOC105377743 | 15.500789 | 1.124522196 | 0.618502311 | 0.003343197 | 0.016527016 |
| KLF2 | 2683.637342 | 1.124409044 | 0.141438333 | 1.66E-16 | 7.79E-15 |
| INKA2 | 171.3506154 | -1.124324315 | 0.170069454 | 3.48E-12 | 1.04E-10 |
| SULF1 | 64.12174169 | -1.122658746 | 0.279211976 | 4.74E-06 | 5.13E-05 |
| GSTO2 | 220.8489974 | -1.122186833 | 0.158898568 | 1.46E-13 | 5.14E-12 |
| FHL2 | 19.08778725 | 1.121330116 | 0.569006271 | 0.002536804 | 0.013105559 |
| SLC2A3 | 1534.42191 | -1.121291316 | 0.087456462 | 1.22E-38 | 2.16E-36 |
| LUCAT1 | 18.23497503 | 1.120805368 | 0.591064548 | 0.002881035 | 0.014592243 |
| GALR2 | 19.93638627 | -1.119859746 | 0.552853353 | 0.00225642 | 0.011898423 |
| LACC1 | 37.84067975 | 1.119707258 | 0.386216416 | 0.000245647 | 0.001753451 |
| PORCN-DT | 6.454392555 | -1.119648374 | 1.814354709 | 0.009877375 | 0.040398324 |
| F8 | 68.98796428 | -1.117978269 | 0.289489417 | 8.56E-06 | 8.76E-05 |
| CYB561 | 328.9821575 | -1.117569611 | 0.144003992 | 8.04E-16 | 3.61E-14 |
| ITGB4 | 16.54158104 | 1.117352286 | 0.600766808 | 0.003121446 | 0.015604527 |
| PRICKLE1 | 107.3080591 | 1.116960245 | 0.211810386 | 1.11E-08 | 1.96E-07 |
| TNFRSF19 | 115.0548781 | -1.115382141 | 0.205252858 | 4.75E-09 | 8.97E-08 |
| NIPAL2 | 170.0231577 | -1.115280655 | 0.188340937 | 2.71E-10 | 6.10E-09 |
| SOWAHD | 51.33443534 | 1.114407357 | 0.34507896 | 8.80E-05 | 0.000709374 |
| LOC105375740 | 10.59069669 | -1.11337664 | 0.998744491 | 0.008015633 | 0.033962982 |
| TUBBP5 | 284.5332944 | 1.112988981 | 0.144596875 | 1.21E-15 | 5.28E-14 |
| NAP1L3 | 31.06877968 | 1.111465793 | 0.413616077 | 0.00047098 | 0.003096413 |
| BCAS1 | 8.941556915 | -1.110927745 | 1.151998618 | 0.008684851 | 0.036222438 |
| DRAM1 | 129.0269701 | -1.10982112 | 0.199356616 | 2.33E-09 | 4.65E-08 |
| AVPI1 | 215.2916685 | -1.109397842 | 0.163650165 | 1.17E-12 | 3.72E-11 |
| CD70 | 4234.569536 | -1.107954135 | 0.106768905 | 3.00E-26 | 2.72E-24 |
| TLR4 | 61.97552922 | 1.106157795 | 0.297797631 | 1.51E-05 | 0.0001477 |
| ZBTB7C | 111.3407431 | 1.105732818 | 0.227388834 | 9.60E-08 | 1.46E-06 |
| ALDH2 | 29.77964658 | 1.105590513 | 0.419731784 | 0.000539388 | 0.003479311 |
| AKAP5 | 20.55124931 | 1.105393391 | 0.556030978 | 0.002400133 | 0.012526554 |
| PCDH18 | 11.57378378 | -1.104742937 | 0.881979759 | 0.007229781 | 0.03132302 |
| BMF | 452.5649303 | 1.104254262 | 0.124618048 | 7.29E-20 | 4.46E-18 |
| MIR22HG | 204.3075569 | -1.104107024 | 0.161551053 | 7.47E-13 | 2.43E-11 |

| | | | | | |
|---|---|---|---|---|---|
| PLXND1 | 119.7808077 | 1.103484708 | 0.210840914 | 1.45E-08 | 2.50E-07 |
| RAB31 | 1047.259457 | 1.102937005 | 0.08982544 | 1.09E-35 | 1.75E-33 |
| SNORD81 | 68.98222011 | -1.101102794 | 0.265454768 | 2.83E-06 | 3.21E-05 |
| LOC124904652 | 26.38475018 | -1.098239354 | 0.466382375 | 0.001121017 | 0.006560545 |
| RXRA | 520.0272881 | 1.09535295 | 0.127092217 | 6.45E-19 | 3.59E-17 |
| CD48 | 10317.80525 | -1.094527021 | 0.066070948 | 1.21E-62 | 4.55E-60 |
| PRSS2 | 39.24838674 | 1.094148569 | 0.360110801 | 0.000169434 | 0.001263604 |
| CSRNP1 | 545.3315559 | -1.08891516 | 0.121719085 | 3.78E-20 | 2.41E-18 |
| HOPX | 46.84611082 | 1.088146018 | 0.361065279 | 0.000183359 | 0.001354616 |
| GLA | 2467.194894 | -1.087780948 | 0.078706782 | 2.08E-44 | 4.49E-42 |
| ASRGL1 | 203.8155128 | 1.085936071 | 0.163894856 | 3.14E-12 | 9.41E-11 |
| OAS1 | 324.6556779 | 1.085759323 | 0.12802836 | 2.10E-18 | 1.12E-16 |
| FBXL22 | 6.6808172 | -1.085157319 | 1.635925803 | 0.010583422 | 0.042691151 |
| KREMEN2 | 76.8630154 | -1.082939844 | 0.304226993 | 2.89E-05 | 0.000263966 |
| TEX19 | 389.6293315 | -1.081130921 | 0.129892017 | 8.62E-18 | 4.39E-16 |
| TP63 | 2107.9023 | 1.080350764 | 0.103877728 | 2.27E-26 | 2.07E-24 |
| CD38 | 2550.970649 | 1.079664297 | 0.076149017 | 1.19E-46 | 2.95E-44 |
| IL15RA | 155.814533 | 1.078521217 | 0.18856486 | 9.45E-10 | 1.99E-08 |
| CCDC141 | 330.6758326 | 1.077406261 | 0.132218629 | 3.50E-17 | 1.70E-15 |
| SPEG | 10.83893143 | 1.076001825 | 0.840986147 | 0.007347798 | 0.031683719 |
| CCL5 | 3132.539705 | -1.075531986 | 0.092937905 | 5.67E-32 | 7.43E-30 |
| EVPLL | 8.37756145 | -1.073973466 | 1.175009239 | 0.009780719 | 0.040088217 |
| CLCN2 | 112.6720126 | -1.073278759 | 0.251754405 | 1.76E-06 | 2.11E-05 |
| LAMP3 | 150.8264532 | 1.072228361 | 0.187853547 | 9.94E-10 | 2.08E-08 |
| SEC31A | 4574.093427 | -1.072066308 | 0.066907879 | 8.65E-59 | 2.88E-56 |
| LOC124904057 | 16.7078952 | 1.071936124 | 0.605691917 | 0.003860497 | 0.018559751 |
| ZMAT3 | 876.3145282 | -1.070105449 | 0.155384828 | 5.13E-13 | 1.70E-11 |
| RASA4 | 218.1046747 | -1.070049211 | 0.154353939 | 3.98E-13 | 1.33E-11 |
| ST3GAL1 | 1313.714289 | 1.070034129 | 0.087695854 | 2.90E-35 | 4.30E-33 |
| JCHAIN | 190423.9986 | -1.069981314 | 0.101909139 | 5.63E-27 | 5.47E-25 |
| PSMB9 | 2016.745803 | 1.069836388 | 0.096664221 | 1.73E-29 | 1.91E-27 |
| PDE4B | 1334.291498 | -1.068513247 | 0.104447525 | 1.40E-25 | 1.21E-23 |
| CD109 | 28.54960763 | -1.066893904 | 0.429053185 | 0.000844395 | 0.005136072 |
| USP36 | 2119.945159 | -1.065392285 | 0.095217246 | 4.60E-30 | 5.38E-28 |
| TFAP2A-AS1 | 28.91702464 | 1.062982229 | 0.4229021 | 0.000795645 | 0.004888372 |
| JAKMIP1 | 222.4537433 | 1.062547574 | 0.166632882 | 1.64E-11 | 4.47E-10 |
| PRSS21 | 809.6975594 | -1.062190235 | 0.113449181 | 8.07E-22 | 5.75E-20 |
| TMEM63C | 174.0026333 | -1.062025141 | 0.183198271 | 6.14E-10 | 1.33E-08 |
| TG | 21.2366276 | -1.061226382 | 0.566951328 | 0.003215566 | 0.016014944 |
| PFN2 | 153.2152551 | -1.058772684 | 0.191435942 | 2.82E-09 | 5.53E-08 |
| WWC3 | 1340.988874 | 1.057566785 | 0.087024895 | 5.33E-35 | 7.76E-33 |
| ELMOD1 | 18.16893881 | -1.055956885 | 0.590434145 | 0.003939142 | 0.01885931 |
| HES7 | 24.15394579 | -1.055696147 | 0.477404069 | 0.001693605 | 0.009334231 |
| SAT1 | 491.16239 | -1.054936106 | 0.118328076 | 4.97E-20 | 3.10E-18 |
| TRBJ2-1 | 15.58842672 | 1.052923127 | 0.622039933 | 0.004461004 | 0.02094096 |
| HSPA4L | 291.3060768 | -1.052481112 | 0.158239691 | 2.56E-12 | 7.75E-11 |
| IFNG | 37.50008019 | 1.052475747 | 0.367776723 | 0.00029746 | 0.002069555 |
| LAT2 | 877.2059991 | 1.052305689 | 0.098013344 | 6.61E-28 | 6.74E-26 |
| HSP90B1 | 79606.19969 | -1.052183302 | 0.063129545 | 3.12E-63 | 1.20E-60 |
| LAPTM5 | 3248.12911 | -1.052051325 | 0.07331199 | 1.06E-47 | 2.70E-45 |

| | | | | | |
|---|---|---|---|---|---|
| SMAD3 | 432.6662137 | 1.050659373 | 0.117851572 | 4.79E-20 | 2.99E-18 |
| LINC01934 | 16.92686248 | 1.050139453 | 0.61244397 | 0.004199879 | 0.019936761 |
| PDE4D | 291.0178439 | -1.049666198 | 0.160337986 | 5.39E-12 | 1.57E-10 |
| ABCB1 | 352.1159988 | 1.049309265 | 0.141473034 | 1.13E-14 | 4.45E-13 |
| LOC124901018 | 67.15720633 | -1.048831892 | 0.266498149 | 7.04E-06 | 7.32E-05 |
| SERINC5 | 221.6268388 | -1.048562532 | 0.154646643 | 1.14E-12 | 3.63E-11 |
| FOSB | 338.720203 | -1.047928741 | 0.154404779 | 1.13E-12 | 3.60E-11 |
| RIMS3 | 472.4965475 | 1.047691037 | 0.121550916 | 6.44E-19 | 3.59E-17 |
| APOBEC3F | 485.9612529 | 1.046862372 | 0.11338191 | 2.63E-21 | 1.80E-19 |
| TMEM229B | 16.61929302 | 1.04604965 | 0.689858957 | 0.005674099 | 0.025581456 |
| AVIL | 52.81280486 | 1.045305893 | 0.321229652 | 8.59E-05 | 0.000694398 |
| MPPED2 | 110.0728042 | 1.043666157 | 0.24023511 | 1.17E-06 | 1.44E-05 |
| LRRC8A | 3451.069071 | 1.043042209 | 0.089265485 | 1.49E-32 | 2.01E-30 |
| SH2D2A | 371.1390761 | -1.040811711 | 0.136947589 | 2.90E-15 | 1.21E-13 |
| MKNK2 | 2425.251899 | -1.039053935 | 0.106299267 | 1.39E-23 | 1.12E-21 |
| NREP | 240.1613392 | 1.038537148 | 0.144820463 | 7.07E-14 | 2.56E-12 |
| XPC | 660.2690065 | -1.036226137 | 0.104769178 | 4.78E-24 | 3.90E-22 |
| ZMYM1 | 25.35549844 | 1.03572584 | 0.460039623 | 0.001524495 | 0.008521521 |
| USP2 | 219.236297 | -1.035381696 | 0.156950097 | 4.22E-12 | 1.24E-10 |
| LDHD | 36.18443124 | -1.034158339 | 0.39250311 | 0.000616917 | 0.003926185 |
| ASIC1 | 201.160905 | -1.033589179 | 0.188028352 | 3.57E-09 | 6.88E-08 |
| PGM5 | 6.246866068 | -1.033471795 | 3.545691624 | 0.01078157 | 0.043288767 |
| LOC105379364 | 40.83256644 | -1.031096042 | 0.357535966 | 0.000291496 | 0.002032137 |
| PLCXD3 | 14.6970384 | 1.029859167 | 0.769204812 | 0.006983897 | 0.030402418 |
| TRIB1 | 3880.8978 | -1.025678759 | 0.075284058 | 2.83E-43 | 5.84E-41 |
| ZNF568 | 18.98931829 | -1.025652568 | 0.555752519 | 0.003576015 | 0.017453734 |
| LAIR1 | 998.9508972 | 1.023502171 | 0.106465783 | 6.81E-23 | 5.22E-21 |
| ITPRIPL2 | 61.09270263 | -1.022588314 | 0.294263915 | 4.41E-05 | 0.000384198 |
| CD3E | 220.1119091 | -1.022257108 | 0.156959713 | 7.40E-12 | 2.11E-10 |
| MVB12B | 117.0116653 | 1.02217504 | 0.215739592 | 1.93E-07 | 2.78E-06 |
| SOCS1 | 22.89942209 | 1.018382668 | 0.541872332 | 0.00331441 | 0.016399721 |
| EGR2 | 86.0679266 | -1.014538483 | 0.239628386 | 2.07E-06 | 2.44E-05 |
| PRDM13 | 39.68825153 | 1.013299744 | 0.363111222 | 0.00039086 | 0.002629573 |
| OGFRP1 | 16.52958679 | 1.013012688 | 0.639581544 | 0.005493526 | 0.024916551 |
| HIC1 | 133.3515382 | 1.012352601 | 0.279292497 | 2.36E-05 | 0.000220618 |
| FBXL7 | 482.8349682 | 1.010422073 | 0.116270549 | 3.60E-19 | 2.05E-17 |
| FAM229B | 51.92612418 | -1.008963561 | 0.327762706 | 0.000172931 | 0.001286361 |
| PURPL | 201.9413791 | -1.008076188 | 0.161541561 | 4.27E-11 | 1.08E-09 |
| DERL3 | 3053.583848 | -1.007615307 | 0.116549517 | 5.58E-19 | 3.13E-17 |
| DGKG | 72.5371525 | -1.006828835 | 0.281940196 | 3.11E-05 | 0.000281999 |
| TBC1D8 | 49.04036894 | 1.00630364 | 0.326385444 | 0.000158077 | 0.001187126 |
| SAMD3 | 18.90107875 | 1.005850864 | 0.650001312 | 0.005685813 | 0.0256076 |
| FAS | 688.5581706 | -1.004935281 | 0.120086915 | 5.70E-18 | 2.94E-16 |
| HOXB4 | 184.4314335 | 1.004690732 | 0.165518857 | 1.26E-10 | 2.96E-09 |
| IKZF1 | 1501.076758 | 1.003863706 | 0.083888109 | 5.20E-34 | 7.38E-32 |
| LCP2 | 300.6267366 | 1.00190862 | 0.150380256 | 2.59E-12 | 7.82E-11 |
| FCMR | 553.8250973 | 1.001735236 | 0.113907834 | 1.43E-19 | 8.49E-18 |
| NQO2 | 131.8053256 | 1.001034795 | 0.20015058 | 5.47E-08 | 8.67E-07 |
| BTN3A2 | 2493.606016 | 0.998985147 | 0.073607031 | 5.79E-43 | 1.18E-40 |
| FEZ1 | 109.4666467 | 0.99727658 | 0.214099029 | 2.90E-07 | 4.08E-06 |

| | | | | | |
|---|---|---|---|---|---|
| MXD1 | 1467.018644 | -0.996807411 | 0.097875028 | 2.38E-25 | 2.04E-23 |
| ZNF432 | 50.11509425 | -0.995291315 | 0.322728098 | 0.000161648 | 0.001210751 |
| HSP90B3P | 247.3794989 | -0.995136938 | 0.150582425 | 3.73E-12 | 1.10E-10 |
| ABTB1 | 235.1594426 | 0.995087109 | 0.152000271 | 5.84E-12 | 1.69E-10 |
| FLT3 | 34.2401546 | 0.994771468 | 0.434364964 | 0.001450642 | 0.008161394 |
| SEPTIN3 | 520.2027013 | -0.994711658 | 0.113765898 | 2.35E-19 | 1.37E-17 |
| BCAT1 | 1689.434002 | 0.994621696 | 0.304247737 | 8.40E-05 | 0.000681089 |
| KIAA0040 | 831.9772117 | 0.992156733 | 0.094687425 | 1.13E-26 | 1.06E-24 |
| KDM6B | 252.7760681 | -0.992005679 | 0.14792796 | 2.14E-12 | 6.52E-11 |
| FGFR1 | 534.0136872 | -0.991203822 | 0.121820289 | 4.20E-17 | 2.03E-15 |
| GADD45B | 983.7292276 | -0.989506501 | 0.104108663 | 2.21E-22 | 1.62E-20 |
| HSPA1B | 2027.499827 | -0.987031042 | 0.121548126 | 4.91E-17 | 2.36E-15 |
| SNHG20 | 35.61068516 | 0.985621865 | 0.37847756 | 0.000685592 | 0.004296923 |
| SULT1A1 | 356.4559536 | -0.984856555 | 0.136556599 | 5.81E-14 | 2.13E-12 |
| TCF4 | 4862.035065 | 0.984712743 | 0.102677788 | 9.01E-23 | 6.84E-21 |
| AK8 | 18.0494313 | 0.984051059 | 0.853221519 | 0.008374804 | 0.035192059 |
| CTSH | 219.8646018 | 0.98370502 | 0.155823312 | 2.77E-11 | 7.21E-10 |
| QPRT | 688.943642 | -0.980109125 | 0.119725486 | 2.97E-17 | 1.46E-15 |
| AKAP6 | 124.4946242 | -0.977713381 | 0.211036781 | 3.38E-07 | 4.69E-06 |
| RUNX1 | 341.5170047 | 0.977472564 | 0.159949882 | 9.74E-11 | 2.33E-09 |
| DNAJB4 | 190.9937856 | -0.976888888 | 0.181537274 | 7.12E-09 | 1.29E-07 |
| ZFP14 | 65.6036324 | 0.976594263 | 0.276704126 | 3.59E-05 | 0.000320718 |
| CPS1 | 18.40859178 | -0.974356126 | 0.664134803 | 0.006788724 | 0.029709461 |
| WNT10B | 990.0130503 | -0.974215257 | 0.094418837 | 6.62E-26 | 5.82E-24 |
| MBOAT1 | 225.0737405 | 0.97324838 | 0.153733225 | 2.45E-11 | 6.44E-10 |
| HSPA1A | 1286.051071 | -0.972569263 | 0.149391654 | 7.83E-12 | 2.23E-10 |
| LOC101929322 | 24.7722714 | 0.972382822 | 0.522361582 | 0.003613388 | 0.017596469 |
| DUSP16 | 1903.883084 | -0.971739807 | 0.079054826 | 1.10E-35 | 1.76E-33 |
| REPS2 | 41.6171579 | 0.969609806 | 0.368283179 | 0.000637245 | 0.0040348 |
| FGF9 | 15.64273856 | 0.969404579 | 0.763730175 | 0.008028767 | 0.034010312 |
| PRKCA | 1020.118408 | -0.969399185 | 0.101414894 | 1.24E-22 | 9.24E-21 |
| AIM2 | 1452.961777 | 0.96864557 | 0.096608821 | 1.23E-24 | 1.03E-22 |
| SLCO3A1 | 813.8634909 | 0.967533787 | 0.101583216 | 1.81E-22 | 1.34E-20 |
| CPED1 | 27.61059672 | 0.966623853 | 0.479752377 | 0.002708785 | 0.013856492 |
| CASKIN1 | 13.73080036 | -0.965797032 | 0.691447307 | 0.007594895 | 0.032538546 |
| TNNT1 | 184.660082 | -0.963131094 | 0.172806264 | 2.65E-09 | 5.22E-08 |
| BOLA2B | 1002.221392 | 0.962822557 | 0.102890684 | 8.72E-22 | 6.19E-20 |
| HOMER1 | 569.3616997 | -0.962762045 | 0.160557977 | 1.99E-10 | 4.54E-09 |
| LOC105369147 | 27.82102202 | -0.959228131 | 0.455513404 | 0.002317072 | 0.012162699 |
| SMARCA2 | 1364.886911 | 0.9581086 | 0.094351763 | 3.33E-25 | 2.84E-23 |
| YPEL1 | 78.43616469 | 0.956258394 | 0.24292889 | 7.76E-06 | 8.01E-05 |
| PLPP1 | 153.3125739 | 0.95521212 | 0.201470304 | 2.08E-07 | 2.98E-06 |
| LOC107986196 | 60.67585911 | -0.954277576 | 0.32050584 | 0.000232263 | 0.001671004 |
| ADAMTS10 | 90.74646023 | 0.954261141 | 0.232363416 | 3.76E-06 | 4.15E-05 |
| AGO4 | 353.5556997 | 0.953393375 | 0.12866812 | 1.32E-14 | 5.11E-13 |
| EZR | 17961.87009 | -0.953223018 | 0.083335404 | 3.22E-31 | 4.01E-29 |
| PIK3CG | 4818.165801 | -0.95299524 | 0.086843512 | 5.54E-29 | 5.89E-27 |
| DDIT4 | 4094.565481 | -0.952586571 | 0.099494809 | 1.14E-22 | 8.54E-21 |
| P2RX7 | 754.2488814 | 0.952245276 | 0.09850541 | 4.45E-23 | 3.47E-21 |
| ALOX5AP | 3886.425623 | 0.951267219 | 0.077244882 | 7.71E-36 | 1.25E-33 |

| | | | | | |
|---|---|---|---|---|---|
| LOC105373428 | 346.8976757 | -0.950390603 | 0.126654706 | 6.67E-15 | 2.68E-13 |
| ZNF227 | 270.956139 | 0.948909722 | 0.147216143 | 1.20E-11 | 3.32E-10 |
| COL27A1 | 15.59694599 | -0.948756553 | 0.62350831 | 0.006708577 | 0.029396565 |
| SLC2A6 | 278.3563827 | 0.948232289 | 0.178932798 | 1.18E-08 | 2.08E-07 |
| ZNF215 | 133.617298 | -0.946170842 | 0.209498505 | 6.42E-07 | 8.42E-06 |
| KSR1 | 25.85675934 | 0.945078789 | 0.505958004 | 0.003692841 | 0.017872813 |
| LOC105371773 | 74.49201584 | 0.944816385 | 0.251477484 | 1.60E-05 | 0.000155232 |
| IZUMO4 | 108.3229074 | -0.944015139 | 0.24058557 | 8.56E-06 | 8.76E-05 |
| SOCS3 | 103.4270306 | -0.943482409 | 0.216504501 | 1.32E-06 | 1.62E-05 |
| TFCP2 | 770.9467579 | 0.940904678 | 0.10505787 | 3.56E-20 | 2.29E-18 |
| MLLT3 | 627.3225158 | 0.940782881 | 0.115119582 | 3.24E-17 | 1.58E-15 |
| L3MBTL3 | 25.11411146 | 0.940430906 | 0.502091666 | 0.003682953 | 0.017839913 |
| PLEC | 1694.150557 | 0.939904553 | 0.133393768 | 1.93E-13 | 6.67E-12 |
| PTPRG | 128.8999694 | 0.93911367 | 0.219831037 | 1.88E-06 | 2.24E-05 |
| ELK3 | 1009.756867 | 0.938734231 | 0.103896936 | 1.69E-20 | 1.10E-18 |
| ENC1 | 306.6149395 | -0.938358054 | 0.147119612 | 2.02E-11 | 5.40E-10 |
| NAV2 | 22.25252627 | 0.936985757 | 0.498932713 | 0.003744376 | 0.018066693 |
| MPP2 | 156.5564412 | 0.936918515 | 0.176618458 | 1.19E-08 | 2.08E-07 |
| DDN | 609.6509557 | -0.936136963 | 0.10921197 | 1.18E-18 | 6.40E-17 |
| ARL4C | 5199.932557 | 0.936102265 | 0.066776105 | 1.34E-45 | 3.09E-43 |
| FGD3 | 230.0563666 | 0.935964268 | 0.16451944 | 1.33E-09 | 2.75E-08 |
| DOCK8 | 706.399754 | -0.93579402 | 0.108084651 | 5.39E-19 | 3.03E-17 |
| ITPKC | 136.1495368 | -0.931990438 | 0.204970758 | 5.65E-07 | 7.48E-06 |
| IRAG2 | 3721.5817 | 0.930685511 | 0.078848704 | 4.20E-33 | 5.77E-31 |
| CBLB | 368.6750503 | 0.928152128 | 0.121581694 | 2.50E-15 | 1.05E-13 |
| SH2D5 | 61.7600699 | -0.927897044 | 0.30476302 | 0.000207231 | 0.001507189 |
| IL1RAP | 333.9044143 | 0.927061484 | 0.148162491 | 4.09E-11 | 1.04E-09 |
| TMC5 | 10.55716173 | -0.926256531 | 1.102913224 | 0.01145524 | 0.045466513 |
| FCRL5 | 696.0956575 | 0.926041883 | 0.106627848 | 4.25E-19 | 2.41E-17 |
| APBA1 | 21.77595203 | -0.924175257 | 0.54418528 | 0.005092829 | 0.02343053 |
| CLDN12 | 364.2813412 | -0.92359376 | 0.1623407 | 1.31E-09 | 2.70E-08 |
| JAM3 | 145.418882 | 0.919949848 | 0.189514704 | 1.27E-07 | 1.90E-06 |
| FNBP1L | 153.6293528 | 0.919849528 | 0.176680942 | 2.01E-08 | 3.42E-07 |
| BCL2L11 | 7450.204258 | -0.919122829 | 0.079249833 | 4.83E-32 | 6.38E-30 |
| CR2 | 49.90572684 | -0.918369315 | 0.331139407 | 0.000479562 | 0.003144479 |
| ZNF550 | 90.84176344 | 0.917445839 | 0.240104325 | 1.28E-05 | 0.000126884 |
| FBXO27 | 26.91798586 | 0.915716295 | 0.449068095 | 0.002873221 | 0.014565056 |
| HIPK2 | 1353.063331 | -0.915119967 | 0.098367356 | 1.48E-21 | 1.03E-19 |
| KCNQ1 | 532.9038214 | 0.914897419 | 0.138128732 | 3.89E-12 | 1.14E-10 |
| CAV2 | 26.1994115 | 0.913488455 | 0.52350383 | 0.004720336 | 0.022009171 |
| LOC124907740 | 12.58006916 | 0.912646685 | 0.944337149 | 0.010788939 | 0.043298276 |
| ANKRD13B | 137.6737093 | -0.911275731 | 0.199684227 | 5.37E-07 | 7.16E-06 |
| FBXO6 | 164.0591924 | 0.910473884 | 0.208087004 | 1.24E-06 | 1.53E-05 |
| BTG4 | 71.98593036 | -0.910168198 | 0.292730651 | 0.000175208 | 0.001299391 |
| TMEM54 | 171.5083215 | 0.908570285 | 0.202531107 | 7.55E-07 | 9.76E-06 |
| AGRN | 381.757639 | -0.907712189 | 0.162429048 | 2.54E-09 | 5.01E-08 |
| CYRIA | 44.844334 | 0.906217896 | 0.364636528 | 0.001019087 | 0.006037473 |
| SLC30A4 | 38.10382079 | 0.906198571 | 0.353515616 | 0.000858714 | 0.005212178 |
| TUBB8B | 44.63851111 | 0.905292311 | 0.326987964 | 0.000484744 | 0.003171256 |
| LOC100288073 | 4262.472444 | -0.904323535 | 0.073972746 | 2.65E-35 | 4.02E-33 |

| | | | | | |
|---|---|---|---|---|---|
| TMEM74 | 22.25343622 | 0.903526208 | 0.516645137 | 0.004874957 | 0.022614421 |
| LINC01801 | 15.03209117 | 0.902833594 | 0.665301906 | 0.008471695 | 0.035513032 |
| MAML2 | 53.44804459 | 0.902498148 | 0.299736794 | 0.000237467 | 0.001702536 |
| SESN1 | 1073.887433 | -0.901954653 | 0.094096298 | 1.10E-22 | 8.26E-21 |
| SLC16A14 | 309.1076173 | -0.900013581 | 0.139396083 | 1.23E-11 | 3.40E-10 |
| H2BC12 | 1262.413359 | -0.899029512 | 0.104509403 | 8.73E-19 | 4.78E-17 |
| MYCL | 38.94339069 | 0.898969457 | 0.370556949 | 0.001220509 | 0.007031125 |
| ZNF792 | 71.10156803 | 0.897823848 | 0.255635065 | 4.36E-05 | 0.000380058 |
| RASAL1 | 49.72739801 | -0.897417985 | 0.308284021 | 0.000332004 | 0.002279664 |
| BOLA2 | 752.0842301 | 0.897294356 | 0.105615043 | 2.32E-18 | 1.23E-16 |
| EPHB6 | 111.2852587 | 0.897043286 | 0.217419029 | 3.73E-06 | 4.13E-05 |
| SNHG26 | 51.7346884 | -0.896090188 | 0.334335369 | 0.0006526 | 0.004119971 |
| DEPDC1 | 591.5288949 | 0.895368693 | 0.142867216 | 4.02E-11 | 1.02E-09 |
| UBC | 58333.65936 | -0.895342754 | 0.065128577 | 8.00E-44 | 1.69E-41 |
| SLC1A1 | 14.55073338 | 0.895271306 | 0.681016747 | 0.008977678 | 0.037252513 |
| IFI30 | 9242.083425 | -0.895146628 | 0.083297436 | 7.33E-28 | 7.42E-26 |
| SNX24 | 94.01293862 | 0.893471971 | 0.244376135 | 2.56E-05 | 0.000236799 |
| SLC2A14 | 119.7875598 | -0.893024542 | 0.214616545 | 3.34E-06 | 3.73E-05 |
| PRKCQ | 94.5499715 | 0.891628438 | 0.250654463 | 3.65E-05 | 0.000325095 |
| UBE2L6 | 1840.520213 | 0.891036147 | 0.078818876 | 1.49E-30 | 1.79E-28 |
| ICA1 | 393.1882792 | -0.890805287 | 0.125686081 | 1.65E-13 | 5.75E-12 |
| NCAM1 | 712.1590273 | -0.890481405 | 0.107519444 | 1.38E-17 | 6.95E-16 |
| ABCG1 | 273.3647945 | 0.889831304 | 0.144367313 | 8.09E-11 | 1.96E-09 |
| LOC105369593 | 177.4692785 | -0.889342155 | 0.166972501 | 1.14E-08 | 2.01E-07 |
| INPP5E | 103.7521171 | 0.889142866 | 0.254435888 | 4.56E-05 | 0.000395689 |
| KIAA0513 | 141.3778006 | -0.888281056 | 0.187657661 | 2.47E-07 | 3.50E-06 |
| CDC14A | 107.5904893 | 0.887258333 | 0.225635659 | 8.76E-06 | 8.94E-05 |
| TNFSF10 | 29.97311655 | 0.887082378 | 0.432950412 | 0.00287582 | 0.014570093 |
| CDH17 | 994.3966146 | 0.88700929 | 0.094291751 | 6.17E-22 | 4.42E-20 |
| TMEM169 | 104.1332034 | -0.886758499 | 0.210630025 | 2.75E-06 | 3.13E-05 |
| ST3GAL6 | 1161.166952 | -0.886364994 | 0.139001922 | 2.07E-11 | 5.50E-10 |
| NUAK2 | 675.375647 | 0.885715146 | 0.119811665 | 1.68E-14 | 6.50E-13 |
| TBC1D2 | 32.75531638 | 0.883075057 | 0.397293343 | 0.002031815 | 0.010872972 |
| CD180 | 398.1628484 | -0.882939028 | 0.125585167 | 2.34E-13 | 8.04E-12 |
| NIBAN1 | 1304.371282 | 0.882874814 | 0.095653615 | 3.16E-21 | 2.14E-19 |
| MEF2D | 4628.966116 | -0.882856172 | 0.072037039 | 1.85E-35 | 2.89E-33 |
| LOC124905143 | 17.70425588 | 0.882289588 | 0.62312652 | 0.007978755 | 0.033831563 |
| MLLT11 | 187.8729356 | -0.88220623 | 0.169264014 | 2.07E-08 | 3.50E-07 |
| CTNNA2 | 200.2991164 | 0.882146697 | 0.155790492 | 1.69E-09 | 3.43E-08 |
| SATB1 | 18.22505185 | 0.880956969 | 0.585316214 | 0.007314272 | 0.031562735 |
| STK17A | 1863.846799 | 0.880733933 | 0.081431557 | 3.45E-28 | 3.58E-26 |
| TTYH3 | 233.9843874 | 0.88008127 | 0.173499189 | 4.40E-08 | 7.14E-07 |
| FAM72D | 553.4875257 | 0.879925189 | 0.128768022 | 9.40E-13 | 3.02E-11 |
| KIAA1671 | 96.38372096 | -0.879325388 | 0.248762134 | 4.00E-05 | 0.000352439 |
| CCND2 | 13433.13858 | -0.878466458 | 0.110786993 | 2.77E-16 | 1.28E-14 |
| RPGRIP1L | 110.9682269 | -0.877512217 | 0.233956214 | 1.90E-05 | 0.000181662 |
| RHEBL1 | 314.7119639 | 0.876911937 | 0.129908602 | 1.75E-12 | 5.45E-11 |
| CALHM2 | 46.33822068 | 0.876525381 | 0.317276496 | 0.000518234 | 0.003369981 |
| RPL37AP5 | 49.73216356 | 0.876507248 | 0.392177613 | 0.001958166 | 0.010550579 |
| ENTPD1 | 634.0556888 | -0.874910147 | 0.108723403 | 1.05E-16 | 4.99E-15 |

| | | | | | |
|---|---|---|---|---|---|
| PXDN | 498.6358122 | 0.874753469 | 0.122909208 | 1.31E-13 | 4.63E-12 |
| PRUNE2 | 52.39864972 | 0.874426633 | 0.305222895 | 0.000384574 | 0.002595348 |
| ESR2 | 16.45888575 | -0.873746483 | 0.716367074 | 0.009904175 | 0.040488811 |
| CHST2 | 281.1717707 | -0.871580128 | 0.161341601 | 7.79E-09 | 1.41E-07 |
| JAKMIP3 | 43.48587845 | -0.870906023 | 0.345705645 | 0.001040765 | 0.006144877 |
| CEACAM1 | 34.86204962 | 0.870448565 | 0.408993052 | 0.002513828 | 0.013029619 |
| UBE2Q2P2 | 26.1499167 | 0.869973802 | 0.44466705 | 0.003616898 | 0.017608611 |
| HBD | 34.30166456 | -0.869854135 | 0.407514174 | 0.002600155 | 0.013372996 |
| APOBEC3D | 466.0670115 | 0.869406934 | 0.111904089 | 9.44E-16 | 4.18E-14 |
| PPP4R4 | 14.12222667 | 0.869277793 | 0.758780142 | 0.010833698 | 0.043447694 |
| LOC105374122 | 14.65816461 | 0.868985799 | 0.713582063 | 0.010150191 | 0.041289841 |
| KCTD12 | 1341.857572 | -0.867843826 | 0.125876067 | 6.45E-13 | 2.11E-11 |
| PIEZO1 | 1643.381713 | -0.8667028 | 0.111613371 | 9.89E-16 | 4.37E-14 |
| JADE2 | 289.1773028 | 0.866460935 | 0.168018741 | 2.87E-08 | 4.76E-07 |
| ARAP2 | 188.6389225 | 0.865995703 | 0.183668296 | 2.62E-07 | 3.72E-06 |
| TMEM119 | 28.51616266 | 0.8659849 | 0.439004713 | 0.003528425 | 0.017260379 |
| MIR5047 | 58.30534627 | -0.865486853 | 0.295892444 | 0.000335054 | 0.00229787 |
| SEZ6L2 | 651.6639941 | -0.865386419 | 0.131918018 | 6.50E-12 | 1.86E-10 |
| MAP2K6 | 322.9547126 | 0.863210729 | 0.132266784 | 7.91E-12 | 2.25E-10 |
| LOC105379549 | 12.7398634 | 0.862515659 | 0.829189887 | 0.011654592 | 0.046011936 |
| LARP1B | 1043.380894 | -0.862264162 | 0.092066785 | 9.48E-22 | 6.67E-20 |
| L1CAM | 91.06024731 | 0.862089602 | 0.23598442 | 2.71E-05 | 0.000249363 |
| SLC43A2 | 877.0100825 | -0.862034284 | 0.111317489 | 1.21E-15 | 5.28E-14 |
| KLKB1 | 23.49313946 | 0.861477064 | 0.533177032 | 0.00641897 | 0.028349224 |
| GPR153 | 30.91166905 | -0.860672151 | 0.433904038 | 0.003643798 | 0.017709696 |
| SEMA3C | 205.4659407 | 0.860513699 | 0.214558181 | 6.32E-06 | 6.64E-05 |
| HSPA5P1 | 2111.764589 | -0.860236354 | 0.083596392 | 9.52E-26 | 8.28E-24 |
| ZBED8 | 31.78858677 | 0.859631907 | 0.404823955 | 0.00266665 | 0.013670331 |
| ZFP36L1 | 487.8059889 | -0.857598364 | 0.135599829 | 2.99E-11 | 7.77E-10 |
| CILK1 | 426.0919667 | 0.85733697 | 0.142193283 | 1.90E-10 | 4.35E-09 |
| TET1 | 306.674584 | 0.857239368 | 0.150702694 | 1.47E-09 | 3.00E-08 |
| COX18 | 254.4943795 | -0.856808621 | 0.14634013 | 5.81E-10 | 1.26E-08 |
| C17orf67 | 27.73371329 | 0.85678829 | 0.425734729 | 0.003340221 | 0.016521736 |
| VPS9D1-AS1 | 126.970486 | -0.856549523 | 0.209208601 | 4.89E-06 | 5.28E-05 |
| LOC388242 | 11.16580471 | 0.856239238 | 0.837355789 | 0.011918988 | 0.046792365 |
| LOC613038 | 11.16580471 | 0.856239238 | 0.837355789 | 0.011918988 | 0.046792365 |
| SULT1A2 | 69.0045149 | -0.854770153 | 0.266898326 | 0.000139724 | 0.001064195 |
| APOM | 49.36609969 | -0.854414721 | 0.321357598 | 0.000739069 | 0.004595628 |
| SEMA7A | 716.1588932 | -0.853827961 | 0.097601687 | 2.78E-19 | 1.62E-17 |
| PLEKHA7 | 380.8705516 | -0.853766476 | 0.128323358 | 3.54E-12 | 1.05E-10 |
| GRM3 | 251.1091637 | 0.851400043 | 0.16598438 | 3.37E-08 | 5.55E-07 |
| PTPRO | 40.25533406 | 0.851048562 | 0.344067807 | 0.00117523 | 0.006817909 |
| CRACR2B | 146.9025449 | -0.850192903 | 0.194459982 | 1.45E-06 | 1.76E-05 |
| SESN3 | 69.2978303 | 0.849590938 | 0.285877395 | 0.00028114 | 0.001966279 |
| CHST14 | 219.3331175 | 0.849368834 | 0.157377955 | 8.14E-09 | 1.47E-07 |
| LOC105371480 | 12.21604721 | 0.847700987 | 0.78902169 | 0.011618296 | 0.045892799 |
| RRM2P3 | 507.800487 | -0.847511138 | 0.112371652 | 5.67E-15 | 2.29E-13 |
| IRF2BPL | 141.4446559 | 0.846737017 | 0.182904813 | 4.18E-07 | 5.67E-06 |
| CHST4 | 263.0945103 | -0.845788355 | 0.138509405 | 1.28E-10 | 3.00E-09 |
| DNAAF4 | 18.19771128 | -0.845689396 | 0.619281338 | 0.009115581 | 0.03771912 |

| | | | | | |
|---|---|---|---|---|---|
| SNHG8 | 1355.431106 | -0.844884337 | 0.099852165 | 3.52E-18 | 1.84E-16 |
| SLC39A14 | 8968.505082 | -0.844015608 | 0.067040648 | 3.19E-37 | 5.36E-35 |
| SNORD29 | 12.91917151 | -0.843145581 | 0.813330767 | 0.012006933 | 0.047073593 |
| LOC124900608 | 10.20720136 | 0.84141951 | 1.043756253 | 0.012577375 | 0.04895536 |
| DISP2 | 27.31743101 | -0.840584757 | 0.453574248 | 0.004846129 | 0.022504806 |
| PSRC1 | 72.43748177 | 0.84025715 | 0.29441502 | 0.000419451 | 0.002798087 |
| SEC24D | 3962.117794 | -0.839640156 | 0.067906555 | 5.23E-36 | 8.55E-34 |
| NUDT6 | 83.6985453 | -0.83942522 | 0.24197444 | 5.62E-05 | 0.000477811 |
| FAM72A | 350.1314812 | 0.838943912 | 0.140034246 | 2.50E-10 | 5.63E-09 |
| DYRK3 | 151.9994019 | 0.838714471 | 0.194586079 | 1.90E-06 | 2.25E-05 |
| SPRR2A | 448.0393016 | 0.837727287 | 0.123466727 | 1.43E-12 | 4.48E-11 |
| CCR7 | 28.30729896 | -0.837008282 | 0.543446963 | 0.007875447 | 0.033491723 |
| VPS35L | 969.2771373 | -0.836191062 | 0.089172781 | 9.11E-22 | 6.44E-20 |
| USO1 | 2896.183654 | -0.835634592 | 0.091475867 | 8.35E-21 | 5.48E-19 |
| PVT1 | 340.5239354 | -0.835443881 | 0.138423027 | 2.06E-10 | 4.68E-09 |
| LOC124902966 | 9.119244793 | 0.835012094 | 1.224476695 | 0.012847175 | 0.049759457 |
| HSPA5 | 124298.6526 | -0.834044504 | 0.061489393 | 8.84E-43 | 1.78E-40 |
| SLC25A35 | 28.42813768 | 0.833567872 | 0.471803057 | 0.005445033 | 0.024741969 |
| ABCA1 | 65.43089975 | 0.833215078 | 0.280029301 | 0.000290824 | 0.002028269 |
| SEL1L3 | 429.1247035 | 0.832923854 | 0.142625479 | 6.53E-10 | 1.41E-08 |
| CPEB3 | 179.1233811 | 0.832178607 | 0.200917713 | 3.83E-06 | 4.22E-05 |
| ZNF724 | 62.3324107 | 0.83156277 | 0.277046831 | 0.000272236 | 0.001913277 |
| CALHM1 | 419.3644038 | 0.831382206 | 0.124385448 | 2.99E-12 | 8.98E-11 |
| SQSTM1 | 60981.00799 | -0.831196598 | 0.098550464 | 9.84E-19 | 5.37E-17 |
| PNRC1 | 1865.019743 | -0.83080935 | 0.094563157 | 2.00E-19 | 1.18E-17 |
| PRKCG | 48.64646947 | 0.830312683 | 0.311846345 | 0.000745563 | 0.004629224 |
| CSF2RB | 9.709319406 | -0.829681804 | 1.164849472 | 0.01330944 | 0.051388979 |
| YOD1 | 468.1020717 | -0.828858538 | 0.197707158 | 3.12E-06 | 3.52E-05 |
| FAM110A | 127.051971 | 0.828193676 | 0.246571344 | 8.37E-05 | 0.000678871 |
| SPG7 | 852.3277537 | -0.827396991 | 0.108853682 | 3.97E-15 | 1.64E-13 |
| APOL1 | 158.0103102 | 0.82639325 | 0.186830332 | 1.13E-06 | 1.41E-05 |
| MAST3 | 219.2124321 | 0.825010097 | 0.162458008 | 4.62E-08 | 7.44E-07 |
| LMF1 | 86.1910172 | -0.824773296 | 0.276860843 | 0.000301447 | 0.002093284 |
| LOC124901083 | 367.2898211 | -0.82403728 | 0.129932575 | 2.82E-11 | 7.33E-10 |
| ACSL1 | 506.2560261 | 0.823588744 | 0.107413063 | 2.28E-15 | 9.60E-14 |
| FBXO16 | 53.77195885 | -0.82350063 | 0.314667284 | 0.00088124 | 0.005324436 |
| SAMD9L | 1790.727664 | 0.823471815 | 0.099455406 | 1.55E-17 | 7.79E-16 |
| BRD2 | 9571.690836 | -0.823212474 | 0.066552424 | 5.13E-36 | 8.45E-34 |
| ZNF362 | 116.1528193 | 0.822831188 | 0.249902824 | 0.000104167 | 0.00082519 |
| PDCD1 | 30.30863952 | -0.822344794 | 0.444265093 | 0.004974346 | 0.022976995 |
| CDC25A | 2666.605065 | -0.821234704 | 0.071283032 | 1.41E-31 | 1.80E-29 |
| CRYBG1 | 367.9953897 | 0.819817084 | 0.121442849 | 1.89E-12 | 5.84E-11 |
| FAM78B | 247.5106596 | -0.818884019 | 0.152676067 | 1.03E-08 | 1.85E-07 |
| DOK3 | 260.024611 | -0.818767515 | 0.16360586 | 7.01E-08 | 1.09E-06 |
| SNHG4 | 147.8649985 | -0.81853555 | 0.184383081 | 1.11E-06 | 1.38E-05 |
| MAGEH1 | 960.3219504 | -0.817436434 | 0.09652219 | 3.25E-18 | 1.70E-16 |
| RENBP | 492.3575823 | -0.815630303 | 0.110918774 | 2.58E-14 | 9.86E-13 |
| HEY2 | 32.19460916 | 0.814954641 | 0.401671695 | 0.003522407 | 0.017235805 |
| DAPP1 | 962.7759118 | -0.814387034 | 0.112557648 | 6.08E-14 | 2.22E-12 |
| LDLRAD4 | 316.5129195 | 0.814097235 | 0.14949917 | 6.51E-09 | 1.19E-07 |

| | | | | | |
|---|---|---|---|---|---|
| BANK1 | 15.2719155 | -0.8140186 | 0.663927149 | 0.011508556 | 0.045636285 |
| HOXC6 | 27.5914215 | 0.812344697 | 0.462389002 | 0.005801911 | 0.026042428 |
| FAM72C | 405.5791291 | 0.812083463 | 0.128686811 | 3.50E-11 | 9.00E-10 |
| GSN | 86.56508402 | 0.810974397 | 0.235599191 | 6.44E-05 | 0.000539967 |
| BDH2 | 455.6560844 | -0.80903571 | 0.117722324 | 8.51E-13 | 2.75E-11 |
| ACVR1B | 499.6136221 | 0.808471717 | 0.119829768 | 2.01E-12 | 6.17E-11 |
| LMO7 | 233.5295686 | 0.807969906 | 0.165154893 | 1.22E-07 | 1.84E-06 |
| SERTAD1 | 571.4124669 | -0.807854221 | 0.142181689 | 1.77E-09 | 3.57E-08 |
| SORBS3 | 197.6688946 | -0.807816204 | 0.163748112 | 1.03E-07 | 1.56E-06 |
| SMAGP | 613.6650866 | 0.807779864 | 0.105662332 | 2.79E-15 | 1.17E-13 |
| ZSCAN31 | 21.24877118 | -0.807512753 | 0.729906035 | 0.011548652 | 0.045721995 |
| RSAD2 | 181.1439592 | 0.807417413 | 0.174588395 | 4.66E-07 | 6.26E-06 |
| TMSB4XP2 | 70.70805998 | 0.807198865 | 0.28058698 | 0.000418651 | 0.002794828 |
| MFSD3 | 77.11275645 | -0.807071009 | 0.279096345 | 0.000414284 | 0.002768874 |
| HOXB3 | 28.22727152 | 0.806243661 | 0.440590547 | 0.005195465 | 0.023845709 |
| TIGAR | 1384.770587 | -0.805954618 | 0.100829029 | 1.81E-16 | 8.43E-15 |
| TPRN | 372.119485 | 0.805164787 | 0.143381578 | 2.53E-09 | 5.01E-08 |
| SH3YL1 | 164.5879532 | -0.805132713 | 0.181667744 | 1.13E-06 | 1.41E-05 |
| KIF20A | 1089.372702 | 0.803767293 | 0.098073634 | 3.39E-17 | 1.65E-15 |
| EPB41L4B | 26.09698673 | -0.803218275 | 0.566148055 | 0.009835013 | 0.040272622 |
| STOX1 | 119.1346504 | 0.802912068 | 0.211250512 | 1.67E-05 | 0.000161396 |
| ID3 | 2067.196992 | -0.801660902 | 0.117439053 | 1.20E-12 | 3.79E-11 |
| C3orf80 | 31.30690201 | 0.801324258 | 0.451764219 | 0.005621317 | 0.025403015 |
| SDCBP2 | 21.73181931 | -0.801034443 | 0.540312925 | 0.008953211 | 0.037176718 |
| H1-10-AS1 | 23.43475639 | -0.800839884 | 0.478955527 | 0.007026577 | 0.030556806 |
| MIR3652 | 641.8950294 | -0.800720474 | 0.118417984 | 1.90E-12 | 5.85E-11 |
| SNORA4 | 24.00837052 | -0.800573072 | 0.517402306 | 0.008153285 | 0.034445092 |
| SYT13 | 82.87969039 | 0.800411177 | 0.240351655 | 9.85E-05 | 0.000785623 |
| OSTC | 6749.445066 | -0.800051369 | 0.077340658 | 6.17E-26 | 5.45E-24 |
| ZBTB42 | 99.83536728 | 0.799327127 | 0.231438244 | 6.45E-05 | 0.000539967 |
| FCGBP | 148.4205818 | 0.799068759 | 0.218399087 | 2.97E-05 | 0.000270823 |
| NT5C1A | 66.98434859 | -0.798666125 | 0.29033415 | 0.000622713 | 0.003954639 |
| TNFRSF14-AS1 | 24.06407999 | 0.798525554 | 0.467749697 | 0.006528329 | 0.028744236 |
| CEL | 57.03478945 | -0.797974918 | 0.306069171 | 0.000923646 | 0.005553742 |
| EIF3EP3 | 1200.529245 | -0.797906038 | 0.088917414 | 4.06E-20 | 2.57E-18 |
| ST6GALNAC5 | 21.01041849 | 0.795647118 | 0.6930289 | 0.011829035 | 0.046534168 |
| SMURF2P1 | 33.35158275 | 0.795352132 | 0.391143106 | 0.003631642 | 0.017670452 |
| SORL1 | 199.5367338 | 0.795301064 | 0.164944634 | 1.81E-07 | 2.62E-06 |
| CELSR1 | 80.80274773 | 0.794811781 | 0.24491382 | 0.000131006 | 0.001007831 |
| NICN1 | 92.63663201 | -0.794780687 | 0.231208902 | 6.88E-05 | 0.000572089 |
| CAPG | 1468.960577 | 0.794633024 | 0.101478062 | 6.67E-16 | 3.04E-14 |
| DGCR11 | 14.65485219 | 0.794514648 | 0.793404974 | 0.013391619 | 0.051625704 |
| DEXI | 213.9561882 | -0.794050241 | 0.157169488 | 5.86E-08 | 9.21E-07 |
| PLCL1 | 33.39710723 | 0.794016083 | 0.452695079 | 0.006041405 | 0.026963661 |
| LOC107987192 | 9.138739743 | -0.793931462 | 1.146866475 | 0.013939158 | 0.053285187 |
| MRPL1 | 1316.816234 | -0.793480172 | 0.094119935 | 4.83E-18 | 2.50E-16 |
| NCAM2 | 173.3073561 | -0.792434685 | 0.215489835 | 2.71E-05 | 0.000249091 |
| METTL7A | 1387.892792 | 0.792099713 | 0.102148578 | 1.24E-15 | 5.39E-14 |
| CASP8 | 1515.758387 | 0.791908114 | 0.100601118 | 4.67E-16 | 2.14E-14 |
| STAT5A | 361.7051247 | 0.791891951 | 0.135248922 | 6.42E-10 | 1.39E-08 |

| | | | | | |
|---|---|---|---|---|---|
| CCNG1 | 7753.360676 | -0.790624748 | 0.097880716 | 1.00E-16 | 4.78E-15 |
| RASA4DP | 96.51495383 | -0.789394383 | 0.221800637 | 4.51E-05 | 0.000391706 |
| USP6NL | 562.5683203 | 0.78914049 | 0.128618833 | 1.12E-10 | 2.64E-09 |
| ZNF551 | 321.0566097 | 0.788803264 | 0.136258654 | 9.25E-10 | 1.95E-08 |
| BFSP2 | 17.17025923 | 0.787947747 | 0.611214131 | 0.011312333 | 0.045023136 |
| PAIP2B | 120.2112018 | 0.786902336 | 0.216561362 | 3.37E-05 | 0.000302251 |
| ZNF530 | 151.8568184 | 0.786412562 | 0.190527131 | 4.50E-06 | 4.90E-05 |
| MINAR1 | 72.69879168 | -0.785706111 | 0.284367016 | 0.000601452 | 0.003837635 |
| SLC22A23 | 75.09005294 | 0.785334308 | 0.304419452 | 0.001006203 | 0.005973403 |
| FAM72B | 276.5884087 | 0.785253611 | 0.15200758 | 3.09E-08 | 5.11E-07 |
| MTSS1 | 31.85262226 | 0.784616724 | 0.419825221 | 0.005038692 | 0.023243215 |
| SDC1 | 190359.9876 | -0.784404833 | 0.081006575 | 5.13E-23 | 3.95E-21 |
| SUGCT | 8.640869249 | -0.784114959 | 1.697928699 | 0.013407405 | 0.051675055 |
| CCDC110 | 235.5910521 | -0.783528211 | 0.21812584 | 3.86E-05 | 0.000341118 |
| LIG4 | 518.9173949 | -0.783517114 | 0.138436677 | 1.96E-09 | 3.94E-08 |
| PLXNC1 | 3403.80757 | 0.783444733 | 0.070517118 | 1.57E-29 | 1.74E-27 |
| TBC1D16 | 408.4182947 | -0.782155727 | 0.118549825 | 5.85E-12 | 1.69E-10 |
| TBXAS1 | 35.94837402 | -0.780620491 | 0.390785747 | 0.004065719 | 0.019363602 |
| MGAT3 | 1952.218556 | -0.780099442 | 0.105887666 | 2.60E-14 | 9.88E-13 |
| NME7 | 160.6323055 | -0.779369046 | 0.176011404 | 1.23E-06 | 1.52E-05 |
| C12orf56 | 43.16130686 | 0.779182172 | 0.353817728 | 0.002599665 | 0.013372996 |
| SELENON | 242.9619578 | 0.778339633 | 0.14398582 | 8.66E-09 | 1.56E-07 |
| DNAH17 | 15.7762061 | -0.778329921 | 0.704918361 | 0.0138071 | 0.052869389 |
| FHIT | 119.5427364 | -0.778029602 | 0.199076986 | 1.18E-05 | 0.000118166 |
| CDIP1 | 1440.891656 | -0.777999361 | 0.0990665 | 6.20E-16 | 2.84E-14 |
| HMCES | 1566.196877 | -0.777931507 | 0.099540663 | 8.13E-16 | 3.63E-14 |
| HMOX1 | 2322.87558 | -0.777931345 | 0.103332009 | 7.39E-15 | 2.94E-13 |
| SLC7A6 | 2172.465486 | -0.777903748 | 0.079596634 | 2.18E-23 | 1.72E-21 |
| CNTNAP1 | 344.4723703 | -0.777669649 | 0.14182593 | 5.75E-09 | 1.06E-07 |
| MAP3K1 | 1709.671823 | 0.777412608 | 0.104358241 | 1.28E-14 | 5.02E-13 |
| ITGB7 | 7521.676215 | 0.777370088 | 0.093208888 | 1.05E-17 | 5.33E-16 |
| RARA | 464.1213719 | 0.77711794 | 0.139336657 | 3.27E-09 | 6.35E-08 |
| NFXL1 | 2357.119228 | 0.775912676 | 0.082270223 | 5.74E-22 | 4.12E-20 |
| SEMA4B | 1205.654422 | 0.775581523 | 0.110249469 | 2.79E-13 | 9.47E-12 |
| NLRP7 | 56.88722944 | 0.775565629 | 0.291799408 | 0.000842174 | 0.005124358 |
| SMTN | 59.5034334 | 0.775404162 | 0.306270079 | 0.001173067 | 0.006810374 |
| E2F5 | 508.8930304 | -0.775348579 | 0.112486909 | 7.64E-13 | 2.47E-11 |
| GPR18 | 91.46547222 | 0.775332501 | 0.238172572 | 0.000133208 | 0.001020689 |
| EMC3-AS1 | 52.79692952 | -0.774207823 | 0.307121303 | 0.001242117 | 0.007124201 |
| MGST1 | 2062.047232 | -0.7730251 | 0.088510611 | 3.59E-19 | 2.05E-17 |
| LYL1 | 40.16029657 | 0.772375333 | 0.404615433 | 0.00486668 | 0.022588125 |
| ISG20 | 1163.071572 | 0.772141514 | 0.093148024 | 1.65E-17 | 8.25E-16 |
| CDKN2D | 51.62480725 | 0.77211482 | 0.325316272 | 0.00179589 | 0.009810546 |
| COL18A1 | 92.23901312 | 0.770546754 | 0.24680271 | 0.000209995 | 0.001524731 |
| PXN | 708.7738897 | 0.770172301 | 0.106129547 | 5.74E-14 | 2.11E-12 |
| ZNF546 | 54.77767819 | 0.76994935 | 0.324688818 | 0.001786797 | 0.009763955 |
| ENPEP | 116.6629529 | -0.769818531 | 0.233128697 | 0.000112729 | 0.000882913 |
| REEP3 | 647.9767107 | 0.769489224 | 0.140142612 | 5.22E-09 | 9.70E-08 |
| DNAH3 | 17.5514648 | -0.768702051 | 0.780233012 | 0.014199691 | 0.053995619 |
| PLPP5 | 908.6123865 | -0.768542061 | 0.094404152 | 5.82E-17 | 2.79E-15 |

| | | | | | |
|---|---|---|---|---|---|
| TNS1 | 9.827193931 | 0.768063486 | 1.326220048 | 0.013513398 | 0.051991003 |
| PARP12 | 473.3447024 | 0.767700018 | 0.127641185 | 2.54E-10 | 5.70E-09 |
| SLAMF7 | 4117.403045 | -0.764822529 | 0.067930244 | 3.17E-30 | 3.77E-28 |
| TUBA4A | 4418.889515 | -0.763861944 | 0.086457688 | 1.51E-19 | 8.96E-18 |
| EIF3E | 19712.08128 | -0.763845969 | 0.070872464 | 6.60E-28 | 6.74E-26 |
| SDC4 | 351.9582076 | 0.763254385 | 0.13246127 | 1.15E-09 | 2.39E-08 |
| MYLK | 751.7175199 | -0.762897601 | 0.104880577 | 4.95E-14 | 1.84E-12 |
| ATXN7L1 | 31.48014061 | 0.76234629 | 0.411107716 | 0.005415595 | 0.024653485 |
| RNVU1-14 | 36.54691213 | 0.762194114 | 0.532430125 | 0.009846723 | 0.04029199 |
| MAN1C1 | 563.0810614 | 0.761095984 | 0.121055855 | 4.59E-11 | 1.15E-09 |
| PGGHG | 182.6617686 | -0.760809816 | 0.194102584 | 1.18E-05 | 0.0001175 |
| GLMP | 2431.401208 | -0.759611972 | 0.083595303 | 1.63E-20 | 1.06E-18 |
| SNORA68 | 17.48933751 | 0.759304184 | 0.587751939 | 0.011963197 | 0.046944638 |
| IRS1 | 749.8372678 | 0.7588372 | 0.099938208 | 4.59E-15 | 1.88E-13 |
| CKAP2L | 539.5997118 | 0.7586559 | 0.120425707 | 4.15E-11 | 1.05E-09 |
| RRM2 | 19820.27348 | -0.758552846 | 0.06856937 | 3.28E-29 | 3.55E-27 |
| TMEM170B | 142.1390857 | 0.758127095 | 0.243279075 | 0.000215669 | 0.001560038 |
| KIF3C | 199.7652508 | 0.757467029 | 0.157988893 | 2.24E-07 | 3.21E-06 |
| NME4 | 1161.698835 | -0.757057179 | 0.104781007 | 7.58E-14 | 2.73E-12 |
| SNX18 | 2422.888103 | 0.755551524 | 0.115641857 | 9.15E-12 | 2.58E-10 |
| ATP8B3 | 64.16747066 | 0.755025689 | 0.272282254 | 0.000642865 | 0.004064437 |
| CEP68 | 579.7694418 | 0.754187029 | 0.102036153 | 2.15E-14 | 8.27E-13 |
| PTPN18 | 791.6191148 | 0.753411944 | 0.097685806 | 1.81E-15 | 7.72E-14 |
| RNF149 | 515.7360575 | 0.753365933 | 0.133117636 | 2.12E-09 | 4.25E-08 |
| PPCDC | 339.3918135 | 0.753364169 | 0.135986342 | 4.33E-09 | 8.24E-08 |
| LINC02551 | 42.70437135 | 0.753231632 | 0.345727728 | 0.00292272 | 0.014747302 |
| ARMCX3 | 1278.58556 | -0.752978001 | 0.094136307 | 1.85E-16 | 8.60E-15 |
| REEP2 | 177.8714942 | -0.752678559 | 0.193614727 | 1.36E-05 | 0.000134049 |
| KLF6 | 4346.697527 | -0.75199137 | 0.07173047 | 1.59E-26 | 1.46E-24 |
| ARF4 | 4411.974842 | -0.75136097 | 0.111039599 | 1.95E-12 | 5.99E-11 |
| RAC3 | 1524.948206 | -0.750937805 | 0.091366847 | 3.14E-17 | 1.54E-15 |
| KLHL32 | 100.976759 | 0.750182304 | 0.245125829 | 0.000261281 | 0.001844515 |
| FAAP24 | 180.1476995 | 0.750106153 | 0.166334603 | 8.96E-07 | 1.14E-05 |
| ZNF296 | 370.9659256 | 0.749723081 | 0.15023686 | 8.51E-08 | 1.30E-06 |
| PKN3 | 49.17638976 | 0.747903216 | 0.343011536 | 0.002980278 | 0.014989154 |
| LOC105379362 | 141.2992252 | -0.747569924 | 0.195527872 | 1.79E-05 | 0.000171763 |
| BHLHE41 | 458.0992311 | 0.747408244 | 0.146218419 | 4.41E-08 | 7.14E-07 |
| AMPD3 | 336.9934546 | -0.746892979 | 0.128101876 | 8.33E-10 | 1.77E-08 |
| RFFL | 482.8764094 | 0.746796408 | 0.115706577 | 1.61E-11 | 4.41E-10 |
| APOL2 | 303.9880875 | 0.745943667 | 0.145519572 | 4.18E-08 | 6.80E-07 |
| FAM111A-DT | 13.60049536 | 0.744661615 | 0.739909737 | 0.01495327 | 0.056146091 |
| ASCC3 | 2236.290265 | -0.743698705 | 0.087973085 | 4.26E-18 | 2.21E-16 |
| TNFAIP8 | 1180.496147 | 0.743594315 | 0.134223398 | 4.22E-09 | 8.06E-08 |
| PRKCQ-AS1 | 168.1798299 | 0.743593433 | 0.180307762 | 5.09E-06 | 5.48E-05 |
| TFAP2A | 23.05822193 | 0.743130337 | 0.517155559 | 0.010776977 | 0.043280362 |
| TEC | 1232.965467 | 0.742831385 | 0.08354374 | 9.13E-20 | 5.56E-18 |
| FCER1G | 43.62454823 | 0.742423293 | 0.367717073 | 0.004110258 | 0.019554216 |
| RBPMS2 | 15.25797239 | 0.740700969 | 0.745131282 | 0.015076121 | 0.056530808 |
| GPNMB | 239.4396951 | -0.740255904 | 0.148176031 | 8.52E-08 | 1.31E-06 |
| MANF | 6204.97689 | -0.740107099 | 0.068638718 | 6.58E-28 | 6.74E-26 |

| | | | | | |
|---|---|---|---|---|---|
| IDI1 | 8806.786668 | -0.738635866 | 0.107384466 | 5.69E-13 | 1.87E-11 |
| CLIP2 | 47.0814177 | 0.738355838 | 0.369936971 | 0.004384083 | 0.020624602 |
| ZFPM2 | 8.424179327 | 0.737532045 | 1.558194767 | 0.013186492 | 0.050959717 |
| DKFZP586I1420 | 261.7066729 | 0.737470285 | 0.172410122 | 2.59E-06 | 2.98E-05 |
| SNORD68 | 27.5896769 | -0.737122891 | 0.460202372 | 0.008924182 | 0.037097988 |
| FNBP1 | 2004.457328 | 0.736731036 | 0.07772201 | 4.00E-22 | 2.92E-20 |
| FAT1 | 11.88040931 | 0.736697253 | 0.904942452 | 0.015280711 | 0.057152498 |
| SNX10 | 366.8524545 | 0.736652473 | 0.198869566 | 2.77E-05 | 0.000254443 |
| RGCC | 392.5628524 | -0.736560385 | 0.136015204 | 9.26E-09 | 1.67E-07 |
| SSTR3 | 1618.577227 | 0.735626563 | 0.084031437 | 3.21E-19 | 1.85E-17 |
| SESN2 | 2627.712654 | -0.734342361 | 0.110028441 | 3.77E-12 | 1.11E-10 |
| DDIT3 | 1906.24835 | -0.733971161 | 0.091246644 | 1.39E-16 | 6.50E-15 |
| TCAF1 | 195.7212708 | 0.733809637 | 0.169817042 | 2.19E-06 | 2.56E-05 |
| CALR | 44845.41128 | -0.733790813 | 0.065313122 | 9.20E-30 | 1.04E-27 |
| PRDM11 | 203.8618304 | 0.733659072 | 0.175875975 | 4.28E-06 | 4.68E-05 |
| COPS4 | 1083.021509 | -0.733143356 | 0.096966293 | 6.19E-15 | 2.50E-13 |
| ADAM28 | 261.5268623 | 0.733023471 | 0.149478292 | 1.37E-07 | 2.04E-06 |
| MDFIC | 1014.150944 | 0.732729306 | 0.161600292 | 8.07E-07 | 1.04E-05 |
| PLS1 | 220.9341643 | 0.732728725 | 0.186310951 | 1.14E-05 | 0.000114597 |
| ZNF385A | 46.24319364 | 0.732606821 | 0.325305484 | 0.002587017 | 0.013321253 |
| IQSEC1 | 937.4233147 | -0.732059053 | 0.104403428 | 3.67E-13 | 1.23E-11 |
| ABCB6 | 265.6926678 | -0.731779718 | 0.190267672 | 1.70E-05 | 0.000164464 |
| KRBOX5 | 181.3499199 | -0.731555161 | 0.201352764 | 3.71E-05 | 0.00032895 |
| TRIP6 | 682.4091702 | -0.731391473 | 0.109891482 | 4.49E-12 | 1.32E-10 |
| SPPL2A | 679.1737331 | 0.731098285 | 0.105583244 | 6.70E-13 | 2.19E-11 |
| PLXNB2 | 1116.225036 | -0.730777234 | 0.101771745 | 1.11E-13 | 3.94E-12 |
| GPSM2 | 432.8363335 | 0.730689274 | 0.115969107 | 4.49E-11 | 1.13E-09 |
| ITGA7 | 24.28105105 | 0.730474812 | 0.510309701 | 0.011266474 | 0.04488188 |
| COL9A2 | 43.42915682 | -0.730344253 | 0.365788619 | 0.004646404 | 0.021699509 |
| PATL2 | 13.17642681 | 0.730184142 | 0.746206947 | 0.015536244 | 0.057883206 |
| IKBIP | 542.2633401 | -0.729837837 | 0.132016101 | 4.85E-09 | 9.11E-08 |
| TMEM198 | 124.9400475 | -0.72921556 | 0.224806261 | 0.000158675 | 0.001190541 |
| CMTM7 | 409.2464426 | -0.72909977 | 0.145515925 | 8.40E-08 | 1.29E-06 |
| C16orf95 | 187.0440278 | -0.728540586 | 0.198465824 | 3.39E-05 | 0.000303903 |
| ZFP41 | 70.87577193 | 0.728305705 | 0.265829514 | 0.000743749 | 0.004619622 |
| ISL1 | 21.88140881 | 0.728257301 | 0.550226194 | 0.012656946 | 0.049205692 |
| BCL6 | 91.68812948 | -0.728164118 | 0.246484189 | 0.000393471 | 0.002644058 |
| ZNF425 | 64.20738903 | -0.727887907 | 0.288680064 | 0.00134431 | 0.007625093 |
| ABHD8 | 58.73169455 | 0.726681291 | 0.29705771 | 0.001659603 | 0.009170179 |
| MXD3 | 74.69938082 | 0.726340428 | 0.325661549 | 0.002742067 | 0.014003058 |
| TPRG1 | 225.9868423 | 0.72618593 | 0.159384335 | 7.53E-07 | 9.74E-06 |
| CHST15 | 1331.705938 | 0.726101612 | 0.083916091 | 8.03E-19 | 4.44E-17 |
| CDH15 | 34.89073446 | -0.725706455 | 0.380505256 | 0.005632096 | 0.025445083 |
| APOC1 | 54.45784238 | -0.725195775 | 0.304238629 | 0.001991352 | 0.010687903 |
| SPATA2L | 216.1379059 | -0.725078033 | 0.193722735 | 2.61E-05 | 0.000240973 |
| ITGB8 | 246.6165396 | -0.723943684 | 0.214985672 | 9.93E-05 | 0.000791265 |
| SH3BP5 | 1051.626161 | 0.722389666 | 0.095139961 | 4.81E-15 | 1.96E-13 |
| MTSS2 | 20.5522689 | -0.722088833 | 0.549372545 | 0.013337293 | 0.051462098 |
| RAB43 | 465.406416 | 0.722045227 | 0.126200106 | 1.64E-09 | 3.33E-08 |
| TXK | 77.65014463 | 0.721853304 | 0.252332952 | 0.000527163 | 0.003415218 |

| | | | | | |
|---|---|---|---|---|---|
| TMEM178B | 21.10852698 | 0.720795916 | 0.536579257 | 0.012667169 | 0.049227417 |
| ZNF85 | 77.14849148 | 0.720305767 | 0.310889155 | 0.002280807 | 0.011997795 |
| ITGB8-AS1 | 225.6676022 | -0.720223681 | 0.157527905 | 7.27E-07 | 9.44E-06 |
| UBAC1 | 1524.063832 | 0.71965662 | 0.087358239 | 2.82E-17 | 1.39E-15 |
| ADGRG5 | 640.3331782 | 0.719570457 | 0.121915451 | 5.61E-10 | 1.22E-08 |
| SCOC | 341.4218573 | -0.719329387 | 0.231660212 | 0.000245883 | 0.001754412 |
| KDELR3 | 488.1267398 | -0.719308919 | 0.114742763 | 5.88E-11 | 1.45E-09 |
| IGHV1OR15-1 | 119.9881362 | 0.719244602 | 0.200422889 | 4.60E-05 | 0.000398892 |
| LOC102724760 | 119.9881362 | 0.719244602 | 0.200422889 | 4.60E-05 | 0.000398892 |
| PABPC1P3 | 27.89880592 | -0.71868729 | 0.600137046 | 0.013808872 | 0.052869389 |
| RSL1D1 | 5647.884187 | -0.717964264 | 0.066883451 | 1.17E-27 | 1.18E-25 |
| TJP3 | 101.5829302 | 0.717872934 | 0.230240181 | 0.000240289 | 0.00172088 |
| GPSM1 | 73.25119987 | 0.716513768 | 0.249815611 | 0.000529446 | 0.003427445 |
| ARID3B | 441.0180016 | -0.716418601 | 0.116490032 | 1.25E-10 | 2.94E-09 |
| SYTL3 | 182.151833 | -0.716324898 | 0.164805637 | 2.09E-06 | 2.46E-05 |
| PXMP4 | 112.7590612 | 0.716246062 | 0.202993559 | 5.79E-05 | 0.000490573 |
| SGMS1 | 559.980646 | 0.716189436 | 0.127967816 | 3.32E-09 | 6.45E-08 |
| ARNTL | 288.3733932 | 0.716135379 | 0.135138711 | 1.77E-08 | 3.02E-07 |
| FTL | 128584.9825 | -0.715995685 | 0.081425699 | 2.27E-19 | 1.34E-17 |
| HABP4 | 51.06790023 | 0.715671454 | 0.300020403 | 0.001982853 | 0.010661027 |
| LETM2 | 77.38842046 | -0.715574717 | 0.28219662 | 0.001332267 | 0.007569185 |
| PTPRJ | 106.8364725 | -0.715456057 | 0.218215239 | 0.000140778 | 0.00107064 |
| ELF4 | 329.4048386 | 0.71386817 | 0.134773278 | 1.82E-08 | 3.10E-07 |
| HIBCH | 935.1909037 | -0.71355641 | 0.102790304 | 6.15E-13 | 2.02E-11 |
| PSMB8-AS1 | 234.7826224 | 0.713380486 | 0.154141159 | 5.58E-07 | 7.40E-06 |
| LOC732229 | 186.0681183 | -0.713359255 | 0.18163102 | 1.27E-05 | 0.000126119 |
| D2HGDH | 115.9682708 | -0.713311383 | 0.202959071 | 6.32E-05 | 0.000531432 |
| SCNN1B | 123.0180647 | 0.712969547 | 0.195130481 | 3.66E-05 | 0.000325832 |
| MCPH1-DT | 41.20515367 | -0.712665247 | 0.345695567 | 0.004222792 | 0.020007136 |
| PDIA4 | 11979.98591 | -0.712643704 | 0.063941479 | 1.28E-29 | 1.43E-27 |
| EPS8 | 43.43780285 | -0.712347633 | 0.339817666 | 0.00394493 | 0.018881802 |
| HMGCS1 | 28065.39591 | -0.711784961 | 0.075068932 | 4.16E-22 | 3.03E-20 |
| RAB30 | 3618.861147 | -0.711543256 | 0.12216073 | 9.02E-10 | 1.90E-08 |
| SPOUT1 | 1014.308928 | 0.711156636 | 0.1110234 | 2.40E-11 | 6.30E-10 |
| MPG | 549.0884669 | -0.711147185 | 0.134044079 | 1.81E-08 | 3.08E-07 |
| TRIAP1 | 1468.885456 | -0.711136292 | 0.082758956 | 1.42E-18 | 7.67E-17 |
| TNFAIP8L1 | 99.97929865 | 0.710094019 | 0.217393241 | 0.000149836 | 0.001134549 |
| TMEM217 | 29.76978327 | -0.709659412 | 0.413825569 | 0.008114731 | 0.034332468 |
| TNFRSF11A | 1528.892728 | 0.708419193 | 0.079981129 | 1.35E-19 | 8.07E-18 |
| RAB37 | 1753.5347 | 0.707981473 | 0.083541771 | 3.93E-18 | 2.04E-16 |
| ABCD2 | 339.7475811 | 0.707853464 | 0.18366277 | 1.65E-05 | 0.000159638 |
| DNAJB1 | 6182.15646 | -0.707194148 | 0.10295037 | 9.92E-13 | 3.18E-11 |
| RARG | 74.08243426 | 0.70701416 | 0.282423278 | 0.00148639 | 0.008332734 |
| RPL34 | 21789.24854 | -0.706733429 | 0.078704632 | 4.66E-20 | 2.93E-18 |
| LOC122539214 | 27.99694943 | 0.706367103 | 0.470362524 | 0.010770512 | 0.043274467 |
| MKLN1-AS | 48.18286467 | -0.706065101 | 0.33283378 | 0.003714085 | 0.017948258 |
| PTP4A1 | 5277.769506 | -0.705636187 | 0.131619084 | 1.32E-08 | 2.29E-07 |
| GABARAPL1 | 224.7861894 | -0.705607383 | 0.153481798 | 6.63E-07 | 8.68E-06 |
| RPS21P4 | 48.34009761 | 0.705591885 | 0.379494129 | 0.006248467 | 0.027730576 |
| NR6A1 | 120.6695341 | -0.704621777 | 0.205551099 | 8.60E-05 | 0.000694494 |

| | | | | | |
|---|---|---|---|---|---|
| GALNS | 672.5531621 | -0.704377727 | 0.119028272 | 5.38E-10 | 1.18E-08 |
| MAL2 | 48.25773536 | 0.703934414 | 0.31799006 | 0.0030389 | 0.015249094 |
| IRF2 | 1104.691681 | 0.703808209 | 0.102609831 | 1.10E-12 | 3.51E-11 |
| WNT10A | 1536.413458 | -0.703783248 | 0.130211119 | 1.05E-08 | 1.87E-07 |
| BTN3A1 | 856.4212354 | 0.703542651 | 0.093123014 | 6.96E-15 | 2.79E-13 |
| HELZ2 | 304.5657274 | 0.70347463 | 0.196832056 | 5.02E-05 | 0.000430842 |
| SPINT2 | 1953.458992 | -0.702573937 | 0.111558844 | 5.01E-11 | 1.25E-09 |
| PUM3 | 1648.249138 | 0.701923196 | 0.07921637 | 1.36E-19 | 8.09E-18 |
| ZNF442 | 25.07463269 | 0.701343402 | 0.469049854 | 0.011242744 | 0.044805827 |
| FAM111A | 747.3158669 | 0.70092349 | 0.124094752 | 2.57E-09 | 5.08E-08 |
| PPP1R15A | 3695.523641 | -0.700885976 | 0.081605554 | 1.50E-18 | 8.09E-17 |
| PRAF2 | 245.177178 | -0.700666151 | 0.170504416 | 6.20E-06 | 6.53E-05 |
| HECW2 | 39.01055439 | 0.700483511 | 0.38222995 | 0.00657521 | 0.028928587 |
| SELENOK | 1707.225957 | -0.700108541 | 0.07786059 | 4.19E-20 | 2.65E-18 |
| HMGA1 | 7718.870289 | -0.698826736 | 0.091308754 | 3.24E-15 | 1.34E-13 |
| SH3GLB2 | 2106.823145 | 0.698602465 | 0.111919904 | 6.98E-11 | 1.71E-09 |
| CBY2 | 84.69391031 | 0.698353778 | 0.24614852 | 0.00060483 | 0.003856345 |
| TBC1D4 | 597.0881416 | -0.698268994 | 0.110069637 | 3.69E-11 | 9.42E-10 |
| MFGE8 | 830.844094 | 0.697522101 | 0.108318381 | 1.99E-11 | 5.34E-10 |
| RAB15 | 54.01034174 | 0.697184482 | 0.304264339 | 0.002577385 | 0.013279554 |
| RCAN3 | 292.7602727 | 0.69693553 | 0.138510686 | 7.73E-08 | 1.19E-06 |
| EIF2AK3 | 8364.157791 | -0.696589509 | 0.062049193 | 5.28E-30 | 6.13E-28 |
| ORAI2 | 1926.395965 | 0.696553206 | 0.098621397 | 2.72E-13 | 9.24E-12 |
| OCIAD1 | 5924.291895 | -0.696539588 | 0.081968605 | 3.60E-18 | 1.88E-16 |
| MTAP | 476.0556834 | -0.696534409 | 0.117960481 | 5.74E-10 | 1.25E-08 |
| LOC101929028 | 172.4511542 | -0.695548349 | 0.184325624 | 2.42E-05 | 0.000224731 |
| TRIM47 | 158.1338417 | -0.695330097 | 0.200606236 | 7.83E-05 | 0.000640734 |
| FTLP3 | 5080.267129 | -0.694604658 | 0.092497599 | 1.06E-14 | 4.19E-13 |
| EPAS1 | 34.77186655 | 0.694501114 | 0.402453968 | 0.008165874 | 0.034464645 |
| CLK1 | 1252.882368 | -0.694105218 | 0.092787942 | 1.23E-14 | 4.81E-13 |
| RAD51L3-RFFL | 435.1956769 | 0.693432834 | 0.116797677 | 4.77E-10 | 1.05E-08 |
| PABPC3 | 660.9341866 | -0.693319646 | 0.117388755 | 5.97E-10 | 1.30E-08 |
| SNAR-G1 | 10.53848811 | 0.693256784 | 1.128265952 | 0.015733662 | 0.058492879 |
| HJURP | 2021.494476 | 0.692916196 | 0.081507494 | 3.18E-18 | 1.68E-16 |
| FZD4 | 37.94548726 | 0.692629203 | 0.370593754 | 0.006398511 | 0.028273287 |
| GPC1 | 108.7396264 | 0.69240118 | 0.226007892 | 0.000308343 | 0.00213583 |
| FAM86B2-DT | 103.5604071 | -0.689823965 | 0.220464765 | 0.000249013 | 0.001774552 |
| PLCG2 | 2265.354633 | -0.689815565 | 0.072713877 | 4.19E-22 | 3.04E-20 |
| PTPA | 3794.15037 | 0.689547004 | 0.099018232 | 5.55E-13 | 1.83E-11 |
| CDK19 | 161.8146364 | 0.689415085 | 0.175176095 | 1.26E-05 | 0.000124851 |
| LOC653653 | 163.8969273 | 0.688332384 | 0.17954544 | 1.94E-05 | 0.000184402 |
| LOC105374748 | 21.62318159 | -0.687183343 | 0.498132068 | 0.013707406 | 0.052597197 |
| STT3B | 14250.40871 | -0.687011328 | 0.090965834 | 7.24E-15 | 2.90E-13 |
| IL22RA1 | 29.91045208 | 0.686459631 | 0.414213578 | 0.009340829 | 0.038522299 |
| UPF3AP1 | 132.5226845 | -0.686396434 | 0.199272284 | 8.61E-05 | 0.000695344 |
| LINC02328 | 149.5358381 | 0.685610159 | 0.202196357 | 0.000101955 | 0.000809888 |
| SERPINE2 | 3594.767138 | 0.685232143 | 0.07395607 | 3.40E-21 | 2.28E-19 |
| KLRF1 | 126.9862902 | 0.685095606 | 0.198116933 | 8.08E-05 | 0.000658136 |
| UPRT | 122.1112552 | -0.683611942 | 0.231753548 | 0.000438599 | 0.002912276 |
| LINC01145 | 78.0064744 | 0.682977054 | 0.238139139 | 0.000575076 | 0.003689735 |

| | | | | | |
|---|---|---|---|---|---|
| RAB38 | 51.5515464 | -0.682221024 | 0.299558049 | 0.002859878 | 0.014514804 |
| LOC107987300 | 14.86193972 | -0.68190454 | 0.660786511 | 0.017605522 | 0.064073559 |
| BMPR2 | 696.8714099 | 0.681639903 | 0.143588359 | 3.29E-07 | 4.58E-06 |
| HIC2 | 248.84186 | -0.681357223 | 0.143715706 | 3.50E-07 | 4.83E-06 |
| IL10RA | 618.8537986 | 0.680791033 | 0.11120821 | 1.59E-10 | 3.68E-09 |
| TTC3 | 3154.33035 | -0.680555633 | 0.075825905 | 5.00E-20 | 3.10E-18 |
| SQLE | 8062.998332 | -0.680492661 | 0.064825851 | 1.64E-26 | 1.51E-24 |
| DUBR | 31.14959349 | 0.679159488 | 0.422020624 | 0.010055583 | 0.041001725 |
| PPM1D | 1291.627058 | -0.678127305 | 0.114923019 | 6.41E-10 | 1.38E-08 |
| LINC02693 | 346.5929075 | 0.678041501 | 0.13240047 | 5.13E-08 | 8.20E-07 |
| CNOT6L | 861.2333782 | -0.677925486 | 0.142893167 | 3.44E-07 | 4.76E-06 |
| CCM2 | 1222.808123 | 0.677397991 | 0.095259428 | 2.00E-13 | 6.91E-12 |
| SLC27A2 | 310.2414433 | 0.675659815 | 0.130624978 | 3.86E-08 | 6.31E-07 |
| KIR3DX1 | 339.8229801 | 0.674911075 | 0.126328889 | 1.54E-08 | 2.65E-07 |
| LRRC23 | 110.8158632 | -0.674881053 | 0.205070358 | 0.000152813 | 0.00115255 |
| TMEM243 | 876.6622947 | -0.674330785 | 0.114549464 | 6.84E-10 | 1.47E-08 |
| UPF3A | 871.9230542 | -0.674169723 | 0.092407796 | 5.36E-14 | 1.98E-12 |
| KCNH2 | 186.5193919 | -0.674101423 | 0.178726918 | 2.61E-05 | 0.00024075 |
| KRT18 | 87.69599706 | 0.673740978 | 0.243481633 | 0.000795713 | 0.004888372 |
| LOC105377673 | 21.73530803 | 0.673639791 | 0.622669732 | 0.017172667 | 0.062764685 |
| LOC124906285 | 32.40903389 | -0.672715013 | 0.405117235 | 0.009940003 | 0.0405778 |
| BTN3A3 | 624.1098513 | 0.671540171 | 0.101997773 | 7.99E-12 | 2.26E-10 |
| NAT8L | 403.0445296 | -0.671362818 | 0.148879628 | 1.11E-06 | 1.38E-05 |
| GFM2 | 2559.329049 | -0.671092486 | 0.081091008 | 2.34E-17 | 1.17E-15 |
| LRRK1 | 136.5937805 | 0.66907635 | 0.191977986 | 7.74E-05 | 0.000634344 |
| LPAR4 | 22.48939735 | 0.668931628 | 0.531787157 | 0.015564933 | 0.057958848 |
| UBFD1 | 1989.254261 | -0.668856474 | 0.080470349 | 1.75E-17 | 8.74E-16 |
| CHMP7 | 2233.560495 | 0.668804842 | 0.086089628 | 1.41E-15 | 6.08E-14 |
| DENND1B | 1521.612279 | 0.668776697 | 0.159020011 | 4.18E-06 | 4.57E-05 |
| IL6R | 2390.277949 | -0.668448401 | 0.085505309 | 1.02E-15 | 4.49E-14 |
| SDF2L1 | 1917.124192 | -0.668061661 | 0.11758948 | 2.37E-09 | 4.73E-08 |
| TRPC4 | 287.7131754 | -0.667361866 | 0.144669163 | 6.64E-07 | 8.68E-06 |
| TTC30A | 118.2355245 | 0.66681473 | 0.205023629 | 0.000172884 | 0.001286361 |
| PRG2 | 115.1799616 | -0.666740638 | 0.221274804 | 0.000389698 | 0.002624814 |
| BBX | 566.5528833 | 0.666565026 | 0.140824864 | 3.69E-07 | 5.07E-06 |
| JAG1 | 697.9993977 | -0.666255552 | 0.113578502 | 7.83E-10 | 1.67E-08 |
| NECTIN2 | 118.2672025 | 0.666165238 | 0.226685326 | 0.000484203 | 0.003168915 |
| RPS15AP19 | 3197.167805 | -0.665999954 | 0.093427453 | 1.80E-13 | 6.25E-12 |
| DNAJC3 | 5324.27249 | -0.665995873 | 0.111182695 | 3.70E-10 | 8.22E-09 |
| FTO | 813.8135348 | -0.665662808 | 0.103045893 | 1.85E-11 | 4.99E-10 |
| RUNX2 | 691.4679668 | 0.665524503 | 0.102767728 | 1.66E-11 | 4.49E-10 |
| LOC105369484 | 105.1359956 | 0.665015212 | 0.228977456 | 0.000542132 | 0.003495144 |
| NRARP | 57.55873312 | 0.664908522 | 0.288591345 | 0.002773605 | 0.014143262 |
| STEAP1 | 117.4996968 | 0.664602559 | 0.200229636 | 0.00013891 | 0.00105923 |
| PCDHA11 | 40.4594716 | -0.664024778 | 0.352536235 | 0.006931105 | 0.030225624 |
| HLA-F | 1081.727256 | 0.663934185 | 0.106840871 | 9.18E-11 | 2.20E-09 |
| MIR210HG | 21.12668927 | 0.663899614 | 0.526477967 | 0.016045367 | 0.059409369 |
| SAYSD1 | 95.86225259 | 0.663880447 | 0.234248273 | 0.000672473 | 0.00421928 |
| LPP | 87.80238281 | 0.663183248 | 0.26329639 | 0.001588093 | 0.008834244 |
| CLCN4 | 457.4357531 | 0.66284233 | 0.115036122 | 1.46E-09 | 3.00E-08 |

| | | | | | |
|---|---|---|---|---|---|
| ZFP36L2 | 1778.525499 | 0.662758876 | 0.093810595 | 2.85E-13 | 9.64E-12 |
| DNAJB9 | 2308.565143 | -0.66264028 | 0.146129295 | 9.68E-07 | 1.22E-05 |
| EHD1 | 5736.109622 | -0.661396839 | 0.095608061 | 8.94E-13 | 2.88E-11 |
| RPL29P33 | 1098.78346 | -0.661394792 | 0.090292658 | 4.35E-14 | 1.63E-12 |
| BRD3 | 284.1271493 | 0.660678884 | 0.14121301 | 4.88E-07 | 6.54E-06 |
| ZNF594 | 48.48978086 | 0.660484954 | 0.305910224 | 0.003885366 | 0.01865859 |
| ESAM | 33.18039882 | 0.660396618 | 0.390298868 | 0.009477321 | 0.039029434 |
| RASSF5 | 630.697432 | 0.660365917 | 0.102288171 | 1.91E-11 | 5.14E-10 |
| CHROMR | 276.6524871 | 0.659687952 | 0.137438147 | 2.72E-07 | 3.85E-06 |
| KCNK6 | 387.5738919 | -0.659437819 | 0.15622091 | 4.17E-06 | 4.57E-05 |
| STAC | 41.92577568 | -0.658908417 | 0.345511096 | 0.006656534 | 0.029234391 |
| LOC105379444 | 24.76714639 | -0.658759197 | 0.480116037 | 0.01479785 | 0.055719583 |
| CIITA | 315.8270466 | 0.658717512 | 0.146584278 | 1.19E-06 | 1.47E-05 |
| ARID3A | 930.3675933 | -0.658287574 | 0.101547536 | 1.64E-11 | 4.48E-10 |
| NEIL2 | 236.9574847 | -0.658257242 | 0.153533338 | 3.17E-06 | 3.56E-05 |
| TYRO3 | 303.5428939 | -0.65734629 | 0.131642254 | 1.06E-07 | 1.61E-06 |
| ANLN | 668.7838096 | 0.65677174 | 0.11762294 | 4.11E-09 | 7.86E-08 |
| SORBS1 | 67.67488899 | -0.656771275 | 0.277596485 | 0.002432364 | 0.012675668 |
| RRM2B | 1099.899837 | -0.656547882 | 0.133536531 | 1.53E-07 | 2.25E-06 |
| CENPV | 619.1484396 | -0.656508504 | 0.121538433 | 1.23E-08 | 2.15E-07 |
| DPH3 | 1028.20128 | -0.656279759 | 0.105029584 | 7.48E-11 | 1.82E-09 |
| CAMK4 | 223.9501814 | 0.656161793 | 0.176983267 | 3.41E-05 | 0.000305572 |
| PIK3AP1 | 657.6440329 | 0.656144456 | 0.106457093 | 1.28E-10 | 3.00E-09 |
| MAGED2 | 4306.263842 | -0.656096563 | 0.067806826 | 7.35E-23 | 5.61E-21 |
| TROAP | 977.5058703 | 0.656024017 | 0.106849982 | 1.51E-10 | 3.51E-09 |
| FUT11 | 138.0289071 | 0.655952602 | 0.18920559 | 8.53E-05 | 0.000690229 |
| ARRB1 | 31.598346 | -0.655913634 | 0.405999915 | 0.010877886 | 0.043594621 |
| TMEM203 | 773.8390795 | 0.65576237 | 0.11571826 | 2.63E-09 | 5.18E-08 |
| VWA5A | 105.4619147 | 0.654945832 | 0.216704682 | 0.000381422 | 0.002576085 |
| ZBTB18 | 125.2683364 | 0.654819424 | 0.199834967 | 0.000164549 | 0.001229824 |
| IGHV3-30 | 366.2075688 | -0.653742027 | 0.146508006 | 1.45E-06 | 1.76E-05 |
| DNAAF4-CCPG1 | 2512.108399 | 0.65293581 | 0.089438834 | 5.26E-14 | 1.95E-12 |
| ZHX2 | 447.4850175 | -0.652849682 | 0.136496411 | 3.08E-07 | 4.31E-06 |
| OVOL2 | 13.36376992 | 0.652194527 | 0.75435684 | 0.019401338 | 0.069189048 |
| RNFT2 | 143.0753109 | 0.65134334 | 0.216810919 | 0.0004105 | 0.002749395 |
| FADS2 | 605.0715748 | -0.651186947 | 0.11088345 | 8.09E-10 | 1.72E-08 |
| ZNF814 | 38.86893076 | 0.650792014 | 0.358273463 | 0.007899827 | 0.033558529 |
| TMEM184B | 1514.9338 | -0.650658089 | 0.099568864 | 1.19E-11 | 3.32E-10 |
| MARVELD1 | 245.1937182 | -0.650537477 | 0.15458172 | 4.54E-06 | 4.94E-05 |
| TMEM132A | 218.0191386 | -0.649985873 | 0.173585051 | 3.09E-05 | 0.000280145 |
| FABP5P2 | 274.0337448 | -0.649674763 | 0.147683833 | 1.88E-06 | 2.23E-05 |
| SLC16A13 | 130.8414426 | 0.649211984 | 0.192157816 | 0.00011783 | 0.000916653 |
| B4GALT1 | 2431.57335 | -0.649185774 | 0.086792549 | 1.43E-14 | 5.55E-13 |
| SLC9A3-AS1 | 41.43419392 | -0.648831238 | 0.420104839 | 0.012412586 | 0.04841188 |
| SCARNA12 | 39.44254781 | 0.648664538 | 0.559920389 | 0.017216108 | 0.06287038 |
| OSTCP2 | 26.42115654 | -0.648086209 | 0.443254664 | 0.013777206 | 0.052792549 |
| AMDHD1 | 26.24854251 | -0.647957099 | 0.452958168 | 0.014212642 | 0.054009998 |
| STAT2 | 906.2631058 | 0.647252867 | 0.089230133 | 7.60E-14 | 2.73E-12 |
| CDCA7 | 1248.312144 | -0.647237837 | 0.097727837 | 6.53E-12 | 1.87E-10 |
| GTSE1 | 881.2654138 | 0.647172173 | 0.095152141 | 1.94E-12 | 5.98E-11 |

| | | | | | |
|---|---|---|---|---|---|
| ZKSCAN2 | 118.1907121 | -0.647142512 | 0.204750463 | 0.000255284 | 0.001811786 |
| LOC105369618 | 54.2873092 | -0.647038297 | 0.302939665 | 0.004255456 | 0.020133008 |
| RBM47 | 878.4119089 | 0.645803765 | 0.104474936 | 1.17E-10 | 2.77E-09 |
| CTDSP1 | 2087.320149 | 0.6451289 | 0.104445684 | 1.21E-10 | 2.86E-09 |
| CTTN | 148.4607515 | -0.644792294 | 0.177178168 | 4.67E-05 | 0.000404705 |
| SOGA1 | 199.0681994 | 0.644724356 | 0.166040045 | 1.75E-05 | 0.00016835 |
| ARHGDIB | 6088.234581 | 0.644343128 | 0.063854514 | 1.19E-24 | 1.00E-22 |
| RBIS | 1913.246954 | -0.6441057 | 0.084985915 | 6.68E-15 | 2.68E-13 |
| SLC27A3 | 69.55057917 | 0.643993812 | 0.304013409 | 0.004442891 | 0.020872908 |
| LDLR | 3489.13975 | -0.643768017 | 0.082914098 | 1.60E-15 | 6.84E-14 |
| RUFY3 | 751.2413758 | -0.643624966 | 0.101675503 | 4.56E-11 | 1.14E-09 |
| MBNL2 | 690.6902641 | -0.643406587 | 0.134170737 | 2.93E-07 | 4.12E-06 |
| JRK | 289.8502916 | 0.64263426 | 0.139768541 | 7.70E-07 | 9.93E-06 |
| STK17B | 4507.361885 | 0.64209575 | 0.134532039 | 3.25E-07 | 4.51E-06 |
| NINJ2 | 79.76718843 | -0.641759114 | 0.256699429 | 0.001835556 | 0.009986277 |
| ZIC2 | 106.2743066 | -0.641164134 | 0.230514538 | 0.000833313 | 0.005084734 |
| SLC1A4 | 4690.569736 | 0.640693286 | 0.074834465 | 2.16E-18 | 1.15E-16 |
| SCART1 | 42.29085349 | 0.64066214 | 0.357330486 | 0.008622408 | 0.036022735 |
| MAGED1 | 6036.985272 | -0.640575558 | 0.070150592 | 1.34E-20 | 8.77E-19 |
| FABP5 | 1826.22715 | -0.640104134 | 0.082621762 | 1.70E-15 | 7.25E-14 |
| IRAK4 | 780.4102007 | 0.639727087 | 0.100154722 | 3.18E-11 | 8.24E-10 |
| ARRDC3 | 2254.745039 | -0.63930827 | 0.146295311 | 2.22E-06 | 2.58E-05 |
| PNP | 4064.265157 | -0.639148937 | 0.074203105 | 1.41E-18 | 7.64E-17 |
| RAPGEF2 | 973.3863998 | -0.639135154 | 0.114704791 | 4.73E-09 | 8.93E-08 |
| LINC01055 | 30.29652253 | -0.639113512 | 0.476810052 | 0.016530899 | 0.060726683 |
| PIK3R3 | 168.8266448 | -0.638879483 | 0.167443397 | 2.37E-05 | 0.000221164 |
| PLEKHG4 | 605.5575039 | -0.638520964 | 0.114484995 | 4.70E-09 | 8.90E-08 |
| TMEM250 | 740.3115583 | 0.638362756 | 0.11595942 | 6.93E-09 | 1.27E-07 |
| CHRM4 | 11.37296557 | 0.638290021 | 0.864029773 | 0.019058884 | 0.068176554 |
| CGAS | 248.4428091 | 0.638085779 | 0.141967163 | 1.25E-06 | 1.54E-05 |
| LRRC34 | 50.29123971 | -0.6379701 | 0.338143308 | 0.007422831 | 0.031959514 |
| CFH | 12.36737858 | 0.637831259 | 0.764415653 | 0.02010117 | 0.071199009 |
| HSP90B2P | 72.79112609 | -0.63780469 | 0.264999146 | 0.002364732 | 0.012374325 |
| TNFRSF17 | 6267.594282 | -0.637393157 | 0.077383986 | 3.52E-17 | 1.71E-15 |
| PIP5KL1 | 15.89536091 | 0.637053814 | 0.605868213 | 0.019944656 | 0.070745927 |
| CCPG1 | 3323.300641 | 0.637047454 | 0.107315586 | 5.46E-10 | 1.19E-08 |
| SLC25A30 | 466.4865133 | 0.637029923 | 0.144316785 | 1.80E-06 | 2.15E-05 |
| ABLIM1 | 919.3570799 | 0.636529131 | 0.091508294 | 6.98E-13 | 2.28E-11 |
| ZNF670-ZNF695 | 64.40576068 | 0.63564284 | 0.26737712 | 0.002535004 | 0.013104079 |
| LOC606724 | 1065.516694 | 0.635522263 | 0.130562424 | 2.14E-07 | 3.06E-06 |
| KCNMB4 | 387.7600267 | 0.635319345 | 0.117876752 | 1.33E-08 | 2.30E-07 |
| RNF170 | 519.8236347 | -0.634847414 | 0.123010779 | 4.63E-08 | 7.44E-07 |
| BTG2 | 5803.060969 | -0.634089947 | 0.075939341 | 1.34E-17 | 6.78E-16 |
| ZNF532 | 49.00320226 | 0.63398742 | 0.334380904 | 0.007158772 | 0.031062002 |
| H3-5 | 1097.119654 | -0.632811433 | 0.089946865 | 3.99E-13 | 1.33E-11 |
| MPZL2 | 164.6565037 | 0.632560302 | 0.206642026 | 0.000354817 | 0.002412391 |
| PANX2 | 47.05111844 | 0.631858856 | 0.35117344 | 0.008610708 | 0.035982535 |
| NT5E | 259.0858404 | 0.631720202 | 0.137000924 | 7.39E-07 | 9.58E-06 |
| LILRP1 | 10.46842495 | 0.631699138 | 1.02071162 | 0.017481631 | 0.063732507 |
| TMEM50B | 1215.515829 | -0.631616105 | 0.097968126 | 2.20E-11 | 5.82E-10 |

| | | | | | |
|---|---|---|---|---|---|
| ZNF773 | 62.16780611 | 0.631555264 | 0.281489396 | 0.003474834 | 0.017051202 |
| EIF3H | 11424.02529 | -0.631395198 | 0.064449055 | 2.33E-23 | 1.83E-21 |
| ACSM3 | 96.49514219 | -0.630470164 | 0.23621697 | 0.001215066 | 0.007010095 |
| NOMO3 | 3763.905226 | -0.629797596 | 0.072366699 | 6.60E-19 | 3.66E-17 |
| PFKFB3 | 888.1010243 | 0.629169091 | 0.091720233 | 1.35E-12 | 4.25E-11 |
| MPHOSPH9 | 619.4592503 | 0.628444517 | 0.118053413 | 1.92E-08 | 3.26E-07 |
| CHST11 | 1219.707205 | 0.628198357 | 0.096602665 | 1.54E-11 | 4.21E-10 |
| IGHV3-33 | 401.9633727 | -0.627757342 | 0.139469927 | 1.30E-06 | 1.59E-05 |
| CENPA | 920.7560027 | 0.627684945 | 0.101677954 | 1.30E-10 | 3.03E-09 |
| ZNF695 | 127.2227739 | 0.627527659 | 0.190975609 | 0.000173801 | 0.001292273 |
| RAET1K | 5.797688895 | 0.627385961 | 3.420564635 | 0.01073742 | 0.043193519 |
| LRRC8C | 975.0628175 | 0.626965064 | 0.121236974 | 4.42E-08 | 7.15E-07 |
| LINC01611 | 37.86388715 | 0.626066226 | 0.394976979 | 0.012487644 | 0.048660784 |
| SKA1 | 1412.277161 | 0.625706585 | 0.096069569 | 1.45E-11 | 3.99E-10 |
| GOLGA4 | 314.4675128 | -0.625631939 | 0.150551513 | 5.86E-06 | 6.22E-05 |
| NOMO1 | 5466.590621 | -0.625532775 | 0.067072924 | 2.25E-21 | 1.54E-19 |
| SPIB | 4787.063148 | 0.625260793 | 0.091827573 | 1.85E-12 | 5.74E-11 |
| H2BC12L | 123.7318378 | -0.624663555 | 0.216352465 | 0.000626883 | 0.003975006 |
| LOC102724334 | 123.7318378 | -0.624663555 | 0.216352465 | 0.000626883 | 0.003975006 |
| NOMO2 | 4748.746331 | -0.62450458 | 0.067188728 | 3.21E-21 | 2.16E-19 |
| LINC02267 | 22.45911132 | -0.624485696 | 0.506461375 | 0.018675348 | 0.067067815 |
| MICAL2 | 182.345634 | -0.624476608 | 0.190945188 | 0.000182445 | 0.001349018 |
| TMEM231 | 333.2945244 | -0.623867473 | 0.136559476 | 9.57E-07 | 1.21E-05 |
| IFTAP | 263.7379505 | -0.623686309 | 0.145509508 | 3.42E-06 | 3.81E-05 |
| ZDHHC14 | 320.6425989 | 0.623424141 | 0.143820827 | 2.73E-06 | 3.11E-05 |
| HTATSF1P2 | 33.20417684 | 0.622842286 | 0.378462581 | 0.011563965 | 0.045761695 |
| ELF3-AS1 | 23.32715855 | -0.622280355 | 0.519501173 | 0.019448512 | 0.069298329 |
| STAG3L5P | 40.64814352 | -0.622217284 | 0.365130178 | 0.010732294 | 0.043191061 |
| LINC02864 | 328.2010829 | 0.622168937 | 0.133738955 | 6.24E-07 | 8.20E-06 |
| GREB1 | 156.523535 | -0.622104585 | 0.191955681 | 0.000203895 | 0.001485424 |
| SLC19A2 | 802.0442517 | -0.622103449 | 0.157202372 | 1.39E-05 | 0.000135949 |
| NTHL1 | 674.0296296 | -0.621947789 | 0.11151407 | 4.95E-09 | 9.27E-08 |
| RASA4CP | 75.72632998 | -0.621842988 | 0.245469252 | 0.001772655 | 0.009694115 |
| TRIM59 | 375.0492006 | 0.621828661 | 0.164069657 | 2.70E-05 | 0.000248537 |
| CIT | 293.2479165 | 0.621635482 | 0.147867932 | 4.81E-06 | 5.20E-05 |
| IGF2BP3 | 863.2582772 | -0.621328474 | 0.109034027 | 2.34E-09 | 4.68E-08 |
| LUC7L | 406.0395277 | -0.620870887 | 0.118190916 | 2.91E-08 | 4.83E-07 |
| BRD3OS | 346.297397 | 0.620774237 | 0.146367995 | 4.16E-06 | 4.56E-05 |
| ZNFX1 | 982.488868 | 0.620587873 | 0.097066806 | 3.21E-11 | 8.29E-10 |
| PIGY | 958.1668947 | -0.620526402 | 0.095465607 | 1.66E-11 | 4.49E-10 |
| PYURF | 958.1668947 | -0.620526402 | 0.095465607 | 1.66E-11 | 4.49E-10 |
| LOC105372903 | 36.51482233 | -0.620277513 | 0.368451828 | 0.011103777 | 0.044356194 |
| HDAC7 | 1446.65401 | 0.620254265 | 0.09958276 | 9.37E-11 | 2.24E-09 |
| KLF13 | 5193.43779 | 0.620206517 | 0.085026262 | 6.05E-14 | 2.21E-12 |
| IFI16 | 2161.785973 | 0.620176163 | 0.095166134 | 1.42E-11 | 3.91E-10 |
| NPL | 61.41351583 | 0.619732094 | 0.288532742 | 0.00448224 | 0.021029248 |
| CDC25C | 323.965925 | 0.61914164 | 0.125142199 | 1.45E-07 | 2.15E-06 |
| CEP55 | 1362.874997 | 0.61891591 | 0.105249264 | 7.99E-10 | 1.70E-08 |
| IPO5 | 12052.63991 | -0.618683536 | 0.063463148 | 4.22E-23 | 3.31E-21 |
| CDKN2C | 1407.352834 | 0.61790821 | 0.086294662 | 1.61E-13 | 5.62E-12 |

| | | | | | |
|---|---|---|---|---|---|
| KIAA0895L | 231.7227934 | -0.617824133 | 0.166530068 | 3.83E-05 | 0.000338896 |
| MANBA | 429.0677132 | -0.617753262 | 0.113776445 | 1.13E-08 | 2.00E-07 |
| ZNF586 | 310.1676771 | 0.617633065 | 0.139679331 | 1.83E-06 | 2.19E-05 |
| KLHL15 | 600.3362506 | -0.617615485 | 0.145552424 | 4.12E-06 | 4.52E-05 |
| CDC20P1 | 19.55819092 | 0.61743418 | 0.536346877 | 0.020332375 | 0.07184473 |
| LINC02210-CRHR1 | 39.50337027 | 0.61725489 | 0.343840364 | 0.009237566 | 0.038132762 |
| RIDA | 495.0760493 | -0.617070538 | 0.156068741 | 1.42E-05 | 0.000138661 |
| IGHJ3P | 150.18929 | -0.616918337 | 0.196004999 | 0.000289356 | 0.002018847 |
| TRAP1 | 6989.58976 | -0.616559623 | 0.071650599 | 1.60E-18 | 8.60E-17 |
| SOCS2 | 409.1236765 | -0.616073857 | 0.123831599 | 1.28E-07 | 1.92E-06 |
| UQCRBP1 | 591.0302906 | -0.615829859 | 0.158582648 | 1.97E-05 | 0.000186681 |
| CENPT | 369.6827836 | -0.615602865 | 0.126191904 | 2.14E-07 | 3.07E-06 |
| MTMR10 | 1032.736435 | 0.615593755 | 0.095094933 | 1.93E-11 | 5.18E-10 |
| CECR7 | 17.24537199 | 0.615375202 | 0.580458299 | 0.021205936 | 0.074457256 |
| VSIG10 | 21.43151224 | 0.6152825 | 0.579943139 | 0.020923375 | 0.073614401 |
| TGFBR2 | 287.7211423 | 0.614885355 | 0.141996874 | 2.82E-06 | 3.20E-05 |
| CDKN2A | 1405.458531 | 0.614854916 | 0.129395348 | 3.89E-07 | 5.31E-06 |
| ATXN1 | 1163.278936 | 0.614395507 | 0.110277843 | 5.01E-09 | 9.35E-08 |
| EPB41L5 | 69.90612289 | 0.614221208 | 0.271215939 | 0.003483755 | 0.017080462 |
| LOC124904996 | 90.08756024 | -0.614133355 | 0.227320108 | 0.001148097 | 0.006691885 |
| CARD10 | 38.28877453 | 0.614130237 | 0.368121971 | 0.011524313 | 0.045677849 |
| C11orf1 | 5518.218844 | -0.6138466 | 0.066570668 | 6.41E-21 | 4.23E-19 |
| TRAF2 | 723.2296709 | 0.613821753 | 0.11851811 | 4.39E-08 | 7.12E-07 |
| ALPK2 | 305.8299734 | 0.613548118 | 0.147202144 | 5.73E-06 | 6.10E-05 |
| C8orf44 | 18.05531418 | -0.612839303 | 0.755632109 | 0.021632567 | 0.075679023 |
| STK38 | 1028.674696 | 0.612380887 | 0.089101964 | 1.29E-12 | 4.07E-11 |
| GOLGA1 | 197.5721516 | 0.611921148 | 0.191120978 | 0.000237291 | 0.001702221 |
| TMPO-AS1 | 90.17060202 | 0.611891344 | 0.230422968 | 0.001298667 | 0.00740257 |
| LOC102723728 | 1175.60895 | -0.611814816 | 0.083139198 | 3.87E-14 | 1.45E-12 |
| CCDC69 | 5409.070865 | 0.610610309 | 0.073120841 | 1.41E-17 | 7.08E-16 |
| TMEM63A | 407.5474737 | 0.610355723 | 0.131819255 | 7.14E-07 | 9.28E-06 |
| POGZ | 1796.432949 | -0.609794692 | 0.08009404 | 5.58E-15 | 2.26E-13 |
| IMPDH2 | 7715.568447 | -0.609697747 | 0.068267858 | 9.39E-20 | 5.71E-18 |
| ST6GAL1 | 3535.099867 | 0.609697419 | 0.069474812 | 3.53E-19 | 2.02E-17 |
| IFT20 | 341.1708324 | -0.609194752 | 0.139044283 | 2.35E-06 | 2.73E-05 |
| DCBLD2 | 175.2048284 | -0.60900767 | 0.191387118 | 0.000261874 | 0.001847202 |
| FAM43A | 82.0097253 | 0.60863525 | 0.28937406 | 0.005168946 | 0.02373658 |
| RPS19 | 53083.81081 | -0.608501431 | 0.107362725 | 1.70E-09 | 3.45E-08 |
| KIF23 | 1134.485311 | 0.608477676 | 0.099781333 | 2.16E-10 | 4.90E-09 |
| SLC5A6 | 2633.947966 | -0.607586547 | 0.098983392 | 1.75E-10 | 4.03E-09 |
| LGALSL | 725.2665921 | -0.607515851 | 0.106925067 | 2.76E-09 | 5.43E-08 |
| IL2RB | 1127.437856 | -0.607486787 | 0.114574166 | 2.37E-08 | 3.99E-07 |
| RREB1 | 265.5203657 | 0.607206464 | 0.147784621 | 7.63E-06 | 7.88E-05 |
| GLDC | 6711.627975 | -0.607132684 | 0.069380242 | 4.55E-19 | 2.57E-17 |
| UGDH | 1534.047654 | -0.60672253 | 0.09621741 | 5.88E-11 | 1.45E-09 |
| ARHGAP9 | 111.7189513 | -0.606708905 | 0.238644807 | 0.001808399 | 0.009857113 |
| JUNB | 1832.771373 | -0.6067003 | 0.107658665 | 3.81E-09 | 7.31E-08 |
| DSCR4 | 66.21315538 | -0.606113655 | 0.267439494 | 0.0036542 | 0.01774531 |
| KIF18A | 477.8250108 | 0.605662628 | 0.127950381 | 4.34E-07 | 5.86E-06 |
| SLC6A9 | 649.8707195 | 0.605333985 | 0.135466356 | 1.56E-06 | 1.88E-05 |

| | | | | | |
|---|---|---|---|---|---|
| SLC31A1 | 2010.185376 | -0.60525184 | 0.079323879 | 5.11E-15 | 2.08E-13 |
| SH3BP1 | 1184.29829 | 0.604980551 | 0.108509725 | 5.04E-09 | 9.39E-08 |
| SEC11C | 7014.392911 | -0.604904372 | 0.070770478 | 2.73E-18 | 1.44E-16 |
| HRK | 167.6106098 | 0.604887626 | 0.18305049 | 0.000172527 | 0.001285459 |
| LOC124906065 | 46.59599084 | -0.604843613 | 0.315868052 | 0.007844487 | 0.033417707 |
| AP1S2 | 1833.296794 | 0.604772918 | 0.274403833 | 0.004090241 | 0.01946433 |
| TSPAN33 | 581.5505065 | 0.604712509 | 0.128121415 | 4.72E-07 | 6.34E-06 |
| LSP1 | 7486.696482 | 0.604694096 | 0.074982909 | 7.13E-17 | 3.41E-15 |
| NBR2 | 36.96021278 | 0.604506421 | 0.390795695 | 0.014187908 | 0.053962661 |
| RPS6KA4 | 412.0600437 | 0.604105019 | 0.133887988 | 1.27E-06 | 1.56E-05 |
| VANGL1 | 321.6191121 | 0.603529244 | 0.127940043 | 4.77E-07 | 6.40E-06 |
| LOC124907737 | 15.22375124 | -0.603409618 | 0.623493717 | 0.022988391 | 0.079379854 |
| FRMD6 | 1396.130487 | 0.603347871 | 0.09034067 | 5.07E-12 | 1.48E-10 |
| ZFP36 | 1556.071319 | -0.603307467 | 0.103376932 | 1.15E-09 | 2.39E-08 |
| CEP85L | 181.6041708 | -0.603225248 | 0.173524771 | 9.52E-05 | 0.000762001 |
| ASPM | 1406.314637 | 0.603072464 | 0.115383052 | 3.44E-08 | 5.65E-07 |
| HDAC6 | 439.5741465 | 0.602821754 | 0.121322264 | 1.37E-07 | 2.04E-06 |
| ZNF671 | 108.305026 | 0.602596309 | 0.206973981 | 0.000632911 | 0.004010292 |
| SLC7A5 | 19816.08882 | -0.6025881 | 0.08828982 | 1.86E-12 | 5.76E-11 |
| CPNE7 | 191.4549718 | -0.601981425 | 0.167051596 | 6.10E-05 | 0.000514245 |
| ZDHHC12 | 1413.25597 | 0.601972323 | 0.134934344 | 1.62E-06 | 1.95E-05 |
| PLPPR2 | 278.7267872 | -0.601895115 | 0.139162508 | 3.08E-06 | 3.47E-05 |
| P3R3URF-PIK3R3 | 143.6641538 | -0.601729177 | 0.181665855 | 0.000172587 | 0.001285459 |
| CMTM3 | 265.152471 | 0.601618035 | 0.16011803 | 3.33E-05 | 0.00029999 |
| ATP8A1 | 1500.794163 | 0.601499452 | 0.125567401 | 3.33E-07 | 4.62E-06 |
| GABARAPL2 | 1937.961548 | -0.601346015 | 0.079522392 | 8.63E-15 | 3.42E-13 |
| N4BP3 | 521.8308196 | 0.601252452 | 0.123858521 | 2.46E-07 | 3.49E-06 |
| FZD7 | 267.6070184 | 0.600927545 | 0.158606031 | 2.95E-05 | 0.000268777 |
| SSNA1 | 1465.625713 | 0.600797056 | 0.110469546 | 1.10E-08 | 1.95E-07 |
| ALG1 | 910.3490292 | -0.600718213 | 0.124279079 | 2.79E-07 | 3.94E-06 |
| RAB27A | 1489.089402 | 0.600680148 | 0.122732549 | 1.98E-07 | 2.86E-06 |
| FERMT1 | 221.7464096 | -0.600236269 | 0.160231118 | 3.56E-05 | 0.000318152 |
| KCNH5 | 49.66014076 | -0.600074216 | 0.325613051 | 0.009013348 | 0.037358581 |
| NDC80 | 1534.142438 | 0.599917206 | 0.088216621 | 2.20E-12 | 6.69E-11 |
| H2BC15 | 16.88397476 | 0.599617526 | 1.242481669 | 0.01441958 | 0.054651753 |
| TRNV | 58.62170058 | 0.598689156 | 0.287699869 | 0.005545958 | 0.025128038 |
| SFXN5 | 172.8581948 | 0.598551677 | 0.171504164 | 9.08E-05 | 0.000729685 |
| LOC105369767 | 38.4719179 | 0.598322617 | 0.353980354 | 0.011610559 | 0.045883172 |
| RASA3 | 5737.78733 | -0.59798951 | 0.067230897 | 1.29E-19 | 7.74E-18 |
| RPL7 | 37941.64882 | -0.597963107 | 0.068775998 | 1.00E-18 | 5.46E-17 |
| ETV3 | 1418.544795 | -0.597943348 | 0.097654995 | 1.95E-10 | 4.45E-09 |
| MYADM | 2168.887901 | 0.597406928 | 0.101422152 | 8.10E-10 | 1.72E-08 |
| KIFC3 | 13.92532866 | -0.597164784 | 0.675804049 | 0.023410371 | 0.080515867 |
| FCHO1 | 294.1248755 | 0.597157867 | 0.143701339 | 6.38E-06 | 6.69E-05 |
| PRRT3 | 172.9156159 | -0.596552199 | 0.184550308 | 0.000233289 | 0.001676991 |
| UAP1 | 9899.497337 | -0.59633634 | 0.064583688 | 6.20E-21 | 4.11E-19 |
| FABP5P7 | 1227.372696 | -0.596280691 | 0.093337267 | 3.56E-11 | 9.11E-10 |
| CALU | 11819.49905 | -0.59603515 | 0.075570105 | 6.98E-16 | 3.16E-14 |
| CTDSP2 | 8362.940958 | 0.595949123 | 0.082480219 | 1.08E-13 | 3.84E-12 |
| MEGF9 | 508.2800856 | 0.595652377 | 0.115269981 | 4.85E-08 | 7.79E-07 |

| | | | | | |
|---|---|---|---|---|---|
| PROB1 | 195.8437303 | -0.595547885 | 0.158425132 | 3.34E-05 | 0.000300499 |
| ZNF45 | 249.5812832 | 0.595534881 | 0.148354477 | 1.18E-05 | 0.000117772 |
| SPRED1 | 285.7952003 | 0.595116799 | 0.232007191 | 0.001724724 | 0.009481602 |
| P2RX1 | 709.0316524 | -0.5948032 | 0.135714155 | 2.46E-06 | 2.83E-05 |
| CNN2 | 3353.924233 | 0.594551334 | 0.075598946 | 8.07E-16 | 3.61E-14 |
| LINC00674 | 396.8267955 | 0.593929079 | 0.117505998 | 8.95E-08 | 1.37E-06 |
| ZNF875 | 189.0497234 | 0.593876367 | 0.157852372 | 3.31E-05 | 0.000298255 |
| LRRC14B | 2591.050494 | -0.593734933 | 0.094864078 | 8.60E-11 | 2.07E-09 |
| ZSCAN30 | 128.3055647 | -0.592879653 | 0.20713248 | 0.000758607 | 0.004696748 |
| SEC13 | 3909.481839 | -0.592426962 | 0.076555146 | 2.28E-15 | 9.60E-14 |
| ZNF718 | 165.171746 | 0.59210907 | 0.208283433 | 0.000794537 | 0.004887745 |
| ITM2B | 7588.00706 | 0.591953867 | 0.080314831 | 3.70E-14 | 1.39E-12 |
| CRELD1 | 256.7297659 | -0.591807349 | 0.153600125 | 2.39E-05 | 0.000223197 |
| ZNF320 | 293.4369442 | 0.591739955 | 0.166464007 | 7.25E-05 | 0.000599108 |
| MEIS1 | 93.73243186 | 0.591076159 | 0.242776985 | 0.002468684 | 0.012830194 |
| MZB1 | 16988.37014 | -0.590818151 | 0.077896808 | 7.46E-15 | 2.96E-13 |
| BMP6 | 695.9828256 | 0.590159202 | 0.106540989 | 6.47E-09 | 1.19E-07 |
| RPS6KA5 | 237.8746427 | 0.590045797 | 0.165469362 | 7.09E-05 | 0.000587179 |
| PIK3C2A | 1277.090507 | 0.589942632 | 0.119756593 | 1.74E-07 | 2.53E-06 |
| ZNF83 | 185.5927609 | 0.58984662 | 0.164616022 | 6.63E-05 | 0.000554225 |
| FDXACB1 | 2474.725752 | 0.589675068 | 0.073201811 | 1.74E-16 | 8.13E-15 |
| LPIN3 | 139.0714 | -0.589118498 | 0.181979836 | 0.000234316 | 0.00168367 |
| NDFIP2 | 1457.5169 | -0.589109203 | 0.138324631 | 4.27E-06 | 4.67E-05 |
| HLA-DMA | 1524.801809 | -0.588565575 | 0.09702215 | 2.92E-10 | 6.55E-09 |
| MX2 | 14.39642667 | 0.58828463 | 0.648618295 | 0.023516851 | 0.080849973 |
| LERFS | 76.71217103 | 0.587951991 | 0.265306837 | 0.004311297 | 0.020337436 |
| SLC39A8 | 4030.180627 | -0.587836719 | 0.092469096 | 4.53E-11 | 1.14E-09 |
| ZNF816 | 198.4812272 | 0.587817181 | 0.160233524 | 4.82E-05 | 0.000415314 |
| ESPNL | 22.55155725 | -0.587783877 | 0.501027367 | 0.022564763 | 0.078370237 |
| SP4 | 618.5251884 | 0.587708194 | 0.167079877 | 8.35E-05 | 0.000677786 |
| LOC124900954 | 408.8294932 | 0.587680843 | 0.118721689 | 1.57E-07 | 2.31E-06 |
| PPP1R18 | 732.4404416 | 0.587496195 | 0.102766312 | 2.34E-09 | 4.68E-08 |
| DDAH2 | 272.2816518 | 0.587210073 | 0.144562688 | 1.00E-05 | 0.00010099 |
| DHRS1 | 200.7897167 | -0.586976846 | 0.154826692 | 3.02E-05 | 0.000274538 |
| DDR1 | 52.50127481 | 0.586766892 | 0.315700831 | 0.009017299 | 0.037365498 |
| PELI1 | 3176.584135 | -0.58633019 | 0.106330905 | 7.62E-09 | 1.38E-07 |
| BMERB1 | 55.38191436 | -0.586248128 | 0.294264743 | 0.007075402 | 0.030738768 |
| YPEL5 | 1720.231986 | -0.586204583 | 0.101388275 | 1.67E-09 | 3.38E-08 |
| TMEM154 | 349.8676748 | 0.586166145 | 0.13962911 | 5.48E-06 | 5.85E-05 |
| CD68 | 42.81631672 | -0.585558882 | 0.33275363 | 0.011121917 | 0.044397934 |
| TANC2 | 178.3021502 | -0.585524471 | 0.168759185 | 0.000104301 | 0.000825873 |
| CDC20 | 2328.652119 | 0.585297207 | 0.082360543 | 2.64E-13 | 8.98E-12 |
| PNPLA2 | 714.6467113 | 0.585140482 | 0.131808921 | 1.89E-06 | 2.24E-05 |
| PADI2 | 22.64192038 | 0.584174985 | 0.517316216 | 0.02269298 | 0.07867354 |
| PREB | 3875.955511 | -0.582990457 | 0.080624595 | 1.10E-13 | 3.92E-12 |
| CYP51A1 | 13257.18068 | -0.582806968 | 0.081691918 | 2.34E-13 | 8.04E-12 |
| NME2P1 | 103.209747 | 0.582339804 | 0.248036138 | 0.003195378 | 0.015941907 |
| ZNF611 | 147.8970172 | 0.58211637 | 0.213423755 | 0.001140812 | 0.006658389 |
| S100A6 | 7695.082105 | -0.581699225 | 0.079811583 | 7.13E-14 | 2.58E-12 |
| RRAD | 9.925369887 | -0.58168079 | 1.09249033 | 0.018034471 | 0.065321677 |

| | | | | | |
|---|---|---|---|---|---|
| MFSD12 | 605.3016902 | -0.581275382 | 0.125345622 | 7.76E-07 | 9.99E-06 |
| GPR137 | 351.8093685 | 0.581160019 | 0.146599592 | 1.54E-05 | 0.000149456 |
| TOX2 | 358.9196974 | 0.580821895 | 0.147436885 | 1.68E-05 | 0.00016262 |
| CTSB | 3698.763941 | -0.580244692 | 0.079131496 | 5.13E-14 | 1.91E-12 |
| GFI1B | 29.91233332 | 0.580119348 | 0.427498348 | 0.019322224 | 0.068998897 |
| ARHGEF2 | 5474.746308 | 0.579968337 | 0.085076227 | 2.07E-12 | 6.33E-11 |
| PHGDH | 2067.690166 | 0.579381386 | 0.089444565 | 2.10E-11 | 5.57E-10 |
| TRAM2 | 4478.856892 | 0.579354125 | 0.080770332 | 1.72E-13 | 5.99E-12 |
| PLEKHG2 | 1755.98317 | -0.579253491 | 0.101483052 | 2.70E-09 | 5.32E-08 |
| ITFG1 | 1528.610044 | -0.579134276 | 0.086013392 | 3.75E-12 | 1.11E-10 |
| ZNF71 | 94.43044057 | 0.579063864 | 0.25160136 | 0.003639384 | 0.017693209 |
| ZNF8 | 105.7544505 | 0.578579824 | 0.21036063 | 0.001103634 | 0.006474129 |
| EEF1A1P4 | 5530.446663 | -0.578321978 | 0.09054296 | 3.81E-11 | 9.69E-10 |
| FLI1 | 1969.190889 | 0.578282639 | 0.099862173 | 1.54E-09 | 3.14E-08 |
| KRT10-AS1 | 62.06232814 | -0.578260108 | 0.288410703 | 0.007119239 | 0.030913705 |
| FABP5P1 | 289.8088471 | -0.578101798 | 0.147707941 | 1.88E-05 | 0.000179817 |
| RAB29 | 1145.305601 | 0.577846323 | 0.094634121 | 2.28E-10 | 5.15E-09 |
| TRIM56 | 370.9047892 | 0.57778041 | 0.172121106 | 0.000154237 | 0.001161772 |
| MEAK7 | 190.9577269 | -0.577534005 | 0.170524107 | 0.000141765 | 0.001077203 |
| CYP4V2 | 85.25316933 | 0.577409472 | 0.255199755 | 0.00395288 | 0.018904175 |
| RPS2P29 | 581.4933596 | -0.5773383 | 0.158399122 | 5.62E-05 | 0.000477811 |
| LINC02381 | 51.31118189 | -0.577114516 | 0.315809796 | 0.010060405 | 0.041009161 |
| KYNU | 13.60299061 | 0.577063925 | 0.76162895 | 0.022727824 | 0.078778569 |
| RPL3P4 | 5228.254634 | -0.576890245 | 0.087587264 | 9.50E-12 | 2.68E-10 |
| B9D1 | 531.8769201 | -0.576591655 | 0.111874274 | 5.77E-08 | 9.09E-07 |
| MFSD9 | 375.2543041 | 0.576191181 | 0.119038849 | 2.82E-07 | 3.97E-06 |
| DMAC1 | 602.7144407 | -0.575979331 | 0.126932205 | 1.27E-06 | 1.55E-05 |
| IGFLR1 | 294.7135023 | -0.575773528 | 0.175108113 | 0.000206305 | 0.001501718 |
| NXF1 | 1614.283676 | -0.575660746 | 0.086187718 | 5.47E-12 | 1.59E-10 |
| ABHD17C | 340.1515182 | 0.575633514 | 0.13086798 | 2.32E-06 | 2.70E-05 |
| TOMM7 | 5336.405709 | -0.575450882 | 0.079813322 | 1.35E-13 | 4.78E-12 |
| GLS | 10225.22791 | -0.575428349 | 0.114782182 | 1.19E-07 | 1.79E-06 |
| SELPLG | 7966.310814 | 0.575122505 | 0.073055518 | 1.30E-15 | 5.64E-14 |
| LOC105370203 | 17.43015714 | -0.575019563 | 0.596574989 | 0.025436942 | 0.086047063 |
| TMEM208 | 1758.890484 | -0.574941753 | 0.087464258 | 1.15E-11 | 3.20E-10 |
| ZNF706 | 1765.052793 | -0.574480873 | 0.077028275 | 2.05E-14 | 7.88E-13 |
| PERP | 2361.464688 | -0.574365869 | 0.086369388 | 6.78E-12 | 1.93E-10 |
| TMEM170A | 662.494323 | -0.574289219 | 0.130341348 | 2.26E-06 | 2.63E-05 |
| SIPA1L1 | 1067.47076 | 0.574144925 | 0.08781674 | 1.41E-11 | 3.90E-10 |
| ZNF254 | 16.00016854 | -0.574057357 | 0.679485148 | 0.024910516 | 0.084616597 |
| APLP1 | 119.0655319 | -0.573248649 | 0.202141038 | 0.000896406 | 0.005408732 |
| FURIN | 726.1943676 | 0.572881239 | 0.118782454 | 3.11E-07 | 4.35E-06 |
| YEATS2 | 709.1150157 | 0.572504181 | 0.095604933 | 4.82E-10 | 1.06E-08 |
| TMOD1 | 53.30461617 | 0.572357644 | 0.307110158 | 0.009443053 | 0.038897561 |
| ARL6 | 244.7631413 | 0.572340188 | 0.193502684 | 0.000597983 | 0.003818315 |
| LOC100507002 | 82.67152134 | 0.572051847 | 0.243815919 | 0.003355512 | 0.01657916 |
| MIR7847 | 412.7006291 | 0.571895333 | 0.145422731 | 1.76E-05 | 0.000169546 |
| ZNF226 | 83.79146882 | 0.571624376 | 0.238512738 | 0.002915167 | 0.014722062 |
| CCDC57 | 396.787178 | -0.571209123 | 0.141215835 | 1.15E-05 | 0.000115427 |
| NEK2 | 956.3408724 | 0.571080321 | 0.101923396 | 4.72E-09 | 8.92E-08 |

| | | | | | |
|---|---|---|---|---|---|
| KIF18B | 566.4086506 | 0.5706581 | 0.111553729 | 6.92E-08 | 1.08E-06 |
| ZNF629 | 78.43092738 | -0.570640715 | 0.242397195 | 0.003314605 | 0.016399721 |
| TNRC6C | 146.0623745 | 0.57038666 | 0.187943126 | 0.000475314 | 0.003122538 |
| MOGS | 1759.744549 | -0.570349348 | 0.104916729 | 1.24E-08 | 2.16E-07 |
| TMSB4XP4 | 742.9418285 | 0.570325587 | 0.11062229 | 5.62E-08 | 8.90E-07 |
| ZFP64 | 499.0753667 | 0.570244118 | 0.111984483 | 7.96E-08 | 1.23E-06 |
| DISP3 | 9.380011966 | 0.5700232 | 1.038455065 | 0.018084382 | 0.065465065 |
| CORO7-PAM16 | 1056.884523 | -0.56992207 | 0.134051736 | 4.69E-06 | 5.09E-05 |
| SUCLG2 | 2054.178545 | -0.569909892 | 0.096282473 | 7.41E-10 | 1.59E-08 |
| RNF122 | 933.3536288 | -0.56990888 | 0.099671115 | 2.50E-09 | 4.95E-08 |
| MC1R | 46.74367261 | -0.56918068 | 0.341848185 | 0.013588594 | 0.052210713 |
| EPM2AIP1 | 310.879188 | -0.569141839 | 0.147961922 | 2.53E-05 | 0.000234708 |
| PABPC1 | 36181.08286 | -0.568830858 | 0.071989674 | 6.46E-16 | 2.95E-14 |
| TCF19 | 728.3453318 | 0.568512497 | 0.104698665 | 1.29E-08 | 2.25E-07 |
| CLIP1 | 251.4600707 | 0.568102782 | 0.153591512 | 4.55E-05 | 0.000395362 |
| SERTAD3 | 203.8431677 | -0.567932102 | 0.163296731 | 0.000107495 | 0.000846514 |
| KIF14 | 618.6623333 | 0.567615258 | 0.119038878 | 4.06E-07 | 5.52E-06 |
| KHDC4 | 351.8266085 | -0.567593617 | 0.132078308 | 3.81E-06 | 4.21E-05 |
| NEIL3 | 176.7130955 | 0.567366676 | 0.193730478 | 0.000660833 | 0.004159816 |
| GNA13 | 7875.731471 | -0.567044307 | 0.124660234 | 1.21E-06 | 1.50E-05 |
| UCK1 | 512.2503222 | 0.566976038 | 0.110012984 | 5.79E-08 | 9.11E-07 |
| KRT7 | 36.40034562 | -0.566835326 | 0.375458089 | 0.017407595 | 0.063475957 |
| VDR | 1255.56403 | 0.566632985 | 0.089368264 | 5.32E-11 | 1.32E-09 |
| NIPSNAP3B | 15.39639638 | -0.566447121 | 0.666867649 | 0.025597994 | 0.086426295 |
| JDP2 | 30.10745717 | 0.566007681 | 0.391053467 | 0.018656575 | 0.067014294 |
| NCOA3 | 7543.689957 | 0.565823025 | 0.07415169 | 5.49E-15 | 2.23E-13 |
| NAT9 | 339.0439409 | -0.565661703 | 0.128846487 | 2.58E-06 | 2.97E-05 |
| NUDT9 | 332.5801136 | -0.565457054 | 0.133765121 | 5.26E-06 | 5.63E-05 |
| ARHGEF19 | 92.69048149 | 0.565452603 | 0.225350518 | 0.002265808 | 0.011937026 |
| ZNF235 | 41.11604411 | 0.565437826 | 0.369129157 | 0.016538447 | 0.060741523 |
| KRT8P12 | 33.3003771 | 0.565428758 | 0.401586557 | 0.019565743 | 0.069658711 |
| CYHR1 | 140.5055933 | 0.565295456 | 0.199050619 | 0.000882858 | 0.005330703 |
| LOC105372733 | 78.62769244 | -0.565118659 | 0.240344136 | 0.003421019 | 0.016830051 |
| CA13 | 49.67777128 | -0.565058579 | 0.35913508 | 0.015919749 | 0.059045256 |
| DTNB-AS1 | 63.2856822 | 0.564933551 | 0.290095237 | 0.00830208 | 0.034937334 |
| SLC12A4 | 830.7047855 | -0.56476384 | 0.114910185 | 2.06E-07 | 2.96E-06 |
| VPS37B | 1527.894549 | 0.564608932 | 0.088268008 | 3.74E-11 | 9.54E-10 |
| FABP5P10 | 45.45279351 | -0.564605594 | 0.326147435 | 0.012697689 | 0.049312823 |
| RPL29P26 | 2379.052456 | -0.564545419 | 0.108016391 | 4.06E-08 | 6.62E-07 |
| RNR1 | 42537.64931 | 0.564263869 | 1.225678633 | 0.013352008 | 0.051507401 |
| CHAC1 | 938.9244302 | -0.563526258 | 0.122412147 | 9.58E-07 | 1.21E-05 |
| GM2A | 3111.283882 | -0.563052458 | 0.075721413 | 2.52E-14 | 9.67E-13 |
| FABP5P11 | 99.92680221 | -0.563007744 | 0.221938165 | 0.002161603 | 0.011478832 |
| CHCHD7 | 1048.251821 | -0.562716878 | 0.08852938 | 4.91E-11 | 1.22E-09 |
| PAK4 | 404.6310018 | 0.562384976 | 0.131208503 | 4.08E-06 | 4.48E-05 |
| ZNF571 | 46.59960505 | 0.562153018 | 0.320221753 | 0.011875526 | 0.046664053 |
| EME1 | 128.2197791 | 0.562021264 | 0.198758677 | 0.000920448 | 0.005540283 |
| KRT81 | 252.8692762 | 0.561973462 | 0.160417442 | 9.79E-05 | 0.000781236 |
| ARHGAP15 | 708.2240825 | -0.561944635 | 0.109460144 | 6.53E-08 | 1.02E-06 |
| PALD1 | 432.1547396 | -0.561581246 | 0.122041646 | 9.74E-07 | 1.23E-05 |

| | | | | | |
|---|---|---|---|---|---|
| S100A4 | 9283.570238 | -0.560661699 | 0.069027011 | 1.13E-16 | 5.32E-15 |
| SYNPO | 79.3518969 | -0.560210732 | 0.239189802 | 0.003590439 | 0.017504403 |
| IDH1 | 1133.942288 | 0.560148316 | 0.096478939 | 1.51E-09 | 3.08E-08 |
| KCNJ11 | 37.19370018 | 0.560077373 | 0.388678946 | 0.018951391 | 0.067834071 |
| GRSF1 | 2989.697037 | -0.559947412 | 0.078527158 | 2.48E-13 | 8.48E-12 |
| CCP110 | 387.6040976 | -0.55973737 | 0.14394701 | 2.22E-05 | 0.000208134 |
| TRPS1 | 90.51889226 | -0.559658459 | 0.243403363 | 0.003930376 | 0.01883296 |
| EDEM1 | 5672.597231 | -0.559540554 | 0.075467898 | 2.98E-14 | 1.12E-12 |
| MDM4 | 111.5653972 | 0.559226725 | 0.218537877 | 0.001999394 | 0.010722669 |
| ERLIN2 | 1218.162856 | -0.55884989 | 0.087200317 | 3.52E-11 | 9.04E-10 |
| LOC102725101 | 1522.657802 | -0.558834576 | 0.12857321 | 3.12E-06 | 3.52E-05 |
| LOC112268316 | 1522.206104 | -0.558705482 | 0.128559071 | 3.13E-06 | 3.52E-05 |
| PDIA3P1 | 1178.923997 | -0.558492715 | 0.094651768 | 8.81E-10 | 1.86E-08 |
| H3-3B | 17064.23059 | -0.558387013 | 0.067498348 | 3.24E-17 | 1.58E-15 |
| C6orf223 | 6.185137994 | 0.558249692 | 2.32125337 | 0.009094683 | 0.03764164 |
| C12orf75 | 2404.562392 | 0.558154024 | 0.075198003 | 2.79E-14 | 1.06E-12 |
| FOXO1 | 1747.442779 | 0.557816672 | 0.095826645 | 1.37E-09 | 2.83E-08 |
| PFKP | 4876.347007 | 0.557544549 | 0.073876441 | 1.07E-14 | 4.24E-13 |
| THEM6 | 304.229368 | 0.557310048 | 0.160994786 | 0.000116382 | 0.000907829 |
| TICAM2 | 83.70619978 | -0.557093438 | 0.235586442 | 0.003368794 | 0.016626815 |
| TESK1 | 248.607665 | 0.556666771 | 0.175455556 | 0.000316271 | 0.002183152 |
| MIR10527 | 27.7970013 | -0.556494093 | 0.489791729 | 0.026129897 | 0.087777191 |
| FAHD1 | 381.7325806 | -0.55632541 | 0.121108115 | 1.01E-06 | 1.27E-05 |
| PRKX | 854.0147083 | -0.555805669 | 0.093466629 | 6.55E-10 | 1.41E-08 |
| DEK | 1396.022637 | 0.555406061 | 0.11534759 | 3.39E-07 | 4.69E-06 |
| PLAAT2 | 478.1678296 | 0.555394718 | 0.12382618 | 1.66E-06 | 2.00E-05 |
| ZNF460 | 118.8134877 | 0.555316045 | 0.239225001 | 0.003728474 | 0.017999996 |
| SURF2 | 470.0101729 | 0.555191329 | 0.118754504 | 6.78E-07 | 8.84E-06 |
| RPS2P55 | 4309.146808 | -0.555131344 | 0.087627459 | 5.73E-11 | 1.41E-09 |
| WNT5B | 554.2763266 | 0.555010662 | 0.117783887 | 5.65E-07 | 7.49E-06 |
| RPS27 | 60699.295 | -0.555005024 | 0.071603515 | 2.26E-15 | 9.53E-14 |
| CORO7 | 1246.616482 | -0.554934338 | 0.126107326 | 2.52E-06 | 2.90E-05 |
| ZBTB20 | 74.62975806 | 0.554812039 | 0.302035785 | 0.010427664 | 0.042210344 |
| LRP1 | 282.5405782 | -0.554411394 | 0.143991488 | 2.67E-05 | 0.000246368 |
| IGKJ2 | 10550.91519 | -0.554359544 | 0.239740768 | 0.003877359 | 0.018630472 |
| HSPBAP1 | 163.1693728 | 0.554333962 | 0.171880004 | 0.000268062 | 0.001887771 |
| ZNF587B | 69.77442515 | 0.554243101 | 0.263583876 | 0.006235451 | 0.027679905 |
| CORO1A | 20798.72046 | 0.554168374 | 0.098481015 | 4.24E-09 | 8.08E-08 |
| WDR5B | 56.12341755 | 0.553996592 | 0.281320516 | 0.008328166 | 0.035021574 |
| MTRR | 3399.017651 | 0.553928799 | 0.091400497 | 3.19E-10 | 7.13E-09 |
| CRIP1 | 1700.132389 | 0.553843592 | 0.106358294 | 4.52E-08 | 7.29E-07 |
| SMARCC2 | 1405.945153 | 0.553438972 | 0.093210727 | 6.89E-10 | 1.48E-08 |
| BBS9 | 272.762986 | -0.553051508 | 0.140870052 | 1.99E-05 | 0.000188725 |
| G3BP2 | 6276.32925 | -0.553012548 | 0.098885677 | 5.31E-09 | 9.85E-08 |
| SPRR2B | 26.52510298 | 0.552826715 | 0.423077077 | 0.022856309 | 0.079081458 |
| VEGFA | 1111.999514 | -0.551890662 | 0.096646292 | 2.73E-09 | 5.37E-08 |
| KC6 | 34.06759246 | -0.551551996 | 0.384059881 | 0.020286913 | 0.07173953 |
| ZNF462 | 13.1046982 | 0.551489514 | 0.692377594 | 0.025565387 | 0.086349875 |
| RPS2P12 | 632.0174741 | -0.551381988 | 0.166811443 | 0.000209508 | 0.001521832 |
| LOXL1-AS1 | 53.71745646 | 0.551129919 | 0.298938193 | 0.010554117 | 0.042612649 |

| | | | | | |
|---|---|---|---|---|---|
| IGKV3-20 | 96725.19326 | -0.5509607 | 0.080882552 | 2.32E-12 | 7.02E-11 |
| RPS15AP38 | 1395.659248 | -0.550918206 | 0.087090843 | 6.12E-11 | 1.50E-09 |
| DBIL5P | 38.32652515 | 0.550891235 | 0.365279152 | 0.018091491 | 0.065465065 |
| ERBB2 | 57.72938098 | 0.55072343 | 0.279962714 | 0.008377083 | 0.035193097 |
| HIGD1C | 386.2299929 | 0.55047302 | 0.128862745 | 4.47E-06 | 4.87E-05 |
| RESF1 | 1213.156276 | 0.550332632 | 0.09384141 | 1.08E-09 | 2.24E-08 |
| NAA80 | 54.01204133 | -0.549841541 | 0.299197393 | 0.010994847 | 0.04399209 |
| LOC124900458 | 24.62602564 | -0.549700066 | 0.505220766 | 0.026450098 | 0.08873234 |
| NECTIN3 | 138.8635714 | 0.549155103 | 0.224513332 | 0.002782998 | 0.014186983 |
| SLCO4A1-AS2 | 98.89398172 | 0.548986151 | 0.217632178 | 0.002312671 | 0.012143277 |
| RPL3P7 | 392.8677013 | -0.548966665 | 0.141061556 | 2.27E-05 | 0.000212411 |
| RPL14 | 30376.55016 | -0.548906006 | 0.062537234 | 4.93E-19 | 2.78E-17 |
| TTC8 | 151.1252325 | -0.548880017 | 0.173280745 | 0.000335984 | 0.002303338 |
| SHISA3 | 14.78611044 | 0.548859381 | 0.638360529 | 0.026555435 | 0.089033971 |
| SRGAP2B | 216.1337153 | 0.548844709 | 0.165726376 | 0.000201104 | 0.001468944 |
| TTC32 | 860.3397049 | 0.548426001 | 0.134177477 | 9.94E-06 | 0.000100388 |
| FAM227B | 35.89554873 | -0.548213239 | 0.372436036 | 0.019501209 | 0.069443231 |
| NTRK1 | 28.88751146 | -0.547577632 | 0.433323694 | 0.024290287 | 0.0829164 |
| PCGF5 | 1738.192007 | 0.547251697 | 0.151663674 | 6.88E-05 | 0.000572089 |
| PTDSS2 | 388.9467019 | -0.547044532 | 0.141492298 | 2.59E-05 | 0.000239699 |
| ATP6V1G2 | 19.17236609 | -0.54695609 | 0.520996542 | 0.028006429 | 0.092749538 |
| AURKA | 2667.359364 | 0.546799724 | 0.080952709 | 3.56E-12 | 1.05E-10 |
| TMEM106A | 120.1497992 | -0.546797586 | 0.218350703 | 0.002463028 | 0.01281233 |
| KNL1 | 435.0802575 | 0.54677466 | 0.138008863 | 1.68E-05 | 0.000162712 |
| MYO1F | 546.0334604 | 0.546608371 | 0.110410756 | 1.79E-07 | 2.60E-06 |
| FKTN | 542.2317564 | -0.546566653 | 0.121228255 | 1.54E-06 | 1.86E-05 |
| CTSO | 113.1724826 | 0.546127611 | 0.211552309 | 0.001982974 | 0.010661027 |
| DNLZ | 218.0269549 | 0.54608569 | 0.187782587 | 0.00076609 | 0.004737991 |
| FAM81A | 134.2518978 | -0.546045012 | 0.192878042 | 0.000981515 | 0.005852927 |
| EEF1A1P12 | 4429.394256 | -0.545898715 | 0.101031869 | 1.59E-08 | 2.73E-07 |
| NRROS | 3691.1547 | 0.545854756 | 0.104657547 | 4.47E-08 | 7.21E-07 |
| TRIM14 | 714.1684134 | 0.545501054 | 0.101628431 | 1.95E-08 | 3.32E-07 |
| ZNF136 | 249.2948571 | 0.544807086 | 0.152251855 | 7.71E-05 | 0.000632724 |
| LINC00963 | 116.0750331 | 0.544693585 | 0.214862944 | 0.002276999 | 0.011985044 |
| APRT | 3634.244898 | -0.544582583 | 0.105817871 | 6.50E-08 | 1.02E-06 |
| MAST4 | 171.2791455 | 0.543996338 | 0.18491947 | 0.000695468 | 0.004349377 |
| BLVRA | 671.6274366 | 0.543979756 | 0.097721598 | 6.42E-09 | 1.18E-07 |
| ETFBKMT | 33.05941193 | 0.543957329 | 0.425992725 | 0.02385787 | 0.081730313 |
| PPP1R16B | 744.7832979 | 0.543910587 | 0.096039351 | 3.65E-09 | 7.03E-08 |
| ADAM8 | 745.397412 | 0.543440824 | 0.131325424 | 8.21E-06 | 8.43E-05 |
| SEPTIN11 | 4474.4132 | -0.543191218 | 0.073631741 | 4.07E-14 | 1.53E-12 |
| TMEM19 | 810.8206158 | 0.543159653 | 0.109208142 | 1.58E-07 | 2.31E-06 |
| PYROXD2 | 176.3571166 | -0.542792283 | 0.172296732 | 0.000363633 | 0.002466525 |
| PLXNA3 | 102.6045868 | -0.542781996 | 0.225140891 | 0.003204323 | 0.015974326 |
| ARHGAP31 | 494.1563718 | 0.54277525 | 0.119577265 | 1.34E-06 | 1.64E-05 |
| LRRC27 | 28.93701723 | -0.542517506 | 0.415734333 | 0.024019909 | 0.08217776 |
| XYLT1 | 796.8134747 | -0.542372485 | 0.097706379 | 7.04E-09 | 1.28E-07 |
| BBLN | 1090.549447 | 0.54207629 | 0.156718687 | 0.000126182 | 0.000975056 |
| CSRP2 | 23.98943504 | 0.541947816 | 0.451231 | 0.025792298 | 0.086896642 |
| COX5AP1 | 20.54390901 | 0.541914827 | 0.681962877 | 0.02551447 | 0.086228352 |

| | | | | | |
|---|---|---|---|---|---|
| MAP1LC3B | 4375.174495 | -0.541862879 | 0.075083028 | 1.37E-13 | 4.82E-12 |
| GOT2 | 6934.858223 | -0.541789741 | 0.068743578 | 8.40E-16 | 3.74E-14 |
| RNF157 | 112.7512792 | -0.541715167 | 0.220653634 | 0.00290179 | 0.014677475 |
| ODC1 | 42714.64366 | -0.541625939 | 0.06121239 | 3.09E-19 | 1.78E-17 |
| SDAD1 | 1295.445028 | -0.541624678 | 0.083314458 | 2.05E-11 | 5.46E-10 |
| LIPH | 174.5384569 | -0.541374342 | 0.178431496 | 0.000528377 | 0.003421805 |
| CALML4 | 70.65882146 | -0.540763399 | 0.274192962 | 0.008656567 | 0.036130581 |
| CXXC5 | 1454.696236 | 0.540329281 | 0.082586676 | 1.53E-11 | 4.21E-10 |
| IGKV3-11 | 5001.153509 | -0.540314416 | 0.098505993 | 1.05E-08 | 1.86E-07 |
| FUT1 | 32.85703673 | 0.540220702 | 0.398706138 | 0.022831287 | 0.079026464 |
| PALM2AKAP2 | 3478.791016 | 0.540212479 | 0.103833188 | 4.85E-08 | 7.79E-07 |
| GAA | 1890.259599 | -0.540196814 | 0.108279258 | 1.51E-07 | 2.23E-06 |
| RBKS | 103.7280473 | -0.539965622 | 0.206717237 | 0.001893945 | 0.010258821 |
| REL | 253.8557455 | -0.539781696 | 0.173401142 | 0.000410734 | 0.002749395 |
| PDIA3 | 12638.76885 | -0.539521409 | 0.060794143 | 1.95E-19 | 1.15E-17 |
| HERC5 | 100.6441527 | -0.539449593 | 0.215124748 | 0.002527796 | 0.013090266 |
| OGFOD1 | 1565.141333 | -0.538734385 | 0.083082416 | 2.27E-11 | 5.97E-10 |
| ZFAND5 | 2942.888067 | -0.538714102 | 0.08951863 | 4.47E-10 | 9.90E-09 |
| RGS19 | 445.507386 | 0.53870663 | 0.144804198 | 4.68E-05 | 0.000405146 |
| TPD52L1 | 39.39427205 | 0.53863589 | 0.338978199 | 0.017036167 | 0.062318401 |
| PHKB | 1191.326212 | -0.538518745 | 0.093627934 | 2.25E-09 | 4.51E-08 |
| ARHGAP45 | 1963.18216 | 0.538446445 | 0.112906531 | 4.38E-07 | 5.92E-06 |
| SMCO2 | 23.63346984 | 0.538295221 | 0.46156788 | 0.02671832 | 0.089406984 |
| RPS20 | 30356.53221 | -0.538293516 | 0.074467073 | 1.27E-13 | 4.51E-12 |
| MCTP2 | 723.5745405 | 0.538237802 | 0.106127904 | 9.67E-08 | 1.47E-06 |
| TSHZ1 | 321.9984907 | 0.538205725 | 0.127472969 | 5.78E-06 | 6.15E-05 |
| MRPS18C | 789.2560066 | -0.538077948 | 0.096379659 | 5.99E-09 | 1.10E-07 |
| ZNF396 | 15.27693504 | 0.537971516 | 0.604260152 | 0.028412556 | 0.093771723 |
| RORA | 588.3637509 | 0.537927446 | 0.174201823 | 0.000437005 | 0.002904878 |
| ZNF232 | 226.6115859 | 0.537433085 | 0.148241819 | 6.71E-05 | 0.000560035 |
| TUFM | 10709.86573 | -0.53715568 | 0.071309441 | 1.30E-14 | 5.05E-13 |
| ZFHX3 | 110.9929989 | 0.537134501 | 0.211163843 | 0.002243788 | 0.011839146 |
| TMEM70 | 1582.686689 | -0.536950145 | 0.079604399 | 3.93E-12 | 1.16E-10 |
| ATP6V1H | 1052.199441 | -0.536344541 | 0.088374162 | 3.33E-10 | 7.42E-09 |
| EPS8L1 | 99.21540164 | 0.535589894 | 0.218713079 | 0.002945814 | 0.014846526 |
| NAGK | 1027.685445 | 0.535433334 | 0.09927408 | 1.74E-08 | 2.98E-07 |
| ZNF567 | 216.3341734 | 0.535289521 | 0.167542853 | 0.00031479 | 0.002174402 |
| PAPSS2 | 361.0020827 | 0.535219997 | 0.132992191 | 1.36E-05 | 0.000133934 |
| SLX1B | 295.2961644 | 0.53521102 | 0.17371849 | 0.000463857 | 0.003061212 |
| SLX1A | 295.7545634 | 0.534939581 | 0.174472284 | 0.000487248 | 0.003186436 |
| WHRN | 325.5431199 | -0.534921285 | 0.135497597 | 1.90E-05 | 0.000181368 |
| MFSD13A | 137.5151728 | -0.534564868 | 0.197851578 | 0.001513816 | 0.008467296 |
| DOLPP1 | 998.5222512 | 0.53429002 | 0.102886191 | 5.21E-08 | 8.29E-07 |
| PHF19 | 3420.269221 | 0.534278073 | 0.095811995 | 6.22E-09 | 1.15E-07 |
| PCGF3 | 335.6336231 | 0.534238956 | 0.131781378 | 1.22E-05 | 0.000121524 |
| MIR4721 | 257.3044405 | -0.534076525 | 0.210408986 | 0.002378065 | 0.012429481 |
| CYTH4 | 44.25533909 | 0.534020953 | 0.325949168 | 0.016131202 | 0.059625192 |
| MTRF1 | 245.642904 | 0.533809477 | 0.139598558 | 3.14E-05 | 0.000284671 |
| HSDL1 | 1494.042687 | -0.533333745 | 0.098800778 | 1.71E-08 | 2.94E-07 |
| SLC44A2 | 53.57411793 | -0.53322222 | 0.296928223 | 0.012468688 | 0.048600648 |

| | | | | | |
|---|---|---|---|---|---|
| HELQ | 260.4746837 | -0.532802613 | 0.138695752 | 2.95E-05 | 0.000269127 |
| PTPRR | 87.90000464 | 0.53232688 | 0.239153658 | 0.005081743 | 0.02339196 |
| HSD17B7P2 | 251.5700453 | -0.531536188 | 0.151641205 | 0.000109878 | 0.000862975 |
| BBC3 | 660.1906889 | -0.531281277 | 0.144015551 | 5.47E-05 | 0.000466769 |
| LINC00641 | 216.5697131 | -0.530911002 | 0.153896395 | 0.000131878 | 0.001013194 |
| TUBA4B | 78.47552715 | -0.530904986 | 0.251358066 | 0.006853094 | 0.029938199 |
| CPQ | 18.52722013 | 0.530304526 | 0.583563666 | 0.029114923 | 0.095634128 |
| ZNF264 | 267.7589652 | 0.530284934 | 0.15135732 | 0.000107775 | 0.000848338 |
| ALDH18A1 | 3659.58003 | -0.53025658 | 0.067112255 | 7.47E-16 | 3.36E-14 |
| FAM88B | 44.65010626 | 0.529843293 | 0.321560831 | 0.01629736 | 0.06009825 |
| RASSF3 | 1739.204846 | 0.52977445 | 0.113273467 | 7.25E-07 | 9.42E-06 |
| DLG3 | 975.2083457 | 0.529396451 | 0.088623467 | 5.99E-10 | 1.30E-08 |
| ORAI1 | 200.8934608 | 0.529178669 | 0.2276748 | 0.004079504 | 0.019418574 |
| SLC16A9 | 230.7351715 | -0.528770207 | 0.166741141 | 0.000354367 | 0.002410284 |
| TLE5 | 7343.174497 | 0.528768262 | 0.079096176 | 6.04E-12 | 1.74E-10 |
| USP18 | 29.83531821 | 0.528400277 | 0.389776957 | 0.023999857 | 0.082135478 |
| DHFR2 | 133.0653032 | 0.528398773 | 0.190392604 | 0.001211449 | 0.006992886 |
| MED18 | 83.7956728 | 0.527889777 | 0.247487524 | 0.006378489 | 0.028206407 |
| RPS15A | 28811.9415 | -0.527800245 | 0.070926168 | 2.55E-14 | 9.74E-13 |
| SORT1 | 2995.595199 | 0.527603276 | 0.070907881 | 2.65E-14 | 1.01E-12 |
| RPSAP47 | 1083.858483 | -0.527032796 | 0.089536582 | 1.05E-09 | 2.20E-08 |
| NFATC1 | 196.1270058 | 0.526959455 | 0.1564701 | 0.000181052 | 0.001339862 |
| GRHPR | 609.3857749 | -0.526723366 | 0.11354318 | 9.08E-07 | 1.16E-05 |
| TMEM42 | 84.25735315 | -0.526670937 | 0.243788202 | 0.0061936 | 0.027529406 |
| DAP | 2411.86563 | -0.526580631 | 0.084972752 | 1.54E-10 | 3.58E-09 |
| ZNF441 | 124.7987389 | 0.526363398 | 0.19770699 | 0.0016844 | 0.009289927 |
| MAPKAPK2 | 1343.491791 | 0.526088582 | 0.091978453 | 2.78E-09 | 5.45E-08 |
| GBGT1 | 14.5083336 | 0.52573515 | 0.621647808 | 0.029244232 | 0.095931495 |
| MRPS25 | 2164.321353 | -0.525563812 | 0.079394577 | 9.64E-12 | 2.71E-10 |
| HELB | 135.1620382 | 0.525235262 | 0.192791603 | 0.001440293 | 0.008111072 |
| PRKD2 | 2032.394394 | -0.525102965 | 0.086775623 | 3.76E-10 | 8.35E-09 |
| CASC9 | 281.9428683 | -0.524704195 | 0.173803863 | 0.000585167 | 0.003748924 |
| LOC102724858 | 60.43369717 | -0.524660523 | 0.296837268 | 0.013463783 | 0.051834668 |
| AZGP1 | 29.79900051 | 0.524568315 | 0.395538695 | 0.025154775 | 0.08527902 |
| ARRDC1 | 245.861089 | 0.524481271 | 0.174703906 | 0.000619429 | 0.003937831 |
| ATF5 | 1180.78297 | -0.524306623 | 0.095776705 | 1.16E-08 | 2.05E-07 |
| DYNC2H1 | 14.3689676 | -0.52405557 | 0.662827176 | 0.028811191 | 0.094870394 |
| PRDM1 | 3481.685965 | -0.523769149 | 0.083780105 | 1.03E-10 | 2.45E-09 |
| SRGAP2D | 84.88130141 | 0.523503524 | 0.237944443 | 0.0055777 | 0.025219068 |
| RABAC1 | 2038.514125 | -0.523415391 | 0.134205875 | 2.43E-05 | 0.000225434 |
| KIFC2 | 38.93030605 | -0.523261821 | 0.379098059 | 0.02406347 | 0.082271888 |
| STAP2 | 41.03756402 | -0.523084526 | 0.371219279 | 0.023615037 | 0.081074862 |
| GYG1 | 2881.320945 | -0.522731554 | 0.081293824 | 3.47E-11 | 8.92E-10 |
| MPP7 | 28.25793817 | 0.522666765 | 0.441760954 | 0.028092704 | 0.092911144 |
| ZNF652 | 575.1153274 | 0.522241263 | 0.143894905 | 6.88E-05 | 0.000572089 |
| ZNRF2 | 909.0184476 | 0.522146931 | 0.146541689 | 8.85E-05 | 0.000712252 |
| PGRMC2 | 1603.865361 | -0.522056123 | 0.103269366 | 1.17E-07 | 1.76E-06 |
| ANXA5 | 11038.62175 | -0.52187895 | 0.071133788 | 6.18E-14 | 2.25E-12 |
| TGFBR3L | 24.59855441 | -0.52183043 | 0.471049199 | 0.030403118 | 0.098630722 |
| ZNF467 | 146.1933122 | 0.521707169 | 0.193903089 | 0.001608227 | 0.008931898 |

| | | | | | |
|---|---|---|---|---|---|
| IGKC | 466566.6229 | -0.521280634 | 0.073272349 | 2.94E-13 | 9.91E-12 |
| LOC102724745 | 79.18948327 | 0.520732106 | 0.233805672 | 0.00535264 | 0.024411817 |
| CD44 | 5427.036428 | 0.520656523 | 0.0752931 | 1.26E-12 | 3.99E-11 |
| LINC01943 | 105.8823985 | -0.520628632 | 0.213944802 | 0.003235861 | 0.016092878 |
| NDOR1 | 265.0542873 | 0.520517933 | 0.144750744 | 8.00E-05 | 0.000652909 |
| IGHJ1 | 58.4490431 | -0.520441436 | 0.284397879 | 0.012398607 | 0.048368254 |
| LRRC28 | 378.8690787 | 0.520372292 | 0.118046476 | 2.65E-06 | 3.03E-05 |
| GLUL | 1706.864411 | 0.520268141 | 0.083019965 | 9.90E-11 | 2.36E-09 |
| LOC105376156 | 112.8722919 | -0.520113657 | 0.205959896 | 0.002578742 | 0.013282593 |
| NEUROG2-AS1 | 23.91428153 | -0.520051528 | 0.496441133 | 0.03063696 | 0.09930655 |
| P4HB | 45709.06162 | -0.520036668 | 0.078694573 | 1.08E-11 | 3.02E-10 |
| ZNF446 | 66.17801694 | 0.519967444 | 0.298100688 | 0.014401904 | 0.054620623 |
| HTR2C | 187.7206444 | 0.519818487 | 0.182313656 | 0.001001135 | 0.005954028 |
| CLCN7 | 807.5596281 | -0.519636941 | 0.129337806 | 1.52E-05 | 0.000148216 |
| SLC27A5 | 485.7901713 | -0.519631108 | 0.15163444 | 0.000151636 | 0.001144676 |
| CREM | 236.2287 | -0.519592635 | 0.147962269 | 0.000110332 | 0.000865707 |
| NSMCE2 | 776.9157513 | -0.519497881 | 0.10390985 | 1.51E-07 | 2.23E-06 |
| SPRED2 | 437.6095867 | 0.519493814 | 0.116475297 | 2.09E-06 | 2.46E-05 |
| LYPD3 | 7.048032621 | 0.519473673 | 1.115241101 | 0.015318357 | 0.057256204 |
| RPL29P11 | 1968.28779 | -0.519469118 | 0.080171502 | 2.55E-11 | 6.67E-10 |
| GLDCP1 | 514.8996996 | -0.519222939 | 0.119759855 | 3.73E-06 | 4.13E-05 |
| ZNF253 | 269.6685988 | 0.519126805 | 0.143795367 | 7.50E-05 | 0.000617231 |
| SPC24 | 191.8404541 | 0.518893427 | 0.167103074 | 0.000457806 | 0.003027043 |
| SLC29A3 | 245.0191551 | 0.518613891 | 0.142339388 | 6.69E-05 | 0.00055893 |
| KDM5B | 1455.695113 | -0.518566889 | 0.079284399 | 1.68E-11 | 4.55E-10 |
| EEF1A1P24 | 120.6631268 | -0.518137535 | 0.218873826 | 0.003947502 | 0.018888888 |
| UTS2R | 152.5224437 | 0.517685855 | 0.192876911 | 0.001643832 | 0.009103373 |
| RPSAP15 | 5050.972217 | -0.51762677 | 0.077444746 | 6.45E-12 | 1.85E-10 |
| MAPKAPK3 | 1599.376237 | -0.517255809 | 0.089908211 | 2.41E-09 | 4.80E-08 |
| FBXO48 | 135.7731431 | 0.516681812 | 0.192874614 | 0.001671417 | 0.009223689 |
| NELFB | 1233.070081 | 0.516614032 | 0.110647055 | 7.98E-07 | 1.03E-05 |
| HNRNPDL | 6519.833783 | -0.516419106 | 0.081763461 | 7.35E-11 | 1.79E-09 |
| SLC5A3 | 2752.476005 | -0.516338583 | 0.10673237 | 3.49E-07 | 4.82E-06 |
| CCDC50 | 2100.527816 | 0.516190168 | 0.102828557 | 1.37E-07 | 2.04E-06 |
| FECH | 1304.258523 | -0.516114329 | 0.082492121 | 1.06E-10 | 2.52E-09 |
| MEX3A | 96.56957458 | 0.5160207 | 0.247863429 | 0.007499992 | 0.032203658 |
| SPTBN1 | 984.625646 | 0.515763482 | 0.092509987 | 6.60E-09 | 1.21E-07 |
| IGF2R | 5506.242351 | -0.515707751 | 0.07210166 | 2.37E-13 | 8.14E-12 |
| NAB2 | 650.5707069 | -0.515623059 | 0.116159434 | 2.43E-06 | 2.81E-05 |
| SLC10A7 | 393.979907 | -0.515561547 | 0.158024133 | 0.000269426 | 0.001896603 |
| POF1B | 91.10378509 | 0.515447946 | 0.230587859 | 0.005305456 | 0.024215755 |
| TBC1D32 | 50.03247233 | -0.515205856 | 0.297055013 | 0.0150284 | 0.056379289 |
| REST | 1334.713841 | 0.515178889 | 0.120957406 | 5.27E-06 | 5.64E-05 |
| PHB2 | 11013.08429 | -0.514866499 | 0.090449904 | 3.43E-09 | 6.64E-08 |
| MIIP | 165.8924199 | 0.514680417 | 0.18468517 | 0.001242426 | 0.007124201 |
| SF3B2 | 7418.509001 | -0.514658438 | 0.075195494 | 2.14E-12 | 6.53E-11 |
| TAP2 | 1995.542624 | 0.514610993 | 0.083480938 | 1.93E-10 | 4.42E-09 |
| FCSK | 209.6381685 | -0.514386977 | 0.16470484 | 0.000443008 | 0.00293803 |
| SMG6 | 620.618219 | 0.514383809 | 0.106860309 | 3.91E-07 | 5.33E-06 |
| NEU1 | 3013.211036 | -0.514198395 | 0.099033782 | 5.65E-08 | 8.93E-07 |

| | | | | | |
|---|---|---|---|---|---|
| CCNB2 | 1917.095578 | 0.514177353 | 0.07826556 | 1.48E-11 | 4.06E-10 |
| NPAS1 | 32.38119742 | -0.513821583 | 0.437919225 | 0.030172115 | 0.098101861 |
| PIM3 | 3435.364498 | -0.513809107 | 0.115425511 | 2.35E-06 | 2.73E-05 |
| CENPF | 374.7446848 | 0.513370409 | 0.182607557 | 0.001138289 | 0.006645904 |
| ITPR2 | 944.910194 | 0.513365146 | 0.107376094 | 4.59E-07 | 6.17E-06 |
| DCLRE1C | 407.8360372 | 0.513280333 | 0.122596841 | 7.31E-06 | 7.58E-05 |
| CRTAP | 3348.212911 | -0.51317987 | 0.080119618 | 4.21E-11 | 1.07E-09 |
| RPL3P2 | 1982.377018 | -0.513141607 | 0.11711973 | 3.16E-06 | 3.56E-05 |
| PCM1 | 1976.707895 | -0.513135676 | 0.083505608 | 2.18E-10 | 4.93E-09 |
| TMEM44 | 93.48991016 | 0.512820328 | 0.234838807 | 0.006145297 | 0.027363876 |
| RPL30 | 28290.10188 | -0.512598733 | 0.067583863 | 1.29E-14 | 5.05E-13 |
| USP33 | 736.9947808 | 0.512433378 | 0.100006763 | 7.99E-08 | 1.23E-06 |
| ZXDB | 140.3161653 | -0.512428679 | 0.205540136 | 0.002856803 | 0.014503447 |
| JUND | 7121.981625 | -0.512268692 | 0.237907422 | 0.006708739 | 0.029396565 |
| LOC124901066 | 9.782570597 | -0.511785596 | 0.81526216 | 0.023272878 | 0.080106624 |
| RHOF | 435.9533062 | 0.511710188 | 0.122654629 | 7.92E-06 | 8.16E-05 |
| BTBD6 | 433.4411327 | 0.511426721 | 0.127258915 | 1.52E-05 | 0.000148346 |
| TMEM147-AS1 | 85.81316667 | -0.511330841 | 0.242319827 | 0.007407825 | 0.031910775 |
| PAQR3 | 671.599016 | -0.511148684 | 0.16445935 | 0.000459635 | 0.003034503 |
| C4orf3 | 1463.765365 | -0.510941122 | 0.094428141 | 1.73E-08 | 2.97E-07 |
| ICAM4 | 338.9586415 | -0.510888593 | 0.133856785 | 3.58E-05 | 0.000319771 |
| RAB23 | 151.3962551 | 0.510855459 | 0.207846245 | 0.003081733 | 0.015428263 |
| LILRB4 | 4375.70188 | 0.510741174 | 0.073899403 | 1.33E-12 | 4.18E-11 |
| ZNF101 | 263.2532885 | 0.510167714 | 0.140596215 | 7.29E-05 | 0.000602546 |
| VASH2 | 50.86590213 | 0.5100522 | 0.334420618 | 0.020944239 | 0.073657877 |
| GDAP1 | 259.0456959 | -0.510042204 | 0.146473501 | 0.000123042 | 0.000952912 |
| SFMBT2 | 1659.848427 | 0.509583613 | 0.082179948 | 1.56E-10 | 3.61E-09 |
| NUP93 | 1751.259737 | -0.509084357 | 0.089269206 | 3.34E-09 | 6.47E-08 |
| RALGPS1 | 105.7245782 | 0.509072299 | 0.206744962 | 0.003115665 | 0.015580126 |
| DCAF8 | 2751.745521 | -0.509070529 | 0.072120331 | 4.71E-13 | 1.57E-11 |
| ZNF461 | 47.17483621 | 0.508729624 | 0.313050282 | 0.018273865 | 0.065954257 |
| STX3 | 438.3051416 | -0.508720472 | 0.125454268 | 1.32E-05 | 0.000130466 |
| USP7 | 4830.004388 | -0.50837249 | 0.082201585 | 1.74E-10 | 4.02E-09 |
| RAC2 | 4174.968786 | 0.507954509 | 0.082546047 | 2.07E-10 | 4.70E-09 |
| UQCRB | 7700.555549 | -0.50782195 | 0.077517433 | 1.56E-11 | 4.28E-10 |
| LOC105378512 | 49.58485831 | -0.507741822 | 0.334165505 | 0.021331423 | 0.074800492 |
| SERINC2 | 41.59062855 | 0.507445501 | 0.380399645 | 0.026959117 | 0.089986704 |
| ATP1B1 | 80.8990027 | 0.5073183 | 0.239046022 | 0.007130604 | 0.030955294 |
| MYH11 | 11.78368887 | 0.506915671 | 0.72146528 | 0.027197879 | 0.090569059 |
| WDR5 | 2042.322027 | 0.506640056 | 0.077568199 | 1.85E-11 | 4.99E-10 |
| ECT2 | 1256.306489 | 0.50661607 | 0.119285935 | 5.73E-06 | 6.10E-05 |
| DNMT3A | 111.6731616 | 0.506542357 | 0.197740895 | 0.002417591 | 0.012613867 |
| KLHL21 | 1787.06493 | -0.506267919 | 0.11322317 | 2.13E-06 | 2.50E-05 |
| TIRAP | 104.2381564 | 0.506125059 | 0.213634339 | 0.004034556 | 0.019231052 |
| PIR-FIGF | 114.0142174 | 0.506009892 | 0.202503433 | 0.002892381 | 0.014641144 |
| MYH9 | 3646.667655 | 0.505974953 | 0.09974018 | 1.07E-07 | 1.61E-06 |
| IL17D | 60.6461664 | -0.5058978 | 0.284467905 | 0.014445478 | 0.054725955 |
| PDK3 | 881.8795724 | 0.50551655 | 0.118141651 | 5.04E-06 | 5.42E-05 |
| CLEC1A | 8.498841094 | 0.505323523 | 0.928303914 | 0.018708207 | 0.067171888 |
| ZNF197 | 117.0615253 | -0.505316566 | 0.211249947 | 0.003774637 | 0.018177249 |

| | | | | | |
|---|---|---|---|---|---|
| P2RY8 | 2279.273354 | 0.505171426 | 0.083767743 | 4.59E-10 | 1.02E-08 |
| HLA-E | 9786.13584 | 0.505146729 | 0.076486743 | 6.46E-12 | 1.85E-10 |
| ZNF696 | 112.9535964 | 0.504880366 | 0.211120768 | 0.00380421 | 0.018302162 |
| ARRDC1-AS1 | 74.38791046 | 0.504493265 | 0.261314561 | 0.010874713 | 0.043591994 |
| IGKV3D-20 | 89740.54299 | -0.504150804 | 0.082490097 | 2.76E-10 | 6.19E-09 |
| EEF1A1P6 | 76800.0169 | -0.503941163 | 0.065094404 | 3.53E-15 | 1.46E-13 |
| CLSPN | 556.5895199 | 0.503741138 | 0.108413108 | 9.23E-07 | 1.17E-05 |
| GAS6 | 762.6551032 | -0.503608051 | 0.113718411 | 2.64E-06 | 3.02E-05 |
| MRPS6 | 2053.681489 | -0.503513442 | 0.080042806 | 9.07E-11 | 2.19E-09 |
| IGKV3D-11 | 4431.485928 | -0.503023134 | 0.095011296 | 3.39E-08 | 5.56E-07 |
| ARNTL2 | 533.7193495 | 0.502610453 | 0.115417893 | 3.60E-06 | 4.00E-05 |
| CCNL1 | 1066.062457 | -0.502444797 | 0.112450708 | 2.14E-06 | 2.51E-05 |
| LINC01518 | 68.36159612 | 0.502404291 | 0.266005411 | 0.011906053 | 0.046762786 |
| PIP4K2A | 1461.0414 | 0.50233515 | 0.091214731 | 1.02E-08 | 1.83E-07 |
| RPS4XP17 | 37.47606325 | -0.502212962 | 0.349926419 | 0.025002598 | 0.084829543 |
| GCSHP5 | 925.6329971 | -0.502138356 | 0.104109321 | 3.91E-07 | 5.32E-06 |
| PTDSS1 | 5606.7099 | -0.502011996 | 0.07134144 | 5.78E-13 | 1.90E-11 |
| IP6K2 | 1172.525797 | -0.501945993 | 0.086853547 | 2.16E-09 | 4.33E-08 |
| PIR | 201.0829503 | 0.501723375 | 0.163993492 | 0.000561692 | 0.003611269 |
| SLC48A1 | 280.520833 | 0.50149221 | 0.158297868 | 0.000390665 | 0.002629284 |
| ZNF681 | 138.1780452 | -0.501402808 | 0.209978581 | 0.003898099 | 0.018709364 |
| ESPL1 | 653.2295044 | 0.500850786 | 0.111624639 | 2.01E-06 | 2.37E-05 |
| ENPP1 | 142.8149307 | 0.500004794 | 0.206996072 | 0.003663892 | 0.017767466 |
| FOXP4 | 376.8985249 | 0.500003562 | 0.14984801 | 0.000219014 | 0.001580276 |
| RFESD | 71.4946931 | -0.499954446 | 0.266932529 | 0.012354796 | 0.048235176 |
| INPP5F | 1022.866844 | 0.499747085 | 0.097209129 | 7.72E-08 | 1.19E-06 |
| MRPS28 | 615.9379422 | -0.499719143 | 0.10072469 | 1.98E-07 | 2.85E-06 |
| IGHV3-23 | 51958.95357 | -0.499669821 | 0.104093133 | 6.37E-07 | 8.37E-06 |
| MTLN | 191.9684678 | 0.499301768 | 0.199814639 | 0.002958451 | 0.014892877 |
| MTM1 | 463.4079071 | 0.499134677 | 0.11757686 | 5.92E-06 | 6.27E-05 |
| CDK17 | 1560.569035 | 0.499094547 | 0.114984096 | 3.86E-06 | 4.25E-05 |
| HEATR5B | 886.8638224 | 0.498981156 | 0.091882392 | 1.58E-08 | 2.72E-07 |
| ZNF660-ZNF197 | 116.7549208 | -0.498502076 | 0.211298719 | 0.004210147 | 0.019952688 |
| C16orf74 | 257.8872009 | -0.498372024 | 0.180467122 | 0.001446741 | 0.008144736 |
| SARAF | 7239.38295 | -0.498352591 | 0.089263435 | 6.83E-09 | 1.25E-07 |
| ATP6V0E2 | 577.8257756 | -0.498314896 | 0.128569472 | 2.94E-05 | 0.000267745 |
| RAPGEF1 | 1640.771897 | 0.49830167 | 0.088322735 | 4.78E-09 | 9.00E-08 |
| RPL19P4 | 58.45576086 | 0.498252131 | 0.352398243 | 0.025390911 | 0.085911374 |
| RPL3 | 98139.73188 | -0.498124838 | 0.071964123 | 1.47E-12 | 4.61E-11 |
| OSGEPL1 | 199.0462264 | 0.497625445 | 0.158462047 | 0.000438295 | 0.002911377 |
| IGHJ6 | 667.0959219 | -0.497566042 | 0.111775469 | 2.41E-06 | 2.79E-05 |
| DPY19L1P2 | 45.84398283 | -0.497503895 | 0.311503361 | 0.020354837 | 0.071879753 |
| ZYX | 492.5083046 | 0.497224452 | 0.151973781 | 0.000277558 | 0.001945943 |
| CASP10 | 2036.104477 | 0.496949304 | 0.077873042 | 5.07E-11 | 1.26E-09 |
| EEF1A1P14 | 2150.044178 | -0.496761731 | 0.099712585 | 1.80E-07 | 2.62E-06 |
| LOC124900275 | 265.6147245 | 0.496493821 | 0.136507581 | 7.41E-05 | 0.000610734 |
| RNLS | 178.3069366 | -0.496443424 | 0.166551683 | 0.000734583 | 0.004570885 |
| LINC02820 | 119.0271626 | 0.496250435 | 0.210457447 | 0.004254337 | 0.020133008 |
| INCENP | 1227.424968 | 0.496087542 | 0.098703821 | 1.42E-07 | 2.10E-06 |
| ATP8B1-AS1 | 74.7453791 | 0.496067179 | 0.248614299 | 0.009843366 | 0.040287773 |

| | | | | | |
|---|---|---|---|---|---|
| POFUT2 | 644.7848431 | -0.495811008 | 0.108642395 | 1.43E-06 | 1.74E-05 |
| MARK2 | 829.9103179 | 0.495598732 | 0.111370709 | 2.39E-06 | 2.76E-05 |
| HHLA3 | 41.49732892 | -0.495576744 | 0.328633741 | 0.02314616 | 0.079765582 |
| URM1 | 1773.072706 | 0.495076629 | 0.10922529 | 1.63E-06 | 1.96E-05 |
| CPNE3 | 2094.880397 | -0.494602671 | 0.111965089 | 2.81E-06 | 3.19E-05 |
| CCDC25 | 1572.939061 | -0.494553829 | 0.084635928 | 1.46E-09 | 3.00E-08 |
| TMEM219 | 708.1243828 | -0.49447037 | 0.105634668 | 8.25E-07 | 1.06E-05 |
| LOC105378482 | 116.0083654 | -0.494469611 | 0.20904579 | 0.004314999 | 0.020343359 |
| FCER2 | 110.5882805 | 0.493869552 | 0.214360235 | 0.00493905 | 0.022850531 |
| BARD1 | 868.5322518 | 0.493861452 | 0.135025762 | 6.84E-05 | 0.000569228 |
| RPSAP54 | 2257.902248 | -0.493821043 | 0.089998036 | 1.21E-08 | 2.12E-07 |
| RPS14P3 | 100.7083393 | 0.493610078 | 0.300917362 | 0.018830072 | 0.067539428 |
| RIOK2 | 1491.624135 | -0.493521805 | 0.093185645 | 3.46E-08 | 5.66E-07 |
| PRDX6 | 7251.359985 | -0.493398966 | 0.069126554 | 2.86E-13 | 9.67E-12 |
| CHPF2 | 1286.535543 | -0.493314392 | 0.103503011 | 5.45E-07 | 7.25E-06 |
| HSDL2 | 598.3240297 | -0.493244732 | 0.138527065 | 9.99E-05 | 0.000794199 |
| ELOC | 2305.244266 | -0.492849645 | 0.097551193 | 1.23E-07 | 1.85E-06 |
| JAK2 | 260.7876824 | 0.492781163 | 0.161332189 | 0.000578982 | 0.003710673 |
| GCSH | 1008.984112 | -0.49250719 | 0.093687845 | 4.24E-08 | 6.88E-07 |
| ZSWIM4 | 103.0249058 | 0.492383282 | 0.20526345 | 0.00392962 | 0.01883296 |
| GDPD5 | 112.2987522 | 0.492382636 | 0.224012854 | 0.006441139 | 0.028432629 |
| SNHG12 | 113.2707976 | -0.492049041 | 0.197792947 | 0.00315453 | 0.015756272 |
| USP46 | 496.589847 | 0.491932286 | 0.137383527 | 9.21E-05 | 0.000739357 |
| RPSAP53 | 589.5274176 | -0.491605475 | 0.117649719 | 8.20E-06 | 8.43E-05 |
| ZNF32 | 153.6900465 | 0.491596901 | 0.193472209 | 0.002705847 | 0.013853796 |
| COX6C | 7815.732175 | -0.491578741 | 0.072456876 | 3.50E-12 | 1.04E-10 |
| FBXO22 | 777.0450689 | -0.491514217 | 0.109100317 | 1.89E-06 | 2.24E-05 |
| MSC-AS1 | 143.7042186 | 0.491445034 | 0.186312328 | 0.00206726 | 0.011031972 |
| PPOX | 508.4774291 | -0.491286533 | 0.131877034 | 5.50E-05 | 0.000468605 |
| SRGN | 11092.01934 | -0.491148758 | 0.094547682 | 6.00E-08 | 9.42E-07 |
| OIP5-AS1 | 2725.590807 | 0.491078097 | 0.094999635 | 6.77E-08 | 1.06E-06 |
| C21orf91 | 2185.196732 | 0.490926436 | 0.281760429 | 0.016066873 | 0.059450865 |
| EEF1A1P5 | 95964.76567 | -0.49015188 | 0.06658632 | 5.48E-14 | 2.02E-12 |
| HMGCR | 8311.02162 | -0.490066073 | 0.072099724 | 3.22E-12 | 9.63E-11 |
| LOC107984950 | 69.2544884 | 0.489885325 | 0.25472453 | 0.011668931 | 0.046029813 |
| IGKV3D-15 | 1398.397438 | -0.489571824 | 0.115784183 | 6.82E-06 | 7.13E-05 |
| IGKV3D-7 | 3186.955388 | -0.489319638 | 0.073725026 | 9.88E-12 | 2.77E-10 |
| DGKA | 141.6892904 | 0.489214561 | 0.190270452 | 0.002531455 | 0.01309099 |
| KCNQ1OT1 | 39.78766292 | -0.489200786 | 0.376199483 | 0.030047404 | 0.097861744 |
| SLC27A4 | 655.6669855 | 0.489199587 | 0.110740521 | 2.86E-06 | 3.25E-05 |
| WSB1 | 1828.894848 | -0.489089116 | 0.079015743 | 1.80E-10 | 4.13E-09 |
| CTSS | 7670.779436 | 0.488977937 | 0.072799429 | 5.54E-12 | 1.61E-10 |
| SNX33 | 91.77367496 | 0.488876334 | 0.233065781 | 0.008179813 | 0.034515063 |
| SHISAL2A | 200.133227 | 0.488676879 | 0.222453489 | 0.006500517 | 0.028636341 |
| RPSA | 43821.68586 | -0.488574803 | 0.08908609 | 1.23E-08 | 2.15E-07 |
| ZFAND2A | 556.4878483 | -0.48840012 | 0.14683744 | 0.000241311 | 0.001726772 |
| LCMT1 | 771.8492325 | -0.48800907 | 0.097094238 | 1.48E-07 | 2.19E-06 |
| GSTA4 | 47.93060546 | 0.487960452 | 0.319420646 | 0.022943512 | 0.079288124 |
| AP1G1 | 4890.791299 | -0.487883883 | 0.068156835 | 2.58E-13 | 8.82E-12 |
| LOC107984561 | 11.54550847 | -0.487679263 | 0.683973877 | 0.029764621 | 0.097178347 |

| | | | | | |
|---|---|---|---|---|---|
| LOC105369957 | 12.40213898 | 0.487598521 | 0.652401982 | 0.030848229 | 0.09977564 |
| ZNF776 | 437.6408197 | 0.487536818 | 0.117364631 | 9.27E-06 | 9.39E-05 |
| MACF1 | 767.7867052 | 0.487239357 | 0.149644214 | 0.000300419 | 0.002087625 |
| SLC31A1P1 | 32.51725488 | -0.487070611 | 0.369994979 | 0.03025985 | 0.098276411 |
| ALDH1L2 | 2894.940067 | -0.486944229 | 0.079028487 | 2.16E-10 | 4.90E-09 |
| EMC9 | 301.0651465 | 0.486831179 | 0.155478126 | 0.000469524 | 0.003089192 |
| CASK | 480.9733678 | 0.486617361 | 0.111124507 | 3.40E-06 | 3.79E-05 |
| RSPRY1 | 774.1330413 | -0.486530301 | 0.110471915 | 3.06E-06 | 3.45E-05 |
| NQO1 | 3837.299544 | -0.486296264 | 0.076041198 | 4.91E-11 | 1.22E-09 |
| ACSF3 | 1037.998409 | -0.486141789 | 0.103787731 | 8.39E-07 | 1.08E-05 |
| MED22 | 581.0414364 | 0.486024648 | 0.115282433 | 7.23E-06 | 7.51E-05 |
| TRBC2 | 1667.349371 | 0.485940341 | 0.095374524 | 1.03E-07 | 1.56E-06 |
| USP20 | 379.7844269 | 0.48575781 | 0.123362498 | 2.33E-05 | 0.000217938 |
| USP12 | 1244.629981 | -0.485749489 | 0.098914658 | 2.65E-07 | 3.75E-06 |
| STAG3L4 | 252.812229 | 0.485667989 | 0.142837654 | 0.000185887 | 0.00136979 |
| SGPL1 | 653.7203842 | 0.485609238 | 0.102658065 | 6.61E-07 | 8.66E-06 |
| EIF4E3 | 550.3624569 | 0.485175592 | 0.131900981 | 6.50E-05 | 0.000543791 |
| SNORD141A | 1149.879122 | -0.485097994 | 0.097136818 | 1.79E-07 | 2.60E-06 |
| PAXX | 506.4979006 | 0.48504401 | 0.140583946 | 0.000156025 | 0.0011732 |
| ZNF416 | 149.044969 | 0.484998247 | 0.180348292 | 0.001802332 | 0.009833328 |
| DYNC2I2 | 966.0504433 | 0.484786843 | 0.100487227 | 4.14E-07 | 5.62E-06 |
| RGS1 | 6201.915724 | -0.484451017 | 0.09690587 | 1.85E-07 | 2.68E-06 |
| ZNF490 | 261.0343197 | 0.484384246 | 0.148192238 | 0.000292602 | 0.002038208 |
| TRPV3 | 17.59568876 | -0.484375519 | 0.631132537 | 0.030804805 | 0.099709684 |
| MSMO1 | 3971.662429 | -0.48431618 | 0.095323834 | 1.12E-07 | 1.69E-06 |
| BTN2A2 | 1168.83122 | 0.484108819 | 0.084643816 | 3.19E-09 | 6.20E-08 |
| PIK3R5 | 12.03115034 | 0.484048079 | 0.732859113 | 0.02606395 | 0.087657684 |
| TGFBR3 | 64.30139134 | 0.483976798 | 0.283794954 | 0.017783927 | 0.064603371 |
| DPM2 | 742.9102902 | 0.483873821 | 0.126629155 | 3.80E-05 | 0.000336042 |
| MARCHF2 | 203.368915 | 0.483820904 | 0.181213985 | 0.001969195 | 0.010600107 |
| PRRC2B | 2657.820232 | 0.483779704 | 0.101639249 | 5.66E-07 | 7.49E-06 |
| ABCC4 | 1745.725234 | -0.483778467 | 0.081697377 | 9.71E-10 | 2.04E-08 |
| SHKBP1 | 2012.312807 | 0.483678947 | 0.114464526 | 6.93E-06 | 7.23E-05 |
| IGKV3-15 | 1452.642019 | -0.483627642 | 0.115526179 | 8.32E-06 | 8.53E-05 |
| TRIM36 | 145.3409897 | 0.48355702 | 0.195214524 | 0.003308095 | 0.016381552 |
| SPTAN1 | 3158.707996 | 0.483415447 | 0.08704188 | 8.26E-09 | 1.49E-07 |
| CDC42EP3 | 534.6937866 | 0.483400412 | 0.114378219 | 6.85E-06 | 7.15E-05 |
| DMKN | 82.08092725 | -0.483088957 | 0.245288167 | 0.011203251 | 0.044671127 |
| LPGAT1 | 1269.767772 | 0.4829911 | 0.106458927 | 1.65E-06 | 1.99E-05 |
| SLC30A1 | 3038.802326 | -0.482975236 | 0.149810434 | 0.000349472 | 0.002379788 |
| FMR1 | 614.7995928 | 0.482793548 | 0.179355361 | 0.001927032 | 0.010402249 |
| MAGEA10 | 1797.490151 | -0.482491151 | 0.084790616 | 3.84E-09 | 7.36E-08 |
| BRD7P2 | 139.2369521 | -0.482070681 | 0.183594479 | 0.002229502 | 0.011774409 |
| CFLAR | 1357.894512 | 0.481990292 | 0.083983954 | 2.86E-09 | 5.60E-08 |
| ZNF263 | 1089.406775 | -0.481969874 | 0.097664206 | 2.41E-07 | 3.43E-06 |
| ABHD18 | 300.9323774 | -0.481820189 | 0.132036134 | 7.59E-05 | 0.000623823 |
| PMM2 | 1836.22363 | -0.481574689 | 0.081570966 | 1.10E-09 | 2.29E-08 |
| LILRA2 | 982.1661721 | 0.481559585 | 0.100948328 | 5.49E-07 | 7.30E-06 |
| WASL-DT | 41.93866732 | -0.4813742 | 0.326447539 | 0.026043606 | 0.087608387 |
| ATP1A3 | 2664.550845 | -0.481301568 | 0.081370018 | 1.03E-09 | 2.15E-08 |

| | | | | | |
|---|---|---|---|---|---|
| APOBEC3A | 266.1945396 | 0.480867919 | 0.139798957 | 0.000163311 | 0.001221626 |
| GTF3C5 | 1930.838582 | 0.480819976 | 0.093282285 | 7.65E-08 | 1.18E-06 |
| RNF139 | 1837.036979 | -0.480768795 | 0.092112532 | 5.42E-08 | 8.60E-07 |
| DIPK1A | 1245.266799 | 0.480763828 | 0.108304916 | 2.65E-06 | 3.03E-05 |
| LSR | 2367.849932 | 0.480713332 | 0.103675183 | 1.05E-06 | 1.31E-05 |
| LINC01234 | 112.0227298 | 0.480663717 | 0.212429537 | 0.005778094 | 0.025962441 |
| TANGO2 | 581.626052 | 0.480615594 | 0.116614476 | 1.10E-05 | 0.000110685 |
| ZMYND8 | 480.0958542 | 0.480430221 | 0.117860138 | 1.31E-05 | 0.000129485 |
| AHNAK | 8447.41327 | 0.480359871 | 0.105077496 | 1.40E-06 | 1.70E-05 |
| LAMTOR3 | 1398.256949 | -0.480355397 | 0.109467245 | 3.34E-06 | 3.73E-05 |
| GNPTAB | 1052.453084 | -0.480107016 | 0.091845887 | 5.18E-08 | 8.27E-07 |
| ZBED1 | 723.874828 | 0.480064656 | 0.108769764 | 3.06E-06 | 3.45E-05 |
| TENT4B | 603.0740348 | -0.47982699 | 0.162035051 | 0.000829437 | 0.005064875 |
| GATB | 691.0564171 | -0.479658242 | 0.103827505 | 1.16E-06 | 1.44E-05 |
| GDF11 | 855.637106 | 0.479632363 | 0.090921534 | 3.98E-08 | 6.50E-07 |
| RPSAP55 | 126.4270152 | -0.479503969 | 0.22051454 | 0.007250117 | 0.031375135 |
| POLR3D | 1019.494901 | -0.479391317 | 0.10581367 | 1.77E-06 | 2.12E-05 |
| DNAJC14 | 1416.539692 | 0.479295 | 0.079525503 | 5.10E-10 | 1.12E-08 |
| CLYBL | 415.3007913 | -0.479213505 | 0.12116875 | 2.23E-05 | 0.000209173 |
| SNORD141B | 1153.381783 | -0.479016966 | 0.097659686 | 2.87E-07 | 4.04E-06 |
| SP100 | 1055.475699 | 0.478761663 | 0.089691203 | 2.84E-08 | 4.71E-07 |
| SMURF2P1-LRRC37BP1 | 201.0794266 | 0.47867695 | 0.152905489 | 0.000483442 | 0.003165126 |
| KDELR1 | 6149.341142 | -0.478367759 | 0.078348946 | 3.25E-10 | 7.26E-09 |
| SS18L2 | 783.0657582 | -0.478367464 | 0.102207397 | 8.77E-07 | 1.12E-05 |
| ADAM15 | 2392.432411 | -0.478197884 | 0.103649479 | 1.20E-06 | 1.48E-05 |
| ZFP69 | 144.3389642 | 0.477903563 | 0.178761206 | 0.001984333 | 0.01066502 |
| XPO6 | 1959.896431 | -0.477621067 | 0.106105023 | 2.04E-06 | 2.41E-05 |
| HSPA8P1 | 184.9494264 | -0.477529738 | 0.203609688 | 0.004820936 | 0.022405838 |
| IGHA2 | 24749.5514 | -0.477509167 | 0.0855875 | 7.55E-09 | 1.37E-07 |
| HEXIM1 | 1832.727642 | -0.477313461 | 0.086819203 | 1.20E-08 | 2.11E-07 |
| SQOR | 585.4277444 | 0.477265285 | 0.10941948 | 3.87E-06 | 4.26E-05 |
| STXBP6 | 238.8701656 | -0.477128738 | 0.172112324 | 0.001479633 | 0.008303083 |
| MFSD8 | 388.9064488 | -0.476979064 | 0.132755183 | 9.37E-05 | 0.000751393 |
| GPATCH11 | 1199.250717 | 0.476895573 | 0.091333661 | 5.36E-08 | 8.51E-07 |
| WRN | 987.1394921 | -0.476868852 | 0.095607614 | 1.85E-07 | 2.69E-06 |
| KCNA2 | 160.9684387 | -0.476694107 | 0.169618222 | 0.001343127 | 0.007622012 |
| MIR6797 | 234.6160808 | -0.476688577 | 0.149229646 | 0.000398995 | 0.002677024 |
| RGPD2 | 313.9233281 | 0.476640542 | 0.128339135 | 5.92E-05 | 0.000500016 |
| RPS2P46 | 13166.37468 | -0.476584674 | 0.072021954 | 1.15E-11 | 3.20E-10 |
| TRIM21 | 347.7401876 | 0.47657056 | 0.123881805 | 3.47E-05 | 0.000310574 |
| POLH | 227.3023303 | -0.476434292 | 0.170787791 | 0.001425502 | 0.008046097 |
| EEF2K | 539.205437 | -0.476411486 | 0.107716326 | 2.94E-06 | 3.32E-05 |
| RPS15AP12 | 776.6304052 | -0.476368958 | 0.108526454 | 3.37E-06 | 3.77E-05 |
| ILDR2 | 87.7960504 | -0.47623891 | 0.225997203 | 0.008531303 | 0.035702411 |
| SAPCD2 | 816.9637493 | 0.476193585 | 0.134213693 | 0.000111331 | 0.000873155 |
| TRAFD1 | 788.5715838 | 0.476083584 | 0.10530234 | 1.84E-06 | 2.20E-05 |
| ALDOC | 7455.789767 | 0.476040237 | 0.073047981 | 2.24E-11 | 5.91E-10 |
| LINC01963 | 267.6255925 | -0.476010015 | 0.147190676 | 0.000343909 | 0.002347446 |
| HSD17B7 | 1319.395695 | -0.476001966 | 0.093002008 | 9.60E-08 | 1.46E-06 |
| METTL8 | 576.4352679 | -0.475869658 | 0.114014159 | 8.95E-06 | 9.09E-05 |

| | | | | | |
|---|---|---|---|---|---|
| MEF2C-AS1 | 64.90749162 | -0.47586096 | 0.264486062 | 0.015943888 | 0.059122122 |
| SINHCAF | 1865.381766 | 0.475823187 | 0.091398275 | 5.85E-08 | 9.19E-07 |
| LTA4H | 3011.910678 | -0.475733047 | 0.076485435 | 1.57E-10 | 3.62E-09 |
| RPL10AP2 | 193.702969 | -0.475671837 | 0.253281762 | 0.013736469 | 0.05269704 |
| TMX4 | 3557.71642 | 0.475550612 | 0.084198942 | 5.00E-09 | 9.34E-08 |
| TNFRSF13B | 204.6806278 | -0.47528539 | 0.153809827 | 0.000564203 | 0.003625342 |
| PDPR2P | 107.585416 | -0.47525554 | 0.2115939 | 0.006207258 | 0.027575957 |
| SRGAP2C | 134.775311 | 0.47520386 | 0.197729547 | 0.004135372 | 0.019652094 |
| RASSF1 | 309.8670811 | -0.475159334 | 0.13417249 | 0.000116739 | 0.000910208 |
| GPC6 | 1037.105419 | -0.475095712 | 0.102415661 | 1.07E-06 | 1.33E-05 |
| AK6 | 1286.880802 | -0.474883703 | 0.097279264 | 3.22E-07 | 4.48E-06 |
| EEF1A1P19 | 13193.56152 | -0.474653804 | 0.088884059 | 2.86E-08 | 4.75E-07 |
| COX1 | 154338.5723 | -0.474528143 | 0.078635186 | 5.05E-10 | 1.11E-08 |
| ANKRD17 | 1664.3001 | -0.474396551 | 0.094346229 | 1.46E-07 | 2.16E-06 |
| SNTA1 | 417.2793688 | -0.474381597 | 0.132982847 | 0.000106586 | 0.000842043 |
| TSTD1 | 1206.901481 | -0.47385841 | 0.117547274 | 1.66E-05 | 0.000161008 |
| RPL29 | 26185.84166 | -0.473700167 | 0.082469318 | 2.51E-09 | 4.97E-08 |
| DIS3 | 2922.431867 | -0.473036585 | 0.084545207 | 6.88E-09 | 1.26E-07 |
| RPS7 | 100731.9685 | -0.47280872 | 0.071366327 | 2.09E-11 | 5.55E-10 |
| RRN3 | 1903.530555 | -0.472698375 | 0.100600934 | 8.01E-07 | 1.03E-05 |
| C2orf68 | 1163.403278 | 0.472130855 | 0.085870661 | 1.18E-08 | 2.08E-07 |
| USP38 | 2287.110519 | -0.471955754 | 0.085251306 | 9.68E-09 | 1.74E-07 |
| KIFAP3 | 1130.671653 | -0.471949755 | 0.088117696 | 2.69E-08 | 4.48E-07 |
| TTC30B | 129.6238183 | 0.47187354 | 0.186788653 | 0.003057627 | 0.015329743 |
| CHEK2 | 123.9424931 | 0.471742514 | 0.19868966 | 0.004531564 | 0.021220414 |
| C16orf91 | 296.7854635 | -0.471595891 | 0.154300265 | 0.000643992 | 0.004070075 |
| IGHJ5 | 101.8589471 | -0.471027687 | 0.212205099 | 0.006688231 | 0.029336396 |
| WDR81 | 753.1409059 | 0.471012315 | 0.12614855 | 5.59E-05 | 0.000476322 |
| GOLGA8M | 43.08600674 | 0.470638542 | 0.321418779 | 0.026915491 | 0.08991041 |
| RPS7P11 | 10944.13477 | -0.470598311 | 0.076015776 | 1.91E-10 | 4.38E-09 |
| ND2 | 43309.73066 | -0.470586129 | 0.073573121 | 5.06E-11 | 1.26E-09 |
| ABCA3 | 196.1402906 | -0.470130353 | 0.17193416 | 0.001739271 | 0.009552474 |
| RACGAP1 | 3260.495299 | 0.470076335 | 0.070746254 | 9.65E-12 | 2.71E-10 |
| GALE | 310.4313368 | -0.469975952 | 0.138980649 | 0.00021353 | 0.001545861 |
| METTL9 | 2318.932705 | -0.469970877 | 0.080280169 | 1.53E-09 | 3.12E-08 |
| PDXP | 1334.614631 | 0.469707858 | 0.129281427 | 8.27E-05 | 0.000672166 |
| MSR1 | 796.0379718 | -0.469475276 | 0.130808512 | 9.82E-05 | 0.000783675 |
| EEF1A1P13 | 24209.3897 | -0.469474618 | 0.085622751 | 1.34E-08 | 2.32E-07 |
| BACH1 | 2108.672738 | -0.469331608 | 0.122094587 | 3.65E-05 | 0.000324906 |
| ATP6V1C1 | 2133.101077 | -0.469218822 | 0.092596131 | 1.26E-07 | 1.90E-06 |
| LOC124900868 | 362.8538185 | -0.469183199 | 0.1226385 | 3.94E-05 | 0.00034724 |
| SLC9B2 | 1167.877976 | -0.469159939 | 0.088698678 | 3.87E-08 | 6.31E-07 |
| B3GNT2 | 957.8424836 | 0.468981216 | 0.137448214 | 0.000186903 | 0.00137669 |
| POGLUT2 | 462.8635293 | -0.468926223 | 0.109067222 | 5.31E-06 | 5.68E-05 |
| LGR4 | 355.2007344 | -0.468749419 | 0.152538412 | 0.000602593 | 0.0038435 |
| TRIM68 | 307.0835942 | 0.468738998 | 0.130920064 | 0.000100878 | 0.000801703 |
| MEF2A | 1438.327863 | 0.468407018 | 0.107794874 | 4.24E-06 | 4.64E-05 |
| H1-10 | 3417.465694 | -0.468116007 | 0.144292231 | 0.000349833 | 0.002381315 |
| WASHC5 | 1732.748306 | -0.468082819 | 0.080533877 | 1.97E-09 | 3.97E-08 |
| RPSAP58 | 9599.633556 | -0.468003243 | 0.078477096 | 7.98E-10 | 1.70E-08 |

| | | | | | |
|---|---|---|---|---|---|
| TPX2 | 3003.47375 | 0.467952944 | 0.07878972 | 9.02E-10 | 1.90E-08 |
| RGL1 | 46.99072063 | 0.467932384 | 0.317392498 | 0.027187487 | 0.090569059 |
| RPL29P4 | 3129.659855 | -0.467827126 | 0.073667052 | 6.98E-11 | 1.71E-09 |
| CKB | 803.9566637 | 0.4672718 | 0.133554483 | 0.000138604 | 0.001057822 |
| PDXDC1 | 2164.63542 | -0.4671933 | 0.072435979 | 3.65E-11 | 9.33E-10 |
| CENPI | 203.6830346 | 0.467002156 | 0.164383283 | 0.001257423 | 0.007198012 |
| BAIAP2-DT | 104.7995584 | 0.466684447 | 0.219596929 | 0.008401026 | 0.035281782 |
| SEMA3G | 148.4607003 | -0.466200508 | 0.176190092 | 0.002263502 | 0.011928503 |
| C2CD2 | 10.08084718 | 0.465935506 | 0.711475671 | 0.027648778 | 0.091793308 |
| CD99 | 451.6054996 | -0.465906707 | 0.123301998 | 4.85E-05 | 0.000417427 |
| VIM-AS1 | 92.74264665 | 0.465826048 | 0.23012812 | 0.010547843 | 0.042606171 |
| LOC124906110 | 101.1802278 | -0.465820586 | 0.220826441 | 0.00878795 | 0.036608355 |
| HSPA8 | 149889.135 | -0.465627111 | 0.067182151 | 1.88E-12 | 5.82E-11 |
| KBTBD7 | 628.1242601 | 0.465557492 | 0.123357837 | 4.83E-05 | 0.000416336 |
| MANEAL | 418.3966507 | 0.465394659 | 0.116834119 | 2.08E-05 | 0.000196039 |
| TMEM102 | 225.6863216 | 0.465111942 | 0.161909226 | 0.001161719 | 0.006756614 |
| AFTPH-DT | 175.6099068 | -0.464875463 | 0.161552329 | 0.0011525 | 0.006713034 |
| DDX59 | 331.5848379 | 0.464789383 | 0.12592635 | 6.71E-05 | 0.000560035 |
| BMI1 | 2713.672019 | 0.464789169 | 0.139060971 | 0.00027347 | 0.00192039 |
| ATP6V1B2 | 3072.603548 | -0.464631198 | 0.073379734 | 7.98E-11 | 1.93E-09 |
| CCDC15 | 55.75084994 | 0.464602618 | 0.279664247 | 0.020901338 | 0.073566764 |
| UPF3AP2 | 68.72881423 | -0.464339553 | 0.254964235 | 0.016078371 | 0.059480698 |
| RRN3P2 | 214.4612773 | -0.464135799 | 0.155627959 | 0.000838491 | 0.005107331 |
| RAB11FIP2 | 156.6902874 | 0.464063078 | 0.189288349 | 0.00379139 | 0.018252851 |
| NAIF1 | 514.8464912 | 0.463872754 | 0.107783715 | 5.25E-06 | 5.62E-05 |
| FRMD8 | 1286.631258 | -0.463813914 | 0.099101062 | 9.16E-07 | 1.17E-05 |
| SERTAD2 | 868.8048988 | 0.463784309 | 0.104359009 | 2.73E-06 | 3.11E-05 |
| RASAL3 | 916.03576 | 0.46373838 | 0.10717788 | 4.69E-06 | 5.09E-05 |
| GIT2 | 616.6192782 | 0.463346889 | 0.100447138 | 1.25E-06 | 1.54E-05 |
| AGFG2 | 195.2256255 | 0.463205232 | 0.167507139 | 0.001616473 | 0.008974821 |
| SNHG32 | 4439.727017 | 0.463103969 | 0.073082087 | 7.73E-11 | 1.88E-09 |
| BOLA1 | 75.89942094 | -0.462893596 | 0.270287015 | 0.019686813 | 0.069989024 |
| SETSIP | 67.82127472 | 0.462839227 | 0.256824235 | 0.01660844 | 0.060946885 |
| MKI67 | 525.3963156 | 0.462703474 | 0.1743576 | 0.002180736 | 0.011562709 |
| RNF168 | 517.7518938 | 0.462677084 | 0.128722731 | 9.72E-05 | 0.000775785 |
| ARHGAP11B | 172.0086662 | 0.462600634 | 0.169275503 | 0.001759988 | 0.009638847 |
| HGSNAT | 442.3364016 | -0.462548408 | 0.114748835 | 1.75E-05 | 0.00016821 |
| FAM53B | 300.4926418 | 0.462289448 | 0.148500944 | 0.000544825 | 0.003508642 |
| CALM3 | 6408.410253 | 0.462035245 | 0.067343232 | 2.29E-12 | 6.96E-11 |
| SEC14L1 | 3267.007492 | 0.461875662 | 0.073462527 | 1.05E-10 | 2.50E-09 |
| FASN | 6525.304382 | -0.461763115 | 0.094562021 | 3.36E-07 | 4.66E-06 |
| PPP2CB | 1210.944166 | -0.461619911 | 0.089903521 | 9.09E-08 | 1.39E-06 |
| IQCB1 | 369.8180539 | -0.461398133 | 0.129552937 | 0.000113242 | 0.00088613 |
| ZNF180 | 285.3430256 | 0.461260029 | 0.139649583 | 0.000283009 | 0.001977753 |
| TGS1 | 1135.457829 | -0.461083195 | 0.08953127 | 8.36E-08 | 1.28E-06 |
| TRBC1 | 186.3654331 | 0.460959588 | 0.162626867 | 0.001325613 | 0.007536322 |
| ADH5P4 | 85.50467024 | -0.460724255 | 0.233174555 | 0.012025321 | 0.047124344 |
| MIS18BP1 | 705.9191863 | 0.459820306 | 0.108024832 | 6.47E-06 | 6.77E-05 |
| MRM1 | 204.0243776 | 0.459722844 | 0.153376439 | 0.000800165 | 0.004913635 |
| MTHFR | 389.1773314 | -0.459570652 | 0.134847644 | 0.000201124 | 0.001468944 |

| | | | | | |
|---|---|---|---|---|---|
| ARGLU1 | 1457.723037 | -0.459545299 | 0.087634808 | 5.04E-08 | 8.06E-07 |
| ATP23 | 363.9944289 | 0.459510425 | 0.130306535 | 0.00012757 | 0.000984025 |
| PIGS | 2504.29088 | 0.459256205 | 0.081124093 | 4.93E-09 | 9.24E-08 |
| LPXN | 1313.235301 | 0.459171532 | 0.088620232 | 7.17E-08 | 1.11E-06 |
| KNSTRN | 858.6637014 | 0.459096228 | 0.094357915 | 3.67E-07 | 5.05E-06 |
| HPCAL4 | 295.9265827 | 0.459045586 | 0.142288506 | 0.000375861 | 0.002540512 |
| NSA2P7 | 284.8629084 | -0.458340387 | 0.14476011 | 0.000465593 | 0.003070325 |
| SLC20A1 | 2542.837175 | -0.458299892 | 0.081805629 | 7.03E-09 | 1.28E-07 |
| CDCA3 | 539.5276942 | 0.45804833 | 0.107222111 | 6.18E-06 | 6.52E-05 |
| LPCAT1 | 3517.45921 | -0.45797264 | 0.081011677 | 4.96E-09 | 9.28E-08 |
| LOC124900276 | 171.5950313 | 0.457945465 | 0.173258184 | 0.002338383 | 0.012255987 |
| TOP2A | 9339.931362 | 0.4578937 | 0.082334374 | 8.71E-09 | 1.57E-07 |
| VAMP1 | 49.40946689 | -0.457882118 | 0.317117828 | 0.029786777 | 0.097232349 |
| PGM2L1 | 89.62171311 | 0.457822521 | 0.224694613 | 0.01052823 | 0.042548232 |
| ANKRD10 | 1223.161724 | -0.457739608 | 0.092194336 | 2.25E-07 | 3.22E-06 |
| C1QTNF3-AMACR | 167.0825024 | -0.457620718 | 0.174502511 | 0.002510122 | 0.013014305 |
| INTS10 | 1652.826259 | -0.45742714 | 0.084642467 | 2.17E-08 | 3.66E-07 |
| SLC17A5 | 324.3742789 | -0.457205972 | 0.131766939 | 0.000162397 | 0.001215777 |
| ALDH3B1 | 538.7849368 | -0.457070265 | 0.125344313 | 8.38E-05 | 0.000679676 |
| DPH7 | 399.5673992 | 0.45704668 | 0.122343954 | 5.81E-05 | 0.00049136 |
| ANKRD46 | 485.0174542 | -0.4568382 | 0.178104839 | 0.002902101 | 0.014677475 |
| TMBIM4 | 1984.036344 | 0.456629784 | 0.09006755 | 1.30E-07 | 1.94E-06 |
| GFI1 | 258.0806691 | -0.456559122 | 0.152530452 | 0.000833681 | 0.005085191 |
| NUMBL | 115.9195438 | -0.456535262 | 0.205311152 | 0.007028224 | 0.030556806 |
| LOC124905322 | 46.08011282 | -0.455989228 | 0.29888463 | 0.027201575 | 0.090569059 |
| LGALS3 | 10885.34447 | -0.455597257 | 0.082334504 | 1.05E-08 | 1.87E-07 |
| UBE2H | 5344.379038 | -0.455251968 | 0.070063129 | 2.75E-11 | 7.18E-10 |
| EEF1A1 | 301640.1382 | -0.455196725 | 0.062768573 | 1.54E-13 | 5.39E-12 |
| PPFIA3 | 310.6196662 | -0.455196505 | 0.139655367 | 0.000343114 | 0.002343871 |
| GSTP1 | 15705.65338 | 0.45506692 | 0.085924648 | 3.74E-08 | 6.12E-07 |
| PRUNE1 | 3486.565202 | 0.4550366 | 0.077455149 | 1.41E-09 | 2.90E-08 |
| BRMS1L | 195.41333 | 0.454979153 | 0.179754581 | 0.003209042 | 0.015991648 |
| MRPL13 | 1891.683236 | -0.454945732 | 0.078389355 | 2.20E-09 | 4.41E-08 |
| RGS14 | 595.4046694 | 0.454942314 | 0.121183694 | 5.46E-05 | 0.000465863 |
| STN1 | 356.1938536 | 0.45492312 | 0.129109266 | 0.000132997 | 0.001019979 |
| S100A11 | 3566.139132 | -0.454900066 | 0.083537934 | 1.74E-08 | 2.98E-07 |
| XRCC3 | 316.8794732 | 0.453999962 | 0.137961957 | 0.000306409 | 0.002124136 |
| CUX2 | 382.1855328 | 0.453883445 | 0.130630285 | 0.000158464 | 0.001189477 |
| BIK | 1016.625622 | -0.453852752 | 0.117099634 | 3.45E-05 | 0.000309442 |
| BBS2 | 278.7340683 | -0.453821888 | 0.14498431 | 0.000537254 | 0.003468915 |
| RIOX2 | 2180.411264 | 0.453646629 | 0.081849317 | 9.82E-09 | 1.76E-07 |
| SMYD4 | 208.1103669 | 0.453529311 | 0.155957015 | 0.001081578 | 0.006355511 |
| SHB | 115.446542 | -0.453474678 | 0.194372137 | 0.005477879 | 0.02485277 |
| EFNA3 | 217.8793308 | -0.453219101 | 0.147812143 | 0.000665368 | 0.004183798 |
| MYO5B | 150.4785023 | -0.45319932 | 0.180505283 | 0.003441843 | 0.016918082 |
| FBN1 | 83.99630297 | -0.452892532 | 0.248565211 | 0.016650332 | 0.06107473 |
| EHMT1 | 663.7913778 | 0.452605443 | 0.11040459 | 1.33E-05 | 0.000130668 |
| PDPR | 1745.926827 | -0.452591338 | 0.079708899 | 4.65E-09 | 8.83E-08 |
| CWH43 | 6.059038035 | 0.45113481 | 0.991862121 | 0.010909745 | 0.043702073 |
| NRM | 232.8641644 | 0.45103777 | 0.165382387 | 0.001891189 | 0.010256425 |

| | | | | | |
|---|---|---|---|---|---|
| TOR1A | 1589.528503 | 0.450973624 | 0.079872897 | 5.53E-09 | 1.03E-07 |
| SHISA5 | 1239.133026 | -0.450907014 | 0.097026567 | 1.13E-06 | 1.40E-05 |
| STK32C | 121.2808965 | -0.450418236 | 0.210200079 | 0.008667698 | 0.036168319 |
| TAF9 | 2649.592623 | -0.450143515 | 0.074378031 | 4.90E-10 | 1.08E-08 |
| DDX1 | 15051.55555 | -0.450141583 | 0.066599248 | 4.82E-12 | 1.41E-10 |
| UAP1L1 | 973.8290418 | 0.450121349 | 0.118326386 | 4.58E-05 | 0.000397074 |
| GMPPB | 537.7133461 | -0.450093882 | 0.125280539 | 0.000105159 | 0.000831905 |
| ZNF709 | 89.6221067 | 0.449920215 | 0.217887958 | 0.010285777 | 0.041753118 |
| LAMTOR4 | 1118.792453 | -0.449821845 | 0.123188569 | 8.45E-05 | 0.000683693 |
| FBXW5 | 1138.66441 | 0.449767942 | 0.1039152 | 4.94E-06 | 5.32E-05 |
| SNORD10 | 21.39604713 | 0.449725561 | 0.801349266 | 0.018925287 | 0.067768949 |
| CPNE2 | 142.6653786 | -0.449591238 | 0.185548657 | 0.00441502 | 0.020750861 |
| CD55 | 1900.256881 | -0.449406643 | 0.088799062 | 1.42E-07 | 2.10E-06 |
| ARHGAP12 | 459.4580784 | 0.449025794 | 0.143424797 | 0.000532897 | 0.003444591 |
| WDSUB1 | 109.6805268 | 0.448723939 | 0.213872268 | 0.009579949 | 0.039367816 |
| UNC93B1 | 1806.957675 | 0.448721316 | 0.126925273 | 0.00013376 | 0.001024466 |
| FPGT | 180.6458798 | 0.448712646 | 0.193644289 | 0.00570171 | 0.025670207 |
| TXN | 11476.44415 | -0.448526123 | 0.067521629 | 1.07E-11 | 3.01E-10 |
| CC2D2A | 6.111268277 | -0.448469866 | 0.998634773 | 0.010069047 | 0.04101961 |
| ABHD14A | 513.6516534 | -0.448142775 | 0.121492424 | 7.38E-05 | 0.000608491 |
| ARIH2 | 2638.130057 | -0.448001318 | 0.073890241 | 4.64E-10 | 1.03E-08 |
| TBKBP1 | 73.07732038 | -0.447750376 | 0.245028771 | 0.017119974 | 0.062598519 |
| SPAG6 | 319.6279644 | 0.447453595 | 0.130474482 | 0.000192487 | 0.001413009 |
| TMSB4XP1 | 1590.718576 | 0.447345604 | 0.096215948 | 1.11E-06 | 1.38E-05 |
| ELOVL7 | 407.3547959 | 0.447319649 | 0.151671533 | 0.000962486 | 0.005757297 |
| NLN | 1283.008709 | -0.447120063 | 0.085470492 | 5.77E-08 | 9.09E-07 |
| NAXD | 265.1905376 | -0.447116037 | 0.155671165 | 0.001264401 | 0.007233444 |
| KCNA3 | 1982.618519 | 0.446921981 | 0.11585989 | 3.68E-05 | 0.00032705 |
| SLC2A13 | 235.7347399 | 0.446905974 | 0.172266332 | 0.002759722 | 0.014084911 |
| ZNF714 | 362.6842441 | 0.446765847 | 0.154298536 | 0.001144126 | 0.006671182 |
| WDTC1 | 1218.120821 | 0.446356681 | 0.090607236 | 2.83E-07 | 3.98E-06 |
| TBC1D13 | 686.7508108 | 0.446134272 | 0.107000872 | 1.02E-05 | 0.0001028 |
| UBE2Q2 | 1348.687108 | 0.446100658 | 0.126186248 | 0.000129558 | 0.000998023 |
| S100A5 | 700.5655752 | -0.445993287 | 0.147683634 | 0.000795089 | 0.004888372 |
| EEF1A1P22 | 77.11753163 | -0.445926855 | 0.241483211 | 0.016616343 | 0.060962967 |
| GLYR1 | 1298.950476 | -0.445896691 | 0.0851048 | 5.63E-08 | 8.91E-07 |
| FAM53C | 3241.350148 | -0.445787801 | 0.089822213 | 2.38E-07 | 3.39E-06 |
| DARS2 | 4464.077625 | -0.445761136 | 0.076978581 | 2.43E-09 | 4.83E-08 |
| PLAGL1 | 147.095834 | 0.445582899 | 0.182385142 | 0.004253609 | 0.020133008 |
| GNL3 | 3156.268103 | -0.445396715 | 0.069477219 | 5.07E-11 | 1.26E-09 |
| EIF1B | 927.5158208 | -0.445386502 | 0.09212772 | 4.54E-07 | 6.12E-06 |
| RPUSD3 | 505.9882731 | -0.445170189 | 0.130903958 | 0.000218947 | 0.001580276 |
| TMEM185B | 938.9481603 | 0.445060807 | 0.0916937 | 4.06E-07 | 5.52E-06 |
| STX11 | 658.1132334 | -0.444802272 | 0.112799354 | 2.68E-05 | 0.000246954 |
| EFNA4 | 601.061756 | -0.444654923 | 0.111970405 | 2.41E-05 | 0.000224442 |
| ASTN2 | 151.4850311 | -0.444572259 | 0.174602326 | 0.003255299 | 0.016175611 |
| LRP8 | 793.7056163 | 0.444494952 | 0.097579364 | 1.76E-06 | 2.11E-05 |
| BORCS5 | 523.5342737 | -0.44436013 | 0.122375943 | 9.35E-05 | 0.000750169 |
| EEF1A1P11 | 18993.51215 | -0.444233784 | 0.068815718 | 4.55E-11 | 1.14E-09 |
| ACVR1 | 107.5427537 | 0.44421746 | 0.231839175 | 0.014417456 | 0.054651753 |

| | | | | | |
|---|---|---|---|---|---|
| BTLA | 9.512340089 | 0.44408351 | 0.682331512 | 0.029281023 | 0.096033991 |
| ZNF431 | 235.6564581 | 0.44398176 | 0.191151027 | 0.005718155 | 0.025726496 |
| PPT2 | 488.6900615 | -0.443955297 | 0.112844257 | 2.80E-05 | 0.000256384 |
| PRF1 | 5.72067429 | 0.443684392 | 0.935639837 | 0.012375168 | 0.048298578 |
| RABGGTB | 1822.043054 | -0.443683537 | 0.09147954 | 4.24E-07 | 5.73E-06 |
| SCARB1 | 1815.888847 | -0.44337865 | 0.099264215 | 2.72E-06 | 3.10E-05 |
| ZNF678 | 282.7988086 | 0.443309686 | 0.138165798 | 0.000422181 | 0.002815134 |
| CARD16 | 170.7599431 | 0.44314445 | 0.16759851 | 0.002474501 | 0.012856565 |
| ZFP37 | 123.1474632 | 0.443061445 | 0.201445686 | 0.007794458 | 0.033256085 |
| TTF1 | 554.9792625 | 0.443012545 | 0.106864039 | 1.12E-05 | 0.000112707 |
| GALNT9 | 63.39834664 | 0.442949641 | 0.273076306 | 0.024727942 | 0.084144975 |
| ADAM23 | 1106.478162 | -0.442942923 | 0.087443004 | 1.41E-07 | 2.09E-06 |
| SLC39A13 | 174.840631 | -0.442551372 | 0.189884132 | 0.005815214 | 0.026082339 |
| TOM1 | 443.9894795 | 0.442521197 | 0.142547157 | 0.000605253 | 0.003857622 |
| TRNG | 165.6437943 | 0.442487878 | 0.244702706 | 0.017780106 | 0.064603041 |
| CLK4 | 198.053736 | -0.442344548 | 0.161532572 | 0.001892889 | 0.010256425 |
| RPS2 | 58652.33887 | -0.442248014 | 0.092897593 | 4.20E-07 | 5.69E-06 |
| EIF4E | 2375.467304 | -0.442199199 | 0.113540309 | 3.32E-05 | 0.000298648 |
| PPP1CC | 9494.082306 | -0.44217566 | 0.101816709 | 4.65E-06 | 5.05E-05 |
| TATDN1 | 695.4551502 | -0.44185318 | 0.113446686 | 3.31E-05 | 0.000298255 |
| SLC2A8 | 205.2086282 | 0.440997451 | 0.210824218 | 0.010099804 | 0.041113847 |
| BRF2 | 588.4477168 | -0.440825182 | 0.109594081 | 1.96E-05 | 0.000185957 |
| NOSIP | 1636.454447 | 0.440658977 | 0.10105135 | 4.34E-06 | 4.74E-05 |
| UBE2D3 | 9476.467775 | -0.440544795 | 0.098464306 | 2.64E-06 | 3.02E-05 |
| AAK1 | 555.8503465 | 0.44048647 | 0.112238275 | 2.89E-05 | 0.000263966 |
| FAM117A | 2193.796854 | 0.440411983 | 0.077567041 | 4.69E-09 | 8.89E-08 |
| ADD3 | 4757.238127 | 0.440230053 | 0.100846588 | 4.14E-06 | 4.54E-05 |
| RAP1GDS1 | 1618.188308 | -0.440088624 | 0.078141194 | 6.32E-09 | 1.16E-07 |
| LINC02009 | 9.553305491 | -0.440033063 | 0.841781974 | 0.015954562 | 0.059149036 |
| MIDN | 857.5813546 | -0.439829893 | 0.107964987 | 1.59E-05 | 0.000154455 |
| HNRNPD | 5525.304036 | -0.439234986 | 0.085461206 | 9.64E-08 | 1.47E-06 |
| BLVRB | 847.3263935 | -0.439093255 | 0.101687107 | 5.48E-06 | 5.85E-05 |
| RMDN1 | 1095.802171 | -0.439006308 | 0.094661229 | 1.22E-06 | 1.51E-05 |
| TAF2 | 1386.857948 | -0.438963981 | 0.093199928 | 8.56E-07 | 1.10E-05 |
| PRKDC | 8855.268547 | -0.438922036 | 0.081974335 | 3.03E-08 | 5.01E-07 |
| FZD2 | 89.26167833 | 0.438608256 | 0.254458702 | 0.021425794 | 0.07509344 |
| NAA15 | 3361.848039 | -0.438567395 | 0.082540916 | 3.77E-08 | 6.16E-07 |
| CCDC97 | 1221.469883 | 0.43848251 | 0.093849118 | 1.02E-06 | 1.28E-05 |
| EEF1A1P16 | 81.59206673 | -0.438434742 | 0.230661254 | 0.01545171 | 0.05765138 |
| SIL1 | 1549.689975 | -0.438361317 | 0.092150458 | 6.89E-07 | 8.97E-06 |
| CALCOCO2 | 1467.923766 | 0.438022216 | 0.085440502 | 1.02E-07 | 1.55E-06 |
| LOC105374445 | 66.7869446 | 0.437547994 | 0.259769955 | 0.022758025 | 0.078838764 |
| NDUFAB1 | 3464.82273 | -0.437406861 | 0.072850129 | 6.94E-10 | 1.49E-08 |
| IRAK2 | 285.2982962 | 0.437225194 | 0.132584194 | 0.000319006 | 0.002200887 |
| MIR6747 | 86.82431507 | -0.437032453 | 0.234049358 | 0.016780237 | 0.061486111 |
| SCN9A | 375.3714482 | -0.436934076 | 0.123708169 | 0.000137319 | 0.001048946 |
| PPIP5K2 | 1188.538988 | -0.436792243 | 0.110104112 | 2.48E-05 | 0.000229552 |
| NEK3 | 383.6359143 | -0.436747557 | 0.120732068 | 9.94E-05 | 0.000791748 |
| B9D2 | 197.6700326 | 0.43650702 | 0.160333308 | 0.002042346 | 0.010919194 |
| MVD | 992.1323019 | -0.436427348 | 0.107906162 | 1.82E-05 | 0.000174321 |

| | | | | | |
|---|---|---|---|---|---|
| ZC3H10 | 187.6463053 | 0.4358056 | 0.165030312 | 0.002540928 | 0.013119037 |
| HAGH | 489.5127286 | -0.435769526 | 0.126233982 | 0.000187749 | 0.001382331 |
| MGAT5 | 1198.712055 | -0.435674166 | 0.091864383 | 7.42E-07 | 9.62E-06 |
| PARP8 | 334.4916703 | 0.435546039 | 0.124344536 | 0.000153968 | 0.001160252 |
| SOS1 | 1280.936916 | 0.435527571 | 0.095732995 | 1.85E-06 | 2.20E-05 |
| C1orf159 | 189.1768917 | 0.435265322 | 0.179605217 | 0.004657681 | 0.02174631 |
| CDC16 | 1883.656486 | -0.435112002 | 0.077531208 | 7.19E-09 | 1.31E-07 |
| CSPG5 | 6.921261341 | -0.435106095 | 0.803673726 | 0.018393213 | 0.066302095 |
| YAF2 | 717.1136811 | 0.435071659 | 0.17369367 | 0.003714648 | 0.017948258 |
| GPAT3 | 74.86032254 | -0.434772025 | 0.247576625 | 0.020644866 | 0.072767584 |
| IGHJ4 | 157.9354699 | -0.43466304 | 0.1855432 | 0.005808915 | 0.026067112 |
| MYL6B | 966.2461734 | -0.434321444 | 0.092765985 | 1.01E-06 | 1.27E-05 |
| GNPTG | 367.1344594 | -0.434302487 | 0.124662314 | 0.000168364 | 0.00125671 |
| RIT1 | 753.3259692 | -0.434292574 | 0.108151692 | 2.04E-05 | 0.000192926 |
| TEDC1 | 309.2663975 | 0.434223646 | 0.174581617 | 0.00393242 | 0.018837541 |
| LMNB1 | 2609.116733 | 0.434116829 | 0.076894633 | 5.85E-09 | 1.08E-07 |
| SLC12A6 | 211.9371862 | 0.433939976 | 0.167909952 | 0.002989657 | 0.015028126 |
| MYEF2 | 292.9890346 | 0.433927711 | 0.161142209 | 0.00221169 | 0.011687969 |
| RGPD1 | 307.2069967 | 0.433744909 | 0.129705777 | 0.000274412 | 0.001926226 |
| CDCA8 | 1977.276582 | 0.433620259 | 0.087184038 | 2.33E-07 | 3.32E-06 |
| SURF6 | 650.4681272 | 0.433585522 | 0.120369869 | 0.000106671 | 0.00084233 |
| LOC102723847 | 80.46179688 | -0.433362161 | 0.22570801 | 0.015226946 | 0.056964074 |
| GTF3C1 | 1260.317984 | -0.433358299 | 0.088199537 | 3.19E-07 | 4.45E-06 |
| UNC119 | 293.798017 | 0.43329051 | 0.132899541 | 0.000369698 | 0.00250374 |
| SLC7A5P2 | 199.1342261 | -0.432925254 | 0.219026714 | 0.013636263 | 0.052359016 |
| COPA | 14317.90463 | -0.432858965 | 0.062551771 | 1.67E-12 | 5.22E-11 |
| PMVK | 2214.221949 | -0.432499059 | 0.080685983 | 3.01E-08 | 4.98E-07 |
| RPS23P8 | 14527.3681 | -0.432043375 | 0.074037181 | 1.98E-09 | 3.97E-08 |
| HNRNPA1P8 | 1152.31738 | -0.43182965 | 0.101393765 | 7.21E-06 | 7.49E-05 |
| ISY1-RAB43 | 748.5652881 | 0.431561012 | 0.096720001 | 2.87E-06 | 3.25E-05 |
| GEMIN2 | 248.035321 | 0.431528845 | 0.143267671 | 0.000849674 | 0.005164553 |
| PPA2 | 3169.791369 | -0.431461393 | 0.075912788 | 4.77E-09 | 8.99E-08 |
| NSUN3 | 449.2922807 | 0.431304929 | 0.128276818 | 0.000256394 | 0.001817063 |
| EEF1B2 | 47193.54818 | -0.431278117 | 0.067098075 | 4.80E-11 | 1.20E-09 |
| CPLX1 | 92.86388816 | -0.43121952 | 0.2302893 | 0.016722197 | 0.061299381 |
| GTPBP1 | 1197.468761 | -0.431121181 | 0.124400089 | 0.000182158 | 0.001347472 |
| SSR2 | 24724.83894 | -0.430962683 | 0.067809658 | 7.78E-11 | 1.89E-09 |
| EVI5L | 481.4223338 | 0.430925075 | 0.122087674 | 0.000141479 | 0.001075501 |
| SCFD2 | 721.0538399 | -0.430832303 | 0.099779498 | 5.63E-06 | 6.00E-05 |
| CCT3 | 29288.30403 | -0.430824089 | 0.060651352 | 4.78E-13 | 1.58E-11 |
| IGHA1 | 187273.9984 | -0.430617054 | 0.099782414 | 3.70E-06 | 4.10E-05 |
| SLC20A2 | 927.9748688 | -0.430134148 | 0.089758426 | 5.95E-07 | 7.85E-06 |
| ELAC1 | 215.3735177 | 0.430037574 | 0.165929196 | 0.002971358 | 0.01494916 |
| GGA2 | 1930.707869 | -0.42967791 | 0.080297403 | 3.20E-08 | 5.27E-07 |
| LZTS2 | 180.6411809 | 0.429583901 | 0.194651566 | 0.008162512 | 0.034464645 |
| ADRB1 | 10.6229612 | 0.429560149 | 0.668521417 | 0.029534715 | 0.096591627 |
| GSPT1 | 6400.21169 | -0.429405587 | 0.068046225 | 1.03E-10 | 2.45E-09 |
| CEP192 | 439.9146609 | 0.429345019 | 0.116750071 | 8.04E-05 | 0.000654841 |
| HSPA8P8 | 3778.520835 | -0.429212652 | 0.085066595 | 1.55E-07 | 2.29E-06 |
| SEMA4F | 226.162253 | 0.429116137 | 0.149605889 | 0.001348754 | 0.007647797 |

| | | | | | |
|---|---|---|---|---|---|
| KIF4A | 896.2902122 | 0.428269513 | 0.095956639 | 2.85E-06 | 3.24E-05 |
| KCTD18 | 501.7260742 | 0.428218422 | 0.113106601 | 5.29E-05 | 0.000452249 |
| ZNF605 | 138.1726802 | 0.428103484 | 0.206944589 | 0.0111548 | 0.044502796 |
| DVL3 | 1367.049057 | 0.427962334 | 0.08850532 | 4.76E-07 | 6.39E-06 |
| PCCA | 896.4094581 | -0.427923015 | 0.089706762 | 6.69E-07 | 8.73E-06 |
| C4orf33 | 340.7702963 | -0.427845093 | 0.14630335 | 0.001136777 | 0.006641556 |
| UPP1 | 287.1494607 | 0.427569943 | 0.139430428 | 0.000728205 | 0.004536087 |
| CHST12 | 1550.049702 | 0.427505559 | 0.105326287 | 1.74E-05 | 0.000167948 |
| NFKB1 | 1890.204967 | -0.427301363 | 0.079012299 | 2.35E-08 | 3.95E-07 |
| ITGA4 | 13333.68548 | 0.427219143 | 0.095011571 | 2.44E-06 | 2.81E-05 |
| DCP1A | 1230.808502 | -0.426897267 | 0.09753464 | 4.34E-06 | 4.74E-05 |
| KRT86 | 243.2894287 | 0.42688031 | 0.15028444 | 0.001479662 | 0.008303083 |
| MROH1 | 135.7579924 | 0.426828872 | 0.187417768 | 0.007018704 | 0.030530746 |
| ASPHD2 | 193.2093339 | 0.426708812 | 0.165256898 | 0.003127191 | 0.015628736 |
| ENOPH1 | 1310.785102 | -0.426701809 | 0.116345663 | 8.54E-05 | 0.000690406 |
| PUS7 | 1029.258951 | -0.426682644 | 0.091255775 | 1.06E-06 | 1.32E-05 |
| TOR1B | 986.5080587 | 0.42654191 | 0.102662523 | 1.16E-05 | 0.000116069 |
| ARHGAP11A | 1521.07513 | 0.426534689 | 0.104005758 | 1.44E-05 | 0.000141321 |
| IKZF3 | 500.3823609 | 0.426364693 | 0.139904268 | 0.000763162 | 0.004721568 |
| NUSAP1 | 3377.532709 | 0.426262598 | 0.076671257 | 9.89E-09 | 1.77E-07 |
| CDK6 | 3733.62944 | -0.426220614 | 0.130403143 | 0.000370947 | 0.002510232 |
| CCDC113 | 101.0146178 | -0.426068348 | 0.254903465 | 0.024792475 | 0.084331427 |
| REEP5 | 1998.692702 | 0.426050955 | 0.090680995 | 9.47E-07 | 1.20E-05 |
| RASGRP3 | 2233.206111 | 0.425978136 | 0.092174519 | 1.37E-06 | 1.68E-05 |
| ANKDD1A | 278.625924 | 0.425707371 | 0.13914803 | 0.000743619 | 0.004619622 |
| DERL1 | 5023.426093 | -0.425659543 | 0.0695469 | 3.50E-10 | 7.78E-09 |
| MSC | 259.1612424 | -0.425584908 | 0.146587431 | 0.001244471 | 0.007133567 |
| MPHOSPH6 | 799.7753007 | -0.425258231 | 0.121441235 | 0.000160447 | 0.001202798 |
| MGST3 | 2303.426174 | -0.425153791 | 0.088310913 | 5.31E-07 | 7.08E-06 |
| B3GLCT | 172.9894611 | 0.425124359 | 0.175064655 | 0.004747424 | 0.022117608 |
| ODAPH | 9.609150816 | 0.425035901 | 0.657188217 | 0.030159629 | 0.098098101 |
| CHMP5 | 875.4231655 | 0.424937293 | 0.112277585 | 5.39E-05 | 0.000460496 |
| SORD2P | 555.8215294 | -0.424652611 | 0.102187435 | 1.17E-05 | 0.000116692 |
| GOLGA8S | 653.4137756 | 0.42449892 | 0.151603541 | 0.001669358 | 0.009217845 |
| AGK | 3710.793867 | -0.424471501 | 0.067899432 | 1.52E-10 | 3.54E-09 |
| RPS2P5 | 8300.962306 | -0.424403998 | 0.091365922 | 1.25E-06 | 1.54E-05 |
| PFKL | 3011.310245 | 0.424285167 | 0.091981214 | 1.44E-06 | 1.75E-05 |
| NCK2 | 341.8827756 | 0.424166769 | 0.12510368 | 0.000242311 | 0.001733212 |
| DENND3 | 157.2459235 | 0.424049077 | 0.176235821 | 0.005097593 | 0.02344622 |
| DDX6 | 7316.782458 | 0.423936843 | 0.087453956 | 4.56E-07 | 6.15E-06 |
| RPSAP19 | 2323.726953 | -0.423730922 | 0.098094618 | 5.73E-06 | 6.10E-05 |
| PAQR8 | 256.4406794 | 0.423696467 | 0.140254951 | 0.000849282 | 0.005163983 |
| TRAPPC2L | 1351.670673 | -0.423306384 | 0.098998613 | 6.99E-06 | 7.28E-05 |
| AGMAT | 150.6520613 | -0.423202289 | 0.178709199 | 0.005679634 | 0.025588625 |
| HLA-J | 952.2477565 | 0.423079881 | 0.094208464 | 2.60E-06 | 2.98E-05 |
| ARHGEF18 | 732.9952004 | 0.42292258 | 0.122643678 | 0.000196369 | 0.001437244 |
| NIPA1 | 745.6445223 | 0.422824897 | 0.109142394 | 3.77E-05 | 0.000334262 |
| TMSB10 | 65249.85219 | 0.422767136 | 0.064834277 | 2.61E-11 | 6.84E-10 |
| CMIP | 889.4364434 | -0.422734849 | 0.091576921 | 1.44E-06 | 1.75E-05 |
| DBF4B | 326.5736858 | 0.422677505 | 0.12527095 | 0.000256447 | 0.001817063 |

| | | | | | |
|---|---|---|---|---|---|
| ENTR1 | 659.2495953 | 0.422659789 | 0.104558565 | 1.89E-05 | 0.000181069 |
| ZNF749 | 136.5895676 | 0.422462979 | 0.177820301 | 0.005544895 | 0.025128038 |
| OTUD1 | 127.205873 | 0.422416919 | 0.189552581 | 0.007935761 | 0.033698765 |
| SNHG3 | 750.5074726 | -0.422409112 | 0.094814311 | 3.10E-06 | 3.49E-05 |
| IFNLR1 | 930.1944758 | 0.422325931 | 0.100104901 | 8.86E-06 | 9.02E-05 |
| NAAA | 806.3489833 | -0.422284071 | 0.097715638 | 5.70E-06 | 6.08E-05 |
| CLMN | 155.5271559 | -0.422227869 | 0.168879198 | 0.004047197 | 0.01928069 |
| ZNF440 | 122.929031 | 0.422205876 | 0.186649874 | 0.007368537 | 0.031757329 |
| PYM1 | 733.2823848 | 0.421982475 | 0.103856957 | 1.75E-05 | 0.000168825 |
| YPEL3 | 248.5508798 | 0.42169778 | 0.158841674 | 0.002623038 | 0.013466693 |
| RCBTB2 | 657.8202796 | 0.421490509 | 0.09813102 | 6.36E-06 | 6.68E-05 |
| PPDPF | 783.9924972 | 0.421373921 | 0.115651429 | 9.54E-05 | 0.000763377 |
| ENTPD5 | 403.0093904 | 0.421169155 | 0.117169549 | 0.000113396 | 0.000886936 |
| SIRT5 | 223.0708336 | 0.421118101 | 0.144673532 | 0.001219063 | 0.007025129 |
| SULT1A4 | 138.6131759 | -0.420918191 | 0.177393538 | 0.005680122 | 0.025588625 |
| STS | 6574.538965 | 0.420883999 | 0.078772618 | 3.38E-08 | 5.55E-07 |
| ZNF598 | 626.2431372 | -0.420748982 | 0.12588312 | 0.00029433 | 0.002049423 |
| GAR1 | 1310.373856 | -0.420639208 | 0.12168607 | 0.00019366 | 0.001419392 |
| ABCB9 | 216.5858961 | -0.420460784 | 0.153711985 | 0.002117533 | 0.011258616 |
| KIF11 | 2827.853892 | 0.420421571 | 0.099482616 | 8.45E-06 | 8.66E-05 |
| RPS7P14 | 90.58759155 | -0.420273534 | 0.215421718 | 0.015184556 | 0.056854262 |
| B3GAT3 | 722.5439623 | -0.420263965 | 0.104455778 | 2.11E-05 | 0.000199061 |
| WDR53 | 421.8764842 | 0.420258959 | 0.122725698 | 0.000217456 | 0.001570341 |
| SMIM30 | 721.2970064 | -0.420241788 | 0.144276732 | 0.001197724 | 0.00692983 |
| RPL13P12 | 8641.897158 | -0.420112577 | 0.122528014 | 0.000216083 | 0.001562384 |
| WSB2 | 434.0976249 | 0.420090406 | 0.120264399 | 0.000169621 | 0.001264451 |
| PPIL6 | 8.338367886 | 0.420072256 | 0.763473636 | 0.018295534 | 0.065991201 |
| VOPP1 | 4949.788336 | 0.420036585 | 0.084664402 | 2.60E-07 | 3.68E-06 |
| HAS3 | 59.5557551 | -0.419863354 | 0.262969283 | 0.02919702 | 0.095812923 |
| EVL | 176.2170571 | 0.419858533 | 0.179713683 | 0.006197643 | 0.027540308 |
| PIK3IP1 | 389.2138863 | -0.419777469 | 0.117446648 | 0.000126538 | 0.000977366 |
| CAP1 | 7323.795974 | 0.419767156 | 0.066110348 | 8.24E-11 | 1.99E-09 |
| PGAP6 | 361.2659598 | -0.419545636 | 0.139628934 | 0.000922783 | 0.005550478 |
| RPL14P1 | 4324.168171 | -0.419437956 | 0.08154167 | 9.63E-08 | 1.46E-06 |
| ANKRD13D | 280.090438 | 0.419210577 | 0.148675187 | 0.001632201 | 0.009053433 |
| HOXB7 | 232.2120645 | 0.418823182 | 0.142042003 | 0.001094795 | 0.006426632 |
| KCTD6 | 301.7095011 | -0.418656633 | 0.175427104 | 0.005454627 | 0.024772562 |
| TCAF1P1 | 81.33933486 | 0.418252217 | 0.23808884 | 0.022020555 | 0.076819088 |
| STIL | 1079.007187 | 0.418251776 | 0.090122034 | 1.29E-06 | 1.58E-05 |
| GALT | 194.5652271 | -0.418099208 | 0.156029347 | 0.002491212 | 0.012926855 |
| EIF4BP2 | 193.1709909 | -0.417755806 | 0.153285377 | 0.002188104 | 0.011593177 |
| MIGA2 | 398.6340038 | 0.417644851 | 0.146333299 | 0.001477157 | 0.008294399 |
| ATP6V0C | 1528.425209 | -0.417607635 | 0.117638358 | 0.000139939 | 0.001065198 |
| YTHDF3 | 2698.264225 | -0.417516931 | 0.116957809 | 0.00013055 | 0.001004768 |
| IGHV3-64 | 886.2653488 | -0.417507817 | 0.122971741 | 0.000248276 | 0.001770028 |
| ZNF548 | 287.1843067 | 0.417496379 | 0.142536095 | 0.001168988 | 0.0067908 |
| E2F8 | 469.6538436 | 0.417492142 | 0.112030508 | 6.98E-05 | 0.000579125 |
| FRK | 87.14996964 | 0.417474924 | 0.245961453 | 0.024623957 | 0.083857044 |
| SRGAP2 | 445.1286907 | 0.417432871 | 0.113836974 | 8.78E-05 | 0.000707799 |
| PTTG1 | 2244.477773 | 0.417350544 | 0.089855334 | 1.27E-06 | 1.56E-05 |

| | | | | | |
|---|---|---|---|---|---|
| NAE1 | 2249.206713 | -0.417232369 | 0.084193606 | 2.74E-07 | 3.88E-06 |
| SULT1A3 | 138.7767621 | -0.416946345 | 0.177014661 | 0.006037491 | 0.02695314 |
| TRABD2A | 835.252875 | 0.416904108 | 0.095574216 | 4.79E-06 | 5.19E-05 |
| DDX21 | 11246.13137 | -0.416696572 | 0.084470332 | 3.07E-07 | 4.31E-06 |
| HSP90AA2P | 1234.024102 | -0.416645831 | 0.094034445 | 3.53E-06 | 3.92E-05 |
| ETHE1 | 381.940726 | 0.416450449 | 0.135967441 | 0.000767482 | 0.004744905 |
| SLC7A11 | 1115.105355 | 0.41642673 | 0.149129465 | 0.001768573 | 0.009676583 |
| PRXL2C | 177.1020122 | 0.416251554 | 0.192488354 | 0.009569127 | 0.039332676 |
| NUMB | 707.5809705 | 0.416096883 | 0.107015479 | 3.63E-05 | 0.000323689 |
| RPS7P1 | 20957.66508 | -0.415913892 | 0.073012105 | 4.78E-09 | 9.00E-08 |
| RIPK3 | 92.92531869 | 0.415759979 | 0.220325925 | 0.017499712 | 0.063784994 |
| IL4R | 123.9612516 | 0.415512639 | 0.186639255 | 0.008226271 | 0.034660421 |
| PLD6 | 471.3464565 | 0.415471327 | 0.112836794 | 8.45E-05 | 0.000683693 |
| PHF21A | 185.4001589 | 0.414861542 | 0.176158548 | 0.005980437 | 0.026732893 |
| UPF1 | 2476.026616 | 0.414734495 | 0.094271175 | 4.05E-06 | 4.45E-05 |
| SLC16A10 | 330.8289853 | -0.414624045 | 0.124381774 | 0.000310958 | 0.002148785 |
| RUBCN | 1668.048851 | -0.414539607 | 0.107815386 | 4.45E-05 | 0.000387031 |
| PIDD1 | 177.0961684 | -0.414126056 | 0.206729904 | 0.013995053 | 0.053381571 |
| ZNF134 | 271.4055024 | 0.414121926 | 0.137861846 | 0.000935402 | 0.00561468 |
| PRC1 | 1570.801727 | 0.414110837 | 0.079044712 | 6.14E-08 | 9.63E-07 |
| RPL10P3 | 315.083834 | -0.414021156 | 0.129668166 | 0.000505082 | 0.003291875 |
| DNAJC10 | 3489.576049 | -0.413837107 | 0.085000204 | 4.29E-07 | 5.80E-06 |
| NOCT | 152.656517 | -0.413831971 | 0.181590939 | 0.007304117 | 0.031541597 |
| SLC25A37 | 434.0897252 | -0.413725944 | 0.11903236 | 0.00018463 | 0.00136285 |
| RALY | 8311.928329 | 0.413599563 | 0.079445734 | 6.97E-08 | 1.08E-06 |
| GNL3L | 631.7403646 | 0.413559194 | 0.117516611 | 0.000158082 | 0.001187126 |
| CBX4 | 1331.373986 | -0.413430778 | 0.091027425 | 2.12E-06 | 2.49E-05 |
| COPS5 | 1649.267838 | -0.413200845 | 0.076379749 | 2.44E-08 | 4.10E-07 |
| MED12 | 735.8731855 | 0.412978636 | 0.102891136 | 2.21E-05 | 0.00020739 |
| KIAA1109 | 613.5240699 | -0.412804019 | 0.109577614 | 6.08E-05 | 0.000513454 |
| RPS25P8 | 768.2237394 | -0.412753433 | 0.11367806 | 0.000104931 | 0.000830481 |
| NFE2L3 | 3239.142092 | 0.412678343 | 0.0719669 | 3.75E-09 | 7.21E-08 |
| RPS6P1 | 1308.784869 | -0.412645785 | 0.10141815 | 1.80E-05 | 0.000172213 |
| LILRB1 | 417.4877792 | -0.412531812 | 0.114890403 | 0.000121888 | 0.000945246 |
| POLR3GL | 2195.157644 | 0.412514977 | 0.078533327 | 5.70E-08 | 9.00E-07 |
| TSHR | 172.9876325 | 0.412490565 | 0.16941304 | 0.004949335 | 0.022880381 |
| AJM1 | 105.0021951 | 0.412312955 | 0.208073873 | 0.014468418 | 0.05478889 |
| AMPD1 | 311.6199674 | -0.411997696 | 0.13806135 | 0.001005067 | 0.005968707 |
| SP140L | 627.3750213 | 0.411925015 | 0.10510626 | 3.28E-05 | 0.000295928 |
| RPL39P15 | 2810.924476 | -0.411837041 | 0.074264605 | 1.14E-08 | 2.02E-07 |
| PDK1 | 4645.990956 | -0.411770497 | 0.091050958 | 2.34E-06 | 2.72E-05 |
| ASPHD1 | 94.66057327 | -0.411611088 | 0.236958803 | 0.023884611 | 0.081789562 |
| TTN | 284.237112 | -0.411524946 | 0.155920838 | 0.002845965 | 0.014461141 |
| RPL26P6 | 112.6875806 | 0.411236767 | 0.256921733 | 0.029659944 | 0.096873113 |
| SPIN4 | 388.0743813 | 0.411179389 | 0.18677492 | 0.008844276 | 0.036807578 |
| FHOD1 | 662.2164766 | -0.411158035 | 0.11524519 | 0.000134975 | 0.001032861 |
| PRR12 | 273.3684941 | 0.410973854 | 0.15395222 | 0.00260716 | 0.013401068 |
| C12orf76 | 88.31579989 | -0.410823646 | 0.232812928 | 0.022597992 | 0.078460295 |
| EXOG | 257.2839854 | -0.410774422 | 0.150119011 | 0.002202063 | 0.01165794 |
| MACROD1 | 343.4828005 | -0.410744326 | 0.154955185 | 0.002799566 | 0.01425885 |

| | | | | | |
|---|---|---|---|---|---|
| DPP7 | 2900.229107 | 0.410691194 | 0.102913367 | 2.59E-05 | 0.000239699 |
| SLC25A32 | 1093.150862 | -0.410382229 | 0.109608347 | 6.78E-05 | 0.000564682 |
| SEC24A | 1708.547493 | -0.410381441 | 0.116772169 | 0.000162459 | 0.001215777 |
| BLM | 993.3989807 | 0.410251218 | 0.10000669 | 1.53E-05 | 0.000148986 |
| CCDC163 | 187.4609859 | 0.410183407 | 0.16290104 | 0.004011007 | 0.019145149 |
| PRIMPOL | 97.28689133 | 0.409994018 | 0.213766931 | 0.016746017 | 0.061373708 |
| FAM107B | 3424.909795 | 0.409674619 | 0.082988504 | 3.00E-07 | 4.21E-06 |
| NIN | 449.8846202 | 0.409645897 | 0.131781843 | 0.000668066 | 0.004196192 |
| TM9SF2 | 9376.203218 | -0.409618427 | 0.072047921 | 5.15E-09 | 9.59E-08 |
| TBC1D14 | 1803.65952 | 0.40936668 | 0.080800744 | 1.58E-07 | 2.31E-06 |
| ZNF420 | 215.0019865 | 0.409288681 | 0.154826244 | 0.002825553 | 0.014378517 |
| INHBE | 978.4106912 | -0.409046903 | 0.0920022 | 3.39E-06 | 3.79E-05 |
| RB1 | 4022.159844 | 0.408767775 | 0.12579323 | 0.000419463 | 0.002798087 |
| MORN2 | 266.705625 | -0.408648152 | 0.139655592 | 0.001239033 | 0.007116527 |
| CEP170 | 414.0243408 | 0.408165074 | 0.133031068 | 0.000769368 | 0.004753171 |
| F11R | 2851.396723 | -0.408132482 | 0.069014889 | 1.34E-09 | 2.76E-08 |
| ABCC1 | 1490.100194 | -0.408027252 | 0.081804479 | 2.40E-07 | 3.42E-06 |
| AIG1 | 997.9846681 | -0.407825324 | 0.088427608 | 1.56E-06 | 1.88E-05 |
| UQCRC2 | 6642.440026 | -0.407258341 | 0.064472464 | 1.08E-10 | 2.56E-09 |
| ENDOG | 345.5230197 | 0.406945307 | 0.167164595 | 0.005084904 | 0.023400286 |
| TTC13 | 309.4022681 | -0.406944707 | 0.159421696 | 0.003687883 | 0.017853809 |
| ZNF468 | 568.1338363 | 0.406909058 | 0.117156582 | 0.000189762 | 0.001394783 |
| DOLK | 582.2043644 | 0.406819738 | 0.12870119 | 0.000576631 | 0.003698342 |
| OSER1 | 1596.125259 | -0.406660023 | 0.077705382 | 6.60E-08 | 1.03E-06 |
| PECR | 706.603887 | -0.406452047 | 0.103611681 | 3.36E-05 | 0.000301655 |
| BAIAP2L1 | 156.9842078 | 0.406322253 | 0.172976045 | 0.006373222 | 0.02819752 |
| METAP1 | 2118.908532 | -0.405913527 | 0.07303125 | 1.09E-08 | 1.94E-07 |
| INTS6P1 | 83.66459939 | -0.405699199 | 0.225731904 | 0.021815689 | 0.076181144 |
| CYB561D1 | 255.3989452 | 0.405582417 | 0.147763078 | 0.002148911 | 0.011414936 |
| TRIM9 | 243.8102463 | -0.40545157 | 0.147887678 | 0.002180066 | 0.011562697 |
| GTPBP2 | 395.9382427 | 0.40541354 | 0.121666705 | 0.000319342 | 0.00220233 |
| DEPTOR | 412.9939923 | 0.405380828 | 0.114312715 | 0.000146461 | 0.001110449 |
| AIFM2 | 396.1883538 | 0.405264173 | 0.122681953 | 0.000357229 | 0.002426886 |
| OCIAD2 | 1889.749377 | -0.405240106 | 0.085613356 | 8.70E-07 | 1.11E-05 |
| TRUB2 | 1641.961438 | 0.405176591 | 0.080452357 | 1.88E-07 | 2.71E-06 |
| LMNA | 8836.352547 | -0.405139704 | 0.090004087 | 2.65E-06 | 3.03E-05 |
| POC1B-GALNT4 | 168.999805 | 0.404933701 | 0.186495956 | 0.009817598 | 0.04021082 |
| ARID2 | 1413.527358 | 0.404910569 | 0.107725323 | 6.43E-05 | 0.000539216 |
| DCTN5 | 3323.966722 | -0.404904754 | 0.071194934 | 5.26E-09 | 9.77E-08 |
| DDX19A | 1679.655762 | -0.404865467 | 0.082037446 | 3.20E-07 | 4.47E-06 |
| TAF8 | 430.5540059 | 0.404770902 | 0.113994615 | 0.00014475 | 0.001098922 |
| DUSP14 | 1073.964376 | -0.404636954 | 0.095422355 | 8.62E-06 | 8.80E-05 |
| ADA2 | 192.2437985 | 0.404540499 | 0.171980077 | 0.006396614 | 0.028273287 |
| LEFTY2 | 78.85030152 | -0.404516017 | 0.232038238 | 0.024470866 | 0.083417713 |
| TMCO1 | 18814.57081 | -0.404514912 | 0.073851343 | 1.75E-08 | 2.98E-07 |
| PSLNR | 9.142911986 | -0.404266483 | 0.680450851 | 0.024236512 | 0.08278179 |
| ANAPC2 | 638.5744312 | 0.40425423 | 0.119581341 | 0.000270957 | 0.001905834 |
| TACC3 | 874.1664009 | 0.404238585 | 0.098682505 | 1.61E-05 | 0.000156202 |
| HNRNPA1P7 | 3753.868367 | -0.404109384 | 0.074344211 | 2.19E-08 | 3.70E-07 |
| NT5DC3 | 402.1910412 | -0.403960716 | 0.150151418 | 0.002528937 | 0.01309099 |

| | | | | | |
|---|---|---|---|---|---|
| UNC13A | 711.7171891 | -0.403677355 | 0.122193842 | 0.000358102 | 0.002431865 |
| GATM | 400.070104 | 0.403426323 | 0.141363945 | 0.001557221 | 0.00867924 |
| RPS23 | 39223.83708 | -0.403366542 | 0.079547025 | 1.57E-07 | 2.31E-06 |
| RGS16 | 1442.976188 | -0.403310168 | 0.095333107 | 9.19E-06 | 9.31E-05 |
| GALNT4 | 160.0032418 | 0.403284208 | 0.192115416 | 0.011601983 | 0.045859746 |
| PDLIM2 | 118.5773244 | -0.403197719 | 0.245170879 | 0.029418983 | 0.09630407 |
| RPS18P5 | 6136.876805 | -0.40318553 | 0.135033677 | 0.001046017 | 0.006173782 |
| WDFY2 | 397.1358597 | -0.403163662 | 0.116099524 | 0.000196343 | 0.001437244 |
| ST20 | 110.7438985 | 0.40308687 | 0.224243595 | 0.022089688 | 0.076998215 |
| DCAF11 | 1928.447114 | 0.403034236 | 0.089194209 | 2.41E-06 | 2.79E-05 |
| VKORC1 | 1493.420389 | -0.402988992 | 0.117382361 | 0.000228577 | 0.001646532 |
| ZER1 | 858.2592123 | 0.402595527 | 0.102098557 | 3.10E-05 | 0.000280873 |
| TMEM14C | 1667.583935 | -0.402476314 | 0.09389835 | 7.20E-06 | 7.48E-05 |
| TNRC6B | 634.3400287 | 0.402315955 | 0.110578381 | 0.000104149 | 0.00082519 |
| GSDMD | 261.0193296 | 0.402219543 | 0.156851705 | 0.003675888 | 0.017810674 |
| LOC84773-CYHR1 | 152.6513341 | 0.40220482 | 0.174544341 | 0.00725268 | 0.031375135 |
| SEMA4D | 209.6467399 | 0.402097681 | 0.157228893 | 0.003707167 | 0.017922113 |
| NT5DC2 | 3203.003655 | -0.402073332 | 0.090320784 | 3.39E-06 | 3.78E-05 |
| STRAP | 8224.01557 | -0.402053728 | 0.070204223 | 4.19E-09 | 8.01E-08 |
| MCAT | 589.282149 | 0.402011045 | 0.12242945 | 0.000385437 | 0.002600161 |
| LAMTOR2 | 1837.577362 | -0.401924412 | 0.084484812 | 7.89E-07 | 1.02E-05 |
| SRCAP | 1677.925059 | -0.401912844 | 0.096390651 | 1.19E-05 | 0.000118973 |
| CNOT1 | 9930.367727 | -0.401867828 | 0.061688557 | 3.01E-11 | 7.80E-10 |
| MEST | 1619.603326 | -0.401844467 | 0.082197561 | 4.06E-07 | 5.52E-06 |
| LCT | 109.0518473 | -0.401703848 | 0.220539877 | 0.021433367 | 0.075103525 |
| ZFAND2B | 448.0769363 | 0.401657953 | 0.123435886 | 0.000427325 | 0.002845054 |
| MCM4 | 6775.403541 | -0.4016295 | 0.064048025 | 1.48E-10 | 3.45E-09 |
| EARS2 | 605.0548012 | -0.401588385 | 0.101167947 | 2.81E-05 | 0.000257918 |
| TM7SF2 | 223.4937306 | -0.401471079 | 0.163828218 | 0.00506403 | 0.023335232 |
| MRPL20-AS1 | 172.3769415 | 0.401446429 | 0.169373752 | 0.006157382 | 0.027410636 |
| DET1 | 99.20004666 | 0.401420029 | 0.210088311 | 0.017686408 | 0.064289573 |
| HADHB | 4255.167612 | -0.401286882 | 0.069728828 | 3.56E-09 | 6.87E-08 |
| TOR3A | 1861.348547 | -0.401100273 | 0.081164587 | 3.13E-07 | 4.38E-06 |
| RNF135 | 254.2518794 | 0.400816135 | 0.155228858 | 0.003504155 | 0.017161046 |
| LINC00339 | 115.8116534 | 0.400782039 | 0.193817596 | 0.012674355 | 0.049244291 |
| RPS15AP1 | 299.3546294 | -0.400750444 | 0.198898975 | 0.014483844 | 0.054835315 |
| POLB | 681.0714672 | -0.40065039 | 0.096327604 | 1.26E-05 | 0.000125462 |
| SUSD3 | 432.5110404 | 0.400629848 | 0.140636786 | 0.001607112 | 0.008928571 |
| SPTY2D1 | 1902.479751 | -0.400359566 | 0.103188406 | 4.04E-05 | 0.000354749 |
| DNMBP | 156.4722762 | 0.400105704 | 0.176918971 | 0.008095419 | 0.034267505 |
| INTS6 | 2189.029055 | -0.400054291 | 0.098354979 | 1.87E-05 | 0.000178873 |
| CREBRF | 805.5873391 | -0.399647102 | 0.135181283 | 0.001155349 | 0.006725103 |
| CT45A10 | 368.6031087 | 0.399252496 | 0.132976912 | 0.000997981 | 0.005938847 |
| CCDC32 | 1165.204723 | 0.399154786 | 0.104043823 | 4.89E-05 | 0.000420635 |
| LARS2 | 1236.031389 | -0.399037461 | 0.083669197 | 7.51E-07 | 9.72E-06 |
| TOM1L2 | 318.791297 | 0.398719834 | 0.156804035 | 0.003935356 | 0.018846393 |
| FOXO3 | 2313.202249 | -0.398695537 | 0.074939902 | 4.22E-08 | 6.87E-07 |
| ZNF787 | 799.6811696 | 0.398652718 | 0.133918727 | 0.001085309 | 0.006373108 |
| DPY19L1 | 1614.911824 | -0.398599954 | 0.117502492 | 0.000266139 | 0.001875752 |
| MORF4L2 | 3922.484003 | -0.398578504 | 0.080855099 | 3.34E-07 | 4.63E-06 |

| | | | | | |
|---|---|---|---|---|---|
| CHFR | 692.1326204 | 0.398252796 | 0.112576626 | 0.000156545 | 0.0011766 |
| C8orf76 | 285.8854737 | -0.39825244 | 0.147333076 | 0.002536641 | 0.013105559 |
| TMED7-TICAM2 | 256.2464094 | -0.398205314 | 0.137512035 | 0.001410539 | 0.00796683 |
| DNMT3B | 1483.479687 | -0.397996946 | 0.08426737 | 9.40E-07 | 1.19E-05 |
| LOC124904621 | 9.058168306 | 0.397903192 | 0.642442361 | 0.028092785 | 0.092911144 |
| VPS35 | 4680.386718 | -0.397873987 | 0.076497548 | 8.09E-08 | 1.24E-06 |
| AKAP11 | 882.684873 | 0.397869502 | 0.132285953 | 0.000975307 | 0.005821918 |
| ADGRL3 | 117.0218665 | -0.397651126 | 0.204289354 | 0.016923991 | 0.061973515 |
| MAP3K2-DT | 101.4762606 | -0.39724725 | 0.207695075 | 0.01825938 | 0.065915714 |
| DNAJC5 | 1539.674557 | 0.39723545 | 0.095393973 | 1.23E-05 | 0.000122817 |
| TENT5C | 8149.343996 | -0.397063071 | 0.086898213 | 1.98E-06 | 2.34E-05 |
| TERF2IP | 2831.628071 | -0.397053087 | 0.069653812 | 4.95E-09 | 9.27E-08 |
| NSA2 | 4703.650312 | -0.396329906 | 0.069266325 | 4.36E-09 | 8.29E-08 |
| TMA7 | 2607.965886 | -0.396287893 | 0.077605515 | 1.35E-07 | 2.03E-06 |
| RNF181 | 2081.25456 | 0.396112324 | 0.092443836 | 7.33E-06 | 7.60E-05 |
| GAMT | 795.8974297 | -0.39610822 | 0.116101011 | 0.000253952 | 0.001805291 |
| PACSIN3 | 194.1143387 | -0.396041597 | 0.176096234 | 0.008587691 | 0.035903679 |
| ATP5MC1 | 5156.004341 | -0.395667727 | 0.084208799 | 1.07E-06 | 1.34E-05 |
| LEF1 | 656.9653646 | -0.395496317 | 0.098922031 | 2.57E-05 | 0.000237928 |
| IL23A | 220.5217067 | -0.395448046 | 0.149969599 | 0.003090196 | 0.015466164 |
| CADM1 | 7981.227806 | 0.39537672 | 0.086369233 | 1.88E-06 | 2.24E-05 |
| GLT8D1 | 789.0453469 | -0.395366272 | 0.106046646 | 7.70E-05 | 0.000632508 |
| PRKAR2B | 196.074585 | -0.395332361 | 0.225144869 | 0.024592371 | 0.083765949 |
| DBP | 183.6448925 | 0.395319217 | 0.19684043 | 0.014876947 | 0.055968736 |
| SNCAIP | 7.738670027 | -0.395183428 | 0.637820944 | 0.029637893 | 0.096837621 |
| ARMC7 | 217.9526991 | 0.394947271 | 0.173140919 | 0.007847967 | 0.033424311 |
| SRRM2 | 4735.546322 | -0.394769447 | 0.113230753 | 0.000193175 | 0.001416861 |
| ZFAND1 | 749.1594878 | -0.394629084 | 0.114118794 | 0.000212967 | 0.001543577 |
| PLEKHA2 | 1350.963976 | 0.394599551 | 0.090280934 | 4.99E-06 | 5.37E-05 |
| ANKRD54 | 311.2351122 | 0.39449429 | 0.133917733 | 0.001218722 | 0.007025129 |
| MLST8 | 1213.445622 | -0.394319582 | 0.098237605 | 2.42E-05 | 0.000225156 |
| E2F1 | 856.7593148 | 0.394306229 | 0.106641899 | 8.62E-05 | 0.000695344 |
| PLA2G12A | 736.3118553 | -0.394272606 | 0.102711116 | 4.93E-05 | 0.000423087 |
| HMMR | 1965.449727 | 0.394152861 | 0.116488616 | 0.00027707 | 0.001943303 |
| PAM16 | 170.3153294 | -0.394130376 | 0.186405171 | 0.011991099 | 0.047025679 |
| ARMC2 | 121.7547169 | 0.394119415 | 0.190767326 | 0.013105201 | 0.050713468 |
| PMPCB | 3128.350361 | -0.39404434 | 0.07791029 | 1.75E-07 | 2.55E-06 |
| EXOSC2 | 1026.351908 | 0.393756745 | 0.090838959 | 5.90E-06 | 6.25E-05 |
| PPP6C | 2746.897078 | 0.393612135 | 0.098742419 | 2.63E-05 | 0.000242841 |
| CNEP1R1 | 267.131983 | -0.393587779 | 0.142586592 | 0.00217243 | 0.011529257 |
| MAP1LC3B2 | 516.1973635 | -0.393161331 | 0.107030768 | 9.61E-05 | 0.000767823 |
| CAMTA1 | 635.3478733 | 0.393140259 | 0.102875097 | 5.30E-05 | 0.00045287 |
| RPS15AP17 | 524.2043849 | -0.392798701 | 0.142214919 | 0.002188491 | 0.011593177 |
| NET1 | 3442.641201 | 0.392796386 | 0.085644079 | 1.83E-06 | 2.18E-05 |
| ZHX1 | 932.272721 | -0.392748894 | 0.099478984 | 3.16E-05 | 0.000285602 |
| IQSEC3 | 243.1472922 | -0.392637393 | 0.164299311 | 0.006060049 | 0.027032938 |
| IGHV3-20 | 1343.395231 | -0.392571255 | 0.137523755 | 0.001643729 | 0.009103373 |
| DCUN1D3 | 87.05457253 | -0.392510541 | 0.232900596 | 0.02878062 | 0.094787751 |
| PPP1R13B | 156.5885299 | 0.392170519 | 0.188234987 | 0.012710625 | 0.049340932 |
| RAB40B | 118.0250246 | -0.391943387 | 0.193305081 | 0.01465935 | 0.055378699 |

| | | | | | |
|---|---|---|---|---|---|
| EXOSC7 | 1051.86377 | -0.391929048 | 0.085618032 | 1.95E-06 | 2.30E-05 |
| ARPC1A | 1723.434092 | 0.391515792 | 0.082819194 | 9.36E-07 | 1.19E-05 |
| SYS1-DBNDD2 | 108.1975695 | 0.391503231 | 0.224149945 | 0.025656561 | 0.086573395 |
| EIF2B1 | 1473.373013 | -0.39136415 | 0.081219626 | 6.04E-07 | 7.95E-06 |
| SGO2 | 843.9520757 | 0.391095057 | 0.137797081 | 0.001714773 | 0.009430053 |
| UGGT2 | 655.7846243 | -0.391030115 | 0.114990965 | 0.000265933 | 0.001875063 |
| OPLAH | 182.053103 | -0.390785943 | 0.180101487 | 0.010619607 | 0.04279724 |
| TNIK | 470.5425061 | 0.390692641 | 0.109094505 | 0.000135706 | 0.001037537 |
| CUL9 | 178.46652 | 0.39056109 | 0.165296929 | 0.006568358 | 0.028905781 |
| SSR3 | 17107.86141 | -0.390423626 | 0.073859206 | 5.38E-08 | 8.54E-07 |
| YWHAZ | 18697.22803 | -0.390204361 | 0.104423094 | 7.32E-05 | 0.000604237 |
| PIGX | 769.413016 | 0.390201713 | 0.106140995 | 9.49E-05 | 0.000760146 |
| SERPINH1 | 299.2672296 | -0.390191718 | 0.165625236 | 0.006742109 | 0.029527846 |
| PIP5K1A | 4034.721885 | -0.389939812 | 0.065761832 | 1.34E-09 | 2.76E-08 |
| MIDEAS | 562.0617061 | 0.389932863 | 0.107098182 | 0.000108362 | 0.000852571 |
| DPF3 | 132.5556648 | 0.389904246 | 0.190258203 | 0.013923117 | 0.053259691 |
| SUCO | 1688.713407 | -0.389715631 | 0.100467342 | 4.26E-05 | 0.000372293 |
| EIF4BP3 | 1647.48279 | -0.389625577 | 0.077271749 | 1.94E-07 | 2.80E-06 |
| STUB1 | 1057.281642 | -0.389165925 | 0.1209561 | 0.000515873 | 0.003357151 |
| MFSD6 | 2242.26892 | 0.388930083 | 0.087558054 | 3.64E-06 | 4.03E-05 |
| LAMP1 | 4600.201752 | -0.388873488 | 0.073496848 | 5.17E-08 | 8.25E-07 |
| ZCCHC4 | 285.5420755 | 0.388831185 | 0.143395157 | 0.002529827 | 0.01309099 |
| MIR3654 | 1676.951848 | -0.388708948 | 0.089163315 | 5.52E-06 | 5.90E-05 |
| FAM83D | 941.5999972 | 0.388341083 | 0.113961302 | 0.000261132 | 0.001844216 |
| LOC105378179 | 1175.526383 | 0.388178419 | 1.082872002 | 4.60E-06 | 5.00E-05 |
| DCAF13 | 2849.785674 | -0.388176178 | 0.076052098 | 1.40E-07 | 2.09E-06 |
| RPS12 | 49491.53495 | -0.388156959 | 0.083223721 | 1.36E-06 | 1.66E-05 |
| ETAA1 | 312.7383059 | 0.388156184 | 0.193384784 | 0.015190942 | 0.056865874 |
| NAGPA | 361.620066 | -0.388078749 | 0.168373698 | 0.007795025 | 0.033256085 |
| NCEH1 | 574.4846287 | 0.38801063 | 0.106798815 | 0.000112342 | 0.000880285 |
| LILRP2 | 6.703438052 | 0.387865767 | 0.675084687 | 0.021647509 | 0.075715998 |
| CAPN12 | 161.4068725 | 0.387856301 | 0.181549878 | 0.011596191 | 0.045847315 |
| MIR6805 | 157.0536283 | 0.387749511 | 0.16498685 | 0.006850612 | 0.029938199 |
| NUP35 | 998.4969243 | 0.387401245 | 0.090418466 | 7.58E-06 | 7.84E-05 |
| EIF2S3 | 7243.970457 | -0.387305175 | 0.083560161 | 1.50E-06 | 1.82E-05 |
| THUMPD1 | 1756.196679 | -0.387283475 | 0.095210246 | 1.87E-05 | 0.000178832 |
| MED28 | 2394.427649 | -0.386951318 | 0.074510047 | 8.76E-08 | 1.34E-06 |
| NECAP1 | 1122.843705 | -0.386366108 | 0.086180453 | 3.11E-06 | 3.50E-05 |
| ATP2A2 | 12145.17505 | -0.386365688 | 0.064599707 | 9.72E-10 | 2.04E-08 |
| SMARCA5 | 4433.391451 | -0.386338795 | 0.076075835 | 1.62E-07 | 2.37E-06 |
| ODF2 | 1113.885174 | 0.386072096 | 0.090502307 | 8.21E-06 | 8.43E-05 |
| SH2B2 | 2676.662101 | 0.38607065 | 0.128112448 | 0.00101519 | 0.006020561 |
| C2orf74-DT | 145.5283054 | 0.386041432 | 0.176035217 | 0.010170553 | 0.041343537 |
| PDXDC2P-NPIPB14P | 229.3896847 | -0.386001407 | 0.143269066 | 0.00272557 | 0.013935249 |
| POU6F1 | 160.796943 | 0.38591329 | 0.170462068 | 0.008595213 | 0.035926456 |
| TADA2B | 510.729585 | 0.385903032 | 0.115040245 | 0.000320014 | 0.002204328 |
| IL16 | 2863.542557 | 0.385879536 | 0.079726059 | 5.43E-07 | 7.23E-06 |
| RPL35P2 | 476.3337067 | 0.385851545 | 0.175117338 | 0.009939081 | 0.0405778 |
| TBCA | 4256.263395 | -0.385602482 | 0.074181115 | 8.62E-08 | 1.32E-06 |
| DOK4 | 225.696293 | 0.385447273 | 0.16963511 | 0.0084279 | 0.035355122 |

| | | | | | |
|---|---|---|---|---|---|
| ETV6 | 774.4390654 | 0.385439863 | 0.106792414 | 0.000125197 | 0.000968309 |
| CENPU | 1495.992256 | 0.385417976 | 0.089585981 | 7.03E-06 | 7.32E-05 |
| ZNF547 | 90.83898291 | 0.385404425 | 0.217647809 | 0.025181367 | 0.085335759 |
| SFR1 | 134.3147272 | 0.385152843 | 0.199167492 | 0.01820651 | 0.065793435 |
| HNRNPA1P12 | 812.0979756 | -0.384346819 | 0.095427572 | 2.36E-05 | 0.000220618 |
| LOC729973 | 370.0732862 | -0.38432518 | 0.149921922 | 0.00395229 | 0.018904175 |
| ZNF28 | 574.0306318 | 0.38423142 | 0.114540857 | 0.000319969 | 0.002204328 |
| SPNS1 | 1245.002327 | -0.384196438 | 0.117580726 | 0.000443156 | 0.00293803 |
| ARHGAP11A-SCG5 | 353.4819589 | 0.384140207 | 0.122291749 | 0.000670477 | 0.004209807 |
| PIP5K1B | 182.3629579 | -0.384077469 | 0.16227974 | 0.006746191 | 0.029538254 |
| MRPL15 | 2815.596198 | -0.384072704 | 0.070768349 | 2.48E-08 | 4.15E-07 |
| MAP4K4 | 1607.845295 | -0.383985148 | 0.086487113 | 3.81E-06 | 4.20E-05 |
| C1GALT1 | 1278.62022 | 0.383891133 | 0.134015425 | 0.001629357 | 0.009040558 |
| EDC4 | 1023.79875 | -0.383867239 | 0.111897207 | 0.000247735 | 0.001766899 |
| MTMR9 | 282.6765667 | -0.383674807 | 0.135146234 | 0.001782227 | 0.009742056 |
| GRIN2D | 121.0655425 | -0.383314309 | 0.230113157 | 0.030705052 | 0.099461165 |
| SLC41A1 | 723.6323234 | 0.383230665 | 0.098786699 | 4.35E-05 | 0.000379326 |
| POLL | 238.9846338 | 0.383072226 | 0.151641905 | 0.004342681 | 0.020463184 |
| PABIR3 | 94.73343847 | 0.383024197 | 0.230634466 | 0.030823483 | 0.099751495 |
| RNF213 | 3060.90686 | 0.38300548 | 0.094015302 | 1.92E-05 | 0.000183215 |
| HADH | 3118.329407 | -0.3830022 | 0.070856913 | 2.81E-08 | 4.67E-07 |
| LDLRAP1 | 3373.512967 | 0.382973737 | 0.07611071 | 2.07E-07 | 2.98E-06 |
| PIK3CD | 153.5042868 | 0.382909244 | 0.184484396 | 0.01349601 | 0.051936359 |
| FAM171B | 658.4632953 | 0.382816673 | 0.149124412 | 0.003881291 | 0.018644195 |
| POLR2C | 3075.951425 | -0.382793808 | 0.083957286 | 2.21E-06 | 2.58E-05 |
| PTGES2 | 857.6733421 | 0.382726664 | 0.1118131 | 0.000251378 | 0.00179067 |
| ARL2BP | 1560.869769 | -0.382671849 | 0.087906616 | 5.84E-06 | 6.19E-05 |
| PPCS | 574.6966857 | 0.382594856 | 0.105543119 | 0.000118431 | 0.000920497 |
| IFIT2 | 228.5685506 | 0.382346424 | 0.151739679 | 0.004486401 | 0.021043066 |
| TMTC4 | 623.6954328 | -0.382193469 | 0.105704162 | 0.000123902 | 0.000958721 |
| GAS5 | 10207.49274 | -0.382139765 | 0.07109145 | 3.31E-08 | 5.46E-07 |
| DEF6 | 619.325718 | 0.382001869 | 0.113812145 | 0.000323895 | 0.002230177 |
| KYAT1 | 348.6210004 | 0.381915152 | 0.125064448 | 0.000908961 | 0.005478759 |
| OSBPL10 | 6.958464709 | -0.381783108 | 0.682405906 | 0.018899594 | 0.067704648 |
| PRXL2B | 340.7295532 | 0.381663871 | 0.137085752 | 0.002113723 | 0.01124181 |
| CSPP1 | 360.0303576 | -0.381590502 | 0.125404153 | 0.000938509 | 0.005631378 |
| SEC61G | 2621.584379 | -0.381588042 | 0.089647757 | 8.91E-06 | 9.06E-05 |
| LINC01215 | 101.2336459 | -0.38157618 | 0.212821049 | 0.024945212 | 0.084702216 |
| ZNF93 | 320.4267952 | 0.381434601 | 0.136085755 | 0.001978305 | 0.010642529 |
| DHRS4 | 472.8678492 | -0.381369367 | 0.109568154 | 0.000208336 | 0.001513958 |
| GPR132 | 380.0580611 | 0.381352794 | 0.138922822 | 0.002357497 | 0.012341223 |
| PPP1R9B | 1162.52492 | 0.381289065 | 0.109851414 | 0.000212067 | 0.001538073 |
| THBS1 | 135.7050377 | 0.381054146 | 0.180418972 | 0.0126247 | 0.049095425 |
| QDPR | 800.4412525 | -0.380906851 | 0.104597202 | 0.000114457 | 0.000894022 |
| ENY2 | 1808.732871 | -0.380819887 | 0.080663517 | 1.01E-06 | 1.27E-05 |
| UBL4A | 1314.423369 | 0.380800827 | 0.091891704 | 1.45E-05 | 0.000141455 |
| GOLM1 | 1637.649541 | 0.380741924 | 0.080104456 | 8.55E-07 | 1.09E-05 |
| NOL9 | 415.6653172 | 0.380724914 | 0.15698341 | 0.005786344 | 0.025992768 |
| FANCE | 309.6900005 | 0.380671092 | 0.133505026 | 0.00173063 | 0.00951105 |
| MRPS33 | 1605.062681 | -0.380664391 | 0.088040786 | 6.74E-06 | 7.05E-05 |

| | | | | | |
|---|---|---|---|---|---|
| SORD | 1390.445212 | -0.380602558 | 0.08337792 | 2.15E-06 | 2.52E-05 |
| FIS1 | 1638.132838 | -0.380568919 | 0.093578807 | 2.04E-05 | 0.000192926 |
| C17orf58 | 204.6640173 | 0.380543481 | 0.148424085 | 0.00399589 | 0.019083515 |
| TRIM5 | 250.2904788 | 0.380507196 | 0.138648685 | 0.002384601 | 0.01244924 |
| TRAF3 | 1846.120401 | 0.380251692 | 0.084241806 | 2.71E-06 | 3.10E-05 |
| EIF4B | 26437.32288 | -0.380220448 | 0.068729717 | 1.37E-08 | 2.36E-07 |
| GLE1 | 1818.334873 | 0.38011704 | 0.079110234 | 6.64E-07 | 8.68E-06 |
| POLR2M | 753.9272361 | 0.380009836 | 0.127997762 | 0.001191695 | 0.006901869 |
| ISG15 | 257.0109724 | 0.379941245 | 0.164284724 | 0.007837991 | 0.033398252 |
| UNC13B | 2004.80722 | -0.379858353 | 0.076747592 | 3.24E-07 | 4.51E-06 |
| NUDCD1 | 1313.773478 | -0.3796737 | 0.088842332 | 8.15E-06 | 8.38E-05 |
| ZNF160 | 376.5502197 | 0.379645944 | 0.123005095 | 0.000814909 | 0.004993549 |
| MALT1 | 2393.839447 | 0.379572027 | 0.112901074 | 0.000310164 | 0.002145867 |
| ZC2HC1A | 248.6523988 | 0.379489287 | 0.178685726 | 0.012207562 | 0.047719719 |
| ARHGAP19-SLIT1 | 131.5557418 | 0.37933418 | 0.185648704 | 0.014720736 | 0.055502175 |
| LRRC45 | 112.7590901 | 0.37892366 | 0.198260907 | 0.019743253 | 0.070160868 |
| EIF2AK2 | 734.2850444 | 0.378891652 | 0.119988695 | 0.000644706 | 0.004073102 |
| ANKRD33B | 2683.294863 | -0.378782991 | 0.077006798 | 3.80E-07 | 5.20E-06 |
| SIAH2 | 1695.572033 | -0.378766435 | 0.082128808 | 1.75E-06 | 2.10E-05 |
| NTMT1 | 963.0647585 | 0.378622617 | 0.117150508 | 0.000504954 | 0.003291875 |
| ZCCHC3 | 1703.834793 | -0.378542496 | 0.096680918 | 3.83E-05 | 0.000338896 |
| DENND4B | 896.0241181 | -0.378535433 | 0.103293251 | 0.000105455 | 0.000833863 |
| SMARCAD1 | 2015.608507 | -0.378481546 | 0.093775229 | 2.31E-05 | 0.000216645 |
| COMMD3-BMI1 | 3254.595333 | 0.378453131 | 0.123476296 | 0.00087272 | 0.005282381 |
| TPT1 | 130508.3277 | -0.378352856 | 0.060801736 | 2.27E-10 | 5.14E-09 |
| RALGAPA2 | 323.2308212 | 0.378266018 | 0.126839504 | 0.001155143 | 0.006725103 |
| ACTR5 | 459.2352334 | 0.378137968 | 0.123902038 | 0.000921186 | 0.005542802 |
| ADO | 947.6780752 | 0.378111281 | 0.096969282 | 4.09E-05 | 0.000359587 |
| ASPH | 267.2617626 | -0.378079195 | 0.14395613 | 0.003436531 | 0.016896764 |
| IRF5 | 315.9331189 | 0.378047278 | 0.1299753 | 0.001461409 | 0.008212648 |
| BTD | 333.2356081 | 0.378030857 | 0.130344215 | 0.001505344 | 0.008428078 |
| RPL10A | 31683.6282 | -0.377972587 | 0.065024705 | 2.95E-09 | 5.76E-08 |
| IGLV3-1 | 9.792253757 | -0.377922834 | 0.610201601 | 0.030197703 | 0.098148202 |
| ATN1 | 433.3094968 | 0.377880142 | 0.137380803 | 0.002356284 | 0.012338605 |
| TRIM44 | 948.923944 | -0.377841432 | 0.089100808 | 9.56E-06 | 9.67E-05 |
| C9orf40 | 288.8793337 | 0.377828478 | 0.145451319 | 0.003658456 | 0.017759322 |
| IFI35 | 373.6588924 | 0.377666863 | 0.134234327 | 0.001963399 | 0.010572194 |
| NIPSNAP1 | 4270.909334 | -0.377611159 | 0.076590457 | 3.62E-07 | 5.00E-06 |
| RNASEH2C | 1289.230903 | 0.377534944 | 0.105338334 | 0.000142235 | 0.001080297 |
| TRDC | 4.384366669 | 0.377508604 | 0.752986744 | 0.008954443 | 0.037176718 |
| FASTKD5 | 2204.137658 | -0.377485519 | 0.079376056 | 8.69E-07 | 1.11E-05 |
| SRSF3P1 | 238.5970954 | -0.377450861 | 0.152430561 | 0.00520453 | 0.023874659 |
| EXTL2 | 220.5393633 | -0.377366658 | 0.158591379 | 0.006697488 | 0.029362126 |
| HASPIN | 707.2762912 | 0.377333543 | 0.121678229 | 0.000790282 | 0.004865027 |
| RNF14 | 1548.213603 | 0.377235316 | 0.092786176 | 2.04E-05 | 0.000192793 |
| LIN9 | 381.9784668 | 0.377089556 | 0.128816075 | 0.001375051 | 0.007786711 |
| SCN4A | 87.95025501 | 0.377072404 | 0.2218324 | 0.02994355 | 0.097595768 |
| SNX29 | 489.05375 | -0.376940244 | 0.118125221 | 0.000589836 | 0.003773252 |
| CEP19 | 158.405487 | 0.376871397 | 0.179113903 | 0.013014765 | 0.050386022 |
| FGF2 | 115.3032803 | -0.376636029 | 0.218504699 | 0.028609639 | 0.094278423 |

| | | | | | |
|---|---|---|---|---|---|
| IGFBP6 | 131.64842 | -0.376634791 | 0.189954908 | 0.017374051 | 0.063380333 |
| ALCAM | 244.3618907 | 0.376616895 | 0.17440418 | 0.011396864 | 0.045276325 |
| PLEKHO1 | 2287.051692 | 0.376369692 | 0.108312509 | 0.000213824 | 0.001547345 |
| CTIF | 358.0936899 | -0.376300229 | 0.138094418 | 0.002593223 | 0.013349239 |
| ESRRA | 1321.295141 | 0.376299757 | 0.11818475 | 0.000600395 | 0.003832305 |
| SNUPN | 394.1488198 | 0.376290468 | 0.121397167 | 0.000795768 | 0.004888372 |
| TFEB | 367.7883508 | 0.37623866 | 0.128546598 | 0.001391493 | 0.007872093 |
| NUP210 | 11931.81376 | -0.37613886 | 0.084508863 | 3.77E-06 | 4.16E-05 |
| C16orf72 | 2362.676358 | -0.376129285 | 0.102500746 | 0.00010354 | 0.000821349 |
| KMT2A | 1149.640535 | 0.375942568 | 0.114168418 | 0.000409456 | 0.002744018 |
| TRAPPC5 | 834.8093022 | 0.375843506 | 0.134963022 | 0.002163734 | 0.011486626 |
| ND4L | 9309.83523 | -0.375757596 | 0.085256507 | 4.61E-06 | 5.01E-05 |
| VPS26B | 3526.365632 | 0.375717145 | 0.073474609 | 1.38E-07 | 2.06E-06 |
| KRTCAP2 | 3872.414115 | -0.37570441 | 0.091838275 | 1.87E-05 | 0.000178832 |
| FKBP11 | 9846.578672 | -0.375551563 | 0.077297394 | 5.21E-07 | 6.96E-06 |
| NAV1 | 490.9557815 | -0.375424116 | 0.111093673 | 0.000304346 | 0.00211152 |
| AMDHD2 | 345.6523773 | -0.375398824 | 0.144444245 | 0.00375834 | 0.018114843 |
| EIF2D | 1032.754935 | -0.375256311 | 0.08822271 | 9.22E-06 | 9.34E-05 |
| CYTH1 | 2315.66943 | 0.375102695 | 0.086408227 | 6.14E-06 | 6.48E-05 |
| SEC61B | 5431.508153 | -0.375095762 | 0.074781149 | 2.39E-07 | 3.41E-06 |
| ELFN1-AS1 | 134.810523 | -0.374983582 | 0.184234524 | 0.015545933 | 0.057906843 |
| CGREF1 | 5.051817547 | -0.374973097 | 0.719888344 | 0.011686456 | 0.046069451 |
| TRANK1 | 426.499408 | 0.374947933 | 0.113224005 | 0.000387556 | 0.002612421 |
| ATP1B3 | 6359.662196 | -0.374714004 | 0.07614251 | 3.82E-07 | 5.22E-06 |
| DPY19L4 | 173.4488858 | 0.374699625 | 0.178491647 | 0.01321034 | 0.051040485 |
| KIF7 | 129.2951612 | -0.374567116 | 0.193288793 | 0.01917701 | 0.068528329 |
| COCH | 795.8272892 | 0.374527322 | 0.097984254 | 5.68E-05 | 0.00048198 |
| ZBTB14 | 355.0038641 | 0.374437911 | 0.128736097 | 0.001475506 | 0.008287815 |
| BRWD3 | 252.7002128 | 0.374436752 | 0.194512991 | 0.019332601 | 0.068998897 |
| TIAM1 | 457.3585614 | 0.374388693 | 0.119334347 | 0.000699724 | 0.004372833 |
| RAB36 | 1781.99091 | 0.37433689 | 0.099753733 | 7.44E-05 | 0.000612496 |
| MTG1 | 279.635953 | 0.374301403 | 0.14637767 | 0.004203897 | 0.019939435 |
| NIPSNAP2 | 1504.457624 | -0.374291865 | 0.099782514 | 7.58E-05 | 0.000622914 |
| EXO5 | 155.9259054 | 0.374128825 | 0.165238285 | 0.009030158 | 0.037401398 |
| LOC84773 | 98.27700744 | -0.374068259 | 0.216467101 | 0.02908919 | 0.095585864 |
| ZHX1-C8orf76 | 229.6813524 | -0.373997414 | 0.150478045 | 0.005165603 | 0.023733813 |
| HNRNPA1P4 | 1139.189605 | -0.373899248 | 0.085312244 | 5.18E-06 | 5.56E-05 |
| DYNC1LI2 | 920.6187054 | -0.373776374 | 0.089810164 | 1.37E-05 | 0.000134748 |
| IFNAR1 | 3484.139187 | 0.373501576 | 0.095721153 | 4.08E-05 | 0.00035848 |
| SPA17 | 143.5622333 | 0.373478825 | 0.184236824 | 0.0156849 | 0.058361714 |
| EIF4A3 | 4272.854358 | -0.372848396 | 0.073189242 | 1.57E-07 | 2.31E-06 |
| CAPRIN2 | 132.9140067 | 0.372754024 | 0.181567361 | 0.014902724 | 0.056017032 |
| RPS18 | 91975.54393 | -0.372737588 | 0.065675699 | 5.83E-09 | 1.08E-07 |
| TMEM263 | 1176.096306 | -0.372728515 | 0.161885773 | 0.008288196 | 0.034895863 |
| FLOT1 | 1551.715484 | 0.372568279 | 0.080408064 | 1.58E-06 | 1.90E-05 |
| LMBR1L | 530.5304537 | 0.372516581 | 0.124160638 | 0.001122498 | 0.006566992 |
| TFRC | 23343.19087 | -0.37226881 | 0.073186929 | 1.65E-07 | 2.41E-06 |
| RASGRP2 | 437.1332126 | 0.372152963 | 0.129351708 | 0.001653572 | 0.009142693 |
| USP53 | 445.2010652 | -0.371499077 | 0.129993617 | 0.001760007 | 0.009638847 |
| TOR2A | 420.9607453 | 0.371462893 | 0.149973634 | 0.005277913 | 0.024128201 |

| | | | | | |
|---|---|---|---|---|---|
| PTGR2 | 188.630297 | -0.371369684 | 0.166431761 | 0.009926614 | 0.04054999 |
| FEZ2 | 484.9866211 | 0.371227211 | 0.119093632 | 0.00076287 | 0.004721451 |
| TRMT1 | 777.017879 | -0.371216028 | 0.10495097 | 0.000176081 | 0.001305306 |
| TUBB6 | 693.8772031 | 0.371156198 | 0.105482658 | 0.000185275 | 0.001366439 |
| ABL1 | 939.8955041 | 0.371073484 | 0.093399636 | 3.09E-05 | 0.000280212 |
| ATF6 | 4651.083971 | -0.371023825 | 0.068520558 | 2.77E-08 | 4.62E-07 |
| MRPL41 | 668.1353177 | 0.371011159 | 0.140774026 | 0.003413158 | 0.016800926 |
| PIH1D1 | 2165.539854 | 0.370928728 | 0.086290619 | 7.58E-06 | 7.84E-05 |
| DDX18 | 4438.83279 | -0.370876196 | 0.077479776 | 7.59E-07 | 9.80E-06 |
| ZNF318 | 277.9407869 | 0.370865057 | 0.132762671 | 0.002134498 | 0.011345333 |
| PDIA6 | 53531.34664 | -0.370855418 | 0.06411995 | 3.82E-09 | 7.33E-08 |
| RNMT | 3359.644794 | -0.370822619 | 0.094489386 | 3.81E-05 | 0.000337211 |
| TBCCD1 | 532.0348292 | 0.370803708 | 0.108805329 | 0.000279595 | 0.001957841 |
| UROD | 1307.409369 | -0.370681641 | 0.086140862 | 7.25E-06 | 7.53E-05 |
| SLC25A20 | 725.2348427 | -0.370627584 | 0.094222995 | 3.68E-05 | 0.000327007 |
| COPS2 | 2583.066751 | -0.370606061 | 0.109189617 | 0.000295544 | 0.002057045 |
| ZEB1 | 473.7292231 | -0.370484596 | 0.126362687 | 0.001403222 | 0.007933269 |
| NUDT22 | 478.7845396 | 0.370433501 | 0.149994775 | 0.005413865 | 0.024652092 |
| VSIR | 503.7867518 | 0.370055805 | 0.115270294 | 0.000561804 | 0.003611269 |
| DDTL | 266.1168607 | 0.370053768 | 0.166389565 | 0.010170051 | 0.041343537 |
| HIGD1A | 4407.860366 | -0.369861749 | 0.112938648 | 0.000453919 | 0.003004781 |
| SYTL1 | 257.9823976 | -0.369756802 | 0.139189548 | 0.003231632 | 0.016081742 |
| BRD7 | 794.2020893 | -0.369612535 | 0.09347674 | 3.38E-05 | 0.000303693 |
| HAUS8 | 486.7506738 | 0.369461642 | 0.110943835 | 0.00036821 | 0.002495615 |
| DALRD3 | 410.5233267 | -0.369420145 | 0.12803889 | 0.001653035 | 0.009142641 |
| C9orf78 | 1927.297786 | 0.369254645 | 0.078053529 | 9.96E-07 | 1.25E-05 |
| DNAJA2 | 1660.434378 | -0.369236691 | 0.092326008 | 2.80E-05 | 0.000256454 |
| NDRG1 | 2579.169846 | 0.369105836 | 0.102252822 | 0.000131812 | 0.001013134 |
| DDA1 | 915.458765 | 0.369097876 | 0.090945737 | 2.18E-05 | 0.000205317 |
| AGAP2 | 301.6927287 | 0.368879625 | 0.129495864 | 0.001821952 | 0.009921416 |
| BRI3 | 638.792986 | -0.368838607 | 0.124404975 | 0.001286493 | 0.007344107 |
| EIF5B | 2321.597539 | -0.368566192 | 0.094148828 | 3.98E-05 | 0.000350849 |
| MTBP | 222.522255 | -0.3685632 | 0.156598123 | 0.007482754 | 0.032153562 |
| RFX3 | 130.6139733 | 0.36851091 | 0.192223818 | 0.020340693 | 0.07185634 |
| PSMD7 | 4450.732776 | -0.368315012 | 0.067890244 | 2.65E-08 | 4.42E-07 |
| APOBEC3B | 2568.430421 | 0.368122802 | 0.078923023 | 1.38E-06 | 1.69E-05 |
| MED25 | 749.3255048 | 0.368050672 | 0.13265634 | 0.002308047 | 0.012126351 |
| PGRMC1 | 2209.663014 | -0.367935967 | 0.094865061 | 4.68E-05 | 0.000404943 |
| HSP90AA1 | 38398.1978 | -0.367916983 | 0.067983915 | 3.14E-08 | 5.19E-07 |
| NACA2 | 228.1556829 | -0.367515388 | 0.214517953 | 0.030844401 | 0.09977564 |
| KICS2 | 397.0500944 | 0.367496426 | 0.131984774 | 0.002232406 | 0.011786149 |
| TMBIM6 | 29949.03538 | 0.36743191 | 0.069833855 | 6.34E-08 | 9.94E-07 |
| ZBTB10 | 376.8234787 | -0.367331582 | 0.165214686 | 0.010276873 | 0.041736542 |
| RPL13 | 54864.63958 | -0.367159273 | 0.105164098 | 0.000274679 | 0.001927319 |
| PCID2 | 1695.747163 | -0.367123344 | 0.079512202 | 1.77E-06 | 2.12E-05 |
| COG5 | 1056.235743 | -0.367110395 | 0.086009697 | 8.84E-06 | 9.00E-05 |
| MAT2B | 2729.250756 | 0.367064278 | 0.111813764 | 0.000458164 | 0.003028254 |
| RAD50 | 472.0071172 | -0.366945709 | 0.116493736 | 0.000702607 | 0.004387684 |
| TCAIM | 405.2094873 | -0.366906506 | 0.124024072 | 0.001310997 | 0.007463028 |
| NUDT21 | 4081.085381 | -0.366888703 | 0.110015815 | 0.000370866 | 0.002510232 |

| | | | | | |
|---|---|---|---|---|---|
| DLGAP5 | 1213.780617 | 0.366829943 | 0.09711586 | 6.95E-05 | 0.000576898 |
| CCDC137 | 402.6962495 | 0.366740894 | 0.125150966 | 0.001426873 | 0.008048585 |
| KLF3 | 1285.141912 | -0.366697516 | 0.118327712 | 0.000835104 | 0.005090286 |
| OAT | 6677.120456 | -0.366575121 | 0.089612867 | 1.93E-05 | 0.00018367 |
| SYNE2 | 163.0242258 | 0.366561523 | 0.177297818 | 0.014783701 | 0.055678414 |
| HLA-K | 163.7042606 | 0.366435365 | 0.164028393 | 0.010087799 | 0.041076248 |
| KLHL30 | 7.870397318 | -0.366408895 | 0.619073405 | 0.024965357 | 0.084736395 |
| PLD3 | 3083.852374 | -0.366380369 | 0.101650571 | 0.000138117 | 0.001054576 |
| ATG4D | 519.387687 | 0.366191595 | 0.128199833 | 0.001801351 | 0.009831071 |
| LY75-CD302 | 533.6799475 | 0.366117586 | 0.103967695 | 0.000185523 | 0.001367691 |
| PSMB8 | 3636.426104 | 0.366112051 | 0.074308246 | 3.79E-07 | 5.19E-06 |
| OGFR | 536.4784761 | 0.366063169 | 0.106439405 | 0.00025445 | 0.001807343 |
| ARHGEF39 | 101.4815555 | 0.365997014 | 0.207893352 | 0.028234 | 0.093253515 |
| AMIGO3 | 142.3812008 | -0.365936534 | 0.187912582 | 0.019624847 | 0.069836418 |
| LOC105371675 | 237.5785721 | 0.365896031 | 0.159849583 | 0.008731054 | 0.036402273 |
| GPBP1 | 3402.82281 | -0.365894154 | 0.085191669 | 7.87E-06 | 8.12E-05 |
| TPCN1 | 658.5811112 | -0.365884416 | 0.110600843 | 0.000412402 | 0.002758429 |
| TCEANC2 | 243.8855925 | 0.36579741 | 0.140138304 | 0.0037005 | 0.017897299 |
| ND4 | 73425.73895 | -0.36570347 | 0.091490817 | 2.88E-05 | 0.000263625 |
| PPT1 | 9800.896171 | -0.365699887 | 0.065298904 | 1.00E-08 | 1.79E-07 |
| GCOM1 | 713.1309119 | 0.365629417 | 0.131615379 | 0.002310519 | 0.012135658 |
| RPSAP17 | 390.6850447 | -0.365526719 | 0.127343447 | 0.001759053 | 0.009638847 |
| RHOG | 705.2912282 | 0.365462345 | 0.107685946 | 0.000301523 | 0.002093284 |
| GMPR2 | 1046.656723 | -0.365386295 | 0.085495678 | 8.68E-06 | 8.86E-05 |
| TMEM143 | 258.7208233 | -0.365361071 | 0.135718 | 0.002997977 | 0.015065572 |
| CBX7 | 144.6703419 | 0.365300377 | 0.182346466 | 0.017253245 | 0.062992715 |
| RN7SL2 | 1160.639719 | 0.365293612 | 0.903678783 | 1.58E-06 | 1.90E-05 |
| AZIN1 | 6430.424314 | -0.364973963 | 0.083611023 | 5.76E-06 | 6.13E-05 |
| CAMKMT | 305.6545907 | -0.36494831 | 0.139822441 | 0.003765291 | 0.01814233 |
| TGFB1 | 3849.368984 | -0.364697884 | 0.094013794 | 4.85E-05 | 0.000417427 |
| RPGR | 144.4117365 | 0.364620912 | 0.17329522 | 0.01379346 | 0.052833743 |
| HNRNPA1L3 | 2821.795122 | -0.364542047 | 0.080045624 | 2.38E-06 | 2.76E-05 |
| EDF1 | 3400.315166 | 0.364500178 | 0.077403656 | 1.13E-06 | 1.40E-05 |
| CLN8 | 263.2924893 | -0.364498202 | 0.136933383 | 0.003259313 | 0.016188289 |
| RGS10 | 752.1457132 | -0.364350675 | 0.113651721 | 0.000592207 | 0.003787017 |
| SREK1 | 975.8373627 | -0.364314785 | 0.117725145 | 0.000851789 | 0.00517559 |
| TSR3 | 696.2579648 | -0.364250854 | 0.152851788 | 0.007054576 | 0.030655983 |
| UTP23 | 1005.668528 | -0.364237272 | 0.121809542 | 0.001196038 | 0.006924706 |
| PRRC2C | 2488.984012 | -0.364219564 | 0.127700427 | 0.001856177 | 0.010082627 |
| LOC284685 | 174.533985 | 0.363965852 | 0.163050991 | 0.010176683 | 0.041358745 |
| TBCK | 221.4373354 | -0.36376485 | 0.143900968 | 0.004788942 | 0.022275076 |
| DCAF16 | 263.8967787 | 0.36373959 | 0.140786935 | 0.004071142 | 0.0193841 |
| RASSF8 | 194.1014835 | 0.363535537 | 0.17645303 | 0.015212168 | 0.056933027 |
| STIM1 | 2488.925636 | 0.363526305 | 0.087731355 | 1.53E-05 | 0.000148807 |
| PLEKHA3 | 677.2412113 | 0.363254632 | 0.112247491 | 0.00052474 | 0.003403344 |
| DGUOK-AS1 | 313.816533 | -0.363197386 | 0.138462109 | 0.003684476 | 0.017842303 |
| HNRNPA1P10 | 3671.264355 | -0.363007623 | 0.073536381 | 3.68E-07 | 5.06E-06 |
| UNC5C | 7.20799091 | 0.362804416 | 0.609378274 | 0.025345804 | 0.085792276 |
| MAPKAP1 | 2380.895094 | 0.362786166 | 0.084521094 | 8.00E-06 | 8.24E-05 |
| RPL10 | 67404.68504 | -0.362581739 | 0.105980284 | 0.000203788 | 0.001485424 |

| | | | | | |
|---|---|---|---|---|---|
| KCTD5 | 1203.553265 | -0.362554214 | 0.093036838 | 4.43E-05 | 0.00038592 |
| MAF1 | 2211.987954 | 0.362510783 | 0.099904327 | 0.000126807 | 0.000978573 |
| MSL3P1 | 214.6743421 | 0.362473712 | 0.145472979 | 0.005271928 | 0.02411994 |
| CDKL5 | 201.9705107 | 0.362462114 | 0.176087225 | 0.015360595 | 0.057376926 |
| SHCBP1 | 2031.760159 | -0.362376421 | 0.074043737 | 4.59E-07 | 6.17E-06 |
| RCHY1 | 578.3691601 | -0.362297467 | 0.132077511 | 0.002596032 | 0.013359729 |
| RPS2P7 | 680.82986 | -0.36219734 | 0.166836432 | 0.012054332 | 0.047184644 |
| HNRNPA1P38 | 163.2298412 | -0.36213342 | 0.166935055 | 0.012132582 | 0.047469479 |
| ADK | 1622.250872 | -0.362108316 | 0.079071867 | 2.16E-06 | 2.53E-05 |
| TMSB4XP8 | 6268.48525 | 0.361939589 | 0.095829103 | 6.99E-05 | 0.000579717 |
| FNBP4 | 862.9870144 | -0.361912371 | 0.104048545 | 0.000224534 | 0.001619429 |
| USF2 | 2452.378259 | 0.361865155 | 0.086656903 | 1.34E-05 | 0.000131966 |
| SPRY1 | 266.861539 | 0.361636806 | 0.142798167 | 0.004735716 | 0.022068997 |
| SPATS2L | 693.4950689 | 0.361470681 | 0.100905711 | 0.000151792 | 0.00114535 |
| RRP1B | 3519.625957 | -0.361432976 | 0.072804481 | 3.20E-07 | 4.46E-06 |
| ZBTB34 | 338.4241991 | 0.36137398 | 0.144968864 | 0.00525688 | 0.024073722 |
| P2RX5-TAX1BP3 | 738.5889196 | -0.361351119 | 0.099534457 | 0.000128381 | 0.000989834 |
| GGH | 3371.535791 | -0.361293796 | 0.093136894 | 4.87E-05 | 0.000418598 |
| QARS1 | 7879.140383 | -0.361276172 | 0.069523562 | 9.72E-08 | 1.47E-06 |
| CRLF3 | 802.8246543 | 0.36118378 | 0.113257567 | 0.000622758 | 0.003954639 |
| RHOT2 | 742.185457 | -0.361174786 | 0.10554327 | 0.000279098 | 0.001955154 |
| ATP8B2 | 6414.501973 | -0.361092915 | 0.080334328 | 3.18E-06 | 3.57E-05 |
| SSB | 4699.728649 | -0.360999535 | 0.085105436 | 1.02E-05 | 0.000102719 |
| RPL39L | 284.0892403 | 0.360944064 | 0.146926496 | 0.005857272 | 0.026256893 |
| CDR2 | 931.8369673 | -0.360859383 | 0.087651921 | 1.76E-05 | 0.000169476 |
| ATP11A | 1299.844384 | 0.360856774 | 0.094442474 | 5.95E-05 | 0.000502627 |
| ASAP1 | 845.3803658 | 0.360768538 | 0.112232128 | 0.000570628 | 0.003665269 |
| RNF4 | 1678.469086 | 0.3607279 | 0.075989446 | 9.53E-07 | 1.21E-05 |
| METTL22 | 375.1635823 | -0.360588057 | 0.144667965 | 0.005369834 | 0.02447734 |
| EEF1A1P9 | 3684.997982 | -0.360537525 | 0.073064984 | 3.77E-07 | 5.17E-06 |
| HOMER3 | 6717.309568 | 0.360366862 | 0.086235197 | 1.34E-05 | 0.000131925 |
| FAHD2A | 1267.35138 | -0.360311055 | 0.082819766 | 6.27E-06 | 6.60E-05 |
| ALDOB | 5.302881371 | 0.360136533 | 0.697312208 | 0.008503557 | 0.035618594 |
| PIM1 | 929.4069624 | 0.360037602 | 0.094254375 | 6.05E-05 | 0.000510928 |
| LYSMD1 | 188.1605758 | -0.360027386 | 0.152435062 | 0.007528972 | 0.032296065 |
| SMAD7 | 305.0872086 | 0.359898875 | 0.130553585 | 0.002490049 | 0.012925712 |
| CARD19 | 310.3966191 | 0.359837787 | 0.139177108 | 0.004126573 | 0.019621049 |
| FKBP14 | 278.5233226 | -0.359761059 | 0.156945459 | 0.008951329 | 0.037176718 |
| NBAS | 4293.07837 | -0.359717226 | 0.06608433 | 2.46E-08 | 4.13E-07 |
| GALNT3 | 1420.367536 | -0.359622643 | 0.122508562 | 0.001458131 | 0.008200861 |
| CDKN3 | 1237.684012 | 0.359606714 | 0.090668154 | 3.29E-05 | 0.000296957 |
| HEMK1 | 291.7667887 | -0.359503475 | 0.132850861 | 0.002948001 | 0.014848902 |
| GRAMD1A | 549.5988909 | -0.359206508 | 0.121243985 | 0.001343326 | 0.007622012 |
| HK1 | 740.4028471 | 0.358990248 | 0.09861074 | 0.000122957 | 0.000952681 |
| PHKA1 | 693.0347696 | -0.358986081 | 0.095564426 | 7.91E-05 | 0.000646363 |
| CLN5 | 630.0825396 | -0.35897183 | 0.10354477 | 0.000237531 | 0.001702536 |
| GNG7 | 2544.639959 | -0.358712233 | 0.081309104 | 4.80E-06 | 5.19E-05 |
| UPF3B | 270.5926172 | 0.358686797 | 0.147363924 | 0.00621847 | 0.0276116 |
| PPP2R5B | 127.2768152 | 0.358663447 | 0.194977169 | 0.025114425 | 0.085175576 |
| CAMSAP3 | 151.1535364 | 0.35852374 | 0.18452062 | 0.020220718 | 0.071549279 |

| | | | | | |
|---|---|---|---|---|---|
| DPCD | 465.5696483 | -0.358431404 | 0.12256629 | 0.001528786 | 0.008537243 |
| EIF4BP6 | 4042.9161 | -0.358137526 | 0.086613479 | 1.64E-05 | 0.000159129 |
| VPS13B | 236.6426164 | -0.358131949 | 0.14410401 | 0.005478029 | 0.02485277 |
| ALPK1 | 110.7493073 | 0.358034192 | 0.198068779 | 0.026604302 | 0.089163285 |
| TOB2 | 623.1798273 | -0.357816871 | 0.098841802 | 0.000135086 | 0.001033255 |
| HTD2 | 523.5337384 | -0.35781081 | 0.105167666 | 0.000301596 | 0.002093284 |
| LCP1 | 31319.03917 | 0.357788974 | 0.060279981 | 1.38E-09 | 2.84E-08 |
| IFT140 | 127.6502856 | -0.357683577 | 0.17987478 | 0.018560076 | 0.066806244 |
| RCC2 | 9436.952024 | 0.357672049 | 0.07461877 | 7.65E-07 | 9.87E-06 |
| NOLC1 | 6753.221282 | -0.35764913 | 0.070923012 | 2.24E-07 | 3.21E-06 |
| FBRS | 1050.024855 | -0.357607611 | 0.116403643 | 0.000947355 | 0.005678556 |
| LY75 | 640.7884341 | 0.357545059 | 0.110050058 | 0.000514052 | 0.003346556 |
| LOC101927297 | 256.6622374 | -0.35752656 | 0.169467976 | 0.014053415 | 0.053556995 |
| CAPN7 | 1048.824459 | -0.357297138 | 0.091554792 | 4.36E-05 | 0.000380232 |
| ZNF581 | 438.5973207 | -0.357285654 | 0.151237816 | 0.007688003 | 0.032888623 |
| SLF2 | 620.1197556 | 0.356829829 | 0.111626735 | 0.000618162 | 0.003932661 |
| HSPD1 | 41155.01915 | -0.356578341 | 0.064063448 | 1.29E-08 | 2.25E-07 |
| GMIP | 299.4488442 | 0.356331503 | 0.143558021 | 0.00556495 | 0.025188299 |
| ZDHHC13 | 569.6142213 | 0.356310285 | 0.113771176 | 0.000770382 | 0.004757741 |
| RPSAP3 | 547.7858995 | -0.356302193 | 0.118942191 | 0.001228597 | 0.007065964 |
| AKTIP | 199.9532735 | 0.356209764 | 0.149834969 | 0.007305494 | 0.031541597 |
| RSPO2 | 5.00839269 | -0.35612044 | 0.661483995 | 0.012800188 | 0.049632972 |
| SLC25A26 | 686.9689812 | -0.356111595 | 0.107315315 | 0.000412975 | 0.002761194 |
| MPZL1 | 4772.357239 | -0.356080765 | 0.073031661 | 5.12E-07 | 6.84E-06 |
| RIOX1 | 328.8212345 | 0.356060752 | 0.122002481 | 0.001548486 | 0.008636113 |
| RPS17P16 | 2130.783229 | -0.35596155 | 0.090146368 | 3.65E-05 | 0.000325035 |
| PSMD6 | 2558.375926 | -0.355929341 | 0.077015207 | 1.80E-06 | 2.15E-05 |
| ARG2 | 121.6127758 | 0.355901164 | 0.188941935 | 0.023127158 | 0.079763593 |
| ZNF764 | 145.8072789 | -0.355890872 | 0.181596104 | 0.019889116 | 0.070606768 |
| SEPTIN6 | 4389.746934 | 0.355777573 | 0.072867053 | 4.96E-07 | 6.63E-06 |
| RNF31 | 440.5165491 | 0.355460747 | 0.131593634 | 0.003000927 | 0.015076024 |
| CARS2 | 1124.80499 | -0.355327446 | 0.097198587 | 0.000119082 | 0.00092514 |
| RPL21 | 26983.87862 | -0.355201146 | 0.074415669 | 8.61E-07 | 1.10E-05 |
| FUS | 6924.639877 | -0.355180616 | 0.067485334 | 6.78E-08 | 1.06E-06 |
| AMACR | 188.3338871 | -0.355163552 | 0.159954918 | 0.01100605 | 0.044006413 |
| LOC124903625 | 8.05136022 | -0.355153583 | 0.643730809 | 0.014545164 | 0.054983323 |
| ABHD14B | 1592.467836 | -0.355139329 | 0.118643917 | 0.001241062 | 0.007124201 |
| PDE12 | 2158.604793 | -0.354963467 | 0.118039566 | 0.001196951 | 0.006927672 |
| CYB5A | 918.0091907 | 0.354934337 | 0.090934221 | 4.39E-05 | 0.000382592 |
| ILKAP | 1198.147151 | 0.354861251 | 0.085219916 | 1.45E-05 | 0.000141461 |
| ZBTB43 | 915.5898059 | 0.354845388 | 0.095573723 | 9.39E-05 | 0.000752536 |
| ZNF737 | 170.1377413 | 0.354662208 | 0.19653036 | 0.027059984 | 0.090253803 |
| EIF3L | 14660.4389 | -0.354653813 | 0.074856162 | 1.01E-06 | 1.27E-05 |
| ZNF791 | 696.6122075 | 0.354580562 | 0.114143467 | 0.000841774 | 0.005123725 |
| EIF4BP7 | 2953.302618 | -0.354342285 | 0.078241259 | 2.80E-06 | 3.19E-05 |
| OXCT1 | 5949.189931 | -0.354300974 | 0.072462735 | 4.87E-07 | 6.52E-06 |
| PYCR1 | 4174.472333 | -0.354268304 | 0.09314509 | 6.74E-05 | 0.000561929 |
| ZBTB8A | 163.1541598 | 0.354227697 | 0.185863677 | 0.022130536 | 0.077109557 |
| SLC35E3 | 618.1348026 | 0.354154817 | 0.109170965 | 0.000532334 | 0.003443532 |
| ZNF266 | 606.736942 | 0.354153683 | 0.100812914 | 0.000203018 | 0.001480906 |

| | | | | | |
|---|---|---|---|---|---|
| GTPBP3 | 947.8654115 | -0.354119474 | 0.127752783 | 0.002480335 | 0.012883013 |
| PUDP | 520.2601326 | -0.354044197 | 0.107221297 | 0.000440839 | 0.002926029 |
| RAB2A | 1903.440378 | -0.354004198 | 0.110406744 | 0.00061081 | 0.003890179 |
| ATP6V0A1 | 2428.462943 | 0.353982938 | 0.073025887 | 5.93E-07 | 7.83E-06 |
| RPL21P28 | 1788.900763 | -0.35380068 | 0.079011474 | 3.60E-06 | 4.00E-05 |
| LEMD3 | 1716.593009 | 0.35363166 | 0.109215096 | 0.000542326 | 0.003495144 |
| ALKBH6 | 273.160872 | 0.353550362 | 0.141241324 | 0.005298075 | 0.024201205 |
| POLR3K | 847.2581314 | -0.353291746 | 0.098120891 | 0.000147463 | 0.00111707 |
| NUB1 | 878.0353427 | 0.353074696 | 0.09269287 | 6.46E-05 | 0.000540829 |
| RPP14 | 539.7054273 | -0.353071331 | 0.104422311 | 0.000330701 | 0.002272521 |
| RPS2P20 | 710.8974916 | -0.35306396 | 0.150254093 | 0.008122619 | 0.034357449 |
| RAD23A | 2918.052046 | 0.353055593 | 0.078127015 | 2.92E-06 | 3.30E-05 |
| UBALD2 | 2018.711886 | 0.352978353 | 0.090550764 | 4.52E-05 | 0.000392476 |
| CMC2 | 1957.653338 | -0.352799064 | 0.080133802 | 5.12E-06 | 5.50E-05 |
| SH3BGRL3 | 1400.570897 | 0.352183642 | 0.094518099 | 9.04E-05 | 0.000726908 |
| TESC | 1006.207345 | -0.352051693 | 0.118064707 | 0.001312215 | 0.007467509 |
| SRM | 4056.437611 | -0.352016576 | 0.103935057 | 0.000327928 | 0.002256019 |
| CETN2 | 920.2562193 | -0.351964067 | 0.086692684 | 2.33E-05 | 0.000217838 |
| PINX1 | 872.7364676 | -0.351907745 | 0.091641776 | 5.80E-05 | 0.00049136 |
| HACD2 | 1878.762259 | -0.351554341 | 0.094340108 | 9.12E-05 | 0.000732587 |
| RPSAP14 | 211.5591731 | -0.351543317 | 0.148089905 | 0.007577244 | 0.032470957 |
| RAB5C | 2458.578966 | 0.351342962 | 0.079337751 | 4.53E-06 | 4.93E-05 |
| DGKD | 2276.022343 | 0.351292227 | 0.091180771 | 5.47E-05 | 0.000466635 |
| UBXN8 | 563.9589869 | -0.351231808 | 0.115102049 | 0.001037023 | 0.006128303 |
| SGCB | 331.7102656 | 0.350960553 | 0.150707895 | 0.008425125 | 0.035352044 |
| MYCBP2 | 2045.525323 | -0.350716185 | 0.086548113 | 2.40E-05 | 0.000223451 |
| SPATA20 | 901.4954466 | 0.35068102 | 0.123771375 | 0.002070254 | 0.011039782 |
| MAP3K14 | 1161.994411 | 0.350679332 | 0.110260333 | 0.000672446 | 0.00421928 |
| NKIRAS2 | 1073.425318 | 0.350573429 | 0.096251137 | 0.000125727 | 0.000971972 |
| MBD1 | 1761.60588 | 0.350463828 | 0.090120193 | 4.71E-05 | 0.000407576 |
| NDUFB10 | 1771.221592 | -0.350422544 | 0.078075573 | 3.47E-06 | 3.86E-05 |
| TRMT2B | 597.8356834 | 0.350309842 | 0.101019341 | 0.000243805 | 0.001742455 |
| REXO5 | 145.8950393 | -0.350247422 | 0.197267124 | 0.029855615 | 0.097438691 |
| HSPE1 | 5405.900538 | -0.350128849 | 0.069115595 | 1.98E-07 | 2.85E-06 |
| NAPA-AS1 | 137.8394103 | -0.349849003 | 0.19599002 | 0.029110138 | 0.095634128 |
| TMF1 | 976.7346189 | -0.349790616 | 0.103759164 | 0.000347376 | 0.00236645 |
| TUBB4B | 14798.6297 | 0.349781334 | 0.074909571 | 1.42E-06 | 1.73E-05 |
| CENPW | 624.0113499 | 0.34978128 | 0.107316326 | 0.00051672 | 0.0033614 |
| PPM1H | 544.8246803 | -0.349736941 | 0.111710789 | 0.000802021 | 0.004923289 |
| JTB | 9207.27148 | -0.349729346 | 0.109620195 | 0.000658296 | 0.004148219 |
| YIPF5 | 2186.882478 | -0.349652497 | 0.089227812 | 4.14E-05 | 0.000362665 |
| HNRNPA1 | 34723.91696 | -0.349644528 | 0.070639314 | 3.66E-07 | 5.04E-06 |
| PSIP1 | 1965.706078 | 0.349630548 | 0.113442746 | 0.000956199 | 0.00572562 |
| HSD17B8 | 860.3363167 | -0.34952068 | 0.103514093 | 0.000345917 | 0.00235929 |
| HINT2 | 676.0018135 | 0.349468774 | 0.128980821 | 0.003067645 | 0.015375515 |
| SNORD77 | 7.925312596 | -0.349460523 | 0.595055271 | 0.022886661 | 0.079170658 |
| AGPAT3 | 1415.594228 | 0.349326448 | 0.097326468 | 0.000155194 | 0.001167971 |
| RPSAP12 | 4444.422419 | -0.349246683 | 0.082053821 | 1.00E-05 | 0.00010099 |
| ATRIP | 161.125037 | -0.349145179 | 0.176375781 | 0.019695325 | 0.070004914 |
| TSC22D4 | 563.8534113 | 0.34906082 | 0.126682106 | 0.002630495 | 0.013492973 |

| | | | | | |
|---|---|---|---|---|---|
| TRMT12 | 263.1818722 | -0.348983654 | 0.140474688 | 0.005765846 | 0.025914133 |
| SYNJ2 | 462.2835898 | 0.348916533 | 0.113982897 | 0.001002673 | 0.005958573 |
| CNPY3-GNMT | 275.4012973 | 0.348871718 | 0.135155464 | 0.004360619 | 0.020530951 |
| PSAT1 | 7254.758118 | 0.348742831 | 0.066293405 | 6.95E-08 | 1.08E-06 |
| LPIN1 | 3904.979442 | 0.348633932 | 0.07147913 | 5.05E-07 | 6.76E-06 |
| EPRS1 | 6822.042748 | -0.348316997 | 0.074252592 | 1.32E-06 | 1.62E-05 |
| TPO | 474.0977667 | 0.348195656 | 0.125225012 | 0.002425145 | 0.012645659 |
| EZH1 | 372.3737903 | 0.348122263 | 0.122560934 | 0.002038699 | 0.01090307 |
| AKAP12 | 3.90462585 | 0.34809998 | 0.702214449 | 0.00292604 | 0.014759753 |
| TSC2 | 467.9880186 | -0.347990694 | 0.116633116 | 0.00131034 | 0.00746174 |
| STK11 | 895.7614177 | 0.347935412 | 0.105850777 | 0.000470763 | 0.003096168 |
| PPIB | 18764.98135 | -0.347933719 | 0.062636131 | 1.38E-08 | 2.38E-07 |
| CENPB | 1664.273121 | 0.347719092 | 0.113339424 | 0.000990813 | 0.005900241 |
| EIF2S3B | 659.6336723 | -0.347693857 | 0.107574153 | 0.000571124 | 0.003667095 |
| CTH | 410.4194494 | -0.347689424 | 0.11399803 | 0.001062127 | 0.006260331 |
| PIGBOS1 | 242.2180747 | 0.34761809 | 0.145368699 | 0.007266561 | 0.031427331 |
| YJU2 | 661.8648803 | 0.347611989 | 0.11417507 | 0.001069133 | 0.006293055 |
| BBS7 | 417.3858404 | -0.347553822 | 0.117551895 | 0.001426348 | 0.008048244 |
| ZNF641 | 340.6831414 | 0.347327618 | 0.127152737 | 0.002823882 | 0.014374242 |
| CYP1A1 | 5.15733928 | 0.34732003 | 0.631286064 | 0.014286336 | 0.054241718 |
| ARSB | 500.987664 | -0.347319893 | 0.112069777 | 0.000903854 | 0.005449872 |
| SOHLH1 | 986.8377559 | 0.347265695 | 0.120916102 | 0.001855857 | 0.010082627 |
| LOC202319 | 183.5265936 | -0.347213332 | 0.17215782 | 0.018437123 | 0.066432723 |
| F8A1 | 229.6412137 | 0.347153838 | 0.156355012 | 0.011293588 | 0.044969201 |
| NCF1C | 7.390197312 | 0.346997589 | 0.573537992 | 0.029562755 | 0.096646826 |
| PML | 385.9853563 | 0.346811694 | 0.121754759 | 0.002003524 | 0.010738171 |
| CIAO3 | 157.398723 | -0.346754078 | 0.170242515 | 0.017608481 | 0.064073559 |
| TBC1D24 | 156.057628 | -0.346747557 | 0.174929826 | 0.019897008 | 0.070620308 |
| PSMG3 | 1025.236996 | -0.346692309 | 0.099446541 | 0.000232944 | 0.001675202 |
| LRPPRC | 14216.82601 | -0.346588696 | 0.063300187 | 2.16E-08 | 3.66E-07 |
| BROX | 514.072093 | 0.346509221 | 0.164852226 | 0.014920341 | 0.056046756 |
| MEA1 | 1413.207573 | 0.34643403 | 0.084939653 | 2.16E-05 | 0.000203708 |
| LIMS1 | 2313.334217 | 0.34641676 | 0.148179955 | 0.008292656 | 0.034906155 |
| TRIOBP | 119.8411792 | 0.34640929 | 0.192436936 | 0.028609498 | 0.094278423 |
| LONP2 | 1391.49574 | -0.3462878 | 0.08040707 | 8.11E-06 | 8.35E-05 |
| IL12RB1 | 713.664823 | 0.346240477 | 0.106219265 | 0.000521283 | 0.003384724 |
| NCBP2AS2 | 465.0421987 | 0.346126545 | 0.128712509 | 0.003234151 | 0.016088996 |
| SWI5 | 453.1551365 | 0.346055795 | 0.114144392 | 0.001117566 | 0.006542558 |
| RAB33B | 247.8443593 | -0.345786823 | 0.167527111 | 0.01638075 | 0.060320761 |
| LUC7L3 | 1838.427468 | -0.345564293 | 0.092940964 | 9.60E-05 | 0.000767823 |
| LOC100130394 | 740.6332605 | -0.345457641 | 0.102045315 | 0.000339371 | 0.00232377 |
| AKT1 | 1580.966387 | 0.345360443 | 0.096934931 | 0.000174576 | 0.001296924 |
| SUMF1 | 373.6189573 | -0.345311483 | 0.117338343 | 0.001516483 | 0.008479477 |
| ABCB10 | 599.7212825 | -0.344983136 | 0.127926628 | 0.00318999 | 0.015919611 |
| RBM25 | 1837.781528 | -0.344949036 | 0.097147863 | 0.000183013 | 0.001352642 |
| TEP1 | 162.6703344 | 0.344941474 | 0.174588916 | 0.020011388 | 0.070953565 |
| ASB6 | 748.9725579 | 0.344865831 | 0.119649952 | 0.001815699 | 0.009890677 |
| COX2 | 78648.49705 | -0.344692329 | 0.080453877 | 8.78E-06 | 8.95E-05 |
| DPH5 | 659.2321788 | -0.344684044 | 0.099612851 | 0.000257333 | 0.001821788 |
| IKBKG | 390.1022925 | 0.344640158 | 0.116270757 | 0.001405898 | 0.0079458 |

| | | | | | |
|---|---|---|---|---|---|
| EML1 | 147.0398553 | -0.344485817 | 0.175203529 | 0.020840059 | 0.073380906 |
| RNF19B | 605.1535003 | 0.344341753 | 0.106503757 | 0.000573733 | 0.003682483 |
| SCRN1 | 566.1757625 | 0.34430982 | 0.105722109 | 0.000532528 | 0.003443532 |
| DDX31 | 769.917454 | 0.344289187 | 0.091583591 | 8.16E-05 | 0.000663834 |
| GPAT4 | 1997.82561 | -0.34423186 | 0.07753602 | 4.44E-06 | 4.84E-05 |
| TTC7A | 1150.517618 | 0.344150925 | 0.096006065 | 0.000160297 | 0.001202196 |
| BUB1 | 2802.274646 | 0.343921298 | 0.073184612 | 1.27E-06 | 1.56E-05 |
| CYTB | 29584.55861 | -0.343321826 | 0.089831881 | 6.40E-05 | 0.000537144 |
| TTK | 1283.281695 | 0.34328982 | 0.104252678 | 0.000467039 | 0.003078686 |
| TPST2 | 2345.443941 | 0.343263448 | 0.097069264 | 0.000191995 | 0.00141 |
| FAHD2B | 827.9468242 | -0.343196787 | 0.096417851 | 0.000179632 | 0.001329921 |
| SKA3 | 1137.410487 | 0.343187918 | 0.090984851 | 7.77E-05 | 0.00063635 |
| MRPS27 | 3576.989165 | -0.34282781 | 0.069331654 | 3.90E-07 | 5.31E-06 |
| RALB | 998.9291624 | 0.342802664 | 0.085008676 | 2.68E-05 | 0.000246954 |
| ARL13B | 438.9154435 | 0.342611552 | 0.142915413 | 0.00730753 | 0.031541597 |
| ARFIP2 | 809.3554858 | -0.342327623 | 0.093989891 | 0.000131204 | 0.001008906 |
| NPM3 | 2169.519368 | -0.34232185 | 0.082874602 | 1.79E-05 | 0.000171486 |
| MPND | 198.5926913 | 0.342320657 | 0.16522915 | 0.016474266 | 0.060582898 |
| KARS1 | 8029.745573 | -0.34228606 | 0.062311634 | 1.99E-08 | 3.38E-07 |
| ERC1 | 2197.608141 | -0.342281827 | 0.079101668 | 7.44E-06 | 7.71E-05 |
| CEP97 | 878.6538679 | 0.342241797 | 0.125024928 | 0.002833753 | 0.014411298 |
| TMEM43 | 2285.729702 | -0.342219927 | 0.081682349 | 1.38E-05 | 0.00013589 |
| PHLDA1 | 152.4069865 | -0.342107433 | 0.172402951 | 0.020116884 | 0.071240099 |
| TCTN3 | 1653.480108 | -0.342081305 | 0.083617928 | 2.13E-05 | 0.000200336 |
| EPN1 | 1000.528978 | 0.342070356 | 0.10389312 | 0.000467259 | 0.003078881 |
| C2orf42 | 532.769307 | 0.342049188 | 0.107124669 | 0.000666658 | 0.004188867 |
| BOLA2-SMG1P6 | 710.3791596 | 0.342025513 | 0.097608841 | 0.000217286 | 0.001569773 |
| TP53RK | 1281.770539 | 0.341945846 | 0.093335885 | 0.000119553 | 0.000928388 |
| MOB4 | 883.6169968 | -0.341891496 | 0.121750939 | 0.002318691 | 0.012167507 |
| ND1 | 23687.75207 | -0.341872662 | 0.08812209 | 5.13E-05 | 0.000439981 |
| PPM1K | 496.8616591 | -0.34159792 | 0.134167875 | 0.005021756 | 0.023171261 |
| DHRS4L2 | 412.0772031 | -0.341475966 | 0.117641639 | 0.001745749 | 0.009575904 |
| SOHLH2 | 252.8627033 | -0.341353038 | 0.137924718 | 0.006003098 | 0.026820342 |
| LINC01806 | 377.3898683 | -0.34131417 | 0.140689913 | 0.006841584 | 0.029925665 |
| SET | 18074.54698 | 0.341235384 | 0.078620364 | 6.93E-06 | 7.23E-05 |
| SNRNP25 | 1402.652143 | -0.34118135 | 0.094534153 | 0.000150445 | 0.001138161 |
| TIMM17A | 5116.985998 | -0.341087277 | 0.082747238 | 1.84E-05 | 0.000176678 |
| KLHL42 | 754.7879173 | 0.340973752 | 0.11424975 | 0.001340372 | 0.007610239 |
| TMEM245 | 1252.061354 | 0.340903965 | 0.092501758 | 0.000110284 | 0.000865707 |
| IL17RA | 558.0564427 | 0.340887796 | 0.117170424 | 0.001688857 | 0.009311029 |
| RPL12P50 | 6.581790053 | 0.340832672 | 0.590757331 | 0.020792601 | 0.073228691 |
| ZNF2 | 218.3081056 | 0.340821512 | 0.1459618 | 0.008683142 | 0.036222438 |
| BMP8A | 2539.236984 | -0.340737285 | 0.086921837 | 4.34E-05 | 0.000378948 |
| ZBP1 | 783.4506695 | 0.340717978 | 0.095302471 | 0.000169397 | 0.001263604 |
| ANKRD40 | 1926.540091 | 0.340715205 | 0.078907251 | 7.74E-06 | 7.99E-05 |
| GAPVD1 | 1201.748344 | 0.340684202 | 0.082897251 | 1.93E-05 | 0.000183964 |
| ELF1 | 4735.404799 | 0.340635271 | 0.08962385 | 6.99E-05 | 0.000579717 |
| PHYKPL | 628.1460718 | -0.340628844 | 0.129139143 | 0.003893957 | 0.018694664 |
| LYSMD3 | 1551.853158 | -0.340595663 | 0.161881342 | 0.015412573 | 0.057546254 |
| HDHD3 | 177.2025628 | 0.340574776 | 0.177023764 | 0.022942336 | 0.079288124 |

| | | | | | |
|---|---|---|---|---|---|
| MSANTD3 | 259.9298347 | 0.340522812 | 0.138036176 | 0.00619285 | 0.027529406 |
| ING1 | 557.4775829 | -0.340364589 | 0.103185479 | 0.000467424 | 0.003078881 |
| P2RX5 | 1075.249719 | -0.340044973 | 0.089482043 | 7.12E-05 | 0.000589653 |
| SEM1 | 1713.711227 | -0.340029885 | 0.099591808 | 0.000310104 | 0.002145867 |
| LSM1 | 1498.604256 | -0.339903575 | 0.089457242 | 7.14E-05 | 0.000590477 |
| ERP27 | 5.21966706 | 0.339800866 | 0.614244647 | 0.014332961 | 0.054394889 |
| ANKRD16 | 194.370276 | 0.339729057 | 0.166973755 | 0.018089795 | 0.065465065 |
| C2CD2L | 358.0125431 | 0.339581225 | 0.130137963 | 0.00418472 | 0.01987025 |
| GOLGA2 | 829.2309243 | 0.339527126 | 0.128002827 | 0.003701001 | 0.017897299 |
| ACTG1 | 97739.00809 | -0.339474866 | 0.085718099 | 3.71E-05 | 0.000328917 |
| LONRF1 | 169.7500703 | 0.339415526 | 0.16437908 | 0.016834889 | 0.061660274 |
| NR1D1 | 221.4101069 | 0.33927105 | 0.150421859 | 0.010759824 | 0.043245816 |
| ATRIP-TREX1 | 160.491431 | -0.339185635 | 0.175812038 | 0.022957243 | 0.079303927 |
| RING1 | 572.4856942 | 0.339177627 | 0.11447489 | 0.001436148 | 0.008090364 |
| PLA2G15 | 476.4915137 | -0.339071963 | 0.12398803 | 0.002947067 | 0.01484852 |
| TMEM68 | 738.7526805 | -0.338794799 | 0.119022084 | 0.002027656 | 0.010857425 |
| REV3L | 503.9270644 | -0.338793207 | 0.111456016 | 0.001132584 | 0.006619293 |
| DCC | 810.4938861 | -0.338756412 | 0.101118186 | 0.000391574 | 0.002632331 |
| GMFG | 3017.552861 | 0.338754604 | 0.07374232 | 2.17E-06 | 2.54E-05 |
| CCSER2 | 451.3174397 | 0.338728513 | 0.116181619 | 0.001669826 | 0.009217845 |
| ARMC1 | 2228.134732 | -0.338535985 | 0.142421828 | 0.007899987 | 0.033558529 |
| SHARPIN | 247.816023 | 0.338488637 | 0.181242243 | 0.025989436 | 0.087475038 |
| GPRIN1 | 298.6358023 | 0.338329671 | 0.145908716 | 0.009208521 | 0.038030992 |
| NDUFAF2 | 1211.428778 | -0.338173786 | 0.086605365 | 4.78E-05 | 0.000412794 |
| G2E3 | 497.3083282 | 0.338043118 | 0.16887287 | 0.01940186 | 0.069189048 |
| HYLS1 | 472.7224066 | 0.337947284 | 0.122897651 | 0.002786263 | 0.014199444 |
| DNTTIP2 | 1943.106544 | -0.337892736 | 0.078948498 | 9.43E-06 | 9.54E-05 |
| MYO1E | 152.3497248 | -0.337592835 | 0.182150465 | 0.026938459 | 0.08995243 |
| CD59 | 6170.46137 | -0.337551648 | 0.066203417 | 1.74E-07 | 2.53E-06 |
| RPS11P5 | 289.3699469 | 0.337549102 | 0.164096444 | 0.017288573 | 0.063108394 |
| PDIA5 | 2072.632406 | -0.337313824 | 0.078476718 | 8.68E-06 | 8.86E-05 |
| GRK2 | 3228.348465 | 0.337289092 | 0.104882367 | 0.000626315 | 0.003974311 |
| EEF1G | 93866.27871 | -0.337278356 | 0.068399527 | 6.30E-07 | 8.28E-06 |
| EML2-AS1 | 146.4839285 | -0.337267594 | 0.169476233 | 0.020333258 | 0.07184473 |
| RNU5A-1 | 8.021062297 | 0.337177724 | 0.655999564 | 0.004453818 | 0.0209129 |
| UCKL1 | 541.9570524 | 0.336994248 | 0.109463487 | 0.001001562 | 0.005954028 |
| SDHC | 6921.921204 | -0.336881694 | 0.076842106 | 5.89E-06 | 6.24E-05 |
| GRK5 | 213.2608236 | -0.336756219 | 0.149539084 | 0.01100382 | 0.044006413 |
| CRELD2 | 1008.458819 | -0.336577752 | 0.105050395 | 0.000666562 | 0.004188867 |
| RNPS1 | 2861.284487 | -0.33651349 | 0.078452873 | 9.04E-06 | 9.17E-05 |
| FAAP100 | 194.9265036 | -0.336455648 | 0.181538953 | 0.027194916 | 0.090569059 |
| MAU2 | 1026.38036 | 0.336450106 | 0.110590337 | 0.001127505 | 0.006591832 |
| RPSAP9 | 1611.569864 | -0.336432243 | 0.099656635 | 0.000363354 | 0.002465599 |
| NCDN | 623.6253906 | -0.336408912 | 0.118529466 | 0.002183185 | 0.011572148 |
| BPGM | 445.0989559 | 0.336360623 | 0.109074438 | 0.000980928 | 0.005851443 |
| EAF1 | 1366.910314 | -0.336282887 | 0.081269002 | 1.76E-05 | 0.000169443 |
| MGRN1 | 516.7865856 | -0.336266346 | 0.119362654 | 0.002320423 | 0.012172905 |
| SLC38A5 | 7085.605756 | -0.336254305 | 0.083622509 | 2.92E-05 | 0.000266526 |
| DENND6B | 460.2265289 | -0.336173046 | 0.117678567 | 0.002056052 | 0.010975538 |
| TOP1MT | 782.3329983 | -0.336115632 | 0.09549794 | 0.000213036 | 0.001543577 |

| | | | | | |
|---|---|---|---|---|---|
| DEAF1 | 331.489909 | 0.336066728 | 0.144479403 | 0.009134146 | 0.037777887 |
| FOXM1 | 1702.190359 | 0.335734724 | 0.090526142 | 0.000102612 | 0.000814735 |
| FDPS | 14718.46404 | -0.335647802 | 0.07040633 | 9.55E-07 | 1.21E-05 |
| ATP6V0D1 | 2041.859584 | -0.335641118 | 0.084178422 | 3.34E-05 | 0.000300499 |
| ERCC6 | 288.2648573 | 0.335545953 | 0.137288198 | 0.006660334 | 0.029243663 |
| HSP90AA3P | 440.55311 | -0.3355307 | 0.143441573 | 0.008828635 | 0.036751314 |
| WDR59 | 775.4691511 | -0.335499262 | 0.103521059 | 0.000586508 | 0.003756124 |
| SEPSECS | 308.4663646 | 0.335419778 | 0.138175261 | 0.006966092 | 0.030353419 |
| TBL3 | 792.3257236 | -0.335363656 | 0.106703969 | 0.0008217 | 0.005026276 |
| CORO2A | 392.4542651 | 0.334971681 | 0.116145594 | 0.001878397 | 0.010196929 |
| TMEM109 | 3347.048031 | 0.334707208 | 0.085274291 | 4.30E-05 | 0.000375827 |
| PSMG4 | 271.9761148 | -0.334600757 | 0.140063139 | 0.007869716 | 0.033491723 |
| TNFRSF14 | 430.4232179 | 0.334546992 | 0.122027147 | 0.002916773 | 0.014725877 |
| RND1 | 305.2123998 | 0.334501351 | 0.129133706 | 0.004512085 | 0.021134914 |
| EXOSC9 | 1048.071697 | -0.334421882 | 0.100154603 | 0.000416657 | 0.002783663 |
| CCDC169-SOHLH2 | 247.5358568 | -0.334377752 | 0.137297985 | 0.006983936 | 0.030402418 |
| F12 | 511.8411991 | -0.334322288 | 0.143321686 | 0.009166861 | 0.037899473 |
| R3HCC1 | 1024.824561 | -0.334209561 | 0.090583104 | 0.000113204 | 0.00088613 |
| ZNF304 | 422.0857655 | 0.334124974 | 0.110648107 | 0.001225453 | 0.007054912 |
| RPS13 | 27834.89052 | -0.334104872 | 0.075416663 | 4.91E-06 | 5.30E-05 |
| CAB39L | 221.769844 | 0.334063077 | 0.149980083 | 0.011713518 | 0.046142628 |
| UBAP2L | 6243.24372 | -0.333951517 | 0.070070845 | 9.64E-07 | 1.22E-05 |
| SLC25A11 | 1620.08905 | 0.333939158 | 0.089049814 | 8.82E-05 | 0.000710226 |
| NCAPH | 1528.959329 | 0.333832139 | 0.078907451 | 1.17E-05 | 0.000117438 |
| CPNE1 | 2380.321919 | 0.333739422 | 0.081411051 | 2.09E-05 | 0.000197444 |
| NUP188 | 3271.517797 | 0.333658169 | 0.07645049 | 6.41E-06 | 6.72E-05 |
| HSPE1-MOB4 | 3159.788551 | -0.333518215 | 0.104907627 | 0.000724194 | 0.004512729 |
| STK38L | 2105.056147 | 0.333413296 | 0.124669497 | 0.003541602 | 0.017310168 |
| HSP90AB1 | 44159.27332 | -0.33324387 | 0.062122077 | 4.42E-08 | 7.15E-07 |
| CEP83 | 193.1424172 | -0.33312511 | 0.171680669 | 0.022814352 | 0.078983632 |
| MAP2K7 | 1171.181383 | 0.333108008 | 0.090385889 | 0.000113548 | 0.000887724 |
| TMEM116 | 244.995156 | 0.332900596 | 0.142413965 | 0.00895258 | 0.037176718 |
| BFAR | 1490.481571 | -0.332741923 | 0.086981132 | 6.63E-05 | 0.000554477 |
| ACOX1 | 786.8455368 | -0.332671569 | 0.099051281 | 0.000393716 | 0.002644676 |
| ZNF584 | 201.1392281 | 0.332575692 | 0.153030646 | 0.01354116 | 0.052074677 |
| TMPO | 5835.521301 | 0.332527112 | 0.114180099 | 0.001700759 | 0.009364342 |
| EMC3 | 2815.352814 | -0.332404422 | 0.073061458 | 2.77E-06 | 3.16E-05 |
| PCYT1A | 2211.177112 | 0.332381311 | 0.071555558 | 1.76E-06 | 2.11E-05 |
| DGAT1 | 156.2466811 | 0.332367074 | 0.169764 | 0.022071462 | 0.076965667 |
| H1-2 | 1185.752572 | -0.332268601 | 0.163401127 | 0.018445157 | 0.066442734 |
| PM20D2 | 2516.759821 | -0.33196679 | 0.132306442 | 0.005730313 | 0.025767808 |
| QKI | 1875.063416 | -0.331766129 | 0.135797103 | 0.006848108 | 0.029938199 |
| BRIP1 | 771.7062962 | 0.331757292 | 0.13411671 | 0.006263207 | 0.027781749 |
| LYRM1 | 361.4145283 | -0.331691827 | 0.146820255 | 0.011047015 | 0.044149816 |
| FKBP8 | 5137.176634 | 0.331606183 | 0.09265904 | 0.000172718 | 0.001285881 |
| C13orf46 | 177.7827663 | -0.3315259 | 0.157765568 | 0.016153168 | 0.059693645 |
| MCFD2 | 2304.877929 | 0.331456702 | 0.075696501 | 5.94E-06 | 6.28E-05 |
| MYEOV | 174.6945955 | -0.331428015 | 0.165184827 | 0.020167291 | 0.071389408 |
| LOC642502 | 2299.107469 | -0.331274883 | 0.078454268 | 1.24E-05 | 0.000123597 |
| SMARCC1 | 5855.419293 | -0.331235668 | 0.063009323 | 7.67E-08 | 1.19E-06 |

| | | | | | |
|---|---|---|---|---|---|
| ARID5A | 1431.31562 | 0.330912159 | 0.097360618 | 0.000336461 | 0.002305699 |
| RBBP6 | 834.3688042 | -0.330629678 | 0.11583128 | 0.002094567 | 0.011157064 |
| ZFAT | 220.7755766 | 0.330618211 | 0.14213668 | 0.009313779 | 0.038419893 |
| HDGFL2 | 666.4651129 | 0.330608227 | 0.100212492 | 0.000480423 | 0.003147746 |
| ZNF44 | 279.1308862 | 0.330451911 | 0.131488199 | 0.005665564 | 0.025549629 |
| ZNF330 | 1418.237059 | -0.330449512 | 0.101114747 | 0.000539467 | 0.003479311 |
| SLC25A13 | 822.1717186 | -0.330371795 | 0.100036546 | 0.000480059 | 0.003146546 |
| TMSB4XP6 | 1487.796797 | 0.330324022 | 0.099844593 | 0.000468161 | 0.003082566 |
| EMSY | 413.9827908 | 0.330199227 | 0.113866566 | 0.001804638 | 0.00983971 |
| RRBP1 | 4355.188653 | 0.330198573 | 0.125170584 | 0.004003512 | 0.019114645 |
| RPS14P1 | 2113.127552 | -0.330025974 | 0.078130659 | 1.24E-05 | 0.000123542 |
| PITPNM1 | 378.122326 | 0.32986336 | 0.142634369 | 0.009697983 | 0.039777349 |
| FOXN2 | 925.1086626 | 0.329803028 | 0.163305469 | 0.019392132 | 0.06918285 |
| MAP4 | 2262.981106 | -0.329734538 | 0.083679008 | 4.16E-05 | 0.000364025 |
| KBTBD6 | 532.9633477 | 0.329728201 | 0.125702054 | 0.00415181 | 0.019724797 |
| NUBPL | 196.4176951 | -0.329719572 | 0.154218654 | 0.014900204 | 0.056017032 |
| TAMM41 | 347.4607917 | -0.32967707 | 0.121500512 | 0.003241977 | 0.016114042 |
| NCSTN | 3844.518393 | -0.329568095 | 0.07433927 | 4.83E-06 | 5.21E-05 |
| FAM216A | 492.9250602 | -0.329552087 | 0.113965214 | 0.001892046 | 0.010256425 |
| SLC35A5 | 1715.338223 | 0.329423592 | 0.11608168 | 0.002208178 | 0.011683172 |
| NDUFAF1 | 796.8087933 | -0.329377918 | 0.108671576 | 0.001215474 | 0.007010095 |
| SND1 | 14561.64581 | -0.329354013 | 0.063944446 | 1.37E-07 | 2.05E-06 |
| CFDP1 | 1198.382023 | -0.329341049 | 0.102275293 | 0.000639648 | 0.004047054 |
| CDAN1 | 213.3125331 | 0.329298915 | 0.161420957 | 0.018567316 | 0.066810659 |
| NUS1 | 4017.737311 | -0.329197701 | 0.111257472 | 0.001510739 | 0.008452819 |
| HNRNPA1P35 | 1553.130714 | -0.329104303 | 0.082102377 | 3.15E-05 | 0.000285326 |
| RPL39 | 19820.05103 | -0.328905506 | 0.086050936 | 6.71E-05 | 0.000560035 |
| PAPSS1 | 1664.261529 | -0.328828771 | 0.079704745 | 1.91E-05 | 0.00018177 |
| ELOB | 2764.583603 | -0.328803757 | 0.104438688 | 0.000824996 | 0.00504287 |
| RNF26 | 1952.094614 | 0.328792153 | 0.093904117 | 0.000231972 | 0.001669602 |
| RPL9 | 41562.1329 | -0.328768405 | 0.071995985 | 2.86E-06 | 3.24E-05 |
| UTP4 | 2658.168451 | -0.328763707 | 0.079720143 | 1.94E-05 | 0.000184307 |
| OXSR1 | 2371.979318 | -0.328622063 | 0.083878837 | 4.59E-05 | 0.000398138 |
| GATD3 | 2269.115132 | -0.328438548 | 0.072739666 | 3.30E-06 | 3.70E-05 |
| VIRMA | 2125.301267 | -0.328368264 | 0.080392335 | 2.29E-05 | 0.000214373 |
| ZNF23 | 197.7544597 | -0.328279651 | 0.155440581 | 0.015993362 | 0.059254823 |
| BMS1 | 1469.553786 | -0.328141483 | 0.080686624 | 2.46E-05 | 0.000228111 |
| HMGB2 | 7382.541094 | 0.328105795 | 0.08913351 | 0.00011835 | 0.000920283 |
| VDAC3 | 4809.310331 | -0.328080164 | 0.065924823 | 3.45E-07 | 4.77E-06 |
| ZNF888 | 226.9594971 | 0.328072233 | 0.166498893 | 0.021674148 | 0.075793865 |
| PRKAR1B | 1124.319683 | -0.328067877 | 0.096569911 | 0.000346907 | 0.002364183 |
| FLNA | 12425.57899 | 0.328062714 | 0.082465581 | 3.50E-05 | 0.000313304 |
| NPM1 | 55304.86066 | -0.327715841 | 0.071459747 | 2.58E-06 | 2.96E-05 |
| EPS8L2 | 813.7688397 | -0.327698826 | 0.094446466 | 0.00026708 | 0.001881621 |
| LDHB | 37455.14738 | -0.327622571 | 0.066308675 | 4.22E-07 | 5.72E-06 |
| EMC1 | 2080.274797 | -0.327375357 | 0.079637315 | 2.05E-05 | 0.000194138 |
| EEF1A1P25 | 2918.804844 | -0.327328476 | 0.094316002 | 0.000255511 | 0.001812656 |
| DSCR8 | 742.5444929 | -0.327298506 | 0.101089253 | 0.000614658 | 0.003913246 |
| ZNF562 | 281.3056052 | 0.327055443 | 0.154379831 | 0.015705844 | 0.058414539 |
| PRDX3 | 8491.777484 | -0.327037804 | 0.093632049 | 0.000240403 | 0.001720986 |

| | | | | | |
|---|---|---|---|---|---|
| SGSM2 | 273.328887 | 0.326882931 | 0.143346851 | 0.01060983 | 0.042777748 |
| MEX3C | 1029.562199 | 0.326864581 | 0.10125963 | 0.000621211 | 0.003947708 |
| DYRK4 | 605.2278516 | -0.326769638 | 0.102823262 | 0.000753097 | 0.004670983 |
| SYNE1 | 247.1897847 | 0.326766083 | 0.142977459 | 0.010420119 | 0.042189668 |
| BAZ2A | 3929.451957 | 0.326530333 | 0.086043123 | 7.48E-05 | 0.000615609 |
| AHCYL2 | 745.530477 | -0.326507931 | 0.103969347 | 0.000853665 | 0.00518517 |
| HDLBP | 10593.50413 | -0.326476936 | 0.071430738 | 2.56E-06 | 2.94E-05 |
| HYAL2 | 522.9767051 | -0.326225666 | 0.127147855 | 0.005056972 | 0.023308912 |
| TPCN2 | 393.5736211 | -0.326179507 | 0.14597829 | 0.012066797 | 0.047222763 |
| PLCB1 | 368.3100772 | 0.326176697 | 0.127972169 | 0.005192424 | 0.02383807 |
| ALDH16A1 | 1534.593513 | 0.326047977 | 0.129330055 | 0.005646528 | 0.025490334 |
| IL12A | 203.3990626 | 0.325892088 | 0.156076708 | 0.017023663 | 0.062298979 |
| RB1CC1 | 1612.527116 | -0.325817538 | 0.113914188 | 0.002107621 | 0.011216248 |
| BRWD1 | 886.9835306 | 0.325698595 | 0.118644902 | 0.002956867 | 0.014889232 |
| ZNF780A | 230.2918062 | 0.325654049 | 0.164933355 | 0.021792878 | 0.076147551 |
| NUS1P1 | 1670.618789 | -0.325644348 | 0.112122131 | 0.001838009 | 0.009996484 |
| TAF7 | 3063.757019 | -0.325563241 | 0.072111544 | 3.34E-06 | 3.73E-05 |
| TUBB4A | 521.4568144 | -0.325467242 | 0.121787195 | 0.003734592 | 0.018024508 |
| SPAG5 | 1810.553418 | 0.32517188 | 0.079414378 | 2.19E-05 | 0.000205826 |
| POLR3C | 4076.982619 | 0.325165934 | 0.0684375 | 1.04E-06 | 1.31E-05 |
| SLC6A6 | 1456.052595 | -0.325011491 | 0.080170641 | 2.65E-05 | 0.000244018 |
| DGKE | 638.4920297 | 0.324997653 | 0.126485254 | 0.004935901 | 0.022844142 |
| LOC101927209 | 137.8084401 | 0.324991374 | 0.17912445 | 0.030934461 | 0.099998517 |
| LOC105372948 | 170.747401 | 0.324935586 | 0.167412103 | 0.023565636 | 0.080985535 |
| RPS27A | 63106.34257 | -0.324894258 | 0.082041429 | 4.35E-05 | 0.000379537 |
| USP49 | 292.9359666 | 0.324815389 | 0.128778552 | 0.005640321 | 0.025468954 |
| DDIAS | 661.8179068 | 0.324757887 | 0.106743154 | 0.001178807 | 0.00683408 |
| TCP11L1 | 260.4270797 | 0.324754953 | 0.145944132 | 0.012256519 | 0.047900281 |
| UNKL | 272.6699046 | -0.324409725 | 0.141680659 | 0.010559471 | 0.042624326 |
| STARD4 | 589.6457882 | -0.324274068 | 0.151404142 | 0.015131162 | 0.056678851 |
| SYNJ1 | 355.2639586 | 0.324196493 | 0.15089293 | 0.014724086 | 0.055502175 |
| RBM39 | 6885.789873 | -0.324034868 | 0.067844706 | 9.51E-07 | 1.20E-05 |
| WDR62 | 467.8256612 | 0.324013354 | 0.116809309 | 0.002737825 | 0.013989646 |
| RAD51D | 624.1952219 | 0.323960584 | 0.112305075 | 0.001963393 | 0.010572194 |
| TOMM6 | 3872.541499 | -0.323778012 | 0.085386892 | 7.86E-05 | 0.000642587 |
| TAB3 | 749.9169994 | 0.323718442 | 0.107468756 | 0.001298533 | 0.00740257 |
| GHDC | 2155.063949 | -0.323661246 | 0.112880157 | 0.002091643 | 0.01114835 |
| RPL10AP6 | 2785.662341 | -0.323508088 | 0.080398928 | 3.01E-05 | 0.000273675 |
| MTFR2 | 587.014449 | 0.323506493 | 0.122705874 | 0.004124897 | 0.019618469 |
| IL10RB | 1764.694952 | 0.323380854 | 0.076721645 | 1.31E-05 | 0.000129485 |
| PLK1 | 2195.954776 | 0.323153053 | 0.094375837 | 0.000316309 | 0.002183152 |
| FAM168A | 492.2567157 | 0.323093368 | 0.110241475 | 0.001701231 | 0.009364342 |
| RNF216P1 | 546.7627285 | -0.323043065 | 0.105452576 | 0.001113835 | 0.006525127 |
| KLHL2 | 254.883814 | 0.32284965 | 0.157908531 | 0.018846339 | 0.067569783 |
| ARHGAP18 | 537.1195298 | 0.322806917 | 0.144644417 | 0.012151673 | 0.047522701 |
| GNB5 | 403.0988494 | 0.32277434 | 0.125787819 | 0.005041708 | 0.023250932 |
| KIF1C | 632.5172007 | -0.322727532 | 0.107578823 | 0.001376027 | 0.007789689 |
| ZNF124 | 213.3576435 | 0.322467307 | 0.157352092 | 0.018765637 | 0.067364124 |
| CRY2 | 555.867583 | -0.322454429 | 0.110657071 | 0.00181021 | 0.00986388 |
| TKFC | 2344.425591 | 0.322382841 | 0.105560206 | 0.001143869 | 0.006671182 |

| | | | | | |
|---|---|---|---|---|---|
| MBD6 | 294.3421655 | 0.322344503 | 0.143295033 | 0.011698991 | 0.04610638 |
| MEX3D | 274.0534786 | 0.322272323 | 0.171658093 | 0.027391341 | 0.091113302 |
| ZBTB17 | 178.0452092 | 0.322146755 | 0.17420083 | 0.028988564 | 0.095345673 |
| GPR15 | 5774.699597 | 0.322099689 | 0.097708045 | 0.000492335 | 0.003218484 |
| CACNB3 | 264.5116939 | 0.322069447 | 0.148560642 | 0.014343258 | 0.054422042 |
| KLHL5 | 1148.923859 | 0.321864689 | 0.113984703 | 0.002365249 | 0.012374325 |
| APOBEC3C | 4250.230076 | 0.321365158 | 0.071520876 | 3.61E-06 | 4.01E-05 |
| MOSPD3 | 200.950219 | 0.321319455 | 0.160642388 | 0.021248341 | 0.074560796 |
| POLR3E | 620.7435419 | -0.32123011 | 0.102838002 | 0.000918209 | 0.00552873 |
| YIF1B | 2888.718208 | -0.321195027 | 0.117077553 | 0.00307717 | 0.015409879 |
| FNDC3A | 2136.167236 | -0.321158115 | 0.111097644 | 0.001987229 | 0.010673959 |
| RPL13A | 92581.23793 | -0.32097504 | 0.076587754 | 1.50E-05 | 0.000146756 |
| TMBIM1 | 2233.664677 | 0.320906018 | 0.082458019 | 5.24E-05 | 0.000448835 |
| IDH2 | 503.455556 | 0.320893462 | 0.118657026 | 0.003419899 | 0.016829325 |
| ACAP1 | 2143.99653 | 0.320876792 | 0.099933589 | 0.000673132 | 0.004221884 |
| MYO1G | 1673.568403 | 0.320769439 | 0.092621191 | 0.000278219 | 0.001949782 |
| TRAF4 | 1265.393114 | -0.320624027 | 0.092603643 | 0.00028192 | 0.001970935 |
| ATP6 | 45999.09096 | -0.32058606 | 0.095499608 | 0.000346636 | 0.002363263 |
| SMG9 | 429.1103347 | 0.320548585 | 0.111486554 | 0.002042974 | 0.010919194 |
| FAM214A | 604.3275386 | 0.320308112 | 0.116243188 | 0.002938865 | 0.01482013 |
| LDHA | 63029.27874 | -0.32014567 | 0.065700833 | 5.99E-07 | 7.89E-06 |
| MDK | 352.5408085 | 0.320132853 | 0.128211371 | 0.006178326 | 0.02748268 |
| METTL18 | 388.9654868 | -0.319966812 | 0.125955076 | 0.005512276 | 0.024995045 |
| AXIN1 | 420.9351388 | -0.319951953 | 0.12231793 | 0.004496907 | 0.021079455 |
| DHX38 | 1232.715556 | -0.319859024 | 0.105044316 | 0.001198908 | 0.006934367 |
| RPS26P3 | 813.0023968 | -0.319846342 | 0.092618686 | 0.000291768 | 0.002033216 |
| TDRKH | 853.1501544 | -0.319843133 | 0.089124268 | 0.000175022 | 0.001298968 |
| PPA1 | 9336.432141 | -0.319810116 | 0.078233146 | 2.33E-05 | 0.000217733 |
| KLHL18 | 963.0934588 | -0.319807042 | 0.100244096 | 0.00073909 | 0.004595628 |
| ZBTB24 | 563.8648016 | 0.319665602 | 0.118411208 | 0.003467512 | 0.017024923 |
| CASP6 | 308.0210719 | 0.319613147 | 0.1378816 | 0.009916424 | 0.040519754 |
| ARHGEF7 | 1560.844486 | -0.319444218 | 0.07829486 | 2.41E-05 | 0.000224568 |
| CHD4 | 7274.012601 | -0.319337166 | 0.072073887 | 5.15E-06 | 5.53E-05 |
| TBC1D8B | 175.1791156 | -0.319303373 | 0.15692173 | 0.019935306 | 0.070727246 |
| CDKN2AIPNL | 481.5621955 | 0.31923571 | 0.114218929 | 0.002619926 | 0.013454705 |
| ZFX | 833.9339017 | 0.318969454 | 0.103144616 | 0.001018285 | 0.006034788 |
| ACBD7 | 361.197384 | 0.318917573 | 0.144566815 | 0.013166117 | 0.050915063 |
| SASS6 | 408.0903594 | 0.318912213 | 0.174844196 | 0.030741628 | 0.099523793 |
| ZDHHC12-DT | 186.0451956 | 0.318801281 | 0.153906388 | 0.018250906 | 0.065915714 |
| HOOK3 | 817.8259565 | -0.318792465 | 0.127203834 | 0.006087147 | 0.027146824 |
| RPL22 | 15734.18398 | -0.318656646 | 0.070447603 | 3.32E-06 | 3.72E-05 |
| CKAP2 | 3577.854098 | 0.318652845 | 0.105012589 | 0.001235249 | 0.007097146 |
| CHD9 | 608.4355641 | -0.318462827 | 0.143410013 | 0.012790068 | 0.049604838 |
| ABHD2 | 2603.797315 | 0.318311553 | 0.073763114 | 8.51E-06 | 8.71E-05 |
| CCDC134 | 1083.158503 | 0.318200059 | 0.105358146 | 0.001305141 | 0.007434581 |
| LSM3 | 2929.915079 | -0.318120608 | 0.082456085 | 6.19E-05 | 0.000521396 |
| ADH5 | 2507.046653 | -0.318083837 | 0.082492619 | 6.18E-05 | 0.000521161 |
| PRKCD | 1058.416164 | -0.317954414 | 0.096192932 | 0.000500501 | 0.003268164 |
| RPSAP26 | 641.0633947 | -0.317883253 | 0.146356592 | 0.014391035 | 0.05459136 |
| TMED4 | 1712.92636 | -0.317602266 | 0.083555986 | 7.75E-05 | 0.000635552 |

| | | | | | |
|---|---|---|---|---|---|
| RN7SL1 | 2188.069043 | 0.317530537 | 0.669453073 | 2.09E-06 | 2.46E-05 |
| LOC102724023 | 2217.091493 | -0.317471021 | 0.073558774 | 8.61E-06 | 8.80E-05 |
| PTPN9 | 559.0033217 | 0.317465228 | 0.109472286 | 0.001915306 | 0.010351857 |
| CARMIL2 | 512.7811804 | -0.317408491 | 0.132937193 | 0.008507187 | 0.035618705 |
| PAK2 | 5476.004751 | 0.317235542 | 0.086654401 | 0.00013032 | 0.001003446 |
| TBCB | 2593.408839 | 0.317197686 | 0.093028401 | 0.00034278 | 0.002342514 |
| DRC3 | 6.411708487 | -0.316892157 | 0.537234085 | 0.02804812 | 0.092834345 |
| LZTFL1 | 950.0356163 | -0.31676653 | 0.112648166 | 0.002538745 | 0.013111674 |
| SMPD3 | 247.0534637 | -0.316687534 | 0.154633255 | 0.019474915 | 0.069363863 |
| SH2B3 | 344.2148624 | 0.31663304 | 0.145030617 | 0.014098642 | 0.053658504 |
| CPTP | 290.312101 | 0.31661365 | 0.138124288 | 0.010821391 | 0.04340839 |
| STK36 | 240.7004074 | -0.316546454 | 0.145417205 | 0.014467305 | 0.05478889 |
| SEPTIN9 | 5059.501889 | 0.316535624 | 0.083463438 | 7.92E-05 | 0.000647031 |
| COX7A2L | 4723.60225 | -0.316479231 | 0.072032397 | 6.10E-06 | 6.44E-05 |
| SUSD1 | 270.7338066 | 0.316324744 | 0.130941014 | 0.007836054 | 0.033398215 |
| MOB2 | 241.1978842 | 0.316320504 | 0.13924083 | 0.011359292 | 0.045147775 |
| RSRC2 | 1522.750394 | -0.31632045 | 0.079790149 | 3.95E-05 | 0.000348068 |
| CEACAM21 | 185.1164289 | 0.316315211 | 0.160089521 | 0.022906847 | 0.07920884 |
| ETS1 | 311.7067026 | 0.316309991 | 0.152090136 | 0.0179319 | 0.065031771 |
| SRSF5 | 2028.528368 | -0.316218237 | 0.079292911 | 3.60E-05 | 0.000320896 |
| FBXO9 | 917.2977004 | 0.316209487 | 0.092943492 | 0.000353049 | 0.002402261 |
| AHCY | 15667.39228 | -0.316008544 | 0.079650508 | 4.01E-05 | 0.000352439 |
| DCPS | 1311.497638 | 0.315938467 | 0.095220625 | 0.000477988 | 0.003135346 |
| TCF25 | 1533.186763 | -0.315899386 | 0.094660388 | 0.00045113 | 0.002987463 |
| CNOT7 | 4193.805647 | -0.315796535 | 0.089172703 | 0.000212099 | 0.001538073 |
| CTU2 | 418.0183582 | -0.315754862 | 0.123335424 | 0.00534706 | 0.024392792 |
| DDR2 | 993.024344 | -0.315673073 | 0.098412029 | 0.000703418 | 0.004389733 |
| RYK | 507.6255588 | 0.315669444 | 0.129343592 | 0.007297752 | 0.031536625 |
| ATL2 | 2633.1936 | -0.315668701 | 0.100909928 | 0.000928816 | 0.005579015 |
| SRXN1 | 2833.475426 | -0.315654771 | 0.084630002 | 0.000103609 | 0.00082152 |
| PDCL | 653.6993453 | 0.315408017 | 0.132892103 | 0.008787588 | 0.036608355 |
| ARL6IP6 | 476.8831238 | 0.315370522 | 0.165578388 | 0.026569781 | 0.089064827 |
| HECTD3 | 498.2326162 | 0.315237968 | 0.106479931 | 0.001594064 | 0.008864612 |
| FDFT1 | 7279.044672 | -0.315228465 | 0.062637571 | 2.82E-07 | 3.97E-06 |
| SPCS1 | 4386.556441 | -0.315014288 | 0.072045248 | 6.72E-06 | 7.02E-05 |
| SNORA103 | 953.6982334 | -0.315012464 | 0.089553109 | 0.000234497 | 0.001684274 |
| CAPN15 | 517.8620314 | -0.314821444 | 0.125557771 | 0.006186875 | 0.027513641 |
| MED27 | 802.5420076 | 0.314697751 | 0.094099428 | 0.000437149 | 0.002904878 |
| RNF19A | 689.5421728 | -0.314686367 | 0.112689146 | 0.002707238 | 0.013853796 |
| TADA2A | 329.9907576 | 0.314630144 | 0.139191487 | 0.011703197 | 0.046112461 |
| BBS1 | 186.694678 | -0.314588066 | 0.164706234 | 0.026695495 | 0.089365143 |
| ISOC2 | 3853.740795 | 0.314532401 | 0.116676611 | 0.003578476 | 0.017460827 |
| HMOX2 | 624.3372171 | -0.314438848 | 0.111153568 | 0.002431215 | 0.012673493 |
| ENOSF1 | 466.2263741 | -0.314377803 | 0.117791309 | 0.003920802 | 0.018807903 |
| TCERG1 | 1954.527613 | -0.314240453 | 0.080988972 | 5.65E-05 | 0.000480039 |
| RCOR3 | 635.7699295 | 0.314209916 | 0.112797651 | 0.002740065 | 0.013996964 |
| CYCSP27 | 164.1279882 | -0.314171611 | 0.166651353 | 0.028159984 | 0.093115609 |
| SSBP1 | 4825.237121 | -0.314141383 | 0.077803284 | 2.94E-05 | 0.000267745 |
| JMY | 859.140776 | -0.314115238 | 0.127422735 | 0.006953491 | 0.030315611 |
| TSC22D4-C7ORF61 | 319.318122 | 0.314060745 | 0.138350805 | 0.011528162 | 0.045682651 |

| | | | | | |
|---|---|---|---|---|---|
| GAGE2A | 1413.827164 | -0.314028151 | 0.0829475 | 8.32E-05 | 0.000675486 |
| HEATR6 | 252.8286804 | 0.314000443 | 0.145418715 | 0.015129734 | 0.056678851 |
| AGA | 332.9788766 | -0.313861195 | 0.123717486 | 0.005714594 | 0.025717157 |
| SLC16A1 | 2423.03446 | -0.313570995 | 0.102038708 | 0.001124829 | 0.00657841 |
| STRBP | 2614.742878 | -0.313544912 | 0.085022464 | 0.000121684 | 0.000944085 |
| C19orf25 | 584.3177353 | 0.313531607 | 0.143037771 | 0.01406716 | 0.053585792 |
| CENATAC | 356.357511 | 0.313522895 | 0.123340771 | 0.005600848 | 0.02531712 |
| LTO1 | 165.1298634 | 0.313424674 | 0.165463775 | 0.02747545 | 0.091340442 |
| NDEL1 | 1265.010871 | -0.313370531 | 0.097359134 | 0.000674407 | 0.004228354 |
| TARS2 | 2426.692753 | -0.313265922 | 0.082498977 | 8.00E-05 | 0.000652573 |
| TAOK3 | 396.5968708 | 0.313172562 | 0.139024069 | 0.011991996 | 0.047025679 |
| OR51B4 | 4.841090439 | -0.313157248 | 0.564722822 | 0.014723147 | 0.055502175 |
| STRADB | 732.1803522 | -0.312774864 | 0.100399769 | 0.00098373 | 0.005862094 |
| PSPH | 1418.456015 | -0.312603447 | 0.088391179 | 0.000224681 | 0.001619819 |
| NSL1 | 398.3147151 | 0.312513973 | 0.132476225 | 0.009173042 | 0.037899473 |
| TPP2 | 3901.367265 | -0.312324243 | 0.079036976 | 4.21E-05 | 0.000367962 |
| LINC01580 | 6.79605658 | -0.312176486 | 0.535983431 | 0.024899418 | 0.084595493 |
| PTPN12 | 172.8296489 | 0.312067556 | 0.157767384 | 0.023076355 | 0.079620093 |
| RPL32 | 36650.70605 | -0.312047281 | 0.062815228 | 3.77E-07 | 5.17E-06 |
| SNX14 | 1339.344937 | 0.311994874 | 0.084085588 | 0.000111416 | 0.000873423 |
| MRPS21 | 3318.118932 | -0.311952159 | 0.07310317 | 1.10E-05 | 0.000110696 |
| PPP1R15B | 4409.680916 | -0.311808505 | 0.08513778 | 0.000136324 | 0.0010418 |
| MGME1 | 751.566075 | 0.311714203 | 0.094670693 | 0.000530795 | 0.003434893 |
| USPL1 | 1008.363039 | -0.311685721 | 0.084405831 | 0.000121252 | 0.000941154 |
| BIRC5 | 3146.645647 | 0.311541599 | 0.07333734 | 1.18E-05 | 0.000117636 |
| MLF2 | 3746.6099 | -0.311402889 | 0.097830725 | 0.00077891 | 0.004805707 |
| NUDT16L1 | 488.5537171 | -0.311347197 | 0.114341617 | 0.00341305 | 0.016800926 |
| ARMCX6 | 439.7454711 | -0.311340538 | 0.111995823 | 0.002862703 | 0.014524884 |
| PEX2 | 887.7341034 | -0.311258083 | 0.102278695 | 0.001257792 | 0.007198012 |
| EHD4 | 989.7855335 | -0.311195068 | 0.104573492 | 0.001562758 | 0.0087073 |
| DNASE2 | 1948.374001 | 0.310997985 | 0.09626597 | 0.000661352 | 0.004161564 |
| SLC9A3R1 | 6083.148174 | 0.31091904 | 0.085082581 | 0.000139746 | 0.001064195 |
| ANGEL1 | 538.2545225 | -0.310892498 | 0.108931402 | 0.002284079 | 0.012011214 |
| ERI1 | 1077.898167 | -0.310822628 | 0.121585523 | 0.005476315 | 0.02485277 |
| ACP1 | 8449.57562 | -0.310554815 | 0.062020758 | 3.10E-07 | 4.34E-06 |
| MAN1A1 | 6292.38465 | -0.31052835 | 0.123756872 | 0.006275008 | 0.027812723 |
| EPOP | 1051.674328 | -0.310406739 | 0.113920815 | 0.003395368 | 0.01673393 |
| DUS1L | 1916.628902 | -0.310387426 | 0.104895149 | 0.001650656 | 0.0091324 |
| OTUD3 | 236.1910295 | -0.310380374 | 0.140813762 | 0.013942091 | 0.053285187 |
| LMAN1 | 12937.07407 | -0.310303668 | 0.107670754 | 0.001750533 | 0.009596067 |
| TBCEL | 863.9720046 | 0.31027281 | 0.108752648 | 0.002271954 | 0.011962126 |
| RPS13P2 | 1582.803813 | -0.310092848 | 0.093924235 | 0.000523503 | 0.003396594 |
| MDH2 | 10413.89721 | -0.309860887 | 0.072337215 | 1.02E-05 | 0.000103118 |
| ZNF655 | 913.1817522 | -0.309802787 | 0.086891555 | 0.000198991 | 0.001455208 |
| ZNF473 | 635.6935536 | 0.30979777 | 0.099179481 | 0.000951364 | 0.005698638 |
| ZNF17 | 207.3636751 | 0.309776249 | 0.166343519 | 0.029938562 | 0.097595768 |
| PRR11 | 1437.137275 | 0.309754603 | 0.104349242 | 0.001580484 | 0.008794741 |
| AUNIP | 310.3860682 | 0.309534986 | 0.123418518 | 0.006280638 | 0.027830557 |
| SLC35D1 | 1435.510771 | -0.309503135 | 0.081082873 | 7.43E-05 | 0.000612413 |
| FTH1P8 | 355.6239076 | 0.30942898 | 0.118085411 | 0.004576376 | 0.021401643 |

| | | | | | |
|---|---|---|---|---|---|
| SCAMP3 | 4362.208068 | -0.309405628 | 0.085013943 | 0.000150236 | 0.001137078 |
| LINC00888 | 690.7934806 | 0.309309318 | 0.112291672 | 0.003075939 | 0.015408168 |
| SERPINI1 | 338.2970748 | -0.309259122 | 0.134729722 | 0.011107109 | 0.044359274 |
| LSM5 | 2256.40967 | -0.309072735 | 0.098947199 | 0.000963517 | 0.005761472 |
| FUT8 | 783.2292705 | 0.309004503 | 0.090441899 | 0.000343451 | 0.002345243 |
| SETD9 | 290.715788 | 0.308999738 | 0.143930961 | 0.015961388 | 0.059159178 |
| ND5 | 25620.5203 | -0.308946238 | 0.123901051 | 0.006626671 | 0.029118006 |
| RAB5B | 2242.499564 | 0.308618934 | 0.076911214 | 3.26E-05 | 0.00029434 |
| WDR76 | 776.0389935 | 0.308408639 | 0.098747721 | 0.000958167 | 0.005733443 |
| NCKAP1 | 452.8697067 | -0.308309044 | 0.127230908 | 0.007952029 | 0.033751295 |
| MLX | 1860.551668 | 0.308302957 | 0.079290055 | 5.55E-05 | 0.000472699 |
| RIN3 | 457.0053081 | 0.308290756 | 0.154676184 | 0.022654692 | 0.078588001 |
| ARL2-SNX15 | 333.8727705 | 0.308270109 | 0.133631298 | 0.010760883 | 0.043245816 |
| EXT1 | 1292.981728 | -0.308089019 | 0.080816376 | 7.64E-05 | 0.000627428 |
| KDM4A | 1243.045924 | 0.307985171 | 0.081115967 | 8.02E-05 | 0.000653988 |
| SARM1 | 587.5387537 | -0.307978162 | 0.129680433 | 0.009133483 | 0.037777887 |
| TRIM39 | 237.3979753 | -0.307886788 | 0.140894828 | 0.014717373 | 0.055502175 |
| PPP4R2 | 2629.430036 | -0.307881305 | 0.128335925 | 0.008522141 | 0.03567269 |
| ARRDC2 | 466.8981848 | 0.307825225 | 0.119037559 | 0.005074092 | 0.02337538 |
| STK3 | 345.0051244 | -0.307816719 | 0.127459152 | 0.008159184 | 0.034461606 |
| POLR1G | 688.5606453 | 0.307777326 | 0.109797674 | 0.002678722 | 0.013728153 |
| H2AZ1 | 15662.56424 | -0.307710808 | 0.079570691 | 6.27E-05 | 0.00052744 |
| SIDT1 | 1139.947659 | 0.307671941 | 0.089687578 | 0.000328039 | 0.002256019 |
| C16orf54 | 198.1848843 | 0.307565137 | 0.150814863 | 0.020589696 | 0.072602683 |
| SCRIB | 734.6966788 | -0.307358649 | 0.129127604 | 0.009019333 | 0.037365498 |
| ZBED5 | 352.4530477 | 0.307315348 | 0.13443722 | 0.011351299 | 0.045129366 |
| RPL7AP11 | 1222.035021 | 0.307096565 | 0.082524003 | 0.000109088 | 0.000857503 |
| TIMM17B | 1655.671807 | -0.307021942 | 0.09810459 | 0.000957852 | 0.005733443 |
| CIPC | 265.0442386 | 0.306989699 | 0.132582859 | 0.010586293 | 0.04269279 |
| KLHL24 | 2309.789831 | 0.306924359 | 0.11310034 | 0.003514093 | 0.017204848 |
| RPL29P12 | 213.6116048 | -0.306875207 | 0.160457092 | 0.027616225 | 0.091702813 |
| OTUD6B-AS1 | 507.7827774 | -0.30686239 | 0.157445279 | 0.025287039 | 0.085643586 |
| THRA | 287.4916769 | 0.306752952 | 0.139002592 | 0.013951773 | 0.053310427 |
| OXNAD1 | 876.036218 | -0.306728355 | 0.101662274 | 0.001390333 | 0.007868103 |
| EIF2B3 | 710.3230813 | -0.306723069 | 0.099057644 | 0.001071639 | 0.006303525 |
| PLBD2 | 930.7553805 | -0.306696882 | 0.114341931 | 0.003868256 | 0.018591894 |
| RFT1 | 1438.401089 | -0.306660821 | 0.08777577 | 0.000257692 | 0.001822901 |
| SYT1 | 788.9830841 | 0.306422049 | 0.105358973 | 0.001920495 | 0.010373975 |
| ZNF639 | 1023.594707 | -0.306379612 | 0.097404823 | 0.000903678 | 0.005449872 |
| TYMS | 10179.26771 | -0.306371453 | 0.06653744 | 2.34E-06 | 2.72E-05 |
| TMED3 | 4510.788894 | -0.30627249 | 0.082644252 | 0.000117047 | 0.000912124 |
| BUD23 | 4991.282261 | -0.306163979 | 0.069722753 | 6.40E-06 | 6.71E-05 |
| MAGEA12 | 2457.681355 | -0.306157343 | 0.088049688 | 0.000280175 | 0.001960316 |
| TRMT61A | 438.6090057 | 0.306144073 | 0.138370222 | 0.013740391 | 0.052700409 |
| E2F4 | 1177.700996 | -0.305974669 | 0.098878707 | 0.001076657 | 0.006328743 |
| TMSB4X | 37893.87346 | 0.305714011 | 0.065374727 | 1.51E-06 | 1.82E-05 |
| DIXDC1 | 1317.70944 | -0.305687419 | 0.111515846 | 0.00327771 | 0.016245019 |
| NDUFAF8 | 989.9820024 | -0.30557673 | 0.139396724 | 0.014722697 | 0.055502175 |
| PRKCB | 582.4184704 | 0.305553536 | 0.100242084 | 0.001231957 | 0.007080586 |
| FAM133B | 609.2311914 | -0.305480906 | 0.113903873 | 0.003901534 | 0.01872066 |

| | | | | | |
|---|---|---|---|---|---|
| PLA2G6 | 261.19187 | 0.305450089 | 0.155201783 | 0.024378922 | 0.083164729 |
| MAPK6 | 2284.996956 | -0.305384144 | 0.102935357 | 0.00164065 | 0.009094476 |
| ZNF766 | 511.8012908 | 0.305332566 | 0.105208885 | 0.001992548 | 0.010689268 |
| DNAJA3 | 2307.602464 | -0.305269256 | 0.073164562 | 1.70E-05 | 0.00016439 |
| DCP1B | 329.1565382 | -0.305242357 | 0.123181421 | 0.006921733 | 0.030192355 |
| CDK9 | 903.3340116 | 0.305178128 | 0.097256691 | 0.000926687 | 0.005568161 |
| XXYLT1 | 871.5357816 | 0.305146688 | 0.098149741 | 0.001017135 | 0.00603003 |
| ATAD2B | 210.7882562 | 0.304969882 | 0.147673843 | 0.019588788 | 0.069726421 |
| ITGAL | 1468.154346 | 0.3049068 | 0.085176885 | 0.000190093 | 0.001396627 |
| NCAPG | 2254.353052 | 0.304415572 | 0.075735832 | 3.22E-05 | 0.000291244 |
| ZNF16 | 224.8282162 | 0.304351967 | 0.15400998 | 0.024056243 | 0.082263422 |
| ZMYM2 | 1542.689575 | -0.304338715 | 0.08631499 | 0.000234687 | 0.001684938 |
| KCNJ4 | 6.902944339 | -0.304079589 | 0.530391978 | 0.022437573 | 0.078022381 |
| NANP | 281.2211398 | -0.304019456 | 0.138327231 | 0.014438216 | 0.054710413 |
| NOL10 | 2732.447423 | -0.303796147 | 0.076600283 | 4.12E-05 | 0.000361345 |
| COX4I1 | 16056.77419 | -0.303783729 | 0.07085607 | 1.03E-05 | 0.000103786 |
| SBDS | 1790.383682 | -0.303709306 | 0.092876827 | 0.000595852 | 0.003806113 |
| SLC25A15 | 823.6499338 | -0.303594201 | 0.097140961 | 0.000974863 | 0.005821277 |
| ZNF273 | 323.76727 | 0.303555657 | 0.162083629 | 0.030038167 | 0.097850064 |
| LOC105375631 | 4.886468523 | 0.303376582 | 0.533289806 | 0.019390134 | 0.06918285 |
| RAP2B | 1526.903762 | 0.303288324 | 0.091372178 | 0.000496915 | 0.003245971 |
| GZF1 | 791.6013525 | -0.303177227 | 0.102917284 | 0.001762765 | 0.009647849 |
| EEF2 | 117008.6091 | -0.303042264 | 0.068936741 | 6.30E-06 | 6.63E-05 |
| RPL39P3 | 17563.16447 | -0.302944235 | 0.07607777 | 3.88E-05 | 0.00034262 |
| HNRNPA1P2 | 276.7549015 | -0.302715376 | 0.135956115 | 0.013561753 | 0.052130715 |
| STAM2 | 469.8989045 | 0.302648012 | 0.12348836 | 0.007488548 | 0.032170476 |
| FOXK1 | 757.7209764 | 0.302406563 | 0.130435283 | 0.010749323 | 0.043219417 |
| TOB1 | 1101.564251 | -0.302253106 | 0.11448559 | 0.00446781 | 0.020967225 |
| COX3 | 135134.7273 | -0.30176516 | 0.071783252 | 1.50E-05 | 0.000146573 |
| RNF40 | 1087.507891 | -0.301706995 | 0.098728488 | 0.001242086 | 0.007124201 |
| FUBP3 | 1450.825335 | 0.301692738 | 0.123620242 | 0.007746572 | 0.033081971 |
| SLC25A10 | 553.4874776 | -0.30167971 | 0.131521104 | 0.011566803 | 0.045762469 |
| CLPTM1L | 13373.92184 | -0.301660479 | 0.088693984 | 0.000374762 | 0.00253407 |
| SDE2 | 623.623546 | 0.301598166 | 0.135316755 | 0.013388784 | 0.051625704 |
| CCNB1 | 4599.807073 | 0.301569878 | 0.069936683 | 9.08E-06 | 9.21E-05 |
| SNRK | 942.6693929 | -0.301506908 | 0.115875931 | 0.004988216 | 0.023028775 |
| BTN2A1 | 303.9492101 | 0.301462552 | 0.132114514 | 0.011858944 | 0.046630639 |
| CCNI | 3941.147697 | -0.301445439 | 0.077209521 | 5.36E-05 | 0.000458495 |
| KAT14 | 419.6421806 | 0.301444744 | 0.114254417 | 0.004451233 | 0.020906427 |
| HNRNPA1L2 | 2431.590444 | -0.301440136 | 0.075429398 | 3.67E-05 | 0.000326384 |
| RPL23A | 35013.05322 | -0.301425192 | 0.068219021 | 5.82E-06 | 6.18E-05 |
| ZNF507 | 899.9918867 | 0.301398598 | 0.113588974 | 0.004274203 | 0.020203357 |
| WDR35 | 10395.40967 | -0.301357402 | 0.078015423 | 6.36E-05 | 0.000534014 |
| CCAR2 | 2459.50263 | -0.301313362 | 0.0805429 | 0.000103285 | 0.000819703 |
| TOMM20P4 | 233.1762256 | -0.301278195 | 0.149541845 | 0.022599755 | 0.078460295 |
| BANP | 258.1818688 | 0.301175711 | 0.142618621 | 0.017911771 | 0.064999611 |
| TTC28 | 402.5624144 | -0.301101598 | 0.143638375 | 0.018577802 | 0.066828374 |
| ZGPAT | 360.5848021 | 0.301091482 | 0.131784418 | 0.011727642 | 0.046177257 |
| CLN3 | 658.0830995 | -0.301040699 | 0.105831738 | 0.002455522 | 0.012780968 |
| FBH1 | 1868.552997 | 0.301008755 | 0.084616069 | 0.000210146 | 0.001525189 |

| | | | | | |
|---|---|---|---|---|---|
| ENTPD6 | 1661.626405 | -0.300724841 | 0.086829467 | 0.000300288 | 0.00208755 |
| CLEC2B | 1083.242168 | 0.300657037 | 0.118949046 | 0.006128617 | 0.027296621 |
| MDN1 | 903.049406 | -0.300496223 | 0.10718476 | 0.002770968 | 0.014133979 |
| ZNF845 | 345.2896092 | 0.300492505 | 0.129658614 | 0.010786268 | 0.043297588 |
| RPUSD2 | 675.8320381 | 0.300444592 | 0.107043491 | 0.002731515 | 0.013961523 |
| AKAP17A | 713.0310649 | -0.300407194 | 0.11698645 | 0.005566531 | 0.025188299 |
| HIVEP2 | 533.8677343 | -0.30020386 | 0.118978353 | 0.006266185 | 0.027787842 |
| MRPL32 | 1589.565538 | -0.300175185 | 0.07690318 | 5.41E-05 | 0.000461669 |
| ZKSCAN8 | 573.4688022 | 0.300101878 | 0.130687383 | 0.011352054 | 0.045129366 |
| PPM1B | 1046.466048 | 0.300007268 | 0.109780563 | 0.003389627 | 0.0167136 |
| FCHSD1 | 238.1039914 | 0.2999668 | 0.142821717 | 0.018484392 | 0.066547654 |
| PTPN23 | 510.4332659 | -0.299959443 | 0.121975843 | 0.007474289 | 0.032125158 |
| AIP | 1183.234794 | 0.299810433 | 0.08627682 | 0.000284015 | 0.001983984 |
| GALNT2 | 3247.046501 | -0.299661186 | 0.076401113 | 5.04E-05 | 0.000432446 |
| GTF3C4 | 2290.487388 | 0.299660439 | 0.08147418 | 0.000132546 | 0.001016967 |
| UBQLN4 | 1787.563425 | -0.299538974 | 0.092655494 | 0.000690974 | 0.004324391 |
| DECR2 | 183.5416353 | -0.299446229 | 0.159064163 | 0.030332728 | 0.098494628 |
| TSPYL4 | 321.3197864 | -0.29941402 | 0.127250584 | 0.00987658 | 0.040398324 |
| LZIC | 525.401781 | 0.299085953 | 0.114604962 | 0.004894383 | 0.022686302 |
| TBC1D7-LOC100130357 | 388.1389507 | -0.299078417 | 0.114966555 | 0.005047811 | 0.023272882 |
| SUPT20H | 668.6620744 | 0.298913733 | 0.105287502 | 0.002491762 | 0.012926855 |
| CHMP3 | 796.0442345 | 0.298906574 | 0.091307575 | 0.000595429 | 0.003805187 |
| STK26 | 1840.181169 | 0.298894905 | 0.12446736 | 0.008644035 | 0.03608697 |
| CNTROB | 644.530816 | 0.298800002 | 0.097344197 | 0.001189468 | 0.006891272 |
| LACTB2 | 791.3530371 | -0.298747887 | 0.14482931 | 0.020355622 | 0.071879753 |
| ALG5 | 3526.052891 | -0.298580546 | 0.073587838 | 2.87E-05 | 0.000262271 |
| DCP2 | 1229.299746 | 0.298541474 | 0.121496797 | 0.007525817 | 0.032290527 |
| ACTL6A | 2691.792082 | 0.298518189 | 0.08173625 | 0.000146091 | 0.00110813 |
| EXOC6 | 621.2238367 | 0.298411469 | 0.122591843 | 0.007994397 | 0.033889589 |
| LCOR | 758.4301821 | 0.298268527 | 0.106117697 | 0.002707128 | 0.013853796 |
| RBM3 | 12428.62938 | -0.298193612 | 0.083359159 | 0.000196723 | 0.001439227 |
| LAMTOR1 | 1447.955503 | 0.298184096 | 0.097355612 | 0.001226204 | 0.007056889 |
| AGO3 | 356.3670873 | 0.298064296 | 0.120461762 | 0.007216451 | 0.031273094 |
| SENP1 | 1487.115119 | 0.298017819 | 0.083595412 | 0.000205765 | 0.001498414 |
| NBEAL1 | 897.2720884 | -0.297944356 | 0.091557803 | 0.000642303 | 0.00406237 |
| CCNG2 | 348.2048782 | -0.297762544 | 0.138813426 | 0.016808346 | 0.061576081 |
| NR2C1 | 226.4692828 | 0.297521887 | 0.145408671 | 0.021093961 | 0.074139257 |
| ATF7IP | 1531.096668 | 0.297398008 | 0.100433911 | 0.001684493 | 0.009289927 |
| TELO2 | 397.3235349 | -0.297369967 | 0.143949369 | 0.020458561 | 0.072199082 |
| NSMF | 996.3417848 | 0.297343919 | 0.101524177 | 0.001894769 | 0.010260075 |
| NUP54 | 892.1653896 | -0.29730994 | 0.105861143 | 0.002755519 | 0.014067606 |
| PDHB | 3526.103465 | -0.296921075 | 0.069513124 | 1.14E-05 | 0.000114386 |
| RPS6 | 98093.0749 | -0.296905041 | 0.064847235 | 2.90E-06 | 3.28E-05 |
| ITGAE | 671.4669008 | 0.29689864 | 0.104424938 | 0.002485582 | 0.012906396 |
| MRPS34 | 2655.960258 | -0.296836345 | 0.081793389 | 0.000163416 | 0.001221879 |
| UBA5 | 2031.681788 | -0.296419656 | 0.07233612 | 2.43E-05 | 0.000225434 |
| SCHIP1 | 6.484781178 | -0.296413948 | 0.511559158 | 0.024971069 | 0.084739172 |
| NLRP2 | 1739.865416 | 0.296355898 | 0.086362349 | 0.000340426 | 0.00232826 |
| TUBAP2 | 4607.783173 | 0.296315057 | 0.073949735 | 3.54E-05 | 0.000316564 |
| BPNT2 | 4591.537599 | -0.296247764 | 0.106994333 | 0.003096365 | 0.01549256 |

| | | | | | |
|---|---|---|---|---|---|
| COLGALT1 | 2215.462348 | -0.296150237 | 0.099811043 | 0.001696814 | 0.00934894 |
| COPB1 | 6427.505721 | -0.296059191 | 0.069680654 | 1.26E-05 | 0.000124851 |
| RPS26P39 | 296.1283806 | -0.296051055 | 0.147515744 | 0.023298304 | 0.080178204 |
| RPL17 | 23735.56283 | -0.295934126 | 0.069568496 | 1.30E-05 | 0.000128714 |
| NORAD | 7852.974148 | -0.295845808 | 0.105589311 | 0.002855096 | 0.014499031 |
| FAH | 1012.481687 | -0.295752744 | 0.088853297 | 0.000500932 | 0.003269746 |
| WAS | 896.9852785 | -0.295713894 | 0.094661628 | 0.001015067 | 0.006020561 |
| TRAF3IP3 | 729.1622609 | 0.295671926 | 0.098731053 | 0.001532253 | 0.008551763 |
| CXXC1 | 1276.37425 | 0.295544491 | 0.091930499 | 0.0007399 | 0.004599013 |
| E2F6 | 1047.152067 | -0.295373437 | 0.095818225 | 0.00116302 | 0.006760662 |
| TLE3 | 1293.62585 | 0.295339975 | 0.085342718 | 0.000306806 | 0.002126036 |
| UBAC2 | 1226.72842 | -0.295264131 | 0.08372269 | 0.000242587 | 0.001734467 |
| PHYH | 357.435093 | -0.295202466 | 0.122800106 | 0.008909344 | 0.037051661 |
| BID | 2113.31469 | 0.29493213 | 0.079625037 | 0.000122641 | 0.000950657 |
| CDCA2 | 1193.686754 | 0.294760631 | 0.084165367 | 0.00026034 | 0.001840719 |
| PPME1 | 573.7451257 | 0.294585778 | 0.101700141 | 0.002110539 | 0.011228325 |
| RPL23AP2 | 6533.92674 | -0.294516185 | 0.063788727 | 2.24E-06 | 2.61E-05 |
| LOC285591 | 397.198487 | -0.294441551 | 0.118604439 | 0.007177996 | 0.031129818 |
| GRIPAP1 | 421.5960537 | 0.294407961 | 0.114180629 | 0.005435041 | 0.02470953 |
| THUMPD3-AS1 | 244.448053 | -0.294169711 | 0.153892271 | 0.029063334 | 0.095519028 |
| SPOPL | 873.0260682 | 0.293987204 | 0.153731319 | 0.028894043 | 0.095088968 |
| SUV39H1 | 1080.198913 | 0.293854983 | 0.100731556 | 0.001989126 | 0.010680838 |
| PRDX2 | 7293.184328 | -0.293803974 | 0.085356547 | 0.00033416 | 0.002292653 |
| ZNF747 | 217.7032336 | -0.293798994 | 0.152082654 | 0.028002702 | 0.092749538 |
| TSPYL1 | 2093.723485 | -0.293792036 | 0.078707987 | 0.000109884 | 0.000862975 |
| KIF13B | 591.3379777 | 0.293786138 | 0.115146333 | 0.005906008 | 0.026434307 |
| ERCC6L | 887.3329057 | 0.293751601 | 0.094545838 | 0.001070071 | 0.006296441 |
| ZNF211 | 227.543993 | 0.293634901 | 0.148553669 | 0.025161333 | 0.085284557 |
| SKA2 | 2380.193585 | 0.293610069 | 0.125592673 | 0.010492313 | 0.042422457 |
| EXOC1 | 701.3626996 | 0.293374677 | 0.094131326 | 0.001034144 | 0.006116214 |
| KHDRBS3 | 278.9025084 | 0.293320952 | 0.138710622 | 0.018254305 | 0.065915714 |
| TUBA1B | 57731.1391 | 0.293313742 | 0.073896524 | 4.12E-05 | 0.000361345 |
| PKHD1 | 327.2723349 | 0.29325848 | 0.124761814 | 0.010239744 | 0.041605267 |
| ATAD5 | 426.2606853 | 0.293242278 | 0.134423373 | 0.015569935 | 0.057958848 |
| PTCH1 | 342.1707 | 0.293136326 | 0.137557823 | 0.017614674 | 0.064082628 |
| PABPC4 | 9163.224073 | -0.29309836 | 0.062896286 | 1.92E-06 | 2.27E-05 |
| MAMLD1 | 3.658575502 | 0.292924968 | 0.564132665 | 0.000525864 | 0.003408076 |
| PSMB4 | 15213.44171 | -0.292679072 | 0.080562523 | 0.000166913 | 0.001246948 |
| HEXA | 4249.073041 | -0.292662724 | 0.081967143 | 0.000208289 | 0.001513958 |
| PHTF2 | 1199.247676 | 0.292560646 | 0.125835004 | 0.010901314 | 0.043678401 |
| FMC1-LUC7L2 | 1764.778239 | -0.292437509 | 0.086402099 | 0.000410622 | 0.002749395 |
| ILF2 | 17227.87467 | -0.29231254 | 0.060077261 | 6.86E-07 | 8.94E-06 |
| MCM6 | 5108.259306 | -0.292198306 | 0.063654938 | 2.64E-06 | 3.02E-05 |
| GPX1 | 3083.684225 | -0.292160488 | 0.104188237 | 0.002867821 | 0.014542331 |
| HLA-G | 1276.015043 | 0.292030052 | 0.121714764 | 0.008902913 | 0.037033808 |
| CACNB1 | 260.9836965 | 0.292001397 | 0.131942228 | 0.014531021 | 0.054941855 |
| CSNK2A2 | 2349.057318 | -0.291940625 | 0.085892548 | 0.000391416 | 0.002632289 |
| NUP214 | 1266.067953 | 0.291853275 | 0.084270739 | 0.000310659 | 0.002147577 |
| NXPH4 | 397.5849585 | 0.291745281 | 0.125037109 | 0.010737893 | 0.043193519 |
| SLC9A8 | 358.0828848 | 0.291710139 | 0.134033727 | 0.015998809 | 0.059262328 |

| | | | | | |
|---|---|---|---|---|---|
| RAVER2 | 238.5462564 | -0.291676142 | 0.152313229 | 0.029173535 | 0.095772151 |
| DDX47 | 2882.440601 | -0.291623848 | 0.073553844 | 4.34E-05 | 0.000378888 |
| PIK3R4 | 1119.726106 | -0.291498649 | 0.085578966 | 0.000388148 | 0.002615392 |
| TMEM115 | 540.1970083 | -0.291480699 | 0.105769344 | 0.003313284 | 0.016399721 |
| GPR107 | 1773.448539 | 0.291404324 | 0.079615748 | 0.000146914 | 0.001113396 |
| ASAH1 | 2177.309363 | -0.291398188 | 0.094738047 | 0.001206707 | 0.00696784 |
| HCP5 | 884.802851 | 0.291347938 | 0.10206464 | 0.00244708 | 0.012740856 |
| PSME1 | 7524.925216 | 0.291335574 | 0.070568119 | 2.15E-05 | 0.000202007 |
| UBN1 | 1016.725114 | -0.291289904 | 0.102306972 | 0.002520338 | 0.013056917 |
| ZNF277 | 608.4198807 | -0.291232539 | 0.123730296 | 0.010270194 | 0.0417192 |
| ATL3 | 1373.724988 | 0.291227501 | 0.115674856 | 0.006553352 | 0.028847075 |
| UVRAG | 1112.816925 | 0.291215449 | 0.086264971 | 0.000425632 | 0.002834872 |
| ZNF761 | 306.0012091 | 0.290938529 | 0.126359211 | 0.011671828 | 0.046030754 |
| CCDC91 | 839.2615271 | 0.290790759 | 0.131304765 | 0.014511637 | 0.054904527 |
| YTHDC1 | 1171.884504 | -0.290740659 | 0.086113608 | 0.000433184 | 0.002880739 |
| UBE2V2 | 2627.253969 | -0.290637578 | 0.145409792 | 0.024849798 | 0.084468264 |
| TAPBPL | 960.1205917 | -0.290549798 | 0.093273603 | 0.001067029 | 0.006284946 |
| DCUN1D1 | 1367.767309 | 0.290461979 | 0.145892424 | 0.024305769 | 0.082952899 |
| RABGEF1 | 1070.304166 | -0.290438811 | 0.091708574 | 0.000890962 | 0.005377759 |
| DNAAF10 | 554.7853692 | 0.290304707 | 0.109519595 | 0.004498444 | 0.021079455 |
| PLAAT3 | 1054.12559 | 0.290294686 | 0.110138409 | 0.004713667 | 0.021983995 |
| PCNA | 10987.42013 | -0.290196476 | 0.070012377 | 2.02E-05 | 0.000191675 |
| HNRNPA1P54 | 271.4820298 | -0.290170458 | 0.137484215 | 0.018891042 | 0.067688015 |
| LOC107985051 | 5.0447856 | 0.290168628 | 0.501085482 | 0.02680255 | 0.089654193 |
| SNORD58B | 201.4156662 | -0.290149642 | 0.146805437 | 0.025534368 | 0.086278761 |
| RPL35P5 | 955.1386566 | 0.290068847 | 0.090603947 | 0.00079128 | 0.004869438 |
| MTCH1 | 1492.809836 | 0.289988006 | 0.103349121 | 0.002851051 | 0.014482739 |
| RPS3A | 49298.68708 | -0.289705696 | 0.066973782 | 1.06E-05 | 0.000106607 |
| C1orf198 | 206.7153571 | 0.289567756 | 0.150827684 | 0.028998602 | 0.095358532 |
| ARHGEF12 | 4787.388584 | 0.289456259 | 0.077482195 | 0.000109076 | 0.000857503 |
| MIEN1 | 1052.413342 | 0.289417782 | 0.10211454 | 0.002636516 | 0.013519855 |
| GFM1 | 2965.801246 | -0.289416207 | 0.084952103 | 0.000385706 | 0.002600961 |
| DDT | 939.489891 | 0.289219674 | 0.115230341 | 0.006782509 | 0.029689763 |
| ZBTB5 | 685.4637254 | -0.289202201 | 0.098950331 | 0.002000161 | 0.010723465 |
| RPL17-C18orf32 | 23347.68702 | -0.289116382 | 0.069542198 | 1.97E-05 | 0.000186803 |
| ZFPL1 | 268.9934088 | 0.289081202 | 0.145123 | 0.024852215 | 0.084468264 |
| SMIM29 | 251.4597552 | 0.289077658 | 0.14850304 | 0.027506563 | 0.09140878 |
| AIMP1 | 1464.043132 | -0.289036601 | 0.093501278 | 0.001158016 | 0.006738361 |
| ACSS1 | 3949.262499 | -0.288983063 | 0.08976886 | 0.000753719 | 0.004673073 |
| NEU3 | 496.1255544 | -0.28896519 | 0.109835505 | 0.004817261 | 0.022394765 |
| NCAPD3 | 3669.844564 | 0.288831261 | 0.066724603 | 8.90E-06 | 9.06E-05 |
| TNFRSF6B | 5.444033738 | 0.288662612 | 0.504837412 | 0.022606393 | 0.07846761 |
| FAM20B | 908.3652013 | 0.288576045 | 0.088301235 | 0.000628386 | 0.003983073 |
| SRR | 252.6821558 | 0.288467496 | 0.141405752 | 0.022207027 | 0.077360508 |
| ARAP1 | 780.8615143 | 0.288418007 | 0.108254501 | 0.004382505 | 0.020622781 |
| BTBD7 | 906.0199358 | 0.288341045 | 0.103379156 | 0.003017145 | 0.015148708 |
| PSENEN | 1128.61163 | -0.288209487 | 0.101229272 | 0.002556548 | 0.01318789 |
| SSBP3 | 919.5157313 | 0.288204211 | 0.097457162 | 0.00179773 | 0.009817499 |
| RXRB | 668.980993 | 0.288198758 | 0.100941653 | 0.002466787 | 0.01282418 |
| GALNT15 | 5.039515183 | -0.288098964 | 0.515783191 | 0.016093451 | 0.05952377 |

| | | | | | |
|---|---|---|---|---|---|
| CD2BP2 | 1516.49044 | -0.288097213 | 0.095095061 | 0.001428054 | 0.008052628 |
| UBR5 | 2751.33295 | -0.288096579 | 0.08041559 | 0.000201115 | 0.001468944 |
| AVEN | 548.6617701 | -0.288083471 | 0.106889957 | 0.004025116 | 0.019201911 |
| TRIM8 | 1006.277365 | 0.288057769 | 0.105644121 | 0.003647455 | 0.017722364 |
| HLA-DPA1 | 6627.843516 | -0.288020621 | 0.065315665 | 6.24E-06 | 6.57E-05 |
| LEPROTL1 | 1150.323669 | -0.287786788 | 0.108759744 | 0.004644818 | 0.021697955 |
| CSK | 2094.2166 | 0.287662798 | 0.096483402 | 0.001657032 | 0.009158894 |
| RPL36AP37 | 5754.795227 | -0.287614819 | 0.071323575 | 3.31E-05 | 0.000298255 |
| IL6ST | 2409.798483 | 0.287597699 | 0.118046447 | 0.008250457 | 0.03474542 |
| ARHGAP5 | 804.2827569 | -0.287504268 | 0.140888433 | 0.02239066 | 0.077890532 |
| SETP14 | 1493.490777 | 0.287500367 | 0.090412596 | 0.000871075 | 0.005276115 |
| CHRNA6 | 3.83315783 | 0.287464538 | 0.540860098 | 0.003575052 | 0.017453734 |
| ASH2L | 1423.015604 | -0.287346449 | 0.080632709 | 0.000216756 | 0.001566596 |
| RABL6 | 1864.17961 | 0.287240404 | 0.094442732 | 0.001367008 | 0.00774623 |
| BUB1B | 1014.284612 | 0.28705549 | 0.10638263 | 0.003962253 | 0.018943769 |
| PFN1P1 | 396.3881288 | 0.28702572 | 0.129689472 | 0.014894476 | 0.056011285 |
| LRCH1 | 437.7180568 | 0.286947784 | 0.118931892 | 0.008855067 | 0.036843634 |
| SLC38A7 | 348.1408708 | -0.286898441 | 0.126590489 | 0.013082874 | 0.050638384 |
| C12orf57 | 2028.79918 | -0.286872193 | 0.102982581 | 0.003099399 | 0.015503264 |
| N4BP1 | 887.5979113 | -0.286476585 | 0.088737405 | 0.00073229 | 0.004558257 |
| LUC7L2 | 1787.60373 | -0.286438897 | 0.088602779 | 0.000719964 | 0.004487983 |
| GLB1 | 2098.397578 | -0.286434183 | 0.090314893 | 0.000912787 | 0.005499908 |
| CNPY3 | 1936.003451 | 0.286297552 | 0.081303647 | 0.000252789 | 0.001799975 |
| YBX3 | 3697.986131 | 0.28623681 | 0.073895193 | 6.37E-05 | 0.000534875 |
| TOP1 | 6171.74811 | -0.286096919 | 0.081756261 | 0.000261051 | 0.001844216 |
| SNHG15 | 975.3732903 | 0.286038495 | 0.11684459 | 0.008128532 | 0.034374065 |
| ARHGAP23 | 1078.808659 | -0.285906344 | 0.101859156 | 0.002903911 | 0.014682343 |
| NOB1 | 2320.079066 | -0.285889753 | 0.075920682 | 9.95E-05 | 0.000791915 |
| ATP8 | 7372.923453 | -0.285862825 | 0.130595098 | 0.015887183 | 0.05894972 |
| RGN | 6.107825075 | -0.285847207 | 0.4941831 | 0.028076663 | 0.092893308 |
| LOC646762 | 2231.268623 | 0.285758671 | 0.072875749 | 5.28E-05 | 0.000452139 |
| PRKAR2A | 1453.533131 | -0.285707667 | 0.080704922 | 0.000237807 | 0.001703806 |
| PPIP5K1 | 373.3605411 | -0.285652205 | 0.127377784 | 0.014034933 | 0.053500453 |
| SNAPC3 | 906.923673 | 0.285564236 | 0.124218117 | 0.012036675 | 0.047155073 |
| C1orf43 | 13017.07157 | -0.285492051 | 0.066492957 | 1.07E-05 | 0.000107535 |
| ACP2 | 723.7326689 | 0.285461034 | 0.103537072 | 0.003360618 | 0.016598924 |
| RPL22P2 | 398.2786167 | -0.285440178 | 0.143627881 | 0.025680424 | 0.086586428 |
| GID8 | 2237.737131 | -0.285392123 | 0.075261571 | 9.00E-05 | 0.00072373 |
| MARS1 | 7419.033775 | 0.285253328 | 0.069503045 | 2.43E-05 | 0.000225722 |
| MBTPS1 | 2924.481431 | -0.285178177 | 0.070137566 | 2.89E-05 | 0.000263966 |
| UBE2C | 2715.695816 | 0.285117694 | 0.07748622 | 0.000139141 | 0.001060524 |
| LMBR1 | 2318.937552 | -0.285104296 | 0.102304542 | 0.003054512 | 0.015318557 |
| SLC39A10 | 1010.887057 | -0.285059332 | 0.120889769 | 0.010572819 | 0.042658318 |
| AMD1 | 4793.186809 | -0.28505474 | 0.095384216 | 0.001646945 | 0.009117694 |
| NME1 | 8644.478716 | -0.285018671 | 0.074635964 | 8.08E-05 | 0.00065809 |
| TRMT112 | 2837.444529 | -0.284893258 | 0.074728254 | 8.31E-05 | 0.000674804 |
| SSH2 | 1463.826735 | 0.284734196 | 0.081646537 | 0.000287654 | 0.002008231 |
| SGO1 | 594.2103032 | 0.284726991 | 0.106443396 | 0.004289361 | 0.020261554 |
| OSGIN2 | 999.3631602 | -0.284725652 | 0.103205507 | 0.003362753 | 0.01660473 |
| ACAT2 | 4068.9261 | 0.284209341 | 0.068687849 | 2.08E-05 | 0.000196039 |

| | | | | | |
|---|---|---|---|---|---|
| ITPK1 | 1813.612831 | 0.284191708 | 0.0845013 | 0.000456216 | 0.003017686 |
| CUL4A | 3457.677824 | -0.284126374 | 0.073136424 | 6.12E-05 | 0.000515867 |
| RPS7P10 | 1220.672059 | -0.284123491 | 0.11398028 | 0.007361056 | 0.031732986 |
| VGF | 6.230296965 | -0.284058125 | 0.491079045 | 0.028021781 | 0.092782635 |
| TNFRSF10C | 4.740041479 | -0.283555511 | 0.502159566 | 0.019851043 | 0.07048606 |
| CLASP2 | 1517.211712 | -0.283495425 | 0.079985931 | 0.000235456 | 0.001689762 |
| ATXN3 | 427.3007105 | 0.283453646 | 0.133513115 | 0.018616941 | 0.066886007 |
| KLHL7 | 1200.691313 | -0.283229359 | 0.085125986 | 0.000520812 | 0.003382933 |
| CIAO2B | 1919.22059 | -0.283198099 | 0.09810769 | 0.002303942 | 0.012108458 |
| CKS2 | 3221.965056 | -0.283186297 | 0.080716249 | 0.000270443 | 0.001902991 |
| ATP11B | 3786.502902 | 0.283123793 | 0.112981308 | 0.006973027 | 0.030370197 |
| NFIA | 292.6935116 | 0.283115451 | 0.144953749 | 0.027704016 | 0.091923824 |
| DPYD | 332.9800743 | 0.283077909 | 0.121448738 | 0.011183026 | 0.04460075 |
| NUGGC | 238.4551681 | 0.283021738 | 0.13838877 | 0.022532441 | 0.078273677 |
| RPLP2 | 24797.15728 | -0.282959798 | 0.08665532 | 0.000639249 | 0.004046007 |
| RPS25P1 | 3017.823369 | -0.282902073 | 0.083864635 | 0.000425142 | 0.002832698 |
| SNORA105B | 677.5426944 | -0.282811656 | 0.146552777 | 0.029338794 | 0.096132432 |
| CENPC | 291.9973037 | -0.282730145 | 0.145480377 | 0.028355055 | 0.093617633 |
| PTPN4 | 505.7013889 | 0.2826539 | 0.136532215 | 0.021235022 | 0.074529162 |
| RPL35 | 33363.99859 | 0.282613503 | 0.066479289 | 1.27E-05 | 0.000126275 |
| STX4 | 289.215767 | -0.282575124 | 0.133283929 | 0.019110862 | 0.06832015 |
| SMIM38 | 4.832812739 | -0.282455682 | 0.515964548 | 0.011663776 | 0.046019961 |
| MTDH | 15785.05687 | -0.282331279 | 0.089934365 | 0.001012678 | 0.006009782 |
| PIGH | 523.8750947 | 0.282207182 | 0.122398817 | 0.012016486 | 0.047100383 |
| HSPE1P3 | 242.3526836 | -0.282042421 | 0.140697942 | 0.025121232 | 0.08518198 |
| DAD1 | 3810.01415 | -0.28175546 | 0.087788121 | 0.000816774 | 0.005001438 |
| PARN | 1139.650382 | -0.281691671 | 0.097765158 | 0.002344069 | 0.01228207 |
| MCTS1 | 1875.075499 | -0.281654146 | 0.075406491 | 0.000114033 | 0.000891116 |
| TBC1D22A | 772.0741285 | 0.281575126 | 0.09340716 | 0.001525468 | 0.00852421 |
| NFATC3 | 1915.023489 | -0.281268539 | 0.076331338 | 0.000138792 | 0.001058794 |
| PCBP2-OT1 | 574.8020153 | 0.281123968 | 0.10785815 | 0.005323297 | 0.024290784 |
| AP1G2 | 323.1638821 | 0.281048104 | 0.125220484 | 0.014067124 | 0.053585792 |
| JPT2 | 1970.891562 | -0.281032906 | 0.076497935 | 0.000144864 | 0.001099307 |
| FMNL2 | 205.7692097 | -0.280812009 | 0.146614398 | 0.030551724 | 0.099075695 |
| ZNF765-ZNF761 | 298.5497578 | 0.280706747 | 0.125018735 | 0.014073205 | 0.053597028 |
| PAN3 | 1026.563776 | 0.280399096 | 0.119292983 | 0.010691928 | 0.043038613 |
| CLOCK | 468.8987853 | 0.280314307 | 0.138386812 | 0.023688671 | 0.081279316 |
| ZMYND19 | 677.1355472 | 0.280281966 | 0.107252736 | 0.005236889 | 0.023997671 |
| ERBIN | 1918.945667 | 0.280079389 | 0.102750068 | 0.003753517 | 0.018105755 |
| ZNF148 | 1311.968404 | 0.280041024 | 0.138263777 | 0.024151512 | 0.082540306 |
| ZNF76 | 364.251353 | 0.279971482 | 0.125668529 | 0.014713452 | 0.055502175 |
| SSBP2 | 971.0470958 | 0.279958541 | 0.098198757 | 0.002575329 | 0.01327291 |
| PPP2R2A | 2154.20657 | -0.279919953 | 0.073157265 | 7.96E-05 | 0.000650229 |
| ZGRF1 | 271.3729438 | -0.279878244 | 0.144291592 | 0.029003995 | 0.095358532 |
| C15orf40 | 308.6350696 | 0.279815711 | 0.132102712 | 0.019255683 | 0.068795267 |
| CDON | 790.4486361 | 0.279782335 | 0.095638355 | 0.002033449 | 0.010878356 |
| DEF8 | 921.2546942 | -0.279768698 | 0.10929072 | 0.006171083 | 0.027464571 |
| EMC8 | 1328.067378 | -0.279704166 | 0.100382692 | 0.003175113 | 0.015854509 |
| RPL5P34 | 1658.862674 | -0.279523788 | 0.07707696 | 0.000175076 | 0.001298968 |
| ABHD3 | 771.8483169 | 0.279232277 | 0.100617726 | 0.003266318 | 0.016202634 |

| | | | | | |
|---|---|---|---|---|---|
| PIGN | 1375.590913 | -0.279013632 | 0.101704215 | 0.00360841 | 0.01757717 |
| DCBLD1 | 296.9628052 | 0.278995943 | 0.127331518 | 0.016211336 | 0.059849327 |
| GTF3C6 | 1223.773448 | -0.278963288 | 0.087048673 | 0.000823564 | 0.005035899 |
| RMC1 | 913.0061262 | 0.278776656 | 0.089568037 | 0.001107627 | 0.006493154 |
| RANBP10 | 516.9676521 | -0.278761725 | 0.116461312 | 0.0097326 | 0.039900434 |
| RPL7AP6 | 2434.428658 | 0.278625575 | 0.094052962 | 0.001822488 | 0.009921416 |
| AP5B1 | 668.9876402 | 0.278547362 | 0.123399213 | 0.013757673 | 0.052743329 |
| DCAF1 | 2575.66101 | -0.278493691 | 0.072045042 | 6.84E-05 | 0.000569228 |
| BAZ1A | 361.5582522 | -0.278478148 | 0.123284692 | 0.013682197 | 0.052523744 |
| NFYC | 1056.758959 | 0.278412848 | 0.098076132 | 0.002693264 | 0.013798597 |
| RNF125 | 797.6127597 | 0.278312696 | 0.108490291 | 0.006023711 | 0.026905496 |
| RAB24 | 576.834004 | 0.278274295 | 0.109865123 | 0.006636872 | 0.029155432 |
| GRK6 | 3990.582301 | 0.278211706 | 0.082375279 | 0.000442791 | 0.002937858 |
| TMED9 | 8597.373774 | -0.278173906 | 0.078960319 | 0.000261534 | 0.001845555 |
| CNST | 589.9897041 | 0.278152285 | 0.105164232 | 0.00472739 | 0.022036128 |
| TGIF2-RAB5IF | 1874.133459 | -0.278145474 | 0.084572999 | 0.000610667 | 0.003890179 |
| PDK2 | 600.9298233 | 0.278080729 | 0.105123914 | 0.004826058 | 0.022423623 |
| SMIM24 | 3.564987027 | 0.277946491 | 0.51964608 | 0.004415714 | 0.020750861 |
| HNRNPA1P15 | 272.5598463 | -0.277903371 | 0.13554121 | 0.0229513 | 0.079299214 |
| SYNRG | 1332.857308 | 0.277788624 | 0.083270946 | 0.000513041 | 0.003342487 |
| NR1H2 | 1384.960432 | 0.277448895 | 0.115871204 | 0.009686249 | 0.039738633 |
| GFOD2 | 335.1278923 | -0.277435973 | 0.135308862 | 0.022971309 | 0.07933669 |
| TNIP1 | 3408.315764 | 0.277315138 | 0.0758351 | 0.00015554 | 0.001170064 |
| H3P47 | 519.8463328 | 0.277270368 | 0.112742871 | 0.008164069 | 0.034464645 |
| PSMC4 | 4574.782961 | 0.277231241 | 0.076197234 | 0.000167603 | 0.001251569 |
| IRF3 | 1129.314763 | 0.277180694 | 0.091640353 | 0.0014995 | 0.008400129 |
| RETREG2 | 2537.262898 | 0.277101017 | 0.084830906 | 0.000658512 | 0.004148219 |
| ORC2 | 735.3035046 | 0.277059911 | 0.104045316 | 0.004575665 | 0.021401643 |
| PLOD3 | 1687.625065 | -0.276952058 | 0.092753277 | 0.001714395 | 0.009430053 |
| CAPZB | 3766.391931 | 0.276917526 | 0.076791923 | 0.000188468 | 0.001387037 |
| DDX3X | 13563.8503 | -0.276830599 | 0.085005115 | 0.000689423 | 0.004316246 |
| ERICH1 | 467.1235415 | -0.276706426 | 0.112915988 | 0.008408811 | 0.035297243 |
| ARMC5 | 486.5612218 | -0.276616703 | 0.140938304 | 0.028188933 | 0.093140192 |
| RERE | 443.8751557 | 0.276508505 | 0.135429376 | 0.023219508 | 0.079986516 |
| REG1A | 4.709463697 | 0.276488632 | 0.487480943 | 0.023552423 | 0.080956195 |
| HBG2 | 5.350408091 | -0.276463441 | 0.48393772 | 0.027674605 | 0.091861441 |
| MRPL28 | 1455.026177 | -0.276448421 | 0.08803072 | 0.001031583 | 0.006105237 |
| MAGEF1 | 442.7459543 | 0.276442815 | 0.113647945 | 0.008765784 | 0.036533594 |
| ZDHHC7 | 1255.967507 | -0.276368584 | 0.084946631 | 0.000698068 | 0.004364058 |
| GAPDH | 208245.2952 | -0.276284074 | 0.073057427 | 0.000153311 | 0.001155808 |
| SCD | 23897.86495 | -0.276217333 | 0.059831498 | 2.48E-06 | 2.86E-05 |
| PSME2 | 5580.558257 | 0.276028132 | 0.070753543 | 5.72E-05 | 0.000484944 |
| RPL10P16 | 9292.03326 | -0.276023643 | 0.093606001 | 0.001920596 | 0.010373975 |
| LOC112694756 | 20140.35259 | -0.276011234 | 0.079446373 | 0.000319838 | 0.002204328 |
| MON1B | 1447.411399 | -0.276004219 | 0.080204252 | 0.00035705 | 0.002426621 |
| CHIC2 | 243.974884 | 0.275937816 | 0.137477326 | 0.025355837 | 0.085809463 |
| PEX12 | 257.8933815 | -0.275937629 | 0.137862964 | 0.02567166 | 0.086586428 |
| RNVU1-3 | 5.961299345 | 0.275927003 | 0.481197684 | 0.026831326 | 0.089733116 |
| KLHL11 | 552.1705485 | 0.275844099 | 0.132692659 | 0.021447326 | 0.075137234 |
| HTRA3 | 5.065849926 | 0.275620978 | 0.480586115 | 0.028074838 | 0.092893308 |

| | | | | | |
|---|---|---|---|---|---|
| RPL10P9 | 3770.556286 | -0.275352046 | 0.100276629 | 0.003638077 | 0.01769182 |
| GK | 293.9915357 | 0.275272673 | 0.133950313 | 0.022667311 | 0.078616026 |
| ELMO2 | 800.6233638 | 0.275163303 | 0.093964989 | 0.002053225 | 0.010963829 |
| SYS1 | 1191.808988 | 0.27513487 | 0.103963429 | 0.00485063 | 0.022519666 |
| PIK3CB | 749.8476279 | 0.274960285 | 0.103481351 | 0.004676824 | 0.021829801 |
| RPS9P2 | 967.6564244 | -0.274849372 | 0.108948181 | 0.006964598 | 0.030353419 |
| ALG11 | 830.9167885 | -0.274786823 | 0.091648238 | 0.001650067 | 0.009132057 |
| COMMD6 | 1461.68438 | -0.274769131 | 0.087473479 | 0.001034608 | 0.006116866 |
| HSPH1 | 5892.186716 | -0.274730628 | 0.088020801 | 0.001108422 | 0.006495618 |
| LOC729998 | 22432.4099 | -0.274730226 | 0.06223964 | 6.38E-06 | 6.69E-05 |
| PRCC | 3192.70803 | -0.274583755 | 0.087466483 | 0.001039055 | 0.006136873 |
| DAZAP2 | 6351.282452 | 0.27452953 | 0.071310915 | 7.31E-05 | 0.000603799 |
| RPN2 | 27041.94306 | -0.274486545 | 0.069456537 | 4.86E-05 | 0.000418057 |
| POLR2K | 1542.555908 | -0.274454768 | 0.09979672 | 0.00359241 | 0.017509082 |
| INTS3 | 1575.436164 | -0.274454721 | 0.093819602 | 0.002092527 | 0.011149627 |
| ADD1 | 3198.733284 | 0.27442854 | 0.068365205 | 3.71E-05 | 0.00032895 |
| ACOT1 | 528.0574176 | 0.274398856 | 0.105719435 | 0.005636308 | 0.025457473 |
| RN7SL3 | 419.7070904 | 0.274347192 | 0.545022315 | 6.20E-06 | 6.53E-05 |
| GALNT7 | 805.1263671 | -0.27430744 | 0.117482669 | 0.011477917 | 0.045546078 |
| TRAF6 | 635.9146473 | -0.274229051 | 0.096938953 | 0.002832391 | 0.014409081 |
| BTG3 | 665.0566759 | 0.274144608 | 0.111444944 | 0.00822543 | 0.034660421 |
| FAM234A | 286.236454 | -0.274049618 | 0.127356575 | 0.018320783 | 0.066068511 |
| SLC50A1 | 1707.365877 | -0.274017252 | 0.086787888 | 0.000981898 | 0.005853195 |
| ADNP2 | 2044.878367 | -0.273997283 | 0.085771677 | 0.000863033 | 0.005232891 |
| FRG1CP | 398.7742631 | 0.273988378 | 0.140099416 | 0.028576 | 0.094221362 |
| RNVU1-2A | 4.688317128 | 0.273929829 | 0.502337853 | 0.010959454 | 0.043870751 |
| UBXN6 | 1590.947527 | 0.273823869 | 0.092463157 | 0.001883392 | 0.010220842 |
| GBP7 | 3.310683266 | 0.273605269 | 0.520434487 | 0.000781204 | 0.004817701 |
| RPS9 | 29711.25847 | -0.273577194 | 0.078420769 | 0.000298137 | 0.002073427 |
| MTFP1 | 1052.831099 | 0.273543361 | 0.109769346 | 0.00754493 | 0.032356503 |
| QSOX2 | 1286.902123 | 0.273383899 | 0.09934572 | 0.00356865 | 0.01743253 |
| AFAP1 | 3.450028265 | 0.273337936 | 0.510373892 | 0.004899037 | 0.022701799 |
| UBL3 | 1370.556457 | 0.27325818 | 0.134219294 | 0.023938458 | 0.081941544 |
| CYTH2 | 1549.094726 | 0.273196047 | 0.088384421 | 0.001221386 | 0.007033838 |
| EIF2AK4 | 987.6636343 | -0.272968101 | 0.089278546 | 0.001374426 | 0.007785716 |
| RPL13AP20 | 2602.895623 | -0.272722946 | 0.073585756 | 0.000132013 | 0.001013725 |
| RAB5A | 1290.988328 | -0.272721846 | 0.115055062 | 0.01056382 | 0.042631946 |
| PREPL | 2712.812633 | -0.27271753 | 0.095683219 | 0.002615141 | 0.013434113 |
| GPD1L | 1073.355826 | -0.272688127 | 0.098677648 | 0.003476441 | 0.017054259 |
| USP4 | 802.9339765 | -0.272615801 | 0.098551891 | 0.003458493 | 0.016985457 |
| OST4 | 5273.050193 | -0.272564812 | 0.080876398 | 0.000468782 | 0.003085479 |
| SLC2A11 | 234.8962443 | -0.272203507 | 0.139911183 | 0.029709768 | 0.097017548 |
| CMC1 | 545.7451335 | -0.272190118 | 0.108782272 | 0.007398471 | 0.03187841 |
| SPAG7 | 1368.994015 | 0.272109639 | 0.084473561 | 0.000783378 | 0.004829388 |
| HAX1 | 7552.519127 | -0.272038731 | 0.069316924 | 5.60E-05 | 0.000476935 |
| C1orf21 | 619.4777335 | -0.271724751 | 0.102243379 | 0.00478396 | 0.022257882 |
| C17orf49 | 1134.874877 | 0.271614 | 0.100866653 | 0.004296357 | 0.020283536 |
| TNKS | 992.8578581 | -0.271447181 | 0.091811813 | 0.00191205 | 0.010340713 |
| ALDOA | 20372.03832 | -0.271401486 | 0.079420165 | 0.000400577 | 0.002686596 |
| SYVN1 | 1496.938855 | -0.271331092 | 0.102298786 | 0.004869547 | 0.022595378 |

| | | | | | |
|---|---|---|---|---|---|
| ANKLE1 | 462.7026208 | 0.271299911 | 0.116044611 | 0.011516621 | 0.045657811 |
| NSMCE1 | 779.1636711 | -0.271220131 | 0.098380191 | 0.003596514 | 0.017524153 |
| STX8 | 824.3465738 | 0.271123654 | 0.094273826 | 0.002462012 | 0.012810895 |
| DAP3 | 8795.023775 | -0.271060344 | 0.061608324 | 6.96E-06 | 7.25E-05 |
| MCEE | 326.7195293 | -0.270753835 | 0.134146093 | 0.025475291 | 0.08612956 |
| LSM14B | 547.8669508 | 0.270649415 | 0.10434969 | 0.005724867 | 0.025750006 |
| SMPD2 | 367.9639865 | 0.270537635 | 0.129321552 | 0.021291306 | 0.074696424 |
| LOC105371632 | 3.50674429 | -0.270506033 | 0.503779525 | 0.005882745 | 0.026343804 |
| PIKFYVE | 932.002617 | 0.270350324 | 0.114490137 | 0.010812323 | 0.043382065 |
| FKBP1A | 4585.164618 | -0.270077512 | 0.067317595 | 3.84E-05 | 0.000339644 |
| SELENOI | 3353.367034 | -0.270029721 | 0.093970622 | 0.002554478 | 0.013181134 |
| GRPEL2 | 1256.476874 | -0.270024542 | 0.136111581 | 0.027482832 | 0.091347446 |
| NIP7 | 1527.904022 | -0.269907083 | 0.102802356 | 0.005271793 | 0.02411994 |
| PIGP | 413.419919 | -0.2698405 | 0.116320622 | 0.012150293 | 0.047522701 |
| GNPDA1 | 1476.38114 | -0.269787127 | 0.080681648 | 0.00051938 | 0.003374896 |
| AP1S1 | 596.2177666 | -0.269748734 | 0.100054173 | 0.004316077 | 0.020343359 |
| NUDCD2 | 694.1909615 | 0.269708777 | 0.102753264 | 0.00525598 | 0.024073722 |
| PSMD2 | 7132.689008 | 0.269609035 | 0.07000077 | 7.38E-05 | 0.000608491 |
| SUN2 | 3184.35895 | 0.269602349 | 0.091370349 | 0.001954243 | 0.010532719 |
| MYG1 | 1607.291612 | 0.269554338 | 0.096885284 | 0.003299226 | 0.016342306 |
| TMEM120B | 327.3933836 | -0.269380085 | 0.122735446 | 0.016665527 | 0.061117521 |
| ZNF510 | 407.324815 | 0.269259076 | 0.121543598 | 0.015754192 | 0.058556629 |
| BIRC3 | 7249.953656 | -0.269227207 | 0.114236745 | 0.011292717 | 0.044969201 |
| NOTCH1 | 887.937418 | 0.269215511 | 0.117441806 | 0.013012393 | 0.050386022 |
| PACS2 | 420.1683965 | 0.269191285 | 0.129836359 | 0.022307202 | 0.077667384 |
| VCL | 1075.294302 | 0.269171341 | 0.086668515 | 0.001177869 | 0.006830932 |
| ALDH5A1 | 504.2611246 | -0.269111129 | 0.108331197 | 0.007873895 | 0.033491723 |
| TRIM16 | 550.4597856 | 0.269093053 | 0.106999911 | 0.007213149 | 0.03126661 |
| KDM2A | 2961.115653 | 0.268984301 | 0.07775391 | 0.000339774 | 0.002324717 |
| MAP3K5 | 547.4877028 | 0.268770678 | 0.108282989 | 0.007877334 | 0.033491723 |
| CARM1 | 1730.874554 | -0.26874404 | 0.090525013 | 0.001899656 | 0.010276895 |
| CDK5RAP1 | 1209.537517 | -0.268738784 | 0.096309218 | 0.003264871 | 0.016202634 |
| NPAT | 1091.812697 | 0.268629127 | 0.122141158 | 0.016490525 | 0.060629813 |
| SPHK2 | 412.6102161 | 0.268564034 | 0.13175321 | 0.024287031 | 0.0829164 |
| RPUSD4 | 1394.676174 | -0.268507575 | 0.084987277 | 0.000992642 | 0.005909105 |
| EXOSC6 | 730.3993197 | -0.268397605 | 0.106537062 | 0.007172804 | 0.03111509 |
| RPL27A | 39609.0962 | -0.268232178 | 0.063122047 | 1.41E-05 | 0.000138643 |
| DDX19B | 1291.103056 | -0.268176384 | 0.089484387 | 0.001714803 | 0.009430053 |
| P3H1 | 726.8530633 | -0.268072575 | 0.104761673 | 0.006470215 | 0.028548422 |
| UBP1 | 2673.563262 | -0.268025479 | 0.077926001 | 0.000368769 | 0.002498426 |
| RPS8 | 73072.73008 | -0.268008288 | 0.066845379 | 4.20E-05 | 0.000367739 |
| CTBP1 | 1870.824133 | 0.26800635 | 0.090635721 | 0.001923778 | 0.010387921 |
| RINT1 | 1098.440908 | -0.267976899 | 0.092508225 | 0.002348813 | 0.012303205 |
| RPS26P7 | 546.431145 | -0.267950321 | 0.108122749 | 0.008002893 | 0.033917301 |
| RPLP0 | 91269.6176 | -0.267900459 | 0.080695155 | 0.000555663 | 0.003575777 |
| DNAJC2 | 854.6058602 | -0.267795528 | 0.105654019 | 0.006881699 | 0.030055583 |
| SLAIN2 | 3905.746187 | 0.267766272 | 0.086782816 | 0.001267935 | 0.007251262 |
| TNIP2 | 333.1909341 | 0.267741172 | 0.127627452 | 0.021067728 | 0.074062087 |
| NDUFA4 | 8722.341988 | -0.267656234 | 0.085007477 | 0.00103725 | 0.006128303 |
| POMP | 4655.790919 | -0.267609267 | 0.073551498 | 0.000176451 | 0.001307488 |

| | | | | | |
|---|---|---|---|---|---|
| NCR3LG1 | 1731.271126 | -0.267468988 | 0.075953418 | 0.000273007 | 0.001917913 |
| ZC3H18 | 1071.475634 | -0.267395134 | 0.101910575 | 0.005356276 | 0.024421968 |
| EPB41 | 3103.61173 | 0.267338923 | 0.081685086 | 0.000660399 | 0.004158591 |
| CHD6 | 833.3484974 | 0.267283705 | 0.098886209 | 0.004207135 | 0.019949332 |
| ZNF346 | 283.796926 | 0.267119457 | 0.132941449 | 0.026129518 | 0.087777191 |
| GTF2E2 | 1173.366799 | -0.267117024 | 0.083587011 | 0.000878465 | 0.005313441 |
| MRPL16 | 1957.323043 | -0.267088208 | 0.07602544 | 0.000280061 | 0.001960312 |
| PRKAG1 | 1922.82629 | 0.267066431 | 0.088667418 | 0.001616998 | 0.00897486 |
| ATF4 | 13189.667 | -0.266818131 | 0.068946602 | 7.02E-05 | 0.000581465 |
| YIF1A | 1073.558619 | -0.266782478 | 0.122293212 | 0.017556493 | 0.063965031 |
| TBC1D1 | 590.8232984 | 0.266745304 | 0.098837272 | 0.004227404 | 0.020023509 |
| SH3BP2 | 694.9496175 | -0.266683866 | 0.100433375 | 0.004919044 | 0.022776227 |
| HPS5 | 440.3353371 | 0.266512744 | 0.126067195 | 0.020446642 | 0.072171728 |
| ACOT8 | 604.242669 | 0.266506899 | 0.105299733 | 0.006967422 | 0.030353419 |
| CBX2 | 633.8036866 | 0.266482561 | 0.122334119 | 0.017608211 | 0.064073559 |
| GFPT1 | 4706.199228 | -0.266464997 | 0.101036527 | 0.005218356 | 0.023924628 |
| NOP53 | 8534.34535 | -0.266423213 | 0.089701874 | 0.001898358 | 0.010273084 |
| NIPSNAP3A | 412.6456265 | -0.266404803 | 0.134659674 | 0.02817562 | 0.093117277 |
| NOP14 | 797.3471983 | -0.266358063 | 0.094685768 | 0.003051128 | 0.01530602 |
| RNF5 | 1343.423056 | 0.266341623 | 0.095450021 | 0.003266351 | 0.016202634 |
| CCNQ | 548.0474619 | 0.266266652 | 0.12648624 | 0.021015325 | 0.073892871 |
| LINC02246 | 3.23318912 | 0.266255169 | 0.506321053 | 0.000861353 | 0.005224537 |
| USP14 | 4927.553726 | -0.266221671 | 0.077492087 | 0.000379093 | 0.002561355 |
| MBD3 | 1770.69965 | 0.266126347 | 0.111052491 | 0.010062247 | 0.041009161 |
| SAP30L | 612.3319564 | 0.266120964 | 0.101643077 | 0.005419354 | 0.024664112 |
| DAGLB | 343.7262112 | 0.265911511 | 0.129319032 | 0.023571492 | 0.08098959 |
| ARHGAP1 | 751.0749643 | 0.265769797 | 0.112616922 | 0.011103247 | 0.044356194 |
| H3C13 | 47.57015424 | 0.26571145 | 0.513107527 | 0.000785103 | 0.00483658 |
| USP39 | 4065.604676 | 0.26558725 | 0.067931903 | 5.84E-05 | 0.000493937 |
| RPL36A | 9580.755347 | -0.265539093 | 0.06779389 | 5.79E-05 | 0.000490415 |
| USP28 | 1061.964981 | 0.26542651 | 0.091160431 | 0.002243811 | 0.011839146 |
| GPATCH4 | 1484.232204 | -0.265375 | 0.098988076 | 0.004570751 | 0.021398134 |
| VPS33A | 1259.941986 | -0.26534874 | 0.082553839 | 0.000829473 | 0.005064875 |
| POLR3A | 734.5529196 | -0.265343943 | 0.103343125 | 0.006319388 | 0.027995098 |
| ARHGAP30 | 6948.270292 | -0.265252481 | 0.070788538 | 0.000115048 | 0.000898237 |
| RHOH | 1128.522917 | 0.265227968 | 0.082323892 | 0.000804603 | 0.004933893 |
| LARP7 | 758.7873699 | -0.264852027 | 0.108508542 | 0.008994636 | 0.037307811 |
| BDH1 | 735.6759066 | 0.264774524 | 0.099212203 | 0.004755392 | 0.022142814 |
| TMA16 | 799.9087035 | 0.264686458 | 0.131516397 | 0.026108302 | 0.087755721 |
| RIPK1 | 737.2831741 | 0.264670954 | 0.093117724 | 0.002764448 | 0.014104875 |
| ZKSCAN1 | 1467.146325 | -0.264659128 | 0.080708529 | 0.000662765 | 0.004168946 |
| SIDT2 | 736.4259088 | -0.264654838 | 0.112919965 | 0.011743273 | 0.046228293 |
| YBEY | 534.9887152 | -0.264630923 | 0.107751413 | 0.008643589 | 0.03608697 |
| TICRR | 578.8622188 | 0.264574295 | 0.10585922 | 0.007635922 | 0.032690053 |
| DCTN6 | 616.1565578 | -0.264570744 | 0.116378375 | 0.013973653 | 0.053327588 |
| RPA3 | 2081.226455 | -0.264464464 | 0.089445899 | 0.001864945 | 0.010127078 |
| ZNF8-ERVK3-1 | 464.8180833 | 0.264412423 | 0.109051865 | 0.009371791 | 0.038629336 |
| PAQR4 | 383.1616807 | -0.264367191 | 0.119451235 | 0.016341408 | 0.060209397 |
| CARD11 | 1046.721447 | 0.264168176 | 0.10275397 | 0.006260853 | 0.027778426 |
| FCF1 | 923.9348935 | 0.264092773 | 0.100483306 | 0.005301565 | 0.024204375 |

| | | | | | |
|---|---|---|---|---|---|
| ATAD3B | 812.4431337 | -0.264074075 | 0.1108342 | 0.010612555 | 0.042778773 |
| H4C3 | 63.44924453 | 0.264072219 | 0.512977161 | 0.000417516 | 0.002788325 |
| P4HA2 | 534.3287261 | 0.263847138 | 0.112556441 | 0.011661615 | 0.046019961 |
| INPP5B | 396.4009756 | 0.263781016 | 0.115993655 | 0.013960001 | 0.053318335 |
| POMT1 | 589.5726474 | 0.263456198 | 0.115104961 | 0.013455295 | 0.051813507 |
| SNORD45B | 4.587180814 | -0.263403937 | 0.478736427 | 0.015697908 | 0.058397567 |
| TM9SF3 | 6341.371692 | -0.263316513 | 0.118940862 | 0.016285022 | 0.060065542 |
| SRSF11 | 2286.512227 | -0.263229318 | 0.116715811 | 0.014742626 | 0.055559969 |
| HARS2 | 867.1277353 | -0.262848317 | 0.092708964 | 0.002905937 | 0.014688299 |
| MSTO1 | 912.4102957 | -0.26273611 | 0.105456683 | 0.007971809 | 0.033810387 |
| NCBP3 | 1640.124426 | 0.2625253 | 0.081953639 | 0.000859629 | 0.005215905 |
| HLA-DRB5 | 2561.214982 | -0.262451811 | 0.087028577 | 0.001637401 | 0.009079368 |
| RELB | 1003.245883 | 0.262396252 | 0.126720431 | 0.023112258 | 0.079728083 |
| ATAD1 | 2240.075406 | -0.262224138 | 0.107564658 | 0.009168777 | 0.037899473 |
| KBTBD8 | 1138.282602 | -0.262196419 | 0.131294641 | 0.027512023 | 0.091409381 |
| RNF144A | 1064.362305 | -0.262108963 | 0.098531335 | 0.004904419 | 0.022714584 |
| BRAF | 1019.045987 | -0.262070479 | 0.111742648 | 0.011716356 | 0.04614331 |
| MKS1 | 344.4067758 | -0.261831243 | 0.126690559 | 0.023474008 | 0.080718703 |
| MRPL34 | 2119.613422 | 0.261719084 | 0.100888933 | 0.005897187 | 0.026401652 |
| PHAF1 | 620.1691722 | -0.261645268 | 0.108117845 | 0.00967958 | 0.039720684 |
| XRN1 | 996.7014012 | 0.261614737 | 0.125152065 | 0.022009332 | 0.076795405 |
| RPS25P2 | 4128.684035 | -0.261607674 | 0.082877398 | 0.001029424 | 0.006094539 |
| XPO7 | 2011.973963 | -0.261580477 | 0.072137309 | 0.000189083 | 0.001390492 |
| XRCC1 | 523.6785211 | 0.261470325 | 0.109274312 | 0.010311609 | 0.041848168 |
| CD37 | 740.3953979 | 0.261172799 | 0.106820496 | 0.009002207 | 0.037330275 |
| RUSF1 | 1275.230942 | -0.261161164 | 0.089250277 | 0.002213064 | 0.011691161 |
| TSPAN14 | 896.8136121 | 0.261115505 | 0.112821741 | 0.012727769 | 0.04938534 |
| CYB5D2 | 326.5678274 | 0.261029901 | 0.127021464 | 0.024021702 | 0.08217776 |
| RBM19 | 398.080413 | -0.261029612 | 0.115051839 | 0.014414726 | 0.054651753 |
| TBC1D7 | 431.0418526 | -0.261007771 | 0.109856223 | 0.010919504 | 0.043731047 |
| WBP2 | 1489.44183 | 0.260835779 | 0.093813162 | 0.003432412 | 0.016881307 |
| NCKAP1L | 1523.537298 | -0.260735682 | 0.078119951 | 0.000546071 | 0.003515357 |
| RPL5P4 | 1219.183515 | -0.260590162 | 0.081232118 | 0.00086372 | 0.005235223 |
| DLD | 3811.102111 | -0.260465798 | 0.097882003 | 0.004942043 | 0.022858267 |
| ACER3 | 500.4984884 | 0.260451191 | 0.119695527 | 0.018052087 | 0.065371808 |
| UBE2S | 3521.993642 | 0.260409103 | 0.109869497 | 0.011005934 | 0.044006413 |
| WASHC3 | 580.7529803 | 0.260407411 | 0.105530723 | 0.008478035 | 0.03553101 |
| ROBO3 | 4.440191396 | 0.260356819 | 0.470514342 | 0.019932567 | 0.070727246 |
| POC5 | 666.2509682 | 0.26030942 | 0.099817286 | 0.005702679 | 0.025670207 |
| UBE2E3 | 1023.081607 | 0.260214374 | 0.094294905 | 0.003662222 | 0.017764342 |
| MRPL51 | 4180.261614 | -0.260163568 | 0.069811641 | 0.000128548 | 0.000990685 |
| CEP78 | 554.6564629 | 0.260121264 | 0.103698756 | 0.007558547 | 0.032398852 |
| RPL32P18 | 1519.882362 | -0.260082726 | 0.089850031 | 0.002446967 | 0.012740856 |
| FARSB | 5968.623597 | -0.259996227 | 0.0655622 | 4.80E-05 | 0.000414315 |
| MYOF | 3.381956076 | 0.259920463 | 0.48725347 | 0.005430387 | 0.024697741 |
| EFR3A | 1208.822835 | 0.259847726 | 0.106670664 | 0.009265534 | 0.038239097 |
| RPS26P25 | 1629.843285 | -0.259837924 | 0.079033445 | 0.000655129 | 0.004132918 |
| NCL | 38598.4019 | -0.259824178 | 0.08550646 | 0.001532374 | 0.008551763 |
| RPS17 | 58553.41158 | -0.259768287 | 0.067210406 | 8.11E-05 | 0.000659717 |
| UTP3 | 1113.335211 | -0.259687248 | 0.10095412 | 0.006365605 | 0.028185421 |

| | | | | | |
|---|---|---|---|---|---|
| RNF44 | 839.8307033 | 0.259612199 | 0.114600975 | 0.014498758 | 0.054867788 |
| OTUD4 | 1282.414248 | -0.259502684 | 0.116699629 | 0.016180415 | 0.059781576 |
| PRDM2 | 1567.823195 | -0.259331822 | 0.089950893 | 0.002552082 | 0.013172698 |
| OIP5 | 319.5281651 | 0.259184686 | 0.12975546 | 0.027701187 | 0.091923824 |
| DDX10 | 1939.703978 | -0.259135725 | 0.090126434 | 0.002630355 | 0.013492973 |
| ACTB | 101862.7806 | 0.2589677 | 0.060905751 | 1.39E-05 | 0.000136207 |
| SACM1L | 1431.21335 | -0.25894056 | 0.087353304 | 0.001950079 | 0.010513547 |
| MLXIP | 652.3598351 | 0.25884638 | 0.119663031 | 0.018771578 | 0.067371485 |
| RPL12P6 | 818.959503 | -0.258814985 | 0.099212149 | 0.005797543 | 0.026029572 |
| RNF115 | 3254.779129 | 0.258622257 | 0.076652468 | 0.000476745 | 0.003129108 |
| SAMD12 | 386.4339583 | 0.258481494 | 0.127181825 | 0.025607665 | 0.086442093 |
| EBPL | 2102.486658 | -0.258449188 | 0.080012546 | 0.000804229 | 0.004933346 |
| WDR48 | 1806.841184 | -0.258435746 | 0.086666185 | 0.001844617 | 0.010029274 |
| RFTN1 | 578.8781563 | -0.25842843 | 0.099252379 | 0.005864842 | 0.026283453 |
| PRMT6 | 841.7395765 | 0.258425425 | 0.093708744 | 0.00369899 | 0.017897299 |
| VAC14 | 1004.781862 | -0.258396451 | 0.100823287 | 0.006599384 | 0.029005464 |
| FBXO45 | 1604.060022 | 0.258050665 | 0.114924485 | 0.015301468 | 0.057217777 |
| NUMA1 | 874.9118975 | 0.258031916 | 0.119080466 | 0.018615698 | 0.066886007 |
| CNIH1 | 3584.826876 | -0.257978082 | 0.102851502 | 0.007695218 | 0.032911358 |
| CHKA | 1196.828747 | -0.257916013 | 0.088228281 | 0.002259448 | 0.011910764 |
| BEND5 | 3.876516145 | 0.257831913 | 0.478726107 | 0.011587662 | 0.045824055 |
| COQ9 | 911.7280623 | -0.257829704 | 0.095133344 | 0.004309153 | 0.020332861 |
| NOL12 | 448.1372341 | 0.257749569 | 0.112465884 | 0.013635042 | 0.052359016 |
| SNAPC5 | 720.0711222 | 0.257748633 | 0.100308612 | 0.006397451 | 0.028273287 |
| TAF1D | 1254.50068 | -0.257733238 | 0.095968801 | 0.00461728 | 0.021575133 |
| ETFA | 5021.166413 | -0.257627244 | 0.078658964 | 0.000689151 | 0.004316102 |
| SNORA22 | 5.830595568 | 0.257495105 | 0.482171616 | 0.007147217 | 0.031019639 |
| TAPT1 | 525.3646039 | 0.257325286 | 0.117969591 | 0.017920359 | 0.06500353 |
| GOT1 | 4359.143135 | 0.257320575 | 0.068740907 | 0.00011709 | 0.000912124 |
| EXOSC3 | 588.156574 | 0.257279666 | 0.101805704 | 0.00727752 | 0.031459015 |
| NEDD1 | 682.7391753 | 0.25724771 | 0.127604672 | 0.026636018 | 0.089217781 |
| SIAH1 | 425.5392206 | -0.257174949 | 0.124472286 | 0.023856567 | 0.081730313 |
| ACSF2 | 503.3388174 | -0.2569972 | 0.115632005 | 0.016440794 | 0.060498348 |
| RPS25 | 43142.83597 | -0.256996515 | 0.0698563 | 0.00015475 | 0.001165133 |
| CIZ1 | 674.7450363 | 0.2569142 | 0.112495347 | 0.01393581 | 0.053285187 |
| MIA2 | 644.1497819 | 0.256843785 | 0.102110296 | 0.007514107 | 0.032256268 |
| HSPE1P2 | 839.8958549 | -0.256789334 | 0.11830918 | 0.018616998 | 0.066886007 |
| ZCCHC9 | 629.4354788 | -0.256713299 | 0.113774263 | 0.015021445 | 0.05636541 |
| PPP4R3A | 3085.021354 | -0.256393939 | 0.082057727 | 0.001152266 | 0.006713034 |
| MYO1C | 1221.490718 | 0.256354387 | 0.091520203 | 0.003274315 | 0.016232843 |
| HLA-DOA | 797.1639888 | -0.256127533 | 0.092191967 | 0.003529731 | 0.01726189 |
| MINPP1 | 928.0820142 | 0.256111346 | 0.102196127 | 0.00772877 | 0.033022235 |
| CBX5 | 4862.974373 | -0.2560646 | 0.105307348 | 0.009536687 | 0.039227271 |
| PELO | 1143.626384 | -0.256045049 | 0.082277263 | 0.001215644 | 0.007010095 |
| UQCRHL | 1971.122826 | -0.256042885 | 0.107431589 | 0.010740788 | 0.043195129 |
| ZC3H12C | 877.9401813 | 0.255989974 | 0.125689321 | 0.025563697 | 0.086349875 |
| GIT1 | 794.232044 | -0.255456027 | 0.097821813 | 0.005795991 | 0.026029349 |
| PRDX1 | 15941.97735 | -0.255426782 | 0.064673861 | 5.20E-05 | 0.000445645 |
| RPL26 | 52018.36366 | -0.255214585 | 0.067194192 | 9.61E-05 | 0.000767823 |
| ARHGAP19 | 800.6593404 | 0.255209166 | 0.100022854 | 0.006852672 | 0.029938199 |

| | | | | | |
|---|---|---|---|---|---|
| SUCLA2 | 1832.619815 | -0.255200842 | 0.108895667 | 0.012043123 | 0.047162091 |
| MMUT | 1153.104245 | -0.25513707 | 0.087955086 | 0.002424947 | 0.012645659 |
| SIAE | 1006.312063 | -0.255129942 | 0.084945981 | 0.001744505 | 0.009574656 |
| SYNC | 4.619406644 | 0.255010399 | 0.478966406 | 0.007961174 | 0.033773553 |
| CAPN1 | 2690.626314 | 0.254796564 | 0.091630384 | 0.003503022 | 0.017160348 |
| EVC2 | 3.79346854 | 0.254702045 | 0.47299295 | 0.012587121 | 0.048982288 |
| NAA20 | 2211.660536 | -0.254585041 | 0.08011049 | 0.00097433 | 0.005820098 |
| SDHAF2 | 907.1190921 | 0.254570309 | 0.088517907 | 0.002611156 | 0.013417624 |
| CDK11B | 453.0593526 | 0.2545614 | 0.129140156 | 0.029888597 | 0.097527952 |
| SPC25 | 1047.342543 | 0.254441262 | 0.105006428 | 0.009785905 | 0.04009998 |
| SNX4 | 1518.627211 | -0.254367429 | 0.11395601 | 0.01580547 | 0.058703211 |
| PCCB | 3569.583074 | -0.25433376 | 0.07683479 | 0.000618707 | 0.003934688 |
| TRAPPC6A | 1002.769139 | -0.254254555 | 0.10119193 | 0.007735132 | 0.033041265 |
| RPL35P1 | 995.6503962 | 0.254226512 | 0.093679576 | 0.004300301 | 0.020296625 |
| IFIH1 | 533.0895489 | 0.253871657 | 0.114273261 | 0.016514265 | 0.060687424 |
| TRMT112P6 | 328.2266898 | -0.253495084 | 0.126918633 | 0.028588791 | 0.094245591 |
| PLEKHA8 | 666.7608654 | -0.253466117 | 0.10545114 | 0.010402915 | 0.042139713 |
| PCNX1 | 870.8557692 | 0.253358208 | 0.093380408 | 0.004273622 | 0.020203357 |
| ADORA2A | 870.0935926 | -0.253343094 | 0.112572812 | 0.015581204 | 0.057988332 |
| NCOA1 | 738.9479569 | 0.253303245 | 0.095101762 | 0.004949466 | 0.022880381 |
| UTP14C | 653.0061331 | -0.253139005 | 0.102597662 | 0.008732173 | 0.036402273 |
| KDELR2 | 4568.584943 | -0.253058091 | 0.07925865 | 0.000932288 | 0.005597933 |
| TRIM38 | 325.1896494 | 0.253047397 | 0.122221165 | 0.023882181 | 0.081789562 |
| INPP5D | 1798.346327 | 0.252953604 | 0.08381434 | 0.001665792 | 0.009201443 |
| IL2RG | 16223.34435 | 0.25284475 | 0.068002755 | 0.000133098 | 0.001020303 |
| SETDB1 | 3141.07157 | 0.252786253 | 0.074074582 | 0.000423874 | 0.002825339 |
| ERFE | 3.832709407 | 0.252410712 | 0.469244085 | 0.01262167 | 0.049094666 |
| UCHL3 | 1294.818944 | -0.252391015 | 0.079914594 | 0.001053612 | 0.006214372 |
| MED21 | 1205.424722 | 0.252390292 | 0.117859131 | 0.020193794 | 0.071468615 |
| PLSCR1 | 422.2536812 | 0.252098494 | 0.111149661 | 0.014709937 | 0.055502175 |
| LEMD2 | 352.9892782 | 0.2520412 | 0.127663814 | 0.03008935 | 0.09797993 |
| ST3GAL5 | 1132.46648 | 0.251842185 | 0.08394764 | 0.001772897 | 0.009694115 |
| H2BU2P | 5.116101717 | 0.25181371 | 0.463112574 | 0.015508112 | 0.057814605 |
| ZFYVE27 | 533.152786 | 0.251544193 | 0.107513213 | 0.012355034 | 0.048235176 |
| NFE2L2 | 5180.791912 | -0.251527028 | 0.086034102 | 0.002278918 | 0.011991501 |
| MCCC2 | 4425.935808 | -0.251492866 | 0.066927706 | 0.000116175 | 0.000906629 |
| HNRNPUL1 | 7478.582979 | 0.251203807 | 0.071795803 | 0.000309029 | 0.002139728 |
| MICOS10 | 2412.66711 | -0.251011562 | 0.105820695 | 0.011323225 | 0.045056132 |
| KRT8P32 | 3.176249968 | -0.250988753 | 0.478440543 | 0.001246144 | 0.007140794 |
| PTEN | 2496.30593 | 0.250800853 | 0.117333739 | 0.020126011 | 0.071257849 |
| MYDGF | 6382.66647 | -0.250650275 | 0.078640346 | 0.000964575 | 0.005765806 |
| SLC36A1 | 299.0831717 | 0.250646469 | 0.125257822 | 0.028403716 | 0.093760418 |
| ZNF480 | 771.3872313 | 0.250625531 | 0.10602243 | 0.011557588 | 0.045746914 |
| GLG1 | 5127.298323 | -0.250598138 | 0.063615287 | 5.66E-05 | 0.000480141 |
| UBR4 | 3495.706345 | 0.250445032 | 0.089006109 | 0.003204643 | 0.015974326 |
| RNF38 | 775.3760556 | 0.250340376 | 0.105880107 | 0.011581906 | 0.045811756 |
| TERF1 | 791.9125993 | -0.250315343 | 0.101081256 | 0.008632842 | 0.036057627 |
| RIOK1 | 711.6352141 | 0.250314614 | 0.092852484 | 0.004572798 | 0.021401643 |
| BRMS1 | 1250.392377 | 0.250159415 | 0.094267398 | 0.005199516 | 0.02385798 |
| GAS7 | 1453.840819 | 0.250099621 | 0.089072891 | 0.00325972 | 0.016188289 |

| | | | | | |
|---|---|---|---|---|---|
| TBL1X | 1693.50421 | 0.24984629 | 0.077293957 | 0.000814193 | 0.004990933 |
| MLEC | 8737.917848 | -0.2498133 | 0.066746478 | 0.000117223 | 0.000912749 |
| MAGT1 | 2096.918274 | -0.249757518 | 0.103694519 | 0.010356644 | 0.042001405 |
| KANSL2 | 1700.915956 | 0.249510495 | 0.07911093 | 0.001068423 | 0.006291019 |
| TUBBP2 | 1828.891582 | 0.249477547 | 0.075718619 | 0.00065651 | 0.004140126 |
| CLASP1 | 995.9794846 | 0.249475877 | 0.084444211 | 0.002069926 | 0.011039782 |
| WDR6 | 2386.772921 | -0.24947352 | 0.086885611 | 0.002709366 | 0.013856492 |
| CNOT10 | 1017.241151 | -0.249314139 | 0.090944109 | 0.004045909 | 0.01927986 |
| PRIM1 | 1054.655472 | 0.249262553 | 0.102246224 | 0.009556732 | 0.039291056 |
| MEPCE | 2804.00096 | -0.249173538 | 0.085487225 | 0.002375001 | 0.012421591 |
| MICB | 724.4861173 | 0.249013612 | 0.095328131 | 0.005866232 | 0.026283453 |
| KLHDC4 | 1076.448333 | -0.249004634 | 0.088549806 | 0.003263666 | 0.016202634 |
| HLA-DQA1 | 3311.554407 | -0.248974039 | 0.074005748 | 0.000519364 | 0.003374896 |
| ELL2P1 | 405.2146738 | -0.2488948 | 0.125968007 | 0.030377552 | 0.098584721 |
| AKR1B1 | 6794.109538 | -0.248662557 | 0.095148184 | 0.005912359 | 0.026450126 |
| TTC5 | 588.5132244 | 0.248592721 | 0.103249896 | 0.01045089 | 0.042294476 |
| ANAPC1P2 | 512.0261082 | 0.248478633 | 0.115794138 | 0.020387475 | 0.071977553 |
| NACC1 | 1905.264347 | 0.248413205 | 0.092544807 | 0.004780535 | 0.022247923 |
| GSR | 1941.438649 | -0.248309153 | 0.074673786 | 0.000595487 | 0.003805187 |
| CAMLG | 776.0197394 | -0.24829892 | 0.102474634 | 0.010055687 | 0.041001725 |
| ZNF544 | 1420.428984 | 0.248281725 | 0.08005203 | 0.00128583 | 0.007343903 |
| PLOD1 | 3084.096521 | -0.248207006 | 0.083693062 | 0.002020228 | 0.010820996 |
| TXNDC16 | 657.1683021 | -0.248174146 | 0.110999935 | 0.016382083 | 0.060320761 |
| ELOA | 1619.112891 | 0.248134064 | 0.087252223 | 0.00294546 | 0.014846526 |
| KRI1 | 388.438682 | 0.248082929 | 0.119596914 | 0.024157394 | 0.082544119 |
| SLC39A7 | 2877.102868 | -0.247974377 | 0.07299501 | 0.000459635 | 0.003034503 |
| INSR | 1376.311758 | -0.247845775 | 0.091271841 | 0.004358914 | 0.020528508 |
| FLNB | 1606.49803 | 0.24783793 | 0.077889872 | 0.000978133 | 0.005836776 |
| CCDC34 | 527.1298439 | -0.247788223 | 0.107737971 | 0.013939605 | 0.053285187 |
| EEF1A1P43 | 1771.651779 | -0.247763905 | 0.091786663 | 0.004593973 | 0.021472019 |
| MET | 1942.683891 | 0.247724685 | 0.076644993 | 0.00082097 | 0.005023583 |
| ARHGAP21 | 1037.630785 | 0.247722211 | 0.094799256 | 0.005870052 | 0.026293766 |
| RPL26L1 | 1115.651537 | -0.24754312 | 0.093483781 | 0.005385432 | 0.024535525 |
| RPS24 | 52464.799 | -0.247449344 | 0.080850627 | 0.001499867 | 0.008400129 |
| COPS9 | 1816.574645 | -0.247370027 | 0.116438685 | 0.021526658 | 0.07532373 |
| SHQ1 | 944.2709544 | -0.247308241 | 0.098984602 | 0.008182092 | 0.034516269 |
| TOMM20 | 5275.201975 | -0.24725088 | 0.090526754 | 0.004201568 | 0.019937364 |
| NOTCH2NLB | 1760.035709 | 0.247172225 | 0.087737149 | 0.003214298 | 0.016013232 |
| PIGM | 1033.083281 | -0.247139979 | 0.091245487 | 0.004491883 | 0.021063073 |
| ADPGK | 1749.889315 | -0.247065471 | 0.082700261 | 0.001892911 | 0.010256425 |
| SCDP1 | 409.2762823 | -0.247051619 | 0.110658515 | 0.016587006 | 0.060906952 |
| INTS8 | 866.444234 | -0.247026118 | 0.091909623 | 0.004774788 | 0.022227153 |
| RNF103-CHMP3 | 850.6940823 | 0.246703526 | 0.088710821 | 0.003582582 | 0.017475937 |
| SAP130 | 527.0383982 | 0.246607062 | 0.116383683 | 0.02186051 | 0.076306885 |
| DSTN | 2552.732315 | -0.246603894 | 0.102627883 | 0.010685876 | 0.043034261 |
| GBE1 | 2802.033679 | -0.246473532 | 0.084766591 | 0.002444423 | 0.01273468 |
| PXMP2 | 503.7102257 | 0.246348666 | 0.118475914 | 0.024117664 | 0.082440899 |
| GTF2H3 | 624.6696284 | 0.246339037 | 0.107101034 | 0.013902641 | 0.053193116 |
| TIMM8B | 599.8458095 | -0.246291742 | 0.115872007 | 0.021768789 | 0.076088139 |
| SLC2A1 | 958.5847048 | 0.24624498 | 0.102472517 | 0.010656922 | 0.042937628 |

| | | | | | |
|---|---|---|---|---|---|
| FBXL20 | 544.6107702 | 0.246179112 | 0.11220864 | 0.01825579 | 0.065915714 |
| KLF9 | 712.7971181 | -0.245933889 | 0.109701215 | 0.016222477 | 0.05985351 |
| NDUFB5 | 4578.610636 | 0.245908567 | 0.083743397 | 0.002211072 | 0.011687969 |
| RPL36A-HNRNPH2 | 10646.42446 | -0.245828659 | 0.065389666 | 0.000117537 | 0.000914779 |
| ZNF428 | 521.2026756 | 0.245793268 | 0.116083047 | 0.022061081 | 0.076944963 |
| H4-16 | 7.320737188 | 0.245768861 | 0.451692337 | 0.018860279 | 0.067591773 |
| CNTNAP3B | 4.128939733 | 0.245647462 | 0.446770042 | 0.026069875 | 0.087660588 |
| PDE4DIP | 1520.593998 | 0.245615408 | 0.090508988 | 0.00439082 | 0.020650687 |
| NOTCH2NLA | 1785.649969 | 0.245557015 | 0.08425055 | 0.002383525 | 0.01244924 |
| WDR61 | 1793.184532 | -0.245536056 | 0.088134631 | 0.003574371 | 0.017453734 |
| MINDY3 | 419.9907664 | 0.245386961 | 0.118332556 | 0.024427804 | 0.083287317 |
| HMGN4 | 2131.098331 | 0.245353915 | 0.098115732 | 0.008149239 | 0.034445092 |
| LOC105374942 | 3.295039973 | 0.245188573 | 0.463723871 | 0.006128141 | 0.027296621 |
| HSPA4 | 3073.8883 | -0.245186231 | 0.072076473 | 0.000456115 | 0.003017686 |
| CREB3 | 980.2390724 | -0.24513767 | 0.097964447 | 0.008214747 | 0.034628717 |
| COQ4 | 578.8347086 | 0.24505296 | 0.107818809 | 0.015051915 | 0.056455276 |
| LOC100129034 | 708.6897691 | 0.244997616 | 0.109670588 | 0.016595875 | 0.060913684 |
| TRNT | 409.9839984 | -0.244988333 | 0.122751671 | 0.029505667 | 0.096533089 |
| PDCD4 | 2541.731718 | -0.244967889 | 0.0981773 | 0.008352202 | 0.035114126 |
| DCTN4 | 1569.873658 | 0.244946537 | 0.087750918 | 0.003483343 | 0.017080462 |
| PALS2 | 790.8909985 | -0.244818058 | 0.120296004 | 0.02687486 | 0.089792002 |
| NUDT2 | 486.4818741 | -0.244689078 | 0.117476979 | 0.024180277 | 0.08260601 |
| NBN | 1692.918539 | -0.244337076 | 0.098073462 | 0.008447354 | 0.035427898 |
| RPL15 | 35585.18656 | -0.244232499 | 0.074942454 | 0.000925981 | 0.005565848 |
| MAN1B1 | 665.5705717 | 0.243920627 | 0.105971353 | 0.014016593 | 0.053451956 |
| WWTR1 | 4.770369721 | 0.24391099 | 0.442377282 | 0.02844034 | 0.093827656 |
| CNOT6 | 2137.319528 | 0.24389532 | 0.10837694 | 0.016119353 | 0.059606841 |
| BLOC1S5-TXNDC5 | 89734.28989 | -0.243685787 | 0.069070179 | 0.000310508 | 0.002147391 |
| MCRS1 | 2434.995868 | 0.243488427 | 0.096901396 | 0.008151342 | 0.034445092 |
| BTRC | 479.9558473 | -0.243481607 | 0.106758609 | 0.014848323 | 0.05588533 |
| MED13L | 1726.502202 | 0.243438151 | 0.086944403 | 0.003408675 | 0.016788402 |
| AGPAT5 | 3187.930282 | -0.24337725 | 0.10059041 | 0.010338309 | 0.041936872 |
| TXNDC5 | 90479.96824 | -0.24329789 | 0.071412278 | 0.000444016 | 0.002942603 |
| MARCHF5 | 776.2283132 | 0.243163126 | 0.117162679 | 0.024511734 | 0.083540582 |
| LSM4 | 4240.083574 | -0.243155124 | 0.079787876 | 0.001575357 | 0.008774671 |
| PBX2 | 771.8976457 | -0.243117474 | 0.095868287 | 0.007493138 | 0.03218221 |
| UIMC1 | 849.767753 | -0.243104668 | 0.090705501 | 0.00493635 | 0.022844142 |
| TIMM10 | 1250.1579 | 0.242927648 | 0.091390735 | 0.005263011 | 0.024091875 |
| TGDS | 673.8163169 | -0.242898447 | 0.119378742 | 0.02718037 | 0.090569059 |
| NOM1 | 814.8745378 | -0.242758516 | 0.100493578 | 0.010456158 | 0.042296028 |
| RLF | 758.0756362 | -0.242689499 | 0.105130402 | 0.01385227 | 0.053012102 |
| ZNF778 | 469.7131618 | -0.2426745 | 0.114936553 | 0.022642026 | 0.078559802 |
| PPFIBP1 | 409.9140165 | -0.242635004 | 0.112195818 | 0.020030184 | 0.070991137 |
| VAV2 | 482.8609666 | 0.242600614 | 0.108042623 | 0.016212577 | 0.059849327 |
| SOAT1 | 892.1236642 | 0.242555183 | 0.095925457 | 0.007604484 | 0.03257157 |
| CASP3 | 3010.897268 | 0.242530199 | 0.084983076 | 0.002907832 | 0.014693588 |
| ATP6V1F | 2399.597485 | -0.242508218 | 0.082800287 | 0.002321824 | 0.012176566 |
| FAM13B | 1029.84915 | 0.242418538 | 0.108024776 | 0.016224079 | 0.05985351 |
| CCDC59 | 850.4222707 | -0.242344484 | 0.098010334 | 0.00896033 | 0.037192243 |
| NTAQ1 | 437.4875284 | -0.242209711 | 0.114261966 | 0.022308574 | 0.077667384 |

| | | | | | |
|---|---|---|---|---|---|
| CHASERR | 1296.118718 | 0.242191404 | 0.081150546 | 0.001930646 | 0.010418511 |
| GAGE10 | 1413.914779 | -0.242179542 | 0.084694177 | 0.002885035 | 0.014608234 |
| UBXN2A | 707.6923766 | 0.241876031 | 0.105041539 | 0.014035489 | 0.053500453 |
| ATG3 | 3536.776056 | 0.241869156 | 0.075687677 | 0.000945776 | 0.005671051 |
| IST1 | 4253.896529 | -0.241364336 | 0.073897468 | 0.000754036 | 0.004673073 |
| TM2D3 | 750.7262379 | -0.241331158 | 0.104727165 | 0.014097908 | 0.053658504 |
| PTTG1IP | 3011.103918 | 0.241155183 | 0.082233579 | 0.002284746 | 0.012011214 |
| MPZL3 | 981.7601927 | -0.241153922 | 0.099870046 | 0.010528335 | 0.042548232 |
| FTH1P20 | 567.8418874 | 0.240993922 | 0.109675267 | 0.018413549 | 0.066361589 |
| ANO10 | 623.4226551 | -0.240814467 | 0.10371664 | 0.013553284 | 0.05210973 |
| ITM2C | 2066.358164 | 0.240733307 | 0.081342093 | 0.00209903 | 0.011177398 |
| CHMP4B | 2885.100722 | 0.240635763 | 0.073334467 | 0.000706929 | 0.004408316 |
| DENND10 | 854.0963994 | 0.240603303 | 0.090497881 | 0.005276573 | 0.024128201 |
| RNF220 | 1218.242801 | 0.240584346 | 0.088137995 | 0.004283144 | 0.020237711 |
| BRCA1 | 813.0104767 | 0.240461716 | 0.104336862 | 0.013963974 | 0.053321749 |
| DDX24 | 3932.2395 | -0.240351556 | 0.066905501 | 0.000227906 | 0.001642383 |
| GALK2 | 1075.505454 | -0.240309311 | 0.092061259 | 0.006126264 | 0.027296621 |
| RPS25P6 | 7614.694045 | -0.240253495 | 0.104284429 | 0.014235638 | 0.054084804 |
| OLMALINC | 438.6541035 | 0.240238127 | 0.111663593 | 0.020724257 | 0.073017691 |
| SNRPEP2 | 613.8271462 | -0.240219076 | 0.110307802 | 0.01944072 | 0.069284823 |
| RPL13AP5 | 5084.164203 | -0.24020048 | 0.10038988 | 0.011244796 | 0.044805827 |
| FES | 932.3299901 | -0.239968087 | 0.105589938 | 0.015426731 | 0.057586701 |
| TMEM39B | 520.3091529 | 0.239933776 | 0.114393731 | 0.023581974 | 0.081009532 |
| AP2M1 | 6788.418553 | 0.239851374 | 0.071371869 | 0.000533316 | 0.003444773 |
| TAPBP | 2132.216487 | 0.239616018 | 0.090710001 | 0.00557426 | 0.025216682 |
| TXNDC11 | 6699.228868 | -0.239531627 | 0.070850386 | 0.00050235 | 0.003277765 |
| CBX6 | 2256.050149 | -0.239415082 | 0.093034159 | 0.006848373 | 0.029938199 |
| RAB1A | 4081.954701 | -0.239167333 | 0.09557255 | 0.008355574 | 0.035119774 |
| LSM10 | 502.1573771 | 0.238986681 | 0.11870248 | 0.028929269 | 0.095186805 |
| IRF4 | 11604.80567 | 0.238964383 | 0.062050055 | 8.17E-05 | 0.000664103 |
| GAGE13 | 2575.561726 | -0.23892952 | 0.076729115 | 0.001276077 | 0.007293012 |
| LPIN2 | 739.5302599 | -0.238651001 | 0.097193187 | 0.009582671 | 0.03936966 |
| TUBA1C | 23069.73723 | 0.238554584 | 0.068541484 | 0.000345195 | 0.002355294 |
| GOLGA7 | 1523.49038 | -0.238461774 | 0.086324123 | 0.003924626 | 0.018821035 |
| MAGEA3 | 2192.951723 | -0.238287072 | 0.075442993 | 0.001098876 | 0.006448401 |
| DOK1 | 406.6913508 | 0.23821371 | 0.119404749 | 0.030170224 | 0.098101861 |
| DENND10P1 | 512.4352527 | 0.238176676 | 0.10869369 | 0.01884609 | 0.067569783 |
| SSR4 | 11673.75035 | -0.238053967 | 0.09669046 | 0.009373475 | 0.038629336 |
| CHTOP | 2868.591372 | -0.238033878 | 0.081199916 | 0.002328497 | 0.012207866 |
| NT5C2 | 2048.859815 | 0.238011423 | 0.078896133 | 0.001750491 | 0.009596067 |
| TLR6 | 3.671020237 | 0.23800272 | 0.441159124 | 0.022738029 | 0.078798169 |
| PIN1 | 1485.339538 | 0.237996778 | 0.118693302 | 0.029526788 | 0.096583943 |
| TPM3 | 23546.42201 | -0.237964676 | 0.064322438 | 0.000151392 | 0.001143831 |
| SLC25A22 | 531.9849223 | -0.237786505 | 0.117171232 | 0.028169939 | 0.093117277 |
| UBL7 | 1106.474584 | 0.237584302 | 0.108915331 | 0.019418988 | 0.069235868 |
| MAFK | 478.1272979 | -0.237560467 | 0.118877146 | 0.030259527 | 0.098276411 |
| LAX1 | 560.1639379 | 0.237515149 | 0.112354883 | 0.022783098 | 0.078900348 |
| NDUFA10 | 4594.601354 | -0.237470098 | 0.069292132 | 0.000429729 | 0.002859959 |
| RNF123 | 662.7912942 | -0.237181366 | 0.108063855 | 0.018942591 | 0.067816592 |
| ANKFY1 | 901.8722665 | 0.237080482 | 0.093811757 | 0.007900798 | 0.033558529 |

| | | | | | |
|---|---|---|---|---|---|
| LSG1 | 1039.692602 | 0.236972568 | 0.094308051 | 0.008111468 | 0.034327049 |
| ZNF35 | 487.6297376 | -0.236896754 | 0.107559748 | 0.018569019 | 0.066810659 |
| RWDD1 | 2193.036501 | -0.236819682 | 0.075875088 | 0.001253236 | 0.007179056 |
| FIP1L1 | 1638.14601 | 0.236785885 | 0.076769798 | 0.001410443 | 0.00796683 |
| DENND5B | 1264.708204 | 0.236534392 | 0.096339164 | 0.009509207 | 0.039142129 |
| OLAH | 3.86743869 | 0.236424169 | 0.447443284 | 0.013333166 | 0.051457643 |
| SNHG7 | 607.1906017 | 0.236387721 | 0.111227029 | 0.02238594 | 0.077889757 |
| PRDM4 | 620.6792427 | 0.236342908 | 0.100696877 | 0.012720319 | 0.049367494 |
| PWWP2A | 808.0620511 | -0.236267518 | 0.100075237 | 0.012336169 | 0.048189813 |
| FADD | 801.2134029 | 0.236192408 | 0.109911981 | 0.021130716 | 0.074208194 |
| TXLNG | 2069.909906 | -0.235996408 | 0.07645315 | 0.001411551 | 0.007969948 |
| PARL | 1693.859405 | 0.235381391 | 0.074405164 | 0.001082802 | 0.006360545 |
| H3C14 | 101.9752733 | 0.235335459 | 0.467375059 | 6.19E-05 | 0.000521488 |
| LYN | 2431.636552 | -0.235323198 | 0.080855941 | 0.002493502 | 0.012932007 |
| MAPRE1 | 9821.165435 | -0.235295069 | 0.0672924 | 0.000331485 | 0.002277004 |
| PHKA2 | 580.1431319 | 0.235223847 | 0.106460292 | 0.018282438 | 0.065964425 |
| PPP3CC | 1064.953442 | -0.235196579 | 0.08882542 | 0.005576553 | 0.025219068 |
| ADCY7 | 827.8154193 | 0.235181693 | 0.092488395 | 0.007525716 | 0.032290527 |
| SERBP1P6 | 506.784175 | -0.235173826 | 0.111911034 | 0.023855185 | 0.081730313 |
| JSRP1 | 3464.506527 | -0.235147721 | 0.09200907 | 0.007299116 | 0.031536625 |
| H3C15 | 101.8174358 | 0.235121891 | 0.467282568 | 5.17E-05 | 0.000442818 |
| TMCC3 | 685.1575751 | 0.234983787 | 0.097738897 | 0.010993712 | 0.04399209 |
| KIR2DL4 | 4.469882141 | 0.234937813 | 0.436610361 | 0.023137239 | 0.079765582 |
| MECP2 | 1078.507267 | 0.234765846 | 0.09948764 | 0.012702037 | 0.049318649 |
| MRPS23 | 2474.563132 | -0.234618817 | 0.069930648 | 0.000559499 | 0.003599128 |
| DLG1 | 2006.426696 | 0.234612712 | 0.085104359 | 0.004015847 | 0.019162972 |
| ERVK3-1 | 403.849251 | 0.234573293 | 0.11440328 | 0.026852099 | 0.089776967 |
| MYD88 | 1104.430225 | 0.234572853 | 0.092940493 | 0.00794316 | 0.033721916 |
| PITPNC1 | 2054.278991 | 0.234525588 | 0.073927889 | 0.001054567 | 0.00621789 |
| RNF103 | 1245.479688 | -0.234429913 | 0.086702238 | 0.004706598 | 0.02195694 |
| RMND5B | 905.1763988 | 0.23426513 | 0.0965788 | 0.010396704 | 0.042124407 |
| SS18 | 2054.509547 | 0.234261085 | 0.10136301 | 0.013816382 | 0.052886449 |
| LRCH3 | 470.6324421 | 0.234246237 | 0.113830723 | 0.026359245 | 0.088478977 |
| CARS1 | 2265.890259 | 0.234099035 | 0.078840234 | 0.002045947 | 0.010928333 |
| AMFR | 1094.047996 | -0.233968009 | 0.101555503 | 0.01451936 | 0.054921746 |
| BRPF1 | 554.4848743 | -0.233958315 | 0.113710459 | 0.026617463 | 0.089172878 |
| RANBP6 | 1024.878399 | -0.23393794 | 0.098968211 | 0.012356138 | 0.048235176 |
| TOE1 | 748.3896099 | 0.233887314 | 0.093584738 | 0.008538206 | 0.035722667 |
| MOB1A | 7618.166088 | 0.233776676 | 0.101152447 | 0.014104332 | 0.053668361 |
| HYOU1 | 18226.73926 | -0.23329265 | 0.070787491 | 0.000701872 | 0.004384675 |
| RPL37A | 65257.24579 | -0.233163988 | 0.084355775 | 0.00396507 | 0.018952006 |
| HLA-C | 42107.42371 | 0.233161733 | 0.078767294 | 0.002070637 | 0.011039782 |
| EIF3C | 6466.613501 | -0.233055411 | 0.062422616 | 0.000134497 | 0.001029659 |
| FLOT2 | 2841.690164 | 0.232906172 | 0.084816106 | 0.004182198 | 0.019863718 |
| UBA6 | 882.8663041 | -0.232755476 | 0.108290395 | 0.021426153 | 0.07509344 |
| EHMT2 | 799.1918441 | 0.232609816 | 0.110454469 | 0.02374551 | 0.081442066 |
| RPS26P8 | 1014.539992 | -0.232408597 | 0.08920461 | 0.006376893 | 0.028206407 |
| SNX11 | 977.041751 | 0.232386822 | 0.097146526 | 0.011505414 | 0.045634278 |
| RPS3 | 71190.88754 | -0.232380236 | 0.070990113 | 0.000754244 | 0.004673073 |
| ASF1B | 2075.65705 | 0.232256177 | 0.092722903 | 0.00844934 | 0.035427898 |

| | | | | | |
|---|---|---|---|---|---|
| AKAP8 | 1096.891898 | -0.232157827 | 0.087673706 | 0.005658096 | 0.025535104 |
| UBXN1 | 1146.371997 | 0.232012359 | 0.093942564 | 0.009293226 | 0.038344245 |
| B4GALT5 | 957.5602079 | 0.232007791 | 0.087241103 | 0.005423557 | 0.024676756 |
| PEX26 | 454.1408623 | 0.231987118 | 0.113877211 | 0.027992575 | 0.092739128 |
| ANO6 | 1239.38939 | 0.231954123 | 0.113565183 | 0.027603517 | 0.091678195 |
| GCLC | 1195.74742 | 0.231915212 | 0.099177934 | 0.013134552 | 0.050804343 |
| ASH1L | 1092.093705 | -0.231719546 | 0.096190151 | 0.011027392 | 0.044081569 |
| LMO4 | 1655.954376 | -0.231669222 | 0.094769435 | 0.010069548 | 0.04101961 |
| SMARCA4 | 2671.545006 | -0.231546542 | 0.099131555 | 0.013455265 | 0.051813507 |
| RAB8A | 2751.159538 | 0.231519466 | 0.073604467 | 0.001161933 | 0.006756614 |
| ELK1 | 1811.261924 | 0.23151427 | 0.088278277 | 0.006054235 | 0.027013962 |
| CIB1 | 1937.434551 | -0.231445688 | 0.100306012 | 0.014525265 | 0.054932084 |
| SPECC1 | 518.6455094 | 0.231445371 | 0.105200959 | 0.01892537 | 0.067768949 |
| ATP5MF-PTCD1 | 1984.543843 | -0.231436206 | 0.113597758 | 0.030225798 | 0.098221082 |
| PPP1CA | 4269.520406 | 0.231407359 | 0.09196541 | 0.008206843 | 0.034603825 |
| SNHG29 | 34069.96789 | -0.231334119 | 0.065960716 | 0.000339501 | 0.00232377 |
| AGTPBP1 | 1074.81694 | -0.23132345 | 0.097201502 | 0.012038608 | 0.047155073 |
| NOP58 | 5682.847222 | -0.231297906 | 0.066813738 | 0.000382914 | 0.002585157 |
| EIF4ENIF1 | 672.1741912 | -0.231258292 | 0.09911506 | 0.013533554 | 0.052056989 |
| PPP2R5A | 938.8163755 | 0.231230523 | 0.094139051 | 0.009653909 | 0.039643524 |
| GOLM2 | 2635.497377 | 0.231180473 | 0.113563751 | 0.027898978 | 0.092469612 |
| TMEM117 | 484.6465463 | -0.231178373 | 0.111481157 | 0.025890016 | 0.087191249 |
| KLHL6 | 2412.418488 | 0.231048167 | 0.086889739 | 0.00543696 | 0.02471177 |
| NDUFB4 | 2790.192327 | 0.231041113 | 0.077968757 | 0.002138144 | 0.011361228 |
| FANCG | 620.6996493 | 0.23066354 | 0.102343355 | 0.016591217 | 0.0609095 |
| DHX29 | 968.261107 | -0.23058084 | 0.100064368 | 0.014603055 | 0.055190116 |
| ASF1A | 1388.43034 | -0.230408132 | 0.099363513 | 0.014097068 | 0.053658504 |
| INIP | 1368.565969 | 0.230257749 | 0.091929527 | 0.008489746 | 0.035571483 |
| RPL27AP6 | 6113.683795 | -0.230234248 | 0.062620754 | 0.00017404 | 0.001293498 |
| RAVER1 | 2542.492925 | 0.230096273 | 0.093660108 | 0.00967453 | 0.039709373 |
| HCCS | 831.2497533 | 0.230077078 | 0.092883247 | 0.009172195 | 0.037899473 |
| RBM26 | 1942.730655 | -0.230052248 | 0.100406189 | 0.01515259 | 0.056746841 |
| MAP3K11 | 613.8765955 | 0.230028856 | 0.103261422 | 0.017750221 | 0.06450799 |
| RPL7AP66 | 1833.657948 | 0.229939878 | 0.083130249 | 0.003979673 | 0.019016554 |
| FANCF | 563.5675275 | 0.229421442 | 0.111850984 | 0.027274869 | 0.090760742 |
| MRPL45 | 2507.960454 | -0.229295685 | 0.076538121 | 0.001944211 | 0.010485175 |
| DOCK11 | 1871.380392 | 0.229282042 | 0.081833024 | 0.003564412 | 0.017416737 |
| TRIM25 | 2421.341785 | 0.229259694 | 0.073797903 | 0.001302358 | 0.007421172 |
| STK35 | 941.1624226 | 0.229228833 | 0.091160656 | 0.008304543 | 0.034939207 |
| SMYD2 | 431.5354409 | 0.229147568 | 0.108847999 | 0.023818167 | 0.081658918 |
| ANAPC13 | 1207.473564 | -0.229136237 | 0.086802869 | 0.005738389 | 0.025797424 |
| TRNT1 | 760.2816685 | -0.228953178 | 0.102898358 | 0.018001788 | 0.065230584 |
| CIAPIN1 | 2181.087498 | -0.228749999 | 0.081219737 | 0.003447851 | 0.0169428 |
| NOTCH2 | 1427.240334 | 0.228699844 | 0.084867569 | 0.004984424 | 0.023017406 |
| TUBG1 | 1768.823241 | 0.228637381 | 0.081112373 | 0.003395682 | 0.01673393 |
| NKAIN2 | 3.122855853 | 0.228564691 | 0.439471257 | 0.007706471 | 0.032951349 |
| KATNB1 | 694.5176908 | -0.228479201 | 0.113438594 | 0.030254462 | 0.098276411 |
| ARFGEF1 | 1248.6832 | -0.22840422 | 0.085755421 | 0.005460109 | 0.024790958 |
| TMOD3 | 1406.445838 | 0.2283677 | 0.112631842 | 0.028941872 | 0.095210182 |
| PARM1 | 2959.152826 | -0.228324035 | 0.068570762 | 0.000623752 | 0.0039595 |

| | | | | | |
|---|---|---|---|---|---|
| RNU5E-1 | 6.291715684 | 0.228147061 | 0.433253536 | 0.016125899 | 0.059618315 |
| TUT4 | 872.8899836 | 0.227995391 | 0.101941119 | 0.017673496 | 0.064256128 |
| ZWINT | 1961.266603 | 0.227968318 | 0.073465268 | 0.001364664 | 0.00773548 |
| CYB561A3 | 1201.875292 | 0.227756786 | 0.10502576 | 0.020760054 | 0.073128938 |
| CPT1A | 620.0670952 | -0.227736298 | 0.097212063 | 0.013388191 | 0.051625704 |
| TRPV2 | 1291.167882 | -0.2276945 | 0.108963501 | 0.025336276 | 0.08577679 |
| UBR7 | 1958.342239 | 0.227564486 | 0.073763664 | 0.001450251 | 0.008161394 |
| ITPR1 | 779.2983217 | 0.227551185 | 0.098812239 | 0.014906853 | 0.056020393 |
| MED16 | 1023.248041 | 0.227524528 | 0.108593936 | 0.024791208 | 0.084331427 |
| ATXN1L | 1408.74402 | -0.227499347 | 0.089175583 | 0.007340113 | 0.031658464 |
| WDCP | 898.9362451 | 0.227305316 | 0.087286888 | 0.006470668 | 0.028548422 |
| TMEM106C | 4570.495743 | 0.227295611 | 0.067137481 | 0.000502671 | 0.003278625 |
| DNPEP | 2413.26675 | 0.227188153 | 0.083270504 | 0.004499028 | 0.021079455 |
| DECR1 | 1521.40716 | -0.227146111 | 0.078955099 | 0.002865917 | 0.014536934 |
| TUBB | 32909.35106 | 0.22712492 | 0.064179885 | 0.000287719 | 0.002008231 |
| KIF2C | 1288.904866 | 0.226935457 | 0.084090853 | 0.004903908 | 0.022714584 |
| EVI2A | 1503.45664 | 0.226821437 | 0.089895631 | 0.008150274 | 0.034445092 |
| NOTCH2NLC | 1594.478928 | 0.226802785 | 0.089278394 | 0.007756213 | 0.033114977 |
| TMEM147 | 3865.306968 | -0.226600292 | 0.105274536 | 0.021804097 | 0.076156019 |
| BRCC3 | 654.176217 | 0.226556128 | 0.110172282 | 0.02720109 | 0.090569059 |
| DSTYK | 661.0370405 | 0.226510494 | 0.099456532 | 0.015807188 | 0.058703211 |
| YIPF2 | 1015.382664 | -0.226373678 | 0.104504485 | 0.021106735 | 0.0741691 |
| HP1BP3 | 3789.918042 | 0.22635447 | 0.069698622 | 0.000832879 | 0.005083882 |
| FRYL | 1878.807402 | -0.226302188 | 0.089091596 | 0.007713334 | 0.03297255 |
| RPSAP45 | 1612.883838 | -0.226273988 | 0.092739965 | 0.010335754 | 0.041936329 |
| SPSB1 | 432.5041769 | 0.226160636 | 0.112894135 | 0.03068182 | 0.099404505 |
| RBFA | 764.2693835 | -0.226055881 | 0.092133816 | 0.009980625 | 0.040714836 |
| MRPL33 | 1613.219369 | -0.226033378 | 0.090758006 | 0.009091542 | 0.037637638 |
| IL18R1 | 3.608689844 | 0.2259772 | 0.432497948 | 0.016473724 | 0.060582898 |
| DYNLRB1 | 1598.780941 | 0.225967247 | 0.081167409 | 0.003804804 | 0.018302162 |
| RERG | 3.002003392 | 0.225902738 | 0.441929039 | 0.001431639 | 0.008070212 |
| RC3H2 | 861.973196 | 0.225645291 | 0.111351188 | 0.029300753 | 0.096052592 |
| RHBDD2 | 939.7214773 | 0.22556511 | 0.106689854 | 0.023824607 | 0.081664829 |
| LBR | 3196.843526 | 0.225518748 | 0.111206552 | 0.029184517 | 0.095790045 |
| PFN1 | 19032.09182 | 0.225436997 | 0.086915759 | 0.006680377 | 0.029309372 |
| LITAF | 5546.447538 | -0.225420271 | 0.072007442 | 0.001253807 | 0.007179952 |
| VPS36 | 2012.257805 | -0.22525756 | 0.094891506 | 0.012393617 | 0.04835968 |
| TXNDC15 | 870.7773855 | -0.22510612 | 0.088522743 | 0.007803575 | 0.033284361 |
| SUGT1 | 1555.135124 | -0.225034452 | 0.098102067 | 0.015327457 | 0.057265494 |
| E2F2 | 2740.716646 | 0.224962744 | 0.080491313 | 0.003672299 | 0.017798264 |
| RACK1 | 62344.28657 | -0.224954999 | 0.071532827 | 0.001203849 | 0.006958293 |
| HIF1A | 1174.234716 | 0.224884903 | 0.111625219 | 0.030185875 | 0.098128176 |
| PAPOLA | 6094.488049 | 0.224862405 | 0.084138361 | 0.005219567 | 0.023924628 |
| MCUR1 | 1565.225702 | 0.224594093 | 0.08850875 | 0.007873548 | 0.033491723 |
| UCK2 | 6145.48848 | -0.224498296 | 0.068596081 | 0.000778981 | 0.004805707 |
| TSPAN3 | 2594.337041 | -0.224443321 | 0.070879004 | 0.001116623 | 0.006539248 |
| TIMM10B | 1377.473525 | -0.224186311 | 0.091130313 | 0.009907606 | 0.040493276 |
| ANAPC1P4 | 451.4303473 | 0.22393219 | 0.10833174 | 0.026857725 | 0.089776967 |
| TRAK1 | 868.7607541 | -0.223539366 | 0.111269714 | 0.030885865 | 0.099860067 |
| H3C8 | 71.33796237 | 0.223533679 | 0.434301859 | 0.008505104 | 0.035618594 |

| | | | | | |
|---|---|---|---|---|---|
| BLOC1S2 | 1449.735873 | -0.223494024 | 0.087653681 | 0.007676778 | 0.032852743 |
| EIF3CL | 6187.002757 | -0.22348534 | 0.063638965 | 0.000325433 | 0.002239876 |
| MAGEA1 | 2581.849815 | -0.223453884 | 0.077657055 | 0.002899012 | 0.014670424 |
| ANXA6 | 12958.18419 | 0.223451314 | 0.065565602 | 0.000471381 | 0.003097875 |
| HSD17B11 | 643.1680481 | -0.223443487 | 0.106609734 | 0.025196247 | 0.085369481 |
| ST7 | 761.9700221 | -0.223361377 | 0.101936913 | 0.020099567 | 0.071199009 |
| NFIL3 | 787.4218518 | 0.223313287 | 0.094357071 | 0.012658738 | 0.049205692 |
| H3-3A | 3114.225508 | 0.223116332 | 0.093225137 | 0.011780753 | 0.046365294 |
| MAD2L1BP | 894.9654238 | -0.223115104 | 0.107724033 | 0.026867532 | 0.089784841 |
| RAB5IF | 2183.100207 | -0.223055112 | 0.095635826 | 0.013976579 | 0.053327588 |
| ETF1 | 7942.028085 | -0.223010194 | 0.071332225 | 0.001287167 | 0.007344274 |
| FKBP1C | 740.8478103 | -0.222998358 | 0.09795547 | 0.016210931 | 0.059849327 |
| TBRG4 | 2181.717746 | -0.222889715 | 0.094145605 | 0.012761232 | 0.049504089 |
| NDUFS5 | 4629.004888 | -0.222709221 | 0.078168773 | 0.003188909 | 0.015918804 |
| UQCC2 | 1057.415722 | 0.222572836 | 0.09081703 | 0.010136843 | 0.041245234 |
| SLC7A6OS | 655.0162791 | -0.222500548 | 0.099544993 | 0.017957089 | 0.065095858 |
| CRCP | 936.5626126 | 0.222453024 | 0.087878399 | 0.008071906 | 0.034184688 |
| MAGEA6 | 3024.270881 | -0.222224811 | 0.071325019 | 0.001336422 | 0.0075903 |
| TMEM9B | 1271.220263 | 0.222159106 | 0.085861516 | 0.006898127 | 0.030104554 |
| COX15 | 1586.878707 | -0.221942068 | 0.078021355 | 0.003221252 | 0.016038649 |
| CTNNB1 | 4036.691529 | -0.221940701 | 0.073444338 | 0.001826461 | 0.009939922 |
| MGA | 1171.235078 | 0.221634785 | 0.098551604 | 0.017167162 | 0.06275781 |
| GINS2 | 2057.840563 | -0.221546363 | 0.079263896 | 0.003755237 | 0.018109003 |
| MRPL9 | 3941.31887 | -0.221519119 | 0.06811642 | 0.000840318 | 0.00511666 |
| SCAMP5 | 813.7210353 | 0.221496074 | 0.109073978 | 0.02949049 | 0.096501665 |
| MRPL44 | 2807.391973 | -0.221376653 | 0.068806136 | 0.000945306 | 0.005670196 |
| SMIM19 | 652.4251765 | -0.221341974 | 0.097382019 | 0.016386329 | 0.060323566 |
| DCAF15 | 763.2189635 | 0.221255965 | 0.101497591 | 0.020502915 | 0.072311399 |
| CLNS1A | 3611.27229 | -0.221232873 | 0.069132741 | 0.001001564 | 0.005954028 |
| BRK1 | 2715.246397 | -0.221232306 | 0.071815592 | 0.001508606 | 0.008443611 |
| ARPC5L | 1859.482143 | 0.221231382 | 0.095163678 | 0.014250499 | 0.05412939 |
| RPL4 | 89090.28676 | -0.221186178 | 0.065010875 | 0.000525509 | 0.003407051 |
| TRA2A | 3011.239667 | -0.221156077 | 0.104053139 | 0.02366149 | 0.081218241 |
| POLR2B | 6061.355607 | 0.221138891 | 0.067454022 | 0.000758057 | 0.004695022 |
| CDK1 | 3105.964545 | 0.221058338 | 0.104685121 | 0.024263013 | 0.082855965 |
| NFS1 | 953.5895864 | -0.221020382 | 0.089257313 | 0.009519496 | 0.039165863 |
| INPPL1 | 852.9798487 | 0.220866139 | 0.098667368 | 0.01783198 | 0.064750767 |
| LIN52 | 732.3331062 | -0.22076905 | 0.092957379 | 0.012562466 | 0.048930326 |
| TRMT10C | 2016.749146 | -0.220579447 | 0.085193781 | 0.006893977 | 0.030101615 |
| TRABD | 1499.902148 | 0.220521581 | 0.107325717 | 0.02790603 | 0.092470095 |
| CISD2 | 1585.648405 | -0.220512695 | 0.10294724 | 0.022758849 | 0.078838764 |
| DCAF7 | 5533.195922 | 0.220324438 | 0.067275601 | 0.000771054 | 0.004760191 |
| COA6 | 546.1293238 | -0.220317487 | 0.109042504 | 0.030361289 | 0.098555008 |
| LOC105374369 | 3.858849021 | -0.22023452 | 0.420633634 | 0.025938029 | 0.087318981 |
| GDE1 | 1654.907612 | -0.220082638 | 0.078372493 | 0.003624596 | 0.017641126 |
| FAM91A1 | 2131.069729 | -0.219975402 | 0.103370403 | 0.023596464 | 0.081027158 |
| STMN1 | 10580.45405 | 0.219972863 | 0.065140903 | 0.000533089 | 0.003444591 |
| RABL3 | 1418.70931 | 0.219804637 | 0.089373533 | 0.009947782 | 0.04059042 |
| TMEM41A | 2171.665624 | -0.219738352 | 0.085403097 | 0.007307553 | 0.031541597 |
| TMEM50A | 4263.376879 | 0.219645149 | 0.082691928 | 0.005659394 | 0.025535104 |

| | | | | | |
|---|---|---|---|---|---|
| ZCRB1 | 1911.309818 | 0.219400682 | 0.082101272 | 0.005431021 | 0.024697741 |
| ZBED4 | 1324.472784 | 0.219390363 | 0.082328774 | 0.005560824 | 0.025182216 |
| RHNO1 | 811.3898332 | 0.219368813 | 0.092407528 | 0.012592471 | 0.048992096 |
| CLTC | 18221.27719 | -0.219326257 | 0.069077171 | 0.001104135 | 0.006474877 |
| SENP5 | 1081.540984 | 0.219319025 | 0.085904708 | 0.007677719 | 0.032852743 |
| WDFY1 | 1417.284092 | -0.219239027 | 0.094611234 | 0.014674655 | 0.055424426 |
| ATG4B | 1876.585687 | 0.219091045 | 0.101909012 | 0.021854456 | 0.076301133 |
| CCNF | 1060.775511 | 0.219090828 | 0.104017704 | 0.024833945 | 0.084439314 |
| ATP2A3 | 4829.407175 | 0.219022416 | 0.102333142 | 0.022853282 | 0.079081458 |
| ASB1 | 1057.44741 | 0.219011239 | 0.086839177 | 0.008410026 | 0.035297243 |
| MORC2 | 727.3578592 | 0.218777005 | 0.108415418 | 0.030662898 | 0.099361791 |
| SEC63 | 3166.89935 | -0.218764952 | 0.082565139 | 0.005830662 | 0.026144375 |
| H1-4 | 164.0977435 | 0.218618745 | 0.440484526 | 0.000271598 | 0.001909568 |
| SRF | 875.9163615 | 0.21856584 | 0.104525552 | 0.025669369 | 0.086586428 |
| PPP1R11 | 908.8213402 | 0.218552135 | 0.090925527 | 0.011623486 | 0.045902829 |
| RPL5 | 59771.58512 | -0.218485039 | 0.066070982 | 0.000706401 | 0.00440661 |
| RAD18 | 856.2756162 | -0.21845949 | 0.106084951 | 0.028033363 | 0.092803241 |
| SCN4B | 3.29871326 | 0.218430206 | 0.422122596 | 0.02009814 | 0.071199009 |
| BAP1 | 1204.632829 | -0.218421188 | 0.098300213 | 0.018858495 | 0.067591773 |
| CACNG6 | 1441.301512 | 0.2184079 | 0.100466889 | 0.021123427 | 0.074197644 |
| SYNE3 | 1014.402978 | 0.218283098 | 0.09256168 | 0.013125226 | 0.050779611 |
| SLC39A6 | 4207.948451 | -0.218282869 | 0.082460462 | 0.005925776 | 0.026502238 |
| GSK3A | 1379.759805 | 0.218221585 | 0.104232964 | 0.025697592 | 0.086627447 |
| RNH1 | 2082.732558 | 0.218153563 | 0.099098016 | 0.019648339 | 0.069895294 |
| ABCF2 | 2239.880208 | -0.218082041 | 0.075920875 | 0.002981039 | 0.014989154 |
| PRPSAP1 | 701.6041707 | 0.218027497 | 0.099137504 | 0.019794505 | 0.070314143 |
| ELL2 | 7542.388245 | -0.217948637 | 0.105547188 | 0.026258671 | 0.088175568 |
| BICD2 | 546.5499532 | 0.217928564 | 0.104379816 | 0.026011743 | 0.087533105 |
| VRK3 | 1212.07365 | 0.217890033 | 0.088281454 | 0.009797969 | 0.040139916 |
| INO80E | 594.631501 | -0.217864732 | 0.101483938 | 0.022785739 | 0.078900348 |
| TLN1 | 2727.957332 | 0.217775139 | 0.082190449 | 0.005815321 | 0.026082339 |
| GBA2 | 544.205974 | 0.217722447 | 0.104657685 | 0.026613483 | 0.089172878 |
| DCAF4 | 797.7759245 | 0.217630692 | 0.095273219 | 0.016036849 | 0.059390529 |
| QRICH1 | 1851.850217 | -0.217490532 | 0.075536368 | 0.002921922 | 0.014747302 |
| NDUFC1 | 967.4786534 | -0.217370533 | 0.09960324 | 0.020917435 | 0.073608457 |
| ARPC1B | 4120.263496 | 0.217273341 | 0.090101974 | 0.011453879 | 0.045466513 |
| TYW1 | 1091.623382 | -0.217184774 | 0.085376847 | 0.007831885 | 0.033388662 |
| SLMAP | 1272.691946 | -0.217019587 | 0.098569839 | 0.019850433 | 0.07048606 |
| RBM15 | 768.9591514 | -0.216874028 | 0.090050069 | 0.011617464 | 0.045892799 |
| RALBP1 | 1263.362115 | 0.216812779 | 0.082915682 | 0.006473637 | 0.028553497 |
| RPL22P1 | 9025.192931 | -0.216796587 | 0.074890391 | 0.002793457 | 0.014231922 |
| TINF2 | 1223.563108 | 0.216674878 | 0.082322085 | 0.006105976 | 0.027216778 |
| USP10 | 2850.503922 | -0.216480811 | 0.068816949 | 0.001229437 | 0.007068449 |
| RPL23 | 44454.30283 | -0.216386362 | 0.060428856 | 0.000201926 | 0.001474176 |
| INPP5K | 1076.398406 | 0.216314964 | 0.099133166 | 0.02122809 | 0.074519935 |
| DNAJC15 | 835.4724152 | -0.21599564 | 0.097510717 | 0.019331214 | 0.068998897 |
| ECE1 | 1492.580615 | 0.215989945 | 0.091945732 | 0.013579201 | 0.052186203 |
| ARMC10 | 1231.460436 | -0.215968739 | 0.089628567 | 0.011656428 | 0.046011936 |
| MRPS31 | 831.6346083 | -0.215950669 | 0.092586577 | 0.0142603 | 0.054154738 |
| SNX12 | 1018.3836 | 0.215859815 | 0.09100363 | 0.01281034 | 0.049661216 |

| | | | | | |
|---|---|---|---|---|---|
| EFCAB14 | 2140.714555 | 0.215833025 | 0.078345332 | 0.004292593 | 0.020271292 |
| KNTC1 | 792.7885505 | 0.215781361 | 0.099803406 | 0.02177148 | 0.076088139 |
| GAGE12J | 2516.740177 | -0.21567955 | 0.076919265 | 0.003724441 | 0.017990555 |
| EIF2B5 | 1875.654384 | 0.215598622 | 0.083621361 | 0.0072431 | 0.031357193 |
| TMEM59 | 4431.146891 | 0.215570259 | 0.079290614 | 0.004791964 | 0.022283147 |
| RPS26P6 | 916.4719013 | -0.215436972 | 0.09075013 | 0.012822015 | 0.049684233 |
| H3P6 | 2961.903058 | 0.215031073 | 0.097380816 | 0.019678345 | 0.069973285 |
| CDC25B | 3897.373797 | 0.214993171 | 0.08678209 | 0.009597152 | 0.039419802 |
| KCMF1 | 3065.511674 | 0.214923258 | 0.068200705 | 0.001200754 | 0.006942725 |
| ERF | 1188.897071 | -0.214835148 | 0.096126418 | 0.018462227 | 0.066481678 |
| TIMELESS | 2220.72032 | 0.214806597 | 0.075526168 | 0.003268988 | 0.016211073 |
| SELENOT | 3448.036536 | 0.214774537 | 0.101622558 | 0.024715924 | 0.084120607 |
| PEX19 | 3351.52952 | -0.214602255 | 0.07008318 | 0.001641687 | 0.009097311 |
| SIGMAR1 | 2924.740536 | -0.214470319 | 0.081744298 | 0.006403398 | 0.028287663 |
| EFHD2 | 1274.459699 | 0.214443775 | 0.105868896 | 0.030640116 | 0.09930655 |
| LRP10 | 2982.989606 | 0.214378749 | 0.097974675 | 0.020632102 | 0.0727374 |
| IGSF3 | 4.973362995 | 0.214290141 | 0.412266662 | 0.028869026 | 0.095024695 |
| UBXN7 | 1837.692394 | 0.214272868 | 0.089928436 | 0.012469404 | 0.048600648 |
| COX19 | 629.1214169 | 0.214268973 | 0.094955491 | 0.017368061 | 0.063371831 |
| CREB3L2 | 2986.252527 | -0.214229717 | 0.072244402 | 0.002245067 | 0.011842165 |
| CD81 | 3948.746341 | -0.213907725 | 0.093237669 | 0.015895819 | 0.058969128 |
| METTL17 | 1398.36918 | -0.213814546 | 0.085652669 | 0.00923455 | 0.038129402 |
| RPL23AP42 | 13698.12968 | -0.21373318 | 0.065210724 | 0.000784268 | 0.004833159 |
| MED10 | 2889.222896 | -0.21367011 | 0.068303224 | 0.001315892 | 0.007485971 |
| ZNF217 | 4825.539589 | 0.213478222 | 0.079934093 | 0.005550642 | 0.025142679 |
| BZW2 | 4850.139007 | -0.213379474 | 0.066626078 | 0.001022288 | 0.006054364 |
| GTPBP6 | 926.3675497 | 0.213184799 | 0.098929884 | 0.022528917 | 0.078273677 |
| NEMP1 | 4144.133223 | 0.213067122 | 0.067638137 | 0.001173538 | 0.006810374 |
| ZNF740 | 1000.866626 | 0.21293997 | 0.086922798 | 0.010454987 | 0.042296028 |
| PDP2 | 804.1060433 | -0.212932684 | 0.091807918 | 0.014894726 | 0.056011285 |
| PAK1 | 1373.209902 | 0.212925978 | 0.079075638 | 0.005217765 | 0.023924628 |
| UBR3 | 1766.62216 | -0.212406854 | 0.095877342 | 0.019464161 | 0.069339821 |
| HLA-B | 56627.87577 | 0.212151481 | 0.083306581 | 0.008075207 | 0.034190306 |
| DAXX | 840.712964 | 0.212122038 | 0.091525859 | 0.014919314 | 0.056046756 |
| GARS1 | 9584.849296 | -0.21210063 | 0.061616005 | 0.000431324 | 0.002869476 |
| RPL4P5 | 2836.022763 | -0.212050818 | 0.073318695 | 0.002873985 | 0.014565056 |
| ABI2 | 1928.292993 | 0.211995933 | 0.078828406 | 0.005282502 | 0.024142804 |
| MRPS24 | 1790.024182 | -0.211904726 | 0.089730564 | 0.013412769 | 0.051684227 |
| TRRAP | 2411.297874 | 0.211893739 | 0.076775864 | 0.004209124 | 0.019952688 |
| PTPN6 | 4650.127487 | 0.211852521 | 0.067390789 | 0.001242154 | 0.007124201 |
| HLA-H | 9005.846693 | 0.21174473 | 0.073136318 | 0.002814836 | 0.014332409 |
| NGRN | 1753.124303 | 0.211494595 | 0.07823196 | 0.005076691 | 0.023378866 |
| HLA-A | 40291.52495 | 0.211325566 | 0.0838411 | 0.008567077 | 0.035834799 |
| LOC124903128 | 2.989897106 | -0.211276427 | 0.422618544 | 0.002017995 | 0.010812384 |
| H2AC17 | 41.43457302 | 0.211163371 | 0.419486149 | 0.009357806 | 0.038583125 |
| ESYT2 | 4349.580731 | -0.211128739 | 0.068094219 | 0.001461649 | 0.008212648 |
| AP1M1 | 2036.541686 | 0.210789505 | 0.095283632 | 0.019672487 | 0.069966821 |
| COX6A1P2 | 2464.849853 | 0.210772237 | 0.093232328 | 0.017388985 | 0.063421451 |
| PRMT7 | 884.8279779 | -0.210537434 | 0.099709283 | 0.025441738 | 0.086047063 |
| MECR | 1014.422684 | -0.210229575 | 0.101030479 | 0.027412595 | 0.091148984 |

| | | | | | |
|---|---|---|---|---|---|
| GPKOW | 590.1700548 | 0.210168897 | 0.103340268 | 0.030362705 | 0.098555008 |
| SRPRA | 7011.740104 | -0.210047618 | 0.063733434 | 0.000752806 | 0.004670847 |
| TRIB2 | 1513.570639 | -0.209990669 | 0.080039299 | 0.006499219 | 0.028636341 |
| MAD1L1 | 931.6235369 | -0.209754053 | 0.090861402 | 0.015511154 | 0.057814605 |
| APEX2 | 770.9058688 | 0.209738552 | 0.091487456 | 0.016065311 | 0.059450865 |
| HEXB | 5154.112642 | -0.209659409 | 0.064509933 | 0.000871987 | 0.005279787 |
| REXO2 | 2961.558753 | -0.209614504 | 0.071038621 | 0.002384071 | 0.01244924 |
| DLGAP4 | 746.7482597 | 0.20957943 | 0.098720016 | 0.024392609 | 0.083183696 |
| PTK2B | 2920.717337 | -0.209476187 | 0.080098383 | 0.006671025 | 0.029283177 |
| ASB8 | 1166.997088 | 0.209464612 | 0.081969751 | 0.007859532 | 0.033460066 |
| RAB11FIP1 | 1787.157624 | -0.209451892 | 0.074355543 | 0.003637289 | 0.01769182 |
| LINC01465 | 4.014183057 | 0.20935815 | 0.410422169 | 0.022637509 | 0.078559802 |
| WBP1L | 974.2442646 | 0.209281683 | 0.091416001 | 0.016222695 | 0.05985351 |
| NRBF2 | 739.3650021 | -0.209247303 | 0.099840176 | 0.026469216 | 0.088762086 |
| GEN1 | 2836.909736 | 0.209203895 | 0.09518091 | 0.019987757 | 0.070884289 |
| ZNF572 | 3.048231341 | -0.209134393 | 0.420229936 | 0.001986879 | 0.010673959 |
| NBPF15 | 1072.798277 | 0.20906387 | 0.086248768 | 0.01134773 | 0.045129366 |
| SLC25A3 | 13477.43291 | -0.208914489 | 0.075709997 | 0.00435812 | 0.020528508 |
| ZC3H15 | 4524.551495 | -0.20882754 | 0.08036715 | 0.007004461 | 0.030484104 |
| ARPC5 | 2277.330632 | 0.208650212 | 0.092356015 | 0.017560967 | 0.063967874 |
| ISG20L2 | 3042.166249 | -0.20864429 | 0.079233736 | 0.006372544 | 0.02819752 |
| ITPRID2 | 1200.550008 | 0.208554644 | 0.099183782 | 0.026464744 | 0.088762086 |
| KBTBD4 | 797.442032 | 0.208440106 | 0.092316779 | 0.017647614 | 0.064188979 |
| CBX3 | 7883.764718 | -0.208352426 | 0.089999924 | 0.015112183 | 0.05663226 |
| SRD5A1 | 1031.598516 | 0.208311586 | 0.092523828 | 0.017918536 | 0.06500353 |
| AAAS | 1503.196141 | 0.208260941 | 0.083512056 | 0.009386642 | 0.038674396 |
| RPS16 | 52225.51742 | -0.208058264 | 0.078588779 | 0.006095063 | 0.027175129 |
| URGCP-MRPS24 | 1815.43762 | -0.208001932 | 0.089144367 | 0.014617903 | 0.055234177 |
| AVL9 | 2473.713621 | -0.207993835 | 0.079879049 | 0.006910412 | 0.030150569 |
| ATP1A1 | 6554.077335 | -0.207929567 | 0.062596657 | 0.0006859 | 0.004297297 |
| RPS14 | 44455.26932 | -0.207895383 | 0.082279745 | 0.008197373 | 0.034572309 |
| ABCF2-H2BK1 | 1437.100427 | -0.20787898 | 0.080325165 | 0.007251206 | 0.031375135 |
| RPL6 | 41375.99304 | -0.207779312 | 0.062263834 | 0.000649981 | 0.004104928 |
| ZNF622 | 1832.127567 | -0.207755475 | 0.076797562 | 0.005140339 | 0.023636549 |
| SNORA79B | 4.965437726 | 0.207732386 | 0.415767053 | 0.009665591 | 0.039682086 |
| YTHDF2 | 3565.760417 | -0.207722521 | 0.07757414 | 0.005565851 | 0.025188299 |
| ERP44 | 3872.772047 | -0.207644642 | 0.075024108 | 0.004256334 | 0.020133008 |
| CYBA | 5858.99131 | -0.20760134 | 0.102178749 | 0.030834538 | 0.099768629 |
| MKRN2 | 1375.080766 | -0.207274397 | 0.085528088 | 0.011501126 | 0.045627719 |
| SRA1 | 995.1076456 | -0.207071101 | 0.100786065 | 0.029402715 | 0.096287218 |
| SNORD3D | 134.5027834 | 0.206980965 | 0.419307337 | 0.00358883 | 0.017501482 |
| NFE2L1 | 6631.382846 | 0.206925693 | 0.070664847 | 0.002557565 | 0.013189206 |
| SNORD3B-1 | 135.1705468 | 0.206839322 | 0.419193536 | 0.003535862 | 0.017286993 |
| WIPF1 | 5804.322658 | 0.206638237 | 0.074622244 | 0.004202309 | 0.019937364 |
| SNORD3B-2 | 135.4977192 | 0.206628776 | 0.418967649 | 0.003517359 | 0.017215969 |
| SEC23B | 4214.9016 | -0.206597228 | 0.066939252 | 0.001553405 | 0.008660756 |
| RCL1 | 925.1195606 | 0.206431272 | 0.089202313 | 0.015318316 | 0.057256204 |
| UBBP4 | 4012.956752 | 0.206359019 | 0.074695848 | 0.004313265 | 0.020341178 |
| RSRC1 | 941.2863459 | 0.20631499 | 0.10106135 | 0.030151414 | 0.098089805 |
| GORASP2 | 6627.572696 | -0.206283304 | 0.06265666 | 0.00074864 | 0.004646669 |

| | | | | | |
|---|---|---|---|---|---|
| PIGO | 1011.034177 | 0.206170991 | 0.097654627 | 0.025792501 | 0.086896642 |
| GPBP1L1 | 1189.496621 | 0.206040004 | 0.097135718 | 0.024949138 | 0.084702216 |
| SZRD1 | 3401.404005 | 0.206026947 | 0.069198459 | 0.002198681 | 0.011643593 |
| CAMSAP1 | 1255.810029 | 0.205999636 | 0.094647737 | 0.021801124 | 0.076156019 |
| SRPRB | 7063.655902 | -0.205872616 | 0.066806751 | 0.001577444 | 0.00878065 |
| ETV5 | 987.3436963 | 0.2057463 | 0.088408732 | 0.0148552 | 0.055899067 |
| SNRPA1 | 2482.431865 | -0.205734199 | 0.073194361 | 0.00375854 | 0.018114843 |
| PIP4K2C | 1430.882964 | 0.205551741 | 0.086700531 | 0.013224604 | 0.051084199 |
| ALAD | 803.9144512 | 0.205272273 | 0.090987201 | 0.017878887 | 0.064907483 |
| TMEM18 | 2709.317734 | -0.205229665 | 0.084431505 | 0.011439766 | 0.045425917 |
| TAF1B | 854.7512275 | 0.205139936 | 0.095748927 | 0.023773213 | 0.081520937 |
| TNPO2 | 894.5905979 | -0.20503214 | 0.097586779 | 0.026404687 | 0.088602183 |
| CERS5 | 876.4227417 | 0.204872246 | 0.088077337 | 0.015089894 | 0.056560972 |
| NR2C2 | 1263.812888 | 0.204637327 | 0.087034837 | 0.013972037 | 0.053327588 |
| KLF10 | 1007.573344 | -0.204601292 | 0.098405925 | 0.027896679 | 0.092469612 |
| MSN | 26876.72863 | 0.204556218 | 0.059626472 | 0.000448928 | 0.002974018 |
| MEMO1 | 1127.268879 | -0.204543124 | 0.089101836 | 0.016340291 | 0.060209397 |
| SNORD3A | 291.4167703 | 0.204462622 | 0.420846949 | 0.00063426 | 0.004017365 |
| ZBTB1 | 1342.255526 | -0.204147753 | 0.097069369 | 0.026406183 | 0.088602183 |
| POLR1D | 2658.931329 | -0.203982065 | 0.077257827 | 0.006274123 | 0.027812723 |
| CHMP1B | 1164.351259 | -0.203968076 | 0.09677887 | 0.0261138 | 0.087757166 |
| USP11 | 1046.528207 | 0.203960721 | 0.093498004 | 0.021715012 | 0.075921433 |
| CDK2 | 1952.344188 | 0.203956 | 0.078863109 | 0.007321192 | 0.031584722 |
| PDE3B | 3.129593481 | -0.203844074 | 0.414410655 | 0.001970879 | 0.010605876 |
| PTGR3 | 955.1014954 | 0.203702984 | 0.094003281 | 0.02252497 | 0.078273677 |
| MMS19 | 1218.499221 | -0.203660733 | 0.093799091 | 0.022437168 | 0.078022381 |
| HMG20B | 993.5960923 | 0.203600627 | 0.088640679 | 0.016192147 | 0.05981216 |
| SMIM7 | 1805.674208 | 0.203546362 | 0.079430911 | 0.007860227 | 0.033460066 |
| C3orf52 | 662.3906428 | 0.203523915 | 0.095133596 | 0.024050939 | 0.082261526 |
| PRG4 | 3.036141836 | 0.203409602 | 0.414369521 | 0.001735514 | 0.009534868 |
| INPP4A | 1553.458285 | 0.203396551 | 0.075345927 | 0.005257655 | 0.024073722 |
| SCARNA16 | 4.559190129 | 0.203366717 | 0.413686068 | 0.004252734 | 0.020133008 |
| PPP2R5E | 1253.09643 | 0.203341606 | 0.098756229 | 0.029222558 | 0.095878561 |
| CMTM6 | 3856.922243 | -0.203298773 | 0.096376902 | 0.026085933 | 0.087697555 |
| CDC34 | 1081.993514 | -0.203105541 | 0.097786249 | 0.028246587 | 0.093277296 |
| NECAP2 | 964.4225399 | 0.203065741 | 0.090497557 | 0.01860802 | 0.066886007 |
| UFD1 | 1699.774481 | 0.202995319 | 0.075898308 | 0.005679468 | 0.025588625 |
| EIF3B | 11820.71204 | -0.202943191 | 0.072703162 | 0.004033481 | 0.019231052 |
| POLR2F | 1267.862869 | -0.202699149 | 0.088030404 | 0.015837971 | 0.058779712 |
| RNF167 | 2077.143018 | 0.202686945 | 0.073810827 | 0.004590515 | 0.021461648 |
| PPIF | 7501.518325 | -0.20265019 | 0.078368047 | 0.007432127 | 0.031983632 |
| G6PD | 1112.694097 | 0.202619266 | 0.09400774 | 0.023230101 | 0.080001204 |
| HAUS1 | 1179.268583 | 0.202533534 | 0.085836427 | 0.013770466 | 0.05278069 |
| STK4 | 3477.391093 | 0.202511527 | 0.084098103 | 0.011961429 | 0.046944638 |
| SRRM1 | 1583.747075 | 0.202416943 | 0.08693554 | 0.014925698 | 0.05605472 |
| ADAR | 10389.89234 | -0.202339863 | 0.062067727 | 0.000865901 | 0.005246606 |
| RPS10 | 16337.22798 | -0.202253618 | 0.094208806 | 0.023896107 | 0.081812749 |
| MGAT1 | 6841.073284 | 0.202238437 | 0.089858352 | 0.018200269 | 0.065784607 |
| CLDND1 | 2019.142786 | 0.202210815 | 0.096376559 | 0.02671414 | 0.089406984 |
| FOXRED1 | 1653.691624 | -0.202203877 | 0.089896653 | 0.018330844 | 0.06609103 |

| | | | | | |
|---|---|---|---|---|---|
| TFDP1 | 7847.473883 | -0.202072867 | 0.06752767 | 0.002211785 | 0.011687969 |
| BRD8 | 911.0458253 | 0.202030578 | 0.091840283 | 0.020895056 | 0.073559602 |
| RPL15P5 | 1758.806371 | -0.201703382 | 0.09431202 | 0.024382245 | 0.083164729 |
| BUB3 | 5309.814202 | 0.201486562 | 0.068584076 | 0.002520603 | 0.013056917 |
| ACTR1A | 2006.61957 | 0.201365065 | 0.081142264 | 0.009841776 | 0.040287773 |
| TMEM230 | 3603.537238 | -0.201335219 | 0.076264192 | 0.006372186 | 0.02819752 |
| NUFIP2 | 4296.049523 | -0.20131865 | 0.097546987 | 0.029288981 | 0.0960419 |
| OPTN | 1280.049498 | 0.201095448 | 0.082123714 | 0.010842416 | 0.043472591 |
| ATP5MF | 4637.626272 | -0.201010089 | 0.076120783 | 0.006356785 | 0.028153567 |
| AMMECR1L | 802.5082717 | 0.200939836 | 0.091829686 | 0.021510177 | 0.075299423 |
| NDUFB9 | 5707.343868 | -0.200840349 | 0.080358539 | 0.009515732 | 0.039159681 |
| PMF1-BGLAP | 784.4730593 | -0.200721875 | 0.093941313 | 0.024565019 | 0.083689247 |
| LRRFIP1 | 3796.394 | 0.199922818 | 0.076029221 | 0.006515169 | 0.028693588 |
| DYNC1LI1 | 1495.443569 | -0.199828477 | 0.078780761 | 0.008578321 | 0.035873166 |
| B2M | 109201.7765 | 0.199808039 | 0.082558238 | 0.012197222 | 0.047690062 |
| KIF3B | 1150.104626 | 0.199403068 | 0.084413017 | 0.013779657 | 0.052792549 |
| CAPNS1 | 9768.786034 | 0.199394244 | 0.080566999 | 0.01015412 | 0.041296125 |
| MRRF | 1040.778933 | -0.199220915 | 0.088361266 | 0.018447575 | 0.066442734 |
| STXBP2 | 1309.846598 | 0.199077349 | 0.093884152 | 0.025571035 | 0.086352107 |
| RPS10-NUDT3 | 15509.14917 | -0.199047102 | 0.090239081 | 0.020649248 | 0.072768218 |
| TCF3 | 5453.695326 | 0.199039966 | 0.094273739 | 0.025775869 | 0.086874412 |
| ME2 | 4682.848622 | -0.198906915 | 0.076394727 | 0.007107187 | 0.030869113 |
| GSE1 | 2467.695437 | -0.198848219 | 0.091920344 | 0.023142049 | 0.079765582 |
| STK39 | 1356.600508 | -0.198847449 | 0.092944212 | 0.024530176 | 0.083586985 |
| GOSR2 | 1360.398421 | 0.19882188 | 0.089263568 | 0.019627756 | 0.069836418 |
| IGBP1 | 1860.961768 | -0.198690258 | 0.088873128 | 0.019329989 | 0.068998897 |
| SNORD3C | 165.9281431 | 0.198530432 | 0.41105068 | 0.002466545 | 0.01282418 |
| EXOC4 | 2498.22005 | -0.198515239 | 0.070570876 | 0.003801631 | 0.018297064 |
| TATDN2 | 1329.428151 | -0.198513736 | 0.083532355 | 0.01342387 | 0.051715496 |
| PSMD5 | 1438.930341 | 0.198499735 | 0.081216878 | 0.011155895 | 0.044502796 |
| CDV3 | 12222.47366 | -0.198499456 | 0.071245001 | 0.004132911 | 0.019645792 |
| CALCOCO1 | 1077.830792 | 0.198461327 | 0.096737059 | 0.030125965 | 0.098043852 |
| LOC105375124 | 3.092233291 | -0.198396954 | 0.402439954 | 0.011536167 | 0.045683009 |
| ARMC8 | 1035.998567 | 0.198385647 | 0.08550654 | 0.015453108 | 0.05765138 |
| RMND5A | 3554.243692 | 0.197931608 | 0.083307444 | 0.013176582 | 0.050932782 |
| HLA-DRB1 | 7105.573951 | -0.197288418 | 0.076648923 | 0.00781886 | 0.033341344 |
| DDX28 | 1142.420755 | -0.197234657 | 0.09457788 | 0.028223048 | 0.093235124 |
| TMEM39A | 1562.36724 | -0.197116238 | 0.081381071 | 0.011880946 | 0.046674759 |
| UFC1 | 5615.452191 | -0.19709308 | 0.065432445 | 0.002031172 | 0.010872892 |
| SRPK1 | 2777.163584 | -0.196924348 | 0.082004369 | 0.012576761 | 0.04895536 |
| WNK1 | 3007.518216 | -0.196691553 | 0.082863972 | 0.013496195 | 0.051936359 |
| RN7SL4P | 617.088509 | 0.196595737 | 0.414855036 | 1.53E-05 | 0.000149093 |
| FAM98A | 2627.523967 | -0.196567768 | 0.085497565 | 0.016516708 | 0.060687424 |
| PINK1-AS | 876.6835759 | -0.196546157 | 0.087211599 | 0.018588002 | 0.066837299 |
| DCUN1D5 | 1996.944417 | -0.19648361 | 0.073674469 | 0.005984155 | 0.026742609 |
| NCBP2 | 2558.065285 | 0.196375307 | 0.075938276 | 0.007440172 | 0.0320103 |
| EIF2AK1 | 5697.508183 | -0.196369965 | 0.063211769 | 0.001484055 | 0.008325035 |
| NBR1 | 2766.860998 | 0.196283836 | 0.081042229 | 0.011783833 | 0.046366881 |
| GAGE12G | 2717.061027 | -0.196059289 | 0.075653278 | 0.007430329 | 0.031983632 |
| RPA2 | 2379.285827 | 0.196030213 | 0.074700337 | 0.006694782 | 0.029357696 |

| | | | | | |
|---|---|---|---|---|---|
| GAGE12B | 2734.109067 | -0.195924473 | 0.0756643 | 0.00747335 | 0.032125158 |
| GAGE12F | 2734.109067 | -0.195924473 | 0.0756643 | 0.00747335 | 0.032125158 |
| RCSD1 | 7590.993836 | 0.195914267 | 0.064996277 | 0.001991677 | 0.010687903 |
| SNRPF | 1812.266409 | -0.195896659 | 0.080280994 | 0.011372308 | 0.045189136 |
| MRPL22 | 1711.326023 | -0.195750459 | 0.082556019 | 0.01369807 | 0.052573023 |
| IVNS1ABP | 913.3968598 | 0.195697747 | 0.088054073 | 0.020027863 | 0.070991137 |
| EPS15 | 1541.710122 | 0.195644439 | 0.087373653 | 0.019097557 | 0.06828668 |
| ADAM22 | 939.7737826 | -0.195362896 | 0.091415442 | 0.024950396 | 0.084702216 |
| RRP36 | 874.5912923 | 0.195317254 | 0.089648831 | 0.022508932 | 0.078254803 |
| COPS6 | 2507.18084 | -0.195295527 | 0.084659703 | 0.016306259 | 0.060118264 |
| SMARCD1 | 3722.213621 | 0.195208301 | 0.074061338 | 0.006499116 | 0.028636341 |
| ENO1 | 96453.02914 | 0.195161837 | 0.071409843 | 0.004832607 | 0.022448031 |
| SGK1 | 8100.544253 | 0.195153529 | 0.067321618 | 0.002912517 | 0.014712966 |
| MIR663A | 4.560096416 | 0.19510588 | 0.397313447 | 0.020326335 | 0.07184473 |
| SERF2-C15ORF63 | 2350.332771 | -0.194928588 | 0.084042874 | 0.015758164 | 0.058558824 |
| HLA-DQA2 | 1620.640269 | -0.194904203 | 0.085531433 | 0.017546571 | 0.063942333 |
| EXTL3 | 1228.169007 | -0.194825372 | 0.084539278 | 0.016360624 | 0.060267373 |
| RPL11 | 41322.19678 | -0.194815681 | 0.061123162 | 0.00113828 | 0.006645904 |
| TXNL1 | 4404.376601 | -0.194814132 | 0.068842785 | 0.003669354 | 0.017788969 |
| SECISBP2 | 1051.794206 | -0.194769894 | 0.089717876 | 0.023015261 | 0.079456792 |
| RPS10P19 | 10764.72424 | -0.19472221 | 0.083134285 | 0.014811057 | 0.05575719 |
| RPS4X | 72868.22404 | -0.194515833 | 0.066518339 | 0.002841457 | 0.014442473 |
| PSEN1 | 2822.372433 | 0.194417169 | 0.0702858 | 0.004401117 | 0.020693495 |
| JMJD6 | 865.2686833 | -0.194368685 | 0.088409796 | 0.02151585 | 0.075301125 |
| H2BC18 | 133.6032591 | 0.194276697 | 0.393930733 | 0.029008994 | 0.095358532 |
| MNS1 | 3.024161102 | -0.194275869 | 0.398874234 | 0.012567579 | 0.048939233 |
| BTF3 | 25341.49107 | -0.194264813 | 0.063821097 | 0.001914407 | 0.01035023 |
| MPC2 | 4652.661429 | -0.194135688 | 0.065899546 | 0.002531716 | 0.01309099 |
| TSR2 | 1391.585447 | 0.193849091 | 0.080964872 | 0.012820173 | 0.049684233 |
| FAM32A | 2413.842778 | 0.193811249 | 0.077127647 | 0.009174487 | 0.037899473 |
| GNS | 4534.640796 | -0.193404911 | 0.064715047 | 0.00220912 | 0.011684587 |
| ECI2 | 1611.738915 | -0.193345285 | 0.077367147 | 0.009708015 | 0.039809069 |
| CCT6A | 10320.67531 | -0.19331009 | 0.062994002 | 0.001698569 | 0.009355636 |
| TERF2 | 1020.640445 | -0.193255768 | 0.089483497 | 0.023670332 | 0.081232489 |
| WDR82 | 7370.666437 | -0.193213487 | 0.067218822 | 0.003203179 | 0.015974326 |
| COX5B | 4993.229351 | 0.19312722 | 0.07833444 | 0.010550053 | 0.042606171 |
| CBWD5 | 729.2968872 | -0.193058402 | 0.091550813 | 0.026928939 | 0.089937983 |
| XRCC5 | 18349.75117 | -0.192792007 | 0.067378444 | 0.003342035 | 0.016525989 |
| DNAJB12 | 1104.407711 | 0.192790798 | 0.085649077 | 0.018809882 | 0.067480988 |
| AASDHPPT | 5173.880411 | -0.192511718 | 0.091409227 | 0.027182014 | 0.090569059 |
| PRKD3 | 2585.927003 | 0.192499285 | 0.081125206 | 0.013614019 | 0.0522968 |
| SMARCB1 | 2639.782238 | 0.192397083 | 0.074311371 | 0.007552769 | 0.032382101 |
| DDHD2 | 1024.223766 | -0.192246609 | 0.092785756 | 0.029580568 | 0.096686805 |
| PEG10 | 8330.706661 | -0.192072936 | 0.079117763 | 0.01186809 | 0.04664542 |
| SELENOS | 2774.31704 | -0.191436838 | 0.08151763 | 0.014709445 | 0.055502175 |
| PPP6R1 | 2821.474068 | 0.19135258 | 0.083811474 | 0.01719713 | 0.062827575 |
| ATRN | 1360.528245 | -0.191285045 | 0.081102158 | 0.014322557 | 0.054367322 |
| CSTF2 | 963.8935026 | 0.191048003 | 0.089183377 | 0.024832215 | 0.084439314 |
| PABPN1 | 1094.204309 | 0.190961504 | 0.088060454 | 0.023312291 | 0.080194467 |
| LOC124900510 | 1903.346362 | 0.190884027 | 0.406818853 | 0.000245598 | 0.001753451 |

| | | | | | |
|---|---|---|---|---|---|
| USP16 | 2192.303345 | -0.190785664 | 0.07836834 | 0.011686961 | 0.046069451 |
| UQCRH | 9908.347694 | -0.190757183 | 0.070468482 | 0.005299955 | 0.024203409 |
| YWHAG | 11690.07482 | -0.190488046 | 0.07005419 | 0.005162023 | 0.023723661 |
| EPC1 | 1562.039936 | -0.190262212 | 0.081566844 | 0.015392511 | 0.057483744 |
| TNFAIP3 | 4052.17862 | 0.189967837 | 0.07118676 | 0.00597668 | 0.026722998 |
| NSD2 | 1356.234541 | 0.189886371 | 0.083109144 | 0.017323087 | 0.063221053 |
| DDX23 | 2661.862089 | 0.189811197 | 0.075106214 | 0.009078191 | 0.037591351 |
| CUEDC2 | 1605.548473 | -0.1893913 | 0.089628088 | 0.026991408 | 0.090077125 |
| TRAPPC10 | 1000.631239 | 0.189316276 | 0.090212858 | 0.027735027 | 0.092009093 |
| RPL31 | 25576.43242 | -0.18909844 | 0.077158755 | 0.011402484 | 0.045288262 |
| SF3B1 | 11863.16303 | -0.189013887 | 0.071554037 | 0.006596119 | 0.028998476 |
| ERGIC3 | 8289.537702 | -0.189012675 | 0.073222999 | 0.007793368 | 0.033256085 |
| DIDO1 | 1905.507353 | -0.188927304 | 0.079081029 | 0.013284125 | 0.051302673 |
| CDS2 | 1983.840414 | -0.188855524 | 0.080285307 | 0.014679033 | 0.055428873 |
| ATMIN | 1874.559885 | -0.188788106 | 0.077806315 | 0.01198755 | 0.047025679 |
| CORO1C | 3143.813043 | -0.188687599 | 0.075572176 | 0.009896689 | 0.040467758 |
| DPF2 | 1031.157029 | 0.188608099 | 0.086232828 | 0.022370733 | 0.077852488 |
| CCNE1 | 935.1849351 | -0.188527292 | 0.091202511 | 0.030135657 | 0.098056965 |
| ZPBP2 | 3.888780471 | 0.188446058 | 0.388965268 | 0.028620763 | 0.09427985 |
| CEBPZ | 3141.932335 | -0.188421619 | 0.074370902 | 0.008924769 | 0.037097988 |
| ZDHHC3 | 1979.403758 | -0.188367753 | 0.075038627 | 0.009553749 | 0.039288119 |
| MKRN1 | 2146.58187 | 0.188350643 | 0.071758013 | 0.007041415 | 0.030606473 |
| ARL6IP1 | 5689.831722 | -0.188092076 | 0.081544985 | 0.016504765 | 0.060669289 |
| SNRPD1 | 3417.549509 | -0.187954649 | 0.086420607 | 0.023233313 | 0.080001204 |
| ILRUN | 2548.652622 | 0.187895552 | 0.075395998 | 0.009928507 | 0.04054999 |
| KDM3B | 1517.402772 | 0.187809034 | 0.081997389 | 0.017187953 | 0.062807297 |
| GEMIN5 | 1486.048614 | -0.187701338 | 0.077756306 | 0.012464262 | 0.048600648 |
| GCN1 | 3337.324446 | -0.18721165 | 0.076618483 | 0.011533338 | 0.045683009 |
| RPRD1B | 1932.41718 | -0.186921569 | 0.082841541 | 0.01899425 | 0.067959385 |
| TYW3 | 1010.709844 | -0.186667363 | 0.089764005 | 0.029365333 | 0.096186338 |
| EMC7 | 3148.337448 | -0.186624083 | 0.073616039 | 0.008946768 | 0.037176718 |
| RNU1-1 | 5.400868021 | 0.186544258 | 0.391662451 | 0.015011633 | 0.056345376 |
| CAMK2N1 | 3.243708314 | 0.186471722 | 0.388497049 | 0.026345613 | 0.088450364 |
| SAR1A | 2693.287557 | -0.186288182 | 0.075213887 | 0.01054062 | 0.04258794 |
| SRI | 1238.403628 | -0.186259121 | 0.081849095 | 0.018092961 | 0.065465065 |
| CAT | 1098.809755 | 0.186209912 | 0.083525955 | 0.020257699 | 0.071650853 |
| BST2 | 4915.253031 | -0.186138123 | 0.075362099 | 0.010773232 | 0.043275356 |
| COA1 | 2919.14616 | -0.185633491 | 0.075746642 | 0.011344564 | 0.045129366 |
| KLHDC3 | 2847.202849 | 0.185466749 | 0.081594903 | 0.018173214 | 0.065714252 |
| RPL24 | 31733.85093 | -0.185038643 | 0.069795983 | 0.006430565 | 0.02839319 |
| THUMPD3 | 1105.769648 | -0.184591453 | 0.086801065 | 0.026677293 | 0.089321478 |
| GPX7 | 1354.175281 | -0.184373411 | 0.077862906 | 0.014208252 | 0.054009998 |
| CELF2 | 4627.299692 | 0.184200487 | 0.088483134 | 0.029303335 | 0.096052592 |
| ARFGAP3 | 1284.993821 | -0.18419557 | 0.08438568 | 0.023023813 | 0.079470474 |
| TGIF2 | 1989.271604 | -0.183941597 | 0.074010254 | 0.010419146 | 0.042189668 |
| VAPA | 5258.834763 | -0.18359119 | 0.064749893 | 0.003727608 | 0.017999996 |
| CBL | 1762.172402 | 0.183119618 | 0.078290393 | 0.015324544 | 0.057265494 |
| PRPF4 | 2596.132388 | -0.182574857 | 0.07109732 | 0.008236785 | 0.03469628 |
| ANAPC1 | 1924.81802 | -0.182454754 | 0.073713462 | 0.010691324 | 0.043038613 |
| AHCYL1 | 1892.20018 | 0.182225893 | 0.082900146 | 0.022121731 | 0.07709439 |

| | | | | | |
|---|---|---|---|---|---|
| DYNC1H1 | 4109.787554 | 0.182030766 | 0.081182456 | 0.019756776 | 0.070194521 |
| SETX | 2553.283965 | 0.181940303 | 0.085762572 | 0.026740539 | 0.089464049 |
| PDE6D | 970.5161508 | 0.181796223 | 0.087363907 | 0.029602791 | 0.096741184 |
| SF3B6 | 3377.273281 | -0.181256662 | 0.0838105 | 0.024354931 | 0.083104302 |
| ATOX1 | 1381.4678 | -0.180933719 | 0.084480885 | 0.025808261 | 0.086932826 |
| COX6B1 | 7491.369108 | 0.18049419 | 0.066167389 | 0.005142481 | 0.023640121 |
| NXPE3 | 3391.102445 | 0.18043913 | 0.085140384 | 0.027038882 | 0.090218173 |
| NELFCD | 3140.676261 | -0.180318036 | 0.072481128 | 0.010389869 | 0.042106566 |
| FLAD1 | 1412.990172 | -0.180017171 | 0.079591463 | 0.019077634 | 0.068229529 |
| LOC105370259 | 3.031943087 | 0.179931202 | 0.386146266 | 0.011534397 | 0.045683009 |
| NUP205 | 3993.964913 | -0.179749634 | 0.067384477 | 0.006216044 | 0.027607906 |
| GAGE1 | 2130.005616 | -0.17963853 | 0.078479212 | 0.017793175 | 0.064623408 |
| EXOSC8 | 1161.557065 | 0.179549176 | 0.08516151 | 0.027847498 | 0.092364514 |
| LOC124905321 | 5.178280207 | 0.179531816 | 0.384600006 | 0.01765786 | 0.064212761 |
| TFG | 2642.946739 | -0.17930352 | 0.076352524 | 0.015227043 | 0.056964074 |
| NAP1L1 | 32611.48732 | -0.179302088 | 0.078611538 | 0.019429115 | 0.069257716 |
| HK2 | 4396.425142 | -0.17929583 | 0.068742085 | 0.007416775 | 0.031941381 |
| RNA5-8SN4 | 952.5250996 | 0.179280715 | 0.394054637 | 0.000255262 | 0.001811786 |
| RNA5-8SN1 | 954.396968 | 0.179215645 | 0.393993226 | 0.000253804 | 0.001804979 |
| RNA5-8SN2 | 954.396968 | 0.179215645 | 0.393993226 | 0.000253804 | 0.001804979 |
| RNA5-8SN3 | 954.396968 | 0.179215645 | 0.393993226 | 0.000253804 | 0.001804979 |
| RNA5-8SN5 | 953.9062094 | 0.179210806 | 0.393986643 | 0.000254344 | 0.001807338 |
| ICE1 | 1989.465967 | -0.179007383 | 0.085694745 | 0.02915781 | 0.095738679 |
| PAIP2 | 5100.900847 | -0.178738776 | 0.077481609 | 0.017034373 | 0.062318401 |
| IFNGR2 | 2065.032047 | 0.178576228 | 0.078438145 | 0.01825901 | 0.065915714 |
| HIF1AN | 1133.025788 | 0.178576056 | 0.08618198 | 0.030488208 | 0.098888238 |
| XBP1 | 3425.621655 | -0.17848159 | 0.078024474 | 0.017910432 | 0.064999611 |
| SMG7 | 1304.424074 | 0.17809087 | 0.085584481 | 0.029935216 | 0.097595768 |
| MICU1 | 1619.927961 | -0.17800154 | 0.08514801 | 0.029426671 | 0.096311029 |
| POM121 | 2573.846736 | -0.177886557 | 0.085067176 | 0.02941571 | 0.09630407 |
| DTL | 2526.278854 | -0.177611653 | 0.079750156 | 0.020935914 | 0.073643555 |
| MESD | 3059.681463 | -0.177576862 | 0.068728197 | 0.007955741 | 0.033758775 |
| CASP2 | 1806.00868 | 0.177383541 | 0.0768728 | 0.016932953 | 0.061993225 |
| ASNSD1 | 2228.518717 | -0.177354999 | 0.083332229 | 0.026859991 | 0.089776967 |
| CUL1 | 2557.691218 | -0.177308474 | 0.069784659 | 0.009005234 | 0.037333883 |
| SHMT1 | 3438.78246 | 0.177091349 | 0.072561275 | 0.011833103 | 0.046539595 |
| AMZ2 | 1681.338475 | -0.176790331 | 0.080083478 | 0.02208338 | 0.076991725 |
| BLMH | 4544.132923 | -0.176775043 | 0.06508834 | 0.005408552 | 0.024634375 |
| PTPN2 | 1453.106355 | -0.176632966 | 0.07746619 | 0.018284301 | 0.065964425 |
| MIR3648-2 | 35.01705876 | 0.176513659 | 0.387702043 | 0.003648451 | 0.017722364 |
| UBA1 | 8944.171441 | 0.17628515 | 0.079516941 | 0.021456547 | 0.075154335 |
| RNU1-2 | 4.174093363 | 0.176262463 | 0.385348872 | 0.006583076 | 0.028948481 |
| ATP5MJ | 4303.651657 | -0.176215058 | 0.07293052 | 0.012845818 | 0.049759457 |
| TXNRD1 | 7403.479794 | -0.175843939 | 0.064096734 | 0.004992897 | 0.023044241 |
| RAB1B | 2499.988218 | 0.175772539 | 0.08266643 | 0.02695232 | 0.089981363 |
| NAXE | 3732.346085 | -0.175470415 | 0.085406866 | 0.030721384 | 0.099495458 |
| CCDC47 | 6614.641003 | -0.175245922 | 0.077317572 | 0.01913626 | 0.068396825 |
| RHOA | 10706.74212 | -0.174929657 | 0.06214003 | 0.004033553 | 0.019231052 |
| VWA8 | 1721.390412 | -0.174912169 | 0.07616559 | 0.0175962 | 0.064069258 |
| HMGN2P41 | 2488.786571 | 0.174849912 | 0.074109659 | 0.01476021 | 0.055612062 |

| | | | | | |
|---|---|---|---|---|---|
| FOXK2 | 1977.168315 | 0.174823029 | 0.078009181 | 0.020236287 | 0.071589741 |
| MPHOSPH10 | 1757.855763 | -0.17481568 | 0.078643185 | 0.021299967 | 0.074711673 |
| H2AC11 | 79.83081824 | 0.174600816 | 0.383416395 | 0.009507539 | 0.039142129 |
| ARNT | 8281.620319 | -0.174543585 | 0.061529182 | 0.003770912 | 0.018164362 |
| APEH | 2163.102571 | -0.174241712 | 0.08086644 | 0.025327565 | 0.085764067 |
| NCAPD2 | 4547.550713 | 0.174206469 | 0.074479943 | 0.01571712 | 0.058443926 |
| POU2AF1 | 6290.681511 | -0.17413558 | 0.078282606 | 0.021333928 | 0.074800492 |
| ANP32A | 4527.47656 | 0.173858091 | 0.080741434 | 0.025269963 | 0.085602494 |
| CNPY2 | 2483.172582 | -0.17379479 | 0.081809882 | 0.027401246 | 0.091128747 |
| ZNF317 | 1538.738676 | 0.173725109 | 0.076839037 | 0.019266689 | 0.06882039 |
| CLIC1 | 13713.81203 | 0.173571726 | 0.063116131 | 0.00488923 | 0.022668488 |
| BAG6 | 3688.63418 | 0.173397875 | 0.080265344 | 0.024668399 | 0.083991874 |
| ASNS | 3656.533744 | 0.173203304 | 0.073073453 | 0.014496185 | 0.054867788 |
| NPEPPS | 2496.047687 | -0.173075732 | 0.069391818 | 0.010379059 | 0.042073342 |
| NOP2 | 2002.340801 | -0.172856217 | 0.08194044 | 0.028452255 | 0.093849085 |
| FASTKD2 | 2916.060639 | -0.172793079 | 0.078857837 | 0.023310912 | 0.080194467 |
| RN7SKP80 | 468.1530191 | 0.172594185 | 0.3861908 | 0.00103218 | 0.00610668 |
| UQCRC1 | 5377.558902 | -0.172271134 | 0.07891319 | 0.023725029 | 0.08138794 |
| GNB1 | 8607.70409 | -0.172171491 | 0.067287624 | 0.008814813 | 0.036702602 |
| TRBJ2-2P | 3.413665147 | 0.172088631 | 0.376452026 | 0.027169383 | 0.090569059 |
| SMG8 | 1916.19281 | -0.171812547 | 0.07440036 | 0.01721127 | 0.062865968 |
| LOC124900412 | 3.160664971 | 0.171650795 | 0.378949611 | 0.012053665 | 0.047184644 |
| GMPS | 2452.014752 | 0.171524389 | 0.073104518 | 0.015476767 | 0.057711277 |
| PSMD3 | 5613.315236 | 0.171284039 | 0.081233963 | 0.028430234 | 0.093812186 |
| CZ1P-ASNS | 3658.453605 | 0.171193079 | 0.07335374 | 0.01605398 | 0.059428555 |
| DNMT1 | 2787.591423 | 0.17110585 | 0.07489003 | 0.018198857 | 0.065784607 |
| CFL1 | 16912.63397 | 0.170759746 | 0.068229705 | 0.0101138 | 0.041161147 |
| FKBP4 | 6899.617781 | -0.170747503 | 0.067979061 | 0.009944286 | 0.040585717 |
| MAX | 1696.431782 | 0.170658802 | 0.07892517 | 0.024882059 | 0.084553102 |
| NUP155 | 1860.413649 | 0.170515105 | 0.074153749 | 0.017594407 | 0.064069258 |
| NRBP1 | 2799.258052 | 0.170375032 | 0.076980242 | 0.021905706 | 0.076449235 |
| EIF2A | 3657.045854 | -0.17036505 | 0.073109159 | 0.016321901 | 0.060163125 |
| HNRNPC | 19846.83776 | -0.170247277 | 0.066071319 | 0.008311824 | 0.034961343 |
| NDUFA6 | 2857.213706 | 0.170010285 | 0.071461567 | 0.014212829 | 0.054009998 |
| CMSS1 | 2728.575011 | -0.169994921 | 0.074242357 | 0.018169363 | 0.065714049 |
| TPM4 | 5418.636529 | 0.169879224 | 0.072597402 | 0.015837924 | 0.058779712 |
| WBP11 | 3879.433989 | -0.169644577 | 0.070996 | 0.013959727 | 0.053318335 |
| OXA1L | 3086.617595 | -0.169513462 | 0.080248572 | 0.02862096 | 0.09427985 |
| FTH1 | 12295.94536 | 0.169440211 | 0.078719589 | 0.02561288 | 0.086442847 |
| EEF1D | 9231.027762 | -0.169289462 | 0.074376057 | 0.018986792 | 0.067946739 |
| MSL1 | 1394.827568 | -0.169221031 | 0.081110983 | 0.030385726 | 0.098592771 |
| PDAP1 | 2365.437965 | -0.169196837 | 0.072193461 | 0.015824646 | 0.058755443 |
| VPS25 | 2051.32934 | -0.168951504 | 0.07852092 | 0.025925034 | 0.087292207 |
| TIMM44 | 1814.304503 | -0.16888697 | 0.078366273 | 0.025678229 | 0.086586428 |
| EML4 | 3795.047714 | 0.168812311 | 0.076908213 | 0.022896992 | 0.079190576 |
| TMEM199 | 1155.872541 | 0.168520055 | 0.080639957 | 0.029917282 | 0.097584776 |
| SIN3A | 1666.749007 | 0.168430972 | 0.075091046 | 0.020480651 | 0.072262312 |
| ERGIC1 | 6887.253614 | 0.168138969 | 0.071810971 | 0.015800476 | 0.058703211 |
| TRIP12 | 4883.310182 | 0.168099935 | 0.069754823 | 0.013172504 | 0.05092839 |
| EMC4 | 2329.232393 | -0.168053406 | 0.075459151 | 0.021473488 | 0.075198461 |

| | | | | | |
|---|---|---|---|---|---|
| SON | 9553.523157 | -0.167890676 | 0.065455208 | 0.008809023 | 0.036687312 |
| SRSF7 | 9348.03934 | -0.167883895 | 0.067893311 | 0.011146432 | 0.044485541 |
| SEC61A1 | 21324.92198 | -0.167648965 | 0.067902126 | 0.011298363 | 0.044977875 |
| NUDC | 2735.429851 | 0.166989933 | 0.071326931 | 0.01596413 | 0.059159178 |
| ANAPC7 | 1288.455009 | 0.166814653 | 0.079754093 | 0.029961863 | 0.097619864 |
| RPL4P4 | 6199.045796 | -0.166720334 | 0.062751333 | 0.006489566 | 0.028609933 |
| ATXN10 | 2858.786448 | -0.166520199 | 0.071896002 | 0.01711857 | 0.062598519 |
| CPD | 3345.592376 | -0.166161222 | 0.076883838 | 0.025445921 | 0.086047063 |
| AKAP13 | 1609.596562 | 0.166160744 | 0.079064662 | 0.029320553 | 0.096090844 |
| RECQL | 2747.313793 | -0.166102129 | 0.07928845 | 0.029936027 | 0.097595768 |
| KHSRP | 7113.451323 | 0.165718462 | 0.070494022 | 0.015516966 | 0.057823823 |
| MFN2 | 2885.751676 | 0.165582728 | 0.075286298 | 0.023060712 | 0.079581976 |
| TOMM40L | 1403.913976 | 0.165543143 | 0.079161605 | 0.030105621 | 0.098014485 |
| RPL37 | 59351.99298 | -0.165251025 | 0.068472465 | 0.014131531 | 0.053760045 |
| PTCD3 | 3861.949822 | -0.165093911 | 0.067308898 | 0.011861806 | 0.046631305 |
| SPATS2 | 1777.779251 | -0.164685475 | 0.074329265 | 0.02224608 | 0.077480968 |
| S100A10 | 2943.072059 | -0.164460021 | 0.071544809 | 0.017982171 | 0.065173138 |
| UBE2I | 2594.347065 | -0.16434884 | 0.071422608 | 0.01794586 | 0.065068772 |
| EIF3I | 6759.228608 | -0.164237387 | 0.068379415 | 0.013752332 | 0.052734528 |
| SUN1 | 2898.972997 | -0.16420987 | 0.070736372 | 0.01697033 | 0.062116932 |
| PEA15 | 2948.359773 | -0.164170971 | 0.069399627 | 0.015078582 | 0.056530808 |
| LIMD1 | 1459.728885 | -0.164011694 | 0.07670663 | 0.027046199 | 0.090225203 |
| SPTLC2 | 1266.072593 | -0.163709476 | 0.078510052 | 0.030882718 | 0.099860067 |
| ESD | 2973.543669 | -0.163625274 | 0.07332756 | 0.021511015 | 0.075299423 |
| FOCAD | 1689.952913 | -0.163492861 | 0.077628114 | 0.029143265 | 0.095709068 |
| PLP2 | 6928.356686 | 0.163067696 | 0.076342515 | 0.027207904 | 0.090572718 |
| TSPYL5 | 1937.128653 | -0.162663323 | 0.073581454 | 0.022682267 | 0.078652146 |
| ATP6V0E1 | 5190.012063 | -0.162613321 | 0.068883832 | 0.01546098 | 0.057664825 |
| ERAP1 | 4565.323821 | -0.162552954 | 0.066927266 | 0.012689843 | 0.049293408 |
| DCTD | 2181.804657 | 0.161464106 | 0.07675929 | 0.029542878 | 0.096600079 |
| UBE2Q1 | 3738.14063 | -0.161142729 | 0.067763497 | 0.014762872 | 0.055612062 |
| ICMT | 2756.337433 | 0.161083657 | 0.068313154 | 0.015454046 | 0.05765138 |
| PSMA2 | 8571.067075 | -0.161014246 | 0.068605879 | 0.015012853 | 0.056345376 |
| DNAJA1 | 7952.115927 | -0.160763389 | 0.070377008 | 0.0187773 | 0.067378057 |
| ISCU | 2808.600563 | -0.160250181 | 0.069958078 | 0.018583179 | 0.066833833 |
| DDOST | 14272.40382 | -0.160176493 | 0.076219711 | 0.029948831 | 0.097595768 |
| NOP56 | 5642.697516 | -0.159745956 | 0.074235189 | 0.026658066 | 0.089274361 |
| ECPAS | 2512.810951 | -0.159697435 | 0.071249579 | 0.021111925 | 0.074172288 |
| NACA | 34708.17268 | -0.159197189 | 0.065873535 | 0.013324734 | 0.051436563 |
| PRR13 | 2676.847201 | 0.159012769 | 0.072137039 | 0.023205801 | 0.079955205 |
| PRCP | 3397.625636 | -0.158564035 | 0.074382992 | 0.027900544 | 0.092469612 |
| TKT | 11859.58936 | -0.158495588 | 0.07149971 | 0.022527489 | 0.078273677 |
| RN7SK | 3036.681396 | 0.158290045 | 0.374109043 | 0.000476722 | 0.003129108 |
| RPL12 | 42849.98504 | -0.157974127 | 0.061213312 | 0.007613893 | 0.032603806 |
| IARS1 | 8547.671056 | -0.157790202 | 0.063036281 | 0.010482117 | 0.042391131 |
| PSMD1 | 7325.054769 | -0.157736077 | 0.0625714 | 0.010281269 | 0.041744605 |
| ESYT1 | 7838.17608 | 0.157466211 | 0.07405197 | 0.02817662 | 0.093117277 |
| ERAP2 | 3819.089324 | 0.157277389 | 0.072876022 | 0.026044233 | 0.087608387 |
| RNA18SN2 | 86634.54653 | 0.156746488 | 0.373500098 | 0.000107349 | 0.000846273 |
| RNA18SN3 | 86634.82812 | 0.156746032 | 0.373499706 | 0.000107349 | 0.000846273 |

| | | | | | |
|---|---|---|---|---|---|
| RNA18SN4 | 86627.3309 | 0.156737718 | 0.373492789 | 0.000107295 | 0.000846273 |
| ACO2 | 6461.376736 | 0.156731857 | 0.070586528 | 0.022314638 | 0.077672882 |
| RNA18SN1 | 86158.25241 | 0.156674937 | 0.373438737 | 0.000107417 | 0.000846284 |
| RNA18SN5 | 86138.74609 | 0.156657871 | 0.373424178 | 0.000107366 | 0.000846273 |
| SLC25A4 | 2356.621829 | 0.155829476 | 0.072938014 | 0.027580472 | 0.091619228 |
| CCNY | 2056.40632 | 0.154836952 | 0.073679092 | 0.030116555 | 0.09803165 |
| SNORA73A | 262.3571146 | 0.154536838 | 0.363847543 | 0.02624383 | 0.088142826 |
| PSMC5 | 5546.74287 | 0.154327961 | 0.065972923 | 0.016424139 | 0.060449906 |
| RNU1-4 | 3.624333137 | 0.154095095 | 0.365705289 | 0.011238811 | 0.044802599 |
| SMAD2 | 2641.695681 | 0.153289062 | 0.071448904 | 0.027214246 | 0.090576418 |
| FXR1 | 3357.286136 | 0.153015536 | 0.07150901 | 0.027387883 | 0.091113302 |
| H1-3 | 33.46418672 | 0.152849104 | 0.36431593 | 0.016674512 | 0.061137523 |
| RNU1-3 | 3.792544837 | 0.152696307 | 0.362301442 | 0.027089245 | 0.090333999 |
| HADHA | 9050.620758 | -0.152329402 | 0.062338787 | 0.012528901 | 0.048810571 |
| RPL38 | 16119.79857 | -0.151576965 | 0.065275234 | 0.018028514 | 0.065313761 |
| PPIA | 48335.62361 | -0.151008156 | 0.061622992 | 0.012327946 | 0.048168557 |
| H2BC7 | 34.39744264 | 0.150629804 | 0.363259658 | 0.013977821 | 0.053327588 |
| SCARNA2 | 45.68333326 | 0.150280392 | 0.367663033 | 0.00036233 | 0.00245961 |
| RPS11 | 68031.72492 | -0.150064021 | 0.067539653 | 0.023237631 | 0.080001204 |
| HSPA9 | 21436.94335 | -0.149584037 | 0.059469855 | 0.010379241 | 0.042073342 |
| PSME4 | 2891.129924 | -0.148607268 | 0.070662274 | 0.030614407 | 0.09926038 |
| RUVBL1 | 3607.805132 | -0.148069594 | 0.068011469 | 0.025501502 | 0.086201348 |
| PCBP2 | 16048.54765 | 0.147720975 | 0.05981277 | 0.011115791 | 0.04438371 |
| HLA-DRA | 18753.67921 | -0.14709729 | 0.06430153 | 0.018151561 | 0.065663376 |
| ATP5F1B | 59861.91626 | 0.147079872 | 0.069834582 | 0.029904796 | 0.097562426 |
| TMEM248 | 3698.802672 | -0.146630967 | 0.068951559 | 0.029366355 | 0.096186338 |
| NSUN2 | 4105.416344 | -0.145810329 | 0.066489304 | 0.024683406 | 0.084026447 |
| TPD52L2 | 2772.224133 | -0.145719904 | 0.068029283 | 0.028561588 | 0.094191775 |
| CDC6 | 4830.242245 | -0.14520608 | 0.064073657 | 0.020489159 | 0.072277605 |
| SNORD15B | 28.81466478 | 0.144471352 | 0.360772001 | 0.00553573 | 0.025094825 |
| ALG9 | 5980.872804 | -0.14269594 | 0.065227709 | 0.025112839 | 0.085175576 |
| HMGN2 | 8002.452209 | 0.142487506 | 0.064250396 | 0.02313887 | 0.079765582 |
| ACAT1 | 6820.874751 | -0.142278022 | 0.065553496 | 0.02787901 | 0.092451325 |
| C1QBP | 15157.67204 | -0.138878304 | 0.061383347 | 0.020308159 | 0.071799997 |
| RPL19 | 80650.76976 | -0.137974524 | 0.061922907 | 0.02294212 | 0.079288124 |
| RMRP | 1631.279453 | 0.137636338 | 0.357944616 | 0.000459264 | 0.003034364 |
| HNRNPK | 25166.42451 | -0.135959418 | 0.064279047 | 0.030732895 | 0.099514127 |
| H4C5 | 86.70549995 | 0.13578519 | 0.353114078 | 0.015569719 | 0.057958848 |
| SF3B3 | 8931.940848 | -0.135610216 | 0.063386825 | 0.028865449 | 0.095024695 |
| PCBP1 | 14859.14959 | -0.135315231 | 0.060990076 | 0.023591803 | 0.081027158 |
| H1-5 | 87.85816769 | 0.134067082 | 0.354552918 | 0.002267206 | 0.011940756 |
| PPIAP22 | 15732.49267 | -0.133701928 | 0.062627467 | 0.029651333 | 0.096863258 |
| RPPH1 | 954.976595 | 0.127875322 | 0.351169009 | 0.0005789 | 0.003710673 |
| LOC124904619 | 183.7958474 | 0.12701292 | 0.35086481 | 0.000256384 | 0.001817063 |
| H3C7 | 27.14277882 | 0.124344795 | 0.346092445 | 0.01646221 | 0.060564285 |
| H2AC21 | 22.62447242 | 0.123179616 | 0.347938532 | 0.001289136 | 0.007353088 |
| H2AC12 | 43.41068933 | 0.123022178 | 0.34773587 | 0.001393536 | 0.00788108 |
| H3C12 | 28.3796924 | 0.121094073 | 0.344897125 | 0.010950114 | 0.043843497 |
| LOC124905331 | 43.07914039 | 0.10718493 | 0.337561887 | 0.010088196 | 0.041076248 |
| H3C2 | 81.92944956 | 0.103097215 | 0.336495367 | 0.00159624 | 0.008873864 |

| | | | | | |
|---|---|---|---|---|---|
| H2BC10 | 37.81222801 | 0.103018903 | 0.33572467 | 0.007716373 | 0.032977402 |
| H2BC3 | 15.15847266 | 0.102201881 | 0.335428481 | 0.006724199 | 0.029456856 |
| SCARNA7 | 9.942884 | 0.091217049 | 0.330509452 | 0.0051688 | 0.02373658 |
| SCARNA5 | 10.3068922 | 0.088501443 | 0.329694692 | 0.001280183 | 0.007314062 |
| H4C2 | 43.60461936 | 0.081700524 | 0.326342412 | 0.010674148 | 0.042997029 |
| SNORD133 | 29.24093075 | 0.073158157 | 0.323711673 | 0.001173474 | 0.006810374 |
| RNU4-1 | 82.488933 | 0.068773636 | 0.321997265 | 0.005447759 | 0.02474786 |
| RNU4-2 | 82.70711221 | 0.047016177 | 0.316037661 | 0.007017219 | 0.030530746 |

**Supplementary Table S3**

| | AMO-1 | | | BM-MNC | | |
|---|---|---|---|---|---|---|
| | log2FoldChange | pvalue | padj | log2FoldChange | pvalue | padj |
| GBP4 | 6.224469149 | 5.90E-77 | 3.00E-74 | 0.725850105 | 1.35E-157 | 2.88E-153 |
| RGS9 | 5.251009442 | 5.64E-30 | 6.51E-28 | 3.376322132 | 3.33E-279 | 7.11E-275 |
| CLIC2 | -4.76969591 | 1.53E-72 | 7.38E-70 | -1.20567701 | 2.06E-121 | 4.41E-117 |
| MAP2 | 3.933701071 | 1.43E-06 | 1.74E-05 | 0.259649733 | 1.61E-11 | 3.44E-07 |
| TNFSF9 | -3.712068328 | 1.12E-211 | 3.23E-208 | -1.192449855 | 1.95E-11 | 4.17E-07 |
| TCEAL9 | -3.527944821 | 3.67E-07 | 5.05E-06 | -1.187320406 | 4.64E-54 | 9.92E-50 |
| EPSTI1 | 3.510372488 | 8.34E-11 | 2.01E-09 | 0.994251705 | 1.01E-12 | 2.16E-08 |
| LY96 | -2.913856489 | 7.21E-29 | 7.61E-27 | -1.565670048 | 0 | 0 |
| AREG | -2.835708899 | 3.36E-05 | 0.000301899 | -3.813613138 | 0 | 0 |
| RGS2 | -2.689502122 | 1.00E-104 | 8.69E-102 | -0.601916592 | 8.16E-51 | 1.74E-46 |
| APOL3 | 2.689404919 | 1.11E-85 | 7.09E-83 | 3.309008404 | 0 | 0 |
| NR4A1 | -2.546387393 | 7.83E-87 | 5.65E-84 | -1.004626805 | 1.55E-178 | 3.32E-174 |
| RNF43 | -2.476777415 | 8.53E-08 | 1.31E-06 | -1.848085545 | 5.80E-08 | 0.001238983 |
| NR4A2 | -2.456137346 | 4.64E-46 | 1.11E-43 | -0.494141535 | 7.30E-76 | 1.56E-71 |
| CYSLTR1 | -2.339376787 | 2.86E-27 | 2.81E-25 | -0.731639782 | 4.90E-58 | 1.05E-53 |
| RNF144B | 2.209670383 | 0.000189098 | 0.001390492 | 0.681272897 | 1.16E-49 | 2.49E-45 |
| ATF3 | -2.073807503 | 2.36E-152 | 4.53E-149 | -0.451269471 | 5.35E-288 | 1.14E-283 |
| GATA3 | 1.956167325 | 3.82E-06 | 4.21E-05 | 1.387247794 | 1.66E-43 | 3.54E-39 |
| DUSP26 | -1.745926356 | 6.98E-05 | 0.000579125 | -3.159885614 | 3.67E-105 | 7.83E-101 |
| IER5 | -1.509266925 | 3.02E-35 | 4.43E-33 | -0.679708655 | 0 | 0 |
| FOS | -1.475895341 | 2.75E-35 | 4.11E-33 | -0.797007171 | 0 | 0 |
| KMT2E-AS1 | -1.442585046 | 3.27E-06 | 3.67E-05 | -1.480924046 | 2.20E-152 | 4.70E-148 |
| DUSP5 | -1.281344761 | 2.85E-52 | 8.22E-50 | -1.597128069 | 0 | 0 |
| JUN | -1.235457871 | 9.61E-47 | 2.41E-44 | -2.86322098 | 0 | 0 |
| TXNIP | -1.196427569 | 2.18E-65 | 8.99E-63 | -1.197487688 | 0 | 0 |
| MOB3B | 1.167674385 | 0.008979153 | 0.037252513 | 1.822037765 | 6.11E-95 | 1.31E-90 |
| CXCR3 | 1.150463326 | 1.16E-07 | 1.76E-06 | 2.032899187 | 1.58E-24 | 3.37E-20 |
| HOPX | 1.088146018 | 0.000183359 | 0.001354616 | 1.685368902 | 0 | 0 |
| CCDC141 | 1.077406261 | 3.50E-17 | 1.70E-15 | 2.31371171 | 6.42E-106 | 1.37E-101 |
| PFN2 | -1.058772684 | 2.82E-09 | 5.53E-08 | -1.518772448 | 4.83E-70 | 1.03E-65 |
| APOBEC3F | 1.046862372 | 2.63E-21 | 1.80E-19 | 0.658999765 | 7.33E-29 | 1.56E-24 |
| HSPA1B | -0.987031042 | 4.91E-17 | 2.36E-15 | -3.589401052 | 0 | 0 |
| TCF4 | 0.984712743 | 9.01E-23 | 6.84E-21 | 0.381815775 | 0 | 0 |
| HOMER1 | -0.962762045 | 1.99E-10 | 4.54E-09 | -0.766265678 | 7.72E-49 | 1.65E-44 |
| DDIT4 | -0.952586571 | 1.14E-22 | 8.54E-21 | -1.705858436 | 0 | 0 |
| ZNF227 | 0.948909722 | 1.20E-11 | 3.32E-10 | 0.295773451 | 2.95E-21 | 6.30E-17 |
| ARL4C | 0.936102265 | 1.34E-45 | 3.09E-43 | 0.341665125 | 5.17E-152 | 1.10E-147 |
| LINC01801 | 0.902833594 | 0.008471695 | 0.035513032 | 1.337291083 | 1.14E-07 | 0.002427537 |
| DEPDC1 | 0.895368693 | 4.02E-11 | 1.02E-09 | 3.002123388 | 0 | 0 |
| UBE2L6 | 0.891036147 | 1.49E-30 | 1.79E-28 | 0.899486838 | 3.42E-258 | 7.31E-254 |
| NCAM1 | -0.890481405 | 1.38E-17 | 6.95E-16 | -0.872196021 | 5.61E-178 | 1.20E-173 |
| ABCG1 | 0.889831304 | 8.09E-11 | 1.96E-09 | 2.456787824 | 4.47E-206 | 9.55E-202 |
| CD180 | -0.882939028 | 2.34E-13 | 8.04E-12 | -0.810733678 | 3.00E-69 | 6.41E-65 |
| CTNNA2 | 0.882146697 | 1.69E-09 | 3.43E-08 | 1.601255931 | 1.77E-75 | 3.77E-71 |
| SEL1L3 | 0.832923854 | 6.53E-10 | 1.41E-08 | 0.508172254 | 4.94065645841247e-323 | 1.05517599982315e-318 |
| SQSTM1 | -0.831196598 | 9.84E-19 | 5.37E-17 | -0.867244032 | 0 | 0 |
| PNRC1 | -0.83080935 | 2.00E-19 | 1.18E-17 | -0.358745578 | 0 | 0 |
| SAMD9L | 0.823471815 | 1.55E-17 | 7.79E-16 | 0.718269431 | 0 | 0 |
| BRD2 | -0.823212474 | 5.13E-36 | 8.45E-34 | -0.390686753 | 8.25E-194 | 1.76E-189 |
| GSN | 0.810974397 | 6.44E-05 | 0.000539967 | 2.115799411 | 1.41E-45 | 3.01E-41 |
| KIF20A | 0.803767293 | 3.39E-17 | 1.65E-15 | 2.093690288 | 1.28E-105 | 2.74E-101 |
| SDC1 | -0.784404833 | 5.13E-23 | 3.95E-21 | -0.836448893 | 7.17E-262 | 1.53E-257 |
| TBXAS1 | -0.780620491 | 0.004065719 | 0.019363602 | -0.443369748 | 5.79E-45 | 1.24E-40 |
| NME7 | -0.779369046 | 1.23E-06 | 1.52E-05 | -0.342373301 | 1.31E-29 | 2.79E-25 |
| CKAP2L | 0.7586559 | 4.15E-11 | 1.05E-09 | 2.287351814 | 1.54E-234 | 3.29E-230 |
| KIF3C | 0.757467029 | 2.24E-07 | 3.21E-06 | 0.279181038 | 2.07E-21 | 4.42E-17 |
| PTPN18 | 0.753411944 | 1.81E-15 | 7.72E-14 | 0.732420305 | 3.61E-151 | 7.70E-147 |
| LINC02551 | 0.753231632 | 0.00292272 | 0.014747302 | 1.577747691 | 4.92E-247 | 1.05E-242 |
| COPS4 | -0.733143356 | 6.19E-15 | 2.50E-13 | -0.250987605 | 1.53E-14 | 3.27E-10 |
| ADAM28 | 0.733023471 | 1.37E-07 | 2.04E-06 | 2.202429331 | 0 | 0 |

| | | | | | | |
|---|---|---|---|---|---|---|
| IQSEC1 | -0.732059053 | 3.67E-13 | 1.23E-11 | -0.676724714 | 8.88E-49 | 1.90E-44 |
| ADGRG5 | 0.719570457 | 5.61E-10 | 1.22E-08 | 1.640460094 | 6.43E-274 | 1.37E-269 |
| SYTL3 | -0.716324898 | 2.09E-06 | 2.46E-05 | -1.308557033 | 7.35E-11 | 1.57E-06 |
| WNT10A | -0.703783248 | 1.05E-08 | 1.87E-07 | -0.930773276 | 8.24E-32 | 1.76E-27 |
| FAM111A | 0.70092349 | 2.57E-09 | 5.08E-08 | 1.164817089 | 0 | 0 |
| KLRF1 | 0.685095606 | 8.08E-05 | 0.000658136 | 0.625650228 | 1.23E-11 | 2.63E-07 |
| IL6R | -0.668448401 | 1.02E-15 | 4.49E-14 | -2.223770292 | 0 | 0 |
| CLCN4 | 0.66284233 | 1.46E-09 | 3.00E-08 | 1.419365805 | 7.50E-65 | 1.60E-60 |
| DNAJB9 | -0.66264028 | 9.68E-07 | 1.22E-05 | -0.782020261 | 0 | 0 |
| ESAM | 0.660396618 | 0.009477321 | 0.039029434 | 0.487915129 | 5.05E-22 | 1.08E-17 |
| ANLN | 0.65677174 | 4.11E-09 | 7.86E-08 | 1.704160244 | 3.59E-112 | 7.67E-108 |
| PIK3AP1 | 0.656144456 | 1.28E-10 | 3.00E-09 | 2.26692331 | 0 | 0 |
| TROAP | 0.656024017 | 1.51E-10 | 3.51E-09 | 1.843016427 | 8.26E-155 | 1.77E-150 |
| JRK | 0.64263426 | 7.70E-07 | 9.93E-06 | 0.327524911 | 6.31E-82 | 1.35E-77 |
| CENPA | 0.627684945 | 1.30E-10 | 3.03E-09 | 2.574646706 | 4.29E-208 | 9.16E-204 |
| GREB1 | -0.622104585 | 0.000203895 | 0.001485424 | -3.846689194 | 1.17E-34 | 2.50E-30 |
| SLC19A2 | -0.622103449 | 1.39E-05 | 0.000135949 | -0.781931884 | 3.45E-15 | 7.36E-11 |
| CIT | 0.621635482 | 4.81E-06 | 5.20E-05 | 1.730693483 | 3.64E-91 | 7.77E-87 |
| CDC25C | 0.61914164 | 1.45E-07 | 2.15E-06 | 2.273983231 | 1.07E-69 | 2.29E-65 |
| CEP55 | 0.61891591 | 7.99E-10 | 1.70E-08 | 2.99652056 | 2.40E-286 | 5.12E-282 |
| TGFBR2 | 0.614885355 | 2.82E-06 | 3.20E-05 | 0.858625057 | 2.47E-136 | 5.28E-132 |
| KIF23 | 0.608477676 | 2.16E-10 | 4.90E-09 | 2.633037163 | 2.51E-257 | 5.36E-253 |
| SEC11C | -0.604904372 | 2.73E-18 | 1.44E-16 | -0.282711963 | 6.27E-31 | 1.34E-26 |
| CMTM3 | 0.601618035 | 3.33E-05 | 0.00029999 | 2.666021342 | 8.66E-216 | 1.85E-211 |
| ETV3 | -0.597943348 | 1.95E-10 | 4.45E-09 | -0.305821616 | 1.50E-10 | 3.21E-06 |
| MEIS1 | 0.591076159 | 0.002468684 | 0.012830194 | 2.254435529 | 5.81E-195 | 1.24E-190 |
| CDC20 | 0.585297207 | 2.64E-13 | 8.98E-12 | 1.801284022 | 3.30E-211 | 7.04E-207 |
| ARL6 | 0.572340188 | 0.000597983 | 0.003818315 | 1.161102838 | 1.63E-83 | 3.47E-79 |
| EME1 | 0.562021264 | 0.000920448 | 0.005540283 | 1.424905039 | 2.43E-41 | 5.19E-37 |
| CORO7 | -0.554934338 | 2.52E-06 | 2.90E-05 | -0.57024629 | 1.58E-61 | 3.38E-57 |
| CRIP1 | 0.553843592 | 4.52E-08 | 7.29E-07 | 0.912976198 | 9.15E-53 | 1.95E-48 |
| VEGFA | -0.551890662 | 2.73E-09 | 5.37E-08 | -1.375142619 | 7.90E-239 | 1.69E-234 |
| AURKA | 0.546799724 | 3.56E-12 | 1.05E-10 | 1.830316785 | 8.41E-112 | 1.80E-107 |
| XYLT1 | -0.542372485 | 7.04E-09 | 1.28E-07 | -0.829050799 | 1.64E-55 | 3.51E-51 |
| MAP1LC3B | -0.541862879 | 1.37E-13 | 4.82E-12 | -0.696358405 | 1.27E-210 | 2.72E-206 |
| NAGK | 0.535433334 | 1.74E-08 | 2.98E-07 | 0.598672697 | 1.93E-57 | 4.13E-53 |
| PHF19 | 0.534278073 | 6.22E-09 | 1.15E-07 | 1.882016652 | 0 | 0 |
| TBC1D32 | -0.515205856 | 0.0150284 | 0.056379289 | -1.544213089 | 2.93E-21 | 6.25E-17 |
| CCNB2 | 0.514177353 | 1.48E-11 | 4.06E-10 | 3.152638657 | 0 | 0 |
| JUND | -0.512268692 | 0.006708739 | 0.029396565 | -0.936465279 | 0 | 0 |
| VASH2 | 0.5100522 | 0.020944239 | 0.073657877 | 2.375007384 | 1.32E-149 | 2.82E-145 |
| ARNTL2 | 0.502610453 | 3.60E-06 | 4.00E-05 | 4.050272217 | 5.54E-136 | 1.18E-131 |
| OSGEPL1 | 0.497625445 | 0.000438295 | 0.002911377 | 0.402617489 | 2.20E-19 | 4.70E-15 |
| MACF1 | 0.487239357 | 0.000300419 | 0.002087625 | 0.750819688 | 0 | 0 |
| RGS1 | -0.484451017 | 1.85E-07 | 2.68E-06 | -0.662425942 | 2.31E-254 | 4.94E-250 |
| ZMYND8 | 0.480430221 | 1.31E-05 | 0.000129485 | 0.294818578 | 9.72E-198 | 2.08E-193 |
| HEXIM1 | -0.477313461 | 1.20E-08 | 2.11E-07 | -0.866525515 | 2.40E-57 | 5.13E-53 |
| SQOR | 0.477265285 | 3.87E-06 | 4.26E-05 | 0.482535942 | 2.03E-108 | 4.33E-104 |
| TSTD1 | -0.47385841 | 1.66E-05 | 0.000161008 | -1.012818196 | 0 | 0 |
| RACGAP1 | 0.470076335 | 9.65E-12 | 2.71E-10 | 2.366925087 | 3.85E-228 | 8.21E-224 |
| TPX2 | 0.467952944 | 9.02E-10 | 1.90E-08 | 2.306901266 | 0 | 0 |
| CENPI | 0.467002156 | 0.001257423 | 0.007198012 | 1.477204705 | 2.10E-31 | 4.48E-27 |
| CCDC15 | 0.464602618 | 0.020901338 | 0.073566764 | 1.981154596 | 1.43E-111 | 3.06E-107 |
| GIT2 | 0.463346889 | 1.25E-06 | 1.54E-05 | 0.319963046 | 9.80E-116 | 2.09E-111 |
| MKI67 | 0.462703474 | 0.002180736 | 0.011562709 | 2.464828436 | 0 | 0 |
| ZNF180 | 0.461260029 | 0.000283009 | 0.001977753 | 0.327097717 | 3.43E-27 | 7.33E-23 |
| BRMS1L | 0.454979153 | 0.003209042 | 0.015991648 | 0.637418469 | 2.42E-86 | 5.17E-82 |
| STN1 | 0.45492312 | 0.000132997 | 0.001019979 | 0.545315138 | 1.27E-106 | 2.70E-102 |
| RIOX2 | 0.453646629 | 9.82E-09 | 1.76E-07 | 0.596333944 | 5.11E-220 | 1.09E-215 |
| WDSUB1 | 0.448723939 | 0.009579949 | 0.039367816 | 0.933224082 | 4.70E-90 | 1.00E-85 |
| ZNF714 | 0.446765847 | 0.001144126 | 0.006671182 | 1.809726182 | 1.37E-262 | 2.92E-258 |
| STX11 | -0.444802272 | 2.68E-05 | 0.000246954 | -0.448804856 | 2.45E-43 | 5.23E-39 |

| | | | | | | |
|---|---|---|---|---|---|---|
| EIF4E | -0.442199199 | 3.32E-05 | 0.000298648 | -1.042966335 | 9.07E-107 | 1.94E-102 |
| MIDN | -0.439829893 | 1.59E-05 | 0.000154455 | -0.498446679 | 7.16E-57 | 1.53E-52 |
| GPAT3 | -0.434772025 | 0.020644866 | 0.072767584 | -0.793274484 | 6.86E-81 | 1.47E-76 |
| TEDC1 | 0.434223646 | 0.00393242 | 0.018837541 | 1.700586771 | 1.64E-235 | 3.50E-231 |
| CDCA8 | 0.433620259 | 2.33E-07 | 3.32E-06 | 2.308397527 | 1.35E-260 | 2.88E-256 |
| ELAC1 | 0.430037574 | 0.002971358 | 0.01494916 | 0.272302714 | 1.62E-47 | 3.46E-43 |
| KIF4A | 0.428269513 | 2.85E-06 | 3.24E-05 | 2.247209078 | 1.72E-176 | 3.68E-172 |
| CHMP5 | 0.424937293 | 5.39E-05 | 0.000460496 | 0.45573438 | 2.16E-120 | 4.61E-116 |
| HOXB7 | 0.418823182 | 0.001094795 | 0.006426632 | 0.353872048 | 9.32E-27 | 1.99E-22 |
| PTTG1 | 0.417350544 | 1.27E-06 | 1.56E-05 | 2.500333653 | 0 | 0 |
| ETHE1 | 0.416450449 | 0.000767482 | 0.004744905 | 0.419572451 | 6.43E-74 | 1.37E-69 |
| ZNF134 | 0.414121926 | 0.000935402 | 0.00561468 | 0.402773464 | 1.93E-21 | 4.11E-17 |
| PRC1 | 0.414110837 | 6.14E-08 | 9.63E-07 | 2.104210214 | 0 | 0 |
| SLC25A37 | -0.413725944 | 0.00018463 | 0.00136285 | -0.327788771 | 5.00E-108 | 1.07E-103 |
| CBX4 | -0.413430778 | 2.12E-06 | 2.49E-05 | -0.679626279 | 2.50E-92 | 5.34E-88 |
| PDK1 | -0.411770497 | 2.34E-06 | 2.72E-05 | -0.699909145 | 9.86E-198 | 2.11E-193 |
| TENT5C | -0.397063071 | 1.98E-06 | 2.34E-05 | -1.662438698 | 0 | 0 |
| SRRM2 | -0.394769447 | 0.000193175 | 0.001416861 | -0.425819357 | 0 | 0 |
| ARPC1A | 0.391515792 | 9.36E-07 | 1.19E-05 | 0.521717641 | 2.22E-187 | 4.74E-183 |
| SGO2 | 0.391095057 | 0.001714773 | 0.009430053 | 1.94729937 | 7.30723090199204e-321 | 1.56060530373844e-316 |
| ZNF93 | 0.381434601 | 0.001978305 | 0.010642529 | 1.070284684 | 1.84E-70 | 3.92E-66 |
| GPR132 | 0.381352794 | 0.002357497 | 0.012341223 | 3.863545301 | 3.30E-236 | 7.05E-232 |
| GOLM1 | 0.380741924 | 8.55E-07 | 1.09E-05 | 0.278935764 | 9.14E-68 | 1.95E-63 |
| ANKRD33B | -0.378782991 | 3.80E-07 | 5.20E-06 | -0.642996765 | 7.47E-12 | 1.60E-07 |
| TPT1 | -0.378352856 | 2.27E-10 | 5.14E-09 | -0.614602609 | 0 | 0 |
| RNF14 | 0.377235316 | 2.04E-05 | 0.000192793 | 0.276632426 | 2.24E-100 | 4.78E-96 |
| ALCAM | 0.376616895 | 0.011396864 | 0.045276325 | 0.887967955 | 1.33E-274 | 2.84E-270 |
| EXO5 | 0.374128825 | 0.009030158 | 0.037401398 | 0.731354046 | 3.82E-12 | 8.15E-08 |
| APOBEC3B | 0.368122802 | 1.38E-06 | 1.69E-05 | 3.611632696 | 0 | 0 |
| HSP90AA1 | -0.367916983 | 3.14E-08 | 5.19E-07 | -0.357998727 | 1.11E-297 | 2.36E-293 |
| DLGAP5 | 0.366829943 | 6.95E-05 | 0.000576898 | 2.096493 | 7.27E-268 | 1.55E-263 |
| KLF3 | -0.366697516 | 0.000835104 | 0.005090286 | -1.932565569 | 0 | 0 |
| RHOG | 0.365462345 | 0.000301523 | 0.002093284 | 0.646200003 | 3.29E-23 | 7.03E-19 |
| CDKL5 | 0.362462114 | 0.015360595 | 0.057376926 | 0.287491575 | 1.23E-33 | 2.62E-29 |
| ARG2 | 0.355901164 | 0.023127158 | 0.079763593 | 0.756989677 | 3.05E-50 | 6.52E-46 |
| ZNF791 | 0.354580562 | 0.000841774 | 0.005123725 | 0.85764303 | 0 | 0 |
| JTB | -0.349729346 | 0.000658296 | 0.004148219 | -0.436865905 | 0 | 0 |
| LPIN1 | 0.348633932 | 5.05E-07 | 6.76E-06 | 1.009935629 | 0 | 0 |
| BUB1 | 0.343921298 | 1.27E-06 | 1.56E-05 | 1.917431182 | 4.15015142506647e-322 | 8.86347839851446e-318 |
| TTK | 0.34328982 | 0.000467039 | 0.003078686 | 2.732618224 | 1.07E-281 | 2.28E-277 |
| RALB | 0.342802664 | 2.68E-05 | 0.000246954 | 1.591530575 | 8.82E-148 | 1.88E-143 |
| HYLS1 | 0.337947284 | 0.002786263 | 0.014199444 | 0.62062617 | 7.40E-40 | 1.58E-35 |
| FOXM1 | 0.335734724 | 0.000102612 | 0.000814735 | 1.423635826 | 1.25E-96 | 2.67E-92 |
| NCAPH | 0.333832139 | 1.17E-05 | 0.000117438 | 2.813927719 | 2.00E-227 | 4.26E-223 |
| EMSY | 0.330199227 | 0.001804638 | 0.00983971 | 0.48464859 | 2.73E-70 | 5.84E-66 |
| SPAG5 | 0.32517188 | 2.19E-05 | 0.000205826 | 1.289127444 | 1.83E-128 | 3.91E-124 |
| DDIAS | 0.324757887 | 0.001178807 | 0.00683408 | 2.186745893 | 2.47E-212 | 5.26E-208 |
| MTFR2 | 0.323506493 | 0.004124897 | 0.019618469 | 1.857583693 | 7.91E-109 | 1.69E-104 |
| METTL18 | -0.319966812 | 0.005512276 | 0.024995045 | -0.642456422 | 9.50E-14 | 2.03E-09 |
| CASP6 | 0.319613147 | 0.009916424 | 0.040519754 | 0.495766672 | 8.30E-66 | 1.77E-61 |
| MED27 | 0.314697751 | 0.000437149 | 0.002904878 | 0.26699022 | 3.28E-52 | 7.02E-48 |
| JMY | -0.314115238 | 0.006953491 | 0.030315611 | -0.310570747 | 5.01E-170 | 1.07E-165 |
| STRADB | -0.312774864 | 0.00098373 | 0.005862094 | -0.509866809 | 1.50E-15 | 3.21E-11 |
| NSL1 | 0.312513973 | 0.009173042 | 0.037899473 | 0.282201765 | 2.06E-64 | 4.40E-60 |
| BIRC5 | 0.311541599 | 1.18E-05 | 0.000117636 | 2.55268575 | 0 | 0 |
| MAN1A1 | -0.31052835 | 0.006275008 | 0.027812723 | -0.315836679 | 5.42E-306 | 1.16E-301 |
| ZNF17 | 0.309776249 | 0.029938562 | 0.097595768 | 0.564620071 | 3.58E-30 | 7.64E-26 |
| PRR11 | 0.309754603 | 0.001580484 | 0.008794741 | 2.775994029 | 3.50786608547285e-322 | 7.49174959874437e-318 |
| FUT8 | 0.309004503 | 0.000343451 | 0.002345243 | 0.381302474 | 7.03E-286 | 1.50E-281 |
| NCKAP1 | -0.308309044 | 0.007952029 | 0.033751295 | -2.983578268 | 1.07E-130 | 2.29E-126 |
| RIN3 | 0.308290756 | 0.022654692 | 0.078588001 | 0.327095043 | 5.16E-149 | 1.10E-144 |
| TMSB4X | 0.305714011 | 1.51E-06 | 1.82E-05 | 0.90976914 | 0 | 0 |

| | | | | | | |
|---|---|---|---|---|---|---|
| NCAPG | 0.304415572 | 3.22E-05 | 0.000291244 | 2.607586159 | 0 | 0 |
| ZMYM2 | -0.304338715 | 0.000234687 | 0.001684938 | -0.411732024 | 7.94E-76 | 1.70E-71 |
| CCNB1 | 0.301569878 | 9.08E-06 | 9.21E-05 | 2.61060301 | 0 | 0 |
| ENTPD6 | -0.300724841 | 0.000300288 | 0.00208755 | -1.213946453 | 5.88E-56 | 1.26E-51 |
| ACTL6A | 0.298518189 | 0.000146091 | 0.00110813 | 0.533480612 | 5.80E-177 | 1.24E-172 |
| NSMF | 0.297343919 | 0.001894769 | 0.010260075 | 0.479244766 | 7.69E-55 | 1.64E-50 |
| NLRP2 | 0.296355898 | 0.000340426 | 0.00232826 | 3.920228433 | 0 | 0 |
| TUBA1B | 0.293313742 | 4.12E-05 | 0.000361345 | 0.659132995 | 1.44E-37 | 3.08E-33 |
| PTCH1 | 0.293136326 | 0.017614674 | 0.064082628 | 1.288135132 | 0 | 0 |
| CCDC91 | 0.290790759 | 0.014511637 | 0.054904527 | 0.771219411 | 0 | 0 |
| ARHGAP5 | -0.287504268 | 0.02239066 | 0.077890532 | -0.81009234 | 2.58E-86 | 5.51E-82 |
| BUB1B | 0.28705549 | 0.003962253 | 0.018943769 | 2.357876507 | 1.17E-262 | 2.50E-258 |
| SGO1 | 0.284726991 | 0.004289361 | 0.020261554 | 2.880703864 | 0 | 0 |
| HSPH1 | -0.274730628 | 0.001108422 | 0.006495618 | -0.251252973 | 7.93E-57 | 1.69E-52 |
| RNF5 | 0.266341623 | 0.003266351 | 0.016202634 | 0.27609703 | 4.79E-73 | 1.02E-68 |
| USP39 | 0.26558725 | 5.84E-05 | 0.000493937 | 1.139177362 | 7.03E-157 | 1.50E-152 |
| P4HA2 | 0.263847138 | 0.011661615 | 0.046019961 | 0.778969351 | 4.35E-74 | 9.29E-70 |
| KBTBD8 | -0.262196419 | 0.027512023 | 0.091409381 | -0.711424894 | 8.79E-13 | 1.88E-08 |
| WASHC3 | 0.260407411 | 0.008478035 | 0.03553101 | 0.550864685 | 8.40E-195 | 1.79E-190 |
| MYOF | 0.259920463 | 0.005430387 | 0.024697741 | 1.207492075 | 2.19E-08 | 0.000467756 |
| SNAPC5 | 0.257748633 | 0.006397451 | 0.028273287 | 0.761781951 | 6.07E-237 | 1.30E-232 |
| EXOSC3 | 0.257279666 | 0.00727752 | 0.031459015 | 1.044028548 | 4.73E-208 | 1.01E-203 |
| SDHAF2 | 0.254570309 | 0.002611156 | 0.013417624 | 0.500581406 | 1.77E-67 | 3.78E-63 |
| SPC25 | 0.254441262 | 0.009785905 | 0.04009998 | 2.747418046 | 2.10E-257 | 4.49E-253 |
| IFIH1 | 0.253871657 | 0.016514265 | 0.060687424 | 0.698713062 | 1.88E-274 | 4.02E-270 |
| NFE2L2 | -0.251527028 | 0.002278918 | 0.011991501 | -0.603293418 | 1.63E-76 | 3.49E-72 |
| GAS7 | 0.250099621 | 0.00325972 | 0.016188289 | 1.978198128 | 2.22E-09 | 4.74E-05 |
| PRIM1 | 0.249262553 | 0.009556732 | 0.039291056 | 0.875228541 | 4.88E-141 | 1.04E-136 |
| MEPCE | -0.249173538 | 0.002375001 | 0.012421591 | -0.625079018 | 2.94E-62 | 6.29E-58 |
| TTC5 | 0.248592721 | 0.01045089 | 0.042294476 | 0.402173406 | 2.84E-95 | 6.06E-91 |
| INSR | -0.247845775 | 0.004358914 | 0.020528508 | -0.853305693 | 1.55E-231 | 3.31E-227 |
| GTF2H3 | 0.246339037 | 0.013902641 | 0.053193116 | 0.666595546 | 2.67E-90 | 5.70E-86 |
| NDUFB5 | 0.245908567 | 0.002211072 | 0.011687969 | 0.556154812 | 2.94E-287 | 6.29E-283 |
| PBX2 | -0.243117474 | 0.007493138 | 0.03218221 | -1.136396052 | 3.17E-98 | 6.77E-94 |
| PTTG1IP | 0.241155183 | 0.002284746 | 0.012011214 | 1.202318621 | 0 | 0 |
| DDX24 | -0.240351556 | 0.000227906 | 0.001642383 | -0.889866647 | 0 | 0 |
| TXNDC11 | -0.239531627 | 0.00050235 | 0.003277765 | -0.331871414 | 2.04E-218 | 4.36E-214 |
| SSR4 | -0.238053967 | 0.009373475 | 0.038629336 | -2.131363247 | 0 | 0 |
| RANBP6 | -0.23393794 | 0.012356138 | 0.048235176 | -0.656105866 | 1.69E-73 | 3.60E-69 |
| ASF1B | 0.232256177 | 0.00844934 | 0.035427898 | 1.688010041 | 1.06E-235 | 2.26E-231 |
| LMO4 | -0.231669222 | 0.010069548 | 0.04101961 | -0.562456796 | 1.94E-264 | 4.13E-260 |
| SPECC1 | 0.231445371 | 0.01892537 | 0.067768949 | 0.874966473 | 5.54E-41 | 1.18E-36 |
| ZWINT | 0.227968318 | 0.001364664 | 0.00773548 | 2.132863684 | 0 | 0 |
| ITPR1 | 0.227551185 | 0.014906853 | 0.056020393 | 0.832081696 | 6.99E-152 | 1.49E-147 |
| TUBB | 0.22712492 | 0.000287719 | 0.002008231 | 0.590637256 | 1.18E-38 | 2.51E-34 |
| TSPAN3 | -0.224443321 | 0.001116623 | 0.006539248 | -0.743451786 | 5.30E-134 | 1.13E-129 |
| ANXA6 | 0.223451314 | 0.000471381 | 0.003097875 | 0.764570228 | 0 | 0 |
| HSD17B11 | -0.223443487 | 0.025196247 | 0.085369481 | -0.866617083 | 1.46E-167 | 3.12E-163 |
| CDK1 | 0.221058338 | 0.024263013 | 0.082855965 | 2.072129767 | 3.75E-197 | 8.00E-193 |
| DCAF7 | 0.220324438 | 0.000771054 | 0.004760191 | 0.883927117 | 9.88E-263 | 2.11E-258 |
| INO80E | -0.217864732 | 0.022785739 | 0.078900348 | -0.342401562 | 2.98E-39 | 6.36E-35 |
| NGRN | 0.211494595 | 0.005076691 | 0.023378866 | 1.090138273 | 2.67E-180 | 5.69E-176 |
| APEX2 | 0.209738552 | 0.016065311 | 0.059450865 | 0.381735332 | 1.84E-32 | 3.92E-28 |
| KBTBD4 | 0.208440106 | 0.017647614 | 0.064188979 | 0.449059821 | 1.25E-72 | 2.67E-68 |
| HAUS1 | 0.202533534 | 0.013770466 | 0.05278069 | 0.645570633 | 3.41E-220 | 7.28E-216 |
| ADAR | -0.202339863 | 0.000865901 | 0.005246606 | -0.533070596 | 5.09E-221 | 1.09E-216 |
| BUB3 | 0.201486562 | 0.002520603 | 0.013056917 | 1.18576582 | 0 | 0 |
| CSTF2 | 0.191048003 | 0.024832215 | 0.084439314 | 0.627544306 | 1.68E-27 | 3.59E-23 |
| COX6B1 | 0.18049419 | 0.005142481 | 0.023640121 | 0.436265576 | 6.40E-273 | 1.37E-268 |
| EXOSC8 | 0.179549176 | 0.027847498 | 0.092364514 | 0.639852845 | 8.22E-192 | 1.76E-187 |
| ANP32A | 0.173858091 | 0.025269963 | 0.085602494 | 0.80148512 | 0 | 0 |
| EEF1D | -0.169289462 | 0.018986792 | 0.067946739 | -0.52823342 | 0 | 0 |

| | | | | | | |
|---|---|---|---|---|---|---|
| NUDC | 0.166989933 | 0.01596413 | 0.059159178 | 0.393537926 | 8.76E-188 | 1.87E-183 |
| AKAP13 | 0.166160744 | 0.029320553 | 0.096090844 | 0.461951287 | 4.26E-306 | 9.11E-302 |
| SUN1 | -0.16420987 | 0.01697033 | 0.062116932 | -0.764122276 | 4.47E-93 | 9.54E-89 |
| PSMC5 | 0.154327961 | 0.016424139 | 0.060449906 | 0.610395344 | 5.77E-302 | 1.23E-297 |

**Supplementary Table S4**

| | Melflufen | | Melphalan | |
|---|---|---|---|---|
| **Cell line** | **EC50, μM** | **EC90, μM** | **EC50, μM** | **EC90, μM** |
| **AMO-1 *TP53* wt** | **0.1** | **0.5** | **9.0** | **≥20** |
| **AMO-1 *TP53* -/-** | **0.6** | **2.5** | **≥20** | **≥20** |

Cell lines were treated with either melflufen or melphalan at an 8-point drug concentration range to determine EC50 or EC90 values using a viability

**Supplementary Table S5**

Baseline demographics

| | Mel - del17p, N=33 | Mel - Not del17p, N=213 | Pom - del17p, N=37 | Pom - Not del17p, N=212 |
|---|---|---|---|---|
| **Age** | 68 (45-85) | 67 (41-91) | 69 (43-83) | 68 (39-87) |
| **Agegroup** | | | | |
| **<65** | 11 (33.3) | 85 (39.9) | 11 (29.7) | 74 (34.9) |
| **65-74** | 19 (57.6) | 94 (44.1) | 19 (51.4) | 106 (50.0) |
| **75+** | 3 (9.1) | 34 (16.0) | 7 (18.9) | 32 (15.1) |
| **Male** | 20 (61) | 119 (56) | 19 (51) | 121 (57) |
| **Race** | | | | |
| **WHITE** | 30 (90.9) | 194 (91.1) | 32 (86.5) | 190 (89.6) |
| **BLACK OR AFRICAN AMERICAN** | 0 (0.0) | 4 (1.9) | 0 (0.0) | 4 (1.9) |
| **ASIAN** | 1 (3.0) | 7 (3.3) | 5 (13.5) | 8 (3.8) |
| **NATIVE HAWAIIAN OR OTHER PACIFIC ISLANDER** | 0 (0.0) | 0 (0.0) | 0 (0.0) | 0 (0.0) |
| **OTHER** | 0 (0.0) | 1 (0.5) | 0 (0.0) | 0 (0.0) |
| **NOT REPORTED** | 0 (0.0) | 0 (0.0) | 0 (0.0) | 1 (0.5) |
| **UNKNOWN** | 2 (6.1) | 7 (3.3) | 0 (0.0) | 9 (4.2) |

Disease characteristics

| | Mel - del17p, N=33 | Mel - Not del17p, N=213 | Pom - del17p, N=37 | Pom - Not del17p, N=212 |
|---|---|---|---|---|
| **Years from diagnosis, median (range)** | 4.5 (1.5-21.5) | 3.9 (0.5-26.3) | 3.8 (1.2-12.5) | 3.9 (0.4-25.2) |
| **Prior treatment regimens, median (range)** | 3 (2-4) | 3 (2-4) | 2 (2-4) | 3 (2-4) |
| **Documented refractory status, n(%)** | | | | |
| **Lenalidomide** | 33 (100) | 213 (100) | 37 (100) | 212 (100) |
| **Pomalidomide** | 0 (0) | 0 (0) | 0 (0) | 0 (0) |
| **Bortezomib** | 15 (45) | 96 (45) | 22 (59) | 103 (49) |
| **Carfilzomib** | 9 (27) | 38 (18) | 9 (24) | 28 (13) |
| **Daratumumab** | 3 (9) | 45 (21) | 7 (19) | 31 (15) |
| **Alkylator refractory, n(%)** | 8 (24) | 70 (33) | 9 (24) | 66 (31) |
| **Melphalan exposed** | 11 (33) | 50 (23) | 17 (46) | 47 (22) |
| **Melphalan refractory** | 2 (6) | 13 (6) | 4 (11) | 19 (9) |
| **Previous stem cell transplant, n(%)** | 19 (58) | 106 (50) | 15 (41) | 105 (50) |
| **International Staging System at Baseline, n(%)** | | | | |
| **I** | 10 (30) | 109 (51) | 14 (38) | 110 (52) |
| **II** | 19 (58) | 75 (35) | 18 (49) | 76 (36) |
| **III** | 4 (12) | 29 (14) | 5 (14) | 26 (12) |
| **High-risk cytogenetics, n(%)** | 32 (97) | 51 (24) | 33 (89) | 53 (25) |
| **Extramedullary disease (EMD), n(%)** | 2 (6) | 28 (13) | 4 (11) | 22 (10) |
| **ECOG at baseline, n(%)** | | | | |
| **0** | 9 (27) | 81 (38) | 11 (30) | 81 (38) |
| **1** | 17 (52) | 113 (53) | 22 (59) | 114 (54) |
| **2** | 7 (21) | 19 (9) | 4 (11) | 17 (8) |
| **TTP following ASCT** | | | | |
| **No transplant** | 14 (42) | 107 (50) | 22 (59) | 107 (50) |
| **<1y** | 4 (12) | 27 (13) | 6 (16) | 26 (12) |
| **1-2y** | 8 (24) | 39 (18) | 4 (11) | 47 (22) |
| **2-3y** | 4 (12) | 19 (9) | 1 (3) | 17 (8) |
| **>3y** | 3 (9) | 21 (10) | 4 (11) | 15 (7) |

### Supplementary Table S6

| | | del(17p) | del(17p); t(4:14) | del(17p); t(14:16) | del(17p); t(4:14); t(14:16) | del(17p); t(14:20) | del(17p); gain 1q (+1q) | del(17p); t(4;14); gain 1q (+1q) | del(17p); t(4;16); gain 1q (+1q) |
|---|---|---|---|---|---|---|---|---|---|
| MflDex | responders | 8 (2 (VGPR), 6 (PR)) | 1 (VGPR) | 0 | 0 | 0 | 2 (1 (VGPR), 1 (PR)) | 0 | 0 |
| | non-responders | 8(2 (SD), 1 (PD), 1 (NE), 4 (MR)) | 0 | 0 | 0 | 0 | 10 (5 (SD), 3 (PD), 2 (MR)) | 1 (SD) | 0 |
| | non -treated | 1 (NE) | 0 | 0 | 0 | 0 | 2 (NE) | 0 | 0 |
| PomDex | responders | 2 (VGPR) | 0 | 0 | 0 | 0 | 2 (1 (VGPR), 1 (PR)) | 0 | 0 |
| | non-responders | 17 (5 (SD), 8 (PD), 2 (MR), 2 (NE)) | 4 (2 (SD), 2 (MR)) | 0 | 3 (SD) | 1 (PD) | 5 (2 (SD), 3 (PD)) | 1 (NE) | 1 (PD) |
| | non -treated | 0 | 1 (PD) | 0 | 0 | 0 | 0 | 0 | 0 |

**Supplementary Table S7**

### Baseline demographics

| | Melflufen - TP53 mutant, N=15 | Melflufen - TP53 wild type , N=62 | Pomalidomide - TP53 mutant, N=18 | Pomalidomide - TP53 wild type, N=50 |
|---|---|---|---|---|
| **Age** | 66 (51-80) | 65 (41-84) | 68 (43-83) | 68 (46-80) |
| **Agegroup** | | | | |
| **<65** | 6 (40.0) | 28 (45.2) | 6 (33.3) | 14 (28.0) |
| **65-74** | 8 (53.3) | 27 (43.5) | 9 (50.0) | 27 (54.0) |
| **75+** | 1 (6.7) | 7 (11.3) | 3 (16.7) | 9 (18.0) |
| **Male** | 6 (40) | 32 (52) | 9 (50) | 25 (50) |
| **Race** | | | | |
| **WHITE** | 15 (100.0) | 55 (88.7) | 15 (83.3) | 44 (88.0) |
| **BLACK OR AFRICAN AMERICAN** | 0 (0.0) | 0 (0.0) | 1 (5.6) | 1 (2.0) |
| **ASIAN** | 0 (0.0) | 4 (6.5) | 2 (11.1) | 3 (6.0) |
| **NATIVE HAWAIIAN OR OTHER PACIFIC ISLANDER** | 0 (0.0) | 0 (0.0) | 0 (0.0) | 0 (0.0) |
| **OTHER** | 0 (0.0) | 1 (1.6) | 0 (0.0) | 0 (0.0) |
| **NOT REPORTED** | 0 (0.0) | 0 (0.0) | 0 (0.0) | 0 (0.0) |
| **UNKNOWN** | 0 (0.0) | 2 (3.2) | 0 (0.0) | 2 (4.0) |

### Disease characteristics

| | Melflufen - TP53 mutant, N=15 | Melflufen - TP53 wild type , N=62 | Pomalidomide - TP53 mutant, N=18 | Pomalidomide - TP53 wild type, N=50 |
|---|---|---|---|---|
| **Years from diagnosis, median (range)** | 4.5 (1.5-8.4) | 4.6 (0.5-17.8) | 4.3 (1.2-12.5) | 4.4 (0.4-13.7) |
| **Prior treatment regimens, median (range)** | 3 (2-4) | 3 (2-4) | 3 (2-4) | 3 (2-4) |
| **Documented refractory status, n(%)** | | | | |
| **Lenalidomide** | 15 (100) | 62 (100) | 18 (100) | 50 (100) |
| **Pomalidomide** | 0 (0) | 0 (0) | 0 (0) | 0 (0) |
| **Bortezomib** | 11 (73) | 30 (48) | 10 (56) | 22 (44) |
| **Carfilzomib** | 0 (0) | 6 (10) | 3 (17) | 7 (14) |
| **Daratumumab** | 1 (7) | 14 (23) | 2 (11) | 7 (14) |
| **Alkylator refractory, n(%)** | 6 (40) | 25 (40) | 6 (33) | 8 (16) |
| **Melphalan exposed** | 5 (33) | 11 (18) | 7 (39) | 15 (30) |
| **Melphalan refractory** | 2 (13) | 5 (8) | 2 (11) | 3 (6) |
| **Previous stem cell transplant, n(%)** | 9 (60) | 33 (53) | 10 (56) | 29 (58) |
| **International Staging System at Baseline, n(%)** | | | | |
| **I** | 6 (40) | 37 (60) | 4 (22) | 31 (62) |
| **II** | 7 (47) | 18 (29) | 13 (72) | 15 (30) |
| **III** | 2 (13) | 7 (11) | 1 (6) | 4 (8) |
| **High-risk cytogenetics, n(%)** | 13 (87) | 16 (26) | 14 (78) | 18 (36) |
| **Extramedullary disease (EMD), n(%)** | 1 (7) | 14 (23) | 2 (11) | 4 (8) |
| **ECOG at baseline, n(%)** | | | | |
| **0** | 6 (40) | 21 (34) | 4 (22) | 20 (40) |
| **1** | 6 (40) | 32 (52) | 12 (67) | 26 (52) |
| **2** | 3 (20) | 9 (15) | 2 (11) | 4 (8) |
| **TTP following ASCT** | | | | |
| **No transplant** | 6 (40) | 29 (47) | 8 (44) | 21 (42) |
| **<1y** | 3 (20) | 4 (6) | 4 (22) | 6 (12) |
| **1-2y** | 4 (27) | 15 (24) | 3 (17) | 19 (38) |
| **2-3y** | 1 (7) | 7 (11) | 2 (11) | 3 (6) |
| **>3y** | 1 (7) | 7 (11) | 1 (6) | 1 (2) |
